# A seven-year longitudinal study of the Alzheimer’s disease blood metabolome

**DOI:** 10.64898/2025.12.05.25341709

**Authors:** Bharadwaj Marella, Patrick Weinisch, Josef J. Bless, Shannon L. Risacher, Colette Blach, Naama Karu, Karel Kalecký, Tingting Wang, Kevin Huynh, Alexandra Kueider-Paisley, J. Will Thompson, Alzheimer’s Disease Neuroimaging Initiative, Alzheimer’s Disease Metabolomics Consortium, Teodoro Bottiglieri, Kwangsik Nho, P. Murali Doraiswamy, Andrew J. Saykin, Peter J. Meikle, Gabi Kastenmüller, Rima Kaddurah-Daouk, Matthias Arnold

## Abstract

**INTRODUCTION:** Metabolic dysregulation is a hallmark of Alzheimer’s disease (AD), but its temporal relationship to disease phenotypes remains unclear.

**METHODS:** We analyzed 506 serum metabolites in 4,063 longitudinal samples from 1,430 Alzheimer’s Disease Neuroimaging Initiative participants using cross-sectional single-timepoint analysis, joint multi-timepoint analysis, and time-interaction analysis.

**RESULTS:** Across 15 AD-related phenotypes, 311 metabolites were significantly associated with disease. Joint multi-timepoint analysis identified 281 metabolites, 243 in cross-sectional analyses, and 19 in time-interaction analysis. Associated metabolites implicated fatty acid and energy metabolism, neurotransmitter pathways, and oxidative stress regulation. Overall, 128 metabolites showed persistent associations over time, consistent with chronic systemic metabolic disruption. Only a small subset, including betaine, choline, and few amino acids, showed steeper declines with worsening AD endophenotypes.

**DISCUSSION:** These findings indicate that most metabolic alterations in AD remain stable once established, while fewer continue to change during progression, clarifying the timing of metabolic dysregulation across the AD trajectory.

## 1. Background

Alzheimer’s disease (AD) is characterized by the accumulation of amyloid-beta plaques and neurofibrillary tau tangles in the brain^1^. Further hallmarks of the disease are dysregulated glucose uptake, impaired cholesterol transport via Apolipoprotein E, and neuroinflammation, leading to neurodegeneration and ultimately dementia.

Previous studies have demonstrated that multiple metabolic pathways are altered during the onset and progression of AD, highlighting disruptions in lipid metabolism, amino acid catabolism, energy pathways, neurotransmission, and oxidative stress response, among others. Trushina et al.^2^ identified alterations in mitochondrial and lipid metabolism in plasma and cerebrospinal fluid (CSF) of individuals with mild cognitive impairment (MCI). Han et al.^3^ demonstrated alterations in sphingolipid and sterol metabolism linked to cognitive decline. Toledo et al.^4^ showed that changes particularly in acylcarnitines, sphingomyelins, and phosphatidylcholines, are associated with AD progression and clinical outcomes. MahmoudianDehkordi et al.^5^ suggested that the gut-brain axis may contribute to AD through altered bile acid metabolism. Another study showed that 18 plasma metabolites, such as amino acids and lipids, were able to predict future dementia risk^6^. Moreover, brain metabolomics studies in humans revealed region-specific changes. Ambeskovic et al.^7^ found increased levels of post-mortem hippocampal N-acetylaspartate, gamma-aminobutyric acid (GABA), and leucine in AD. In contrast, Batra et al.^8^ showed that lower GABA levels in the dorsolateral prefrontal cortex (DLPFC) are associated with AD traits. Disruptions in neurotransmitter systems have been generally well documented in AD^9^, including neurotransmitter level changes in blood^10^. Finally, studies have suggested the role of oxidative stress as a potential driver of neurodegeneration in AD. Knörnschild et al.^11^ reported disrupted glucose oxidation and elevated oxidative stress metabolites (e.g., L-cysteine) in AD mouse models, while Ahmad et al.^12^ demonstrated strong associations between CSF isoprostanes and tau pathology in MCI patients, and Batra et al.^8^ showed disruptions of the glutathione pathway, among others. Dronse et al.^13^ found that elevated morning serum cortisol levels correlated with poorer verbal memory in AD patients. Mosconi et al.^14^ found that higher serum cortisol was linked to lower total brain volume, reduced glucose metabolism in the frontal cortex and increased amyloid-ß burden in regions vulnerable to AD.

Despite the large body of consistent links between AD and metabolic dysregulation, some findings vary across tissues and disease stages. For example, decreased lysine levels have been observed in post mortem hippocampal tissue^15^ and plasma samples of AD patients^16^, while elevated lysine levels have been found in DLPFC^8^ and urine of AD patients^17^. Similarly, lower plasma ceramide levels were observed in patients with MCI compared to cognitively normal individuals and AD patients, whereas higher levels were predictive of greater cognitive decline and greater hippocampal atrophy in MCI patients, suggesting a dynamic evolution of metabolic changes across disease stages^18^.

Metabolic signatures are unique to the individual and signature shifts can reflect disease states and underlying molecular mechanisms, especially in complex, progressive diseases such as AD. While cross-sectional metabolomics captures inter-individual differences at a single time point, it cannot track intra-individual changes over time. Longitudinal metabolomics, however, uses repeated measurements from the same individuals to study temporal trajectories of metabolites across disease progression. This dynamic insight is especially important in the prodromal stage of AD, when subtle metabolic alterations may precede clinical symptoms, and the contribution of biological pathways may shift as disease advances. Notably, Huang et al.^19^ observed changes in pathways including glutathione, tryptophan, and bile acid metabolism in individuals with physio-cognitive decline compared to normal aging controls. Furthermore, contrasting cross-sectional and longitudinal data permits assessment of the temporal robustness of metabolite–phenotype associations^20^.

In this study, we utilized data from the Alzheimer’s Disease Neuroimaging Initiative (ADNI). ADNI provides longitudinal in-depth phenotypic assessments and metabolite profiles generated by the AD metabolomics consortium collected every 6 to 12 months, enabling evaluation of metabolite associations with AD onset, severity, and progression. Our prior work in ADNI showed a significant 3–4.8% reduction in ether lipid species in individuals who progressed to AD^21^. Building on these findings, we extended our investigation to polar metabolites and adopted a broader analytical approach to evaluate the reproducibility of such associations.

To achieve this, we contrasted metabolic alterations observed in cross-sectional and longitudinal analyses. More specifically, we conducted cross-sectional analyses to identify metabolic differences in AD at single time points, complemented by two types of longitudinal analyses: a joint multi-timepoint analysis examining the persistence of metabolic differences over time; and a time-interaction model to identify metabolites that show a changing relationship to the disease over time. By combining results across analyses, we then categorized metabolites by their association patterns with disease, highlighting those that are most consistently linked to AD or are continuing to change along the disease trajectory. Our findings provide a first reference to prioritize metabolic pathways not only cross-sectionally, but longitudinally, allowing for a better understanding of metabolic disruptions in AD and a more solid basis for potential novel, metabolically-guided therapeutic strategies.

## 2. Methods

### 2.1. Study population and diagnostic classification

Data used in the preparation of this article were obtained from the Alzheimer’s Disease Neuroimaging Initiative (ADNI) database (adni.loni.usc.edu). The ADNI was launched in 2003 as a public-private partnership, led by Principal Investigator Michael W. Weiner, MD. The primary goal of ADNI has been to determine whether serial magnetic resonance imaging (MRI), positron emission tomography (PET), biological markers, and clinical and neuropsychological assessments can be combined to measure the progression of mild cognitive impairment (MCI) and early Alzheimer’s disease (AD). For up-to-date information, see www.adni-info.org. Written informed consent was obtained at enrollment, which included permission for analysis and data sharing. Consent forms were approved by each participating site’s institutional review board.

The definition of probable AD in ADNI followed the NINDS-ADRDA criteria. Specifically, individuals with Mini-Mental State Exam (MMSE) scores between 20 and 26 (inclusive) and a Clinical Dementia Rating Scale (CDR) of 0.5 or 1.0 were classified as patients with mild AD and were followed up for a maximum of 2 years into the study^22^. Participants were defined as MCI if they had MMSE scores between 24 and 30, a subjective memory complaint, objective memory loss measured by education-adjusted scores on Wechsler Memory Scale Logical Memory II, a CDR of 0.5, absence of significant levels of impairment in other cognitive domains, and essentially preserved activities of daily living^23^.

In this study, we leveraged data from 1524 ADNI-1, -GO, and -2 participants. Clinical data and biological samples were collected at baseline and every 6-12 months for a maximum of 10 years (maximum number of visits = 13). To group participants into a general longitudinally valid diagnostic group, we screened the diagnoses reported for each individual across all visits where metabolomics data was available, resulting in five longitudinal diagnostic classifications: cognitively normal (CN; *n*=383), stable MCI (*n*=423), MCI converters (CN to MCI; *n*=66), AD converters (CN or MCI to AD; *n*=294), reflecting progression to AD dementia at any time point during the course of the study, and AD (*n*=264). Individuals exhibiting atypical diagnostic trajectories—such as reversion from MCI or AD to CN, or fluctuation between stages—were excluded from this classification to ensure consistency in group assignment (*n*=72). As an additional outcome variable, the diagnostic status recorded at each visit was numerically encoded as CN = 0, MCI = 1, and AD = 2 to capture time-resolved diagnostic trajectories for all participants.

### 2.2. Phenotypic assessments

The available quantitative longitudinal outcomes included: (1) concentrations of amyloid-beta (Aβ_1-42_) and tau (phosphorylated (p-)tau and total (t-)tau) in CSF, (2) measures of neurodegeneration (glucose uptake measured by Fluorodeoxyglucose-Positron Emission Tomography (FDG-PET), grey matter volume and thickness, entorhinal volume and thickness, and hippocampal volume), and (3) neurocognitive scores (cognition subscale of the 13-item AD assessment scale - ADAS-Cog. 13, ADNI composite scores on memory, executive function, visospatial functioning and language). In total, fifteen clinical phenotypes and diagnostic group comparisons were selected for association analysis, representing diverse readouts that capture various aspects of AD. These phenotypes were defined based on the ATN framework^24^—which classifies AD by amyloid, tau, and neurodegeneration biomarkers—and were extended to include cognitive assessments and diagnostic status. Comprehensive descriptions of these phenotypes and their transformations, along with the corresponding covariates specific to each phenotype, are provided in **Supplementary Table S1**.

Aβ_1-42_, t-tau, and p-tau_181p_ in CSF samples were measured via a fully automated Roche Elecsys immunoassay applying a sample preparation procedure with two incubation steps^25,26^.

MRI protocol and data were accessed through LONI (https://adni.loni.usc.edu/). In brief, MRI imaging was acquired using a 1.5 T or 3 T MRI scanner with the sequences of T1 and dual echo T2-weighted imaging (ADNI-1), or a 3 T MRI scanner with fully sampled and accelerated T1-weighted imaging in addition to 2D FLAIR and T2*-weighted imaging (ADNI-GO/2). To process MRI scans and extract whole brain and ROI (region of interest)-based neuroimaging measurements determined by automated segmentation and parcellation including volumes and cortical thickness, we used FreeSurfer V6 (https://surfer.nmr.mgh.harvard.edu/fswiki)^27–29^. After the cortical surface was reconstructed, the cortical thickness at each vertex was calculated by taking the Euclidean distance between the grey/white boundary and the grey/CSF boundary at each vertex on the surface.

For FDG-PET, we used a ROI–based measure representing the average glucose uptake across the left and right angular gyrus, left and right temporal lobes, and bilateral posterior cingulate cortex. These values were derived from preprocessed scans (including co-registration, averaging, spatial standardization, and resolution harmonization) and intensity-normalized using a pons region to calculate standard uptake value ratio (SUVR) means^30,31^.

### 2.3. Metabolomics profiling and data preprocessing

Serum metabolites were quantified using the MXP® Quant 500 platform (Biocrates Life Sciences AG), a targeted, kit-based metabolomics assay. Sample preparation and metabolomics profiling were conducted by the Alzheimer’s Disease Metabolomics Consortium at Duke University using 10 µL aliquots resulting in 624 metabolites belonging to 26 biochemical classes. All procedures, including sample extraction, metabolite detection, identification, quantification, and initial quality control (QC), were performed in accordance with standardized protocols^4,32^. In addition to individual metabolite measurements, the dataset included vendor-defined biologically relevant sums and ratios reflecting functions such as enzyme activity. These composite measures (from the MetaboINDICATOR^TM^ software) were derived from raw concentrations and processed in the same manner as single metabolites. Furthermore, we have aggregated metabolites from the 26 biochemical classes into 11 categories to enhance the comprehension of the associations observed across the various classes (see **Supplementary Table S2**)

A comprehensive quality control procedure was applied to exclude unreliably measured metabolites from the dataset. First, 98 metabolites were removed due to having greater than 30% missingness in at least one cohort (ADNI-1 and ADNI-GO/2 were profiled in two separate runs). Next, an additional 8 metabolites were excluded based on a coefficient of variation (CV) exceeding 30% in quality control reference materials. A further 12 metabolites were removed due to complete missingness across one or more assay plates. In total, 118 metabolites were removed from further analysis. Details of the quality control assessments are provided in **Supplementary Table S3**).

The resulting dataset comprised 506 metabolites across 26 biochemical classes and 4,884 samples from 1,524 subjects. Biological replicates were averaged (n=41), and samples with more than 30% missing data were excluded (n=10). Multivariate outlier detection using Mahalanobis distance identified outliers with *D*^2^ >1.5 × max(D^2^_expected_), leading to the exclusion of 168 samples (6 subjects). Following log transformation and removal of 284 non-fasting samples (15 subjects), missing metabolite values were imputed using random forest–based imputation^33^. Subjects with atypical diagnostic trajectories were removed (subjects = 72; samples = 270). The final three timepoints (48 samples) were excluded to minimize bias. After all filtering steps, the final dataset included 4,063 samples from 1,430 subjects, with approximately 80% of samples collected at baseline, 12 months, and 24 months (hereafter referred to as three main timepoints). The decision to limit timepoints to a maximum of 24 months here was further based on the fact that participants who entered the ADNI studies with mild AD at baseline per study protocol dropped out after 24 months.

### 2.4. Identification of significant medication effects

To account for the influence of medications on the metabolome, we employed the Boruta feature selection algorithm^34,35^. The procedure was iterated 100 times to refine the set of selected and rejected features, and those exceeding 4 standard deviations above the mean importance were retained as covariates in subsequent regression models. A total of 38 and 20 significant medication–metabolite associations were identified at baseline and longitudinally at the three main timepoints, respectively. Several of these medications are commonly used in the treatment of Alzheimer’s disease, although AD-specific anti-dementia drugs were excluded from the analysis (**Supplementary Figure S1; Supplementary Tables S4** and **S5**).

### 2.5. Statistical analysis

Single-timepoint cross-sectional analyses were performed to examine associations between metabolite levels and disease phenotypes at each of the three main timepoints. In parallel, two mixed-effects models for repeated measures (MMRMs) were employed to assess metabolite–phenotype associations longitudinally. The first model incorporated the three main timepoints, with subject and time specified as random effects (except for time-resolved diagnostic status, where we used time as a fixed effect), to evaluate the consistency of associations across time (joint multi-timepoint analysis). The second model included all timepoints, subjects as a random effect, and a time-by-phenotype interaction term to assess temporal variation in associations (time-interaction analysis). To check for potential bias from the additional timepoints, we conducted a sensitivity analysis using only the three main timepoints as well. We further performed paired *t*-tests, as an additional line of evidence, in individuals who transitioned from CN to MCI or from MCI to AD within six to twelve months, with analyses restricted to transitions involving ≥30 individuals to ensure adequate power. To rule out spurious associations from imputation, we performed another sensitivity analysis on non-imputed data (**Supplementary Table S6**).

All regression models were adjusted for sex, age, education, BMI, APOE ε4 status, relevant medications, and phenotype-specific covariates (see section 2.2 and **Supplementary Table S1**). For lipids, we further included log_2_-transformed levels of total cholesterol and HDL cholesterol as covariates. All analyses were performed using the MeTime R package^36^.

For replication of lipid associations, we performed identical analyses on lipid species also available in independently generated lipidomics data^21^ from a largely overlapping set of longitudinal ADNI samples. For this, we first mapped lipid species with significant associations from the MXP® Quant 500 platform to the lipidomics dataset. Given the greater analytical resolution of the lipidomics platform compared to the MXP® Quant 500, e.g. for phosphatidylcholines, and the ambiguity of triglyceride annotations in both platforms, this resulted in several 1:n mappings (see **Supplementary Table S7**). Consequently, replication was defined as follows: if any lipid species in the lipidomics dataset that mapped to a given lipid from the MXP® Quant 500 platform demonstrated a significant association with AD in the same direction, the corresponding lipid in the MXP® Quant 500 platform was considered replicated.

The Li and Ji method^37^ was used for multiple hypothesis testing correction, which in a Bonferroni-like fashion adjusts for the effective number of independent metabolic features among the 506 metabolites at baseline (*n*=122 independent features). A similar correction method was applied to the replication analysis in the lipidomics dataset, where we adjusted for 120 unique lipid species that could be mapped to significant lipids from the MXP® Quant 500 (n= 35 independent features). This approach was chosen because both the metabolites, phenotypes and repeated measures per individual analyzed in this study exhibit strong correlation structures and the conducted association tests are therefore not mutually independent. Because of the strong correlation of the investigated phenotypes (median Spearman correlation = 0.4), no additional correction for the number of phenotypes was applied. Furthermore, this global significance threshold facilitated comparisons across the different analysis types employed in this study. Less stringent false discovery rate (FDR)-adjusted q-values were also calculated and are reported for all analyses in the respective Supplementary Tables.

Beyond single metabolite tests, we conducted p-gain analysis^38^ to assess whether associations were driven primarily by individual metabolites or by informative sums and ratios (see Section 2.3).

### 2.6. Bootstrapping analysis

To assess the robustness of significant time-interaction associations, we used participant-level nonparametric bootstrapping. In 1,000 iterations, ADNI participants were sampled with replacement while retaining all longitudinal observations for each selected participant, and the original models were refitted using the same specifications and covariates. For each association, bias was evaluated from the difference between the original effect estimate and the mean bootstrap effect estimate, and its magnitude was summarized using the absolute difference. Studentized bootstrap 95% confidence intervals were calculated using the effect estimate and standard error from each iteration. Shapiro-Wilk tests were used to assess the normality of the effect estimate distribution across bootstraps. Summary *p* values were computed based on the mean effect estimate and mean standard error across all bootstraps.

## 3. Results

### 3.1. Cohort characteristics

In this study, we leveraged longitudinal metabolomics data generated on 4063 blood samples as well as longitudinal phenotypic data from 1430 participants of the ADNI-1, -GO and -2 cohorts. 1293 participants had metabolomics data available starting at baseline (**Table 1**). Blood samples were collected at annual or biannual follow-ups after study enrollment, spanning a total of 10 time points and seven years, respectively (**Figure 1**). The majority of the data (approximately 80%) was concentrated at three main timepoints: baseline, 12 months, and 24 months.

**Figure 1:**
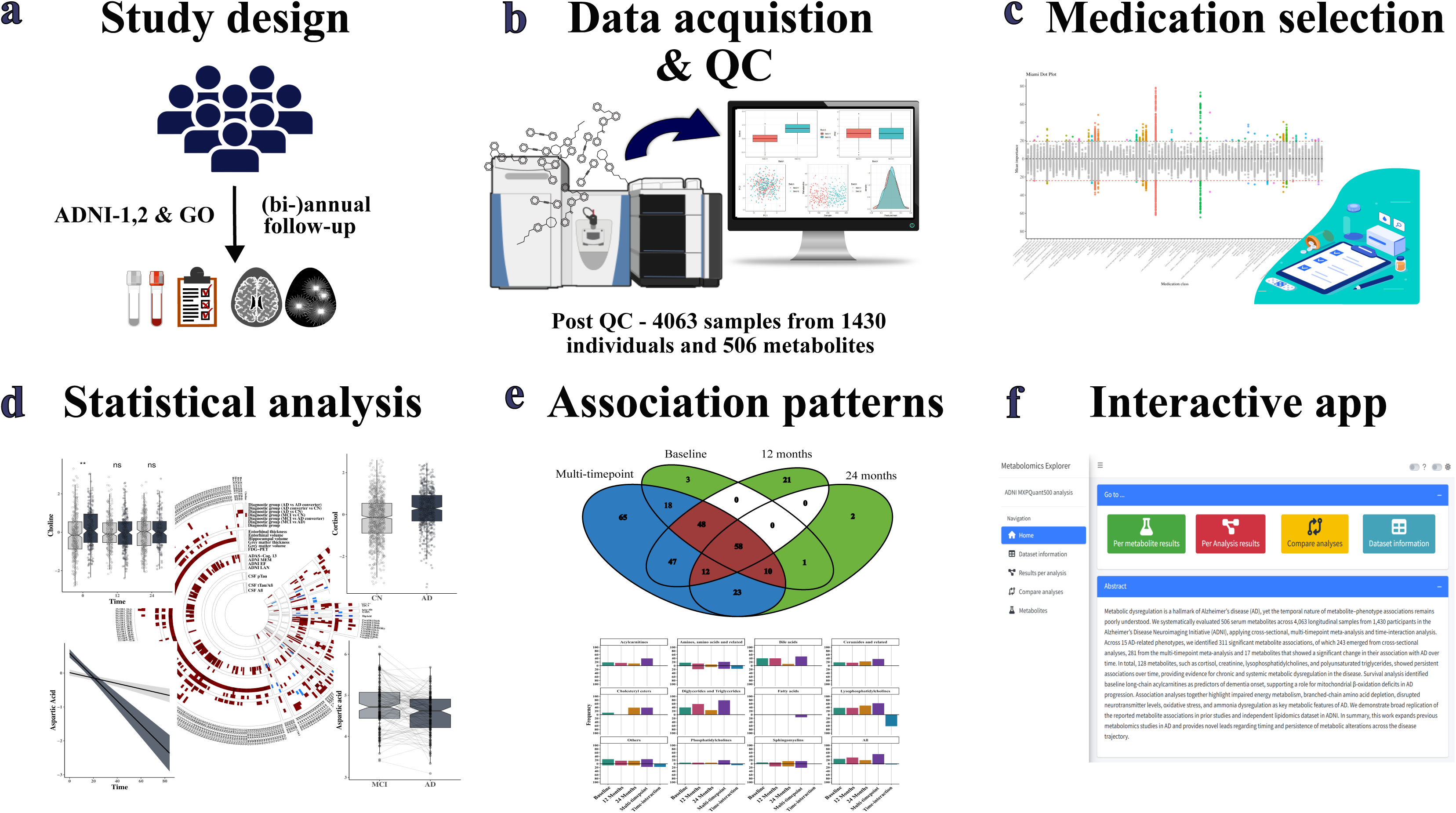
Analysis workflow. **(a)** Participants from the ADNI-1, -GO, & -2 studies were followed up annually or biannually, and blood and CSF samples were collected along with cognitive assessments and neuroimaging. **(b)** Metabolite concentrations were measured on the MXP® Quant 500 platform. After rigorous quality control, the final dataset consisted of 4,063 samples from 1,430 participants. **(c)** Feature selection analysis was performed using boruta backward random forest method to identify medications that are associated with metabolite levels and later used as covariates in downstream analyses. **(d)** The different types of statistical analyses conducted in this study, covering cross-sectional single timepoint analysis, joint multi-timepoint analysis and time-interaction analysis. **(e)** Assessing persistence of associations observed and identifying the patterns of associations found for the different metabolites investigated in this study. **(f)** All study outcomes are available through an interactive web application at https://metabolomics.helmholtz-munich.de/mxtra_ad/.

**Table 1:** Characteristics of the study cohort. Levels of high-density lipoprotein (HDL) cholesterol and total cholesterol are log_2_-transformed and limited to baseline. *CN – cognitively normal controls; MCI – participants with mild cognitive impairment; AD – participants with clinical Alzheimer’s disease; CN-MCI converter - CN to MCI converters; AD converters - CN or MCI to AD converters; BMI - body mass index*.

### 3.2 Identification of persistent, latent, and episodic associations

We first conducted cross-sectional analyses to identify serum metabolites associated with disease phenotypes at each of the three main timepoints separately. We obtained 138 significant associations (at a Bonferroni-adjusted p < 4.1 × 10 ) at baseline, 186 significant associations at 12 months, and 106 significant associations at 24 months, totaling to 243 unique metabolites (48.02%) that were significant in at least one analysis (**Supplementary Table S8; Supplementary Figure S2**). Cortisol was the metabolite associated with the largest number of phenotypes at baseline and at 24 months. At 12 months, triglyceride (TG) (18:1/34:4) showed the highest number of phenotype associations. Associated metabolites covered 10 of the 26 metabolite classes assessed here at baseline and 24 months, and 9 classes at 12 months (**Supplementary Table S9**). Among the tested phenotypes, hippocampal volume (n=98 associations) showed the highest number of associations at baseline, and ADAS-Cog. 13 at 12 months (n=119) and 24 months (n=70) **(Supplementary Table S10).**

Next, on top of the cross-sectional analyses, we employed linear mixed-effects models referred to as joint multi-timepoint analysis that incorporated the three main timepoints simultaneously, modeling time and subject as random effect variables to test the persistence of associations and their effect directions over time. Here, we discovered 281 out of 506 metabolites (55.53%) with significant associations. The majority of metabolites (94.31%) showed higher levels with more advanced disease phenotypes and diagnostic status, while 15 metabolites showed negative associations. This was consistent with our observations in the single timepoint cross-sectional analyses (**Figure 2; Supplementary Figure S2**). Cortisol was the metabolite associated with the largest number of phenotypes in the joint multi-timepoint analysis. Overall, metabolites from 20 of the 26 metabolite classes were associated with disease phenotypes in this joint multi-timepoint analysis, with triglycerides being the most frequently associated in absolute numbers, and fatty acids the least **(Supplementary Table S9).** Among the tested phenotypes, hippocampal volume (n=203 associations), ADAS-Cog 13 score (n=109), and the composite memory score (n=129) exhibited the largest number of significant metabolite associations in the joint multi-timepoint analysis **(Supplementary Table S10).**

**Figure 2:**
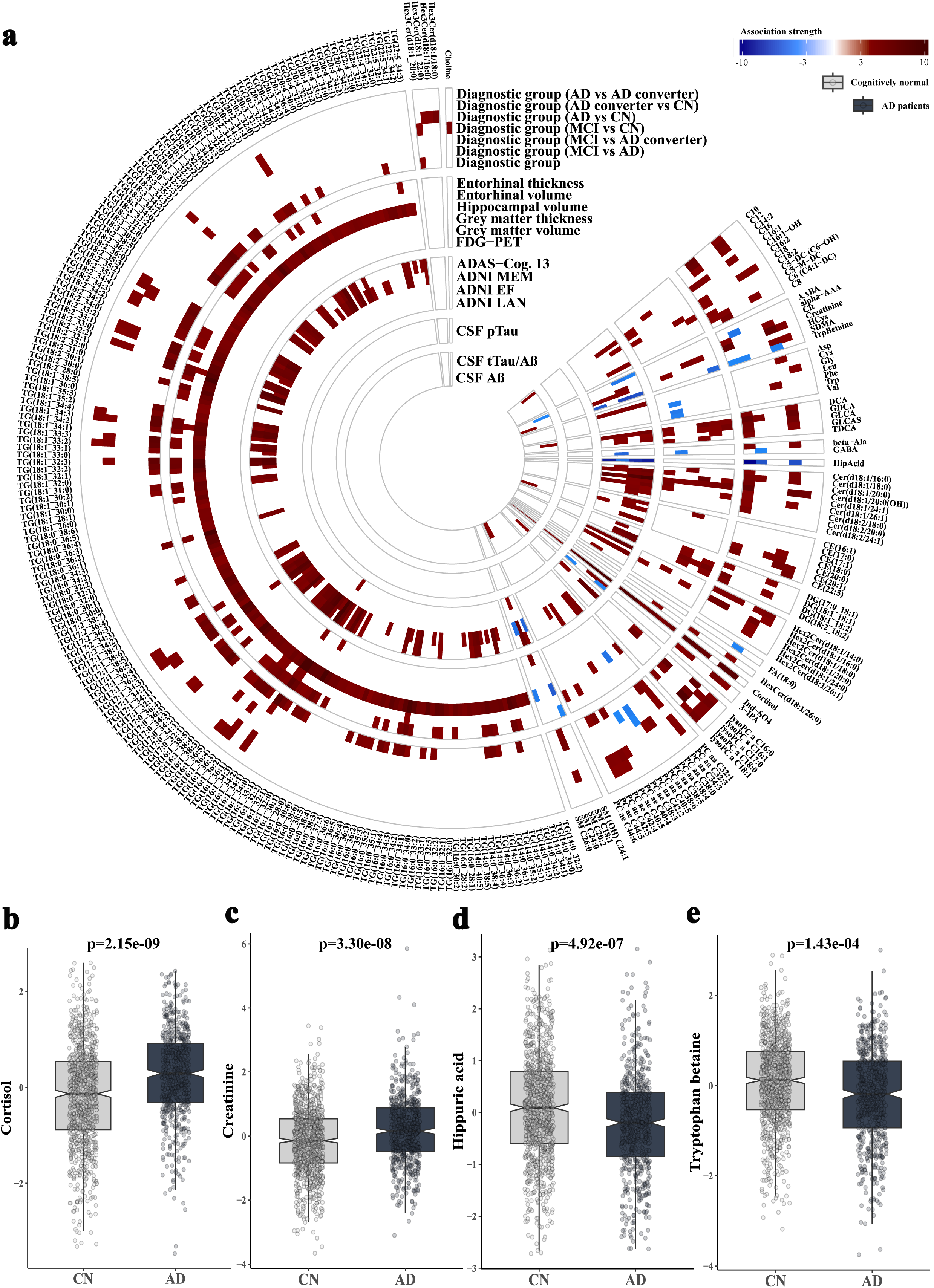
Significant associations in joint multi-timepoint analysis. **(a)** Circular heatmap showing all significant associations obtained. Each individual track represents a particular phenotype, red color constitutes positive association, and blue color represents negative associations. Association strength is calculated as −log_10_(p) × sign(β) where p is the p-value associated with the disease phenotype and β is its estimated effect coefficient. All effect directions across phenotypes were aligned so that positive beta estimates consistently reflect correlation with worse disease outcome. Metabolites are grouped by the biochemical classes provided by the platform vendor. **(b-e)** Exemplary boxplots displaying whitened residual values extracted from the joint multi-timepoint analysis model in controls and AD patients, respectively, for the two most significant positive (cortisol, creatinine) and negative (hippuric acid, tryptophan betaine) associations with AD. Box plots display the median (central line), interquartile range (box bounds), whiskers extending to the smallest and largest values within 1.5 times the interquartile range from the quartiles, and notches indicating the 95% confidence interval for the median. All tests were two-sided and raw *p* values were reported. *CN – cognitively normal controls; AD – participants with clinical Alzheimer’s disease*.

We then categorized all significant metabolite–AD associations based on their recurrence across phenotypic endpoints and the two models (**Figure 3** and **Supplementary Table S11**). Metabolites showing consistent cross-sectional associations at two or more single-timepoint analyses plus were also significant in the joint multi-timepoint analysis were classified as persistently associated metabolites (*n*=128). Those cross-sectionally significant at none or just one single timepoint but also in the joint multi-timepoint analysis were categorized as latent (*n*=153), where significant signals only emerge with large statistical power available. The third category, episodic metabolites (*n*=27), included metabolites that were significant only in single-timepoint cross-sectional analyses but not in the joint multi-timepoint analysis, indicating weak or transient effects. Out of those, 26 were only detected at one single timepoint.

**Figure 3:**
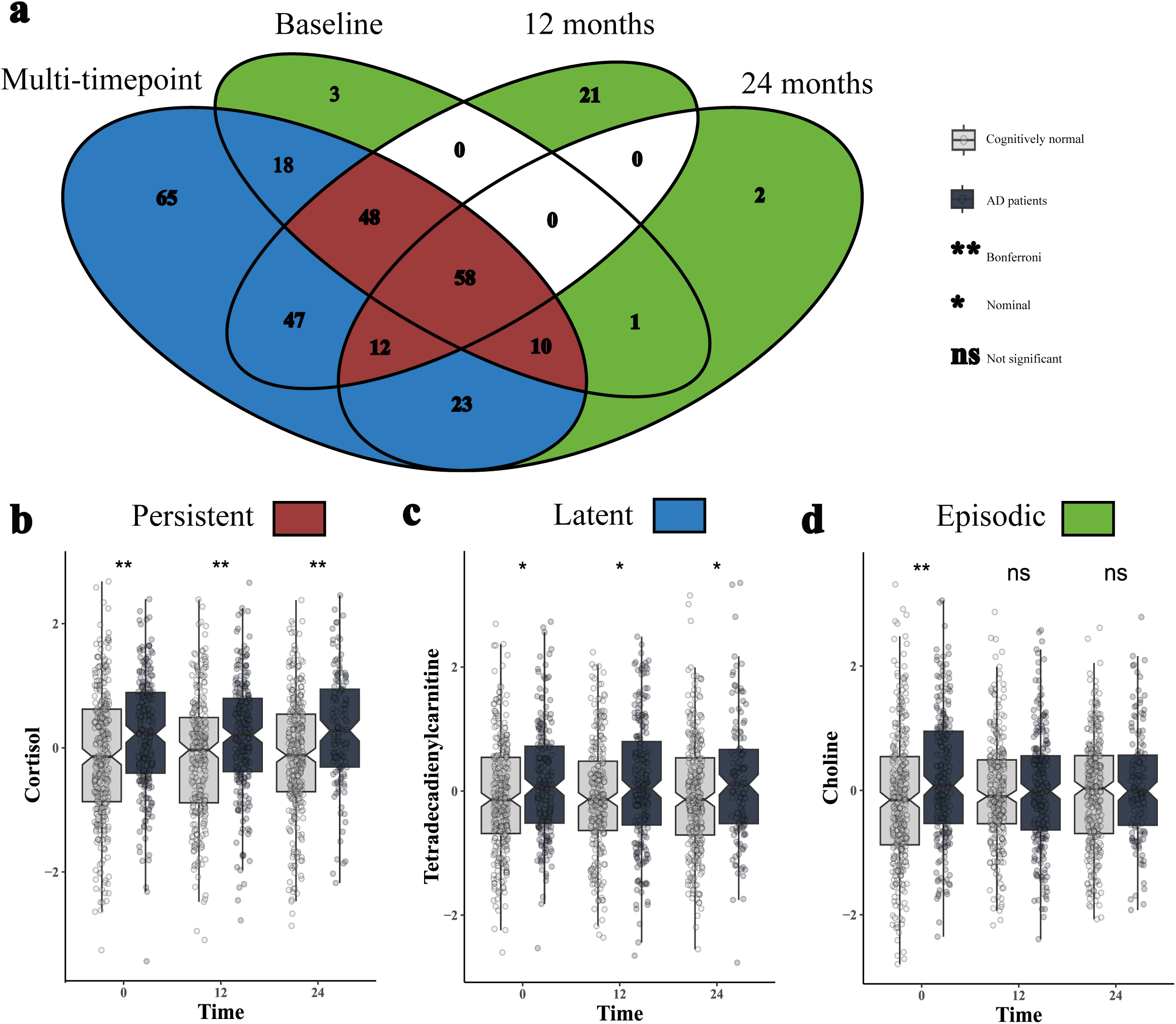
Temporal patterns of associations. **(a)** Venn diagram visualizing the shared significant metabolites across single timepoint cross-sectional analysis and joint multi-timepoint analysis. The Venn diagram is colored according to the respective categories, which are defined as follows: (i) Persistent metabolites (n=128): Significant in the joint multi-timepoint analysis and in all or ≥2 single-timepoint analyses. (ii) Latent metabolites (n=153): Significant in the joint multi-timepoint analysis but in ≤1 single-timepoint analysis. (iii) Episodic metabolites (n=27): Significant in one or more single-timepoint analyses but not in the joint multi-timepoint analysis. **(b-d)** Examples for the association patterns observed across the different models that define the different categories. The y-axis displays metabolite residuals derived from single time point models, while the x-axis represents time in months. Residuals are stratified by diagnostic groups (CN and AD), p-value significance is reported as calculated in the single time point cross-sectional analyses. Box plots display the median (central line), interquartile range (box bounds), whiskers extending to the smallest and largest values within 1.5 times the interquartile range from the quartiles, and notches indicating the 95% confidence interval for the median. All tests were two-sided and raw *p* values were reported. *CN – cognitively normal controls; AD – participants with clinical Alzheimer’s disease*.

The majority of the 128 persistent metabolites were TGs and diglycerides (DGs) (*n* = 95). The rest of the metabolites included: cortisol, creatinine, symmetric dimethylarginine (SDMA), alpha-amino adipic acid (α-AAA), choline, phenylalanine, alanine, tryptophan betaine, indoxyl sulphate, hippuric acid, decanoylcarnitine (C10:0), dodecanoylcarnitine (C12:0), octadecanoylcarnitine (C18:0), cholesteryl esters (CE) 20:1 and CE(22:5), the glycine-conjugated secondary bile acids glycodeoxycholic acid (GDCA), glycolithocholic acid (GLCA) and GLCA sulfate (GLCAS), phosphatidylcholine (PC) aa C32:3, sphingomyelin (SM) C20:2, hydroxy-SM C24:1, lyso-PCs (LPC) C16:0, C16:1, and C18:0, and nine (hexosyl-)ceramides. Of the 153 metabolites in the latent category, the majority were again TGs and DGs (n=93). The other metabolites included: aspartic acid, citrate, cysteine, α-aminobutyric acid (AABA) and γ-aminobutyric acid (GABA), homocysteine, glycine, leucine, valine, tryptophan, deoxycholic acid (DCA), taurodeoxycholic acid (TDCA), fatty acid (FA) C18:0, 3-indolepropionic acid, LPCs C17:0 and C18:1, SMs C18:1, C24:0, and C26:0, five cholesteryl esters, ten acylcarnitines, eleven (hydroxyl-)ceramides, and 15 PCs. Lastly, the 27 metabolites in the episodic category included: 3-methylhistidine, serine, lysine, isoleucine, sarcosine, hydroxy-SM C22:1, dihydroceramide (d18:0/24:0), dihexosylceramide (d18:1/22:0), trihexosylceramide (d18:1/24:1), six PCs and twelve TGs/DGs.

To further substantiate the temporal classification of metabolites, we performed replication analysis using an independent lipidomics dataset generated on largely overlapping ADNI samples (**Supplementary Tables S12–S14** and **Supplementary Figure S3**) and compared results across platforms. Out of 265 lipids showing significant associations, 189 could be mapped to 120 unique corresponding lipid species in the lipidomics dataset (**Supplementary Table S7**). Of these 189 lipids, 129 (68.25%; n=81 unique lipids) could be successfully replicated. Specifically, 76 lipids were classified as persistent, of which 65 (85.53%) were replicated; 98 were classified as latent, of which 63 (64.29%) were replicated; and 15 were classified as episodic, of which 1 (6.67%) were replicated, supporting the proposed temporal classification by reproducibility based on replication success.

### 3.3. Identification of evolving metabolic changes over time

In the time-interaction analysis, we extended this framework by including time-by-phenotype as an interaction term, allowing effect sizes to vary across time and thus capturing whether the strength or effect direction of associations changed longitudinally (**Figure 4a**). As a sensitivity check we performed two additional analyses: a subset time-interaction analysis restricted to the three main timepoints, and a paired t-test analysis on subjects who transitioned from CN to MCI or MCI to AD across any time interval for each metabolite. A metabolite was considered to significantly change longitudinally with disease if it showed: (i) significance in both the full and subset time-interaction analysis; (ii) significance in the full time-interaction analysis (p < 4.1 × 10□□; Figure 3) and paired t-test analysis (p < 0.05); or (iii) significance in all the three analyses. In case of associations with time-resolved diagnostic status, no additional criteria were assessed.

**Figure 4:**
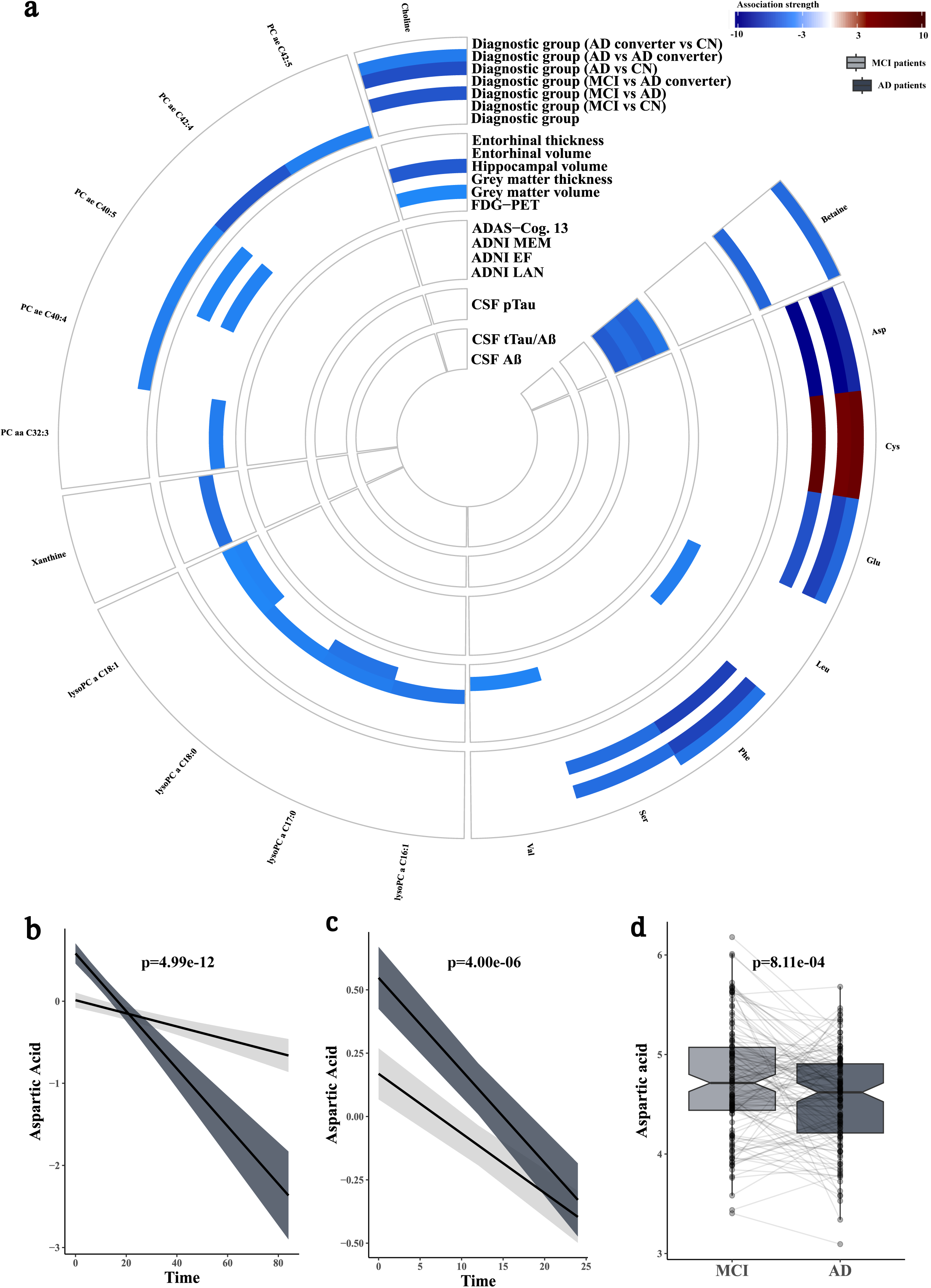
Significant associations in time-interaction analysis. **(a)** Heatmap showing all the significant associations found in time-interaction analysis. Each individual track represents a particular phenotype, red indicates positive associations, i.e. the metabolite levels increase with less desirable phenotypic levels over time, and blue represents negative associations, i.e. metabolite levels tend to decrease with respect to disease over time. Association strength is visualized by −log_10_(p) × sign(β) where p is the p-value for the interaction between disease phenotype and time and β is the effect estimate. All effect directions across phenotypes were aligned so that higher beta estimates consistently reflect a worse disease outcome. Metabolites are grouped by the biochemical classes provided by the platform vendor. **(b & c)** Effect plots of the significant association of aspartic acid visualizing its temporal trajectory in MCI and AD in both the full dataset **(b)** and the dataset subset limited to the three main time points **(c)**. The x-axis represents time in months, and the y-axis shows the model-predicted levels of aspartic acid. **(d)** Boxplot illustrating the results of the paired t-test comparing aspartic acid levels for each subject that converted from MCI to AD over a 6-month interval. The x-axis denotes the diagnostic groups, and the y-axis represents levels of aspartic acid. Ribbons around linear fits represent ±1 standard error around the estimate. Box plots display the median (central line), interquartile range (box bounds), whiskers extending to the smallest and largest values within 1.5 times the interquartile range from the quartiles, and notches indicating the 95% confidence interval for the median. All tests were two-sided and raw *p* values were reported. *MCI – participants with mild cognitive impairment; AD – participants with clinical Alzheimer’s disease*.

Using the aforementioned criteria, we identified 19 metabolites (3.75%) that were significantly associated. These included aspartic acid, leucine, betaine, valine, cysteine, glutamate, phenylalanine, serine, xanthine, choline, the LPCs C16:1, C17:0, C18:0, and C18:1, PC aa C32:3, PC ae C40:4, PC ae C40:5, PC ae C42:4, and PC ae C42:5. Among these, valine, LPC C16:1, LPC C17:0, and PC aa C32:3 were supported by both the full time-interaction analysis and paired t-test analysis (**Supplementary Table S15**), while betaine and the remaining four PCs were significant in the full time-interaction analysis associated with the time-resolved diagnostic status. The remaining metabolites were consistently significant in both the full and subset time-interaction analysis with aspartic acid showing all three lines of evidence. From a phenotype perspective, grey matter volume and grey matter thickness (n=7 metabolites each) exhibited the largest number of metabolite associations **(Supplementary Table S10; Supplementary Figure S2).**

### 3.4. Association patterns observed across the different analyses

To directly compare cross-sectional and longitudinal results, we defined concordant effects as associations with the same effect size direction across single timepoint cross-sectional or joint multi-timepoint analysis and the time-interaction analysis, and discordant effects as associations with opposite effect size directions. Concordant effects were observed for the branched-chain amino acids valine and leucine, which showed consistently lower levels in individuals with reduced grey matter volume in both the joint multi-timepoint analysis and the time-interaction analysis. Cysteine also demonstrated a concordant positive association with AD versus CN, detectable both in the 12-month cross-sectional analysis and the time-interaction analysis. Notably, cysteine synthesis (expressed as the ratio of cysteine over the sum of serine and methionine) was the only MetaboINDICATOR showing a significant p-gain in the time-interaction analysis for the AD versus CN comparison (see **Supplementary Tables S16-S18** and **Supplementary Figure S4**; p-gain = 3371.48). In contrast, discordant effects were identified for PC ae C40:4, PC ae C42:4, and PC ae C42:5, which showed a positive association with AD in the joint multi-timepoint analysis but negative associations in the time-interaction analysis. Similarly, choline, phenylalanine, and aspartic acid were elevated at baseline in individuals with AD but declined more sharply over time in the same individuals, leading to opposite effect directions across the two analyses. Phenylalanine and choline were the only persistent metabolites to exhibit a discordant pattern, starting at higher levels and more steeply decreasing levels in AD (as visualized for aspartic acid in **Figure 4b-d**), while the rest of the metabolites were either latent or episodic in nature.

To assess whether these trends extended to the metabolite-class level, we quantified the proportion of metabolites in each class that were significantly associated with any AD phenotype **(Figure 5)**. This revealed several patterns: Acylcarnitines, ceramides (including hexosyl-ceramides), bile acids, TGs, and DGs showed frequent associations in both single timepoint cross-sectional analyses and in the joint multi-timepoint analysis, but none in the time-interaction analysis. CEs had few associations at baseline but a greater number at later follow-up (24 months), but without further significant evidence from the time-interaction analysis. PCs and LPCs were elevated in disease at baseline but declined over time in relation to less favorable phenotypes. Fatty acids were reduced in individuals with less favorable phenotypes, with no significant associations detected in the time-interaction analysis. Finally, SMs, amino acids and biogenic amines, and several other metabolites (such as hexose, indoles and derivatives, choline, and cortisol; **Supplementary Table S2**) displayed variable or metabolite-specific associations, without a consistent class-level trend besides most metabolites within these classes (except for cysteine) declining over time along the disease trajectory in the time-interaction analysis.

**Figure 5:**
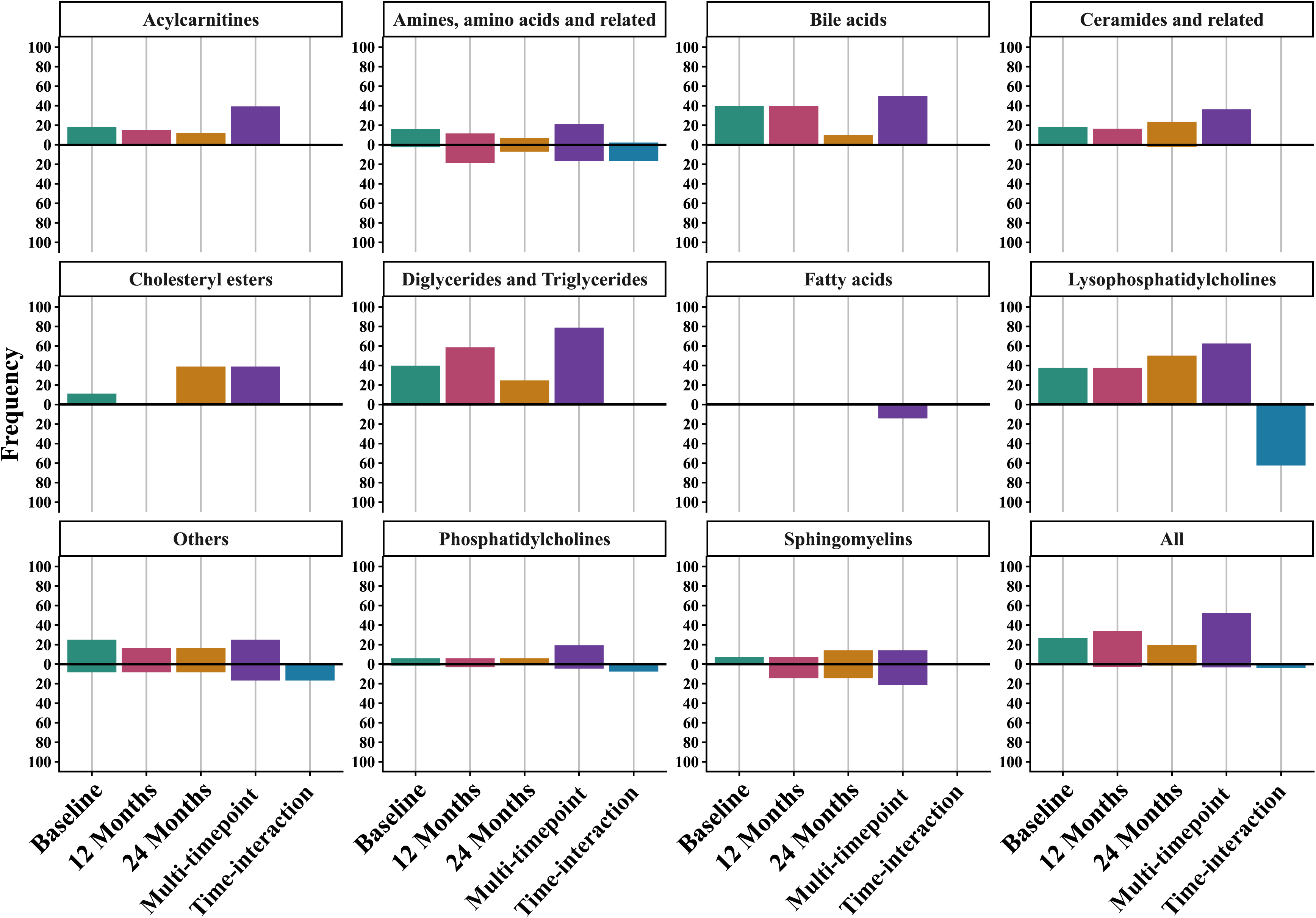
Miami plots representing the frequency (in %) of metabolites significantly associated with any disease phenotype across the different analyses grouped by biochemical classes. Frequencies of positive associations with disease are displayed above zero, negative associations are shown below zero, but frequency values are expressed as positive percentages in both directions. “Others” here combines metabolite classes with only few representatives, such as hexose, indoles and derivatives, choline, and cortisol (**Supplementary Table S2**).

### 3.5. Statistical robustness of time-interaction associations

Across the 44 evaluable time-interaction associations, bootstrap estimates closely agreed with the original estimates, with a mean absolute bias of 1.10×10^−4^, a median absolute bias of 4.28×10^−5^, and a maximum absolute bias of 5.12×10^−4^. Bootstrap-t 95% confidence intervals excluded zero for all associations, and 40 of 44 bootstrap effect distributions (90.9%) showed no evidence for deviations from normality. At the study-wide significance threshold, 43 out of 44 associations were significant in more than 50% of bootstrap iterations (i.e., meeting the post-hoc power threshold to replicate associations) with same effect directions, with a median replication proportion of 81.8%. The same 43 associations remained significant according to the mean-beta/mean-SE summary test **(Supplementary Table S19**). The only exception was the association between PC ae C40:5 and grey matter thickness, which reached the study-wide threshold in only 48.4% of iterations and a bootstrap summary *p* = 5.52×10^−4^.

## 4. Discussion

In this study, we systematically characterized temporal metabolite–AD associations in serum by distinguishing persistent, latent, and episodic associations, as well as metabolites whose associations changed over time. Across all analyses, 311 metabolites were associated with at least one AD-related phenotype. Most were detected in the joint multi-timepoint analysis (n=281), followed by single-timepoint cross-sectional analyses (n=243), whereas 19 emerged from the time-interaction analysis, which captures longitudinal shifts in disease-related level differences. All results are also accessible in an interactive web portal (https://metabolomics.helmholtz-munich.de/mxtra_ad/).

The larger number of associations in the joint multi-timepoint analysis likely reflects increased statistical power through pooling data across the three main timepoints in linear mixed-effects models. Moreover, the relatively small number of metabolites with significant time-varying effects suggests that many AD-related metabolic alterations remain comparatively stable once established. Indeed, over 40% of significant metabolites were classified as persistent, showing associations in the multi-timepoint analysis and cross-sectionally at at least two timepoints.

With respect to metabolites identified in the time-interaction analysis, 16 out of 19 were also significant in either the cross-sectional or multi-timepoint models. Eleven were latent or episodic, while phenylalanine, choline, LPC C16:1 and C18:0, and PC aa C32:3 showed persistently higher levels in more affected individuals but declined more strongly over time. Except for cysteine, all metabolites with evolving associations showed negative time interactions, indicating steeper decreases in more severely affected individuals. This is especially relevant for metabolites such as PC ae C42:4, which this and earlier work found elevated in AD blood samples^39^. If such metabolites decline faster with disease progression, studies focused on later-stage AD may report conflicting results.

These findings are broadly consistent with prior literature^4,40,41^. Our findings on higher levels of sphingomyelins, ceramides, and cholesteryl esters align with reports linking these lipids to poorer AD outcomes^42–45^. Likewise, higher levels of secondary conjugated bile acids are replicated in earlier plasma, serum, and brain studies^5,46,47^. Higher levels of LPCs C16:1 and C18:1 have previously been associated with MCI and memory decline, followed by steep depletion^41,48^. Elevated cortisol levels are consistent with systemic stress-related changes in brain, CSF, and plasma linked to brain atrophy and neuropathological burden^49,50^. We further replicated creatinine, an indicator of muscle breakdown and early marker of frailty and dementia risk^51–53^, to be robustly associated with AD. Hippuric acid, a protective diet- and microbiota-related metabolite^54^, showed negative associations with AD, in line with previous animal studies^55,56^. We also extend prior evidence on the role of PCs in AD, including potentially novel findings^39,45,57,58^. Finally, we replicated nearly three quarters of reported lipid associations in an independent lipidomics dataset, with replication rates highest for persistent (∼90%) and lowest for episodic associations (∼10%), as expected, and provide evidence for time-interaction effect stability through bootstrapping.

Together, these results both replicate and extend previous metabolic findings in AD. Below, we highlight several metabolic domains that appear especially relevant based on present and prior evidence.

### 4.1 Bioenergetic dysfunction as a chronic feature of AD

Our data strengthens evidence for impaired energy metabolism in AD^59^. Individuals with AD showed elevated TGs, medium- and long-chain acylcarnitines, and reduced free fatty acids in the joint multi-timepoint analysis, suggesting a compensatory yet inefficient shift toward lipid-based energy metabolism^59,60^, consistent with impaired mitochondrial β-oxidation or carnitine shuttle dysfunction^61^. This supports the hypothesis that mitochondrial bioenergetic failure in AD may result in reliance on alternative fuel sources when glucose utilization is compromised^62^. Importantly, many of the elevated TGs, DGs, and acylcarnitines, were persistent, indicating that bioenergetic dysfunction is a chronic component of AD pathophysiology in both brain and periphery. Mitochondrial dysfunction, together with elevated homocysteine, may also contribute to muscle breakdown, potentially explaining the sustained increases in creatinine and alanine.

Further, we found levels of the two branched-chain amino acids (BCAAs) valine and leucine were negatively associated with grey matter volume and declined faster over time in AD. This extends earlier findings^63,64^ and is consistent with recent evidence showing lower plasma valine, leucine, and isoleucine in individuals with AD or dementia and a marked decline in BCAA levels after the age of 60 in *APOE* ε4 carriers^65^. In addition, a previous longitudinal study reported decreasing levels of valine over time to be associated with all-cause mortality^66^. Together these findings may reflect increased BCAA use as alternative substrates for the tricarboxylic acid (TCA) cycle. Because BCAA supplementation can promote mitochondrial biogenesis and ATP production in malnourished older adults^67–69^, their depletion in AD may worsen cellular energy supply. BCAA dysregulation is also linked to insulin resistance and metabolic syndrome, both established AD risk factors^70,71^. Through mTORC1 signaling, chronic BCAA depletion could impair autophagy and protein turnover, increase oxidative vulnerability, and promote neurodegeneration^72–74^. Since BCAAs also contribute to glutamate and GABA synthesis, their depletion may further connect impaired bioenergetics to synaptic dysfunction^75^.

### 4.2 Impairment of neurotransmission systems worsens with AD progression

Disrupted neurotransmission, especially in cholinergic, glutamatergic, and GABAergic systems, is central to AD pathology^76,77^. Consistently, choline was elevated in AD but declined steeply over time with advancing disease. This peripheral depletion of choline in AD may reflect increased compensatory transport into the brain, which in rodents depends on cerebral choline demand^78^.

We further found GABA levels to be negatively associated with executive function and grey matter volume and a significant decrease of glutamate levels over time in AD. As noted above, these may be linked to impaired bioenergetics, since BCAA depletion and reduced TCA cycle activity can limit production of α-ketoglutarate, a precursor for both glutamate and GABA^79,80^.

Prior longitudinal studies support a dynamic glutamate trajectory, where peripheral glutamate levels may be elevated years before dementia onset^81^, whereas lower levels closer to diagnosis are associated with faster decline and inverse brain-serum correlations^82–86^. In our data, lower glutamate levels do not appear to be explained by increased conversion to glutamine, as glutaminase activity in the metaboINDICATOR analysis was not significantly associated with AD.

Finally, similar to choline, phenylalanine, aspartic acid, and serine showed initially higher but steeply decreasing levels in AD. In earlier brain metabolomics work, we observed elevated glutamate, serine, and phenylalanine in AD brain tissue^8^, which could point to altered uptake or compartmental redistribution. Phenylalanine is a precursor for tyrosine and catecholamines, and its dysregulation in AD may contribute to dopaminergic and noradrenergic dysfunction^15,85,87–89^. Altered serine and aspartic acid may likewise affect neurotransmission through their roles as NMDA receptor agonists^15,90^.

Overall, these findings indicate that compensatory responses in neurotransmitter-related metabolism are progressively impaired as disease advances.

### 4.3 Oxidative stress response and antioxidant failure

GABA, besides its role in neurotransmission, and AABA have antioxidant functions, additionally implicating oxidative stress, another hallmark of AD^91^. We found lower GABA and AABA levels to be associated with worse cognitive function and less favorable biomarker profiles, consistent with previous work^92^.

Several other metabolites further suggest a role for diet and the gut microbiome in oxidative and inflammatory pathways. Lower tryptophan betaine, an anti-inflammatory marker linked to plant-rich diets^93,94^, was consistently associated with poorer cognitive outcomes. We also detected associations for microbially derived tryptophan metabolites. Indoxyl sulfate, a pro-inflammatory and neurotoxic co-metabolite, was persistently elevated in AD, replicating prior plasma and brain-based studies^49,95,96^. In contrast, lower levels of 3-indolepropionic acid (3-IPA), which has anti-inflammatory, antioxidant, and neuroprotective properties^97–99^, were associated with poorer cognition^49,100^. Because 3-IPA production can be enhanced by low-fat diets, these observations also point toward potentially modifiable aspects of disease^101^.

Conversely, several markers of oxidative damage were elevated in AD. For instance, SDMA, a marker of oxidative stress^102^, and homocysteine was positively associated in the joint multi-timepoint analysis. Hyperhomocysteinemia is an established dementia risk marker and may reflect subclinical vitamin B deficiency, which is itself linked to oxidative stress and AD^16,81,103–105^.

Finally, we observed potentially compensatory increases in cysteine and glycine levels in AD. Both metabolites are building blocks of glutathione, a key antioxidant previously linked to AD, including in brain tissue^8^. Cysteine additionally increased over time, and cysteine synthesis, approximated by the ratio of cysteine to the sum of serine and methionine, was the only MetaboINDICATOR with a significant longitudinal change. This may reflect an adaptive attempt to buffer oxidative stress or vitamin B-related deficits by increasing conversion of homocysteine via serine to cysteine^106–111^.

However, cysteine is also a precursor for coenzyme A, linking it again to mitochondrial bioenergetics, including the TCA cycle and β-oxidation^112,113^. In general, oxidative stress and mitochondrial dysfunction are inherently intertwined. And as we found glutamate, the third glutathione precursor, to decline with disease progression, further work is needed to fully disentangle compensation from failure in this interconnected system.

### 4.4 Limitations and potential for future work

This study has several limitations. First, ADNI-1/GO/2 is enriched for older adults of Caucasian descent, which limit generalizability given that age, ethnoracial background, and genetics affect both metabolism and AD. Second, statistically averaging across participants may mask heterogeneity in individual disease trajectories. Third, CSF measures were largely restricted to baseline, limiting power for longitudinal analyses on CSF amyloid and tau. Fourth, lipid class-level trends were consistent, but species-level interpretation remains constrained by unresolved acyl chain composition and desaturation sites. In addition, the platform used here is lipid-focused, so broader untargeted approaches may capture additional biology. Finally, while in general our results are very consistent with previous reports and we performed ample analyses to provide estimates for replicability and robustness of our findings, validation in independent studies is required to assess transportability of our findings to the general population. In particular, the observed time-interaction effects require external replication, despite their strong bootstrap robustness and consistency with prior evidence. Episodic associations should likewise be interpreted with caution, as they were observed only at specific timepoints, although this does not preclude their biological relevance.

Future work should integrate longitudinal blood metabolomics paired with brain tissue-derived data along with emerging blood biomarkers while accounting for diet and gut microbiome composition^114,115^ to improve temporal mapping between central and peripheral AD metabolism.

### 4.5 Conclusion

In summary, this study systematically characterized temporal evaluation of metabolic alterations in AD by distinguishing persistent, latent, episodic, and time-evolving associations. Our findings highlight widespread and interconnected alterations to lipid, amino acid, oxidative stress, energy, and neurotransmitter metabolism. Overall, our results indicate robust but only partly dynamic metabolic abnormalities in AD and underscore the importance of temporal context in metabolomics. As the identified metabolic alterations are involved in pathways regulated across multiple organ systems, biochemical changes in AD may reflect systemic metabolic dysfunction rather than pathological processes confined to the brain. We speculate that longitudinal effects on metabolic substrates relevant across the periphery and the central nervous system as exemplified here mirror globally reduced metabolic flexibility. While circulating and brain metabolite trajectories likely differ, these peripheral alterations could nevertheless influence the metabolic environment in which neurodegeneration progresses. The predominance of persistent alterations may provide important opportunities here, as it suggests sustained windows for pathway-targeted interventions. Future studies will be essential to clarify mechanisms, refine temporal trajectories, and identify modifiable disease-related factors.

## Consent Statement

All ADNI participants provided written informed consent. Consent forms were approved by each participating site’s institutional review board.

## Data Availability

The results published here are in whole or in part based on data obtained from the AD Knowledge Portal (https://adknowledgeportal.org). The AD Knowledge Portal is a platform for accessing data, analyses, and tools generated by the Accelerating Medicines Partnership (AMP-AD) Target Discovery Program and other National Institute on Aging (NIA)-supported programs to enable open-science practices and accelerate translational learning. The data, analyses, and tools are shared early in the research cycle without a publication embargo on secondary use. Data are available for general research use according to the following requirements for data access and data attribution (https://adknowledgeportal.org/DataAccess/Instructions). ADNI longitudinal metabolomics data can be accessed under https://doi.org/10.7303/9618123. All other data, including the full complement of clinical and demographic data for the ADNI cohorts are hosted on the LONI data sharing platform and can be requested at https://adni.loni.usc.edu/data-samples/adni-data/#AccessData. All summary statistics as well as additional information and visualizations are available online at https://metabolomics.helmholtz-munich.de/mxtra_ad/.

## Code Availability

All R code required to conduct the analyses reported here is available at https://github.com/compneurobio/mxtra-ad. All analyses were conducted using the MeTime R package^36^ available at https://github.com/compneurobio/MeTime/.

## Acknowledgements

This work was supported by the National Institutes of Health/the National Institute on Aging through grants R01AG069901, U01AG088562, and R01AG081322. Metabolomics data is provided by the Alzheimer’s Disease Metabolomics Consortium (ADMC) and funded wholly or in part by the following grants and supplements thereto: NIA U19AG063744, R01AG046171, RF1AG057452, R01AG059093, RF1AG058942, U01AG061359, awarded to Dr. Kaddurah-Daouk at Duke University in partnership with a large number of academic institutions. As such, the investigators within the ADMC, not listed specifically in this publication’s author’s list, provided data along with its pre-processing and prepared it for analysis, but did not participate in analysis or writing of this manuscript. A complete listing of ADMC investigators can be found at: https://sites.duke.edu/adnimetab/team/. The Biocrates Q500 data was generated at Duke University. Data collection and sharing for this project was funded by the Alzheimer’s Disease Neuroimaging Initiative (ADNI) (National Institutes of Health Grant U01 AG024904) and DOD ADNI (Department of Defense award number W81XWH-12-2-0012). ADNI is funded by the National Institute on Aging, the National Institute of Biomedical Imaging and Bioengineering, and through generous contributions from the following: AbbVie, Alzheimer’s Association; Alzheimer’s Drug Discovery Foundation; Araclon Biotech; BioClinica, Inc.; Biogen; Bristol-Myers Squibb Company; CereSpir, Inc.; Cogstate; Eisai Inc.; Elan Pharmaceuticals, Inc.; Eli Lilly and Company; EuroImmun; F. Hoffmann-La Roche Ltd and its affiliated company Genentech, Inc.; Fujirebio; GE Healthcare; IXICO Ltd.; Janssen Alzheimer Immunotherapy Research & Development, LLC.; Johnson & Johnson Pharmaceutical Research & Development LLC.; Lumosity; Lundbeck; Merck & Co., Inc.; Meso Scale Diagnostics, LLC.; NeuroRx Research; Neurotrack Technologies; Novartis Pharmaceuticals Corporation; Pfizer Inc.; Piramal Imaging; Servier; Takeda Pharmaceutical Company; and Transition Therapeutics. The Canadian Institutes of Health Research is providing funds to support ADNI clinical sites in Canada. Private sector contributions are facilitated by the Foundation for the National Institutes of Health (www.fnih.org). The grantee organization is the Northern California Institute for Research and Education, and the study is coordinated by the Alzheimer’s Therapeutic Research Institute at the University of Southern California. ADNI data are disseminated by the Laboratory for Neuro Imaging at the University of Southern California. KH is supported by a National Health and Medical Research Council (NHMRC) investigator grant (#1197190).

## Author Contributions

**Conceptualization:** PJM, GK, RK-D, MA; **Data curation:** BM, PW, SLR, CB, NK, KK, AK-P, JWT, TB, KN, AJS, MA; **Collection of clinical data:** PMD; **Formal analysis:** BM, PW, MA; **Funding acquisition:** GK, RK-D, MA; **Investigation:** BM, PW, JJB, NK, KK, JWT, GK, MA; **Methodology:** BM, PW, TW, KH, JWT, MA; **Project administration:** CB, AK-P, GK, RK-D, MA; **Software:** BM, PW, MA; **Supervision:** PJM, GK, RK-D, MA; **Visualization:** BM, PW, JJB, GK, MA; **Writing – original draft:** BM, PW, JJB, MA; **Writing – review & editing:** All authors

## References

1. Querfurth HW, LaFerla FM. Alzheimer’s disease. N Engl J Med. Jan 28 2010;362(4):329–44. doi:10.1056/NEJMra0909142

2. Trushina E, Dutta T, Persson XM, Mielke MM, Petersen RC. Identification of altered metabolic pathways in plasma and CSF in mild cognitive impairment and Alzheimer’s disease using metabolomics. PLoS One. 2013/5//2013;8(5):e63644. doi:10.1371/journal.pone.0063644

3. Han X, Rozen S, Boyle SH, et al. Metabolomics in early Alzheimer’s disease: identification of altered plasma sphingolipidome using shotgun lipidomics. PLoS One. 2011;6(7):e21643. doi:10.1371/journal.pone.0021643

4. Toledo JB, Arnold M, Kastenmuller G, et al. Metabolic network failures in Alzheimer’s disease: A biochemical road map. Alzheimers Dement. Sep 2017;13(9):965–984. doi:10.1016/j.jalz.2017.01.020

5. MahmoudianDehkordi S, Arnold M, Nho K, et al. Altered bile acid profile associates with cognitive impairment in Alzheimer’s disease-An emerging role for gut microbiome. Alzheimers Dement. Jan 2019;15(1):76–92. doi:10.1016/j.jalz.2018.07.217

6. Tian Q, Yao S, Marron MM, et al. Shared plasma metabolomic profiles of cognitive and mobility decline predict future dementia. Geroscience. Oct 2024;46(5):4883–4894. doi:10.1007/s11357-024-01228-7

7. Ambeskovic M, Hopkins G, Hoover T, Joseph JT, Montina T, Metz GAS. Metabolomic Signatures of Alzheimer’s Disease Indicate Brain Region-Specific Neurodegenerative Progression. Int J Mol Sci. Sep 30 2023;24(19):14769–14769. doi:10.3390/ijms241914769

8. Batra R, Arnold M, Worheide MA, et al. The landscape of metabolic brain alterations in Alzheimer’s disease. Alzheimers Dement. Mar 2023;19(3):980–998. doi:10.1002/alz.12714

9. Akyuz E, Arulsamy A, Aslan FS, et al. An Expanded Narrative Review of Neurotransmitters on Alzheimer’s Disease: The Role of Therapeutic Interventions on Neurotransmission. Mol Neurobiol. Feb 2025;62(2):1631–1674. doi:10.1007/s12035-024-04333-y

10. Pillet LE, Taccola C, Cotoni J, Thiriez H, André K, Verpillot R. Correlation between cognition and plasma noradrenaline level in Alzheimer’s disease: a potential new blood marker of disease evolution. Translational Psychiatry. 2020/12//2020;10(1):1–10. doi:10.1038/S41398-020-0841-7;TECHMETA

11. Knornschild F, Zhang EJ, Ghosh Biswas R, et al. Correlations of blood and brain NMR metabolomics with Alzheimer’s disease mouse models. Transl Psychiatry. Mar 18 2025;15(1):87. doi:10.1038/s41398-025-03293-8

12. Ahmad S, Yang W, Orellana A, et al. Association of oxidative stress and inflammatory metabolites with Alzheimer’s disease cerebrospinal fluid biomarkers in mild cognitive impairment. Alzheimers Res Ther. Jul 30 2024;16(1):171. doi:10.1186/s13195-024-01542-4

13. Dronse J, Ohndorf A, Richter N, et al. Serum cortisol is negatively related to hippocampal volume, brain structure, and memory performance in healthy aging and Alzheimer’s disease. Front Aging Neurosci. 2023/5//2023;15:1154112. doi:10.3389/fnagi.2023.1154112

14. Mosconi L, Williams S, Carlton C, et al. Sex-specific associations of serum cortisol with brain biomarkers of Alzheimer’s risk. Sci Rep. Mar 6 2024;14(1):5519. doi:10.1038/s41598-024-56071-9

15. Xu J, Begley P, Church SJ, et al. Graded perturbations of metabolism in multiple regions of human brain in Alzheimer’s disease: Snapshot of a pervasive metabolic disorder. Biochim Biophys Acta. Jun 2016;1862(6):1084–92. doi:10.1016/j.bbadis.2016.03.001

16. de Leeuw FA, Peeters CFW, Kester MI, et al. Blood-based metabolic signatures in Alzheimer’s disease. Alzheimers Dement (Amst). 2017/1//2017;8(1):196–207. doi:10.1016/j.dadm.2017.07.006

17. Kurbatova N, Garg M, Whiley L, et al. Urinary metabolic phenotyping for Alzheimer’s disease. Sci Rep. Dec 10 2020;10(1):21745. doi:10.1038/s41598-020-78031-9

18. Mielke MM, Haughey NJ, Bandaru VV, et al. Plasma ceramides are altered in mild cognitive impairment and predict cognitive decline and hippocampal volume loss. Alzheimers Dement. Sep 2010;6(5):378–85. doi:10.1016/j.jalz.2010.03.014

19. Huang YL, Chang WJ, Lin CH, et al. Data from multi-national aging cohorts show dysregulated metabolic pathways in people with physio-cognitive decline. Commun Med (Lond). Aug 14 2025;5(1):351. doi:10.1038/s43856-025-01073-5

20. Lawrence E, Vegvari C, Ower A, Hadjichrysanthou C, De Wolf F, Anderson RM. A Systematic Review of Longitudinal Studies Which Measure Alzheimer’s Disease Biomarkers. J Alzheimers Dis. 2017;59(4):1359–1379. doi:10.3233/JAD-170261

21. Wang T, Arnold M, Huynh K, et al. Trajectory of plasma lipidome associated with the risk of late-onset Alzheimer’s disease: a longitudinal cohort study. EBioMedicine. Aug 2025;118:105826. doi:10.1016/j.ebiom.2025.105826

22. Mueller SG, Weiner MW, Thal LJ, et al. The Alzheimer’s disease neuroimaging initiative. Neuroimaging Clin N Am. Nov 2005;15(4):869–77, xi–xii. doi:10.1016/j.nic.2005.09.008

23. Weiner MW, Veitch DP, Aisen PS, et al. Impact of the Alzheimer’s Disease Neuroimaging Initiative, 2004 to 2014. Alzheimers Dement. Jul 2015;11(7):865–84. doi:10.1016/j.jalz.2015.04.005

24. Jack CR, Jr., Bennett DA, Blennow K, et al. A/T/N: An unbiased descriptive classification scheme for Alzheimer disease biomarkers. Neurology. Aug 2 2016;87(5):539–47. doi:10.1212/WNL.0000000000002923

25. Hansson O, Seibyl J, Stomrud E, et al. CSF biomarkers of Alzheimer’s disease concord with amyloid-beta PET and predict clinical progression: A study of fully automated immunoassays in BioFINDER and ADNI cohorts. Alzheimers Dement. Nov 2018;14(11):1470–1481. doi:10.1016/j.jalz.2018.01.010

26. Bittner T, Zetterberg H, Teunissen CE, et al. Technical performance of a novel, fully automated electrochemiluminescence immunoassay for the quantitation of beta-amyloid (1-42) in human cerebrospinal fluid. Alzheimers Dement. May 2016;12(5):517–26. doi:10.1016/j.jalz.2015.09.009

27. Jack CR, Jr., Bernstein MA, Fox NC, et al. The Alzheimer’s Disease Neuroimaging Initiative (ADNI): MRI methods. J Magn Reson Imaging. Apr 2008;27(4):685–91. doi:10.1002/jmri.21049

28. Fischl B, Salat DH, Busa E, et al. Whole brain segmentation: automated labeling of neuroanatomical structures in the human brain. Neuron. Jan 31 2002;33(3):341–55. doi:10.1016/s0896-6273(02)00569-x

29. Fischl B, Dale AM. Measuring the thickness of the human cerebral cortex from magnetic resonance images. Proc Natl Acad Sci U S A. Sep 26 2000;97(20):11050–5. doi:10.1073/pnas.200033797

30. Landau SM, Harvey D, Madison CM, et al. Associations between cognitive, functional, and FDG-PET measures of decline in AD and MCI. Neurobiol Aging. Jul 2011;32(7):1207–18. doi:10.1016/j.neurobiolaging.2009.07.002

31. Jagust WJ, Bandy D, Chen K, et al. The Alzheimer’s Disease Neuroimaging Initiative positron emission tomography core. Alzheimers Dement. May 2010;6(3):221–9. doi:10.1016/j.jalz.2010.03.003

32. St John-Williams L, Blach C, Toledo JB, et al. Targeted metabolomics and medication classification data from participants in the ADNI1 cohort. Sci Data. Oct 17 2017;4(1):170140. doi:10.1038/sdata.2017.140

33. Breiman L. Random forests. Machine Learning. Oct 2001;45(1):5–32. doi:Doi 10.1023/A:1010933404324

34. Kursa MB, Jankowski A, Rudnicki WR. Boruta - A System for Feature Selection. Fundamenta Informaticae. 2010;101(4):271–286. doi:10.3233/Fi-2010-288

35. Kursa MB, Rudnicki WR. Feature Selection with theBorutaPackage. Journal of Statistical Software. 2010/9//2010;36(11):1–13. doi:10.18637/jss.v036.i11

36. Marella B, Weinisch P, Vehovec L, et al. MeTime: an R package for reproducible longitudinal metabolomics data analysis. Bioinform Adv. 2026;6(1):vbag252. doi:10.1093/bioadv/vbag252

37. Li J, Ji L. Adjusting multiple testing in multilocus analyses using the eigenvalues of a correlation matrix. Heredity (Edinb). Sep 2005;95(3):221–7. doi:10.1038/sj.hdy.6800717

38. Petersen AK, Krumsiek J, Wagele B, et al. On the hypothesis-free testing of metabolite ratios in genome-wide and metabolome-wide association studies. BMC Bioinformatics. Jun 6 2012;13(1):120. doi:10.1186/1471-2105-13-120

39. Wilkins JM, Trushina E. Application of Metabolomics in Alzheimer’s Disease. Front Neurol. 2018/1//2017;8(JAN):719. doi:10.3389/fneur.2017.00719

40. Chen T, Wang L, Xie G, et al. Serum Bile Acids Improve Prediction of Alzheimer’s Progression in a Sex-Dependent Manner. Adv Sci (Weinh). Mar 2024;11(9):e2306576. doi:10.1002/advs.202306576

41. Arnold M, Nho K, Kueider-Paisley A, et al. Sex and APOE epsilon4 genotype modify the Alzheimer’s disease serum metabolome. Nat Commun. Mar 2 2020;11(1):1148. doi:10.1038/s41467-020-14959-w

42. Baloni P, Arnold M, Buitrago L, et al. Multi-Omic analyses characterize the ceramide/sphingomyelin pathway as a therapeutic target in Alzheimer’s disease. Commun Biol. Oct 8 2022;5(1):1074. doi:10.1038/s42003-022-04011-6

43. Liu Y, Thalamuthu A, Mather KA, et al. Plasma lipidome is dysregulated in Alzheimer’s disease and is associated with disease risk genes. Transl Psychiatry. Jun 7 2021;11(1):344. doi:10.1038/s41398-021-01362-2

44. Cutler RG, Kelly J, Storie K, et al. Involvement of oxidative stress-induced abnormalities in ceramide and cholesterol metabolism in brain aging and Alzheimer’s disease. Proc Natl Acad Sci U S A. Feb 17 2004;101(7):2070–5. doi:10.1073/pnas.0305799101

45. Huynh K, Lim WLF, Giles C, et al. Concordant peripheral lipidome signatures in two large clinical studies of Alzheimer’s disease. Nat Commun. Nov 10 2020;11(1):5698. doi:10.1038/s41467-020-19473-7

46. Baloni P, Funk CC, Yan J, et al. Metabolic Network Analysis Reveals Altered Bile Acid Synthesis and Metabolism in Alzheimer’s Disease. Cell Rep Med. Nov 17 2020;1(8):100138. doi:10.1016/j.xcrm.2020.100138

47. Greenberg N, Grassano A, Thambisetty M, Lovestone S, Legido-Quigley C. A proposed metabolic strategy for monitoring disease progression in Alzheimer’s disease. Electrophoresis. Apr 2009;30(7):1235–9. doi:10.1002/elps.200800589

48. Lin W, Zhang J, Liu Y, et al. Studies on diagnostic biomarkers and therapeutic mechanism of Alzheimer’s disease through metabolomics and hippocampal proteomics. Eur J Pharm Sci. Jul 15 2017;105:119–126. doi:10.1016/j.ejps.2017.05.003

49. Kalecky K, German DC, Montillo AA, Bottiglieri T. Targeted Metabolomic Analysis in Alzheimer’s Disease Plasma and Brain Tissue in Non-Hispanic Whites. J Alzheimers Dis. 2022;86(4):1875–1895. doi:10.3233/JAD-215448

50. Czech C, Berndt P, Busch K, et al. Metabolite profiling of Alzheimer’s disease cerebrospinal fluid. PLoS One. 2012;7(2):e31501. doi:10.1371/journal.pone.0031501

51. Buergel T, Steinfeldt J, Ruyoga G, et al. Metabolomic profiles predict individual multidisease outcomes. Nat Med. Nov 2022;28(11):2309–2320. doi:10.1038/s41591-022-01980-3

52. Mak JKL, Kananen L, Qin C, et al. Unraveling the metabolic underpinnings of frailty using multicohort observational and Mendelian randomization analyses. Aging Cell. Aug 2023;22(8):e13868. doi:10.1111/acel.13868

53. Low DY, Lefevre-Arbogast S, Gonzalez-Dominguez R, et al. Diet-Related Metabolites Associated with Cognitive Decline Revealed by Untargeted Metabolomics in a Prospective Cohort. Mol Nutr Food Res. Sep 2019;63(18):e1900177. doi:10.1002/mnfr.201900177

54. Ticinesi A, Guerra A, Nouvenne A, Meschi T, Maggi S. Disentangling the Complexity of Nutrition, Frailty and Gut Microbial Pathways during Aging: A Focus on Hippuric Acid. Nutrients. Feb 24 2023;15(5)doi:10.3390/nu15051138

55. Rutledge GA, Sandhu AK, Miller MG, Edirisinghe I, Burton-Freeman BB, Shukitt-Hale B. Blueberry phenolics are associated with cognitive enhancement in supplemented healthy older adults. Food Funct. Jan 7 2021;12(1):107–118. doi:10.1039/d0fo02125c

56. He Y, Wang Y, Liu S, et al. A metabolomic study of the urine of rats with Alzheimer’s disease and the efficacy of Ding-Zhi-Xiao-Wan on the afflicted rats. J Sep Sci. Apr 2020;43(8):1458–1465. doi:10.1002/jssc.201900944

57. Klavins K, Koal T, Dallmann G, Marksteiner J, Kemmler G, Humpel C. The ratio of phosphatidylcholines to lysophosphatidylcholines in plasma differentiates healthy controls from patients with Alzheimer’s disease and mild cognitive impairment. Alzheimers Dement (Amst). Sep 2 2015;1(3):295–302. doi:10.1016/j.dadm.2015.05.003

58. Whiley L, Sen A, Heaton J, et al. Evidence of altered phosphatidylcholine metabolism in Alzheimer’s disease. Neurobiol Aging. Feb 2014;35(2):271–8. doi:10.1016/j.neurobiolaging.2013.08.001

59. Arnold M, Buyukozkan M, Doraiswamy PM, et al. Individual bioenergetic capacity as a potential source of resilience to Alzheimer’s disease. Nat Commun. Feb 24 2025;16(1):1910. doi:10.1038/s41467-025-57032-0

60. Sharmin T, Chatterjee P, Doecke JD, et al. Circulating medium- and long-chain acylcarnitines are associated with plasma P-tau181 in cognitively normal older adults. J Neurochem. Feb 2025;169(2)doi:10.1111/jnc.16244

61. Heggermont WA, Papageorgiou AP, Heymans S, van Bilsen M. Metabolic support for the heart: complementary therapy for heart failure? Eur J Heart Fail. Dec 2016;18(12):1420–1429. doi:10.1002/ejhf.678

62. Duran-Aniotz C, Hetz C. Glucose Metabolism: A Sweet Relief of Alzheimer’s Disease. Curr Biol. Sep 12 2016;26(17):R806–9. doi:10.1016/j.cub.2016.07.060

63. Zhang X, Hu W, Wang Y, et al. Plasma metabolomic profiles of dementia: a prospective study of 110,655 participants in the UK Biobank. BMC Med. Aug 15 2022;20(1):252. doi:10.1186/s12916-022-02449-3

64. Tynkkynen J, Chouraki V, van der Lee SJ, et al. Association of branched-chain amino acids and other circulating metabolites with risk of incident dementia and Alzheimer’s disease: A prospective study in eight cohorts. Alzheimers Dement. Jun 2018;14(6):723–733. doi:10.1016/j.jalz.2018.01.003

65. Amin N, Liu J, Sproviero W, et al. Interplay between age, APOE E4 and the metabolome in plasma and brain in Alzheimer’s disease. Transl Psychiatry. Oct 31 2025;15(1):460. doi:10.1038/s41398-025-03625-8

66. Lacruz ME, Kluttig A, Tiller D, et al. Instability of personal human metabotype is linked to all-cause mortality. Sci Rep. Jun 28 2018;8(1):9810. doi:10.1038/s41598-018-27958-1

67. Buondonno I, Sassi F, Carignano G, et al. From mitochondria to healthy aging: The role of branched-chain amino acids treatment: MATeR a randomized study. Clin Nutr. Jul 2020;39(7):2080–2091. doi:10.1016/j.clnu.2019.10.013

68. Barazzoni R, Short KR, Nair KS. Effects of aging on mitochondrial DNA copy number and cytochrome c oxidase gene expression in rat skeletal muscle, liver, and heart. J Biol Chem. Feb 4 2000;275(5):3343–7. doi:10.1074/jbc.275.5.3343

69. Valerio A, D’Antona G, Nisoli E. Branched-chain amino acids, mitochondrial biogenesis, and healthspan: an evolutionary perspective. Aging (Albany NY). May 2011;3(5):464–78. doi:10.18632/aging.100322

70. Tan HC, Hsu JW, Kovalik JP, et al. Branched-Chain Amino Acid Oxidation Is Elevated in Adults with Morbid Obesity and Decreases Significantly after Sleeve Gastrectomy. J Nutr. Dec 10 2020;150(12):3180–3189. doi:10.1093/jn/nxaa298

71. Vanweert F, de Ligt M, Hoeks J, Hesselink MKC, Schrauwen P, Phielix E. Elevated Plasma Branched-Chain Amino Acid Levels Correlate With Type 2 Diabetes-Related Metabolic Disturbances. J Clin Endocrinol Metab. Mar 25 2021;106(4):e1827–e1836. doi:10.1210/clinem/dgaa751

72. Juricic P, Gronke S, Partridge L. Branched-Chain Amino Acids Have Equivalent Effects to Other Essential Amino Acids on Lifespan and Aging-Related Traits in Drosophila. J Gerontol A Biol Sci Med Sci. Jan 1 2020;75(1):24–31. doi:10.1093/gerona/glz080

73. Tramutola A, Triplett JC, Di Domenico F, et al. Alteration of mTOR signaling occurs early in the progression of Alzheimer disease (AD): analysis of brain from subjects with pre-clinical AD, amnestic mild cognitive impairment and late-stage AD. J Neurochem. Jun 2015;133(5):739–49. doi:10.1111/jnc.13037

74. Di Domenico F, Tramutola A, Foppoli C, Head E, Perluigi M, Butterfield DA. mTOR in Down syndrome: Role in Ass and tau neuropathology and transition to Alzheimer disease-like dementia. Free Radic Biol Med. Jan 2018;114:94–101. doi:10.1016/j.freeradbiomed.2017.08.009

75. Holecek M. Branched-chain amino acids in health and disease: metabolism, alterations in blood plasma, and as supplements. Nutr Metab (Lond). 2018/12//2018;15(1):33. doi:10.1186/s12986-018-0271-1

76. Judd JM, Jasbi P, Winslow W, et al. Inflammation and the pathological progression of Alzheimer’s disease are associated with low circulating choline levels. Acta Neuropathol. Oct 2023;146(4):565–583. doi:10.1007/s00401-023-02616-7

77. Lanari A, Amenta F, Silvestrelli G, Tomassoni D, Parnetti L. Neurotransmitter deficits in behavioural and psychological symptoms of Alzheimer’s disease. Mech Ageing Dev. Feb 2006;127(2):158–65. doi:10.1016/j.mad.2005.09.016

78. Allen DD, Smith QR. Characterization of the blood-brain barrier choline transporter using the in situ rat brain perfusion technique. J Neurochem. Feb 2001;76(4):1032–41. doi:10.1046/j.1471-4159.2001.00093.x

79. Wang R, Reddy PH. Role of Glutamate and NMDA Receptors in Alzheimer’s Disease. J Alzheimers Dis. 2017/4//2017;57(4):1041–1048. doi:10.3233/JAD-160763

80. Salcedo C, Andersen JV, Vinten KT, et al. Functional Metabolic Mapping Reveals Highly Active Branched-Chain Amino Acid Metabolism in Human Astrocytes, Which Is Impaired in iPSC-Derived Astrocytes in Alzheimer’s Disease. Front Aging Neurosci. 2021/9//2021;13:736580. doi:10.3389/fnagi.2021.736580

81. Seshadri S, Beiser A, Selhub J, et al. Plasma homocysteine as a risk factor for dementia and Alzheimer’s disease. N Engl J Med. Feb 14 2002;346(7):476–83. doi:10.1056/NEJMoa011613

82. Rahman ML, Shu XO, Jones DP, et al. A nested case-control study of untargeted plasma metabolomics and lung cancer among never-smoking women within the prospective Shanghai Women’s Health Study. Int J Cancer. Aug 1 2024;155(3):508–518. doi:10.1002/ijc.34929

83. Toyoshima K, Nakamura M, Adachi Y, et al. Increased plasma proline concentrations are associated with sarcopenia in the elderly. PLoS One. 2017/9//2017;12(9):e0185206. doi:10.1371/journal.pone.0185206

84. Francois M, Karpe AV, Liu JW, et al. Multi-Omics, an Integrated Approach to Identify Novel Blood Biomarkers of Alzheimer’s Disease. Metabolites. Oct 6 2022;12(10):949–949. doi:10.3390/metabo12100949

85. Corso G, Cristofano A, Sapere N, et al. Serum Amino Acid Profiles in Normal Subjects and in Patients with or at Risk of Alzheimer Dementia. Dement Geriatr Cogn Dis Extra. Jan–Apr 2017;7(1):143–159. doi:10.1159/000466688

86. Conde R, Oliveira N, Morais E, et al. NMR analysis seeking for cognitive decline and dementia metabolic markers in plasma from aged individuals. J Pharm Biomed Anal. Jan 20 2024;238(115815):115815. doi:10.1016/j.jpba.2023.115815

87. Liu P, Yang Q, Yu N, et al. Phenylalanine Metabolism Is Dysregulated in Human Hippocampus with Alzheimer’s Disease Related Pathological Changes. J Alzheimers Dis. 2021;83(2):609–622. doi:10.3233/JAD-210461

88. Cui M, Jiang Y, Zhao Q, et al. Metabolomics and incident dementia in older Chinese adults: The Shanghai Aging Study. Alzheimers Dement. May 2020;16(5):779–788. doi:10.1002/alz.12074

89. Wang F, Wang G, Li W, Xu C, Zeng Z, Zhou Y. Analysis of serum metabolism in premature infants before and after feeding using GC-MS and the relationship with necrotizing enterocolitis. Biomed Chromatogr. Jan 2023;37(1):e5505. doi:10.1002/bmc.5505

90. Murtas G, Marcone GL, Sacchi S, Pollegioni L. L-serine synthesis via the phosphorylated pathway in humans. Cell Mol Life Sci. Dec 2020;77(24):5131–5148. doi:10.1007/s00018-020-03574-z

91. Zhu Z, Shi Z, Xie C, Gong W, Hu Z, Peng Y. A novel mechanism of Gamma-aminobutyric acid (GABA) protecting human umbilical vein endothelial cells (HUVECs) against H(2)O(2)-induced oxidative injury. Comp Biochem Physiol C Toxicol Pharmacol. Mar 2019;217:68–75. doi:10.1016/j.cbpc.2018.11.018

92. Xu J, Green R, Kim M, et al. Sex-Specific Metabolic Pathways Were Associated with Alzheimer’s Disease (AD) Endophenotypes in the European Medical Information Framework for AD Multimodal Biomarker Discovery Cohort. Biomedicines. Nov 3 2021;9(11):1610–1610. doi:10.3390/biomedicines9111610

93. Rebholz CM, Lichtenstein AH, Zheng Z, Appel LJ, Coresh J. Serum untargeted metabolomic profile of the Dietary Approaches to Stop Hypertension (DASH) dietary pattern. Am J Clin Nutr. Aug 1 2018;108(2):243–255. doi:10.1093/ajcn/nqy099

94. Chen H, Shen J, Tao Y, et al. Circulating metabolomic profile of the MIND diet and its relation to cognition in middle-aged and older adults. Alzheimer’s & Dementia. 2024/12//2025;20(S7)doi:10.1002/alz.088163

95. Zhan M, Li Z, Chen J, et al. Indoxyl sulfate (IS) mediates pro-inflammatory responses in severe pneumonia in patients with rheumatoid arthritis associated interstitial lung disease. Clin Immunol. Mar 2025;272(110430):110430. doi:10.1016/j.clim.2025.110430

96. Stepanova N, Driianska V, Korol L, Snisar L. Association between serum total indoxyl sulfate, intraperitoneal inflammation, and peritoneal dialysis technique failure: a 3-year prospective cohort study. BMC Nephrol. Dec 31 2024;25(1):475. doi:10.1186/s12882-024-03935-x

97. Hendrikx T, Schnabl B. Indoles: metabolites produced by intestinal bacteria capable of controlling liver disease manifestation. J Intern Med. Jul 2019;286(1):32–40. doi:10.1111/joim.12892

98. Bendheim PE, Poeggeler B, Neria E, Ziv V, Pappolla MA, Chain DG. Development of indole-3-propionic acid (OXIGON) for Alzheimer’s disease. J Mol Neurosci. Aug–Oct 2002;19(1-2):213–7. doi:10.1007/s12031-002-0036-0

99. Pappolla MA, Perry G, Fang X, Zagorski M, Sambamurti K, Poeggeler B. Indoles as essential mediators in the gut-brain axis. Their role in Alzheimer’s disease. Neurobiol Dis. Aug 2021;156:105403. doi:10.1016/j.nbd.2021.105403

100. Sathyasaikumar KV, Blanco-Ayala T, Zheng Y, et al. The Tryptophan Metabolite Indole-3-Propionic Acid Raises Kynurenic Acid Levels in the Rat Brain In Vivo. Int J Tryptophan Res. 2024/1//2024;17:11786469241262876. doi:10.1177/11786469241262876

101. Ma YY, Li X, Yu JT, Wang YJ. Therapeutics for neurodegenerative diseases by targeting the gut microbiome: from bench to bedside. Transl Neurodegener. Feb 27 2024;13(1):12. doi:10.1186/s40035-024-00404-1

102. Feliers D, Lee DY, Gorin Y, Kasinath BS. Symmetric dimethylarginine alters endothelial nitric oxide activity in glomerular endothelial cells. Cell Signal. Jan 2015;27(1):1–5. doi:10.1016/j.cellsig.2014.09.024

103. Tyagi N, Sedoris KC, Steed M, Ovechkin AV, Moshal KS, Tyagi SC. Mechanisms of homocysteine-induced oxidative stress. Am J Physiol Heart Circ Physiol. Dec 2005;289(6):H2649–56. doi:10.1152/ajpheart.00548.2005

104. Gokkusu C, Tulubas F, Unlucerci Y, Ozkok E, Umman B, Aydin M. Homocysteine and pro-inflammatory cytokine concentrations in acute heart disease. Cytokine. Apr 2010;50(1):15–8. doi:10.1016/j.cyto.2009.12.015

105. Smith AD, Refsum H, Bottiglieri T, et al. Homocysteine and Dementia: An International Consensus Statement. J Alzheimers Dis. 2018;62(2):561–570. doi:10.3233/JAD-171042

106. Chen T, Pan F, Huang Q, et al. Metabolic phenotyping reveals an emerging role of ammonia abnormality in Alzheimer’s disease. Nat Commun. May 7 2024;15(1):3796. doi:10.1038/s41467-024-47897-y

107. Ren Z, Zhao L, Zhao M, et al. Increased intestinal bile acid absorption contributes to age-related cognitive impairment. Cell Rep Med. May 21 2024;5(5):101543. doi:10.1016/j.xcrm.2024.101543

108. Matsumoto S, Haberle J, Kido J, Mitsubuchi H, Endo F, Nakamura K. Urea cycle disorders-update. J Hum Genet. Sep 2019;64(9):833–847. doi:10.1038/s10038-019-0614-4

109. Braissant O. Ammonia toxicity to the brain: effects on creatine metabolism and transport and protective roles of creatine. Mol Genet Metab. 2010;100 Suppl 1:S53–8. doi:10.1016/j.ymgme.2010.02.011

110. Butterfield DA, Hensley K, Cole P, et al. Oxidatively induced structural alteration of glutamine synthetase assessed by analysis of spin label incorporation kinetics: relevance to Alzheimer’s disease. J Neurochem. Jun 1997;68(6):2451–7. doi:10.1046/j.1471-4159.1997.68062451.x

111. Moreira P, Honda K, Liu Q, et al. Alzheimers Disease and Oxidative Stress: The Old Problem Remains Unsolved. Current Medicinal Chemistry-Central Nervous System Agents. 2005;5(1):51–62. doi:10.2174/1568015053202714

112. Rehman T, Shabbir MA, Inam-Ur-Raheem M, et al. Cysteine and homocysteine as biomarker of various diseases. Food Sci Nutr. Sep 2020;8(9):4696–4707. doi:10.1002/fsn3.1818

113. Ezerina D, Takano Y, Hanaoka K, Urano Y, Dick TP. N-Acetyl Cysteine Functions as a Fast-Acting Antioxidant by Triggering Intracellular H(2)S and Sulfane Sulfur Production. Cell Chem Biol. Apr 19 2018;25(4):447–459 e4. doi:10.1016/j.chembiol.2018.01.011

114. Le Chatelier E, Nielsen T, Qin J, et al. Richness of human gut microbiome correlates with metabolic markers. Nature. Aug 29 2013;500(7464):541–6. doi:10.1038/nature12506

115. Bell LK, Edwards S, Grieger JA. The Relationship between Dietary Patterns and Metabolic Health in a Representative Sample of Adult Australians. Nutrients. Aug 5 2015;7(8):6491–505. doi:10.3390/nu7085295

