## Supplementary Figure S1 for "A seven-year longitudinal study of the Alzheimer’s disease blood metabolome"

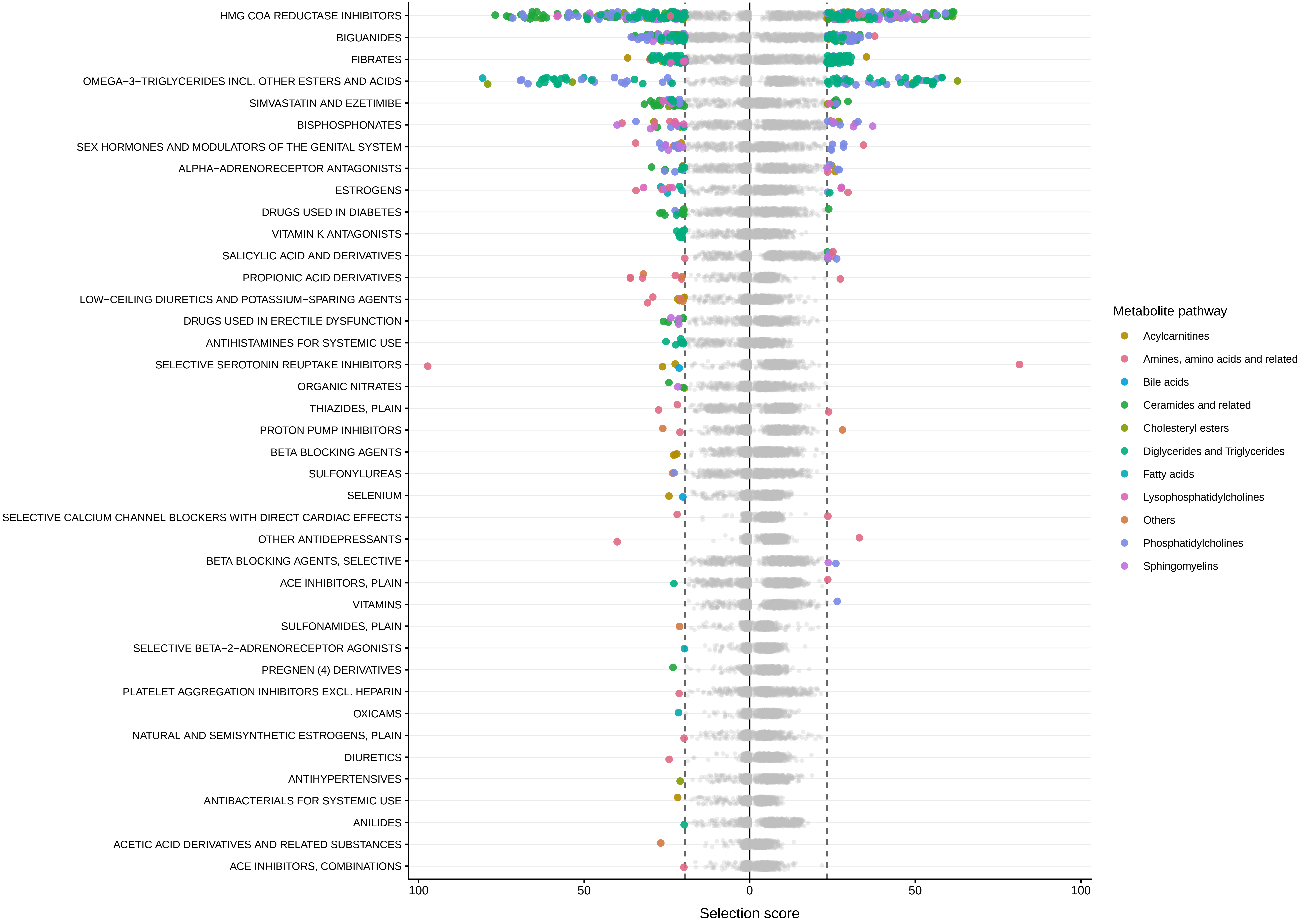

HMG COA REDUCTASE INHIBITORS
BIGUANIDES
FIBRATES
OMEGA−3−TRIGLYCERIDES INCL. OTHER ESTERS AND ACIDS
SIMVASTATIN AND EZETIMIBE
BISPHOSPHONATES
SEX HORMONES AND MODULATORS OF THE GENITAL SYSTEM
ALPHA−ADRENORECEPTOR ANTAGONISTS
ESTROGENS
DRUGS USED IN DIABETES
VITAMIN K ANTAGONISTS
SALICYLIC ACID AND DERIVATIVES
PROPIONIC ACID DERIVATIVES
LOW−CEILING DIURETICS AND POTASSIUM−SPARING AGENTS
DRUGS USED IN ERECTILE DYSFUNCTION
ANTIHISTAMINES FOR SYSTEMIC USE
SELECTIVE SEROTONIN REUPTAKE INHIBITORS
ORGANIC NITRATES
THIAZIDES, PLAIN
PROTON PUMP INHIBITORS
BETA BLOCKING AGENTS
SULFONYLUREAS
SELENIUM
SELECTIVE CALCIUM CHANNEL BLOCKERS WITH DIRECT CARDIAC EFFECTS
OTHER ANTIDEPRESSANTS
BETA BLOCKING AGENTS, SELECTIVE
ACE INHIBITORS, PLAIN
VITAMINS
SULFONAMIDES, PLAIN
SELECTIVE BETA−2−ADRENORECEPTOR AGONISTS
PREGNEN (4) DERIVATIVES
PLATELET AGGREGATION INHIBITORS EXCL. HEPARIN
OXICAMS
NATURAL AND SEMISYNTHETIC ESTROGENS, PLAIN
DIURETICS
ANTIHYPERTENSIVES
ANTIBACTERIALS FOR SYSTEMIC USE
ANILIDES
ACETIC ACID DERIVATIVES AND RELATED SUBSTANCES
ACE INHIBITORS, COMBINATIONS
100
50
0
50
100
Selection score
Metabolite pathway
Acylcarnitines
Amines, amino acids and related
Bile acids
Ceramides and related
Cholesteryl esters
Diglycerides and Triglycerides
Fatty acids
Lysophosphatidylcholines
Others
Phosphatidylcholines
Sphingomyelins

**Supplementary Figure S1: Biocrates MXP® Quant 500 metabolite-medication associations identified by the Boruta random forest algorithm.**
Medication classes are shown on the y-axis and selection scores on the x-axis, with baseline results plotted to the left of zero and longitudinal results to the right.
Each point represents a metabolite-medication pair. Selected relationships are colored by metabolite class or pathway, whereas unselected relationships are shown in grey.
The exact statistics of these associations can be found in **Supplementary Tables S4-S5.**
