## Supplementary Figure S2 for "A seven-year longitudinal study of the Alzheimer’s disease blood metabolome"

**Supplementary Figure S2: Effect plots for the strongest metabolite-phenotype associations across baseline, multi-timepoint, and time-interaction models.** This multi-page figure (starting from the next page) presents effect plots summarizing the strongest associations identified across three analyses: baseline, multi-timepoint, and time-interaction analyses. For each page, the y-axis reflects the appropriate outcome measure for that model: residualized metabolite levels for the baseline analyses (also shown for 12 and 24 months), whitened metabolite levels for the multi-timepoint analysis, and estimated longitudinal effects for the time-interaction models. The x-axis denotes time for the baseline and time-interaction plots, whereas for the multitimepoint analysis represents the evaluated group comparison. Continuous phenotypes were median-split at baseline to create grouping variables. Exact summary statistics for all the said analyses can be found in **Supplementary Table S6.**

### 3−IPA

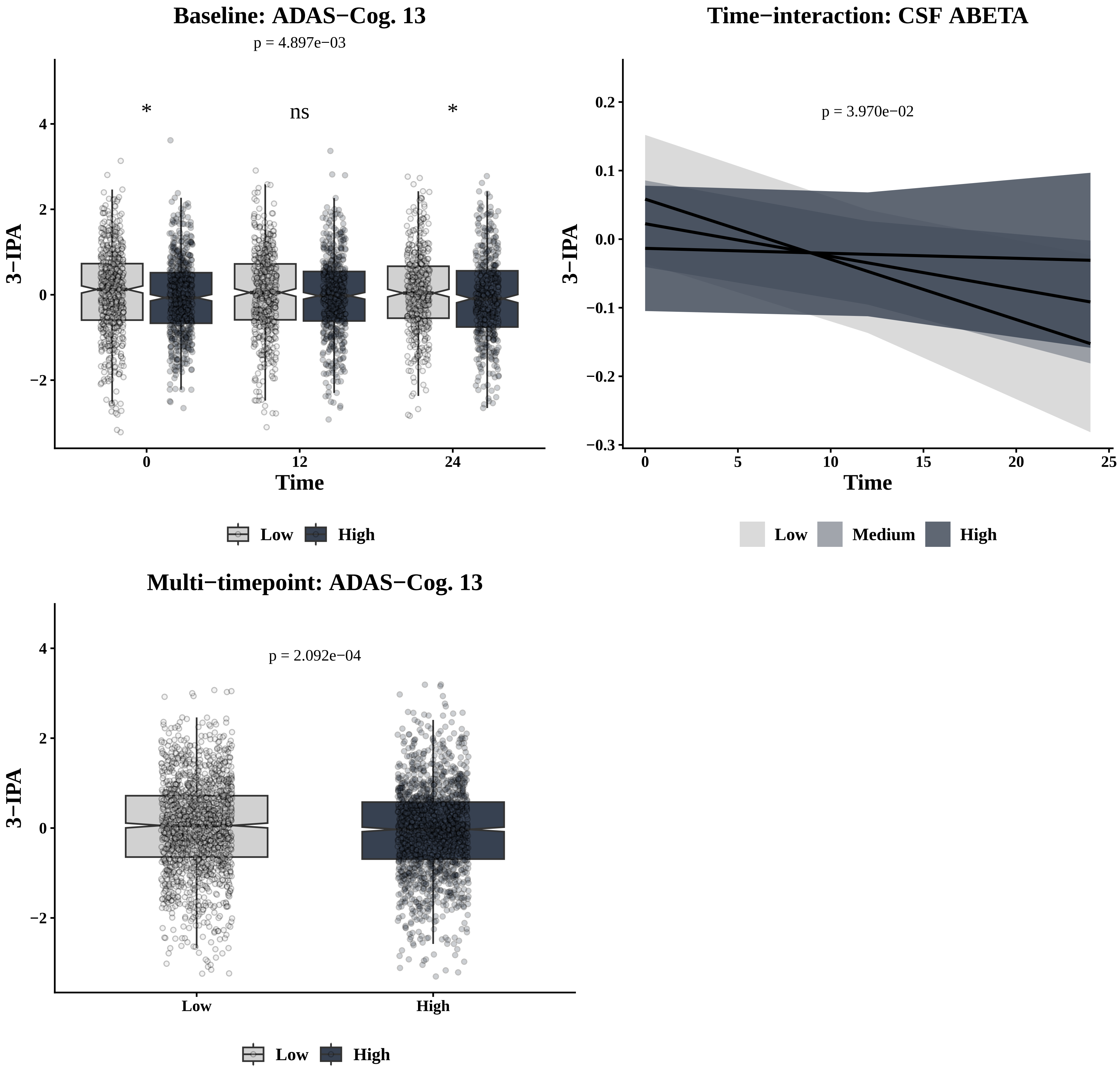

Baseline: ADAS−Cog. 13
p = 4.897e−03
*
ns
*
3−IPA
Time
Low
High
Time−interaction: CSF ABETA
p = 3.970e−02
3−IPA
Time
Low
Medium
High
Multi−timepoint: ADAS−Cog. 13
p = 2.092e−04
3−IPA
Low
High

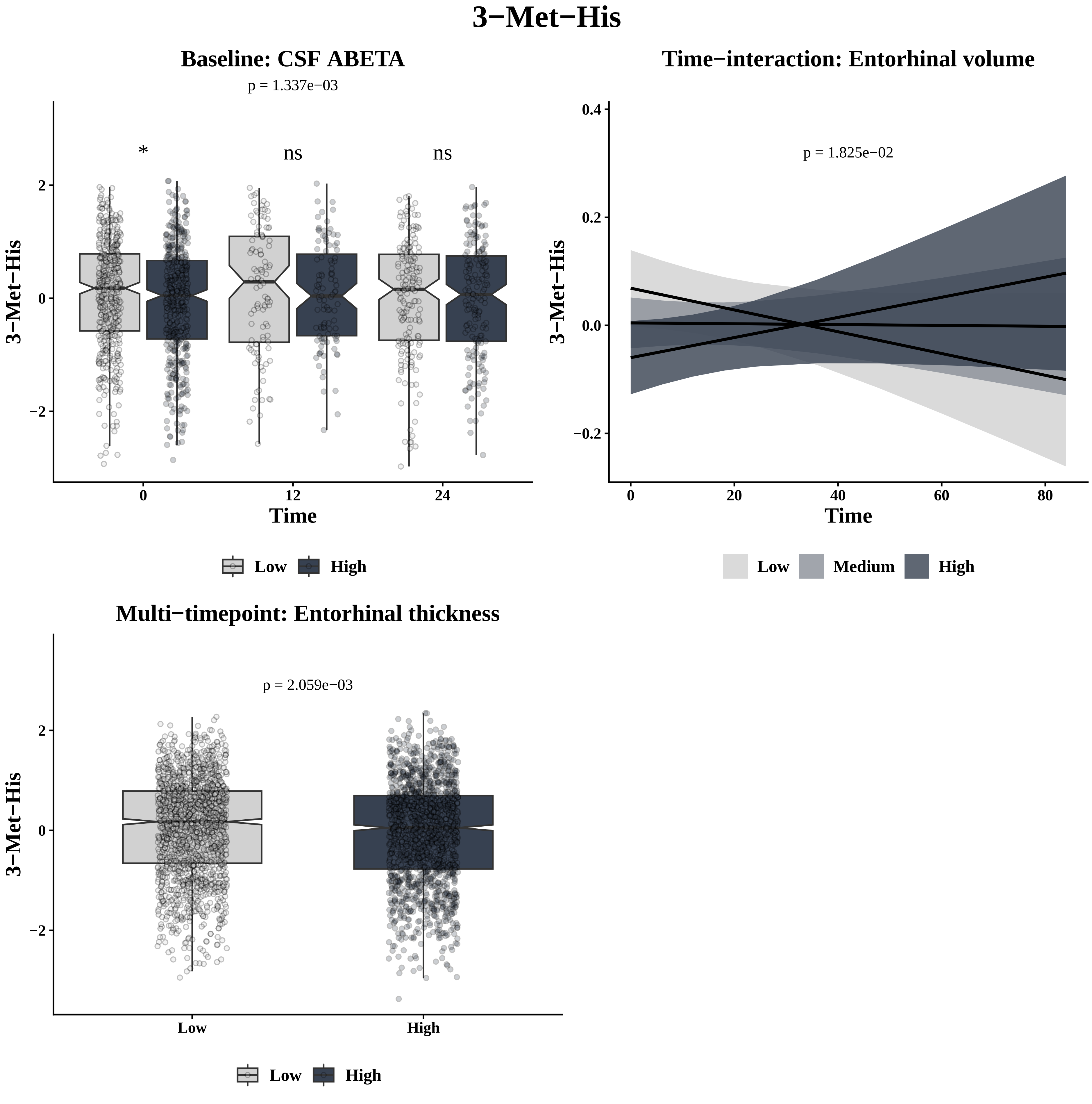
3−Met−His
Baseline: CSF ABETA
p = 1.337e−03
*
ns
ns
3−Met−His
2
0
−2
0
12
24
Time
Low
High
Time−interaction: Entorhinal volume
p = 1.825e−02
3−Met−His
0.4
0.2
0.0
−0.2
0
20
40
60
80
Time
Low
Medium
High
Multi−timepoint: Entorhinal thickness
p = 2.059e−03
3−Met−His
2
0
−2
Low
High
Low
High

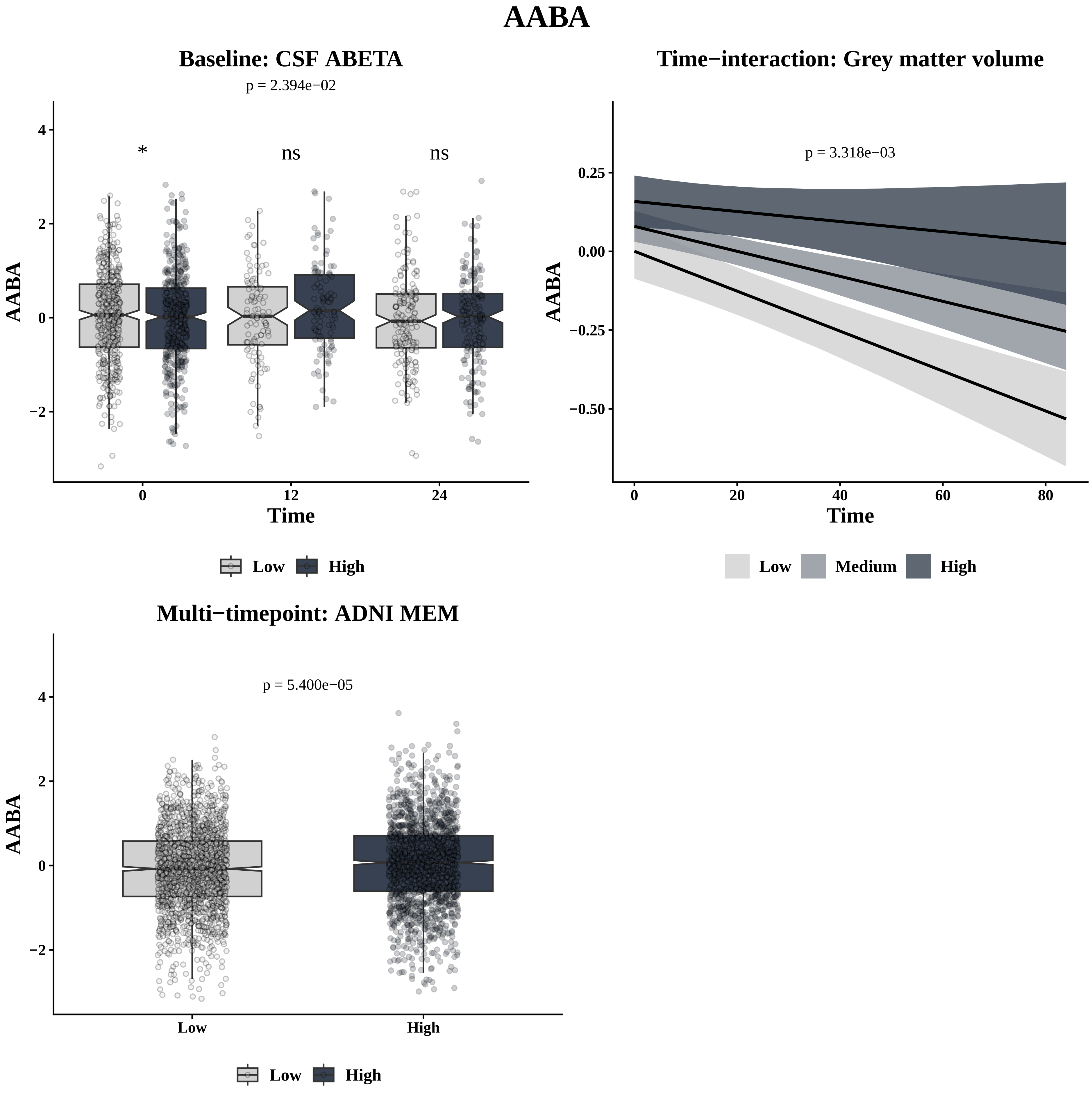
AABA
Baseline: CSF ABETA
p = 2.394e−02
*
ns
ns
AABA
4
2
0
−2
0
12
24
Time
Low
High
Time−interaction: Grey matter volume
p = 3.318e−03
AABA
0.25
0.00
−0.25
−0.50
0
20
40
60
80
Time
Low
Medium
High
Multi−timepoint: ADNI MEM
p = 5.400e−05
AABA
4
2
0
−2
Low
High
Low
High

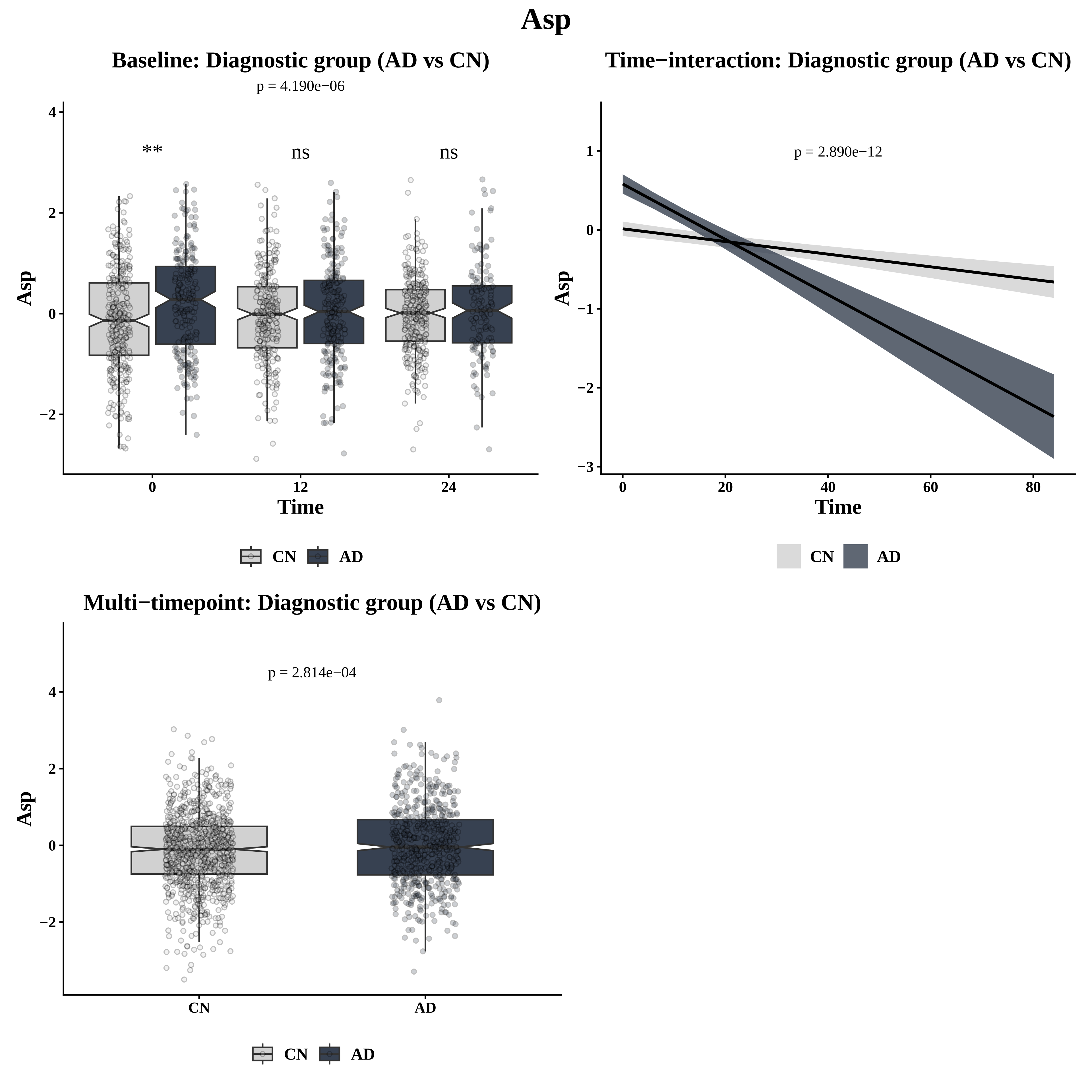
Asp
Baseline: Diagnostic group (AD vs CN)
p = 4.190e−06
**
ns
ns
Asp
Time
0
12
24
4
2
0
−2
CN
AD
Time−interaction: Diagnostic group (AD vs CN)
p = 2.890e−12
Asp
Time
0
20
40
60
80
1
0
−1
−2
−3
CN
AD
Multi−timepoint: Diagnostic group (AD vs CN)
p = 2.814e−04
Asp
4
2
0
−2
CN
AD
CN
AD

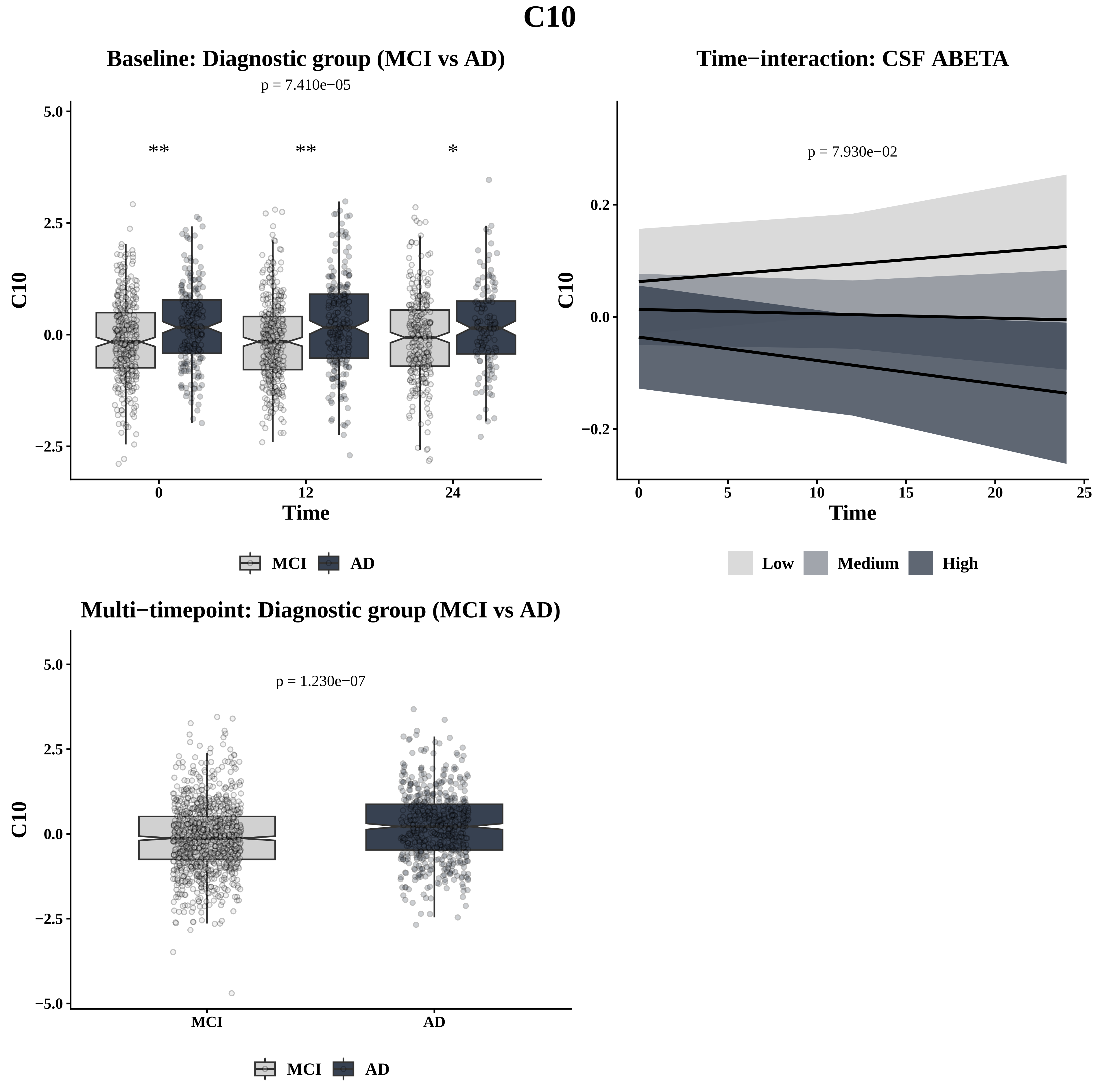
C10
Baseline: Diagnostic group (MCI vs AD)
p = 7.410e−05
**
**
*
C10
Time
MCI
AD
Time−interaction: CSF ABETA
p = 7.930e−02
C10
Time
Low
Medium
High
Multi−timepoint: Diagnostic group (MCI vs AD)
p = 1.230e−07
C10
MCI
AD
MCI
AD

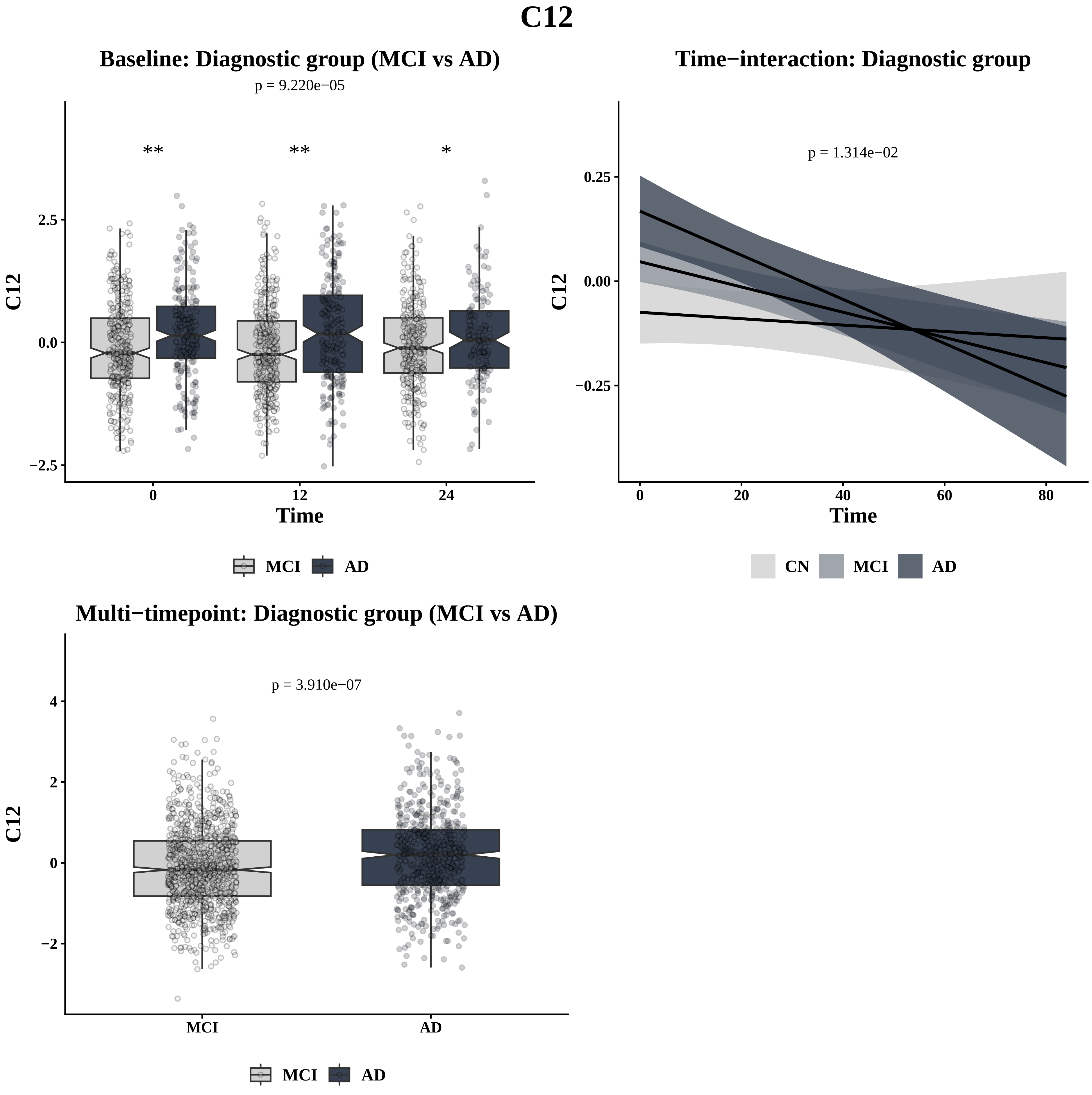
C12
Baseline: Diagnostic group (MCI vs AD)
p = 9.220e−05
**
**
*
2.5
0.0
−2.5
C12
0
12
24
Time
MCI
AD
Time−interaction: Diagnostic group
p = 1.314e−02
0.25
0.00
−0.25
C12
0
20
40
60
80
Time
CN
MCI
AD
Multi−timepoint: Diagnostic group (MCI vs AD)
p = 3.910e−07
4
2
0
−2
C12
MCI
AD
MCI
AD

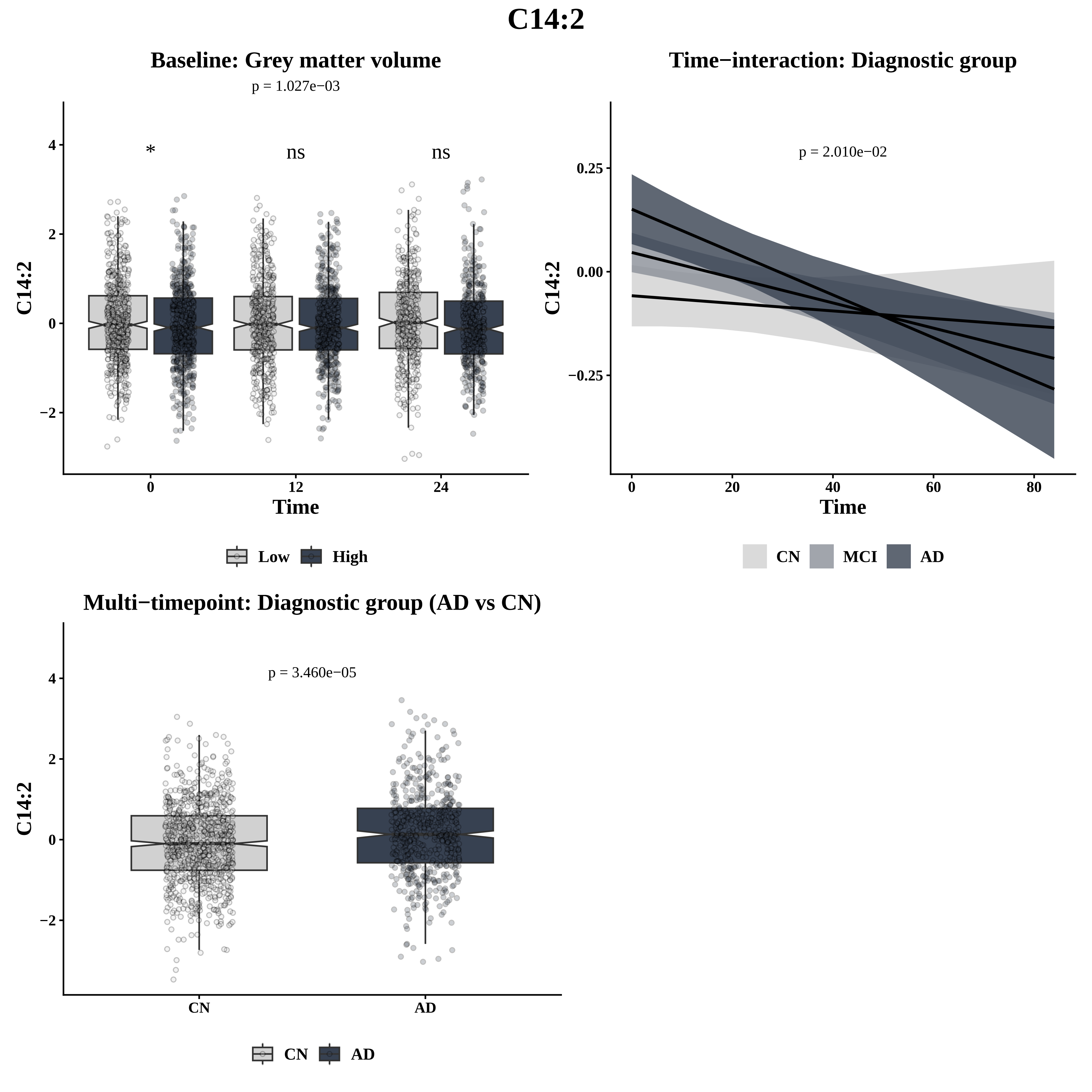
C14:2
Baseline: Grey matter volume
p = 1.027e−03
*
ns
ns
C14:2
Time
0
12
24
Low
High
Time−interaction: Diagnostic group
p = 2.010e−02
0.25
0.00
−0.25
C14:2
Time
0
20
40
60
80
CN
MCI
AD
Multi−timepoint: Diagnostic group (AD vs CN)
p = 3.460e−05
C14:2
CN
AD
CN
AD

# C16

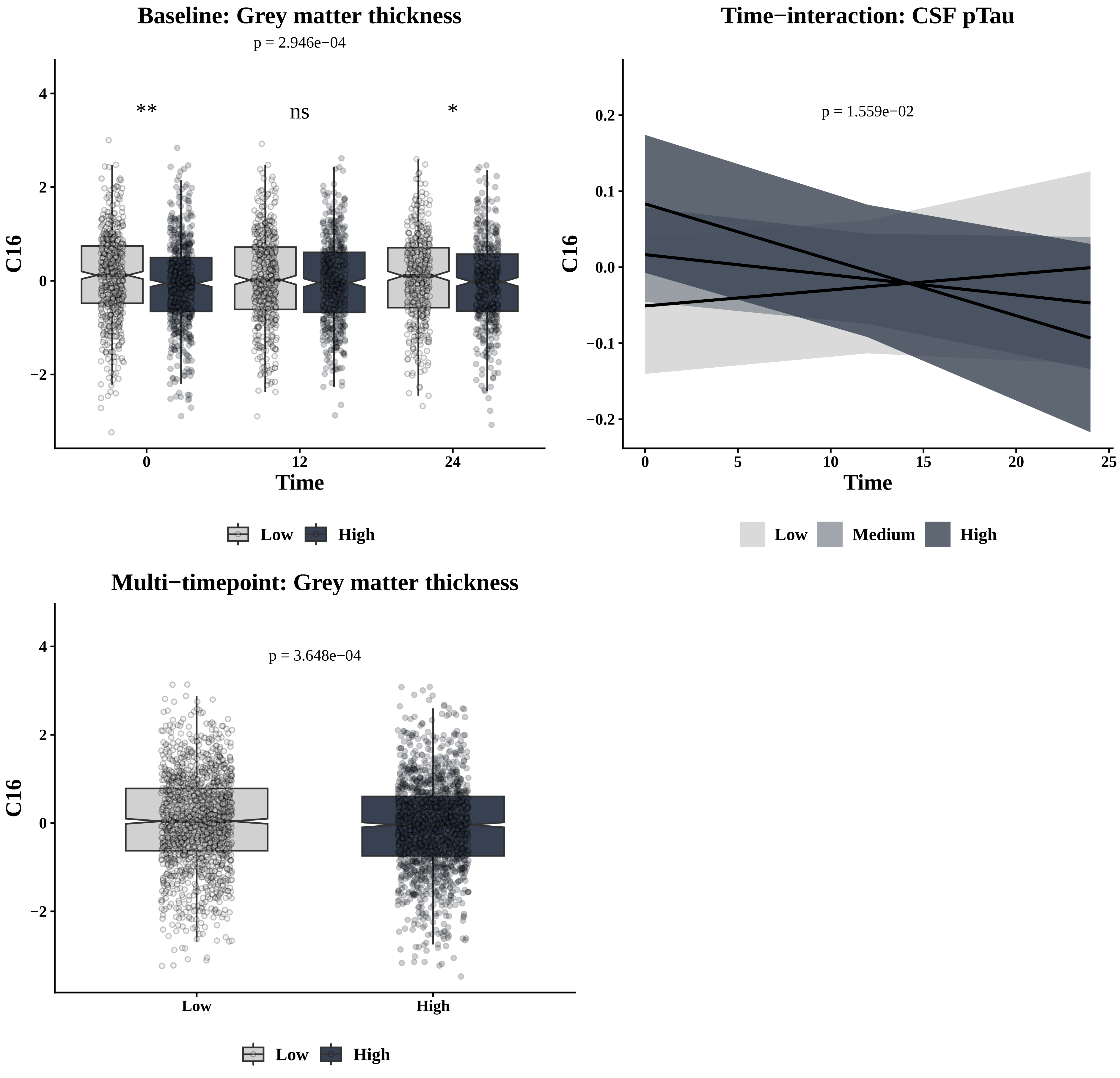

Baseline: Grey matter thickness
p = 2.946e−04
**
ns
*
C16
4
2
0
−2
0
12
24
Time
Low
High
Time−interaction: CSF pTau
p = 1.559e−02
C16
0.2
0.1
0.0
−0.1
−0.2
0
5
10
15
20
25
Time
Low
Medium
High
Multi−timepoint: Grey matter thickness
p = 3.648e−04
C16
4
2
0
−2
Low
High
Low
High

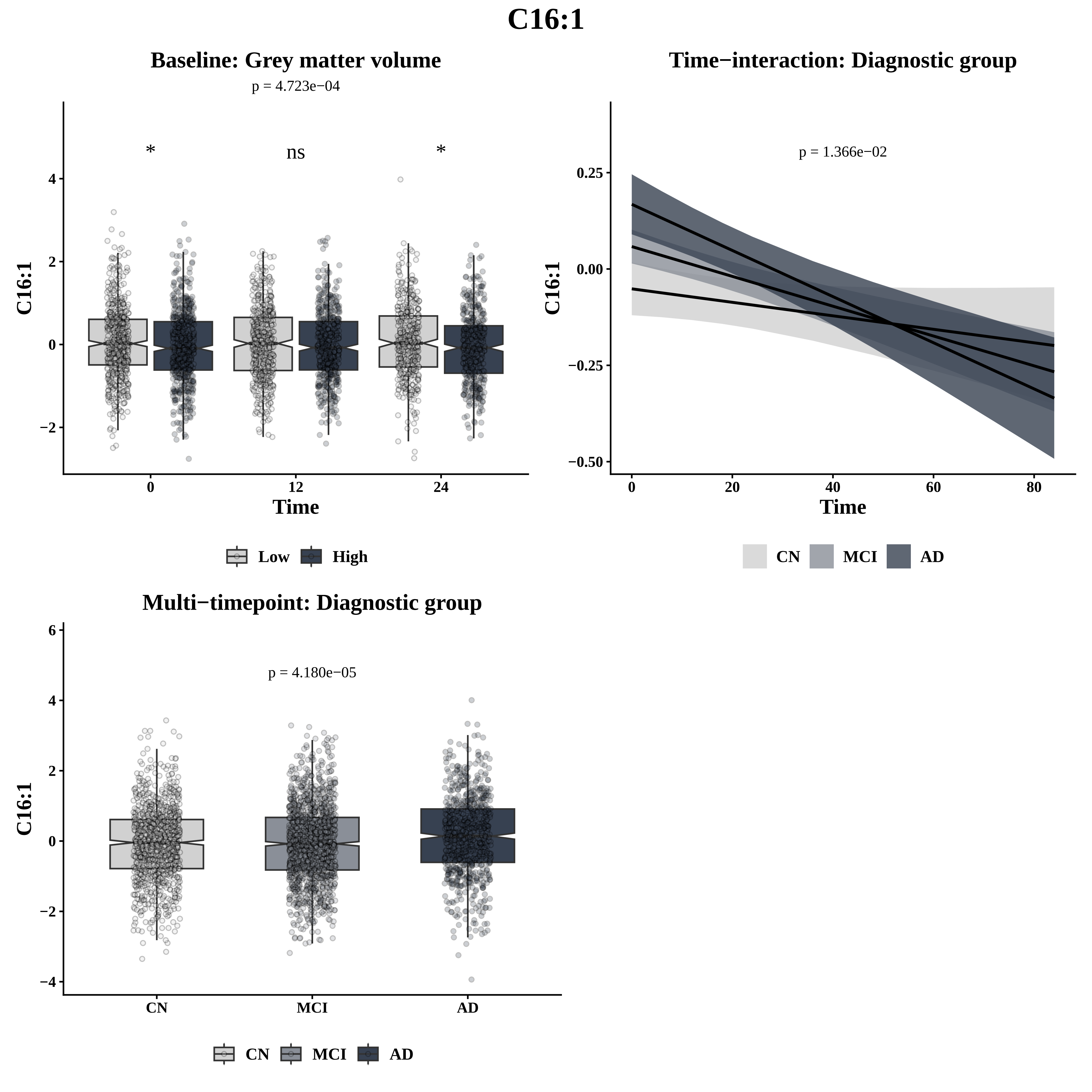
C16:1
Baseline: Grey matter volume
p = 4.723e−04
*
ns
*
C16:1
Time
0
12
24
Low
High
Time−interaction: Diagnostic group
p = 1.366e−02
C16:1
Time
CN
MCI
AD
Multi−timepoint: Diagnostic group
p = 4.180e−05
C16:1
CN
MCI
AD
CN
MCI
AD

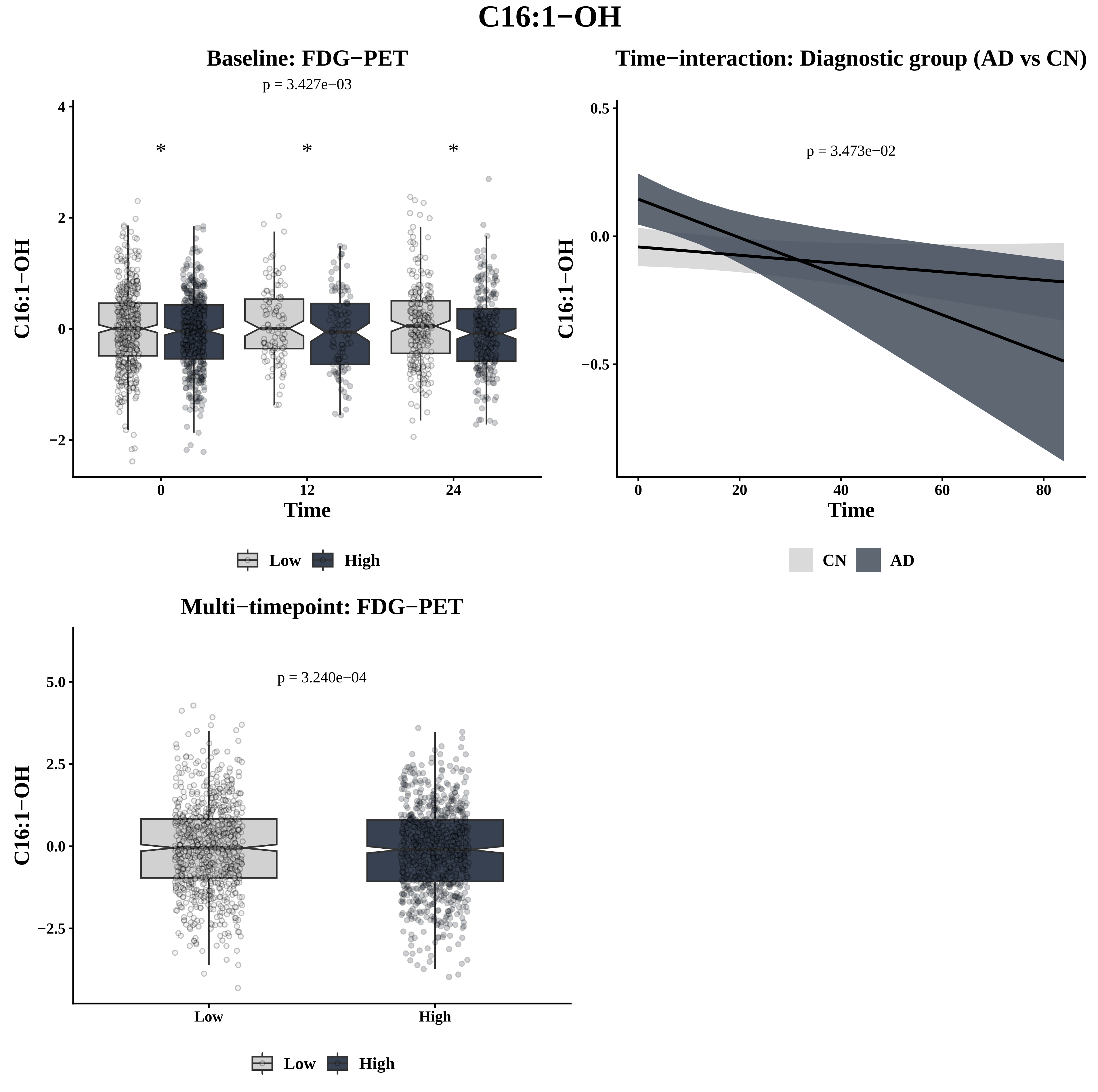

C16:1−OH
Baseline: FDG−PET
p = 3.427e−03
*
*
*
C16:1−OH
Time
0
12
24
4
2
0
−2
Low
High
Time−interaction: Diagnostic group (AD vs CN)
p = 3.473e−02
C16:1−OH
Time
0.5
0.0
−0.5
0
20
40
60
80
CN
AD
Multi−timepoint: FDG−PET
p = 3.240e−04
C16:1−OH
5.0
2.5
0.0
−2.5
Low
High
Low
High

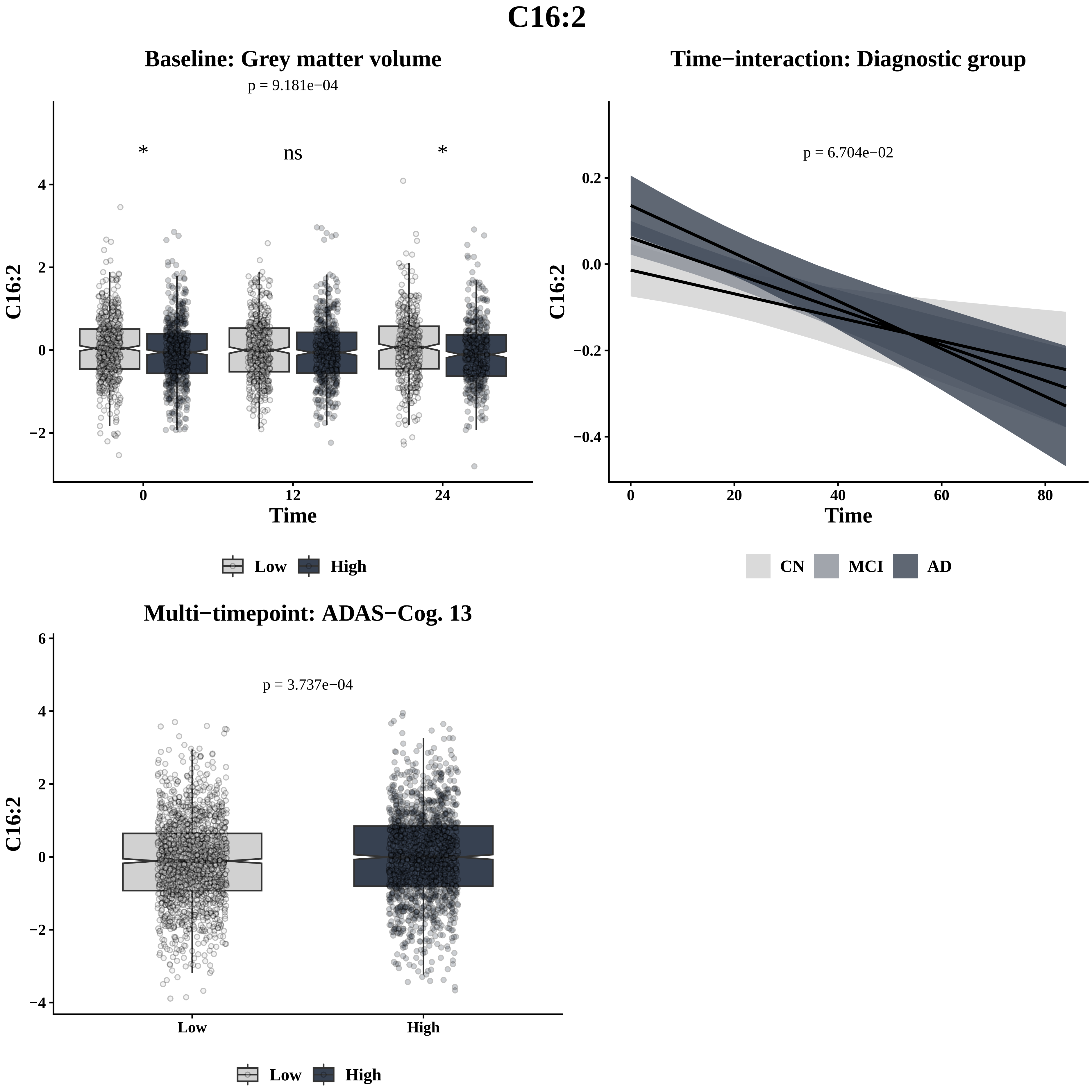
C16:2
Baseline: Grey matter volume
p = 9.181e−04
*
ns
*
C16:2
Time
Low
High
Time−interaction: Diagnostic group
p = 6.704e−02
C16:2
Time
CN
MCI
AD
Multi−timepoint: ADAS−Cog. 13
p = 3.737e−04
C16:2
Low
High
Low
High

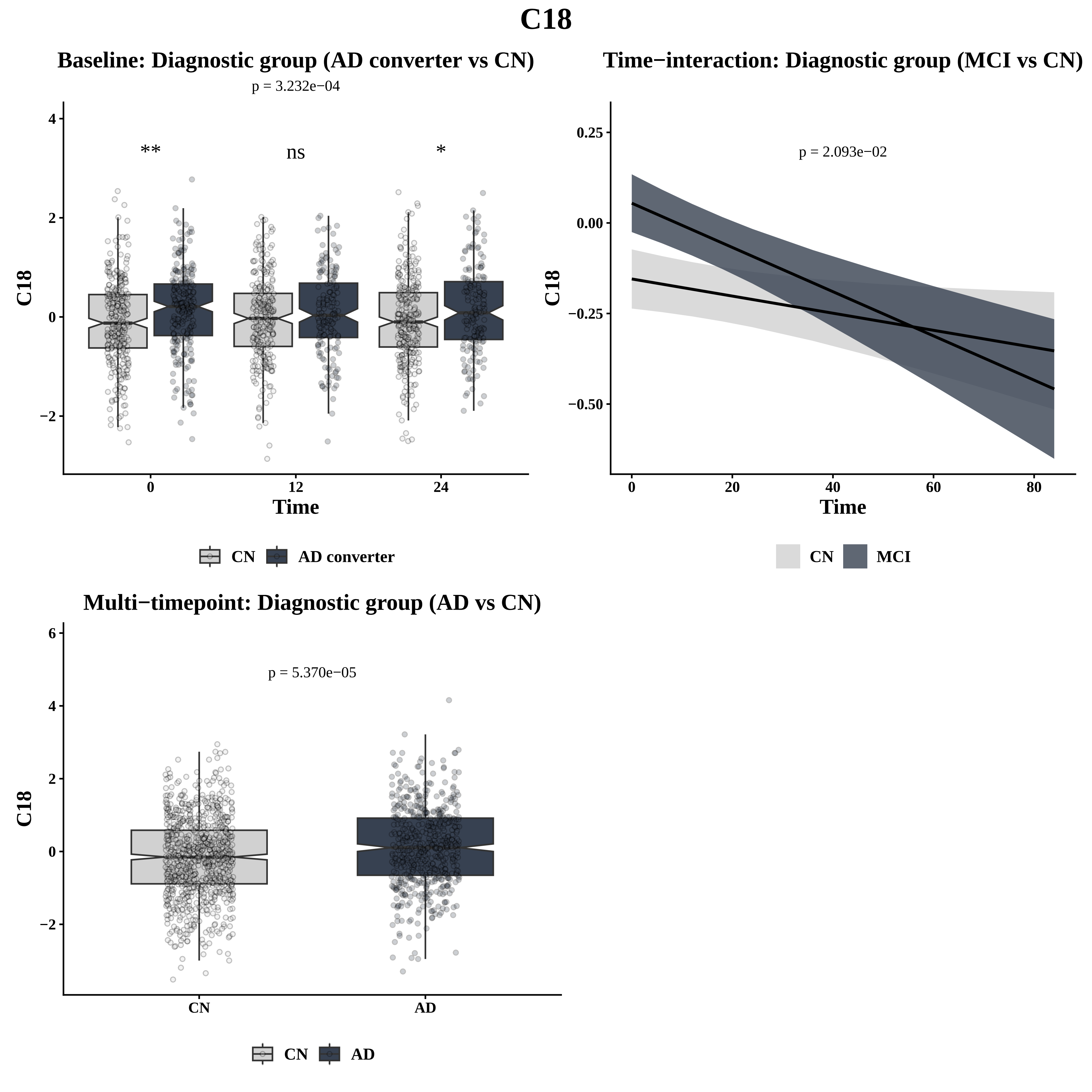

C18
Baseline: Diagnostic group (AD converter vs CN)
p = 3.232e−04
**
ns
*
C18
Time
CN
AD converter
Time−interaction: Diagnostic group (MCI vs CN)
p = 2.093e−02
C18
Time
CN
MCI
Multi−timepoint: Diagnostic group (AD vs CN)
p = 5.370e−05
C18
CN
AD

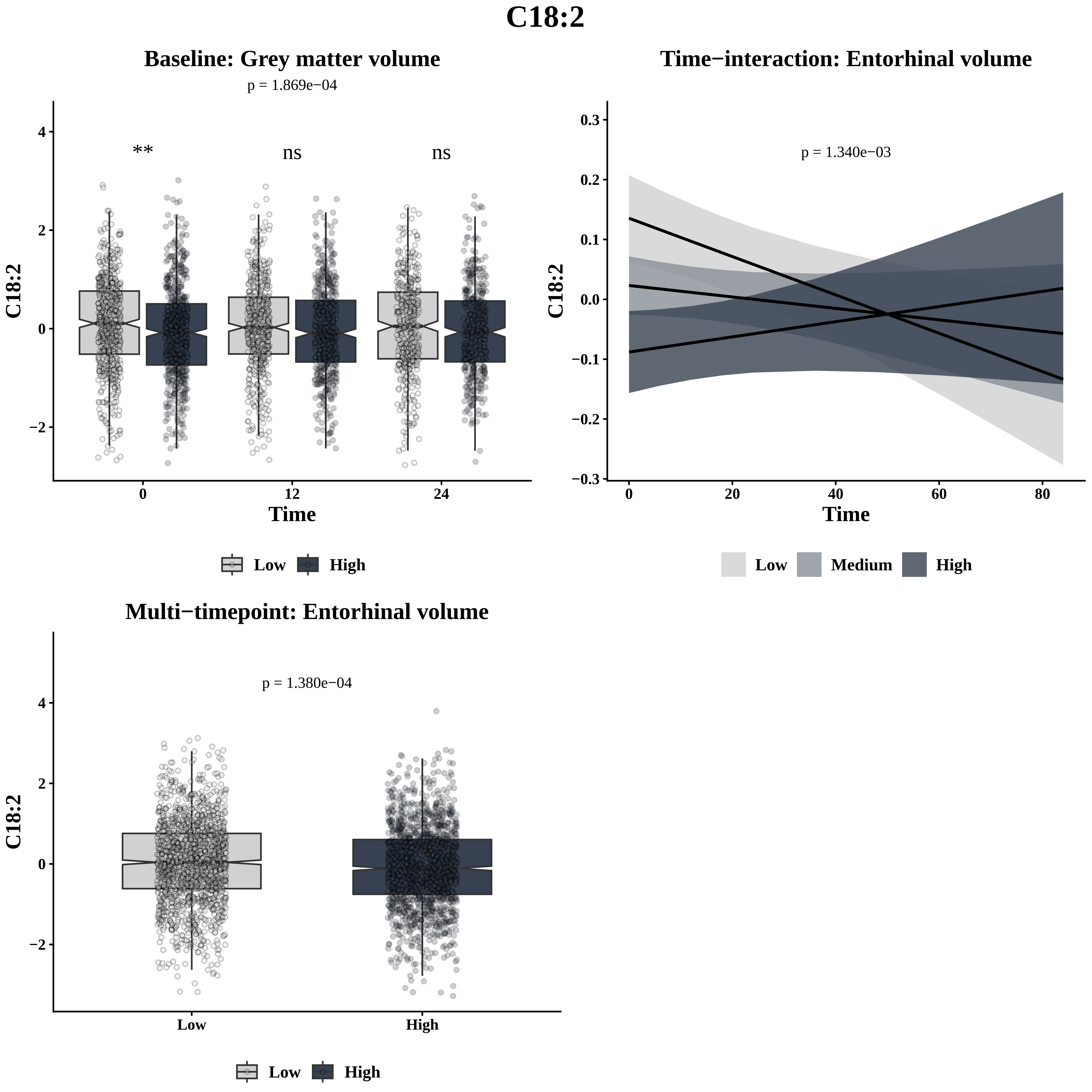
C18:2
Baseline: Grey matter volume
p = 1.869e−04
**
ns
ns
C18:2
Time
0
12
24
Low
High
Time−interaction: Entorhinal volume
p = 1.340e−03
C18:2
Time
Low
Medium
High
Multi−timepoint: Entorhinal volume
p = 1.380e−04
C18:2
Low
High

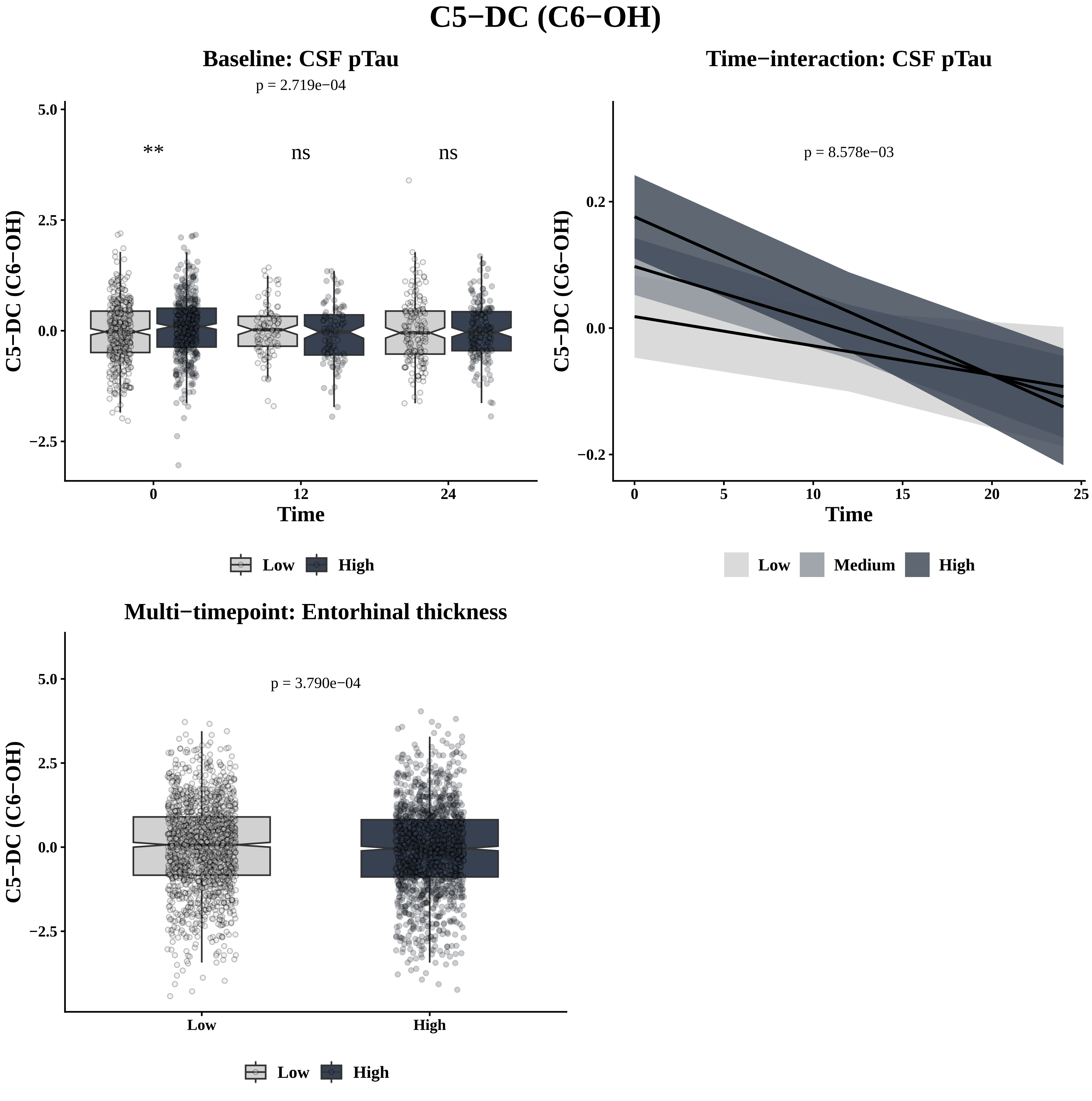
C5−DC (C6−OH)
Baseline: CSF pTau
p = 2.719e−04
**
ns
ns
C5−DC (C6−OH)
5.0
2.5
0.0
−2.5
0
12
24
Time
Low
High
Time−interaction: CSF pTau
p = 8.578e−03
C5−DC (C6−OH)
0.2
0.0
−0.2
0
5
10
15
20
25
Time
Low
Medium
High
Multi−timepoint: Entorhinal thickness
p = 3.790e−04
C5−DC (C6−OH)
5.0
2.5
0.0
−2.5
Low
High
Low
High

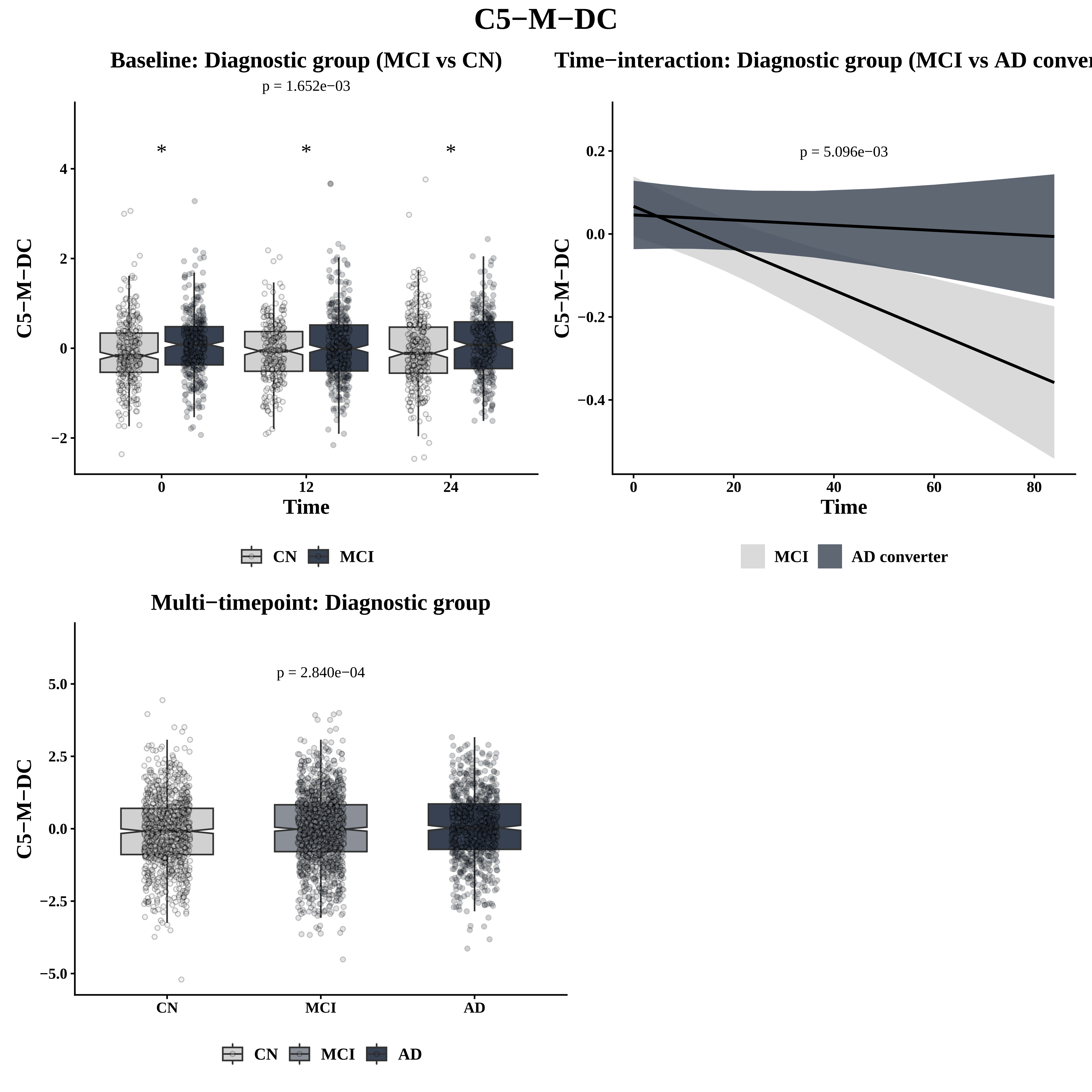
C5−M−DC
Baseline: Diagnostic group (MCI vs CN)
p = 1.652e−03
*
*
*
C5−M−DC
Time
0
12
24
4
2
0
−2
CN
MCI
Time−interaction: Diagnostic group (MCI vs AD conver
p = 5.096e−03
0.2
0.0
−0.2
−0.4
0
20
40
60
80
Time
MCI
AD converter
Multi−timepoint: Diagnostic group
p = 2.840e−04
5.0
2.5
0.0
−2.5
−5.0
CN
MCI
AD
CN
MCI
AD

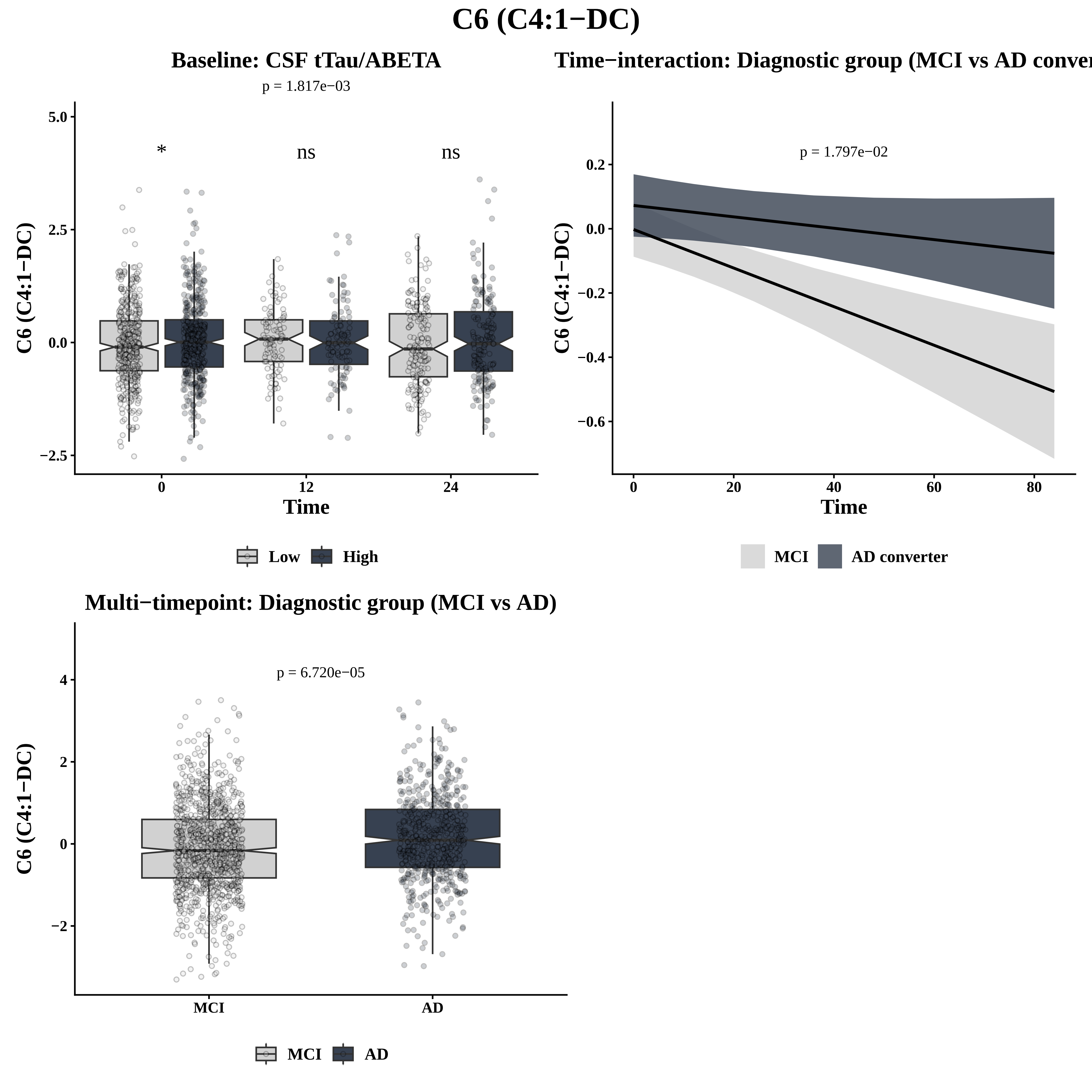
C6 (C4:1−DC)
Baseline: CSF tTau/ABETA
p = 1.817e−03
*
ns
ns
C6 (C4:1−DC)
Time
Low
High
Time−interaction: Diagnostic group (MCI vs AD conver
p = 1.797e−02
C6 (C4:1−DC)
Time
MCI
AD converter
Multi−timepoint: Diagnostic group (MCI vs AD)
p = 6.720e−05
C6 (C4:1−DC)
MCI
AD

# C8

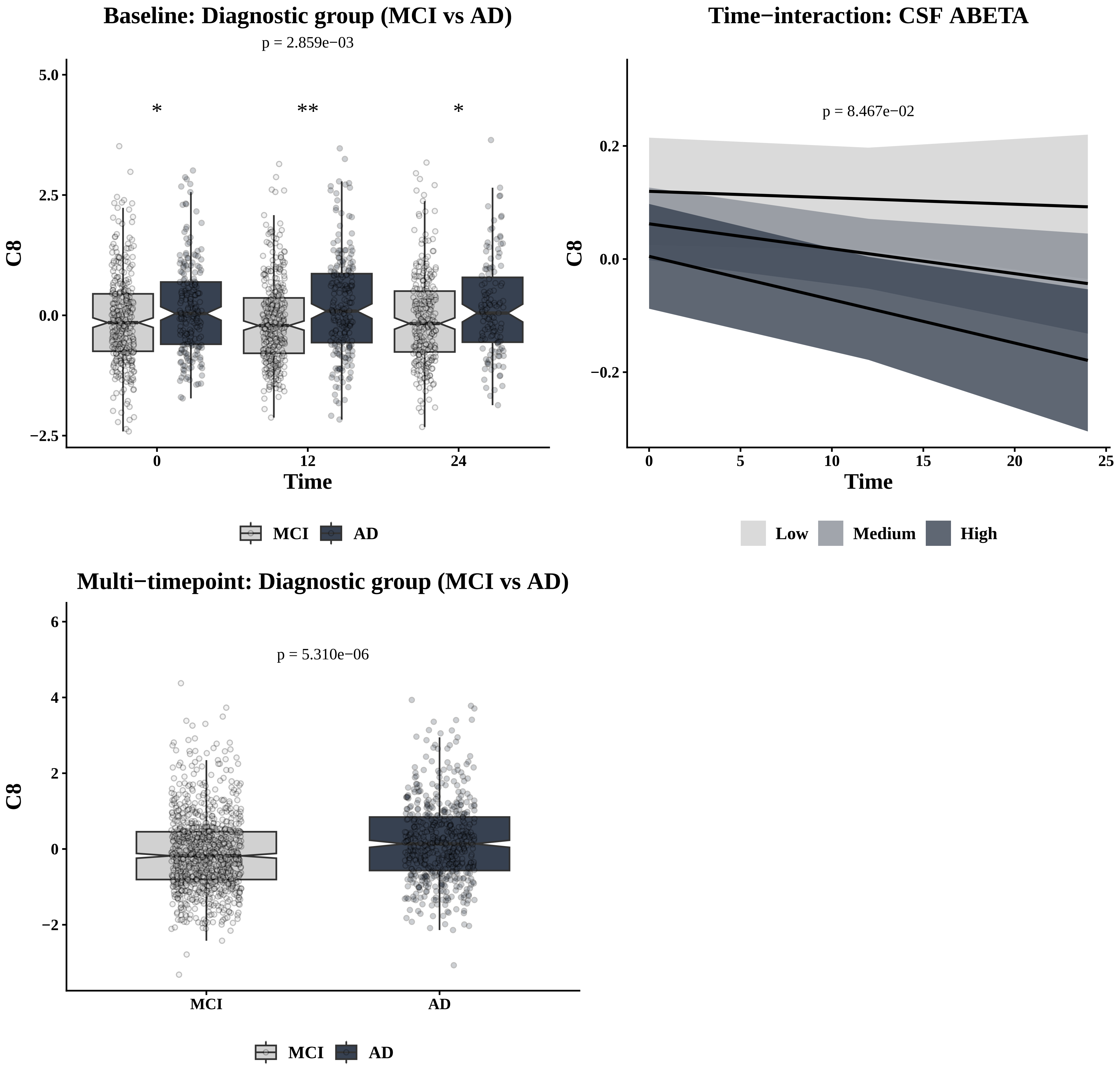
Baseline: Diagnostic group (MCI vs AD)
p = 2.859e−03
*
**
*
C8
5.0
2.5
0.0
−2.5
0
12
24
Time
MCI
AD
Time−interaction: CSF ABETA
p = 8.467e−02
C8
0.2
0.0
−0.2
0
5
10
15
20
25
Time
Low
Medium
High
Multi−timepoint: Diagnostic group (MCI vs AD)
p = 5.310e−06
C8
6
4
2
0
−2
MCI
AD
MCI
AD

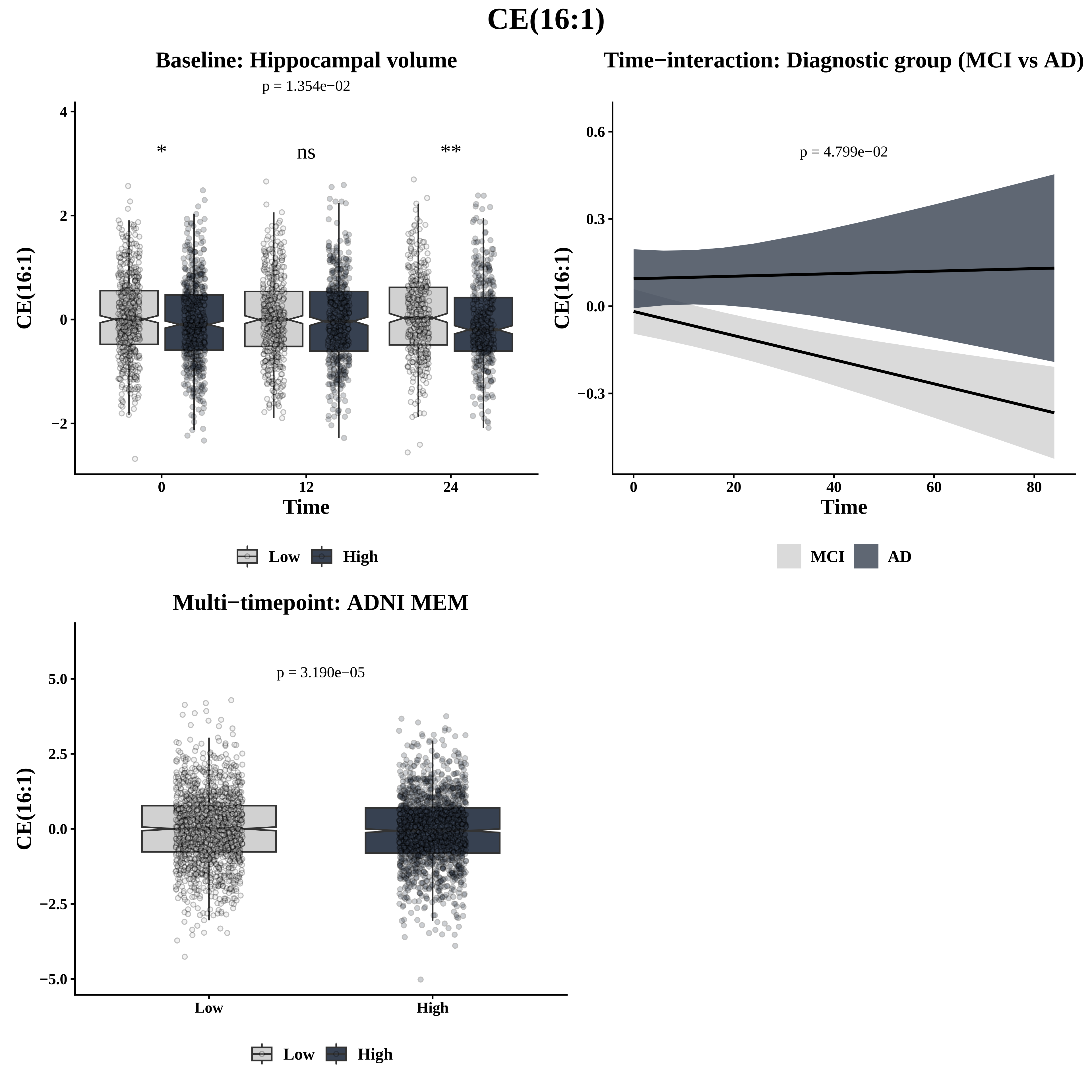
CE(16:1)
Baseline: Hippocampal volume
p = 1.354e−02
*
ns
**
CE(16:1)
Time
0
12
24
4
2
0
−2
Low
High
Time−interaction: Diagnostic group (MCI vs AD)
p = 4.799e−02
CE(16:1)
0.6
0.3
0.0
−0.3
0
20
40
60
80
Time
MCI
AD
Multi−timepoint: ADNI MEM
p = 3.190e−05
CE(16:1)
5.0
2.5
0.0
−2.5
−5.0
Low
High
Low
High

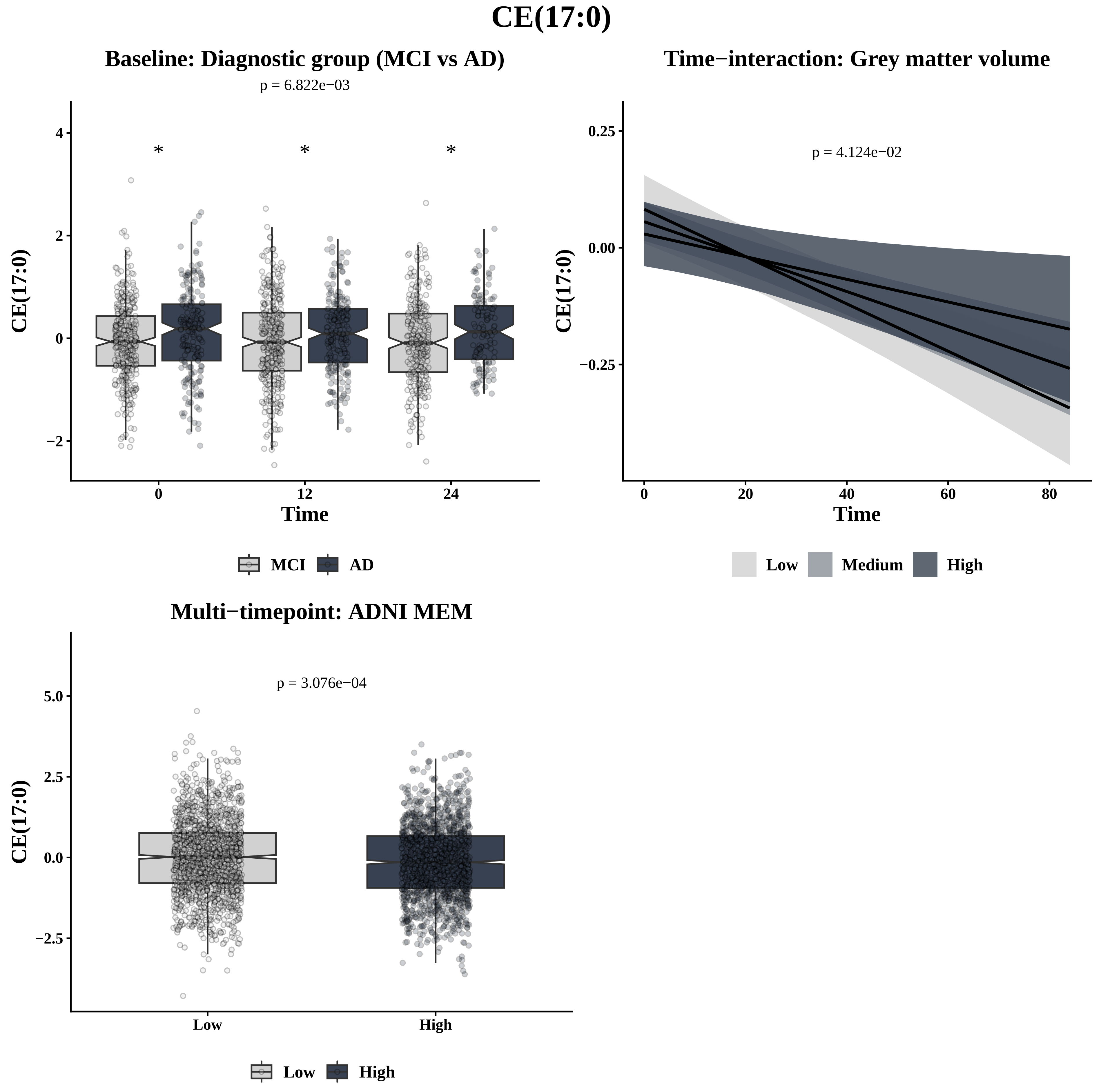
CE(17:0)
Baseline: Diagnostic group (MCI vs AD)
p = 6.822e−03
CE(17:0)
4
2
0
−2
*
*
*
0
12
24
Time
MCI
AD
Time−interaction: Grey matter volume
p = 4.124e−02
CE(17:0)
0.25
0.00
−0.25
0
20
40
60
80
Time
Low
Medium
High
Multi−timepoint: ADNI MEM
p = 3.076e−04
CE(17:0)
5.0
2.5
0.0
−2.5
Low
High
Low
High

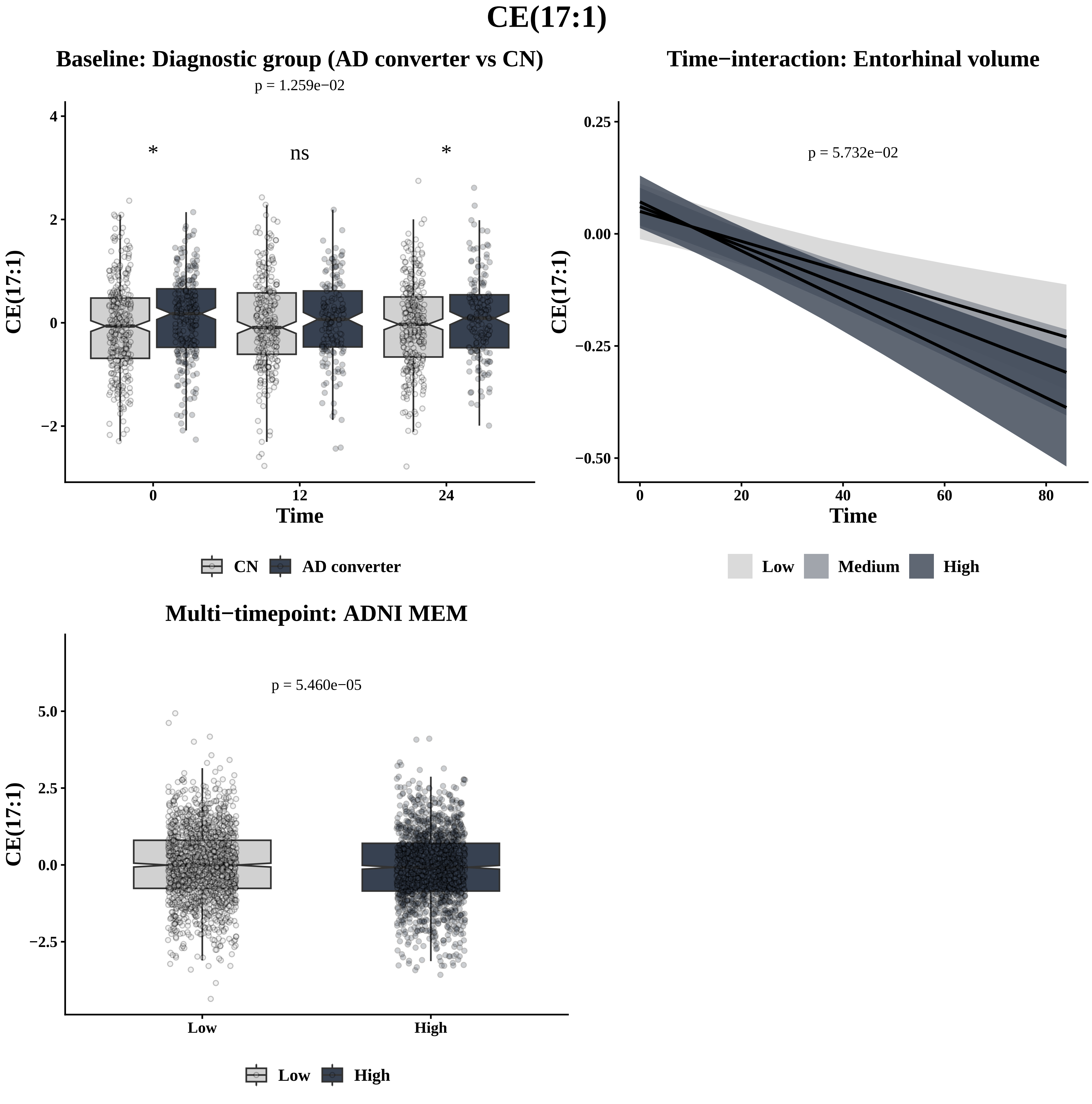
CE(17:1)
Baseline: Diagnostic group (AD converter vs CN)
p = 1.259e−02
*
ns
*
CE(17:1)
Time
0
12
24
4
2
0
−2
CN
AD converter
Time−interaction: Entorhinal volume
p = 5.732e−02
0.25
0.00
−0.25
−0.50
0
20
40
60
80
Time
Low
Medium
High
Multi−timepoint: ADNI MEM
p = 5.460e−05
5.0
2.5
0.0
−2.5
Low
High
Low
High

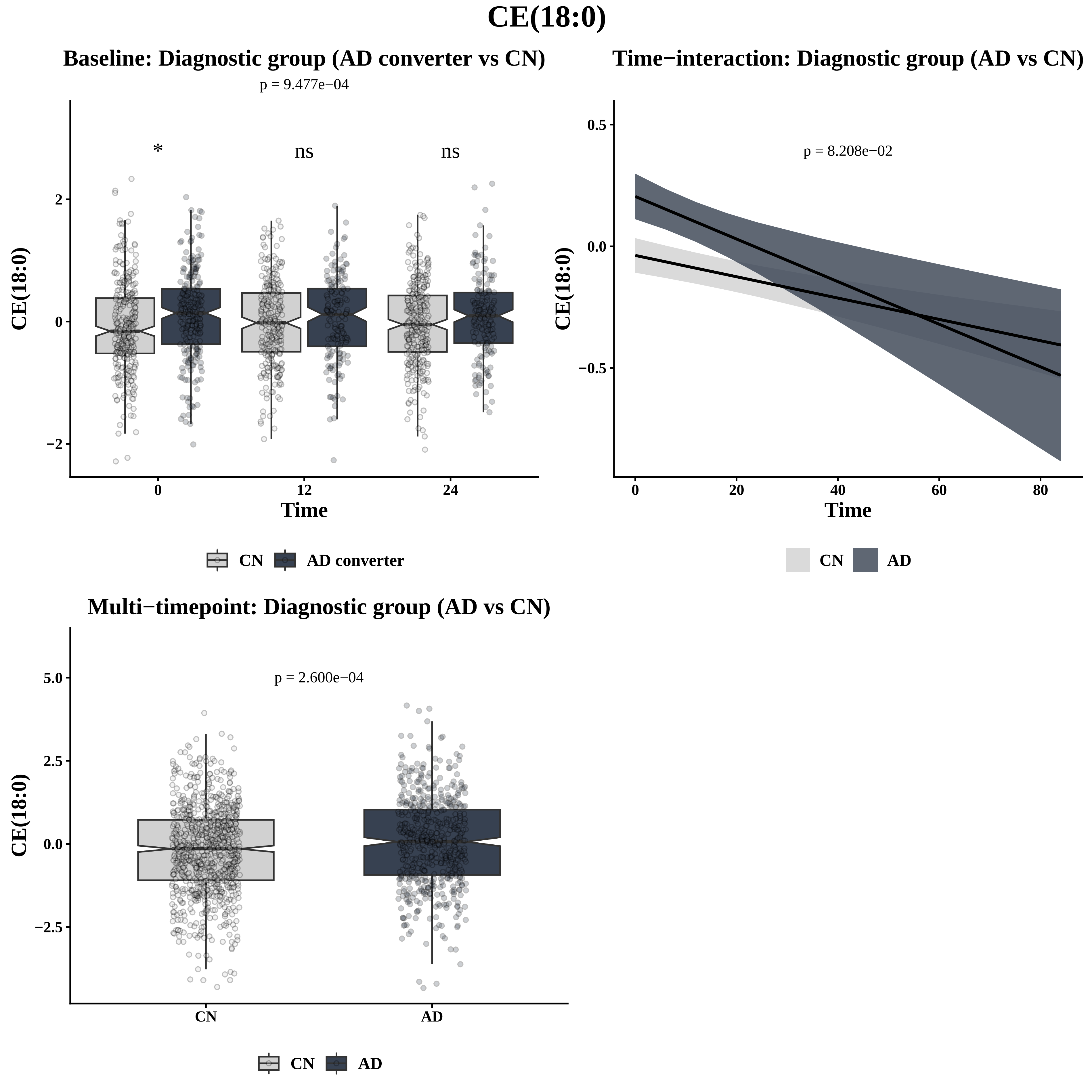
CE(18:0)
Baseline: Diagnostic group (AD converter vs CN)
p = 9.477e−04
*
ns
ns
CE(18:0)
2
0
−2
0
12
24
Time
CN
AD converter
Time−interaction: Diagnostic group (AD vs CN)
p = 8.208e−02
CE(18:0)
0.5
0.0
−0.5
0
20
40
60
80
Time
CN
AD
Multi−timepoint: Diagnostic group (AD vs CN)
p = 2.600e−04
CE(18:0)
5.0
2.5
0.0
−2.5
CN
AD
CN
AD

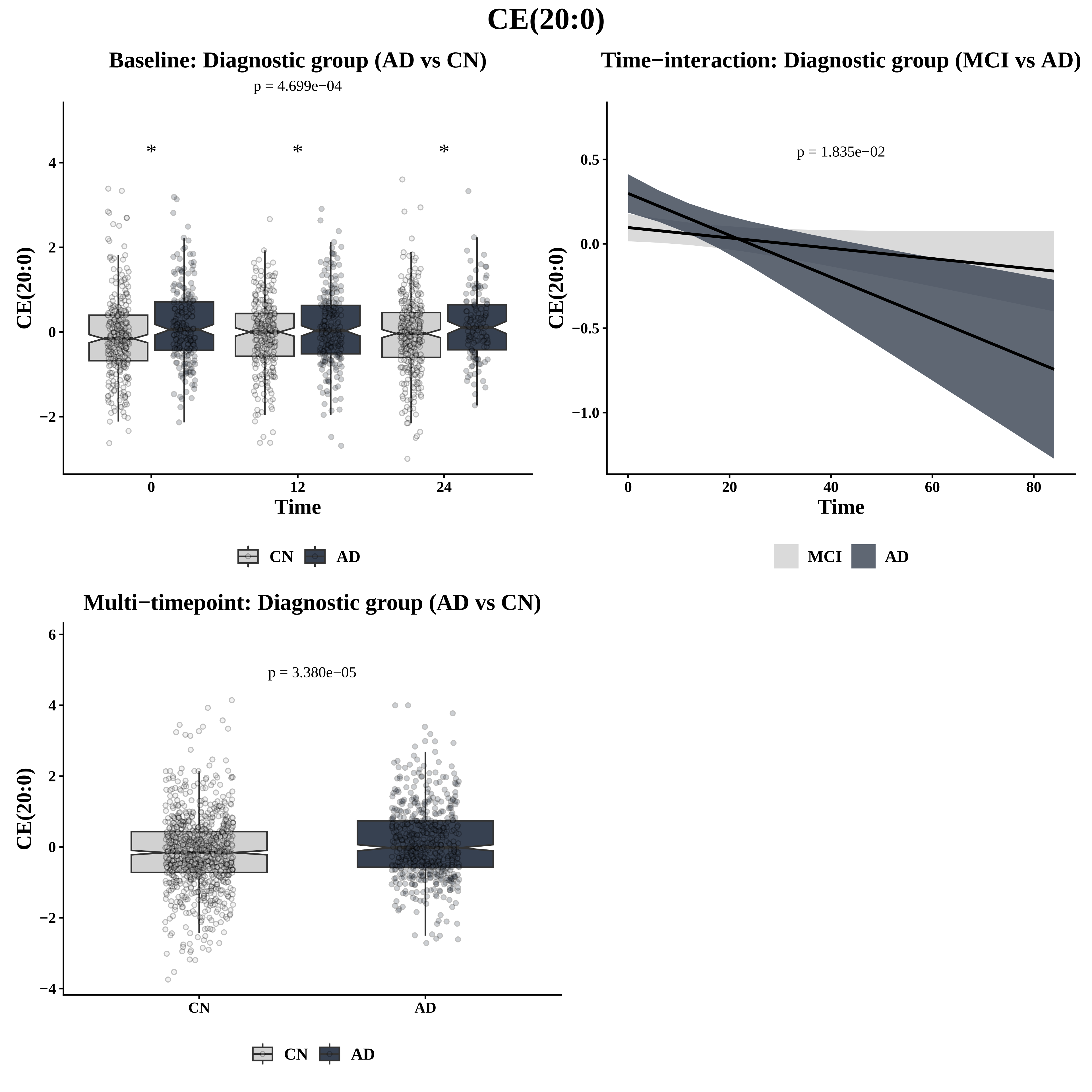
CE(20:0)
Baseline: Diagnostic group (AD vs CN)
p = 4.699e−04
*
*
*
CE(20:0)
Time
0
12
24
CN
AD
Time−interaction: Diagnostic group (MCI vs AD)
p = 1.835e−02
CE(20:0)
Time
MCI
AD
Multi−timepoint: Diagnostic group (AD vs CN)
p = 3.380e−05
CE(20:0)
CN
AD
CN
AD

# CE(20:1)

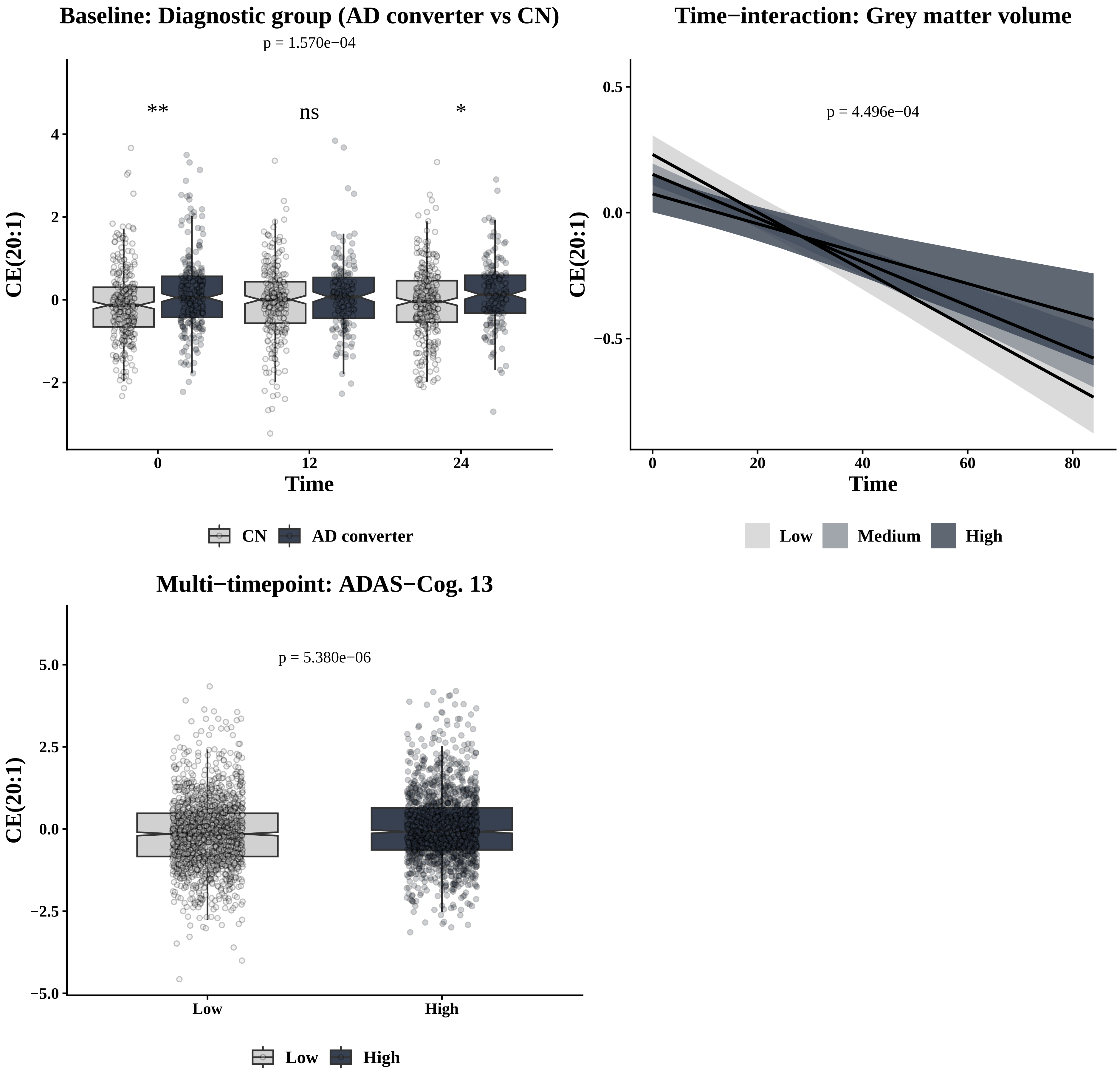
Baseline: Diagnostic group (AD converter vs CN)
p = 1.570e−04
**
ns
*
CE(20:1)
4
2
0
−2
0
12
24
Time
CN
AD converter
Time−interaction: Grey matter volume
p = 4.496e−04
CE(20:1)
0.5
0.0
−0.5
0
20
40
60
80
Time
Low
Medium
High
Multi−timepoint: ADAS−Cog. 13
p = 5.380e−06
CE(20:1)
5.0
2.5
0.0
−2.5
−5.0
Low
High
Low
High

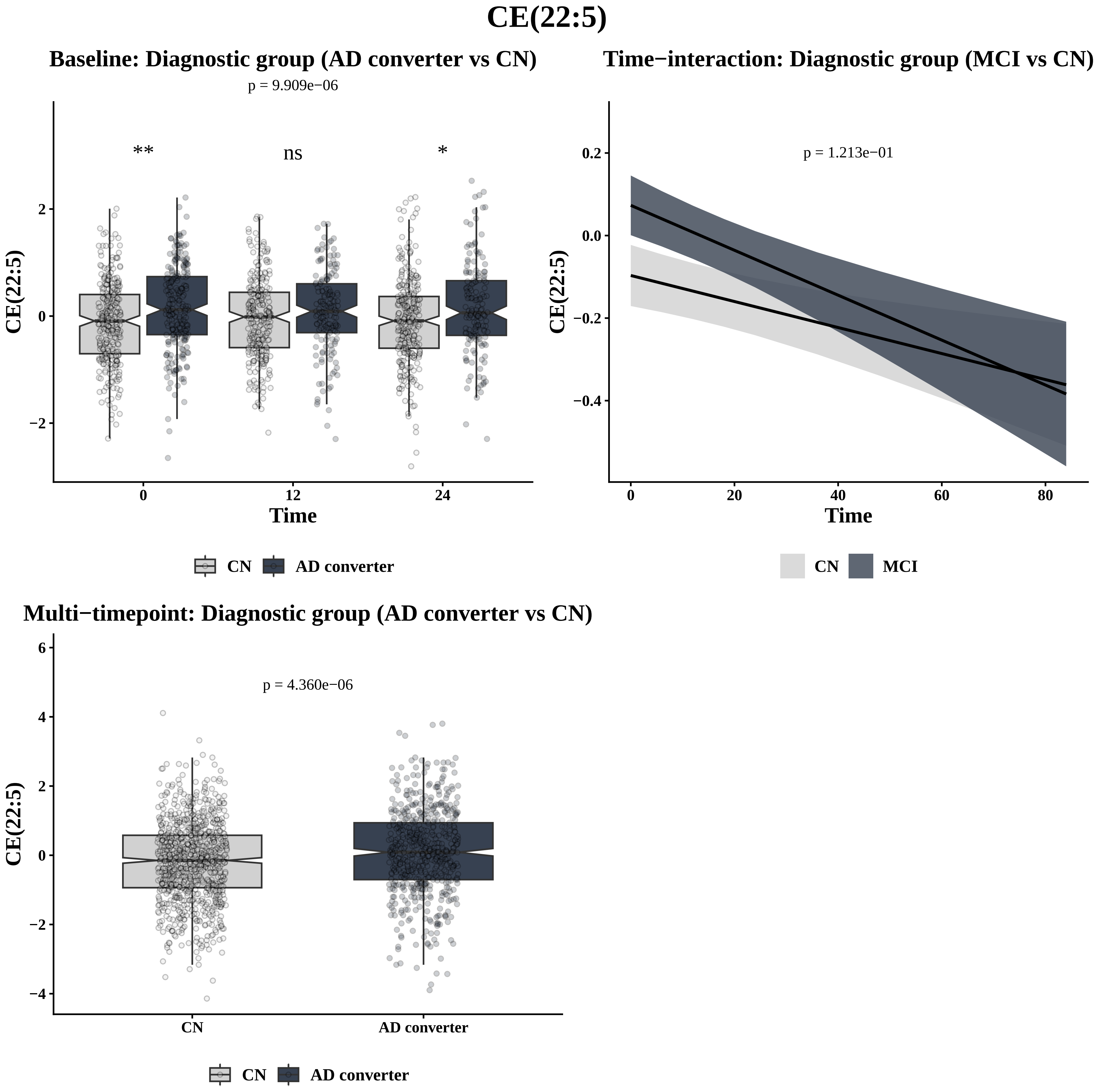

CE(22:5)
Baseline: Diagnostic group (AD converter vs CN)
p = 9.909e−06
**
ns
*
CE(22:5)
2
0
−2
0
12
24
Time
CN
AD converter
Time−interaction: Diagnostic group (MCI vs CN)
p = 1.213e−01
CE(22:5)
0.2
0.0
−0.2
−0.4
0
20
40
60
80
Time
CN
MCI
Multi−timepoint: Diagnostic group (AD converter vs CN)
p = 4.360e−06
CE(22:5)
6
4
2
0
−2
−4
CN
AD converter
CN
AD converter

### Cer(d18:0/24:0)

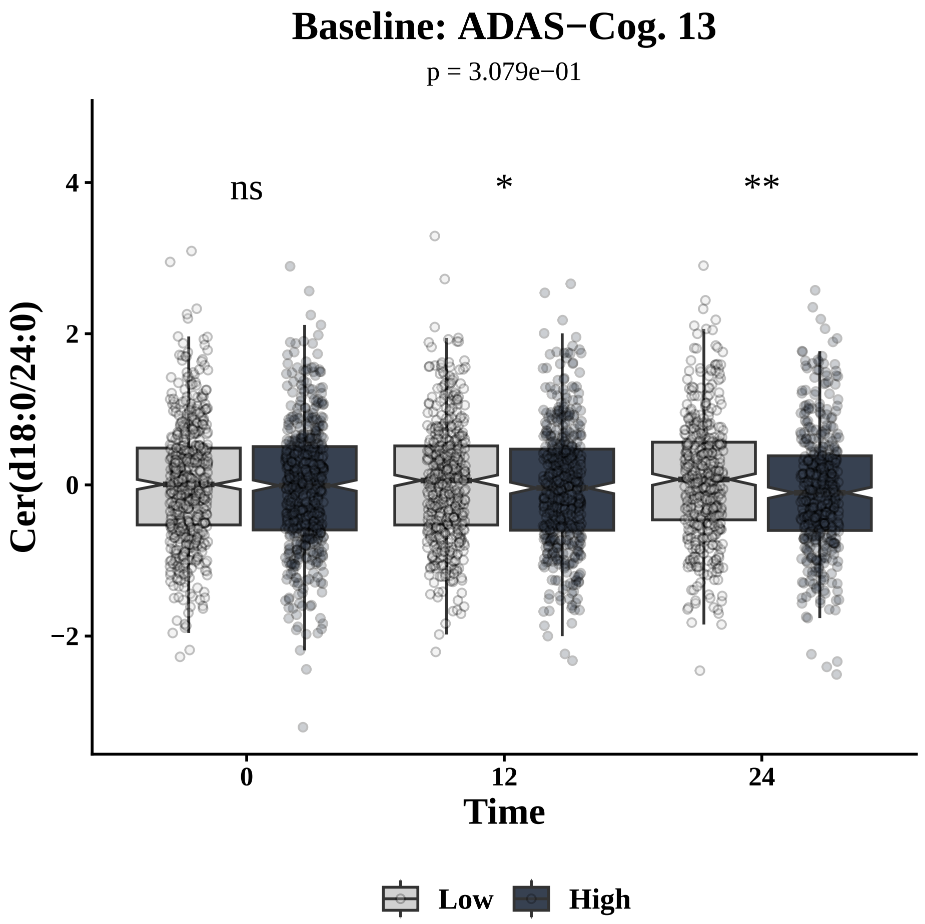
Baseline: ADAS−Cog. 13
p = 3.079e−01
ns
*
**
Cer(d18:0/24:0)
4
2
0
−2
0
12
24
Time
Low
High

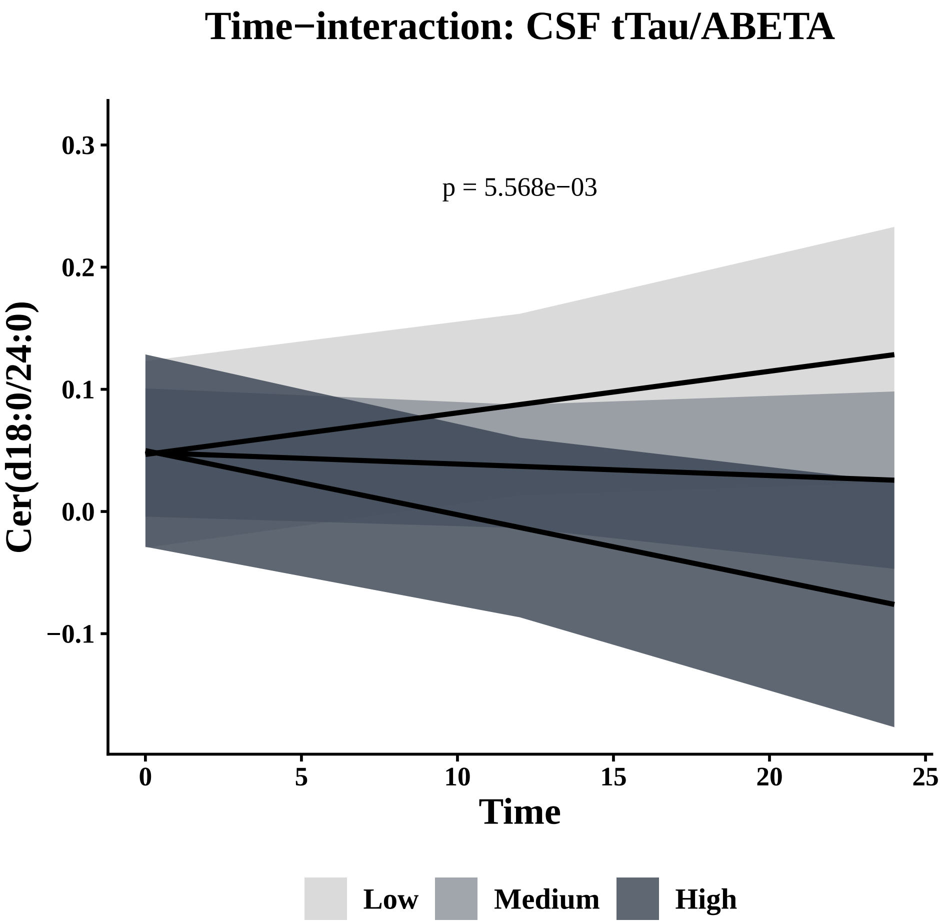
Time−interaction: CSF tTau/ABETA
p = 5.568e−03
Cer(d18:0/24:0)
0.3
0.2
0.1
0.0
−0.1
0
5
10
15
20
25
Time
Low
Medium
High

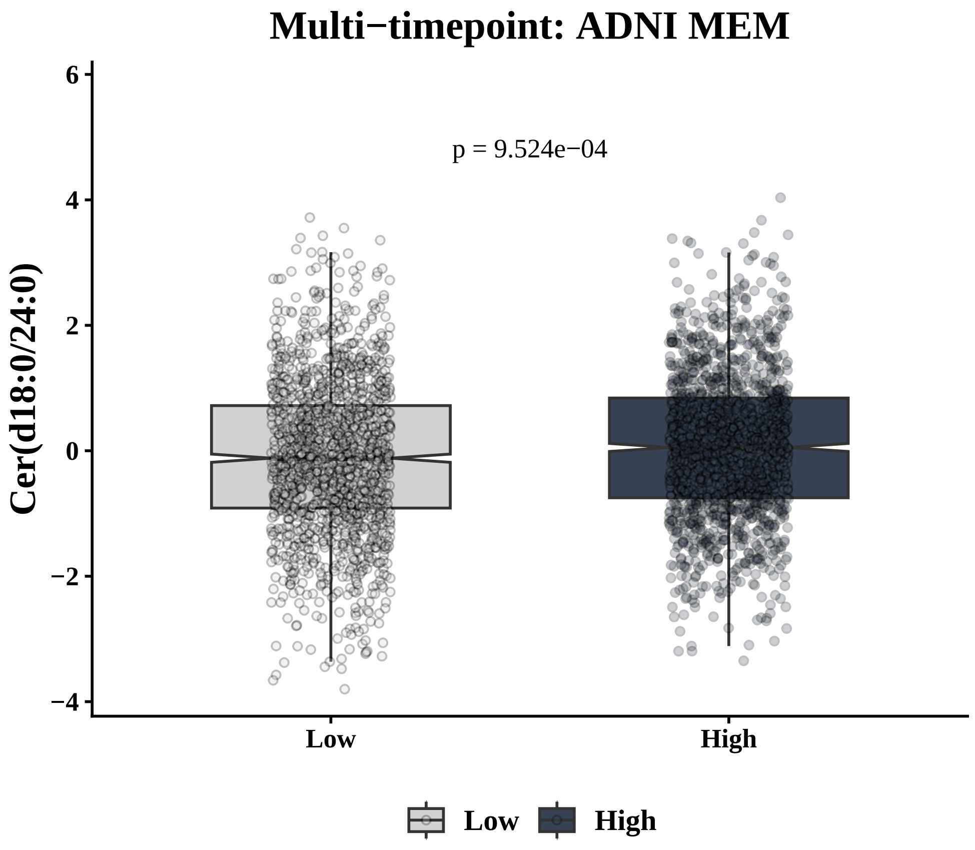
Multi−timepoint: ADNI MEM
p = 9.524e−04
Cer(d18:0/24:0)
6
4
2
0
−2
−4
Low
High
Low
High

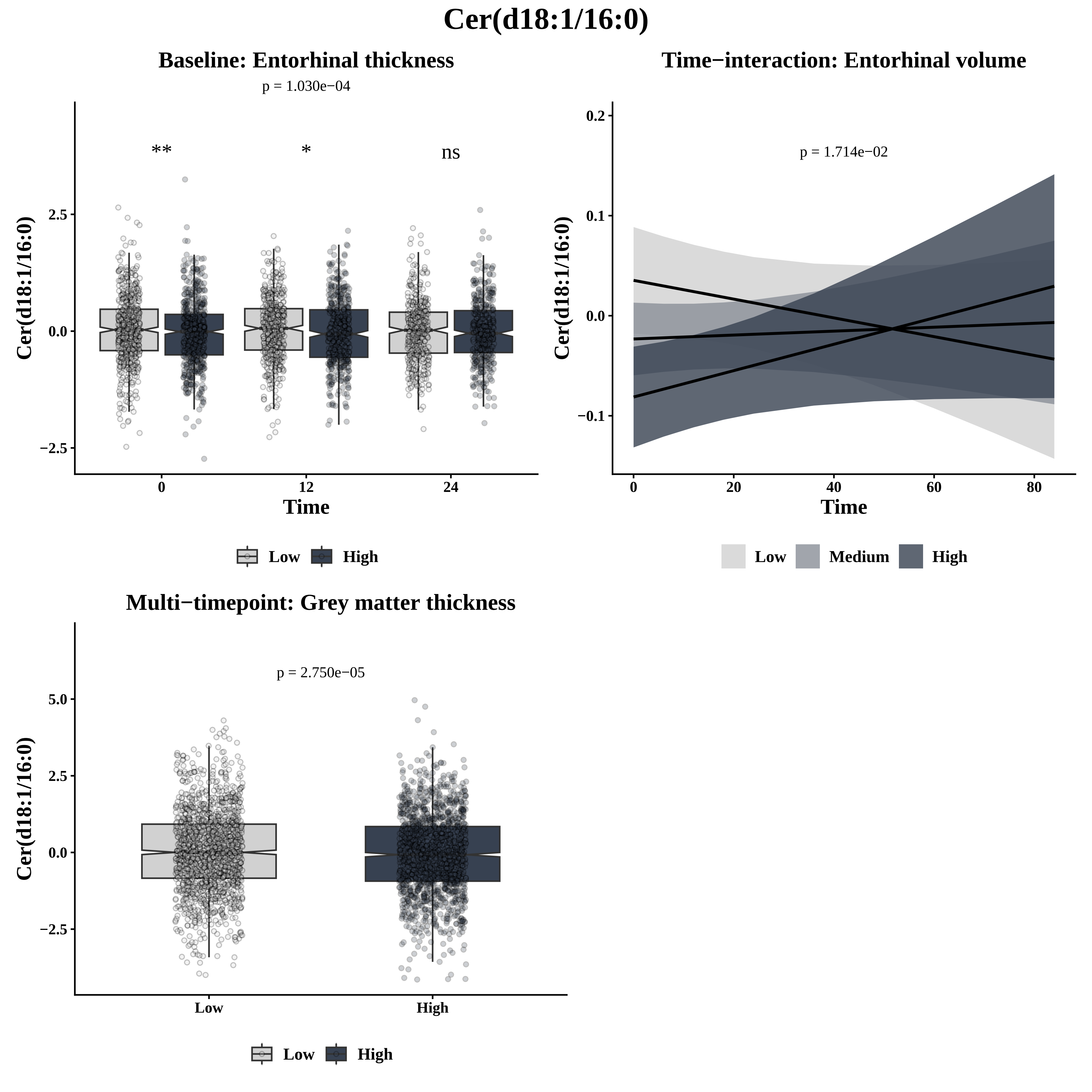

Cer(d18:1/16:0)
Baseline: Entorhinal thickness
p = 1.030e−04
**
*
ns
Cer(d18:1/16:0)
2.5
0.0
−2.5
0
12
24
Time
Low
High
Time−interaction: Entorhinal volume
p = 1.714e−02
Cer(d18:1/16:0)
0.2
0.1
0.0
−0.1
0
20
40
60
80
Time
Low
Medium
High
Multi−timepoint: Grey matter thickness
p = 2.750e−05
Cer(d18:1/16:0)
5.0
2.5
0.0
−2.5
Low
High
Low
High

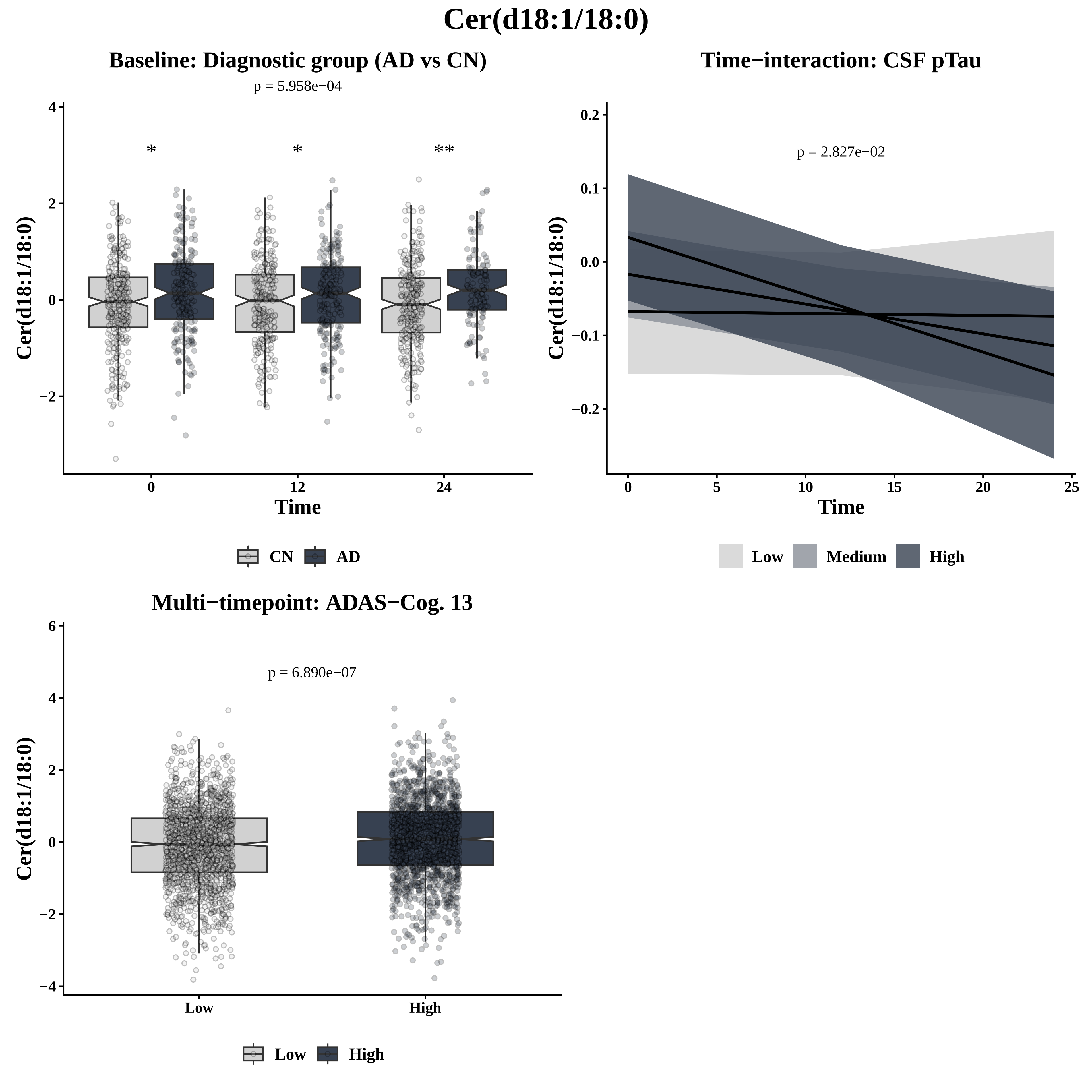
Cer(d18:1/18:0)
Baseline: Diagnostic group (AD vs CN)
p = 5.958e−04
*
*
**
Cer(d18:1/18:0)
Time
CN
AD
Time−interaction: CSF pTau
p = 2.827e−02
Cer(d18:1/18:0)
Time
Low
Medium
High
Multi−timepoint: ADAS−Cog. 13
p = 6.890e−07
Cer(d18:1/18:0)
Low
High
Low
High

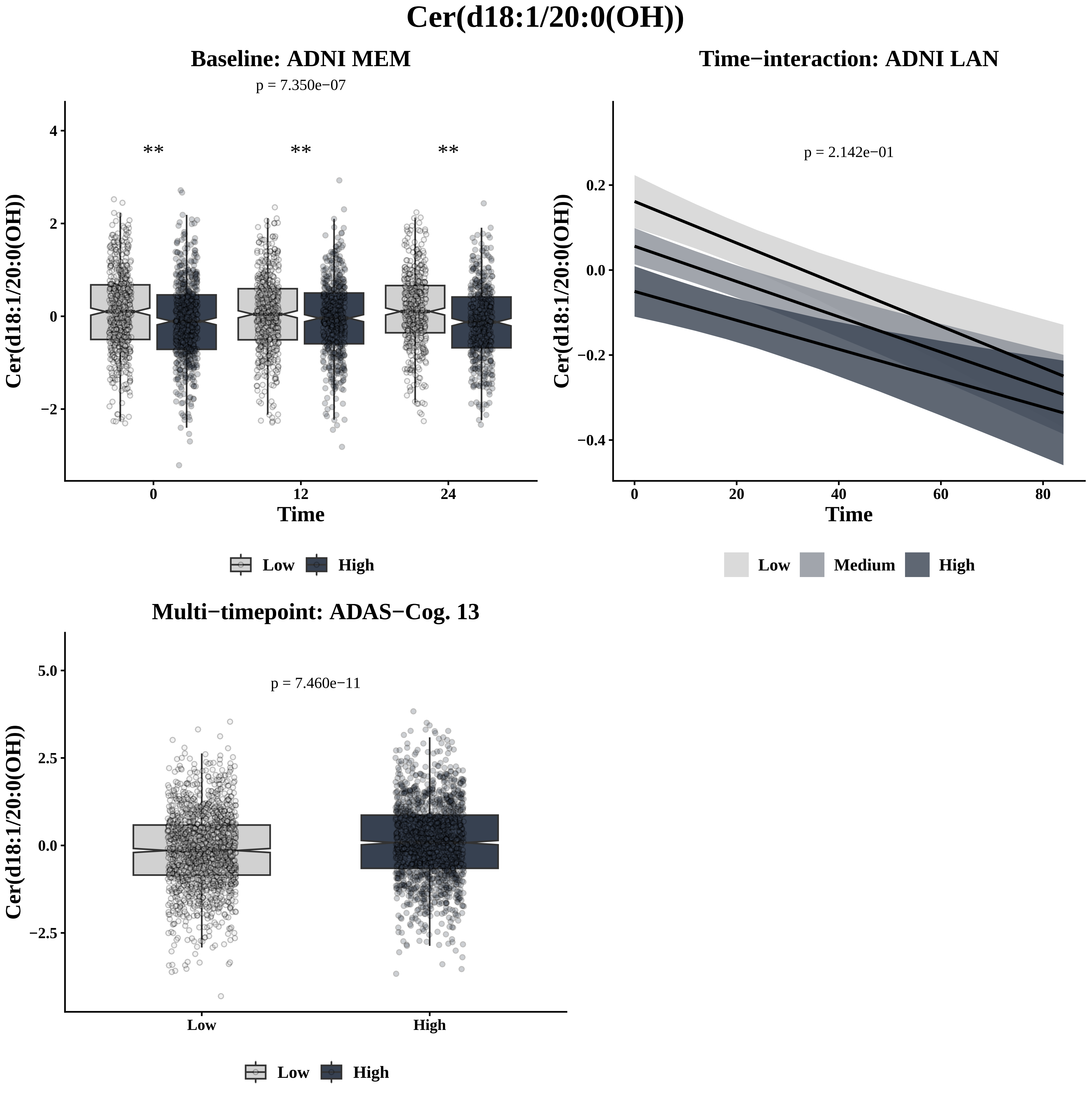
Cer(d18:1/20:0(OH))
Baseline: ADNI MEM
p = 7.350e−07
**
**
**
Cer(d18:1/20:0(OH))
Time
Low
High
Time−interaction: ADNI LAN
p = 2.142e−01
Cer(d18:1/20:0(OH))
Time
Low
Medium
High
Multi−timepoint: ADAS−Cog. 13
p = 7.460e−11
Cer(d18:1/20:0(OH))
Low
High
Low
High

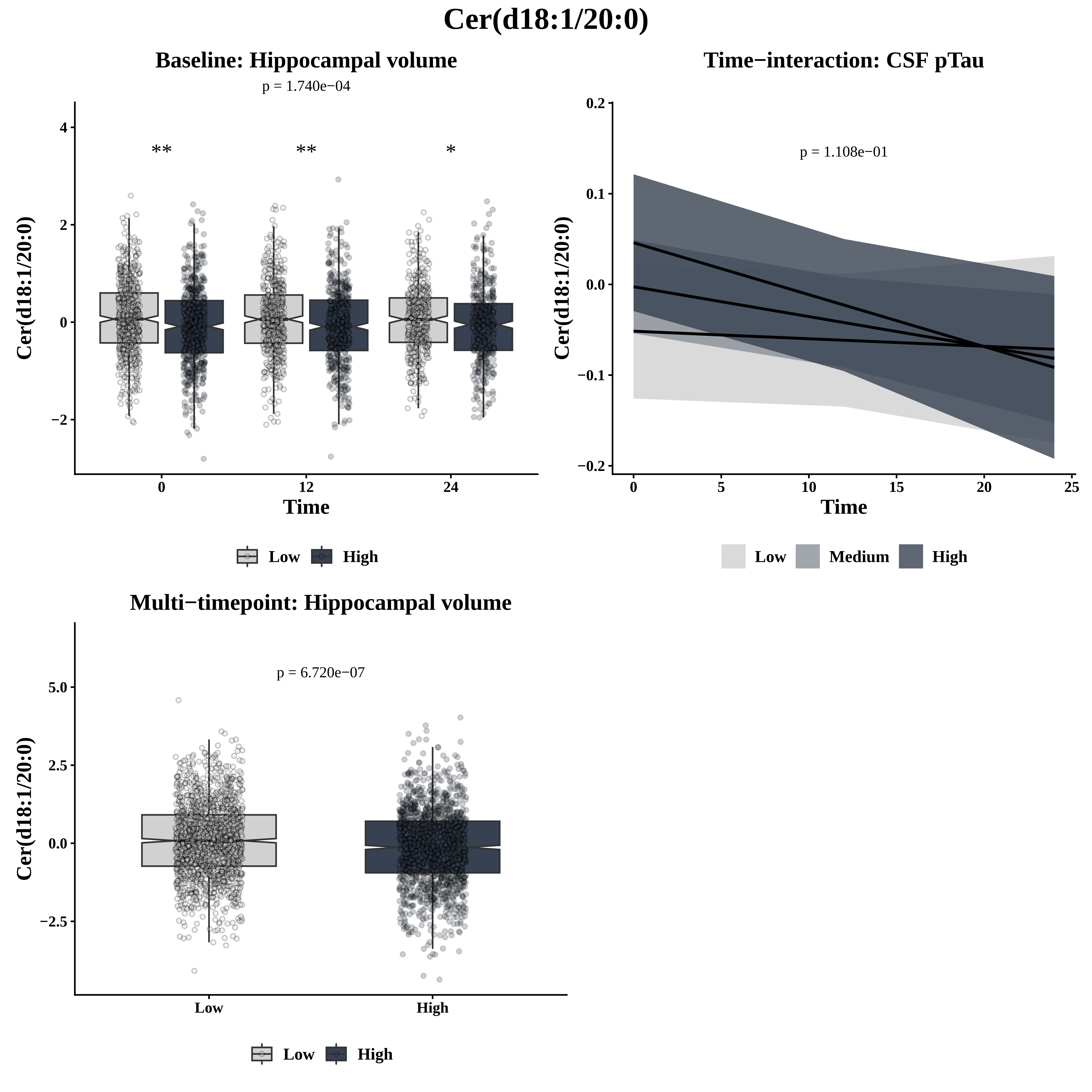

Cer(d18:1/20:0)
Baseline: Hippocampal volume
p = 1.740e−04
**
**
*
Cer(d18:1/20:0)
Time
0
12
24
4
2
0
−2
Low
High
Time−interaction: CSF pTau
p = 1.108e−01
0.2
0.1
0.0
−0.1
−0.2
0
5
10
15
20
25
Low
Medium
High
Multi−timepoint: Hippocampal volume
p = 6.720e−07
5.0
2.5
0.0
−2.5
Low
High

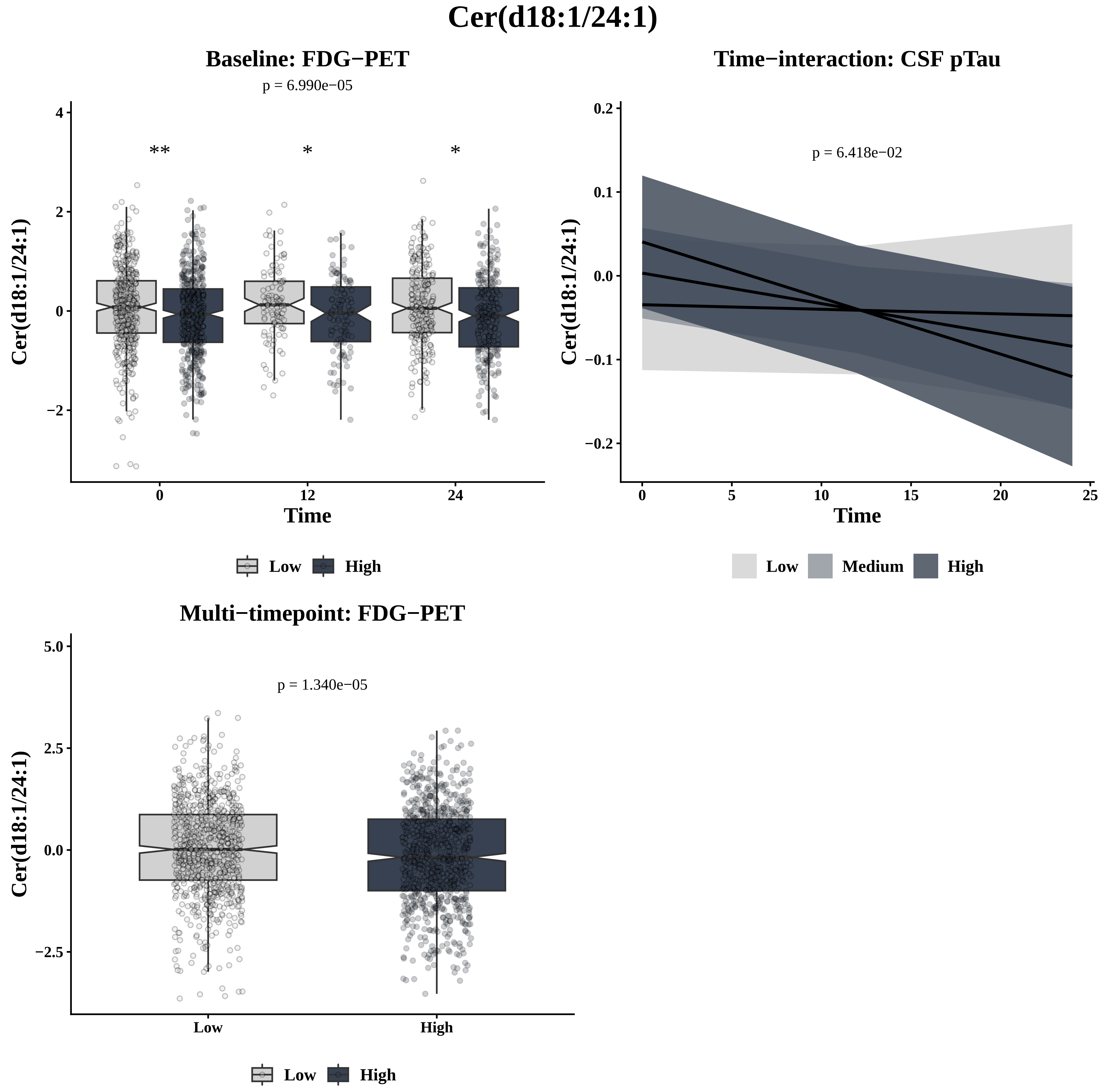
Cer(d18:1/24:1)
Baseline: FDG−PET
p = 6.990e−05
**
*
*
Cer(d18:1/24:1)
4
2
0
−2
0
12
24
Time
Low
High
Time−interaction: CSF pTau
p = 6.418e−02
Cer(d18:1/24:1)
0.2
0.1
0.0
−0.1
−0.2
0
5
10
15
20
25
Time
Low
Medium
High
Multi−timepoint: FDG−PET
p = 1.340e−05
Cer(d18:1/24:1)
5.0
2.5
0.0
−2.5
Low
High
Low
High

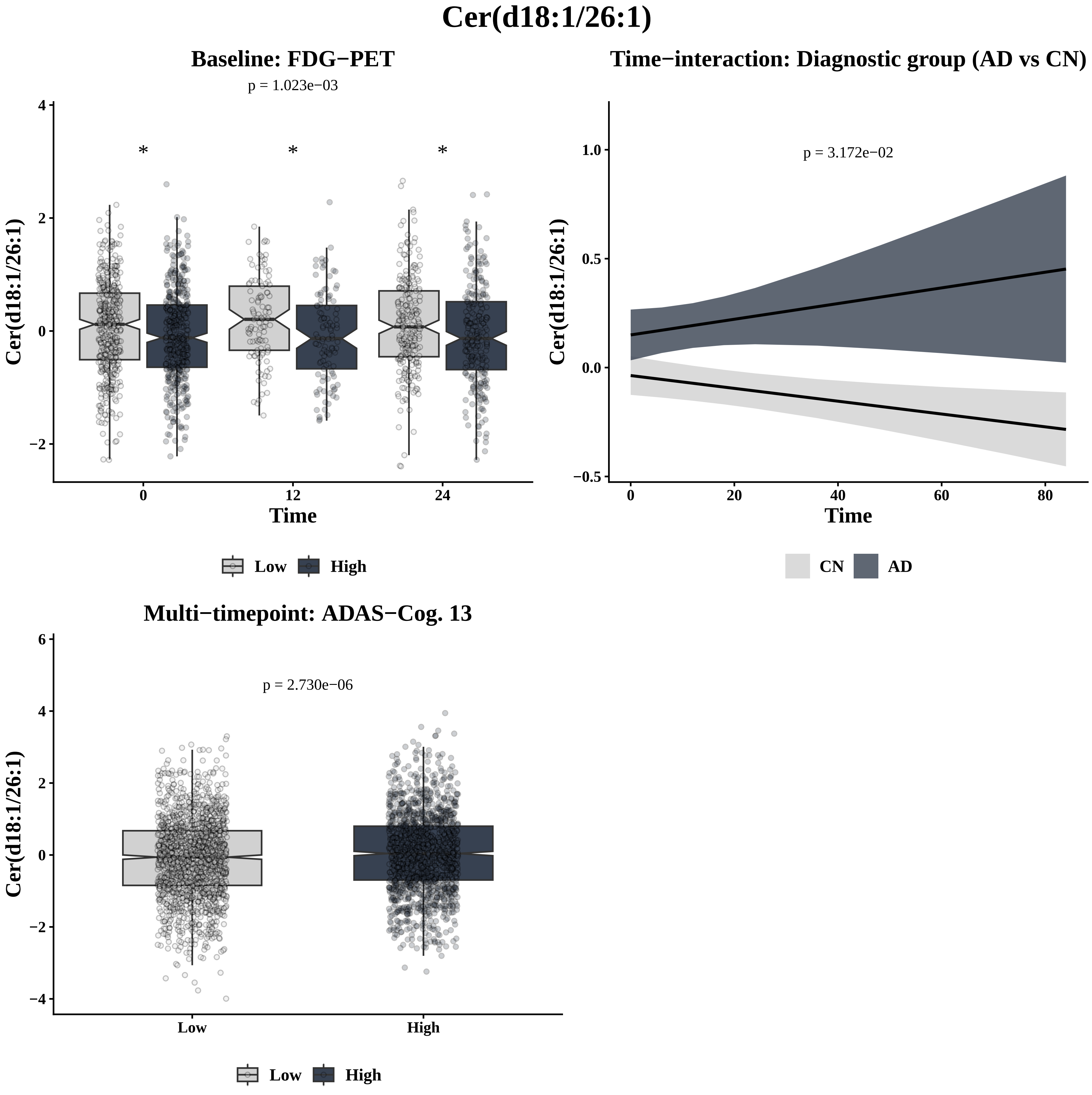

Cer(d18:1/26:1)
Baseline: FDG−PET
p = 1.023e−03
Cer(d18:1/26:1)
Time
0
12
24
*
Low
High
Time−interaction: Diagnostic group (AD vs CN)
p = 3.172e−02
Cer(d18:1/26:1)
Time
0
20
40
60
80
CN
AD
Multi−timepoint: ADAS−Cog. 13
p = 2.730e−06
Cer(d18:1/26:1)
Low
High

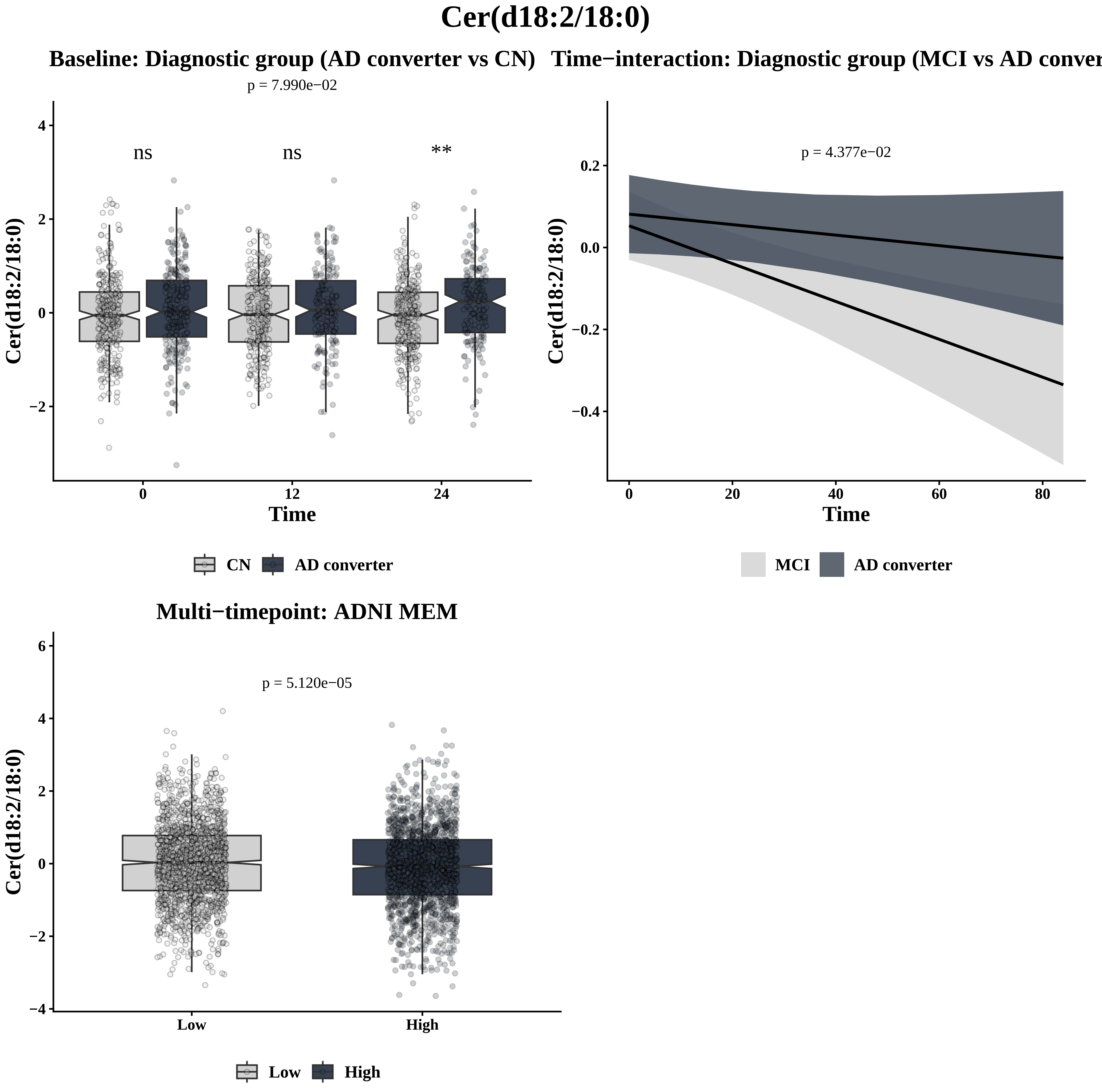

Cer(d18:2/18:0)
Baseline: Diagnostic group (AD converter vs CN)
p = 7.990e−02
ns
ns
**
Cer(d18:2/18:0)
Time
0
12
24
CN
AD converter
Time−interaction: Diagnostic group (MCI vs AD conver
p = 4.377e−02
Cer(d18:2/18:0)
Time
MCI
AD converter
Multi−timepoint: ADNI MEM
p = 5.120e−05
Cer(d18:2/18:0)
Low
High

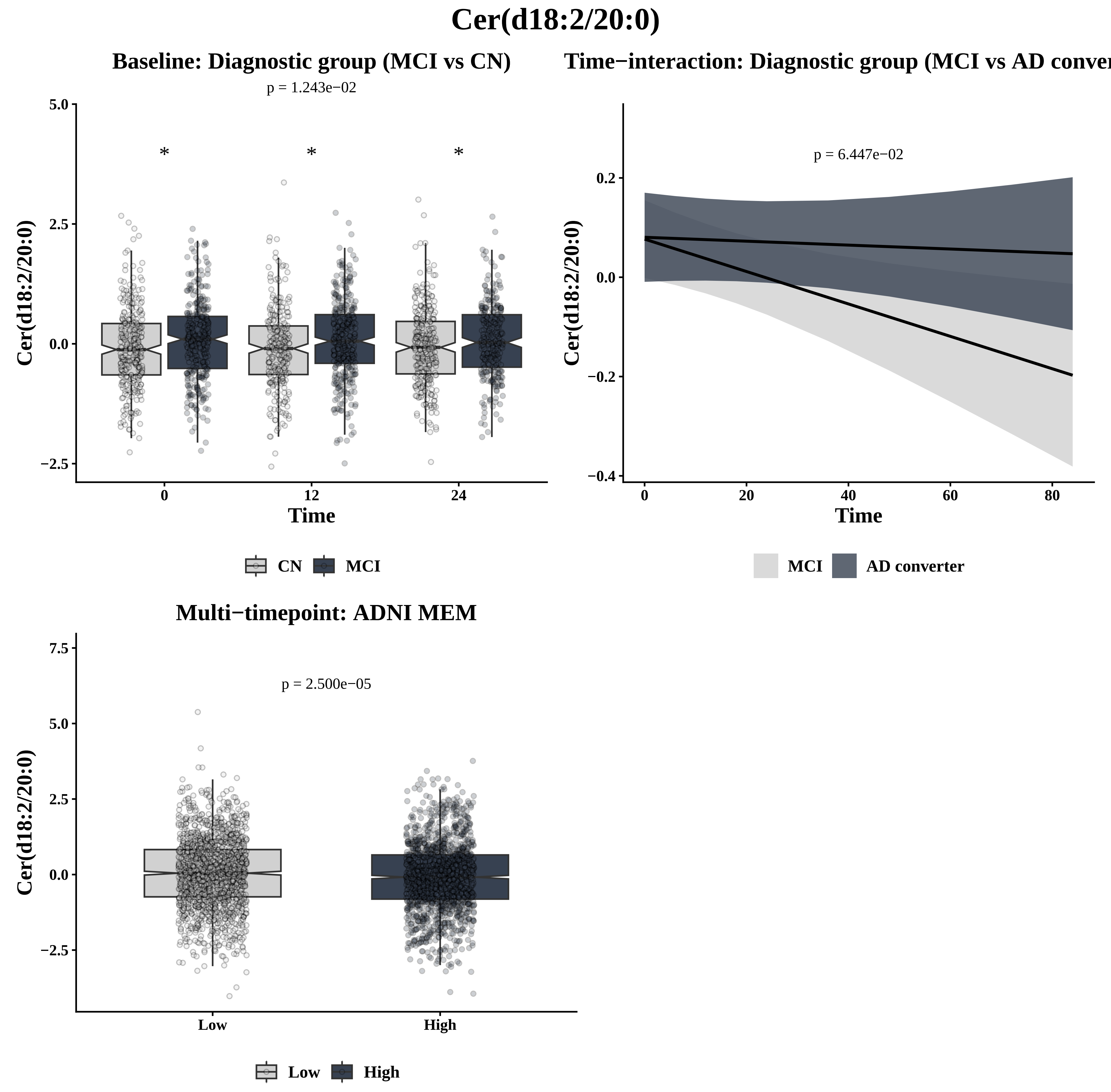

Cer(d18:2/20:0)
Baseline: Diagnostic group (MCI vs CN)
p = 1.243e−02
Time
CN
MCI
Time−interaction: Diagnostic group (MCI vs AD conver
p = 6.447e−02
Time
MCI
AD converter
Multi−timepoint: ADNI MEM
p = 2.500e−05
Low
High

### Cer(d18:2/24:1)

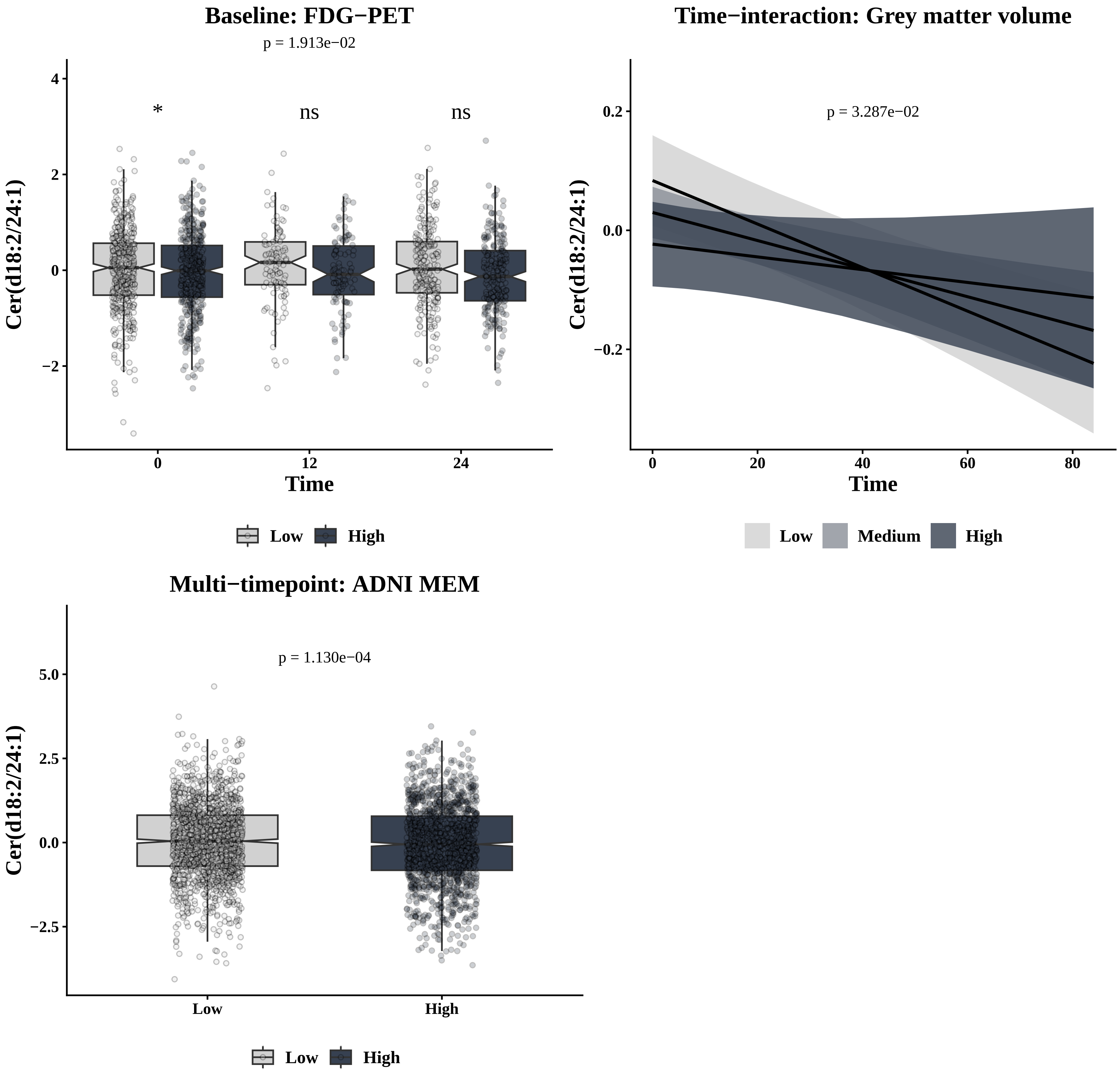

Baseline: FDG−PET
p = 1.913e−02
*
ns
ns
Cer(d18:2/24:1)
Time
0
12
24
Low
High
Time−interaction: Grey matter volume
p = 3.287e−02
Cer(d18:2/24:1)
Time
0
20
40
60
80
Low
Medium
High
Multi−timepoint: ADNI MEM
p = 1.130e−04
Cer(d18:2/24:1)
Low
High
Low
High

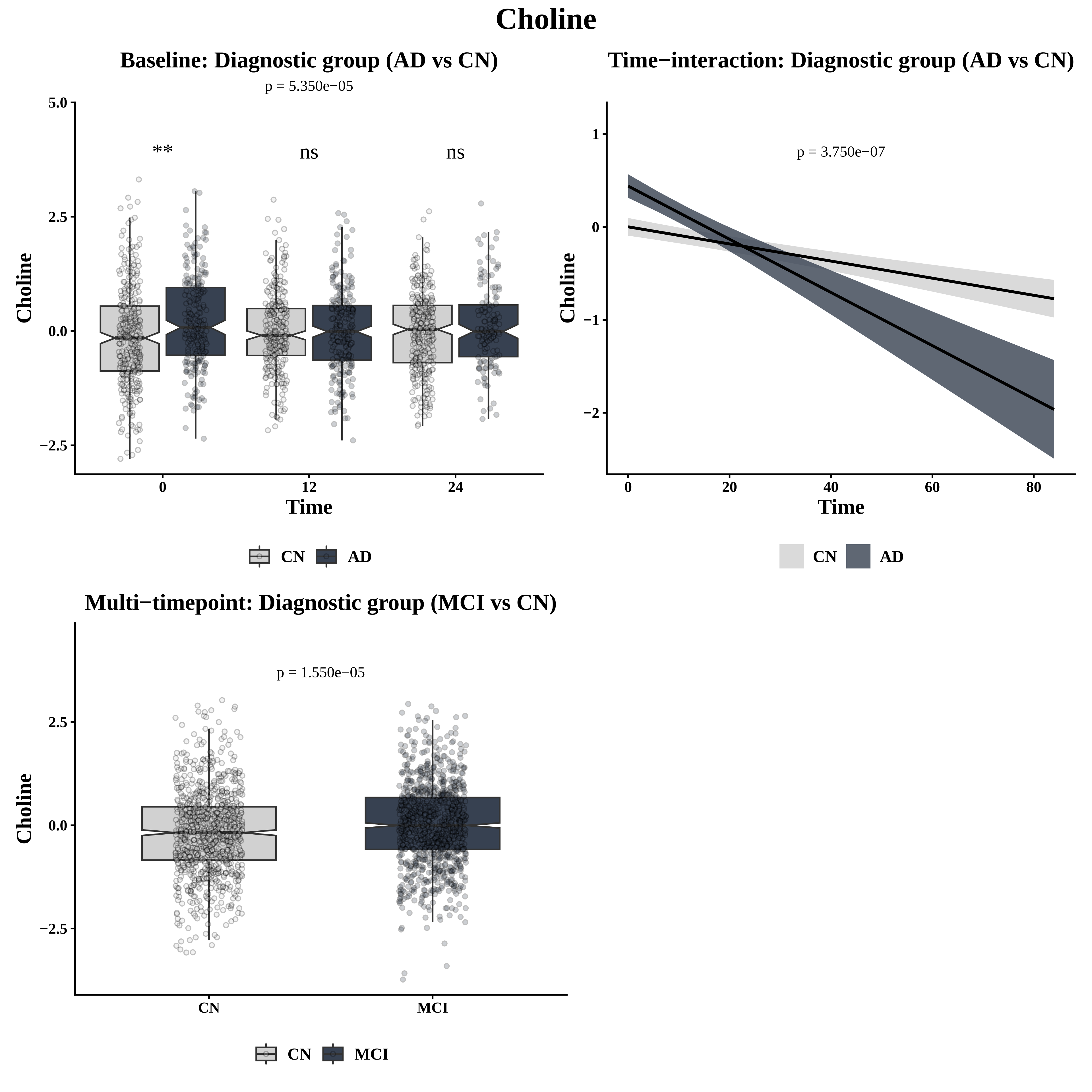
Choline
Baseline: Diagnostic group (AD vs CN)
p = 5.350e−05
**
ns
ns
5.0
2.5
0.0
−2.5
Choline
0
12
24
Time
CN
AD
Time−interaction: Diagnostic group (AD vs CN)
p = 3.750e−07
1
0
−1
−2
Choline
0
20
40
60
80
Time
CN
AD
Multi−timepoint: Diagnostic group (MCI vs CN)
p = 1.550e−05
2.5
0.0
−2.5
Choline
CN
MCI
CN
MCI

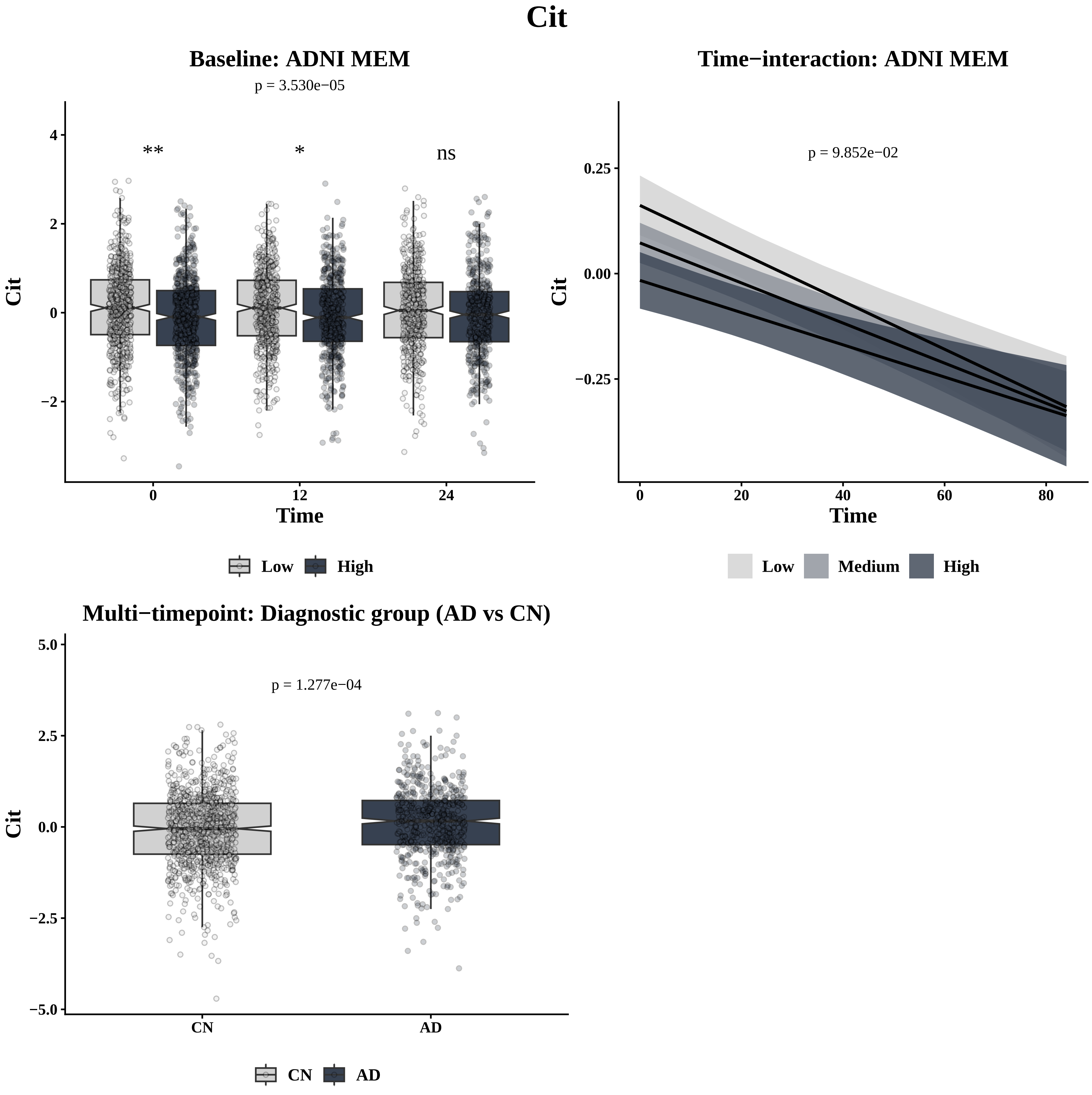

Cit
Baseline: ADNI MEM
p = 3.530e−05
**
*
ns
4
2
0
−2
Cit
0
12
24
Time
Low
High
Time−interaction: ADNI MEM
p = 9.852e−02
0.25
0.00
−0.25
Cit
0
20
40
60
80
Time
Low
Medium
High
Multi−timepoint: Diagnostic group (AD vs CN)
p = 1.277e−04
5.0
2.5
0.0
−2.5
−5.0
Cit
CN
AD
CN
AD

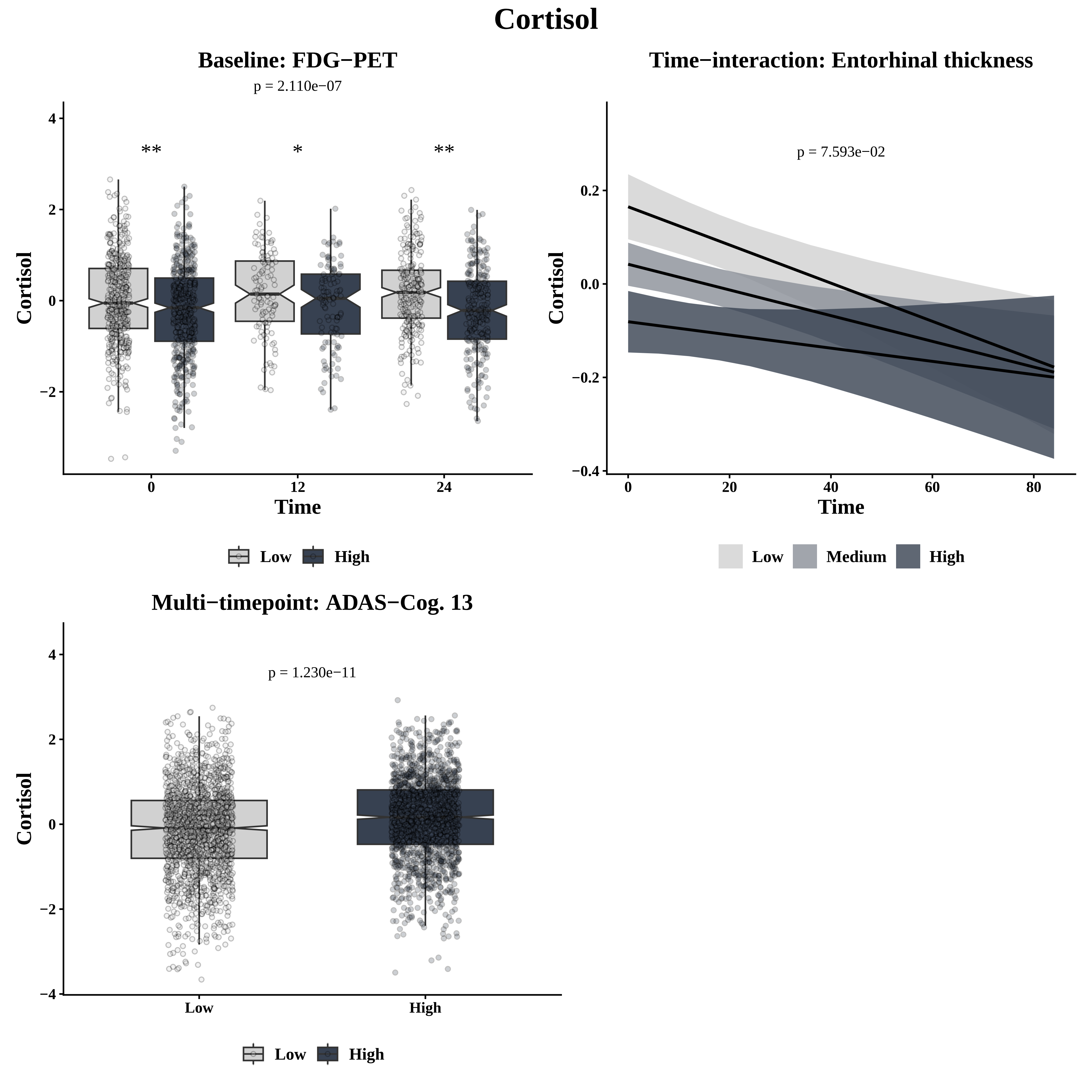

Cortisol
Baseline: FDG−PET
p = 2.110e−07
**
*
**
Cortisol
Time
0
12
24
4
2
0
−2
Low
High
Time−interaction: Entorhinal thickness
p = 7.593e−02
Cortisol
Time
0.2
0.0
−0.2
−0.4
0
20
40
60
80
Low
Medium
High
Multi−timepoint: ADAS−Cog. 13
p = 1.230e−11
Cortisol
4
2
0
−2
−4
Low
High
Low
High

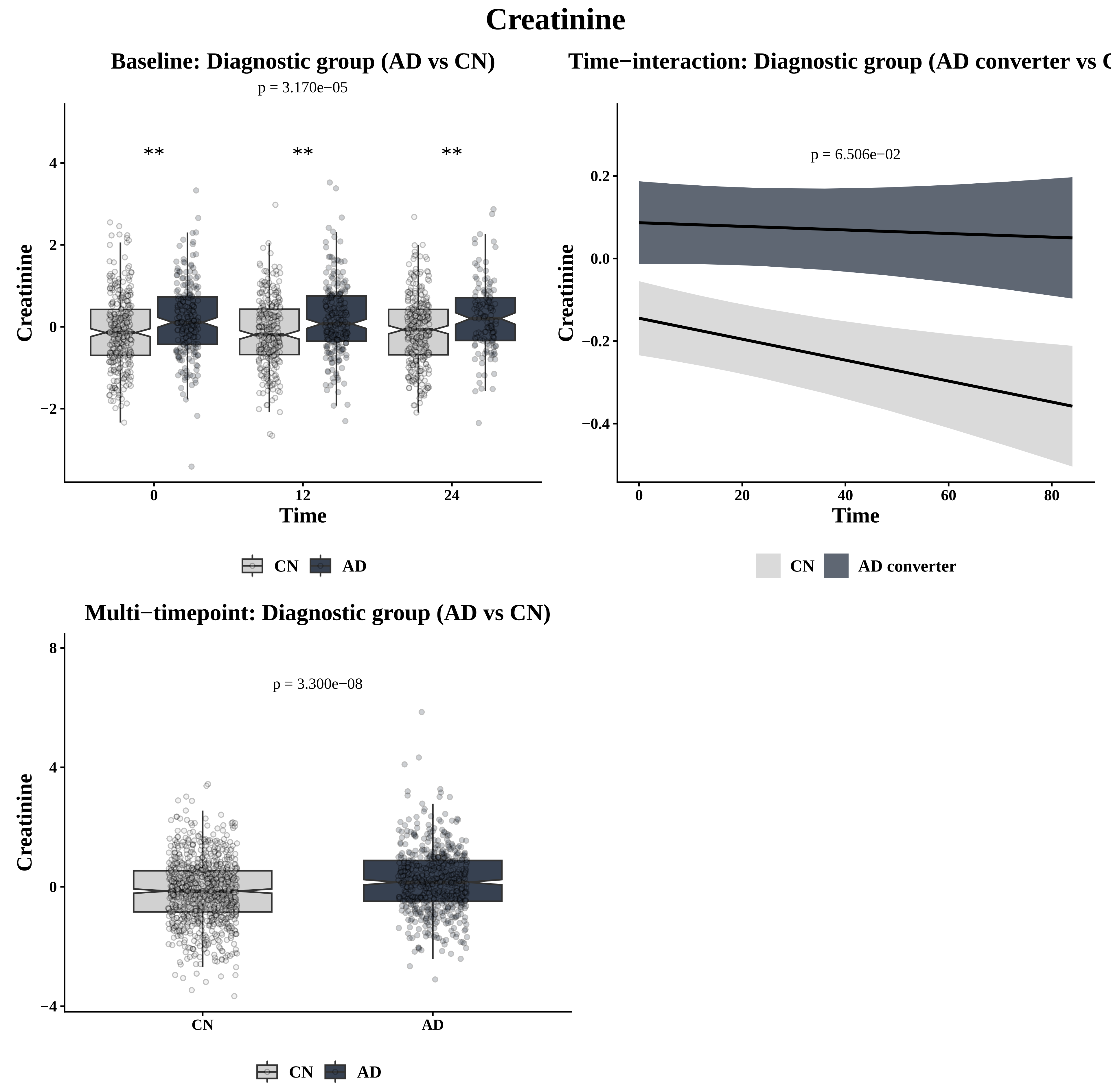
Creatinine
Baseline: Diagnostic group (AD vs CN)
p = 3.170e−05
**
**
**
Creatinine
4
2
0
−2
0
12
24
Time
CN
AD
Time−interaction: Diagnostic group (AD converter vs C
p = 6.506e−02
Creatinine
0.2
0.0
−0.2
−0.4
0
20
40
60
80
Time
CN
AD converter
Multi−timepoint: Diagnostic group (AD vs CN)
p = 3.300e−08
Creatinine
8
4
0
−4
CN
AD
CN
AD

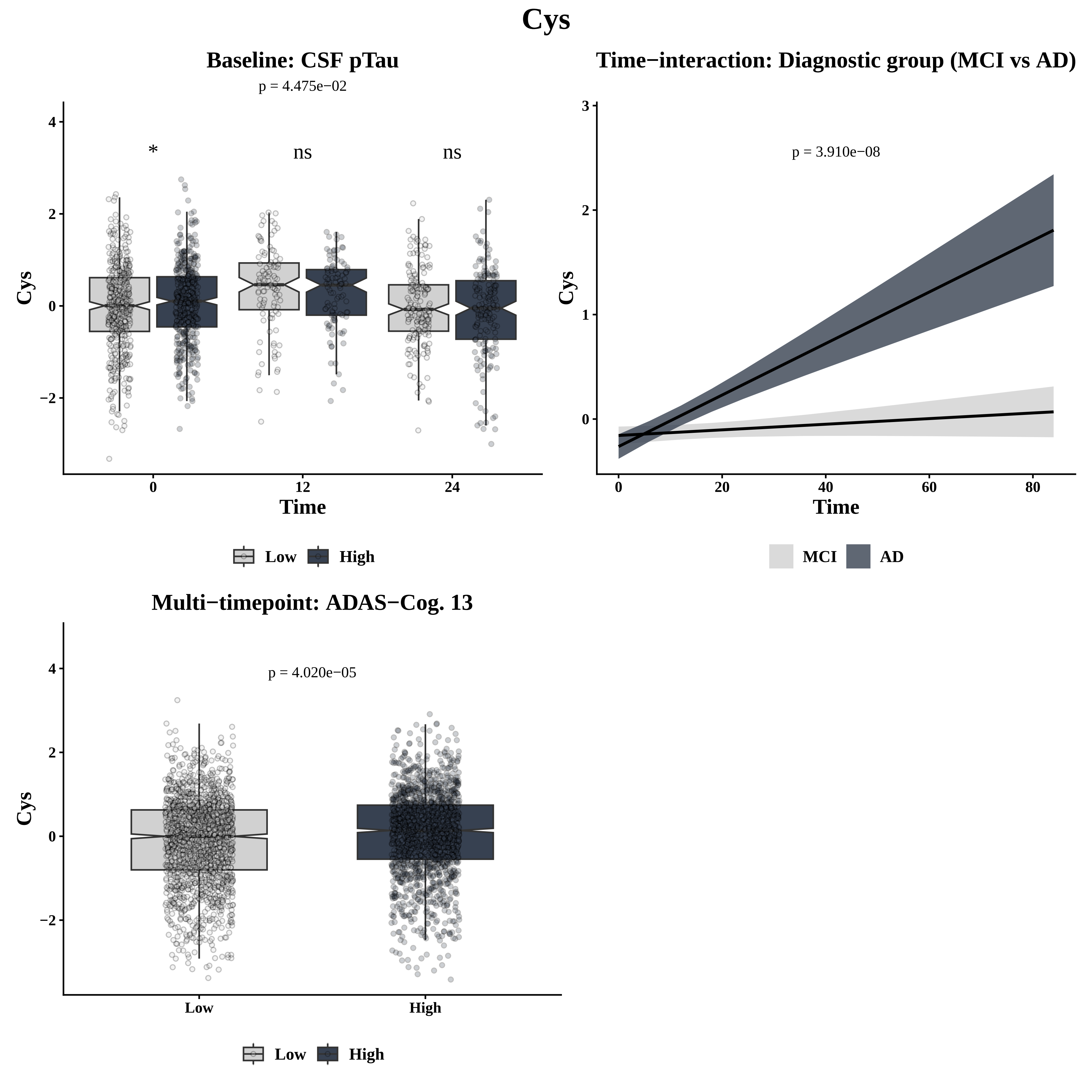

Cys
Baseline: CSF pTau
p = 4.475e−02
*
ns
ns
Cys
Time
0
12
24
Low
High
Time−interaction: Diagnostic group (MCI vs AD)
p = 3.910e−08
Cys
Time
MCI
AD
Multi−timepoint: ADAS−Cog. 13
p = 4.020e−05
Cys
Low
High

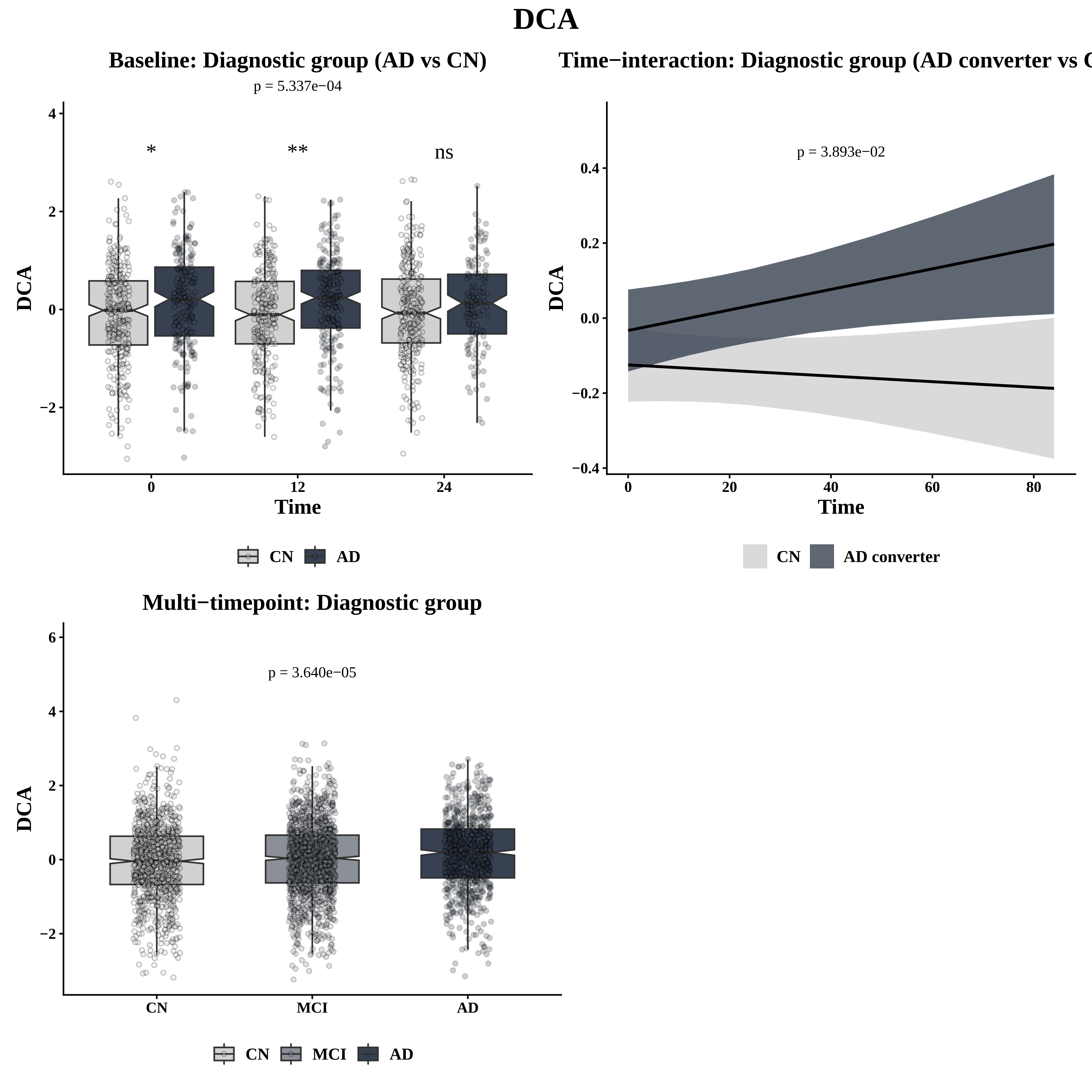
DCA
Baseline: Diagnostic group (AD vs CN)
p = 5.337e−04
*
**
ns
DCA
Time
CN
AD
Time−interaction: Diagnostic group (AD converter vs C
p = 3.893e−02
DCA
Time
CN
AD converter
Multi−timepoint: Diagnostic group
p = 3.640e−05
DCA
CN
MCI
AD

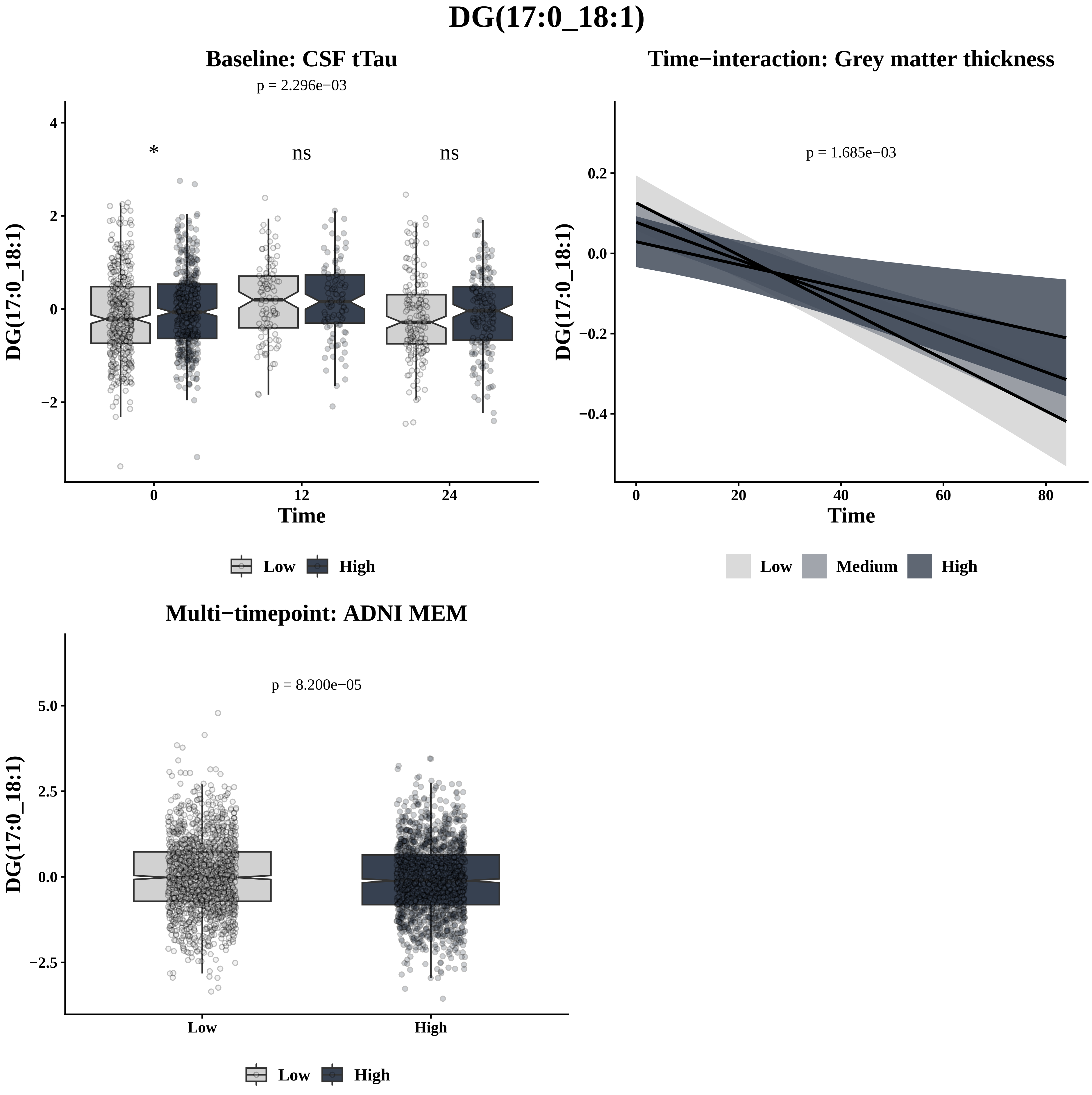
DG(17:0_18:1)
Baseline: CSF tTau
p = 2.296e−03
*
ns
ns
4
2
0
−2
DG(17:0_18:1)
0
12
24
Time
Low
High
Time−interaction: Grey matter thickness
p = 1.685e−03
0.2
0.0
−0.2
−0.4
DG(17:0_18:1)
0
20
40
60
80
Time
Low
Medium
High
Multi−timepoint: ADNI MEM
p = 8.200e−05
5.0
2.5
0.0
−2.5
DG(17:0_18:1)
Low
High
Low
High

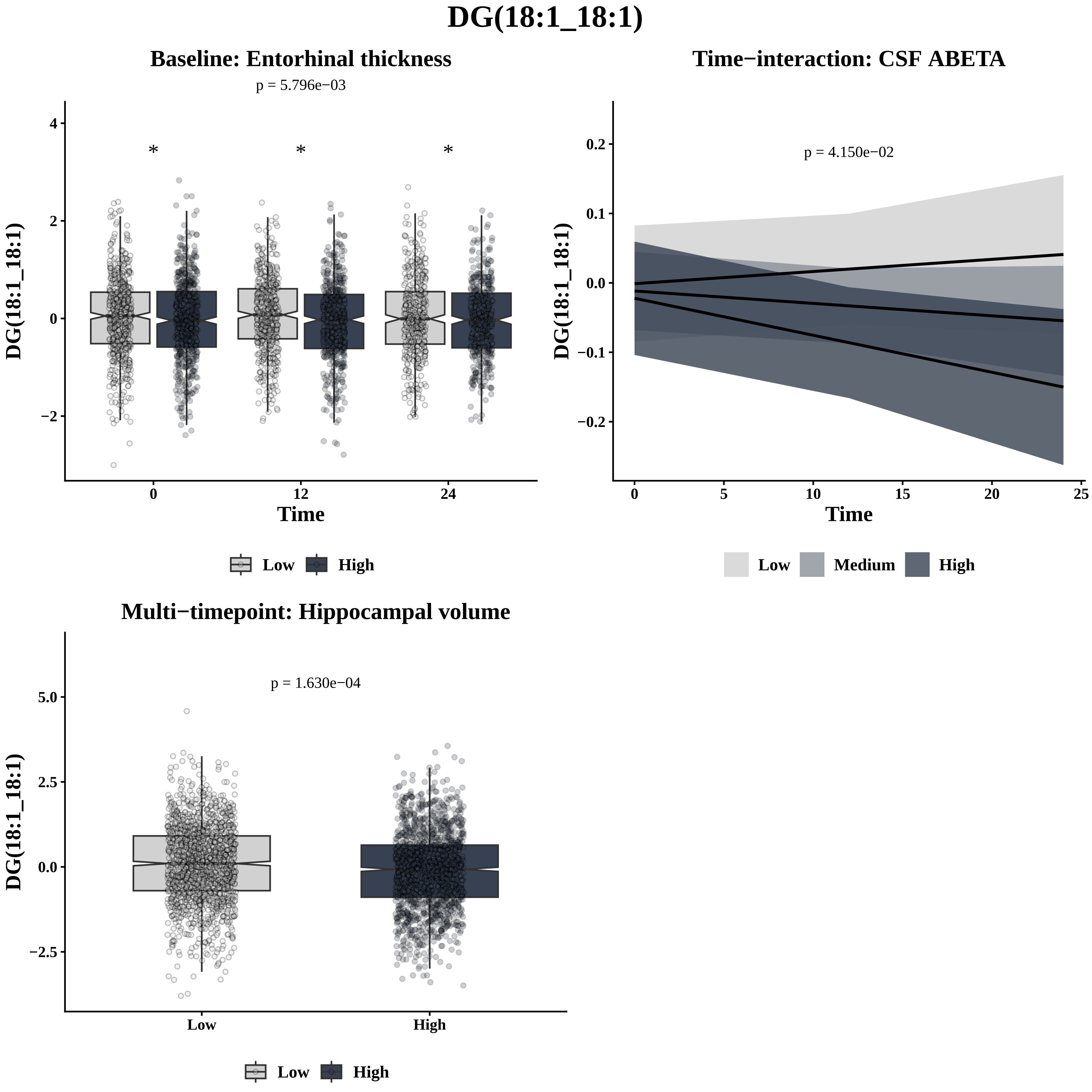
DG(18:1_18:1)
Baseline: Entorhinal thickness
p = 5.796e−03
*
*
*
DG(18:1_18:1)
Time
0
12
24
4
2
0
−2
Low
High
Time−interaction: CSF ABETA
p = 4.150e−02
DG(18:1_18:1)
0.2
0.1
0.0
−0.1
−0.2
0
5
10
15
20
25
Time
Low
Medium
High
Multi−timepoint: Hippocampal volume
p = 1.630e−04
DG(18:1_18:1)
5.0
2.5
0.0
−2.5
Low
High
Low
High

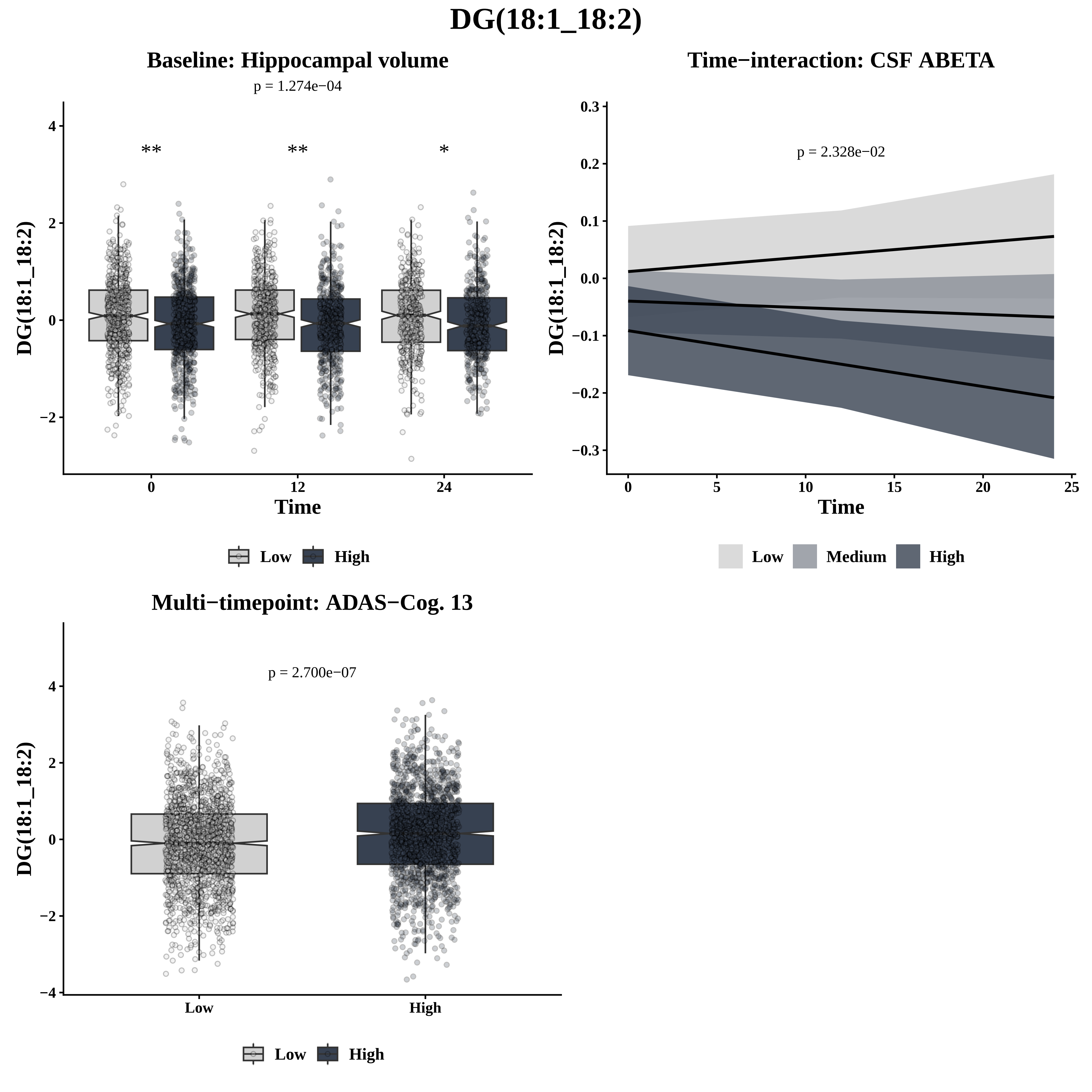
DG(18:1_18:2)
Baseline: Hippocampal volume
p = 1.274e−04
**
**
*
DG(18:1_18:2)
Time
Low
High
Time−interaction: CSF ABETA
p = 2.328e−02
DG(18:1_18:2)
Time
Low
Medium
High
Multi−timepoint: ADAS−Cog. 13
p = 2.700e−07
DG(18:1_18:2)
Low
High
Low
High

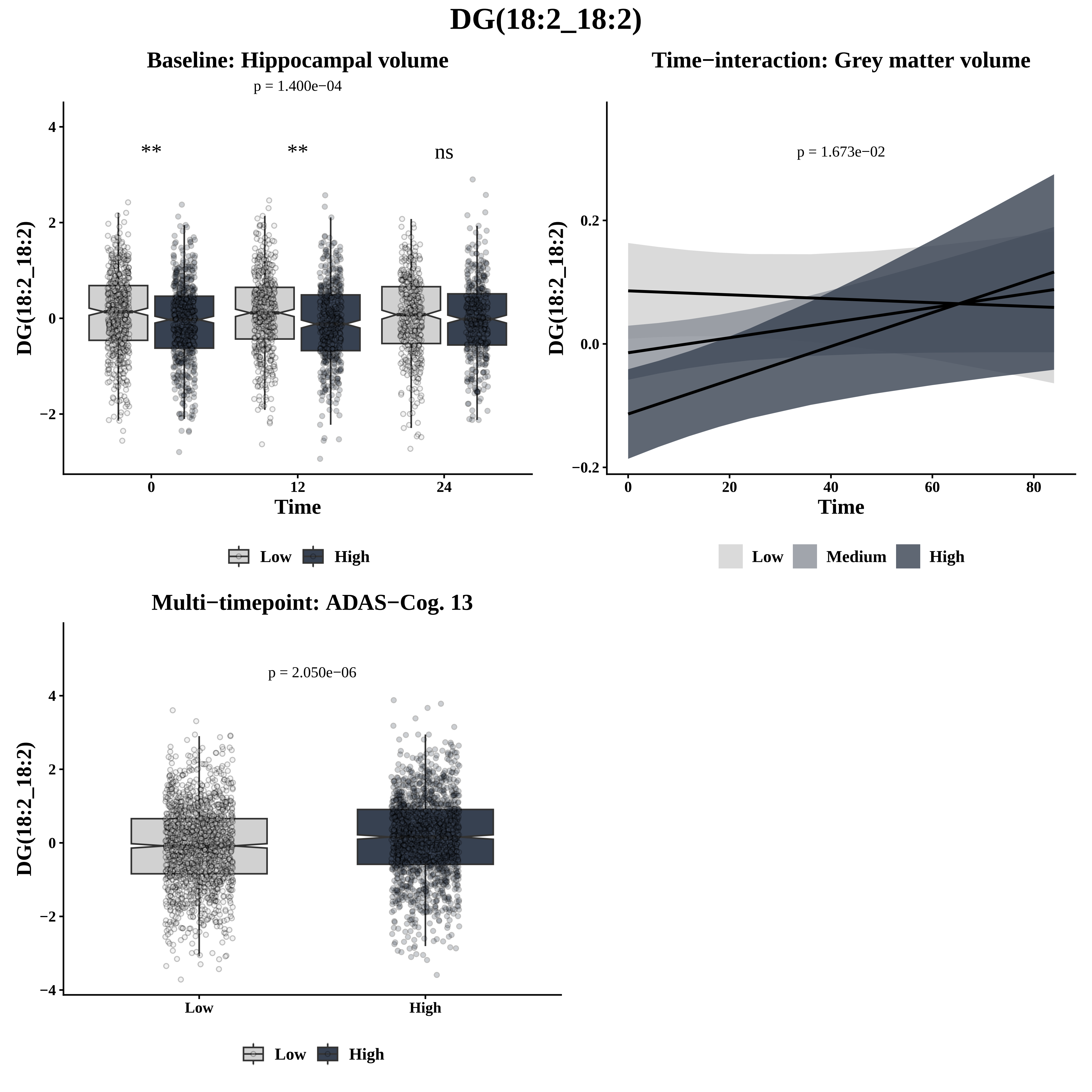

DG(18:2_18:2)
Baseline: Hippocampal volume
p = 1.400e−04
**
**
ns
DG(18:2_18:2)
Time
0
12
24
Low
High
Time−interaction: Grey matter volume
p = 1.673e−02
DG(18:2_18:2)
Time
Low
Medium
High
Multi−timepoint: ADAS−Cog. 13
p = 2.050e−06
DG(18:2_18:2)
Low
High
Low
High

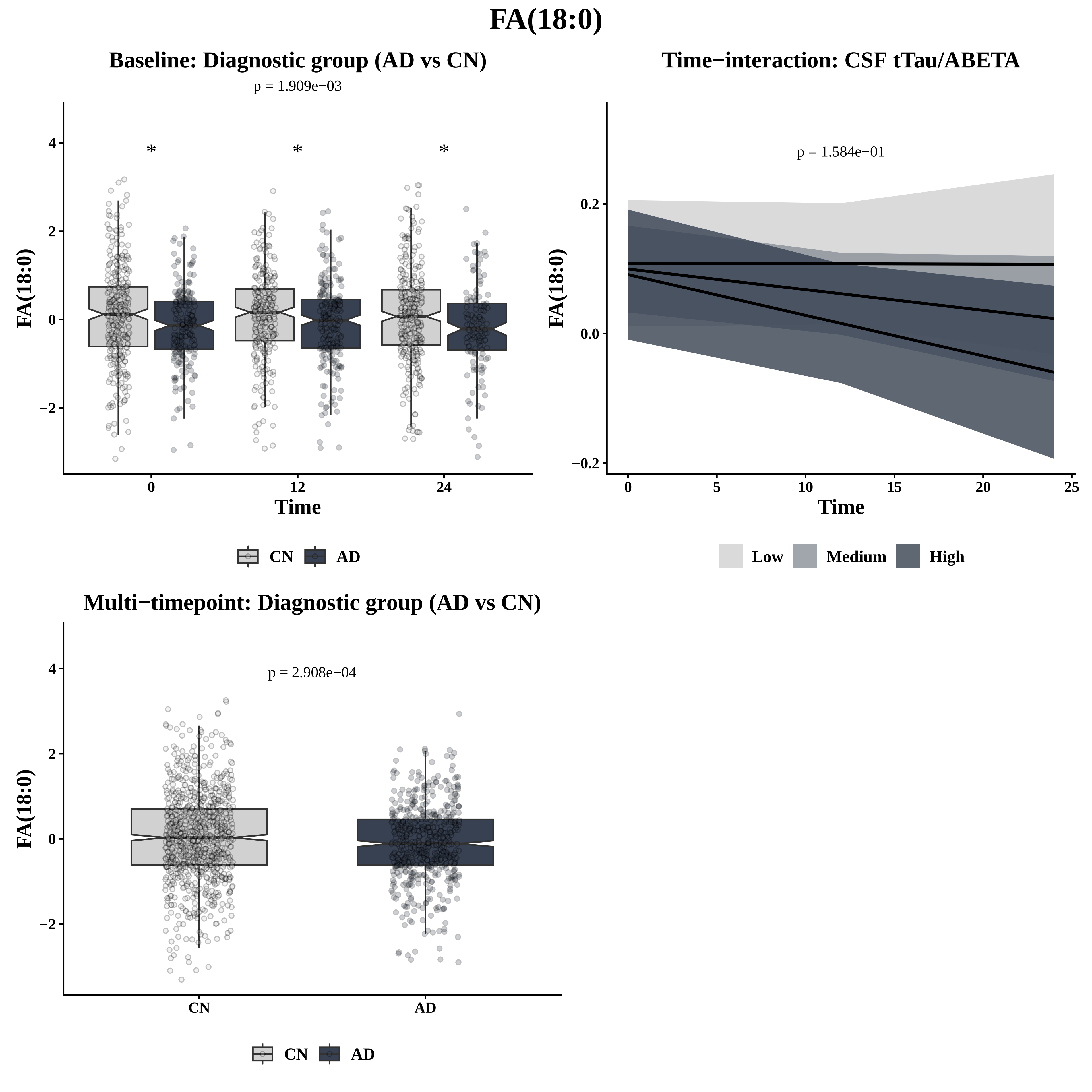
FA(18:0)
Baseline: Diagnostic group (AD vs CN)
p = 1.909e−03
*
*
*
FA(18:0)
Time
0
12
24
CN
AD
Time−interaction: CSF tTau/ABETA
p = 1.584e−01
FA(18:0)
Time
Low
Medium
High
Multi−timepoint: Diagnostic group (AD vs CN)
p = 2.908e−04
FA(18:0)
CN
AD

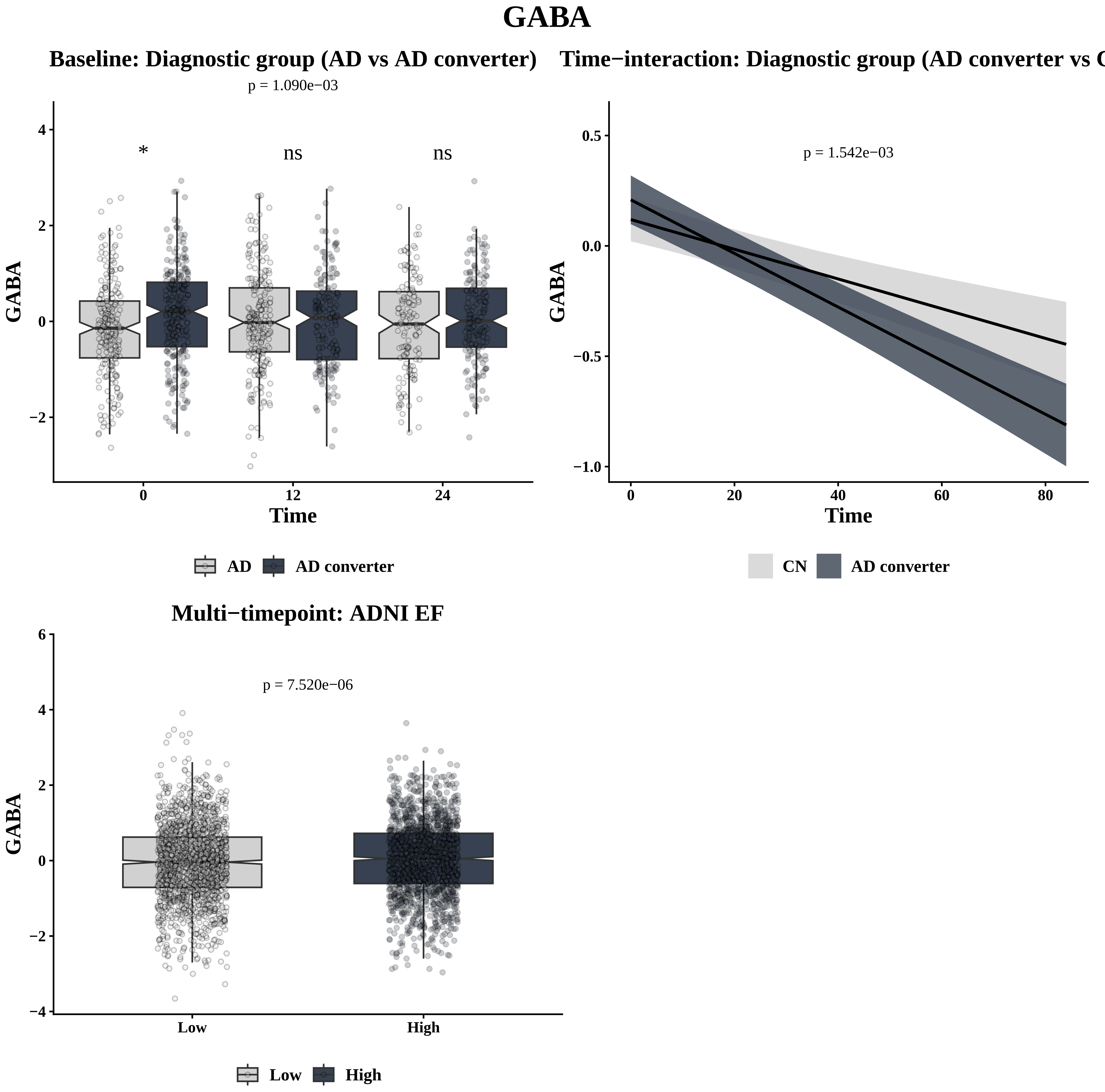

GABA
Baseline: Diagnostic group (AD vs AD converter)
p = 1.090e−03
*
ns
ns
GABA
Time
0
12
24
AD
AD converter
Time−interaction: Diagnostic group (AD converter vs
p = 1.542e−03
GABA
Time
CN
AD converter
Multi−timepoint: ADNI EF
p = 7.520e−06
GABA
Low
High
Low
High

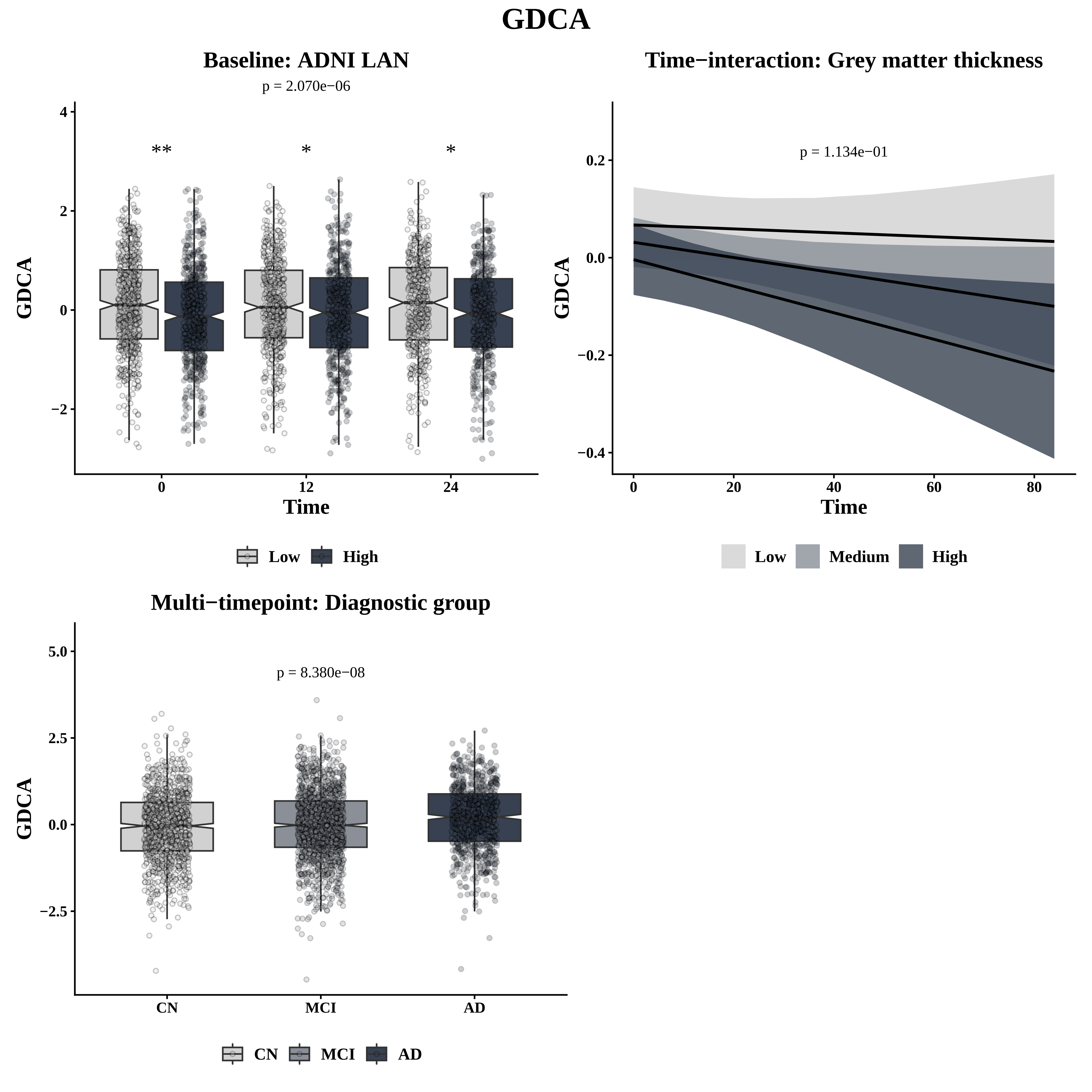
GDCA
Baseline: ADNI LAN
p = 2.070e−06
**
*
*
GDCA
Time
0
12
24
4
2
0
−2
Low
High
Time−interaction: Grey matter thickness
p = 1.134e−01
GDCA
Time
0.2
0.0
−0.2
−0.4
0
20
40
60
80
Low
Medium
High
Multi−timepoint: Diagnostic group
p = 8.380e−08
GDCA
5.0
2.5
0.0
−2.5
CN
MCI
AD
CN
MCI
AD

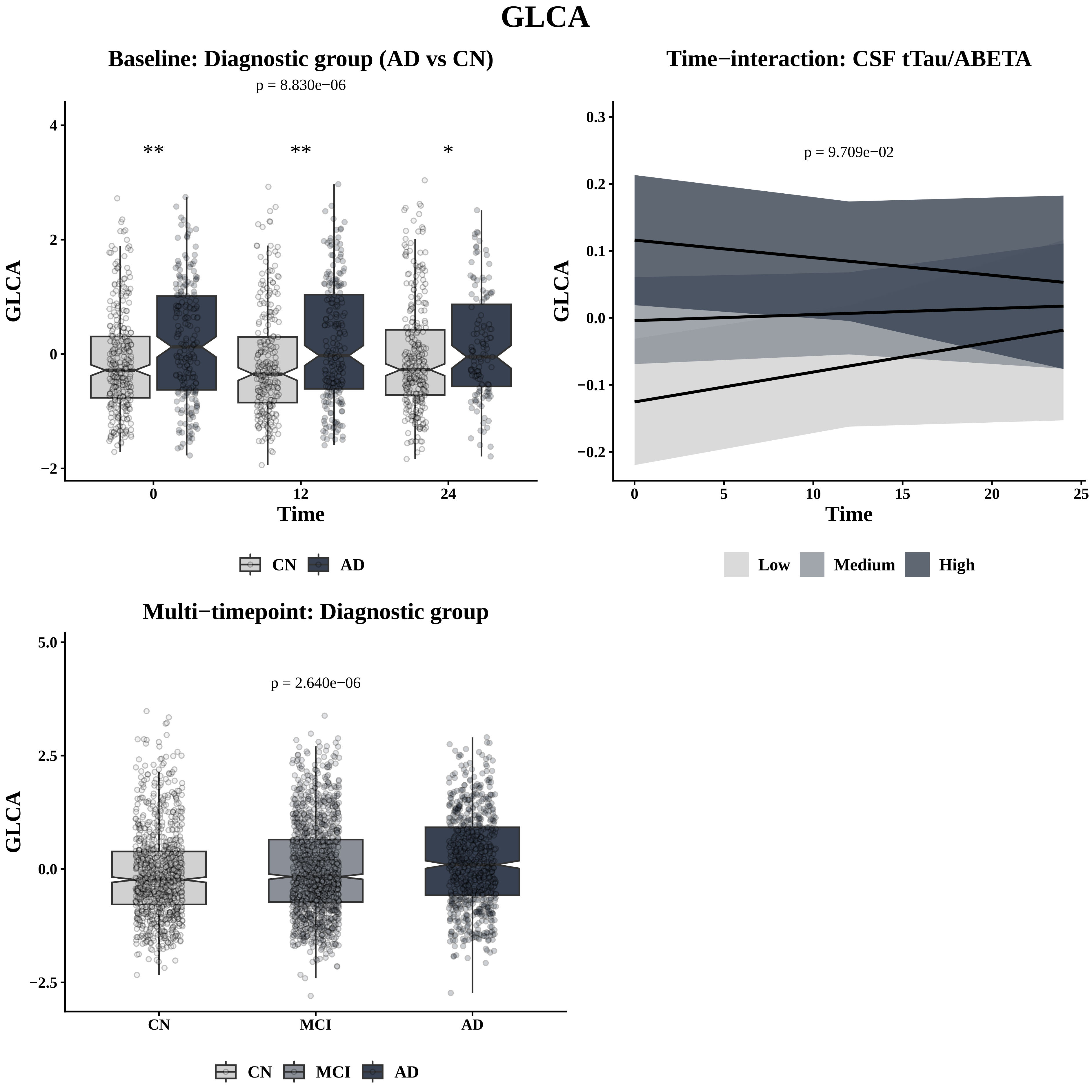
GLCA
Baseline: Diagnostic group (AD vs CN)
p = 8.830e−06
**
**
*
GLCA
Time
CN
AD
Time−interaction: CSF tTau/ABETA
p = 9.709e−02
GLCA
Time
Low
Medium
High
Multi−timepoint: Diagnostic group
p = 2.640e−06
GLCA
CN
MCI
AD

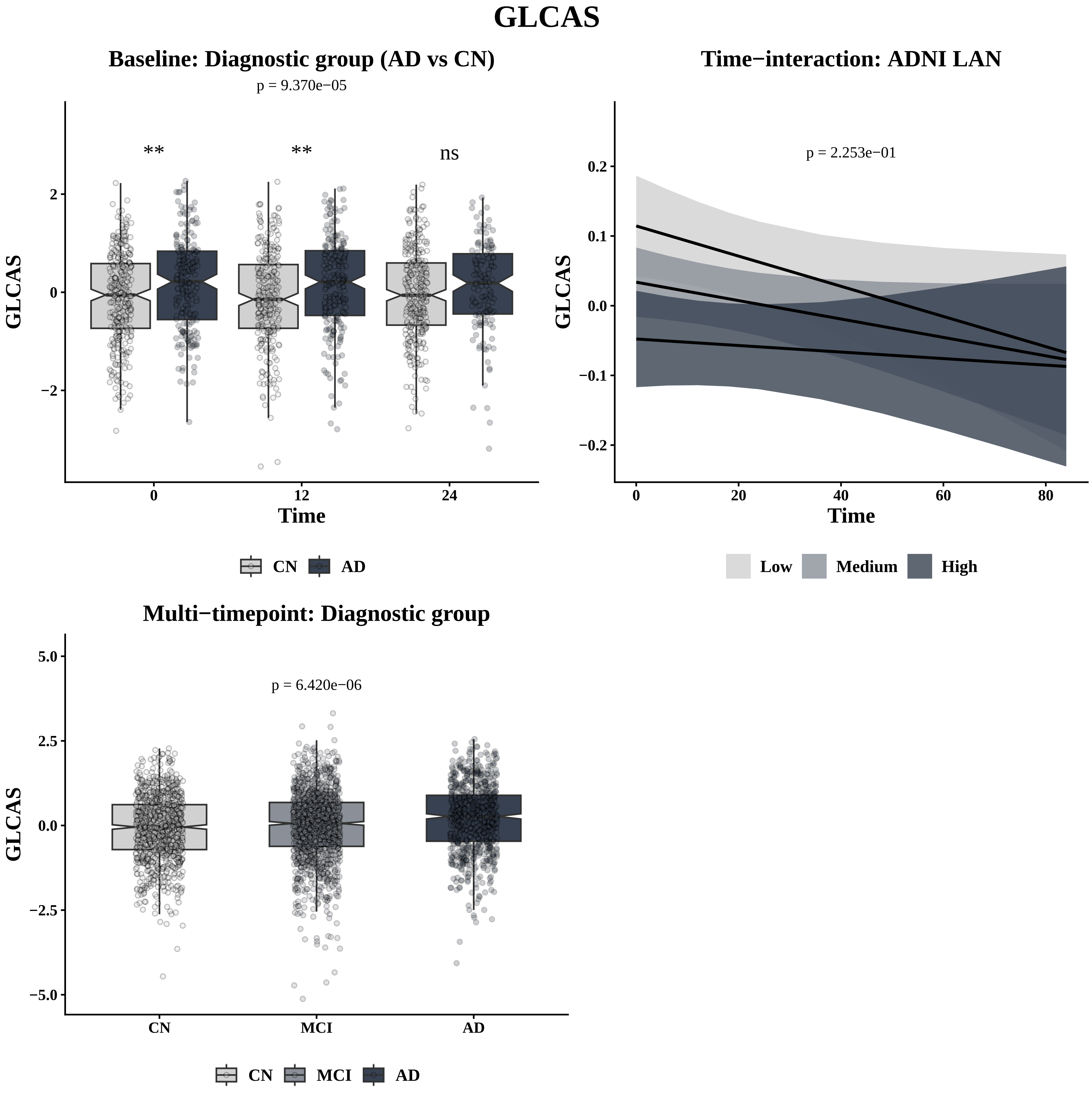
GLCAS
Baseline: Diagnostic group (AD vs CN)
p = 9.370e−05
**
**
ns
GLCAS
2
0
−2
0
12
24
Time
CN
AD
Time−interaction: ADNI LAN
p = 2.253e−01
GLCAS
0.2
0.1
0.0
−0.1
−0.2
0
20
40
60
80
Time
Low
Medium
High
Multi−timepoint: Diagnostic group
p = 6.420e−06
GLCAS
5.0
2.5
0.0
−2.5
−5.0
CN
MCI
AD
CN
MCI
AD

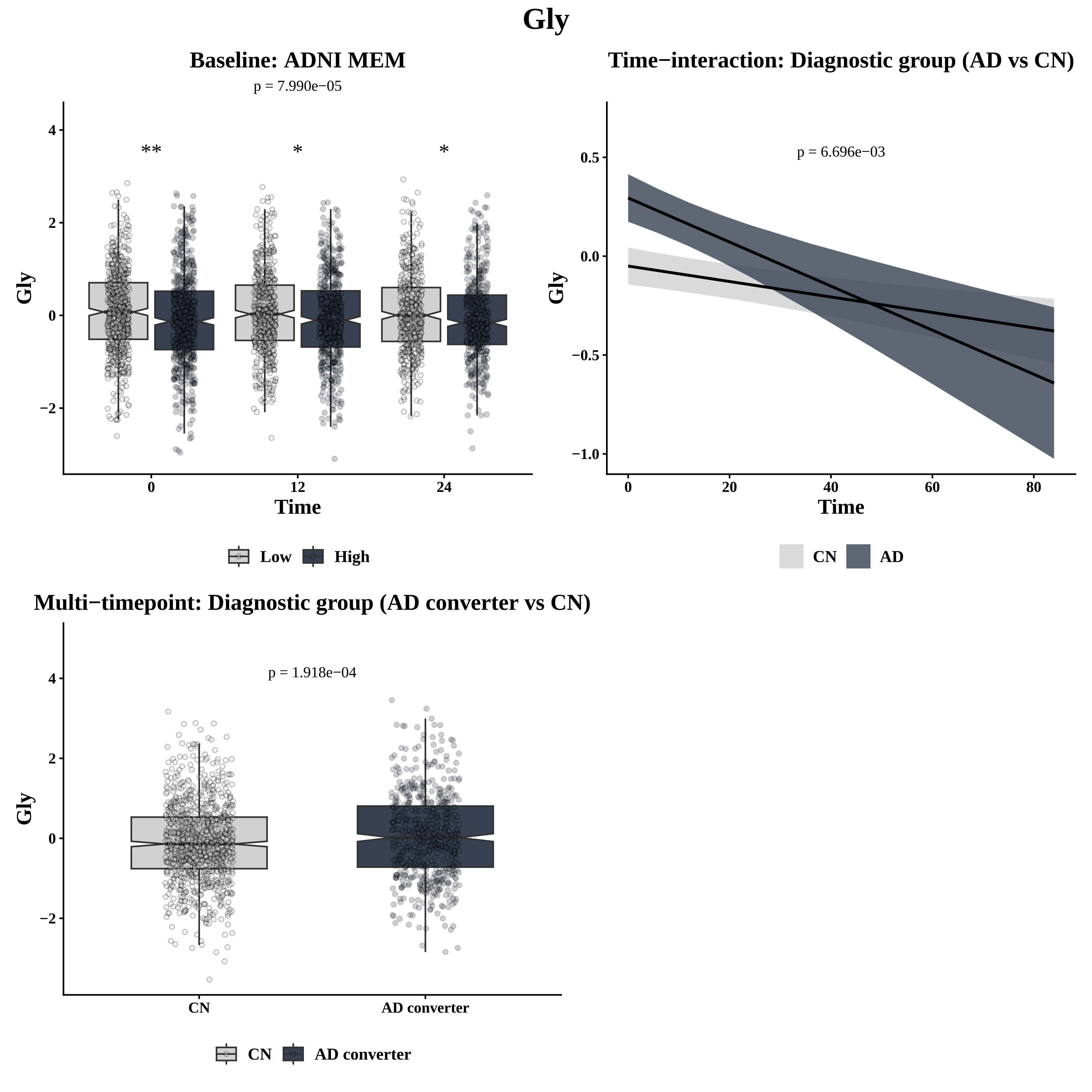

Gly
Baseline: ADNI MEM
p = 7.990e−05
**
*
*
Gly
Time
0
12
24
Low
High
Time−interaction: Diagnostic group (AD vs CN)
p = 6.696e−03
Gly
Time
CN
AD
Multi−timepoint: Diagnostic group (AD converter vs CN)
p = 1.918e−04
Gly
CN
AD converter

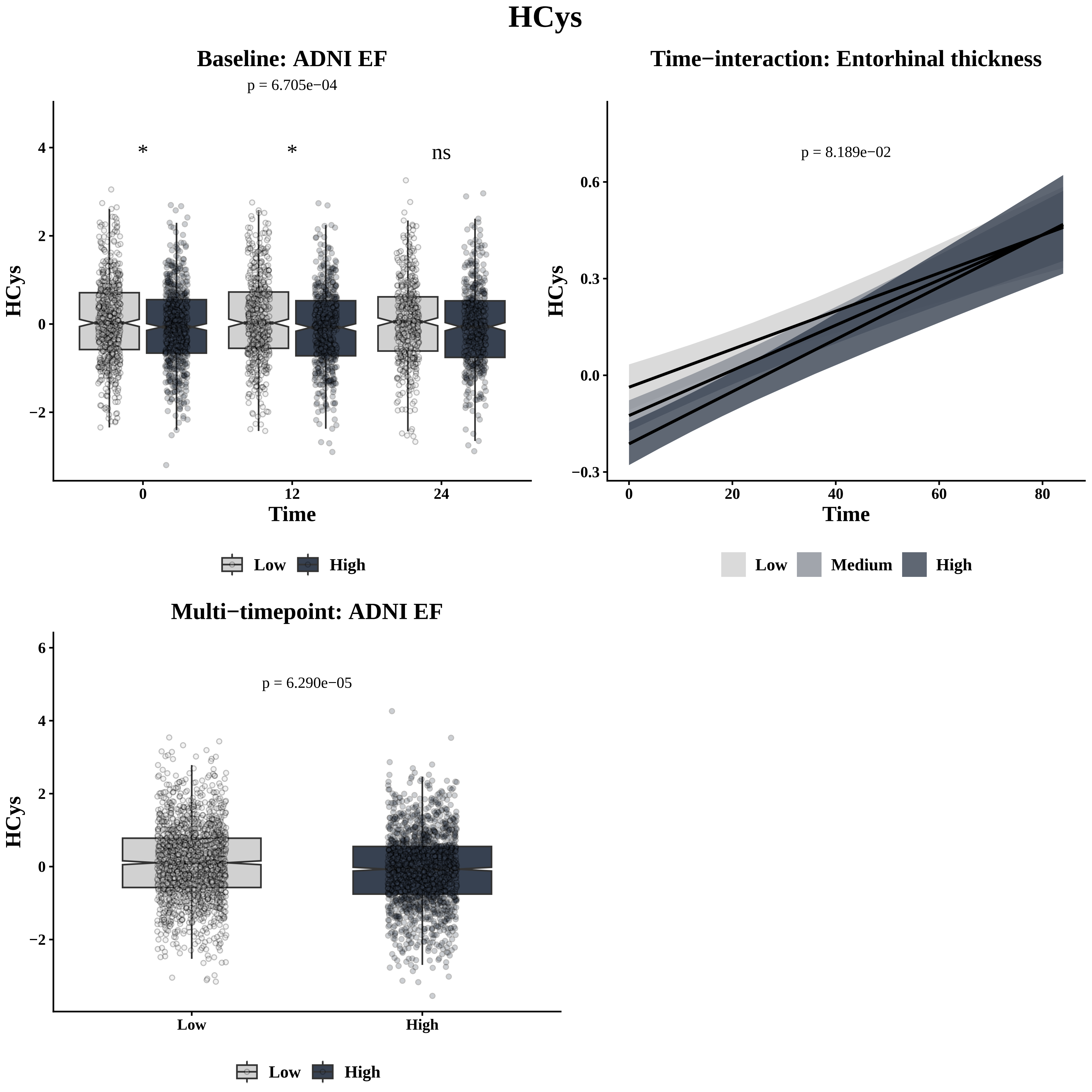
HCys
Baseline: ADNI EF
p = 6.705e−04
*
*
ns
HCys
Time
0
12
24
Low
High
Time−interaction: Entorhinal thickness
p = 8.189e−02
HCys
Time
Low
Medium
High
Multi−timepoint: ADNI EF
p = 6.290e−05
HCys
Low
High

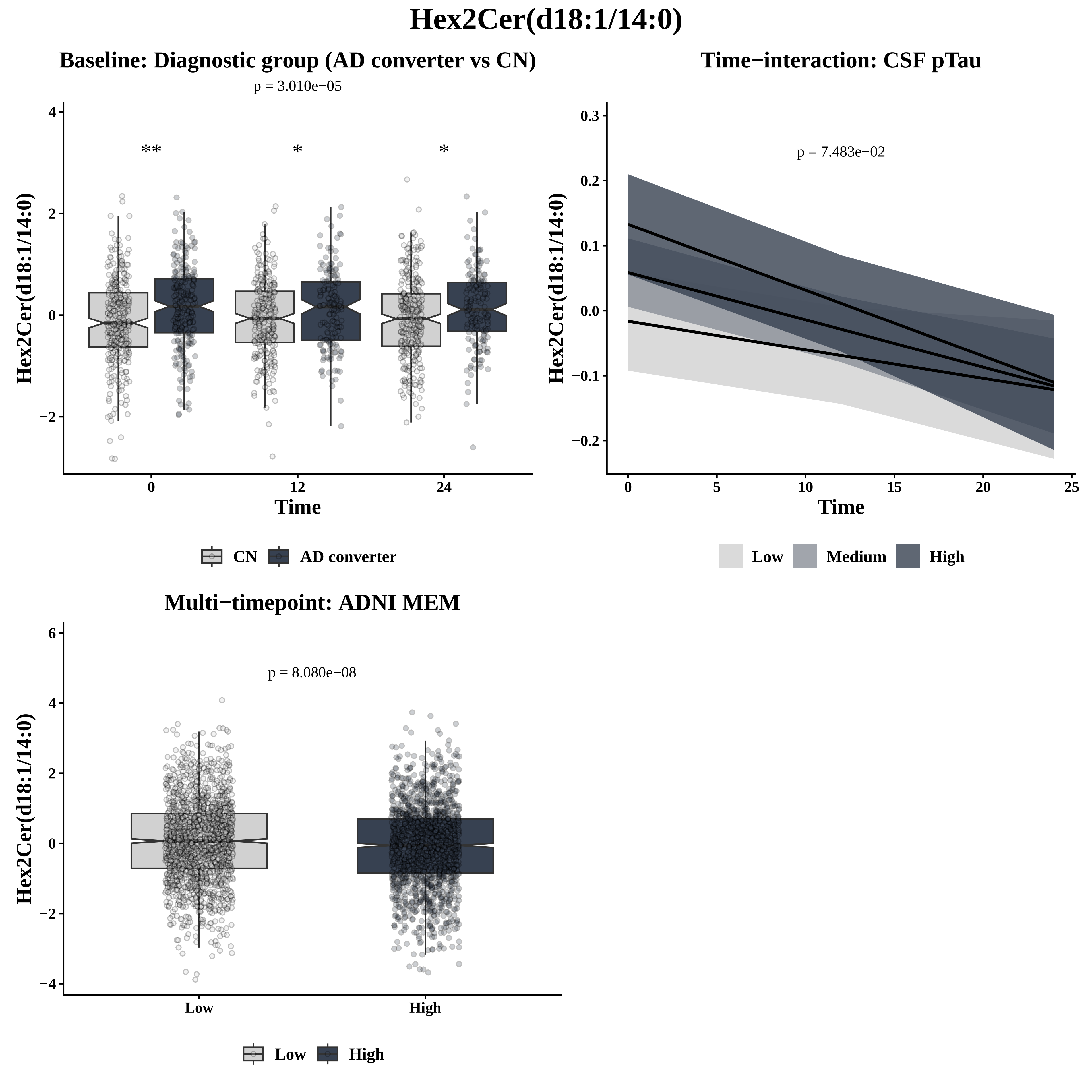
Hex2Cer(d18:1/14:0)
Baseline: Diagnostic group (AD converter vs CN)
p = 3.010e−05
**
*
*
Hex2Cer(d18:1/14:0)
4
2
0
−2
0
12
24
Time
CN
AD converter
Time−interaction: CSF pTau
p = 7.483e−02
Hex2Cer(d18:1/14:0)
0.3
0.2
0.1
0.0
−0.1
−0.2
0
5
10
15
20
25
Time
Low
Medium
High
Multi−timepoint: ADNI MEM
p = 8.080e−08
Hex2Cer(d18:1/14:0)
6
4
2
0
−2
−4
Low
High
Low
High

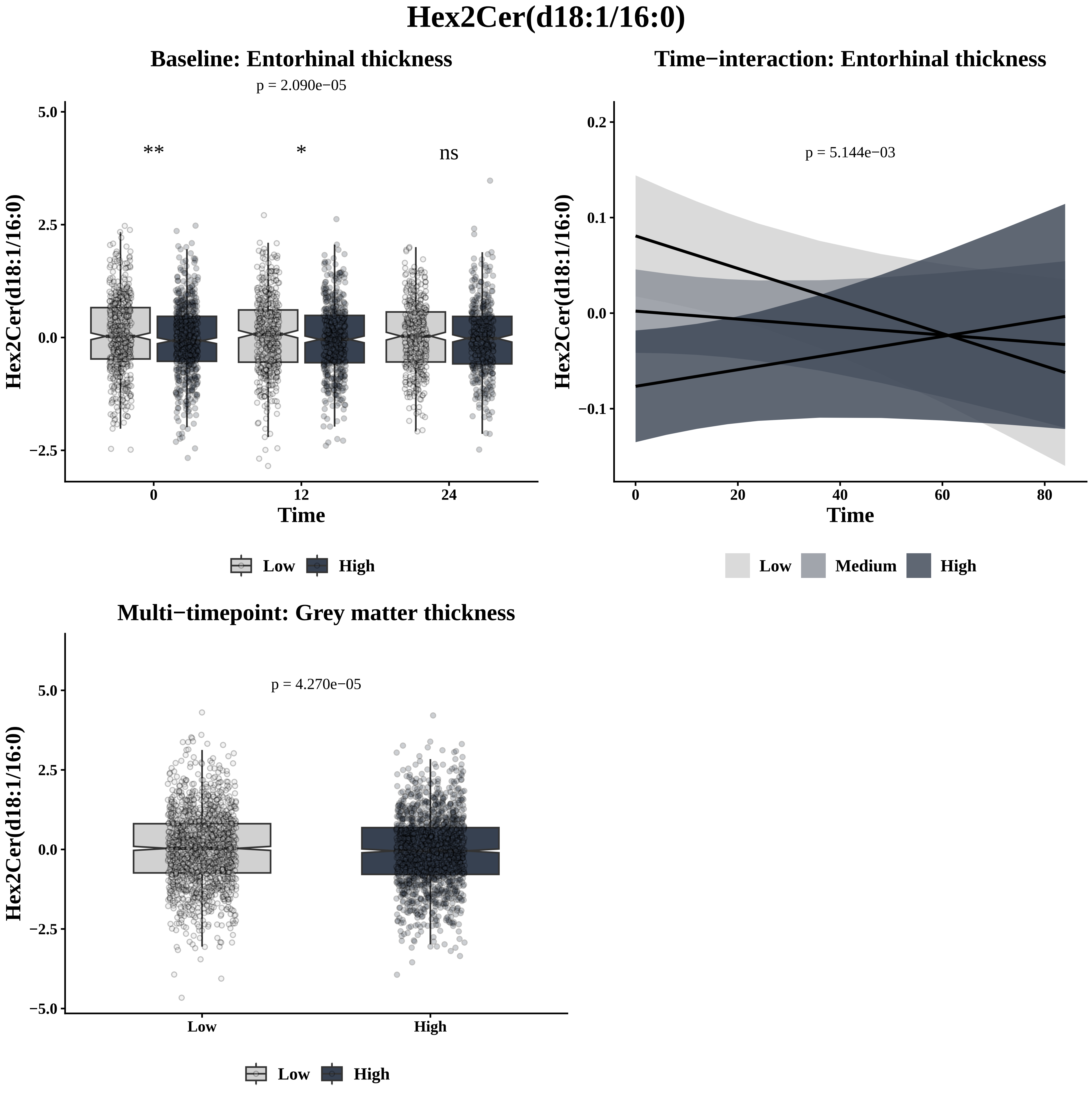

Hex2Cer(d18:1/16:0)
Baseline: Entorhinal thickness
p = 2.090e−05
**
*
ns
5.0
2.5
0.0
−2.5
Hex2Cer(d18:1/16:0)
0
12
24
Time
Low
High
Time−interaction: Entorhinal thickness
p = 5.144e−03
0.2
0.1
0.0
−0.1
Hex2Cer(d18:1/16:0)
0
20
40
60
80
Time
Low
Medium
High
Multi−timepoint: Grey matter thickness
p = 4.270e−05
5.0
2.5
0.0
−2.5
−5.0
Hex2Cer(d18:1/16:0)
Low
High
Low
High

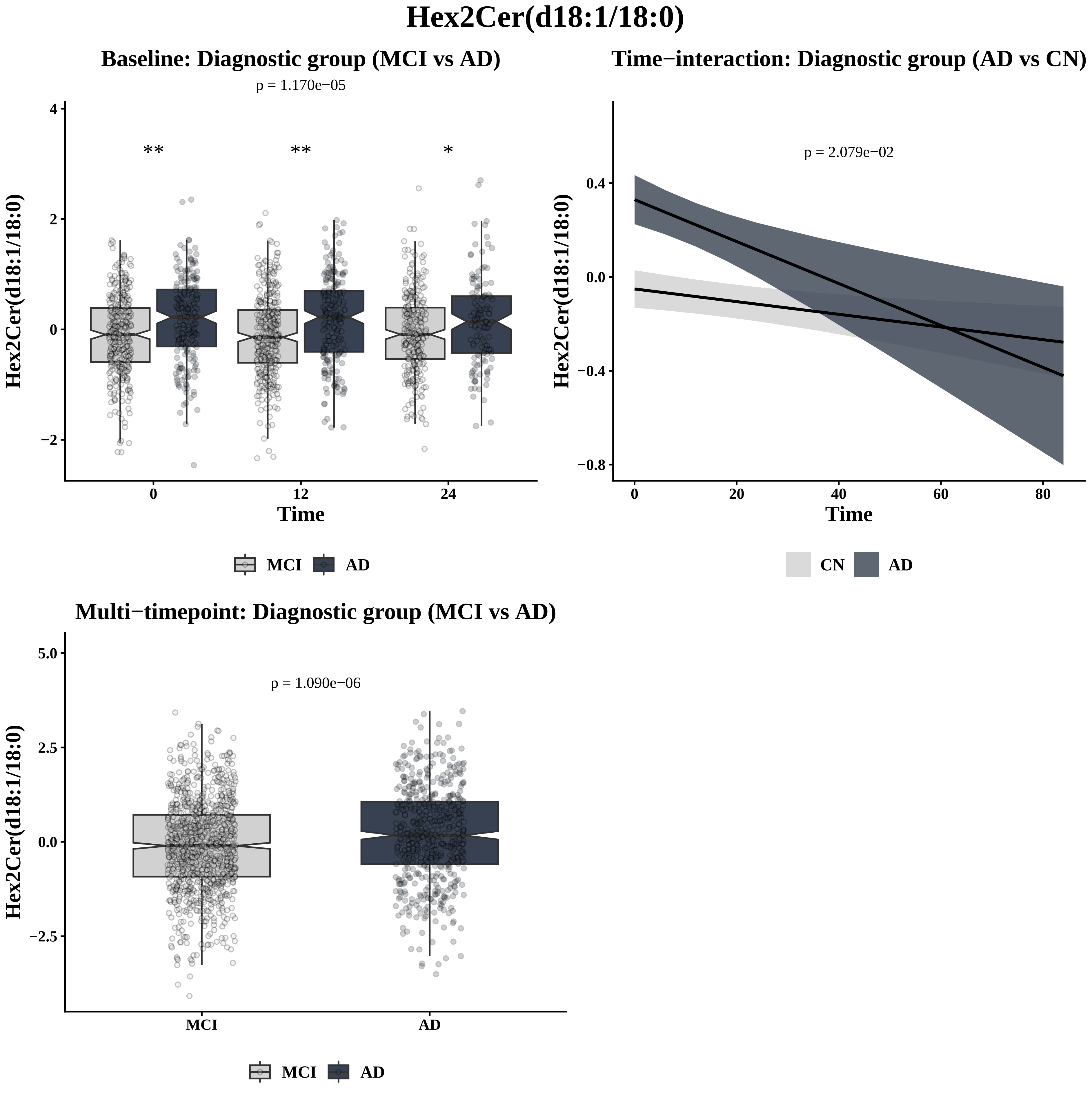
Hex2Cer(d18:1/18:0)
Baseline: Diagnostic group (MCI vs AD)
p = 1.170e−05
**
**
*
Hex2Cer(d18:1/18:0)
Time
0
12
24
4
2
0
−2
MCI
AD
Time−interaction: Diagnostic group (AD vs CN)
p = 2.079e−02
Hex2Cer(d18:1/18:0)
0.4
0.0
−0.4
−0.8
0
20
40
60
80
Time
CN
AD
Multi−timepoint: Diagnostic group (MCI vs AD)
p = 1.090e−06
Hex2Cer(d18:1/18:0)
5.0
2.5
0.0
−2.5
MCI
AD
MCI
AD

### Hex2Cer(d18:1/20:0)

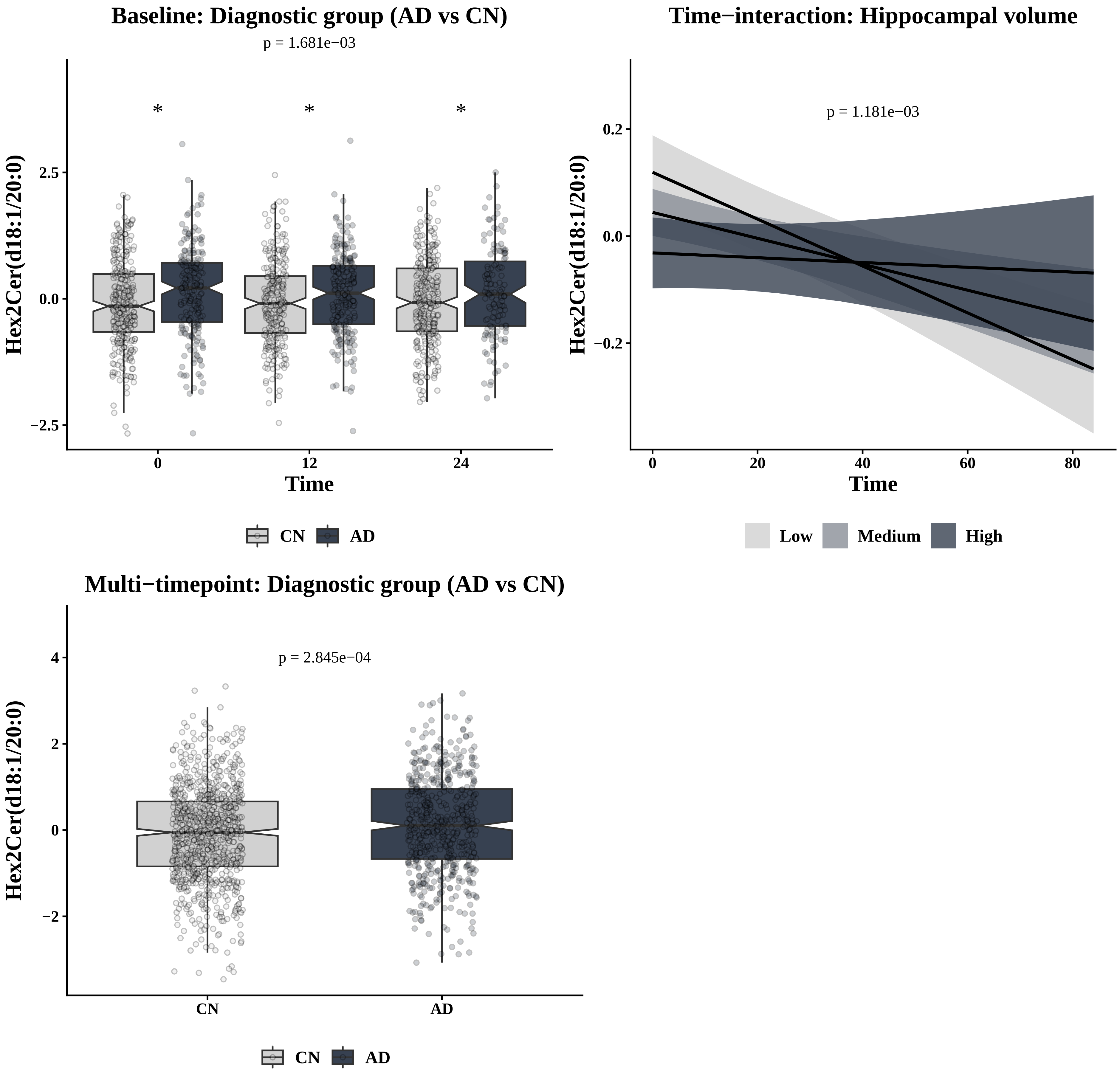

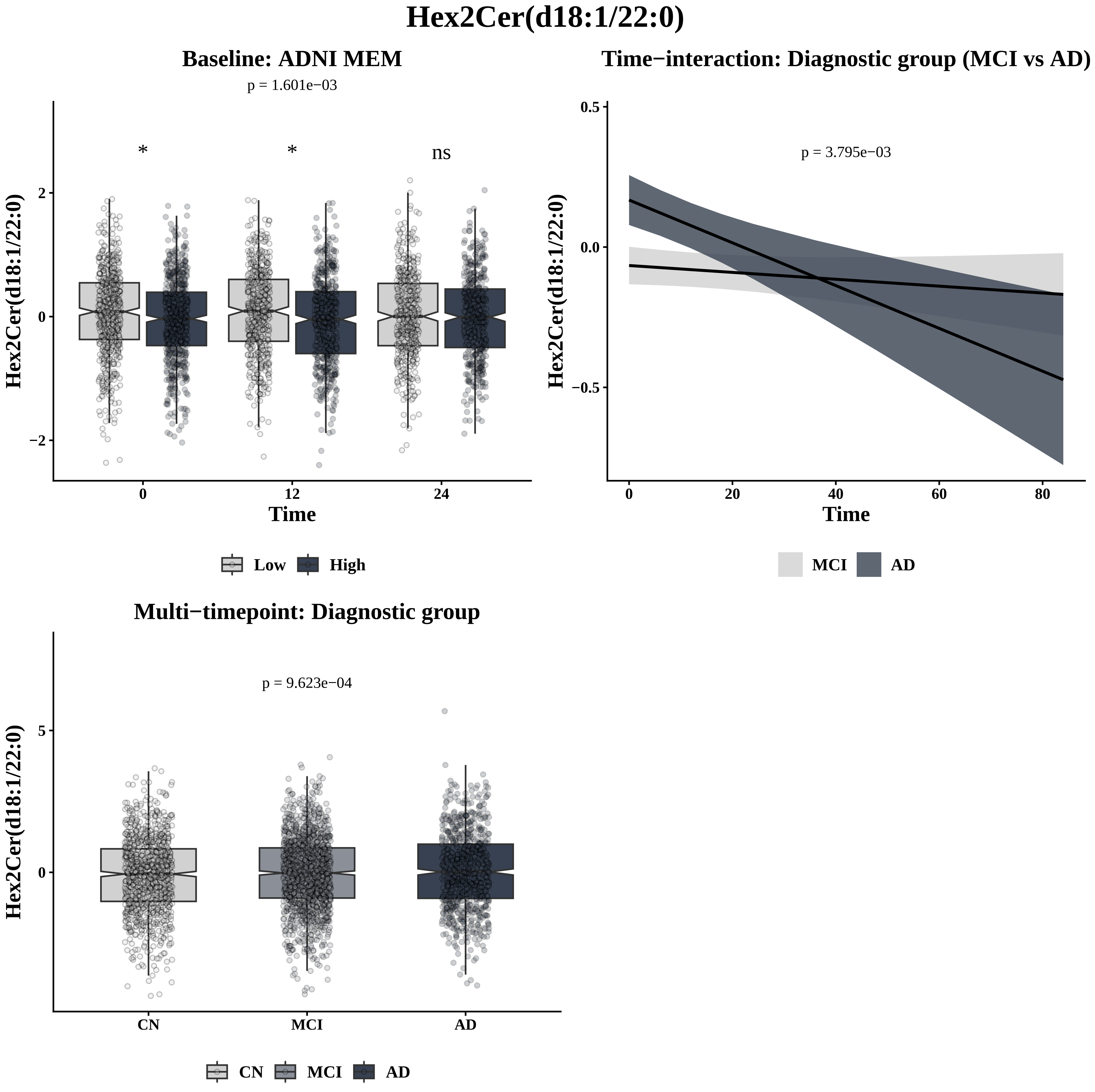

Hex2Cer(d18:1/22:0)
Baseline: ADNI MEM
p = 1.601e−03
*
*
ns
Hex2Cer(d18:1/22:0)
2
0
−2
0
12
24
Time
Low
High
Time−interaction: Diagnostic group (MCI vs AD)
p = 3.795e−03
Hex2Cer(d18:1/22:0)
0.5
0.0
−0.5
0
20
40
60
80
Time
MCI
AD
Multi−timepoint: Diagnostic group
p = 9.623e−04
Hex2Cer(d18:1/22:0)
5
0
CN
MCI
AD
CN
MCI
AD

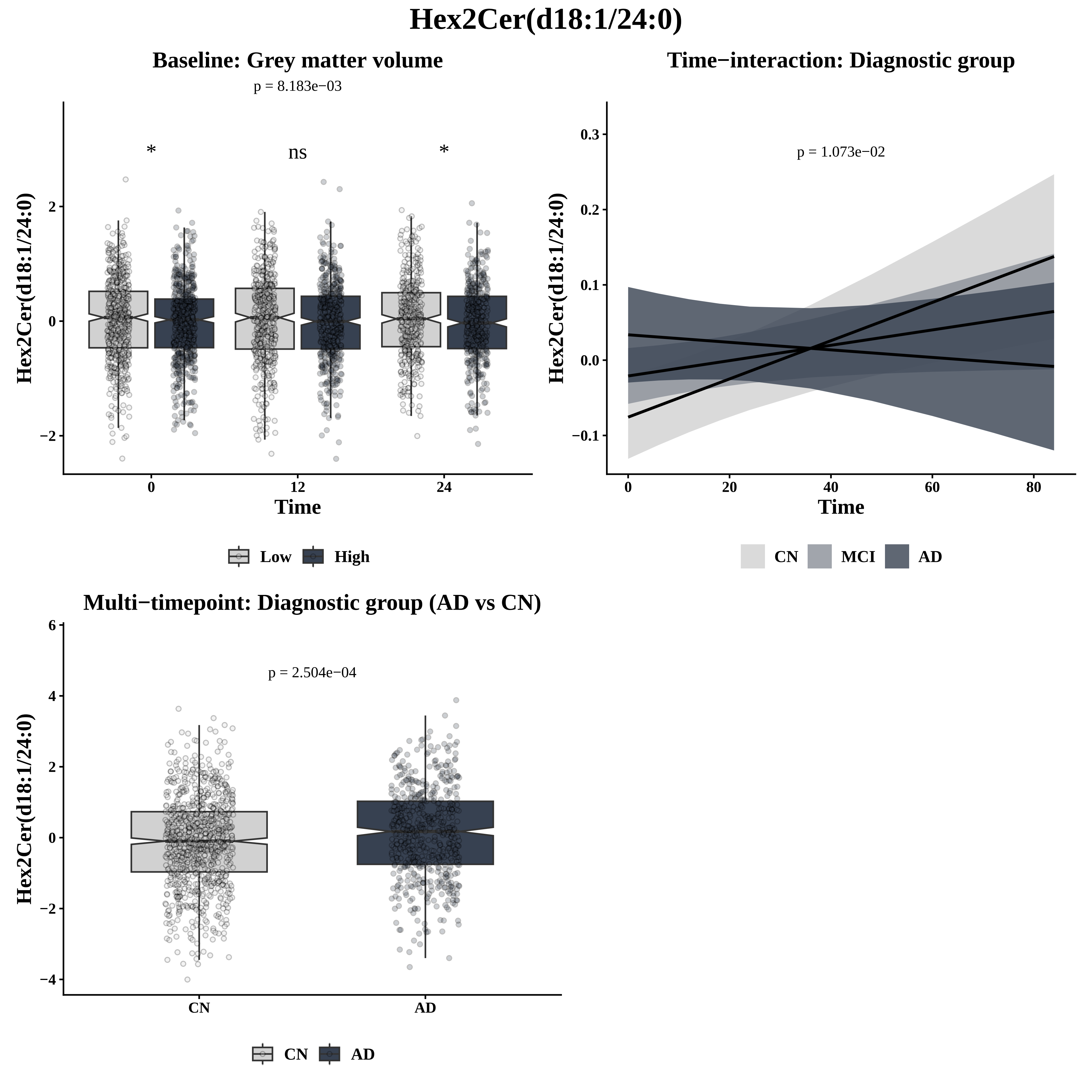

Hex2Cer(d18:1/24:0)
Baseline: Grey matter volume
p = 8.183e−03
*
ns
*
Hex2Cer(d18:1/24:0)
Time
Low
High
Time−interaction: Diagnostic group
p = 1.073e−02
Time
CN
MCI
AD
Multi−timepoint: Diagnostic group (AD vs CN)
p = 2.504e−04
CN
AD

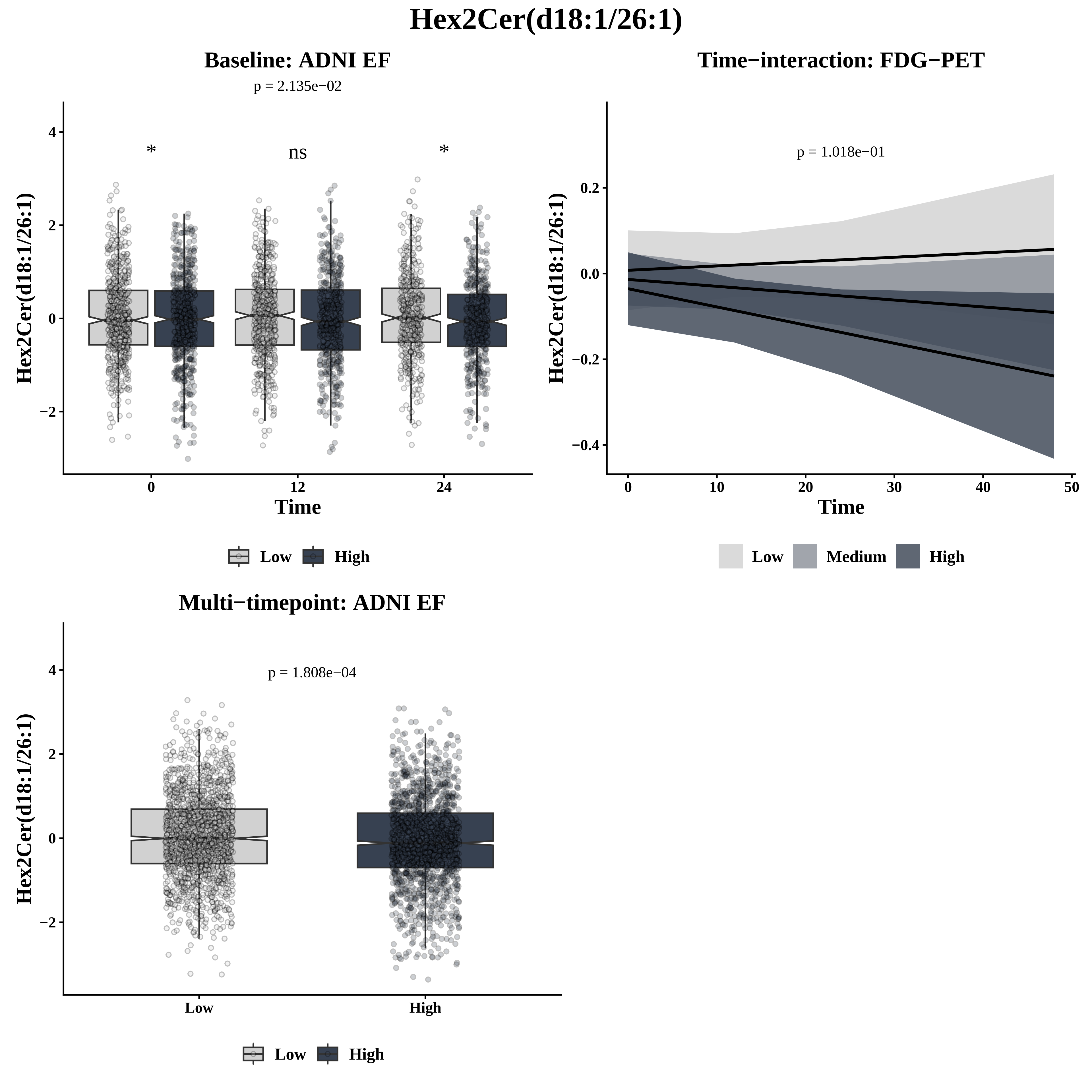
Hex2Cer(d18:1/26:1)
Baseline: ADNI EF
p = 2.135e−02
*
ns
*
Hex2Cer(d18:1/26:1)
Time
Low
High
Time−interaction: FDG−PET
p = 1.018e−01
Hex2Cer(d18:1/26:1)
Time
Low
Medium
High
Multi−timepoint: ADNI EF
p = 1.808e−04
Hex2Cer(d18:1/26:1)
Low
High

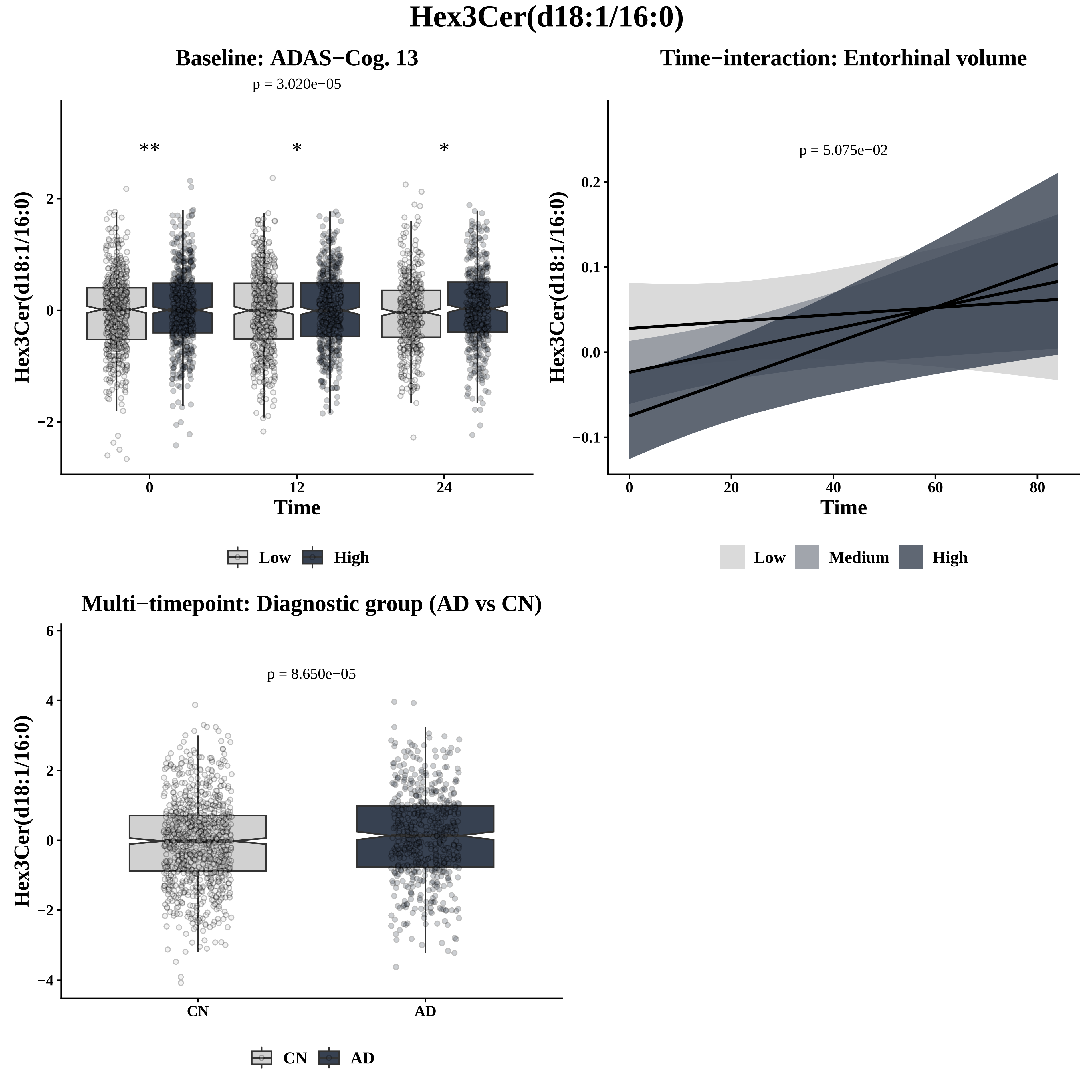
Hex3Cer(d18:1/16:0)
Baseline: ADAS−Cog. 13
p = 3.020e−05
**
*
*
Hex3Cer(d18:1/16:0)
2
0
−2
0
12
24
Time
Low
High
Time−interaction: Entorhinal volume
p = 5.075e−02
Hex3Cer(d18:1/16:0)
0.2
0.1
0.0
−0.1
0
20
40
60
80
Time
Low
Medium
High
Multi−timepoint: Diagnostic group (AD vs CN)
p = 8.650e−05
Hex3Cer(d18:1/16:0)
6
4
2
0
−2
−4
CN
AD
CN
AD

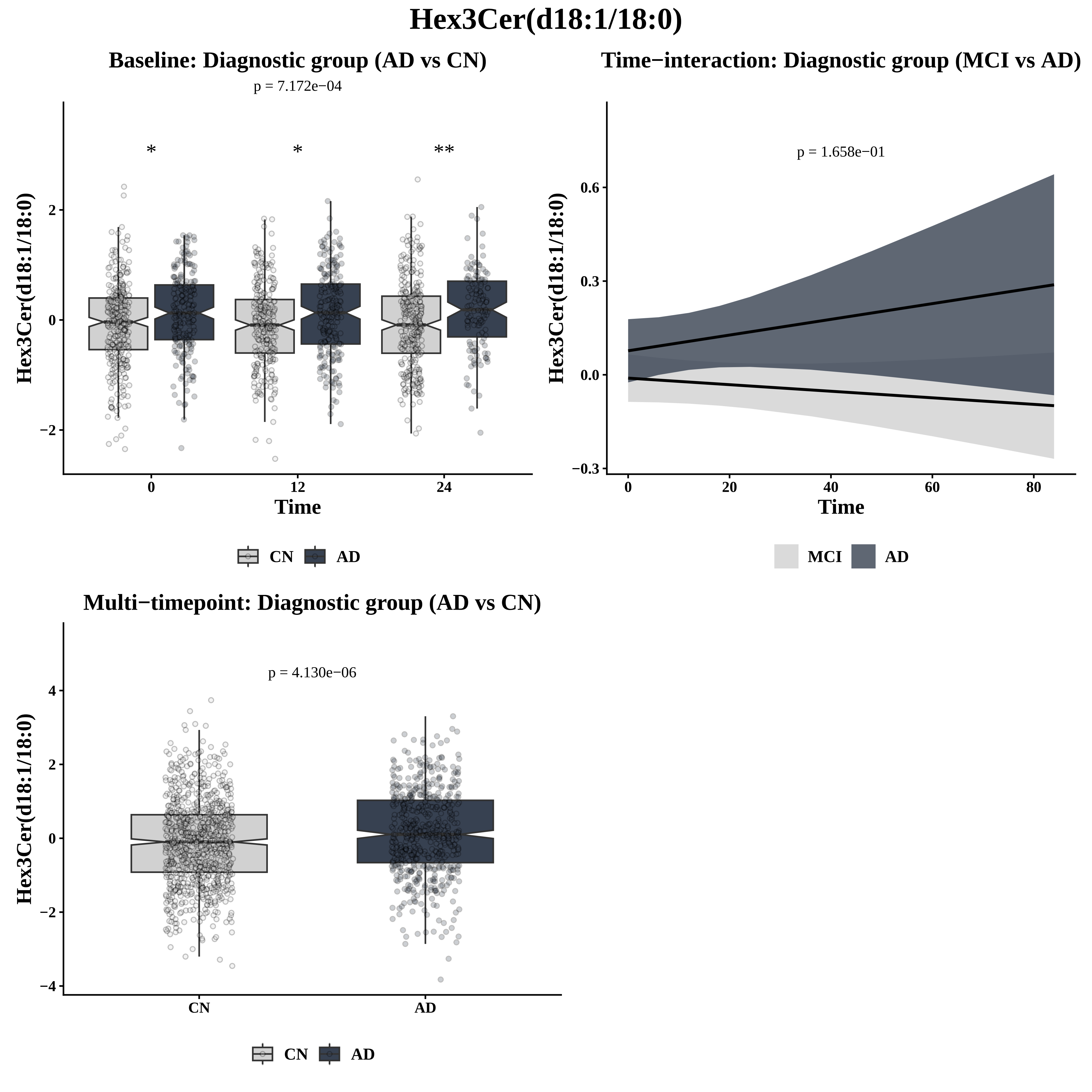

Hex3Cer(d18:1/18:0)
Baseline: Diagnostic group (AD vs CN)
p = 7.172e−04
*
*
**
Hex3Cer(d18:1/18:0)
2
0
−2
0
12
24
Time
CN
AD
Time−interaction: Diagnostic group (MCI vs AD)
p = 1.658e−01
Hex3Cer(d18:1/18:0)
0.6
0.3
0.0
−0.3
0
20
40
60
80
Time
MCI
AD
Multi−timepoint: Diagnostic group (AD vs CN)
p = 4.130e−06
Hex3Cer(d18:1/18:0)
4
2
0
−2
−4
CN
AD
CN
AD

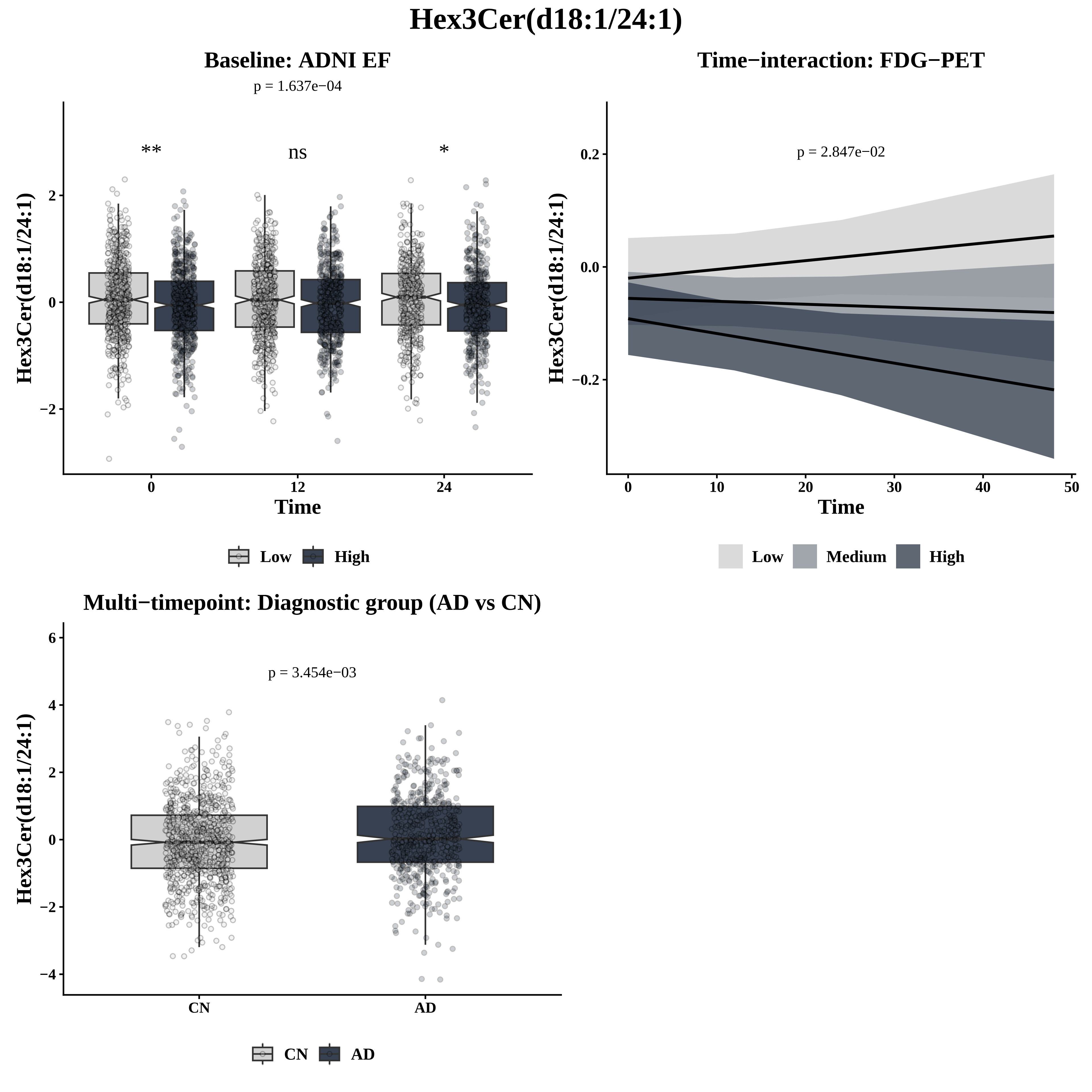

Hex3Cer(d18:1/24:1)
Baseline: ADNI EF
p = 1.637e−04
**
ns
*
Hex3Cer(d18:1/24:1)
2
0
−2
0
12
24
Time
Low
High
Time−interaction: FDG−PET
p = 2.847e−02
Hex3Cer(d18:1/24:1)
0.2
0.0
−0.2
0
10
20
30
40
50
Time
Low
Medium
High
Multi−timepoint: Diagnostic group (AD vs CN)
p = 3.454e−03
Hex3Cer(d18:1/24:1)
6
4
2
0
−2
−4
CN
AD
CN
AD

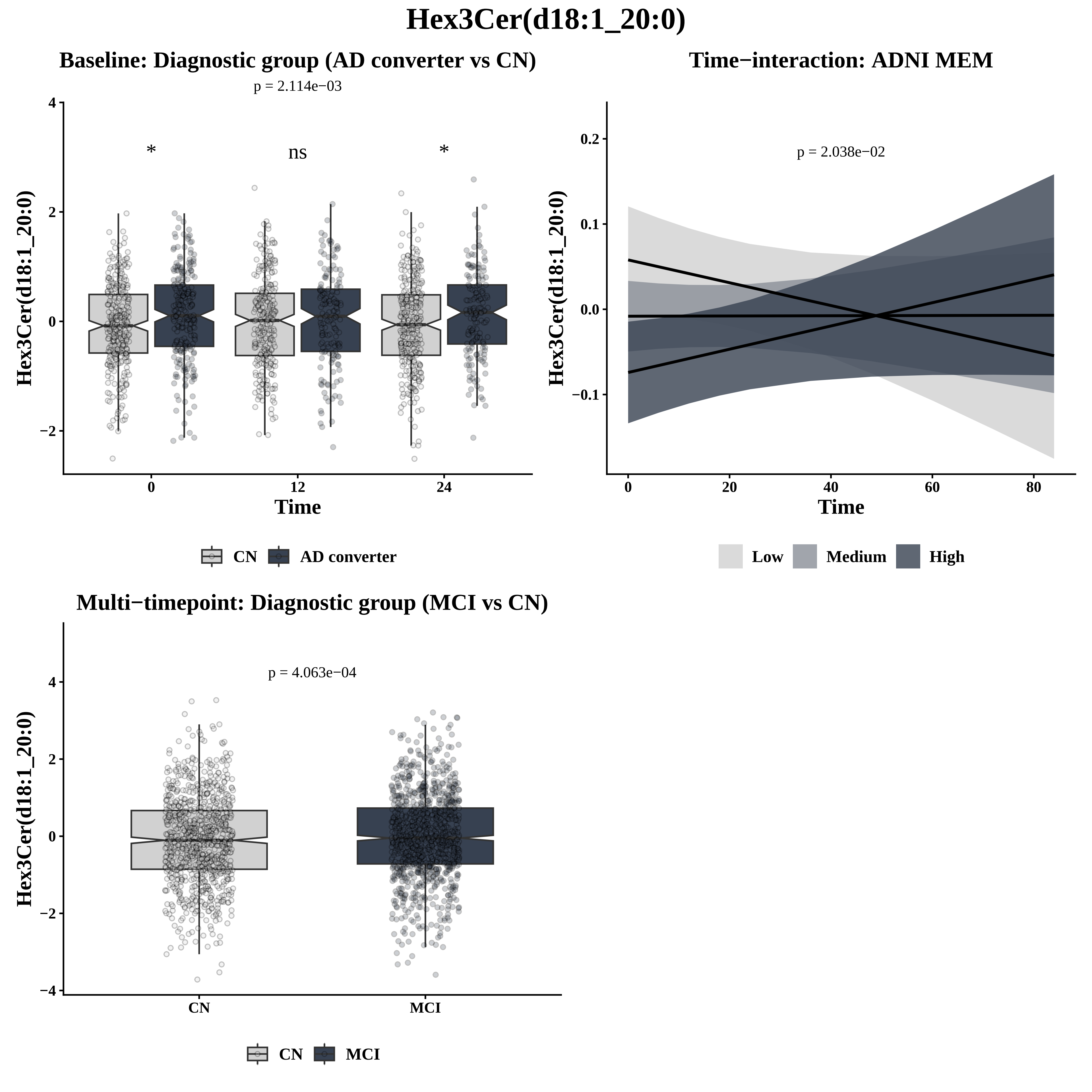

Hex3Cer(d18:1_20:0)
Baseline: Diagnostic group (AD converter vs CN)
p = 2.114e−03
*
ns
*
Hex3Cer(d18:1_20:0)
Time
CN
AD converter
Time−interaction: ADNI MEM
p = 2.038e−02
Hex3Cer(d18:1_20:0)
Time
Low
Medium
High
Multi−timepoint: Diagnostic group (MCI vs CN)
p = 4.063e−04
Hex3Cer(d18:1_20:0)
CN
MCI

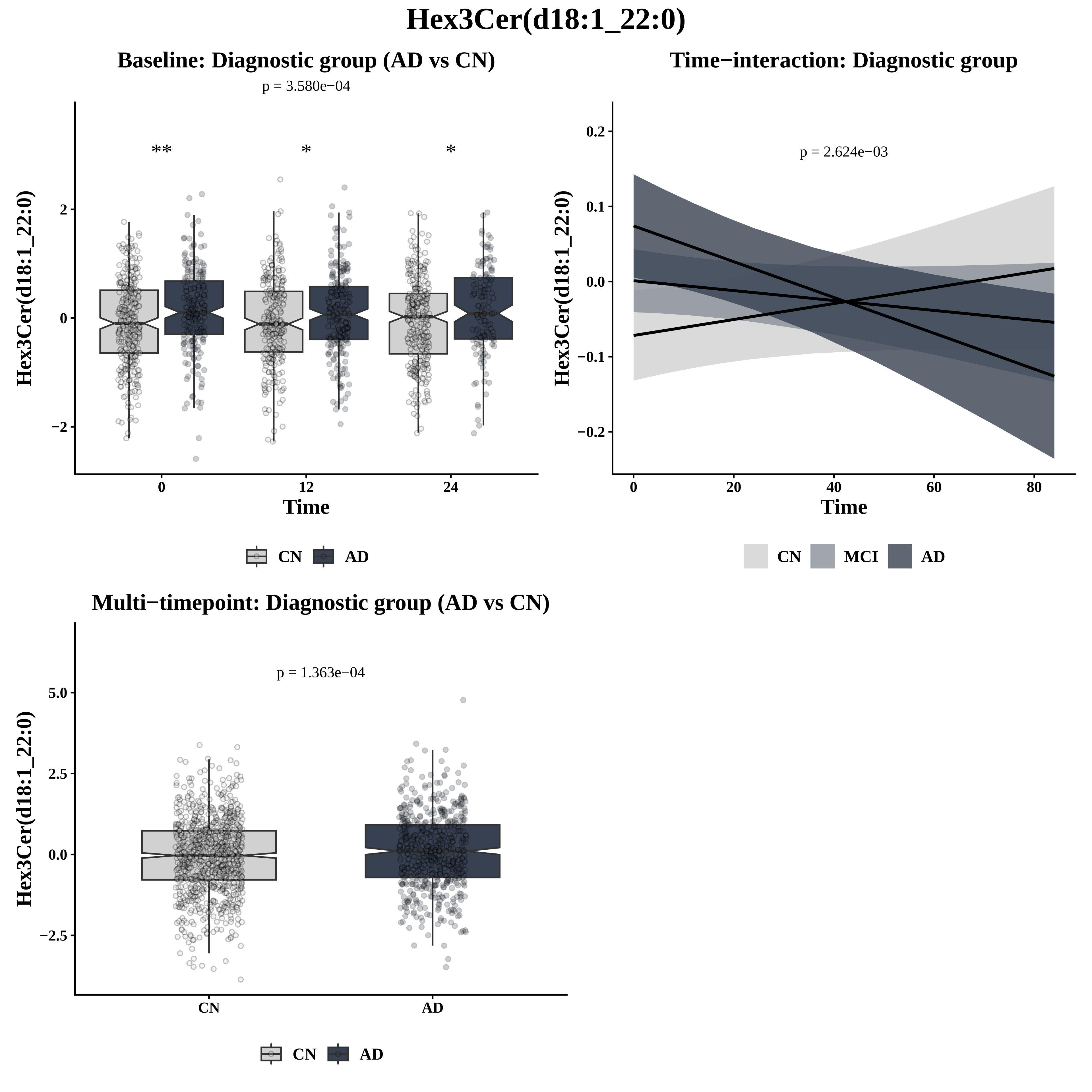

Hex3Cer(d18:1_22:0)
Baseline: Diagnostic group (AD vs CN)
p = 3.580e−04
**
*
*
Hex3Cer(d18:1_22:0)
2
0
−2
0
12
24
Time
CN
AD
Time−interaction: Diagnostic group
p = 2.624e−03
0.2
0.1
0.0
−0.1
−0.2
0
20
40
60
80
Time
CN
MCI
AD
Multi−timepoint: Diagnostic group (AD vs CN)
p = 1.363e−04
5.0
2.5
0.0
−2.5
CN
AD

### HexCer(d18:1/26:0)

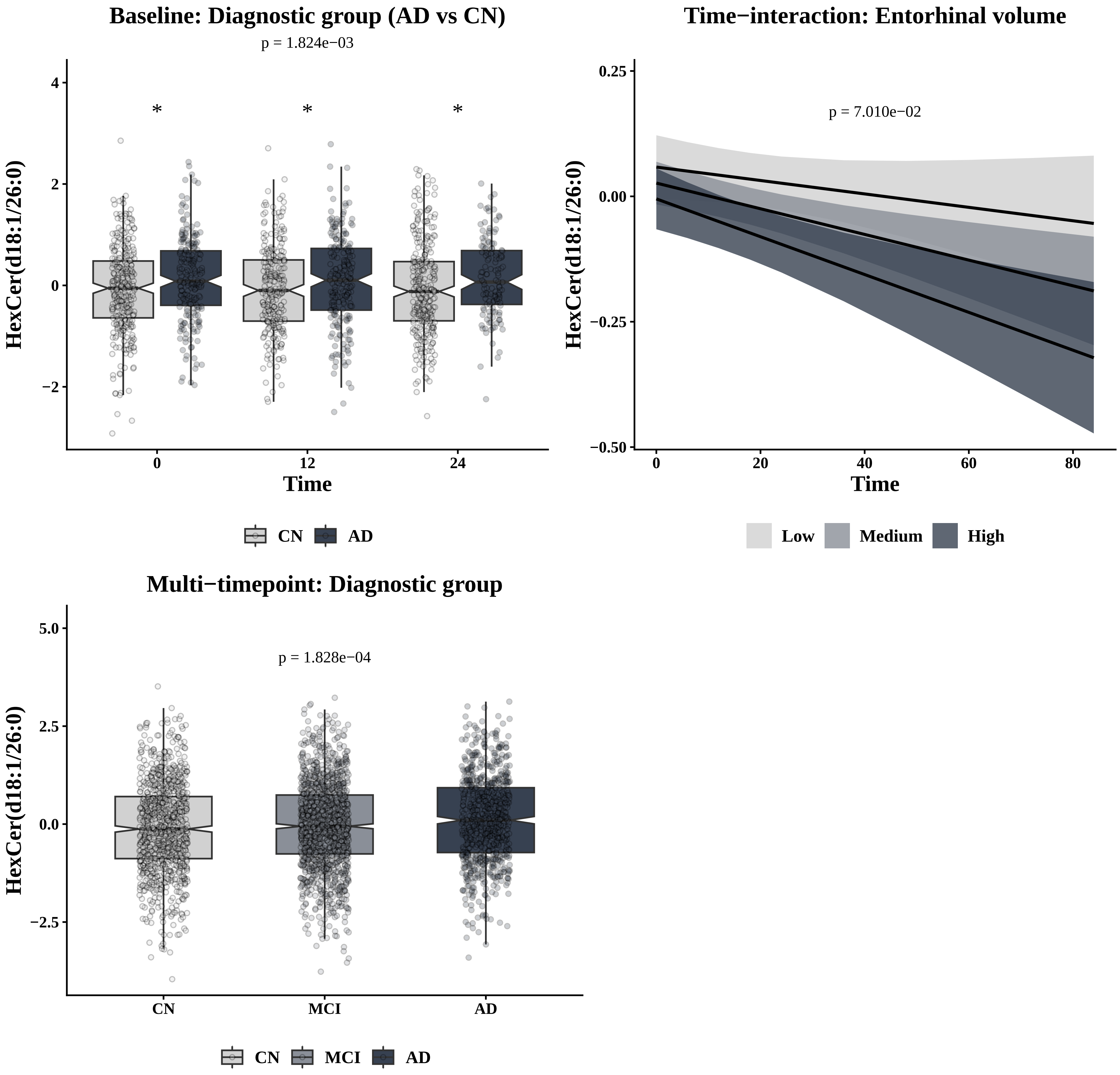

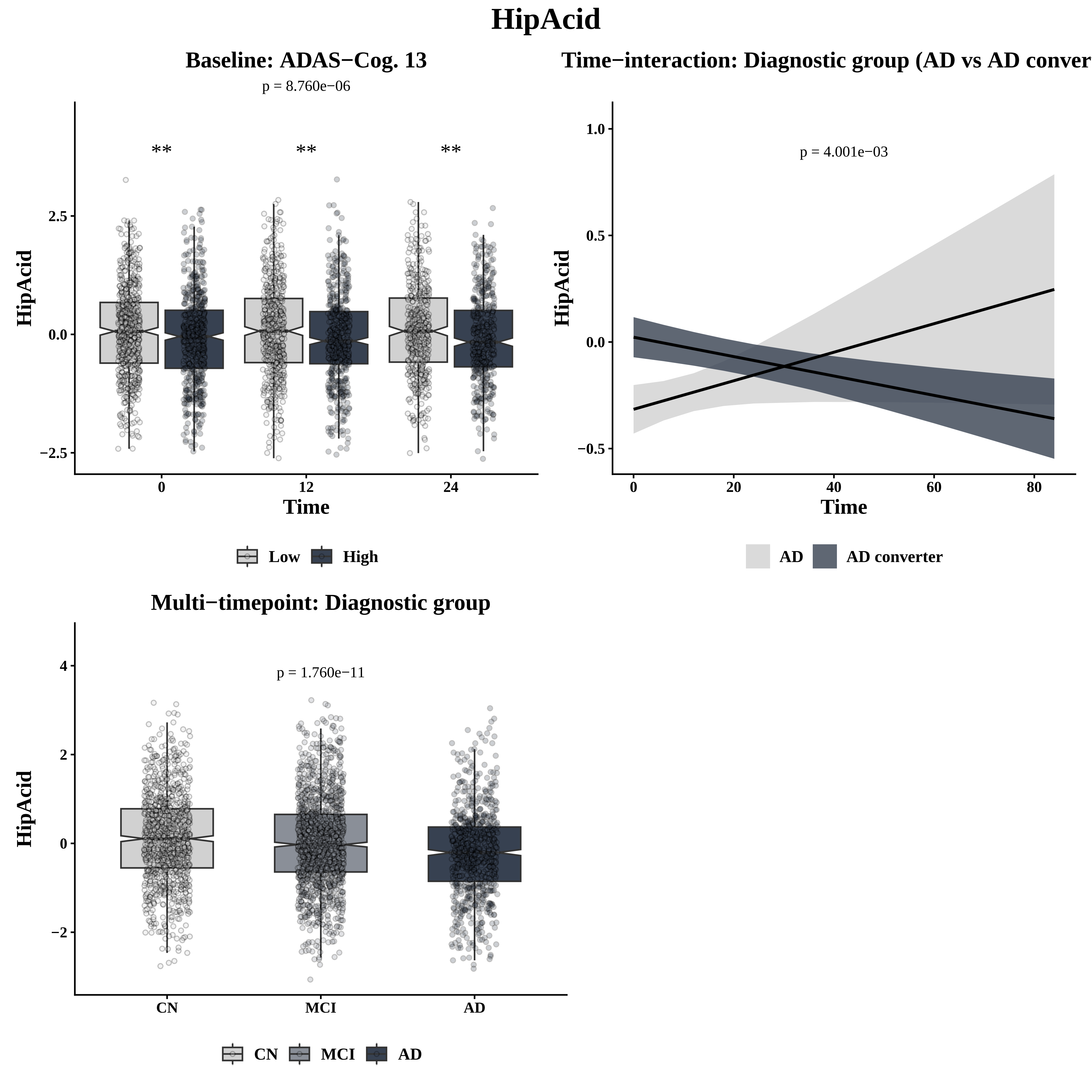

HipAcid
Baseline: ADAS−Cog. 13
p = 8.760e−06
**
**
**
2.5
0.0
−2.5
HipAcid
0
12
24
Time
Low
High
Time−interaction: Diagnostic group (AD vs AD conver
1.0
0.5
0.0
−0.5
p = 4.001e−03
HipAcid
0
20
40
60
80
Time
AD
AD converter
Multi−timepoint: Diagnostic group
4
2
0
−2
p = 1.760e−11
HipAcid
CN
MCI
AD
CN
MCI
AD

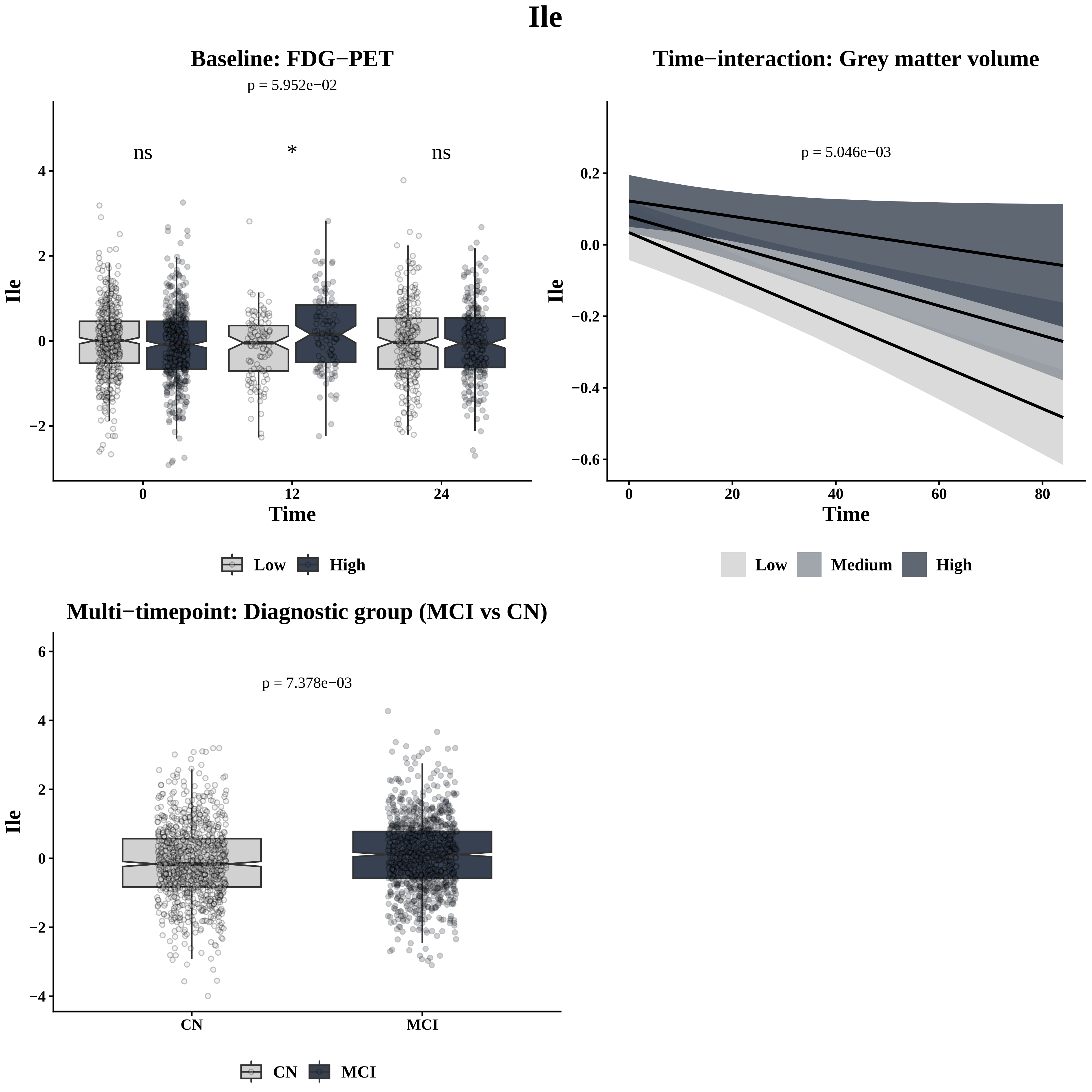
Ile
Baseline: FDG−PET
p = 5.952e−02
ns
*
ns
Ile
Time
0
12
24
4
2
0
−2
Low
High
Time−interaction: Grey matter volume
p = 5.046e−03
0.2
0.0
−0.2
−0.4
−0.6
0
20
40
60
80
Low
Medium
High
Multi−timepoint: Diagnostic group (MCI vs CN)
p = 7.378e−03
6
4
2
0
−2
−4
CN
MCI
CN
MCI

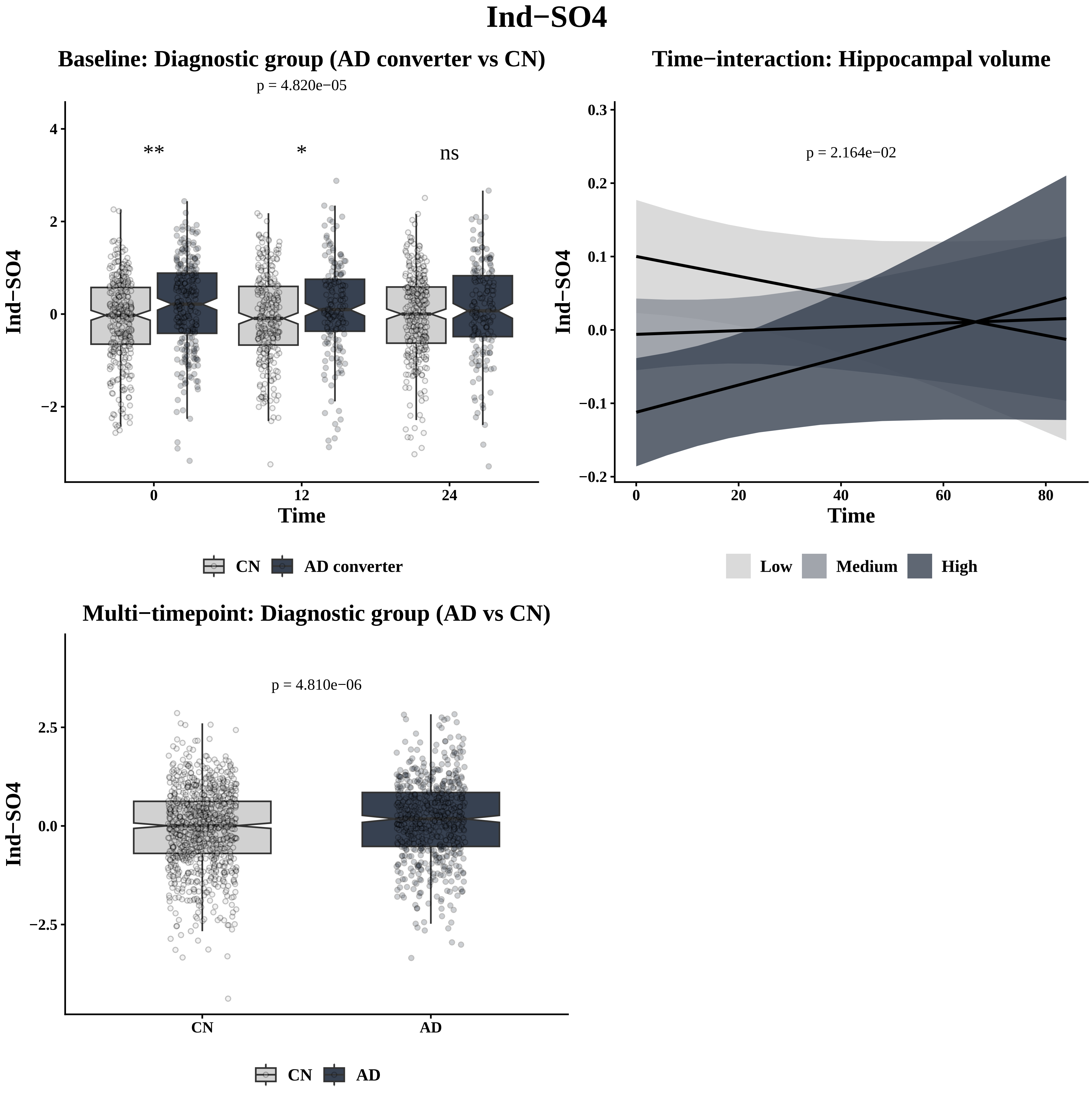
Ind−SO4
Baseline: Diagnostic group (AD converter vs CN)
p = 4.820e−05
**
*
ns
Ind−SO4
Time
0
12
24
4
2
0
−2
CN
AD converter
Time−interaction: Hippocampal volume
p = 2.164e−02
Ind−SO4
Time
0.3
0.2
0.1
0.0
−0.1
−0.2
0
20
40
60
80
Low
Medium
High
Multi−timepoint: Diagnostic group (AD vs CN)
p = 4.810e−06
Ind−SO4
2.5
0.0
−2.5
CN
AD

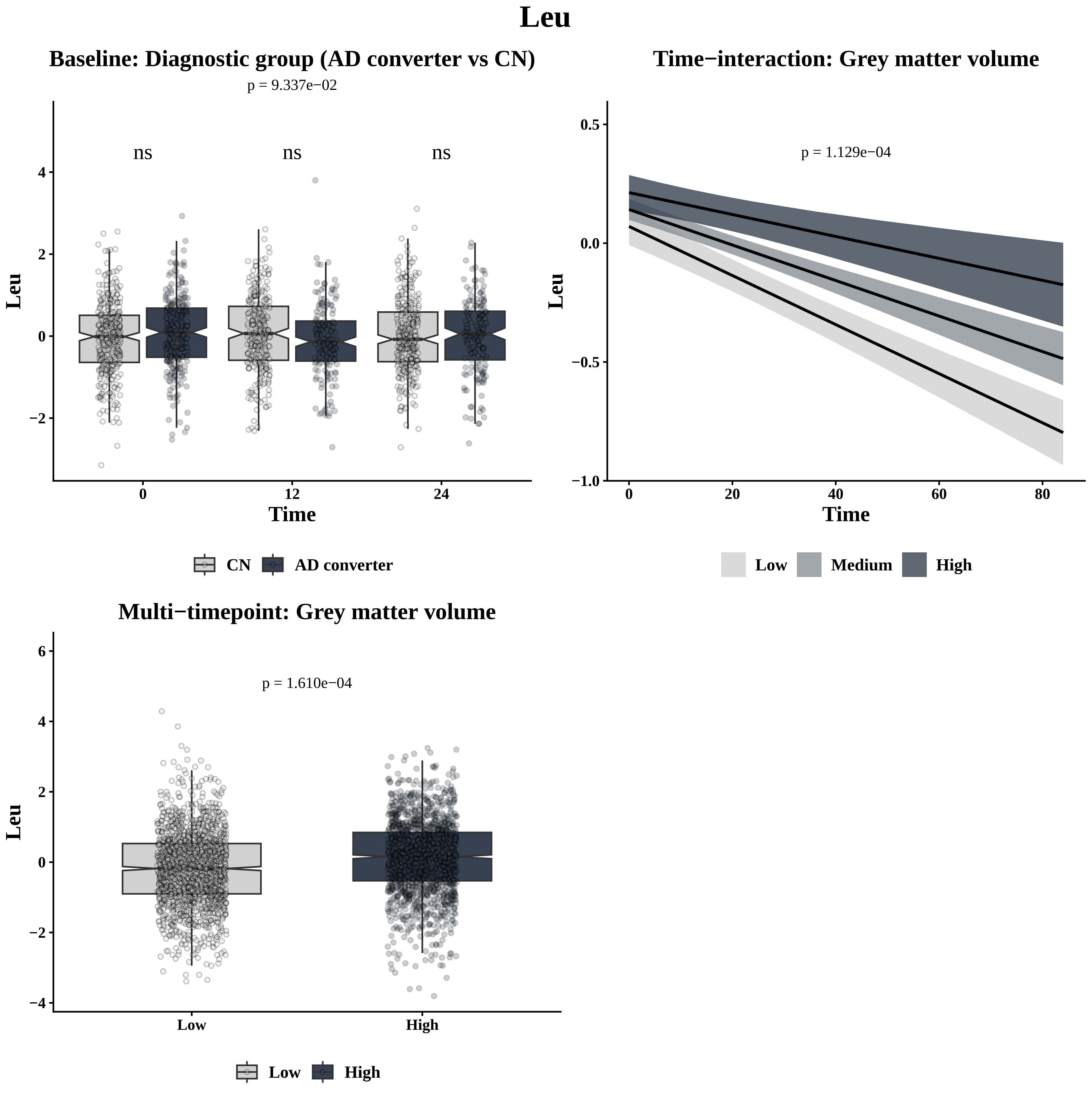
Leu
Baseline: Diagnostic group (AD converter vs CN)
p = 9.337e−02
ns
ns
ns
Leu
4
2
0
−2
0
12
24
Time
CN
AD converter
Time−interaction: Grey matter volume
p = 1.129e−04
Leu
0.5
0.0
−0.5
−1.0
0
20
40
60
80
Time
Low
Medium
High
Multi−timepoint: Grey matter volume
p = 1.610e−04
Leu
6
4
2
0
−2
−4
Low
High
Low
High

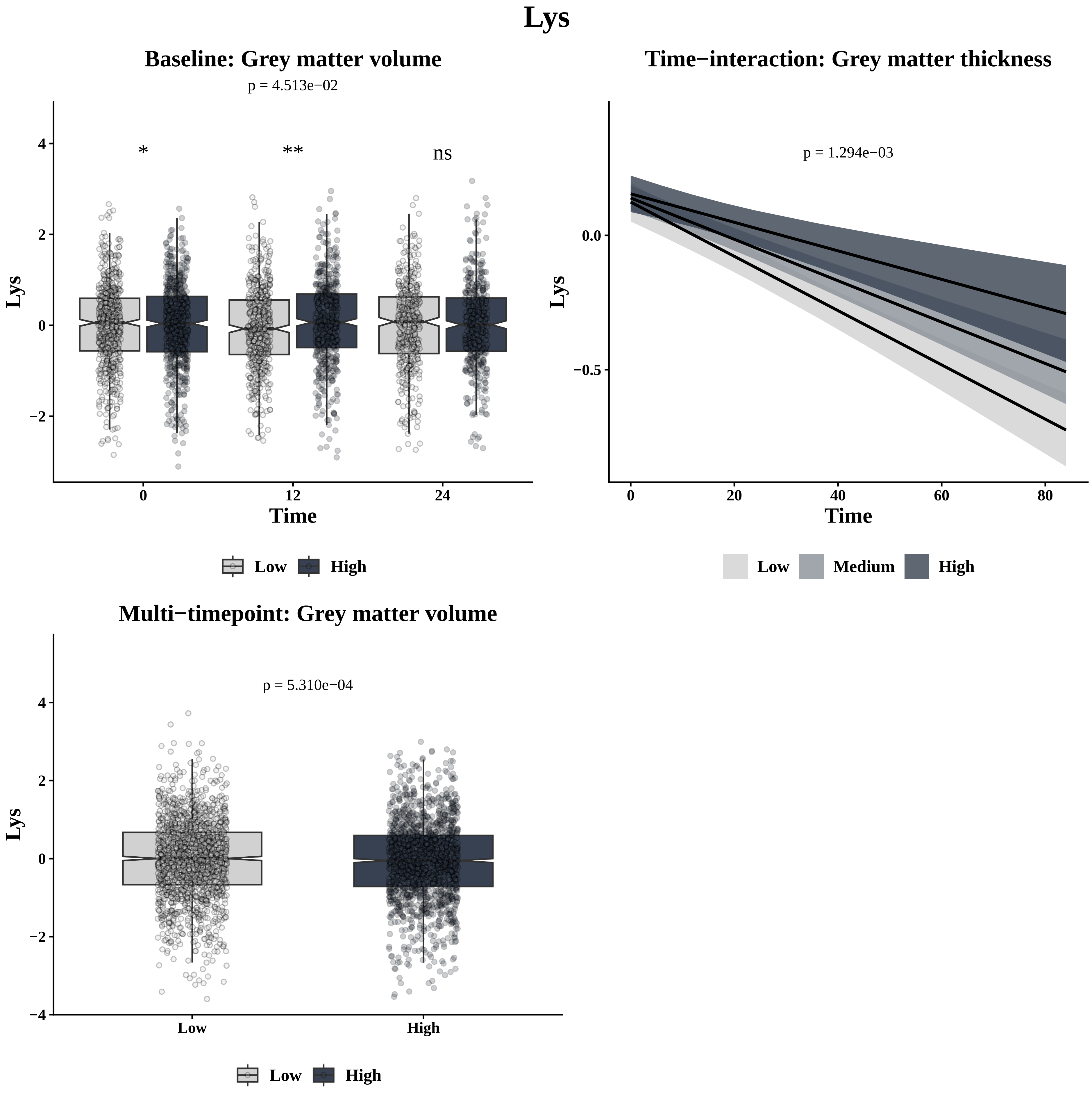
Lys
Baseline: Grey matter volume
p = 4.513e−02
*
**
ns
Lys
Time
0
12
24
4
2
0
−2
Low
High
Time−interaction: Grey matter thickness
p = 1.294e−03
0.0
−0.5
Lys
Time
0
20
40
60
80
Low
Medium
High
Multi−timepoint: Grey matter volume
p = 5.310e−04
Lys
4
2
0
−2
−4
Low
High
Low
High

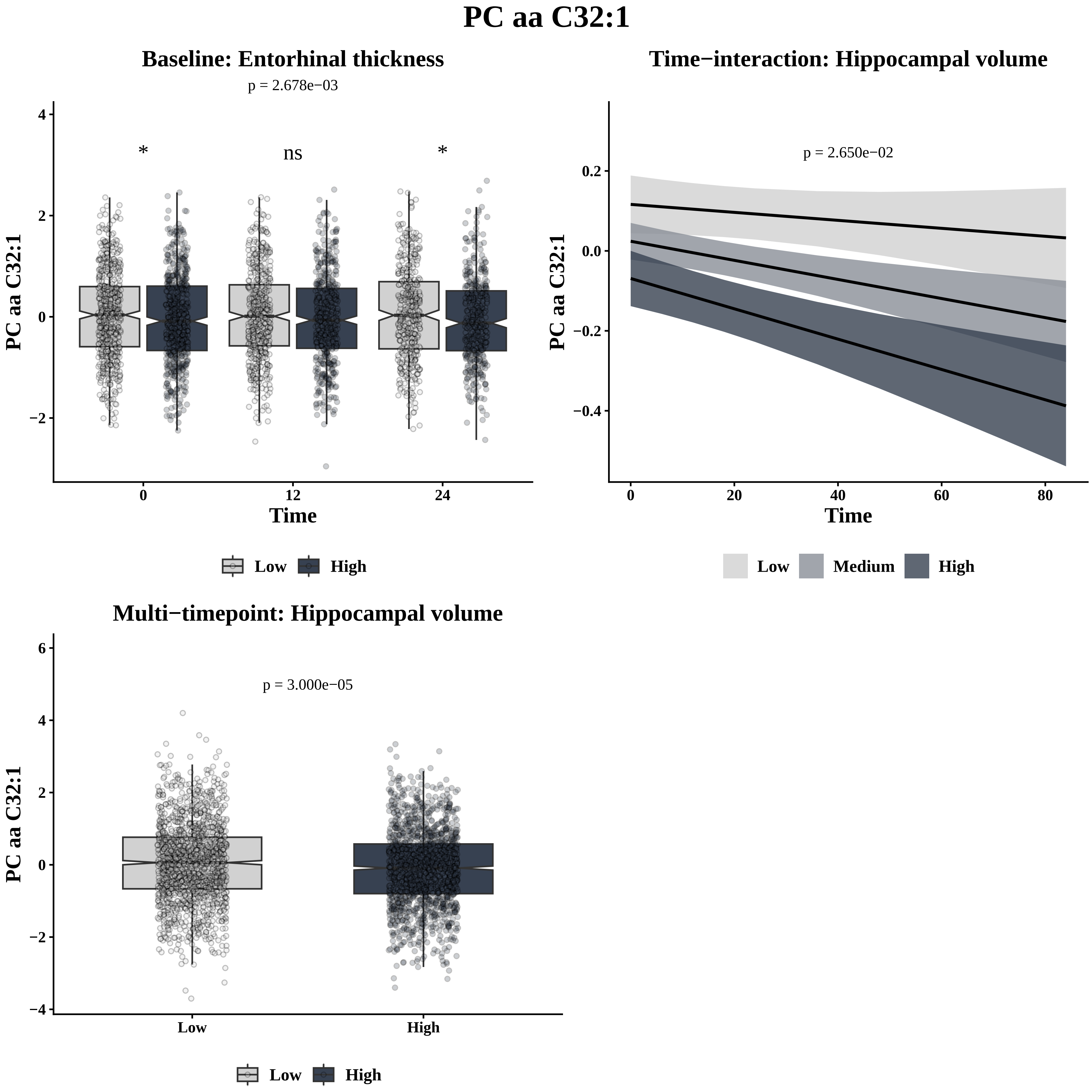
PC aa C32:1
Baseline: Entorhinal thickness
p = 2.678e−03
*
ns
*
PC aa C32:1
Time
0
12
24
4
2
0
−2
Low
High
Time−interaction: Hippocampal volume
p = 2.650e−02
PC aa C32:1
Time
0.2
0.0
−0.2
−0.4
0
20
40
60
80
Low
Medium
High
Multi−timepoint: Hippocampal volume
p = 3.000e−05
PC aa C32:1
6
4
2
0
−2
−4
Low
High
Low
High

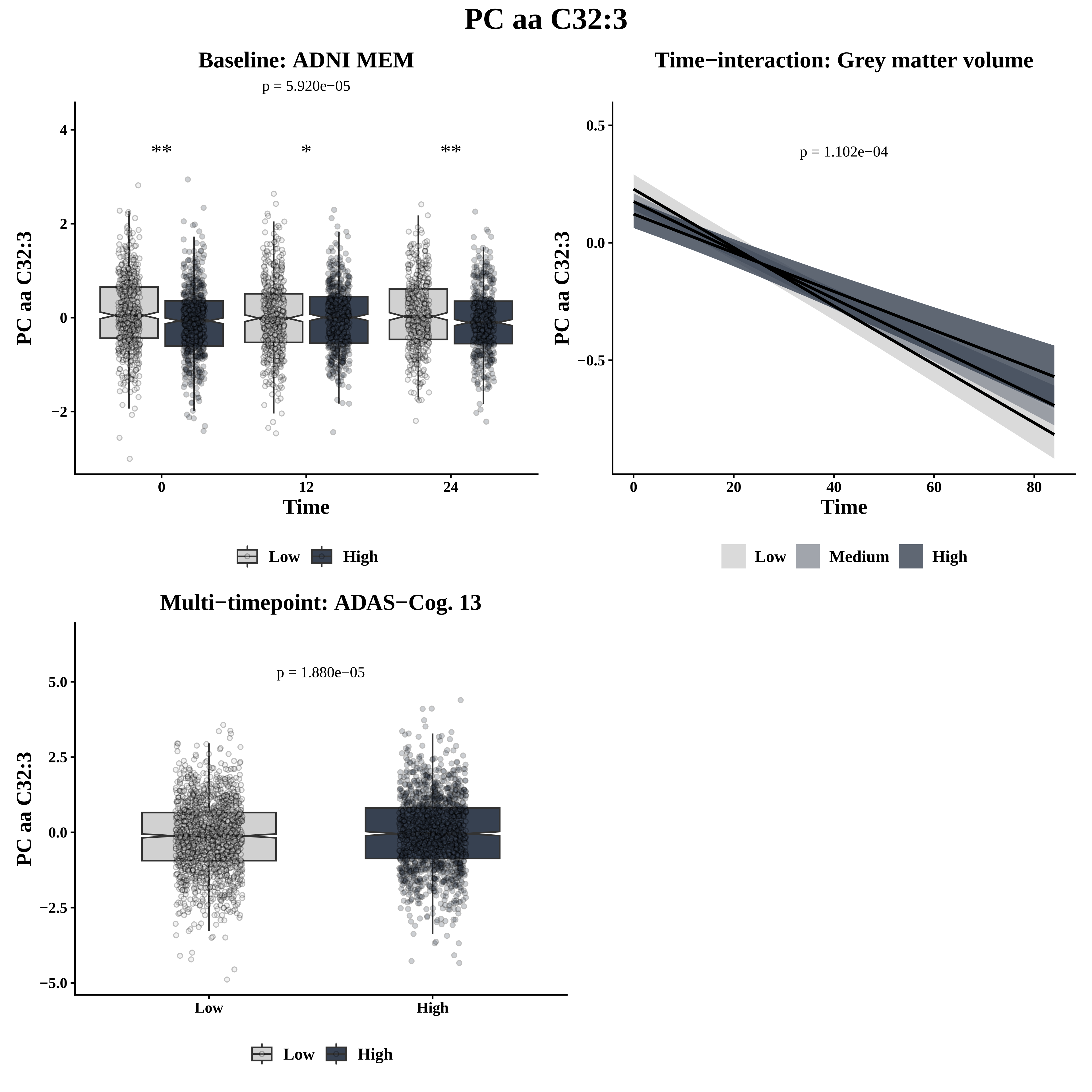
PC aa C32:3
Baseline: ADNI MEM
p = 5.920e−05
**
*
**
PC aa C32:3
Time
0
12
24
4
2
0
−2
Low
High
Time−interaction: Grey matter volume
p = 1.102e−04
PC aa C32:3
Time
0.5
0.0
−0.5
0
20
40
60
80
Low
Medium
High
Multi−timepoint: ADAS−Cog. 13
p = 1.880e−05
PC aa C32:3
5.0
2.5
0.0
−2.5
−5.0
Low
High
Low
High

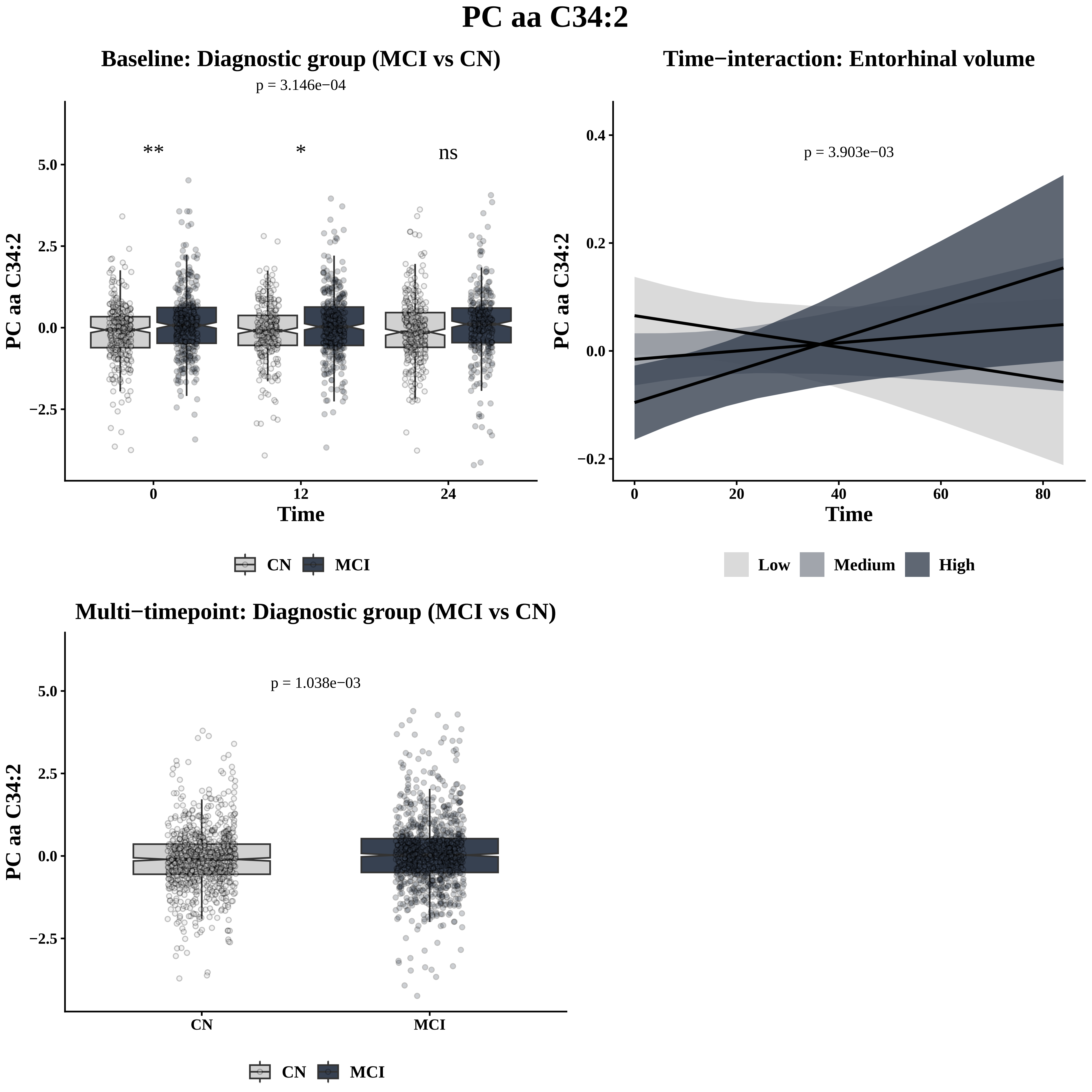
PC aa C34:2
Baseline: Diagnostic group (MCI vs CN)
p = 3.146e−04
**
*
ns
PC aa C34:2
5.0
2.5
0.0
−2.5
0
12
24
Time
CN
MCI
Time−interaction: Entorhinal volume
p = 3.903e−03
PC aa C34:2
0.4
0.2
0.0
−0.2
0
20
40
60
80
Time
Low
Medium
High
Multi−timepoint: Diagnostic group (MCI vs CN)
p = 1.038e−03
PC aa C34:2
5.0
2.5
0.0
−2.5
CN
MCI
CN
MCI

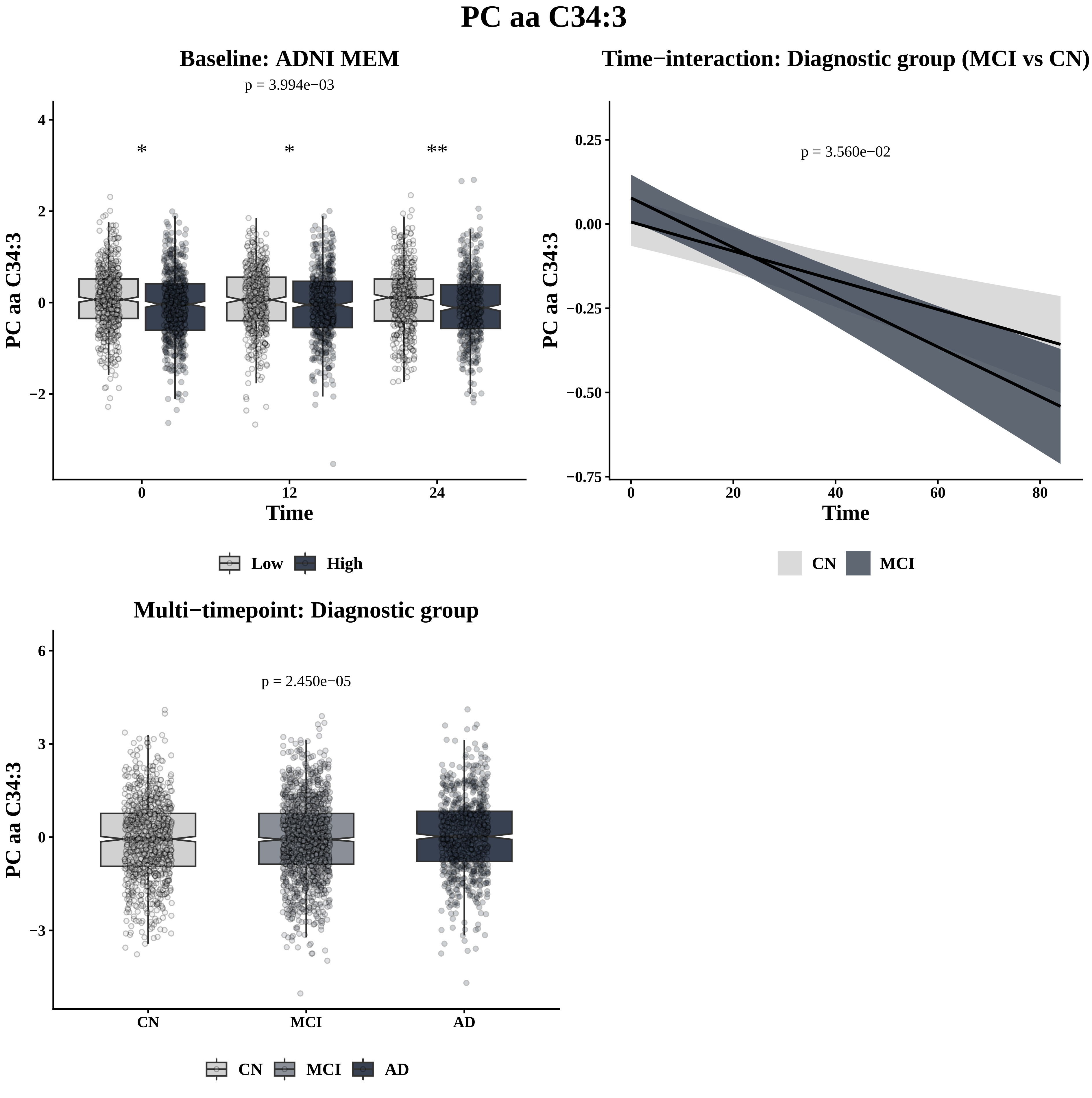
PC aa C34:3
Baseline: ADNI MEM
p = 3.994e−03
*
*
**
PC aa C34:3
4
2
0
−2
0
12
24
Time
Low
High
Time−interaction: Diagnostic group (MCI vs CN)
p = 3.560e−02
PC aa C34:3
0.25
0.00
−0.25
−0.50
−0.75
0
20
40
60
80
Time
CN
MCI
Multi−timepoint: Diagnostic group
p = 2.450e−05
PC aa C34:3
6
3
0
−3
CN
MCI
AD
CN
MCI
AD

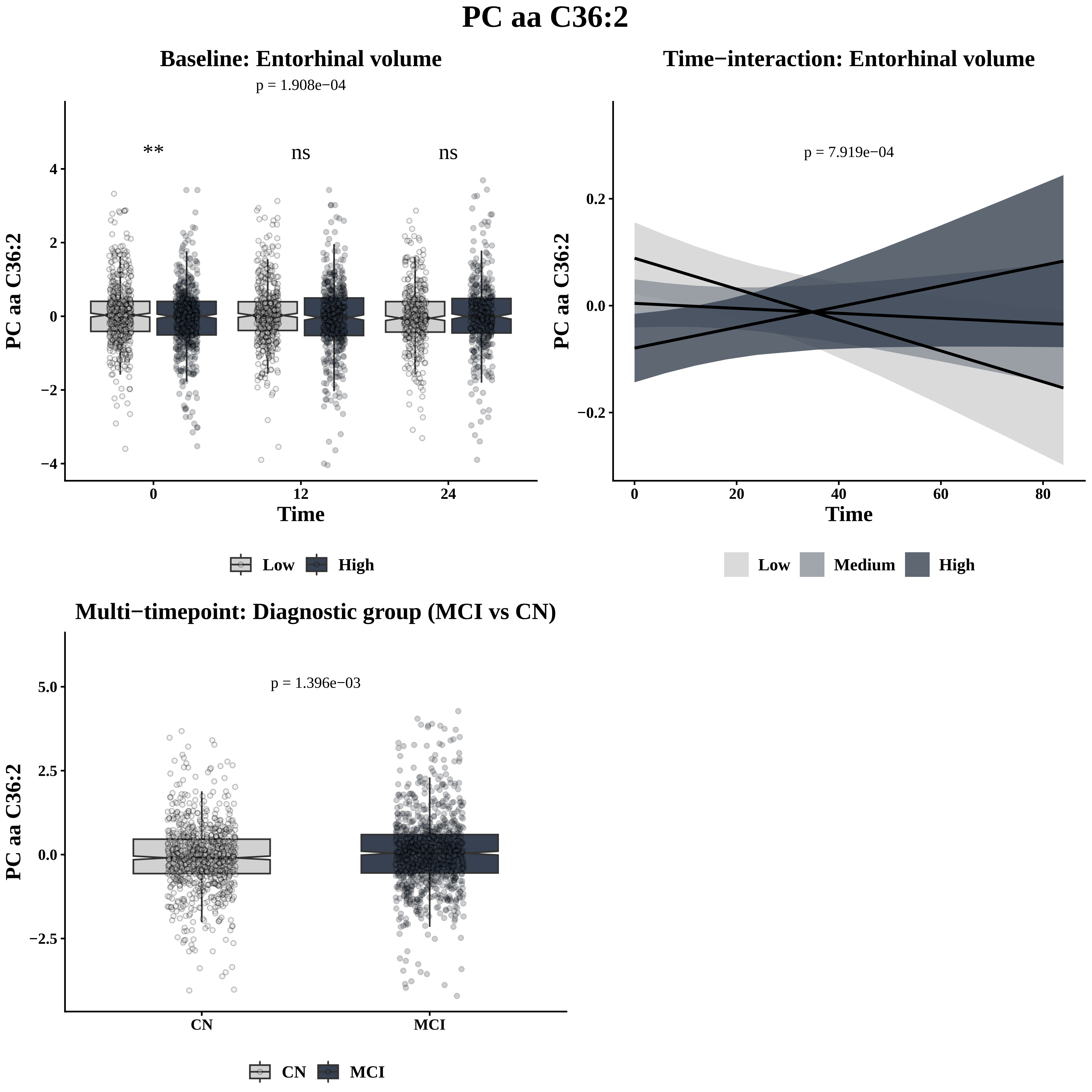

PC aa C36:2
Baseline: Entorhinal volume
p = 1.908e−04
**
ns
ns
PC aa C36:2
Time
0
12
24
4
2
0
−2
−4
Low
High
Time−interaction: Entorhinal volume
p = 7.919e−04
0.2
0.0
−0.2
0
20
40
60
80
Low
Medium
High
Multi−timepoint: Diagnostic group (MCI vs CN)
p = 1.396e−03
5.0
2.5
0.0
−2.5
CN
MCI

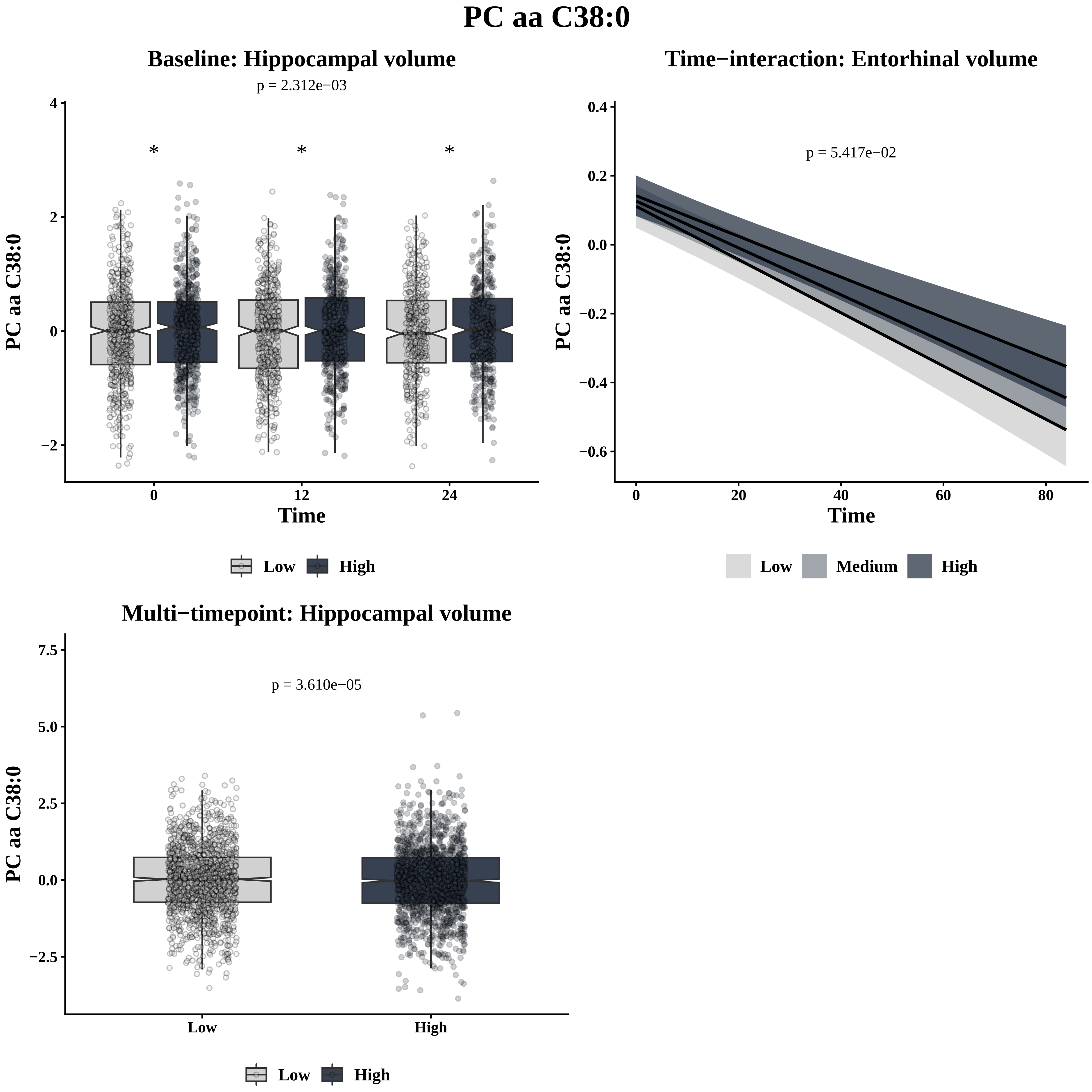
PC aa C38:0
Baseline: Hippocampal volume
p = 2.312e−03
*
*
*
PC aa C38:0
Time
0
12
24
Low
High
Time−interaction: Entorhinal volume
p = 5.417e−02
Time
Low
Medium
High
Multi−timepoint: Hippocampal volume
p = 3.610e−05
Low
High

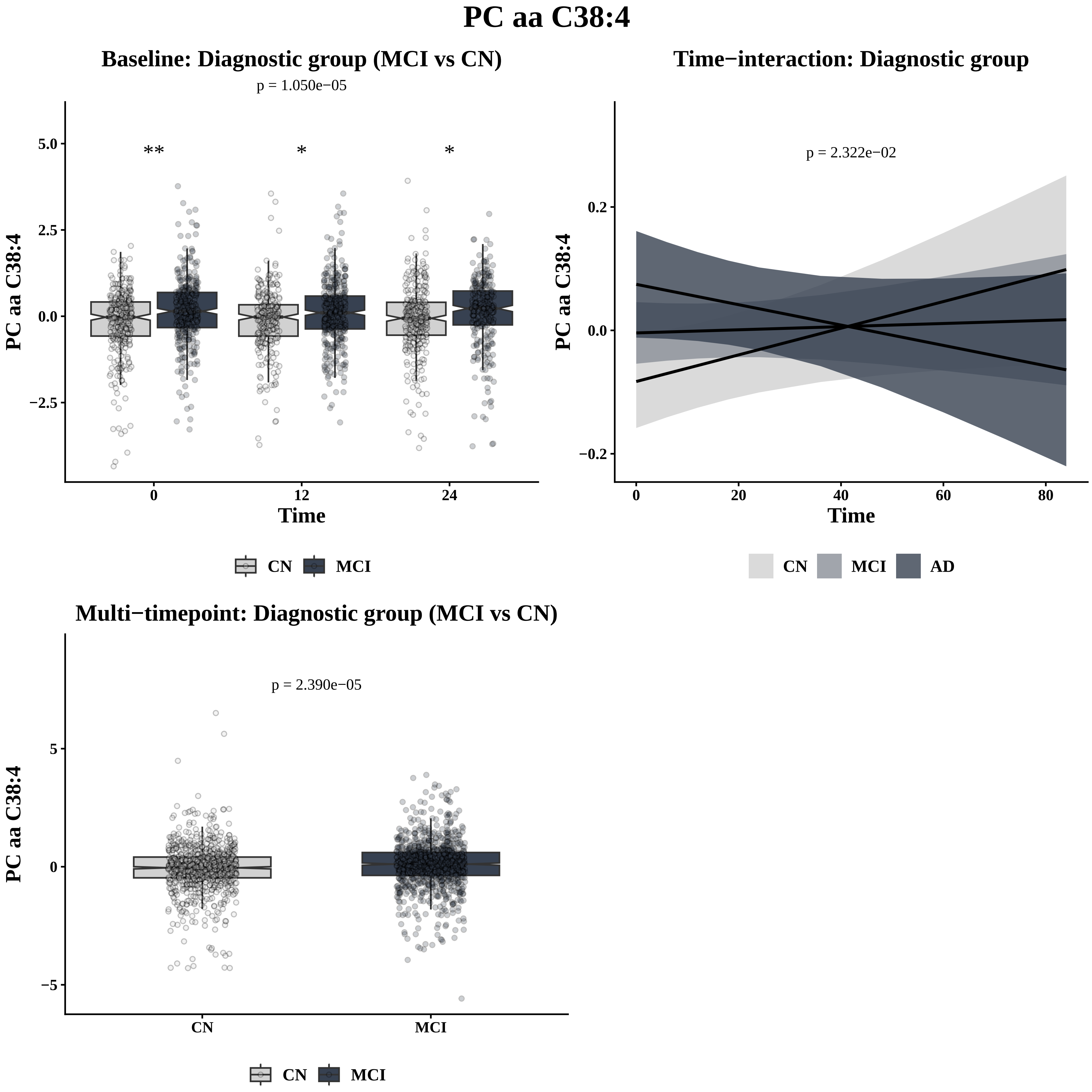
PC aa C38:4
Baseline: Diagnostic group (MCI vs CN)
p = 1.050e−05
**
*
*
5.0
2.5
0.0
−2.5
PC aa C38:4
0
12
24
Time
CN
MCI
Time−interaction: Diagnostic group
p = 2.322e−02
0.2
0.0
−0.2
PC aa C38:4
0
20
40
60
80
Time
CN
MCI
AD
Multi−timepoint: Diagnostic group (MCI vs CN)
p = 2.390e−05
5
0
−5
PC aa C38:4
CN
MCI
CN
MCI

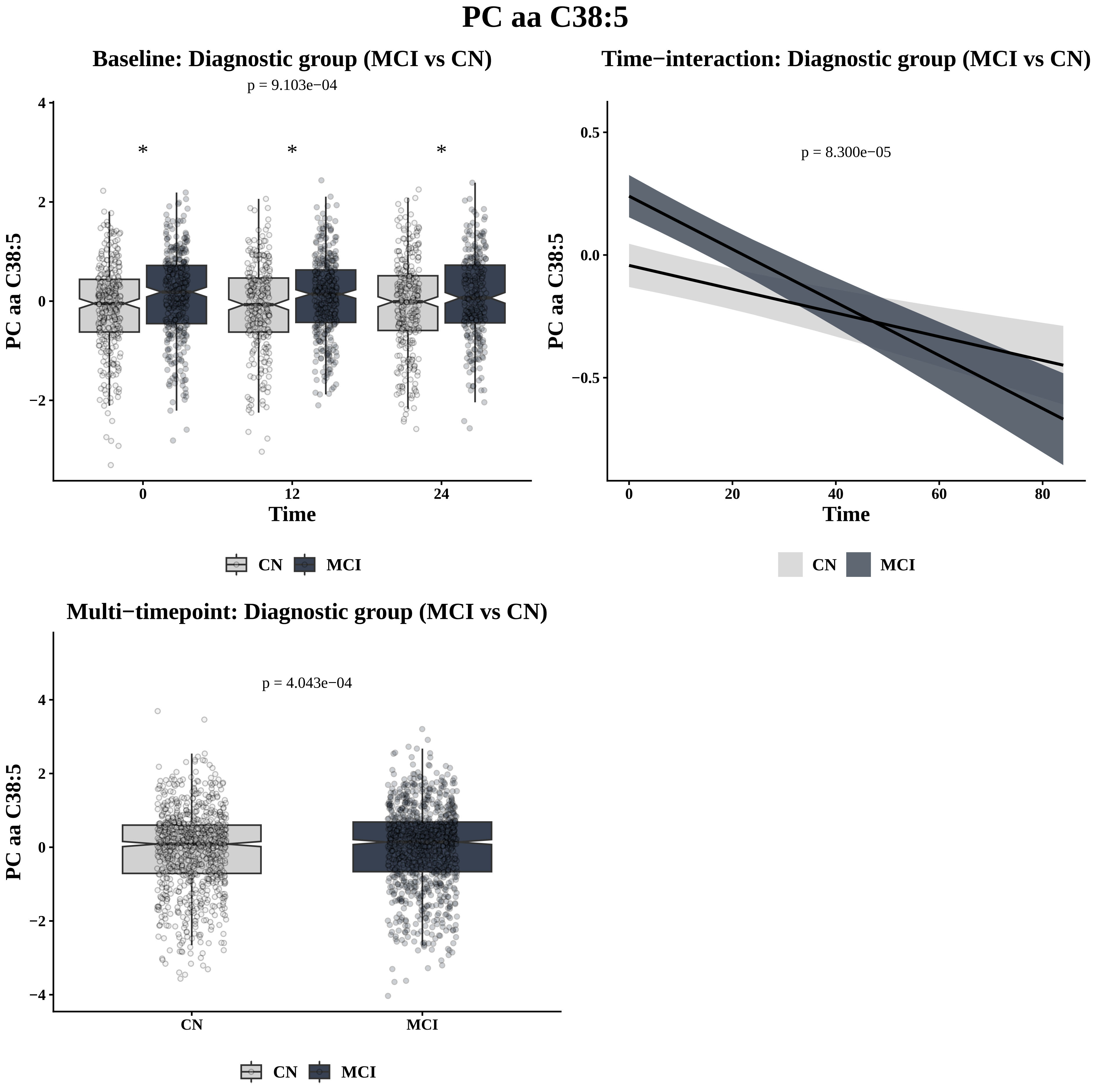
PC aa C38:5
Baseline: Diagnostic group (MCI vs CN)
p = 9.103e−04
*
*
*
PC aa C38:5
4
2
0
−2
0
12
24
Time
CN
MCI
Time−interaction: Diagnostic group (MCI vs CN)
p = 8.300e−05
PC aa C38:5
0.5
0.0
−0.5
0
20
40
60
80
Time
CN
MCI
Multi−timepoint: Diagnostic group (MCI vs CN)
p = 4.043e−04
PC aa C38:5
4
2
0
−2
−4
CN
MCI
CN
MCI

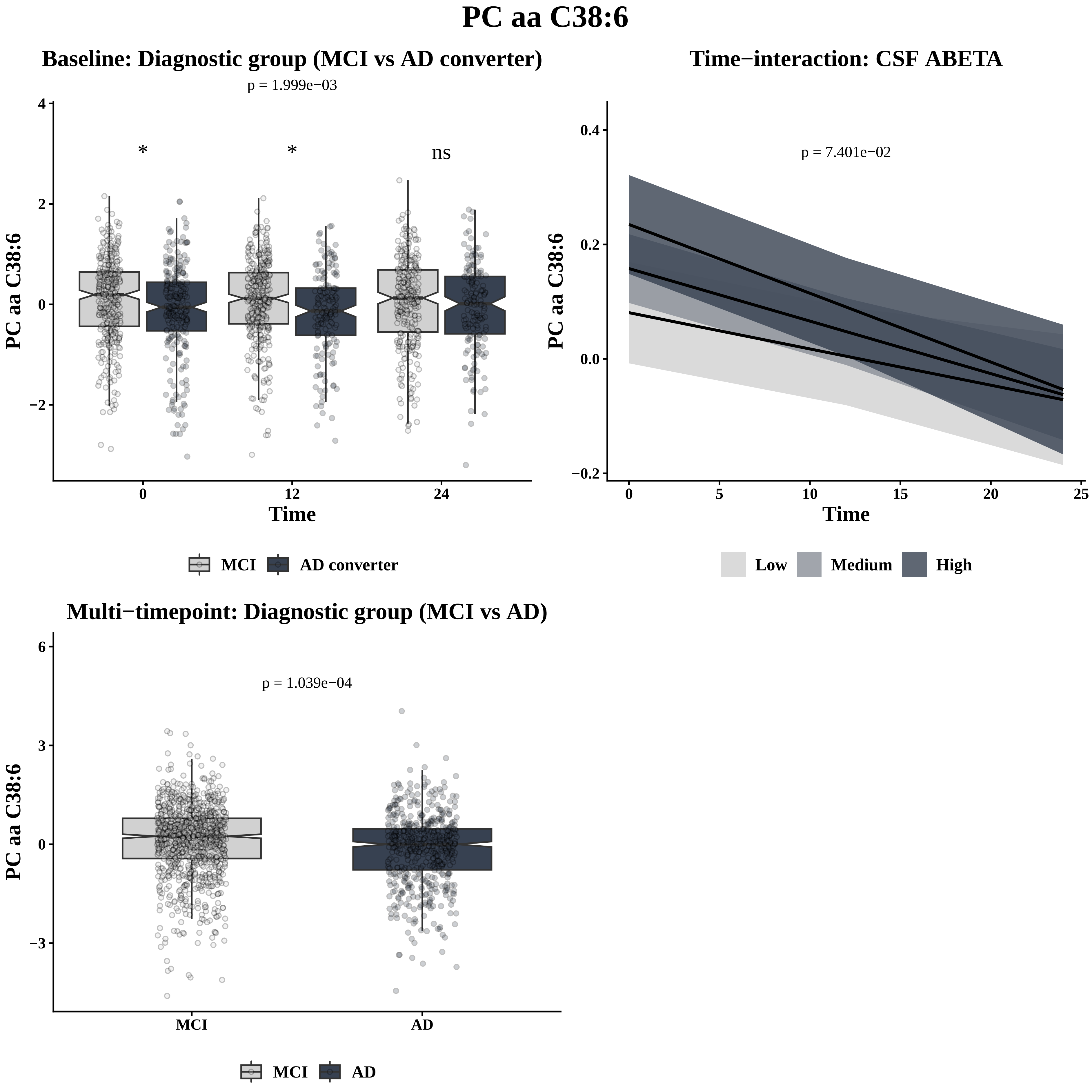
PC aa C38:6
Baseline: Diagnostic group (MCI vs AD converter)
p = 1.999e−03
*
*
ns
PC aa C38:6
Time
MCI
AD converter
Time−interaction: CSF ABETA
p = 7.401e−02
PC aa C38:6
Time
Low
Medium
High
Multi−timepoint: Diagnostic group (MCI vs AD)
p = 1.039e−04
PC aa C38:6
MCI
AD
MCI
AD

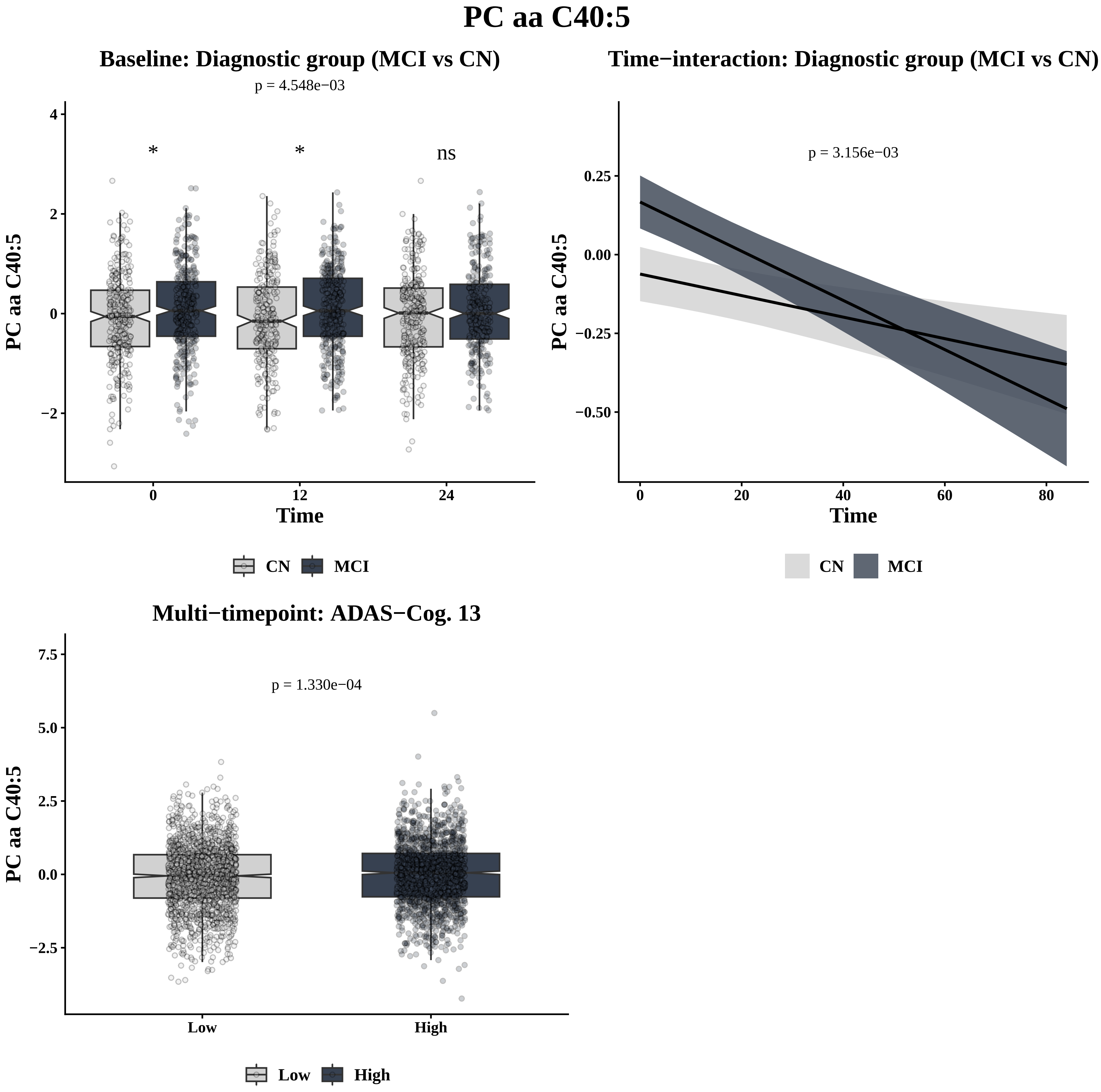
PC aa C40:5
Baseline: Diagnostic group (MCI vs CN)
p = 4.548e−03
*
*
ns
PC aa C40:5
4
2
0
−2
0
12
24
Time
CN
MCI
Time−interaction: Diagnostic group (MCI vs CN)
p = 3.156e−03
PC aa C40:5
0.25
0.00
−0.25
−0.50
0
20
40
60
80
Time
CN
MCI
Multi−timepoint: ADAS−Cog. 13
p = 1.330e−04
PC aa C40:5
7.5
5.0
2.5
0.0
−2.5
Low
High
Low
High

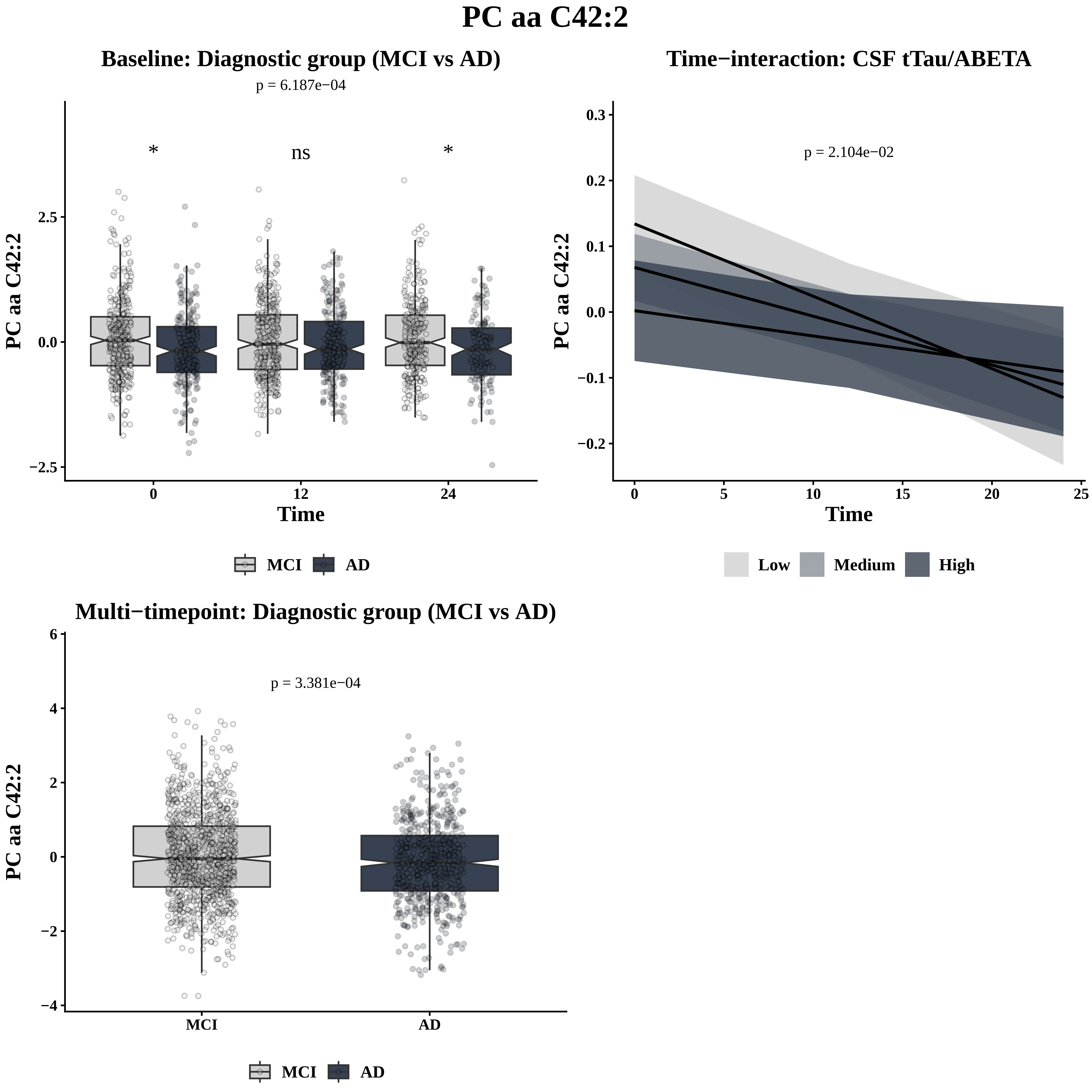
PC aa C42:2
Baseline: Diagnostic group (MCI vs AD)
p = 6.187e−04
*
ns
*
PC aa C42:2
2.5
0.0
−2.5
0
12
24
Time
MCI
AD
Time−interaction: CSF tTau/ABETA
p = 2.104e−02
PC aa C42:2
0.3
0.2
0.1
0.0
−0.1
−0.2
0
5
10
15
20
25
Time
Low
Medium
High
Multi−timepoint: Diagnostic group (MCI vs AD)
p = 3.381e−04
PC aa C42:2
6
4
2
0
−2
−4
MCI
AD
MCI
AD

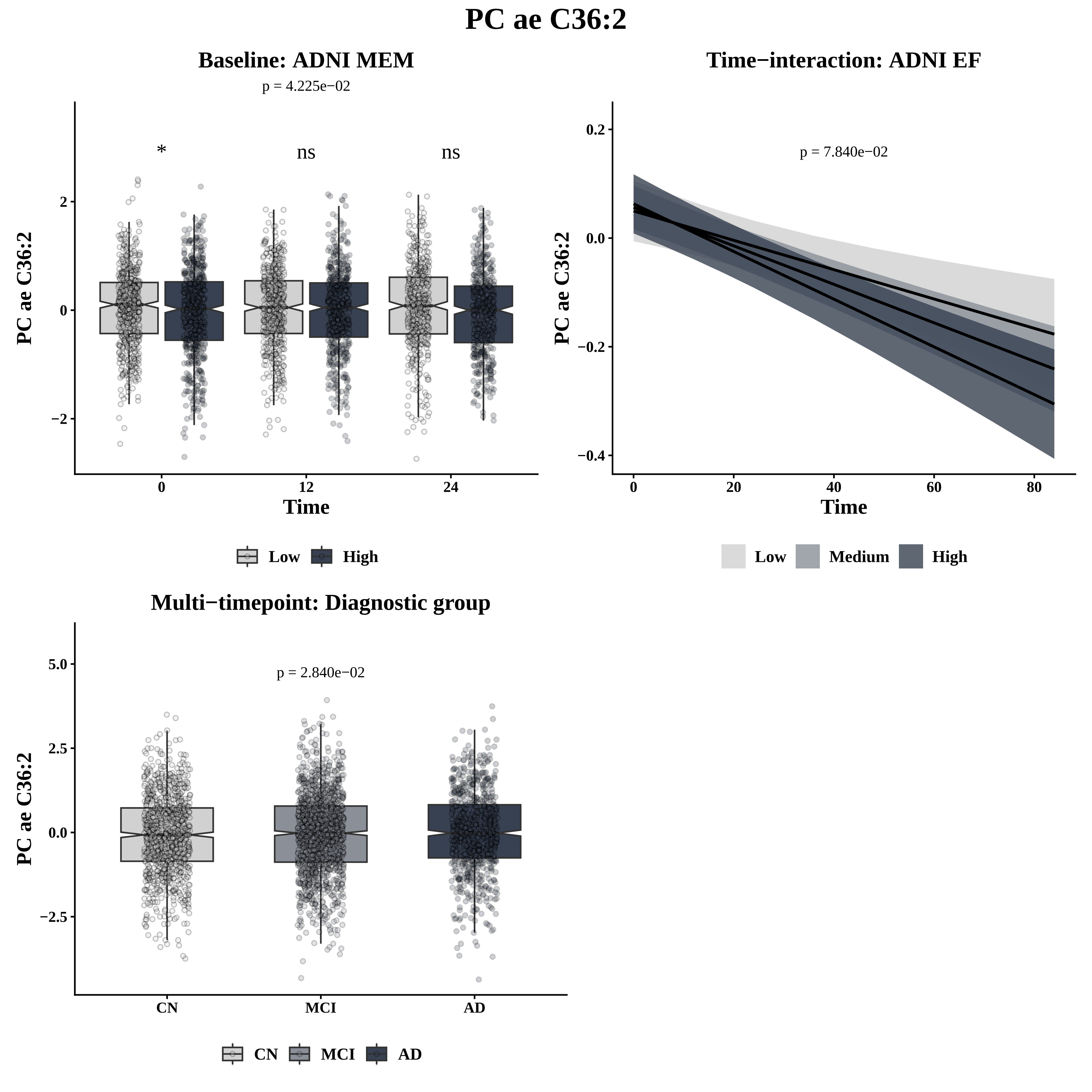

PC ae C36:2
Baseline: ADNI MEM
p = 4.225e−02
*
ns
ns
2
0
−2
PC ae C36:2
0
12
24
Time
Low
High
Time−interaction: ADNI EF
p = 7.840e−02
0.2
0.0
−0.2
−0.4
PC ae C36:2
0
20
40
60
80
Time
Low
Medium
High
Multi−timepoint: Diagnostic group
p = 2.840e−02
5.0
2.5
0.0
−2.5
PC ae C36:2
CN
MCI
AD
CN
MCI
AD

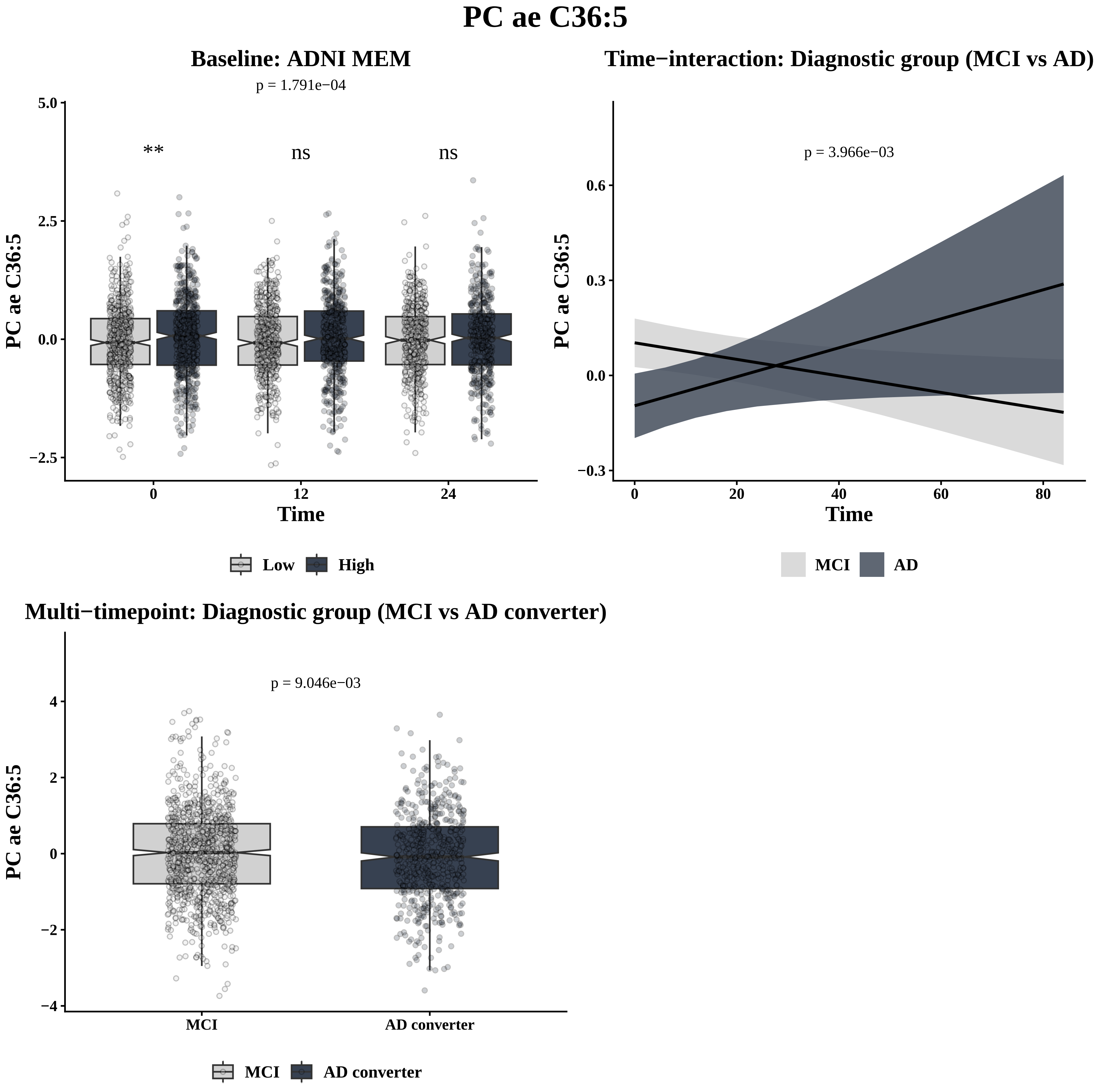
PC ae C36:5
Baseline: ADNI MEM
p = 1.791e−04
**
ns
ns
5.0
2.5
0.0
−2.5
PC ae C36:5
0
12
24
Time
Low
High
Time−interaction: Diagnostic group (MCI vs AD)
p = 3.966e−03
0.6
0.3
0.0
−0.3
PC ae C36:5
0
20
40
60
80
Time
MCI
AD
Multi−timepoint: Diagnostic group (MCI vs AD converter)
p = 9.046e−03
4
2
0
−2
−4
PC ae C36:5
MCI
AD converter
MCI
AD converter

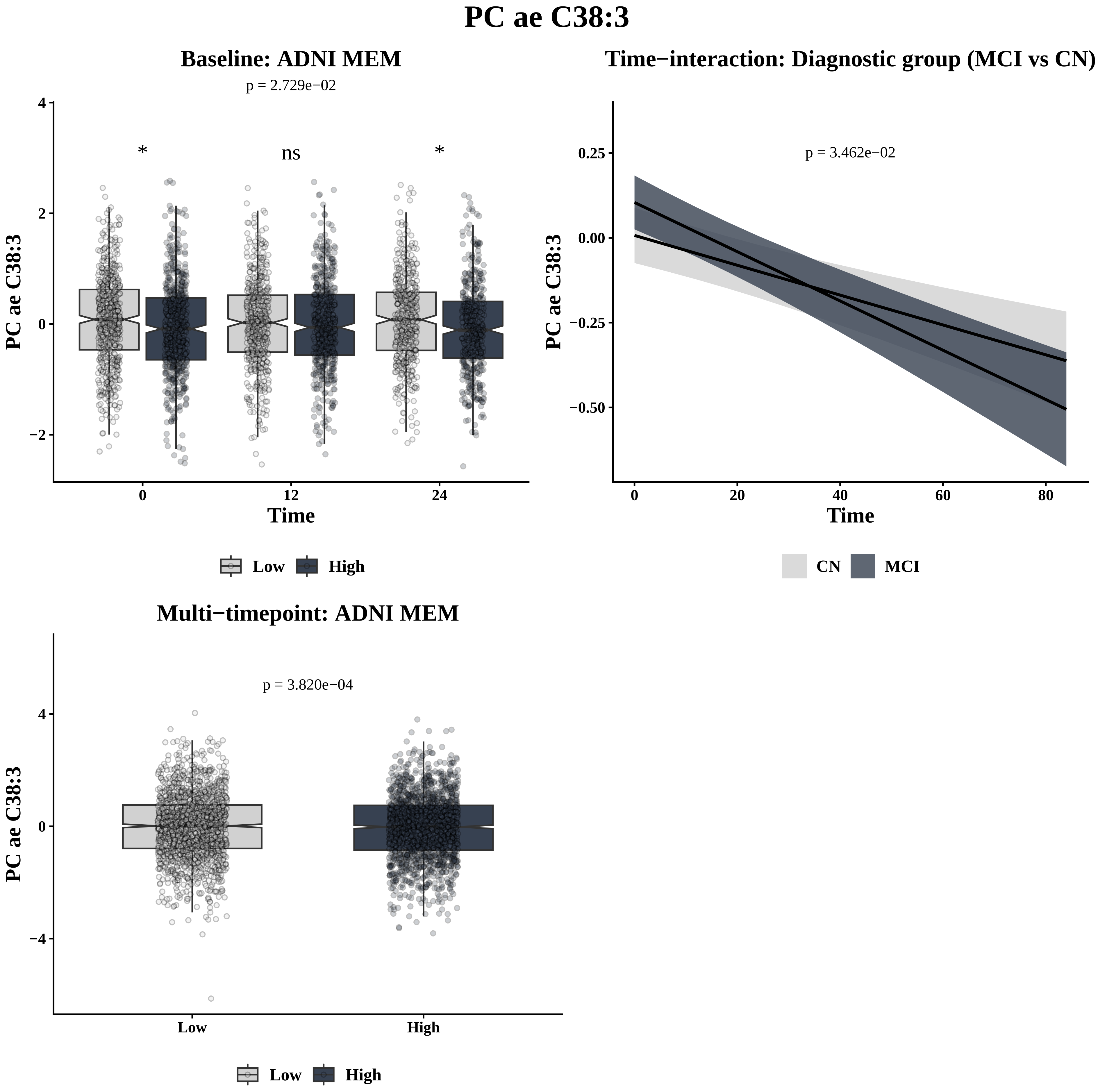
PC ae C38:3
Baseline: ADNI MEM
p = 2.729e−02
*
ns
*
4
2
0
−2
PC ae C38:3
0
12
24
Time
Low
High
Time−interaction: Diagnostic group (MCI vs CN)
p = 3.462e−02
0.25
0.00
−0.25
−0.50
PC ae C38:3
0
20
40
60
80
Time
CN
MCI
Multi−timepoint: ADNI MEM
p = 3.820e−04
4
0
−4
PC ae C38:3
Low
High
Low
High

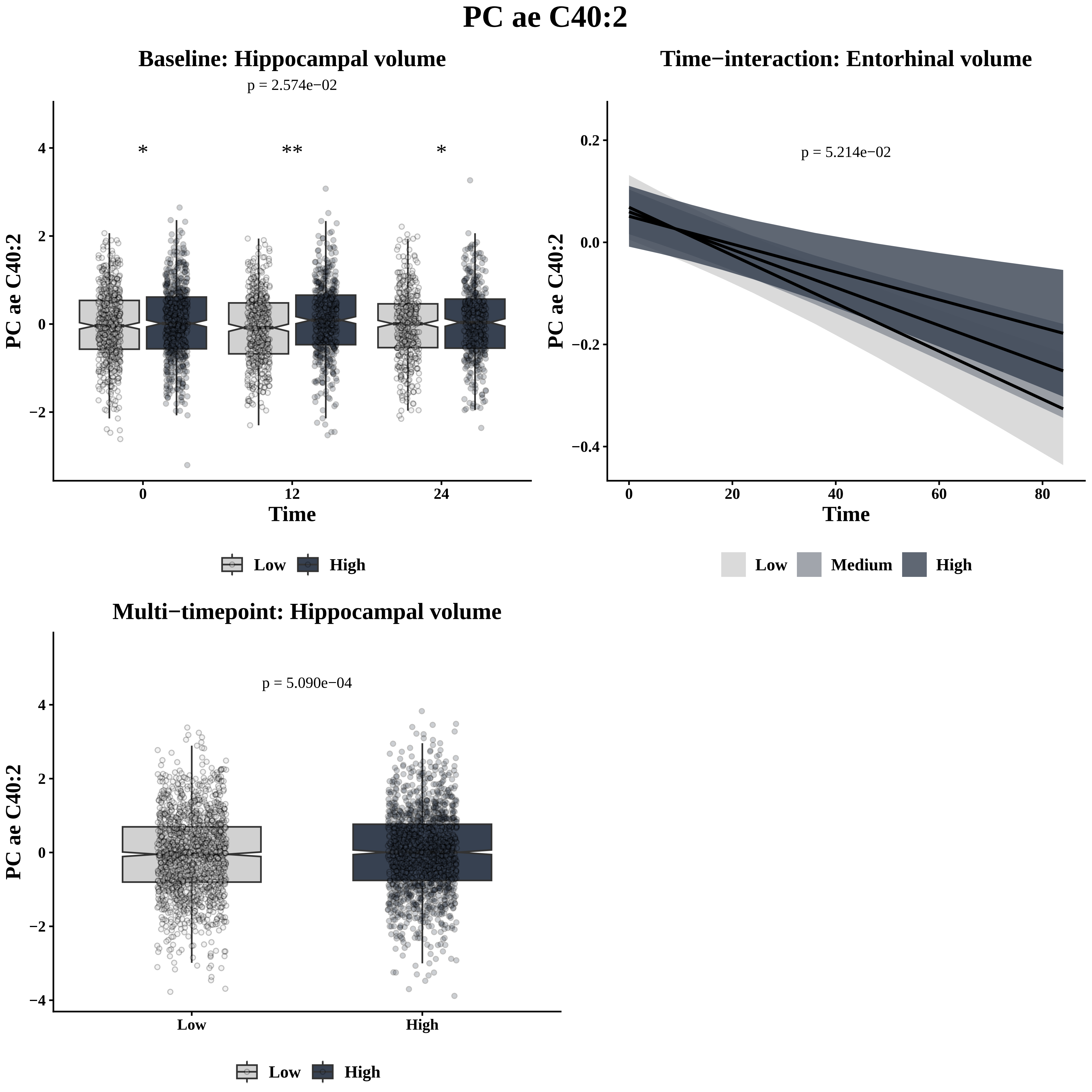
PC ae C40:2
Baseline: Hippocampal volume
p = 2.574e−02
*
**
*
PC ae C40:2
Time
0
12
24
Low
High
Time−interaction: Entorhinal volume
p = 5.214e−02
PC ae C40:2
Time
0
20
40
60
80
Low
Medium
High
Multi−timepoint: Hippocampal volume
p = 5.090e−04
PC ae C40:2
Low
High
Low
High

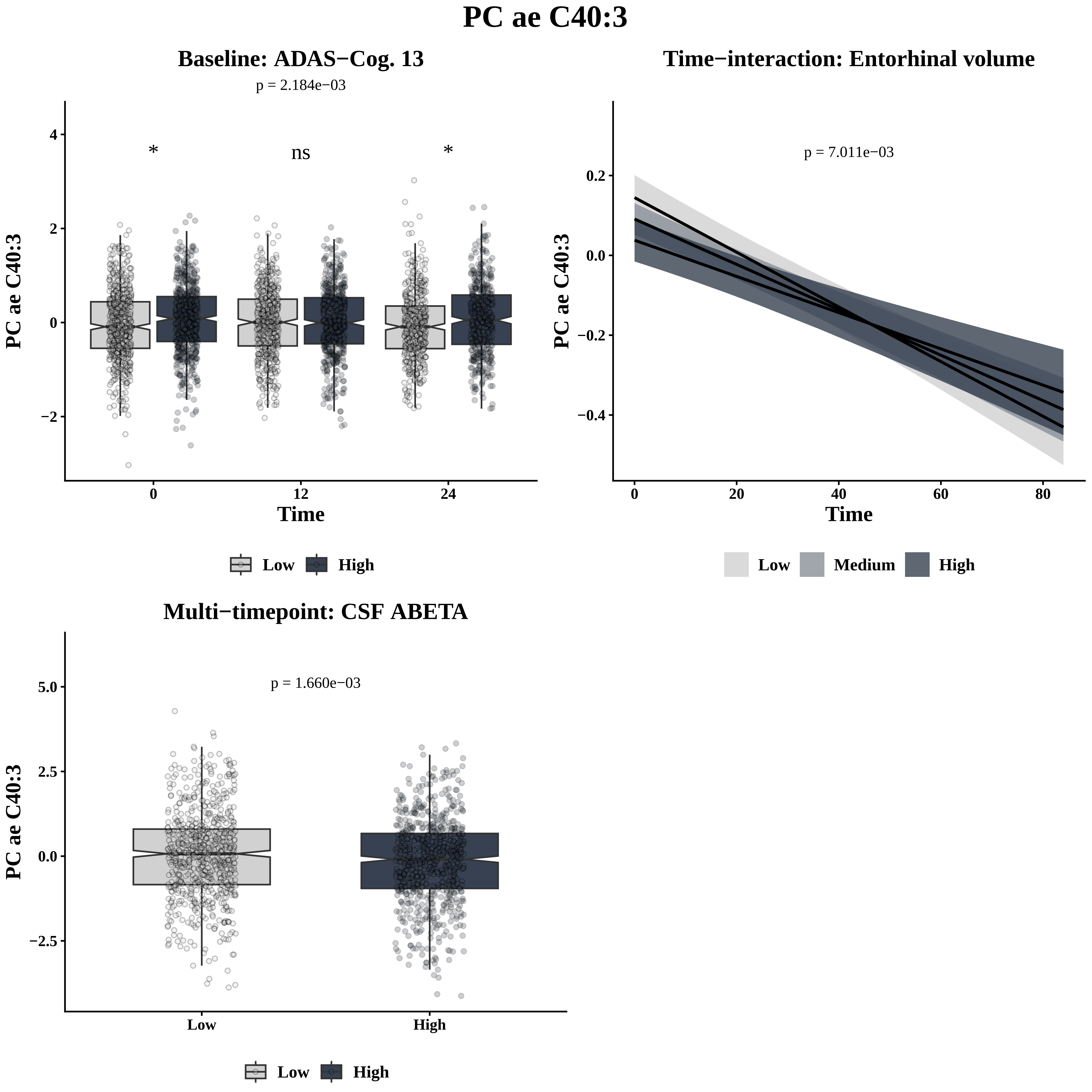

PC ae C40:3
Baseline: ADAS−Cog. 13
p = 2.184e−03
*
ns
*
PC ae C40:3
4
2
0
−2
0
12
24
Time
Low
High
Time−interaction: Entorhinal volume
p = 7.011e−03
PC ae C40:3
0.2
0.0
−0.2
−0.4
0
20
40
60
80
Time
Low
Medium
High
Multi−timepoint: CSF ABETA
p = 1.660e−03
PC ae C40:3
5.0
2.5
0.0
−2.5
Low
High
Low
High

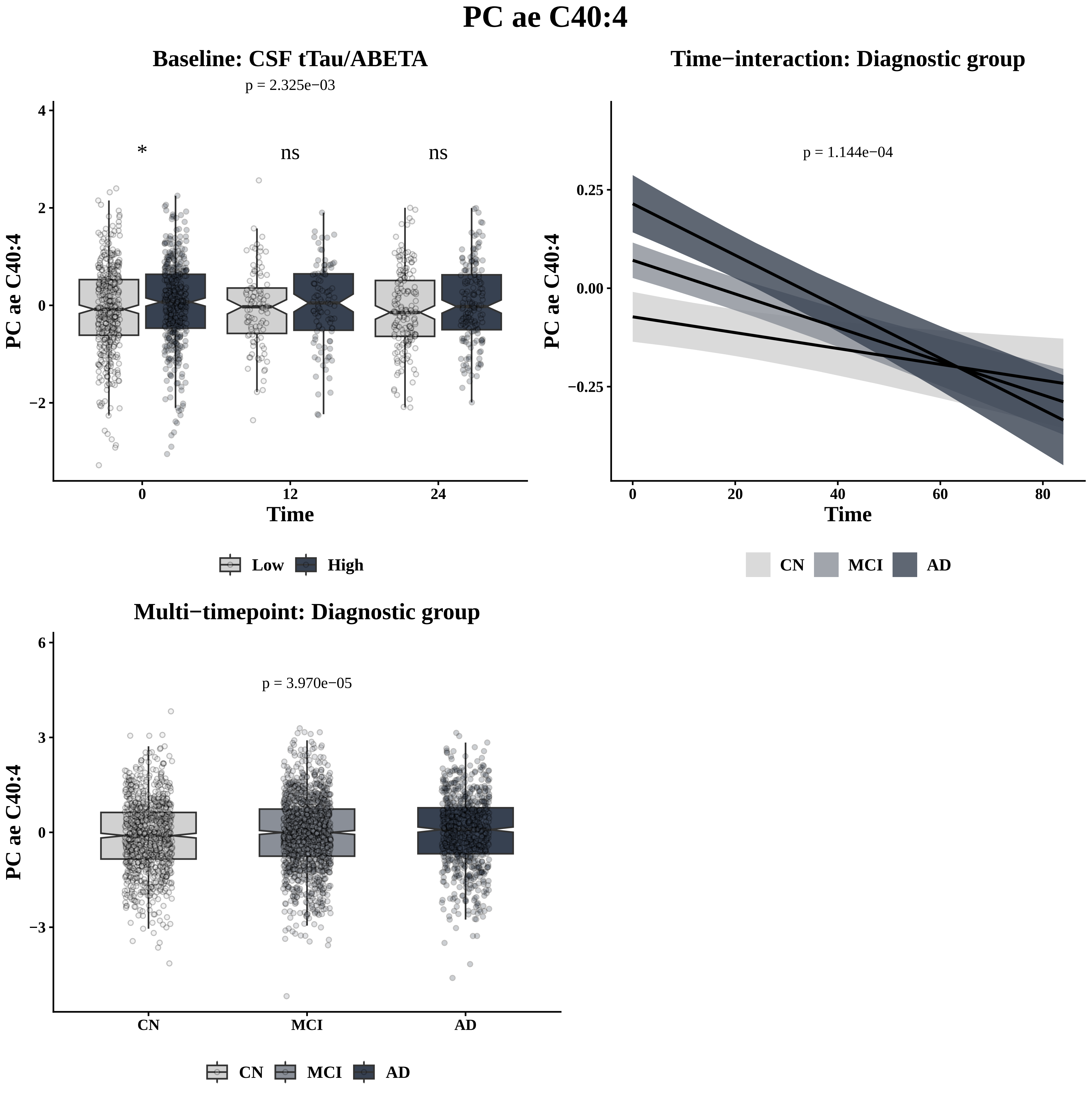
PC ae C40:4
Baseline: CSF tTau/ABETA
p = 2.325e−03
*
ns
ns
PC ae C40:4
Time
0
12
24
4
2
0
−2
Low
High
Time−interaction: Diagnostic group
p = 1.144e−04
0.25
0.00
−0.25
PC ae C40:4
Time
0
20
40
60
80
CN
MCI
AD
Multi−timepoint: Diagnostic group
p = 3.970e−05
6
3
0
−3
PC ae C40:4
CN
MCI
AD
CN
MCI
AD

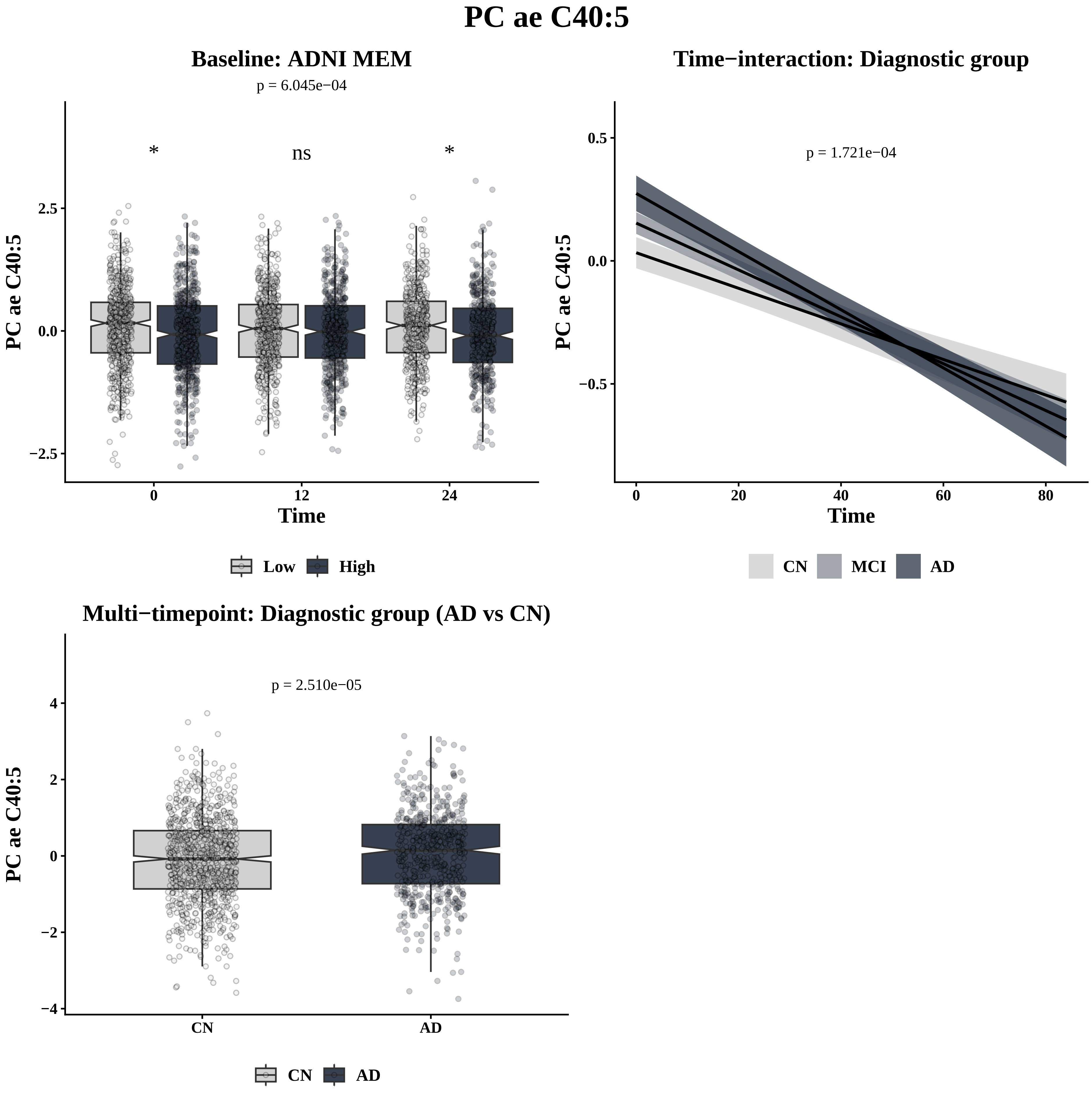
PC ae C40:5
Baseline: ADNI MEM
p = 6.045e−04
*
ns
*
2.5
0.0
−2.5
PC ae C40:5
0
12
24
Time
Low
High
Time−interaction: Diagnostic group
p = 1.721e−04
0.5
0.0
−0.5
PC ae C40:5
0
20
40
60
80
Time
CN
MCI
AD
Multi−timepoint: Diagnostic group (AD vs CN)
p = 2.510e−05
4
2
0
−2
−4
PC ae C40:5
CN
AD
CN
AD

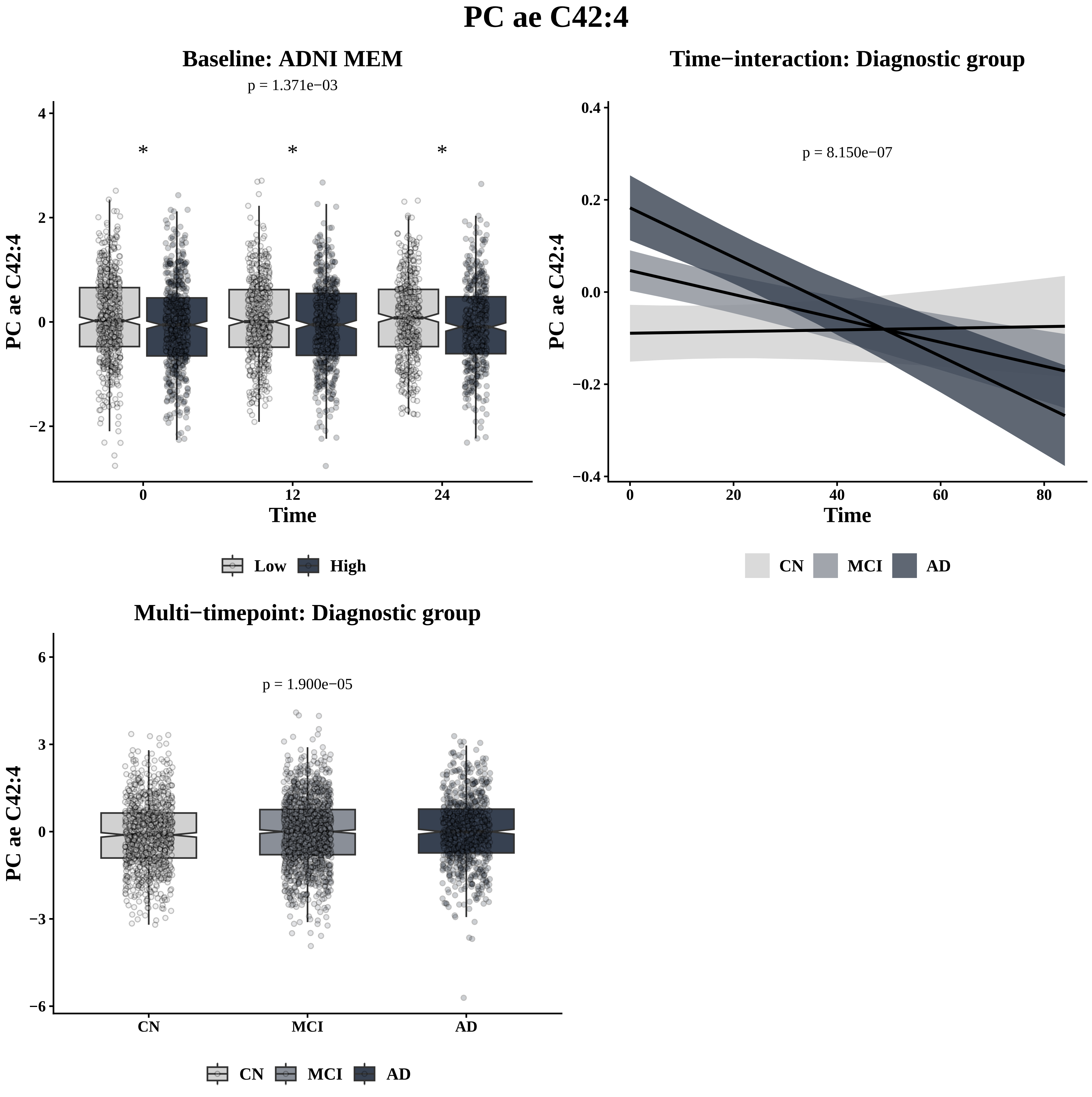
PC ae C42:4
Baseline: ADNI MEM
p = 1.371e−03
*
*
*
PC ae C42:4
Time
4
2
0
−2
0
12
24
Low
High
Time−interaction: Diagnostic group
p = 8.150e−07
PC ae C42:4
Time
0.4
0.2
0.0
−0.2
−0.4
0
20
40
60
80
CN
MCI
AD
Multi−timepoint: Diagnostic group
p = 1.900e−05
PC ae C42:4
6
3
0
−3
−6
CN
MCI
AD
CN
MCI
AD

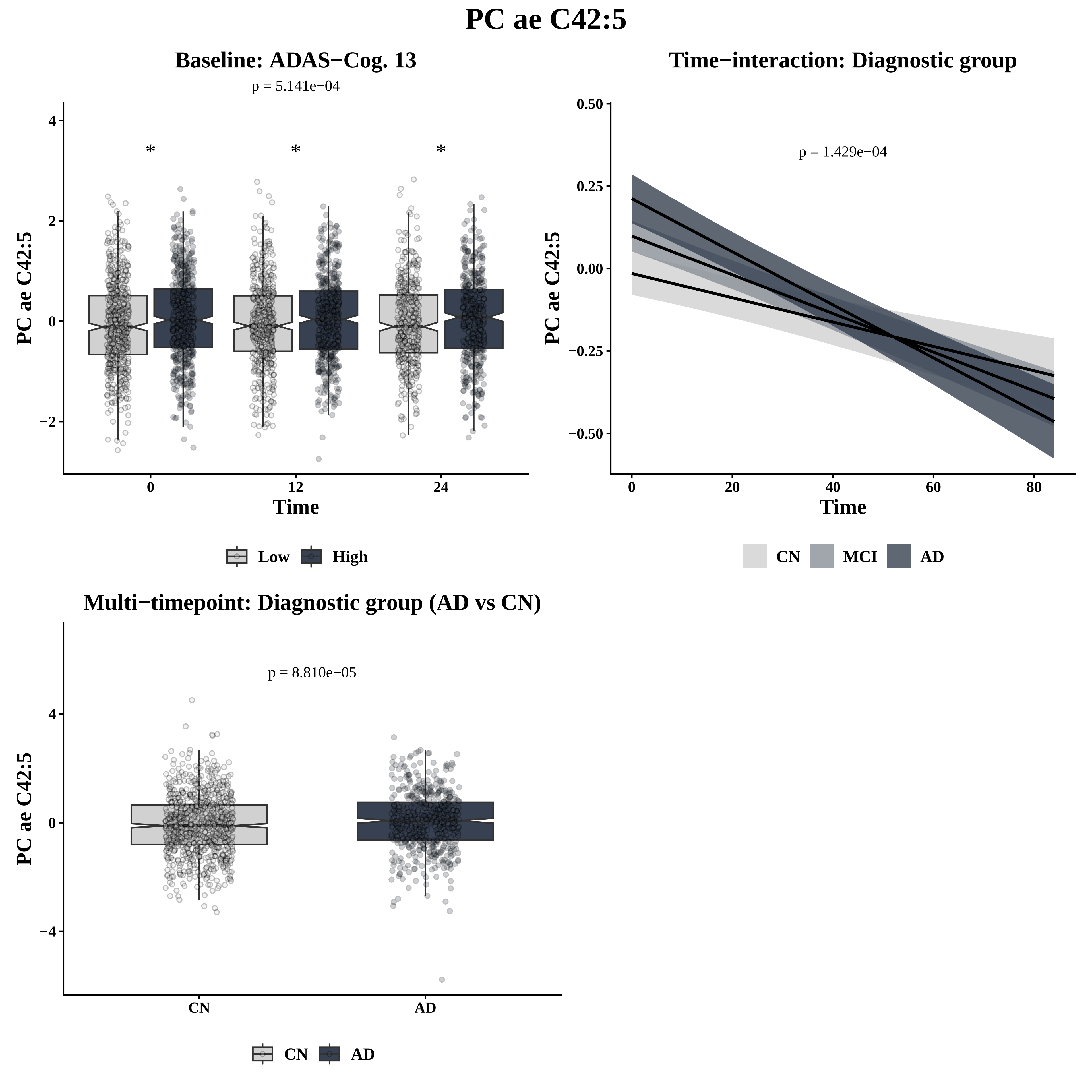
PC ae C42:5
Baseline: ADAS−Cog. 13
p = 5.141e−04
*
*
*
PC ae C42:5
Time
0
12
24
4
2
0
−2
Low
High
Time−interaction: Diagnostic group
p = 1.429e−04
0.50
0.25
0.00
−0.25
−0.50
0
20
40
60
80
CN
MCI
AD
Multi−timepoint: Diagnostic group (AD vs CN)
p = 8.810e−05
4
0
−4
CN
AD

# PC ae C44:5

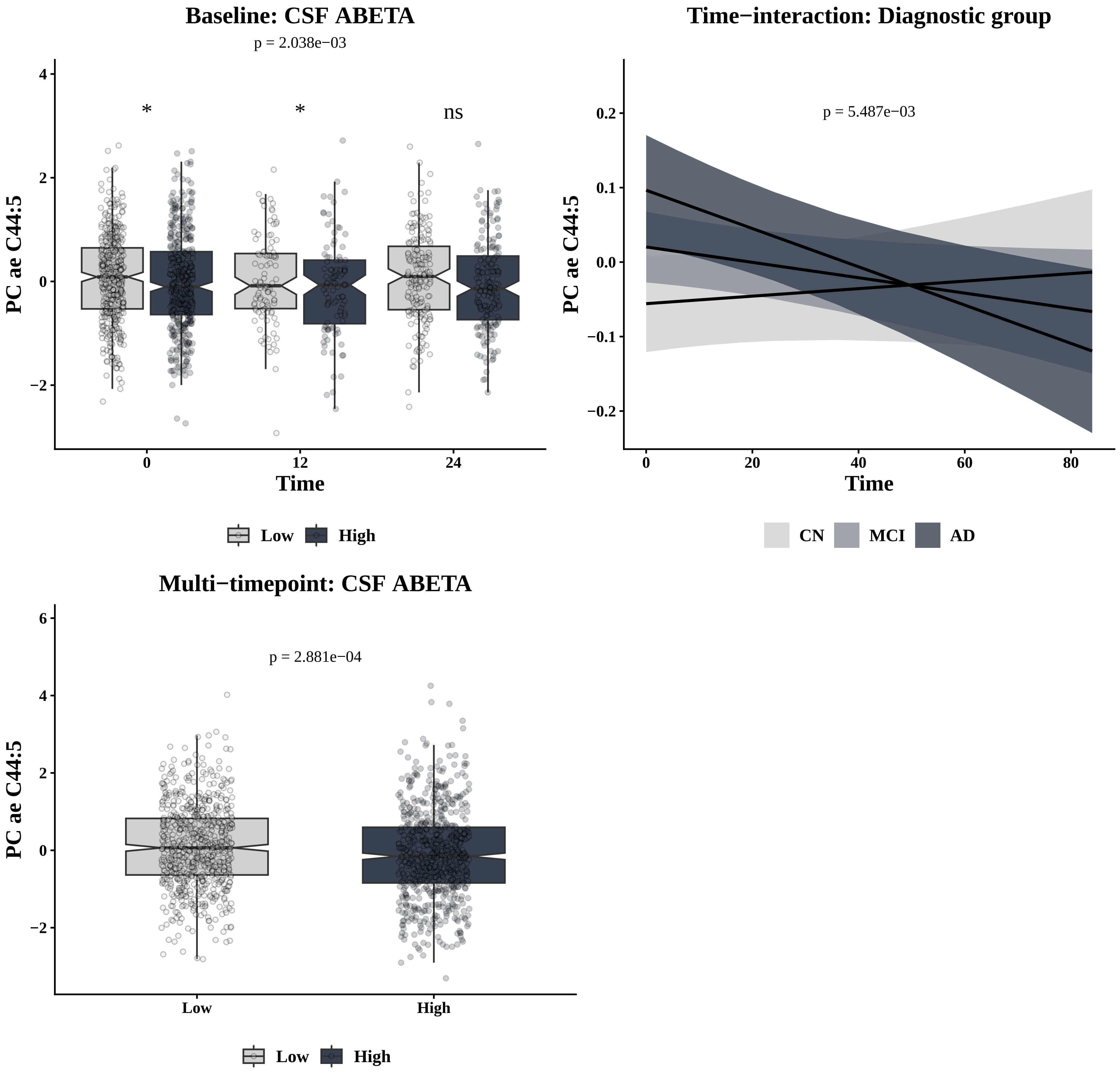

Baseline: CSF ABETA
p = 2.038e−03
*
*
ns
PC ae C44:5
Time
0
12
24
Low
High
Time−interaction: Diagnostic group
p = 5.487e−03
PC ae C44:5
Time
CN
MCI
AD
Multi−timepoint: CSF ABETA
p = 2.881e−04
PC ae C44:5
Low
High

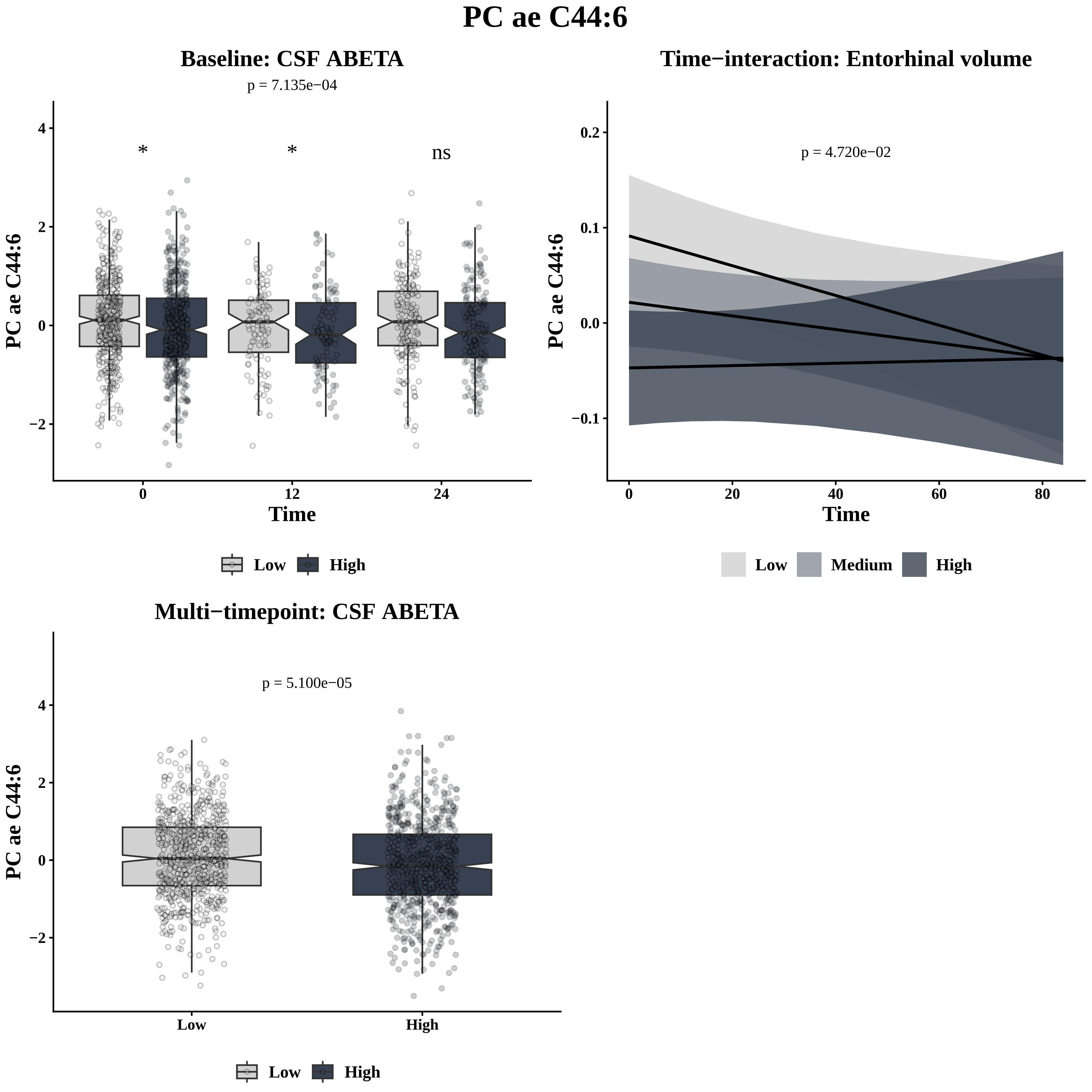
PC ae C44:6
Baseline: CSF ABETA
p = 7.135e−04
*
*
ns
PC ae C44:6
Time
0
12
24
Low
High
Time−interaction: Entorhinal volume
p = 4.720e−02
PC ae C44:6
Time
Low
Medium
High
Multi−timepoint: CSF ABETA
p = 5.100e−05
PC ae C44:6
Low
High
Low
High

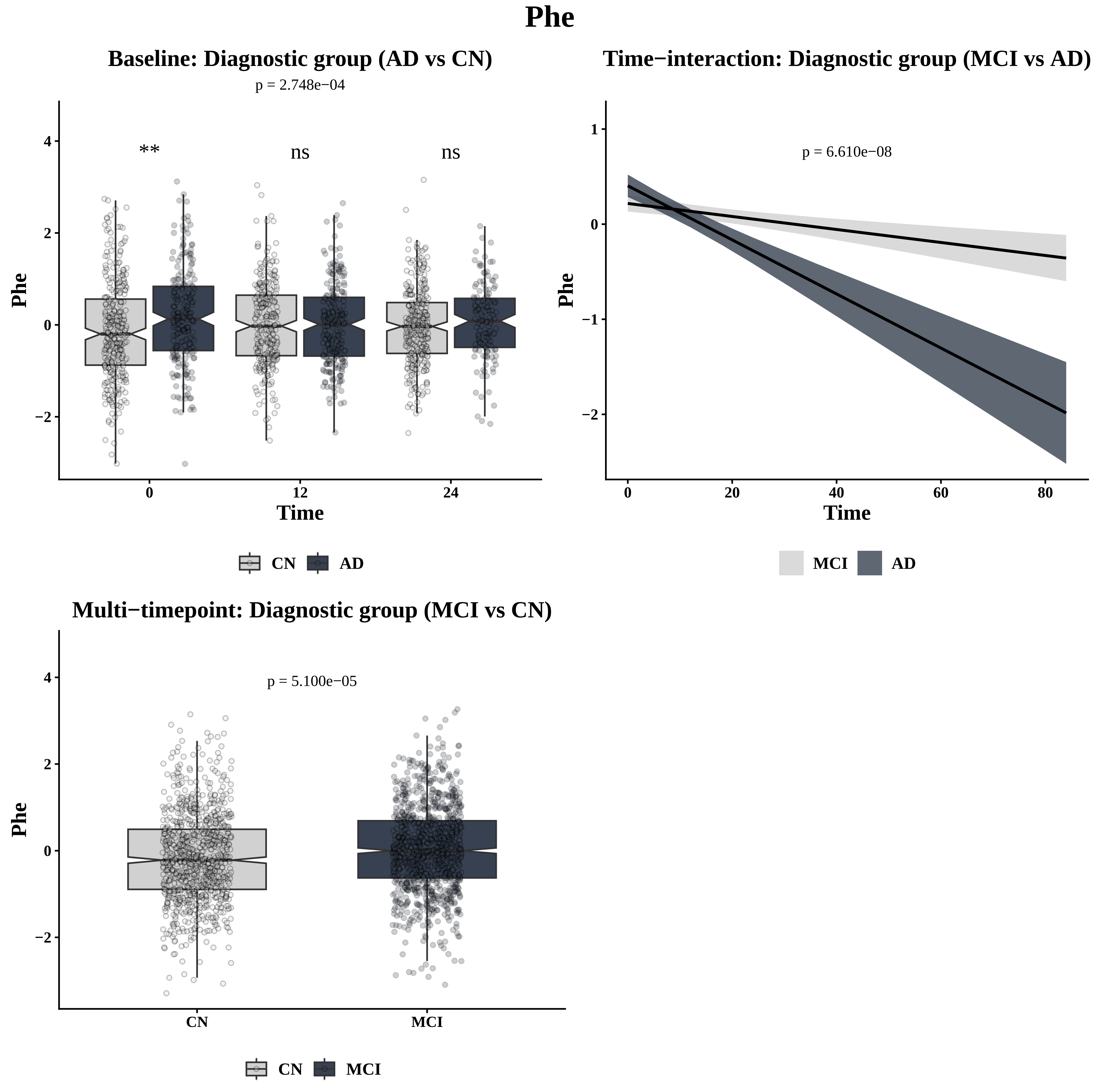
Phe
Baseline: Diagnostic group (AD vs CN)
p = 2.748e−04
**
ns
ns
Phe
Time
0
12
24
CN
AD
Time−interaction: Diagnostic group (MCI vs AD)
p = 6.610e−08
Phe
Time
0
20
40
60
80
MCI
AD
Multi−timepoint: Diagnostic group (MCI vs CN)
p = 5.100e−05
Phe
CN
MCI
CN
MCI

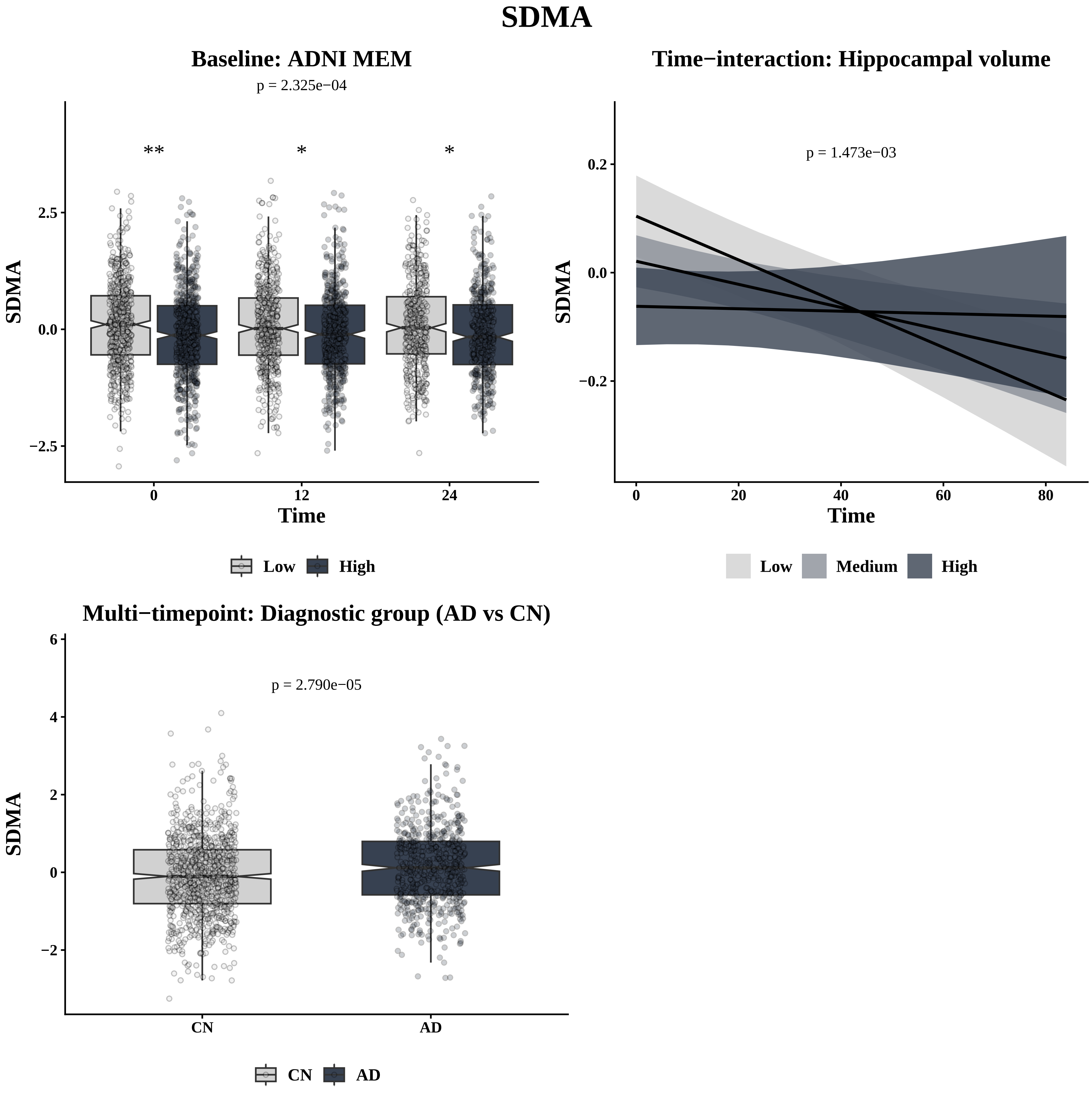
SDMA
Baseline: ADNI MEM
p = 2.325e−04
**
*
*
2.5
0.0
−2.5
SDMA
0
12
24
Time
Low
High
Time−interaction: Hippocampal volume
p = 1.473e−03
0.2
0.0
−0.2
SDMA
0
20
40
60
80
Time
Low
Medium
High
Multi−timepoint: Diagnostic group (AD vs CN)
p = 2.790e−05
6
4
2
0
−2
SDMA
CN
AD
CN
AD

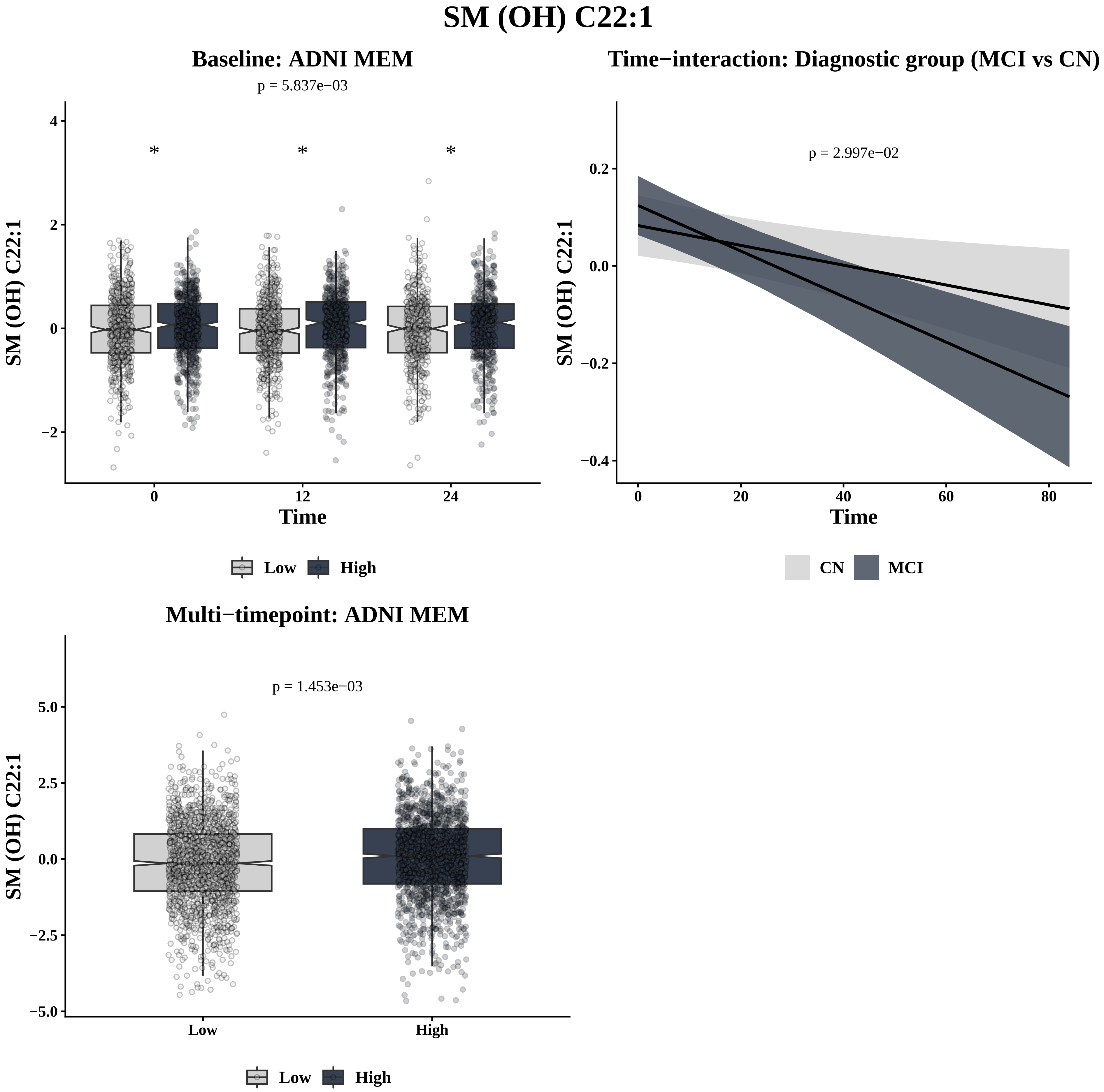

SM (OH) C22:1
Baseline: ADNI MEM
p = 5.837e−03
*
*
*
SM (OH) C22:1
Time
0
12
24
Low
High
Time−interaction: Diagnostic group (MCI vs CN)
p = 2.997e−02
SM (OH) C22:1
Time
CN
MCI
Multi−timepoint: ADNI MEM
p = 1.453e−03
SM (OH) C22:1
Low
High
Low
High

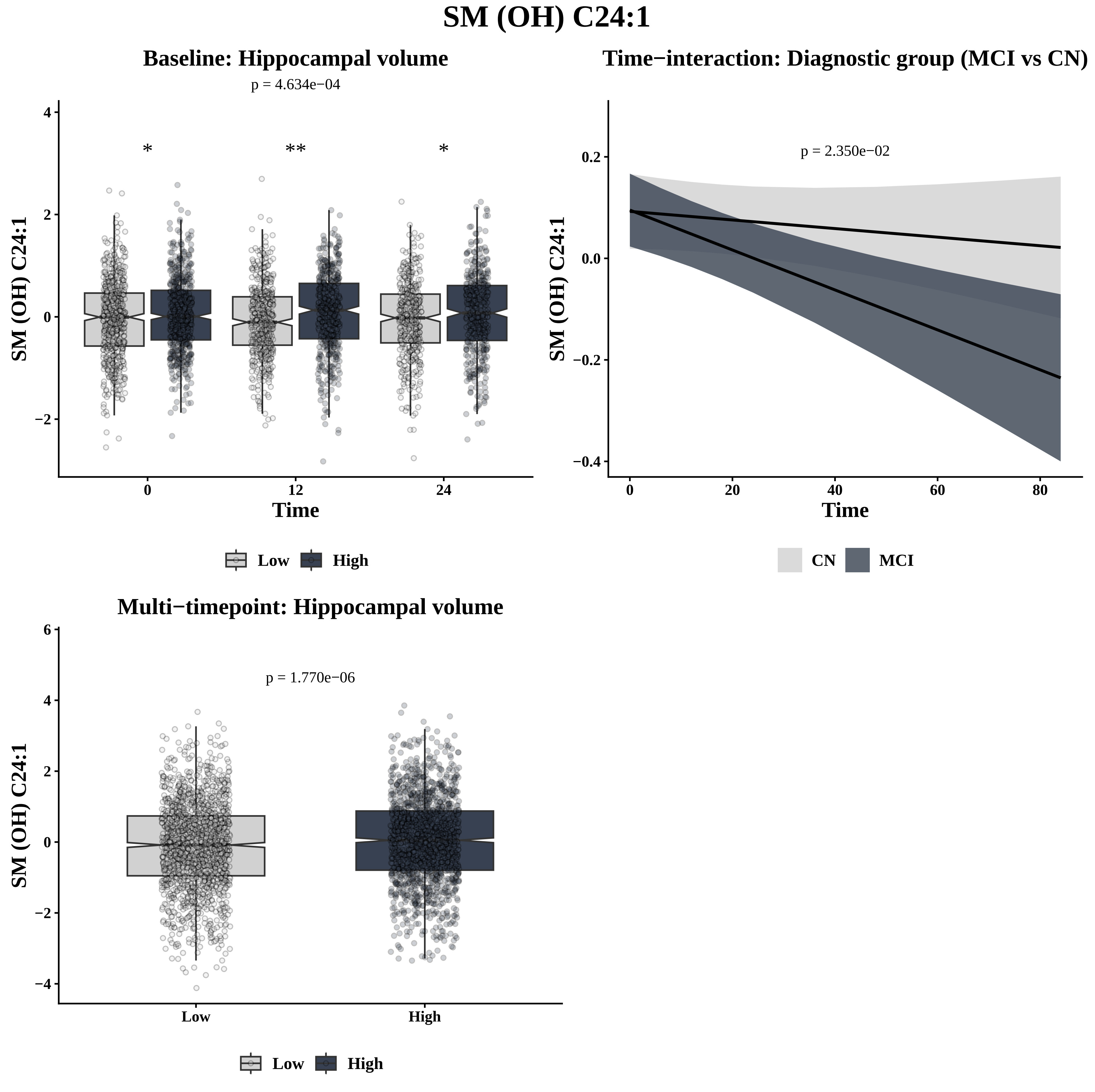

SM (OH) C24:1
Baseline: Hippocampal volume
p = 4.634e−04
*
**
*
SM (OH) C24:1
4
2
0
−2
0
12
24
Time
Low
High
Time−interaction: Diagnostic group (MCI vs CN)
p = 2.350e−02
SM (OH) C24:1
0.2
0.0
−0.2
−0.4
0
20
40
60
80
Time
CN
MCI
Multi−timepoint: Hippocampal volume
p = 1.770e−06
SM (OH) C24:1
6
4
2
0
−2
−4
Low
High
Low
High

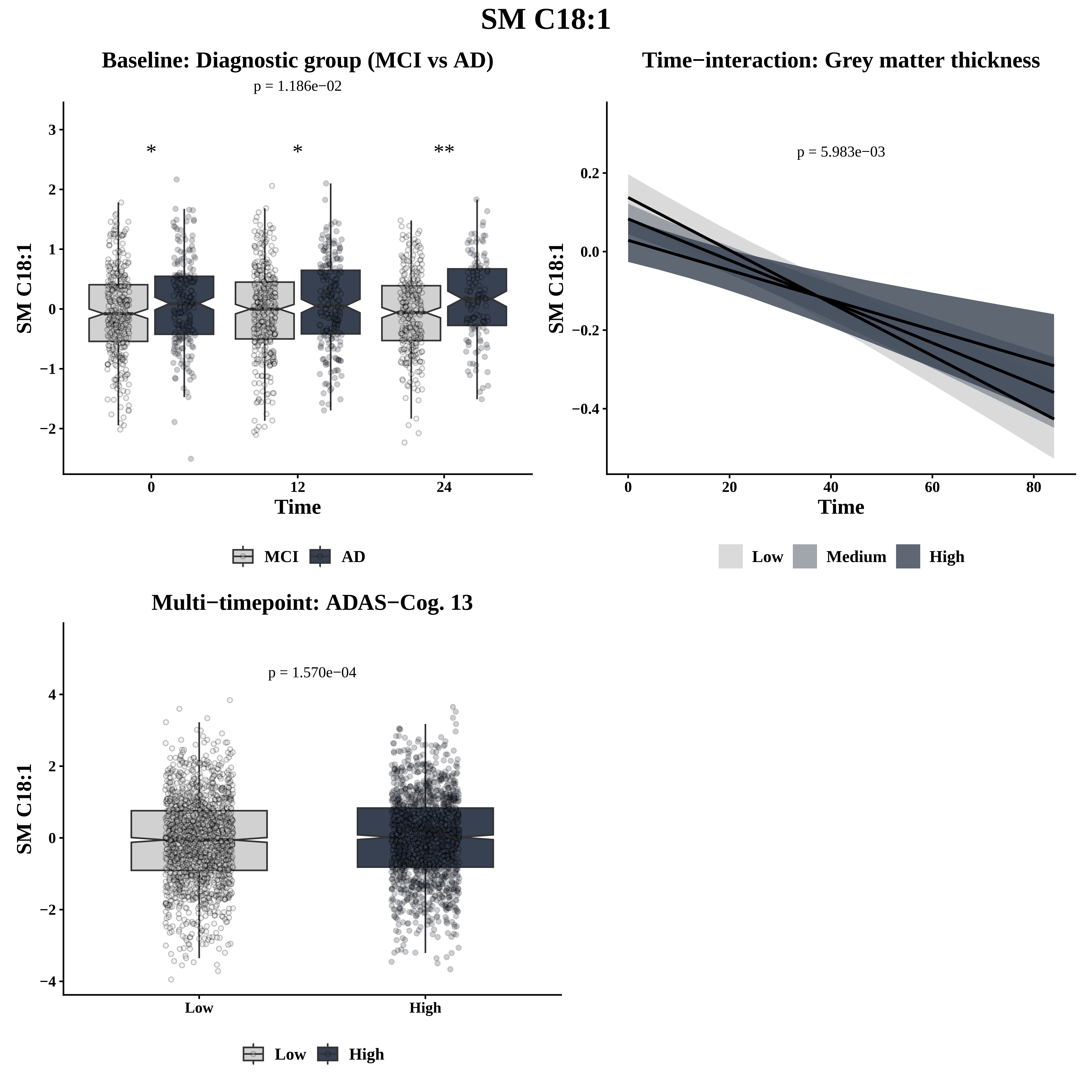
SM C18:1
Baseline: Diagnostic group (MCI vs AD)
p = 1.186e−02
*
*
**
SM C18:1
3
2
1
0
−1
−2
0
12
24
Time
MCI
AD
Time−interaction: Grey matter thickness
p = 5.983e−03
SM C18:1
0.2
0.0
−0.2
−0.4
0
20
40
60
80
Time
Low
Medium
High
Multi−timepoint: ADAS−Cog. 13
p = 1.570e−04
SM C18:1
4
2
0
−2
−4
Low
High
Low
High

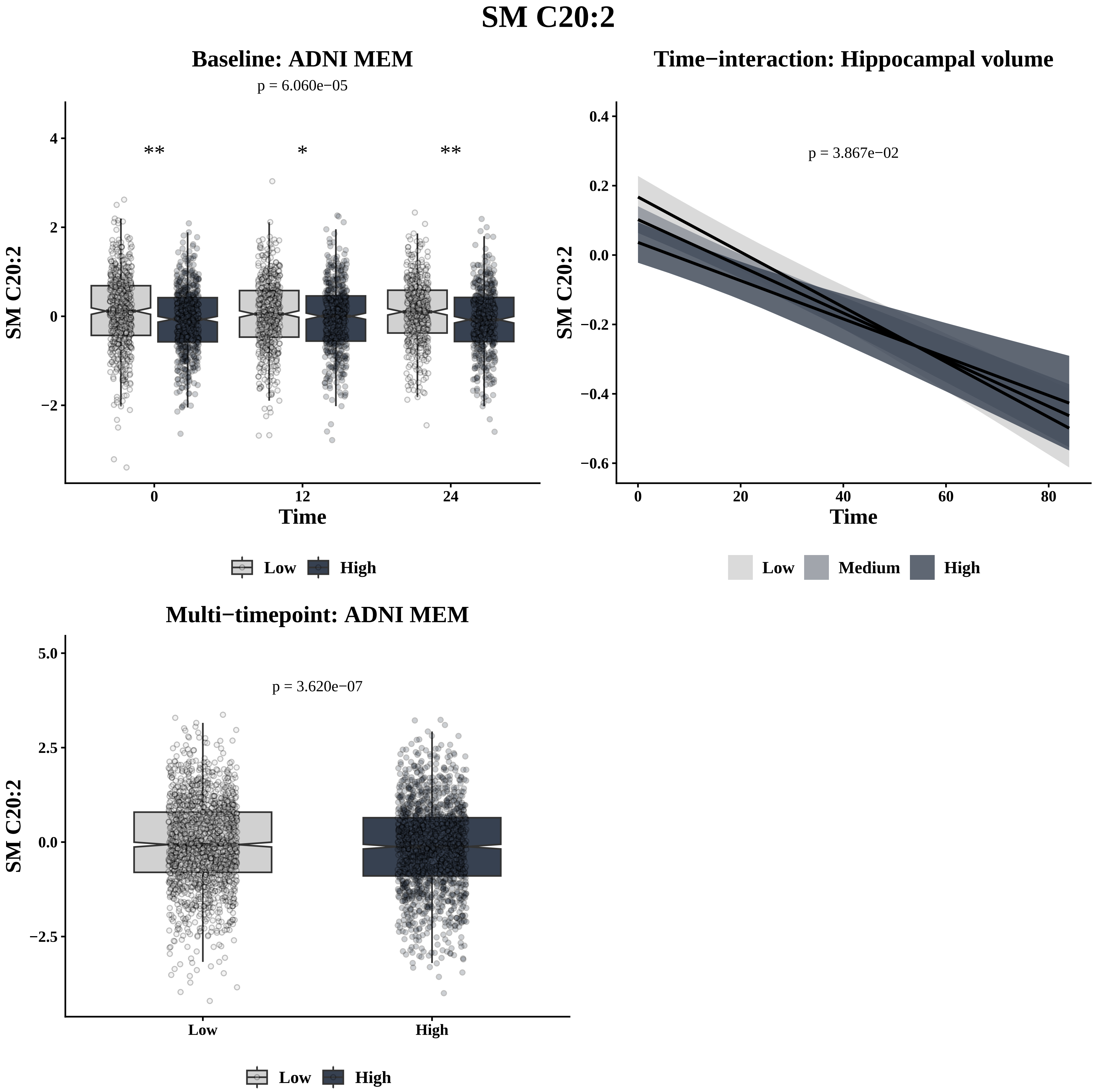

SM C20:2
Baseline: ADNI MEM
p = 6.060e−05
**
*
**
SM C20:2
Time
0
12
24
4
2
0
−2
Low
High
Time−interaction: Hippocampal volume
p = 3.867e−02
SM C20:2
Time
0.4
0.2
0.0
−0.2
−0.4
−0.6
0
20
40
60
80
Low
Medium
High
Multi−timepoint: ADNI MEM
p = 3.620e−07
SM C20:2
5.0
2.5
0.0
−2.5
Low
High
Low
High

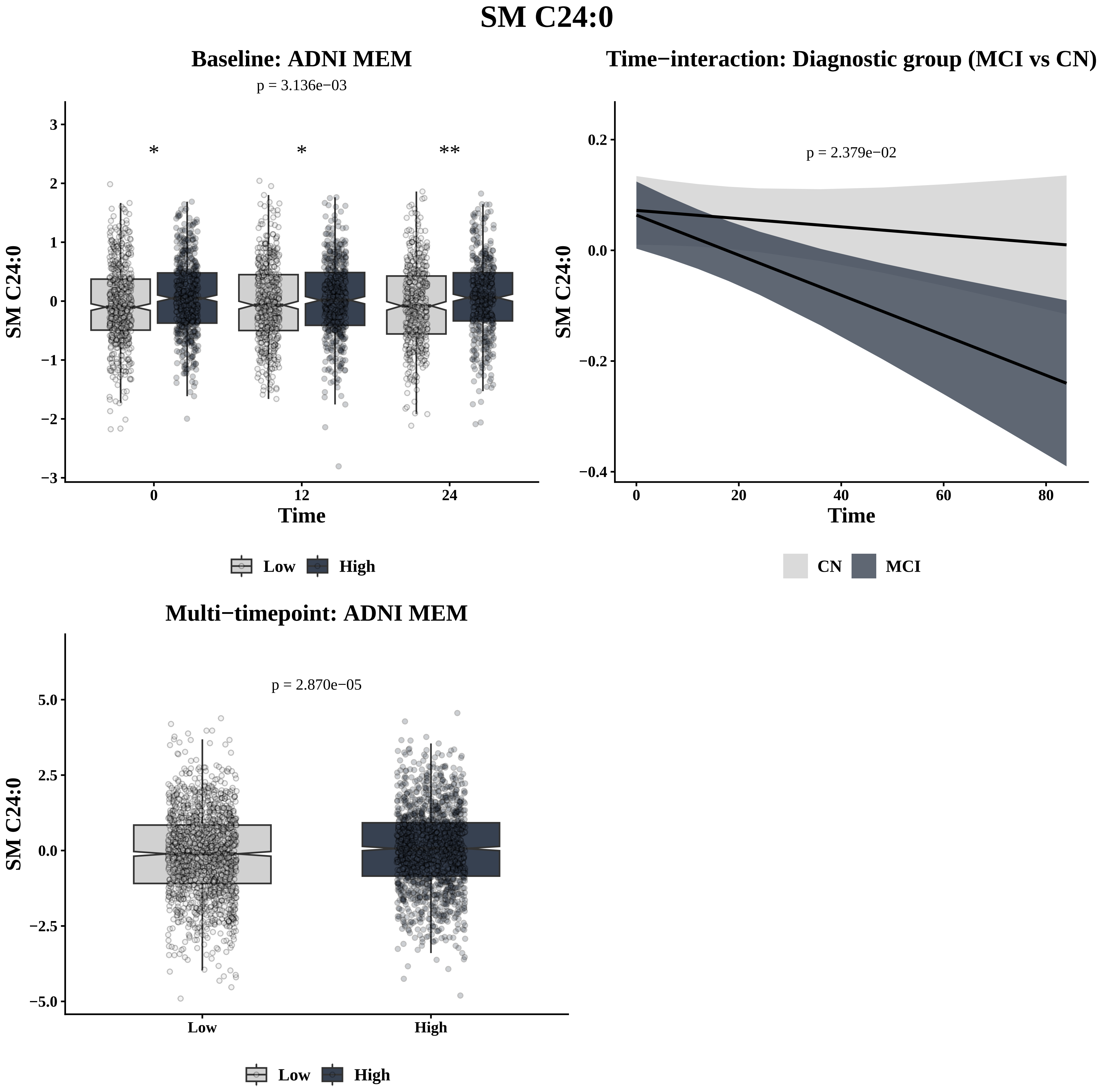
SM C24:0
Baseline: ADNI MEM
p = 3.136e−03
*
*
**
SM C24:0
Time
0
12
24
3
2
1
0
−1
−2
−3
Low
High
Time−interaction: Diagnostic group (MCI vs CN)
p = 2.379e−02
SM C24:0
Time
0.2
0.0
−0.2
−0.4
0
20
40
60
80
CN
MCI
Multi−timepoint: ADNI MEM
p = 2.870e−05
SM C24:0
5.0
2.5
0.0
−2.5
−5.0
Low
High
Low
High

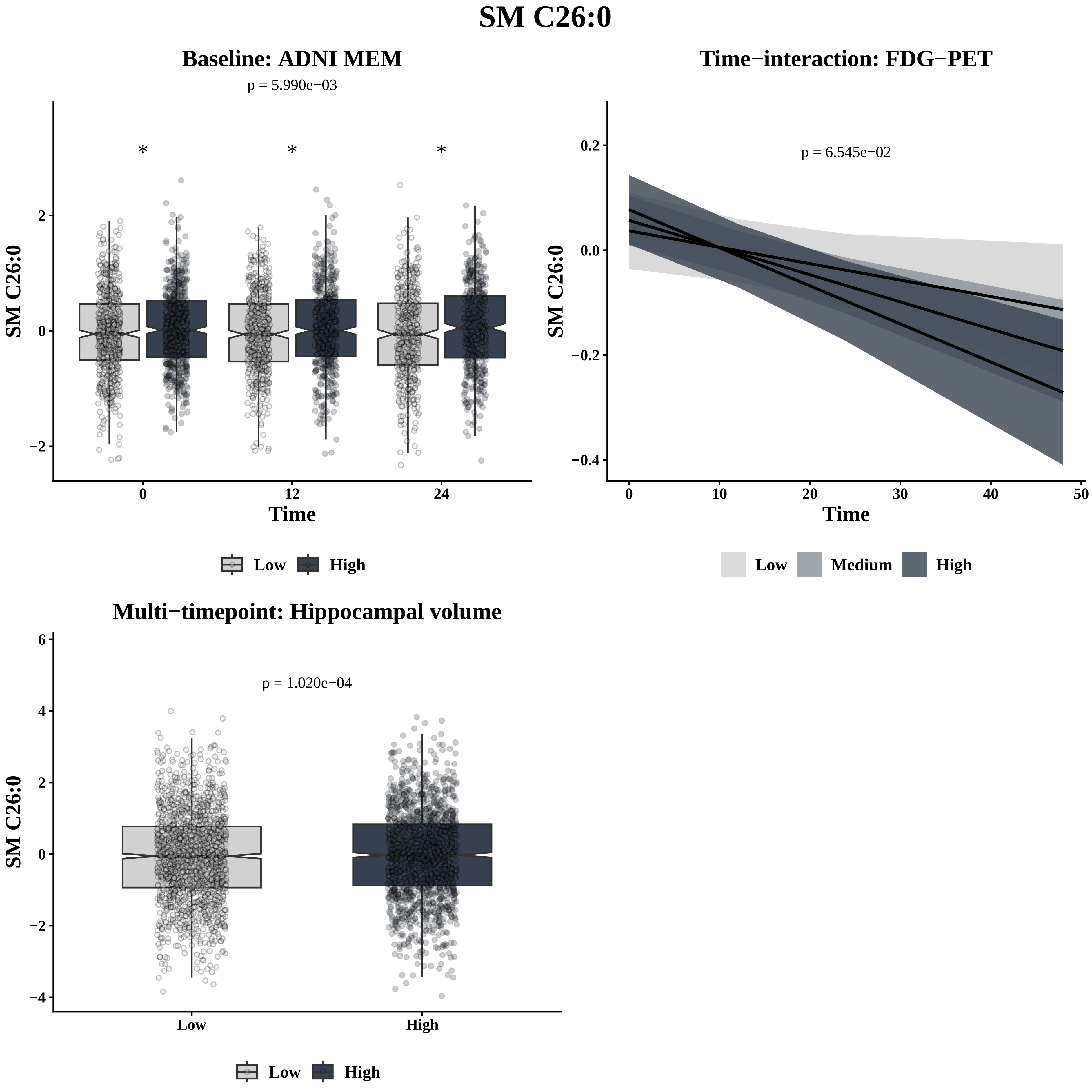
SM C26:0
Baseline: ADNI MEM
p = 5.990e−03
*
*
*
SM C26:0
2
0
−2
0
12
24
Time
Low
High
Time−interaction: FDG−PET
p = 6.545e−02
SM C26:0
0.2
0.0
−0.2
−0.4
0
10
20
30
40
50
Time
Low
Medium
High
Multi−timepoint: Hippocampal volume
p = 1.020e−04
SM C26:0
6
4
2
0
−2
−4
Low
High
Low
High

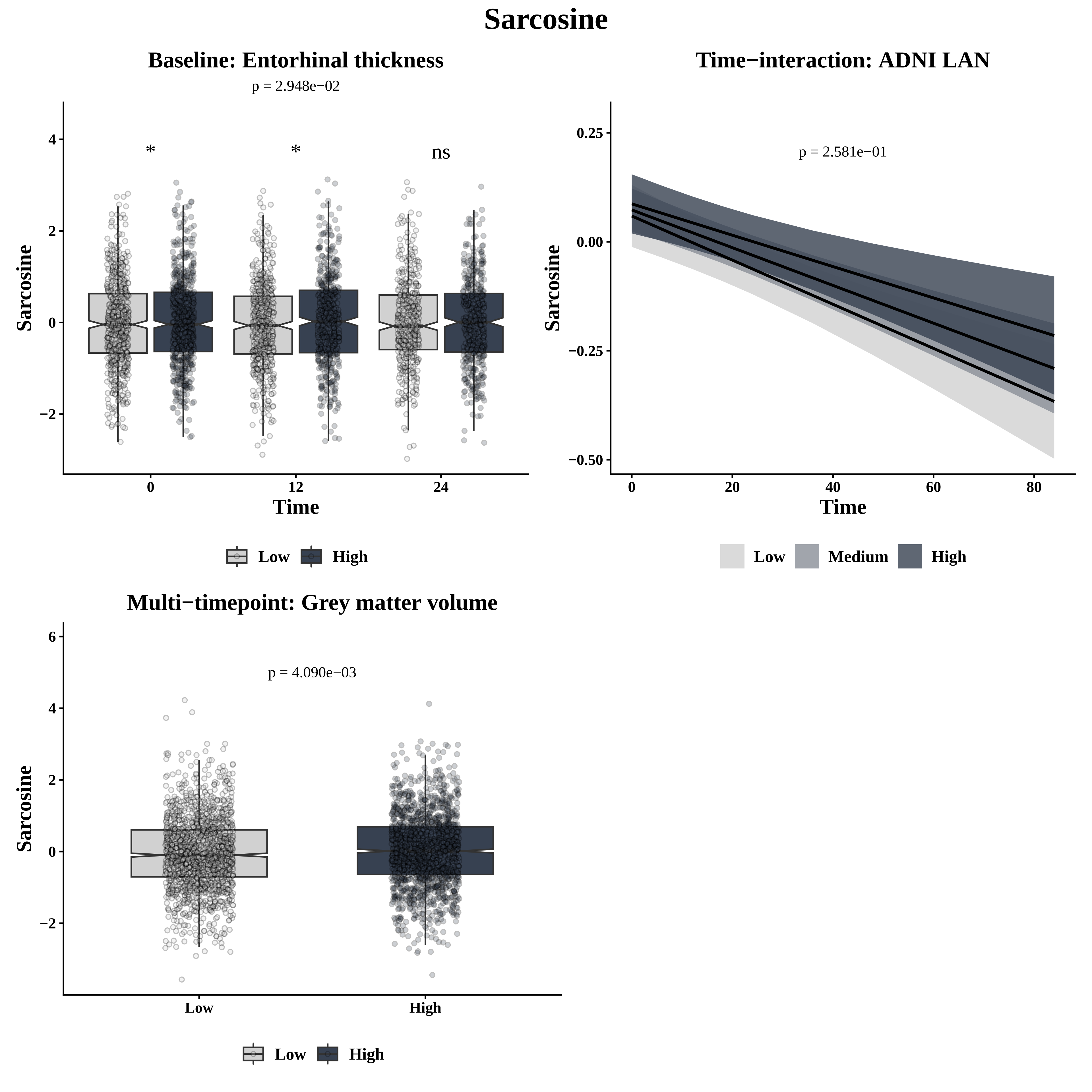

Sarcosine
Baseline: Entorhinal thickness
p = 2.948e−02
*
*
ns
Sarcosine
Time
0
12
24
4
2
0
−2
Low
High
Time−interaction: ADNI LAN
p = 2.581e−01
0.25
0.00
−0.25
−0.50
Sarcosine
Time
0
20
40
60
80
Low
Medium
High
Multi−timepoint: Grey matter volume
p = 4.090e−03
6
4
2
0
−2
Sarcosine
Low
High
Low
High

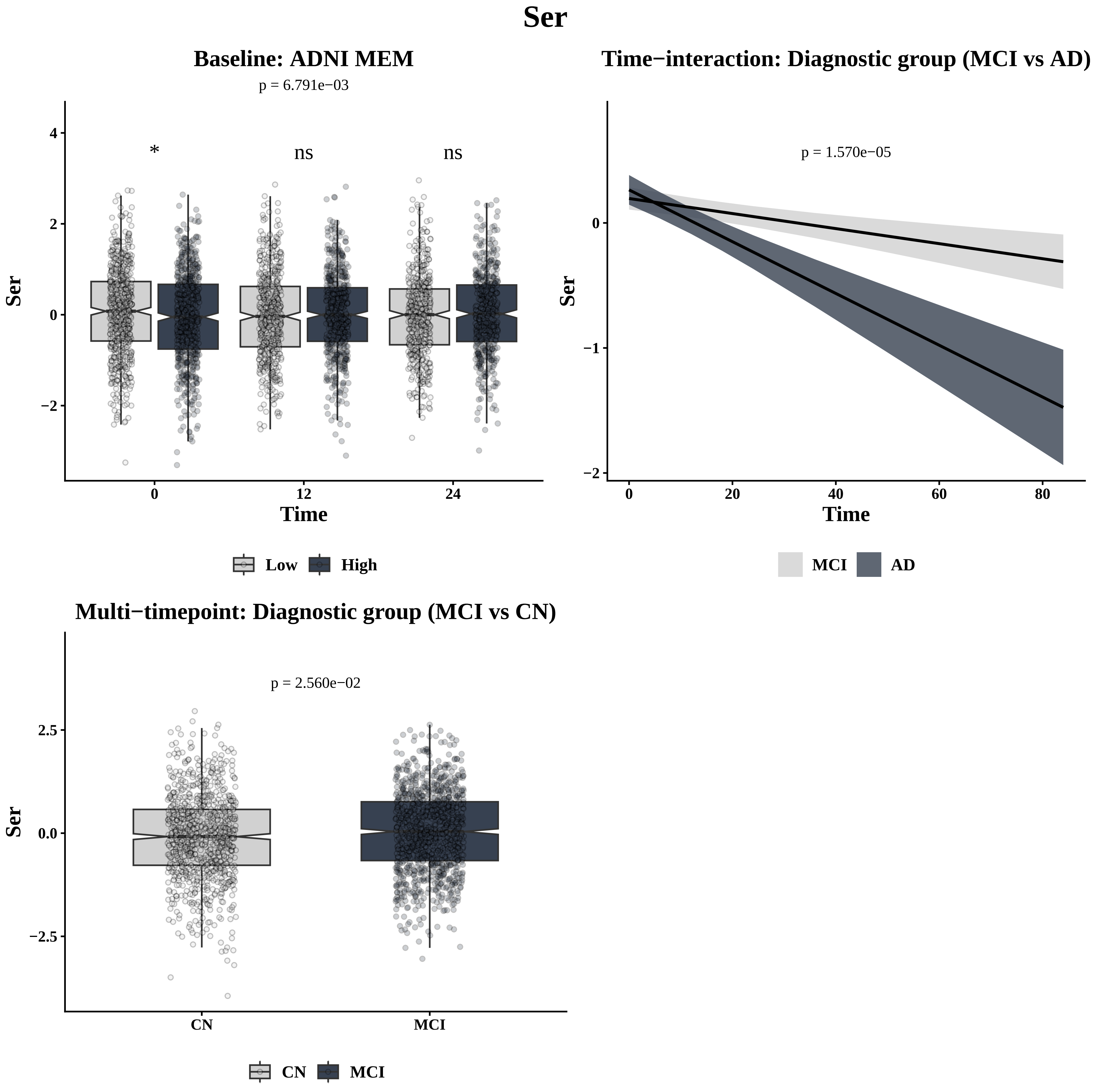

Ser
Baseline: ADNI MEM
p = 6.791e−03
*
ns
ns
4
2
0
−2
Ser
0
12
24
Time
Low
High
Time−interaction: Diagnostic group (MCI vs AD)
p = 1.570e−05
0
−1
−2
Ser
0
20
40
60
80
Time
MCI
AD
Multi−timepoint: Diagnostic group (MCI vs CN)
p = 2.560e−02
2.5
0.0
−2.5
Ser
CN
MCI
CN
MCI

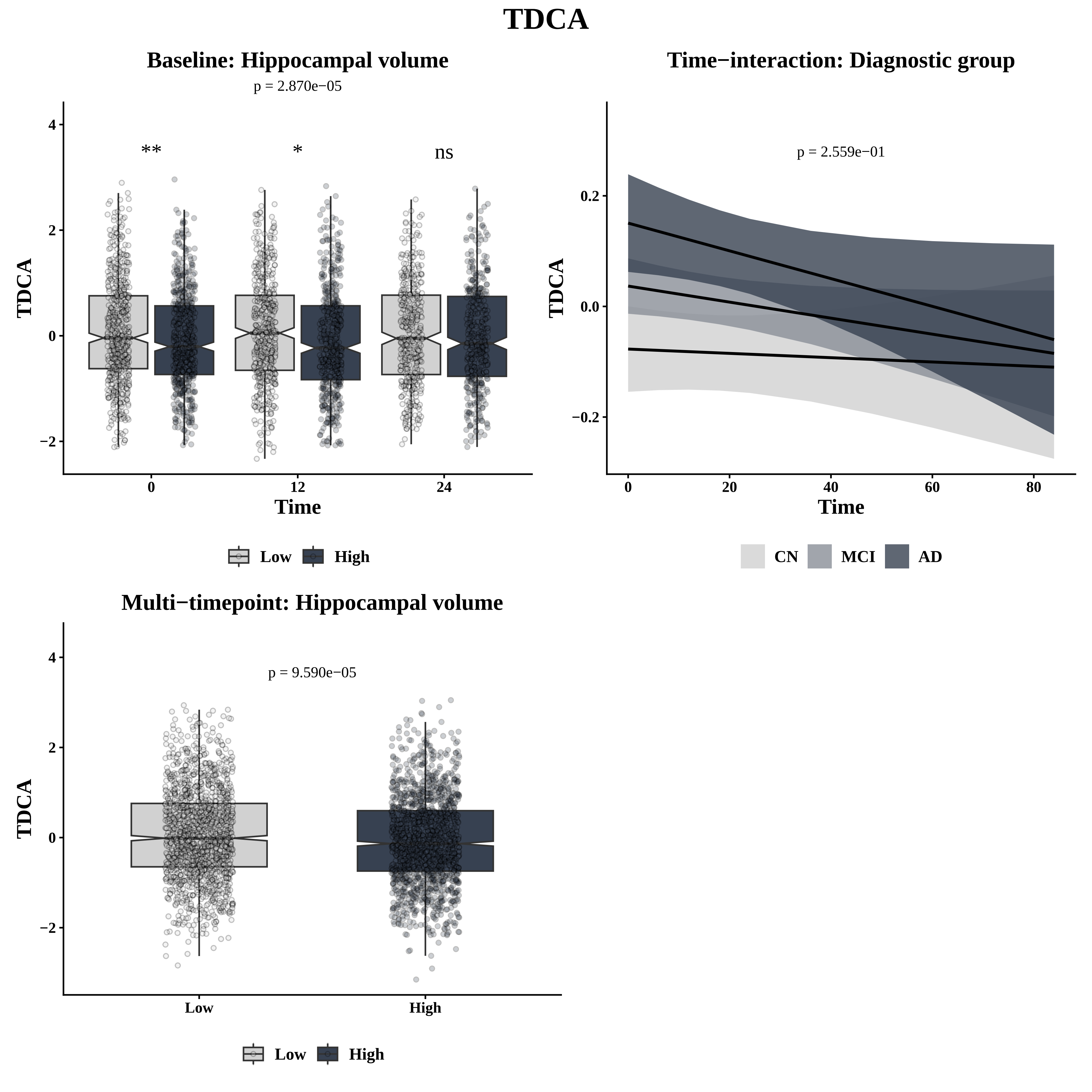

TDCA
Baseline: Hippocampal volume
p = 2.870e−05
**
*
ns
TDCA
Time
0
12
24
Low
High
Time−interaction: Diagnostic group
p = 2.559e−01
TDCA
Time
CN
MCI
AD
Multi−timepoint: Hippocampal volume
p = 9.590e−05
TDCA
Low
High

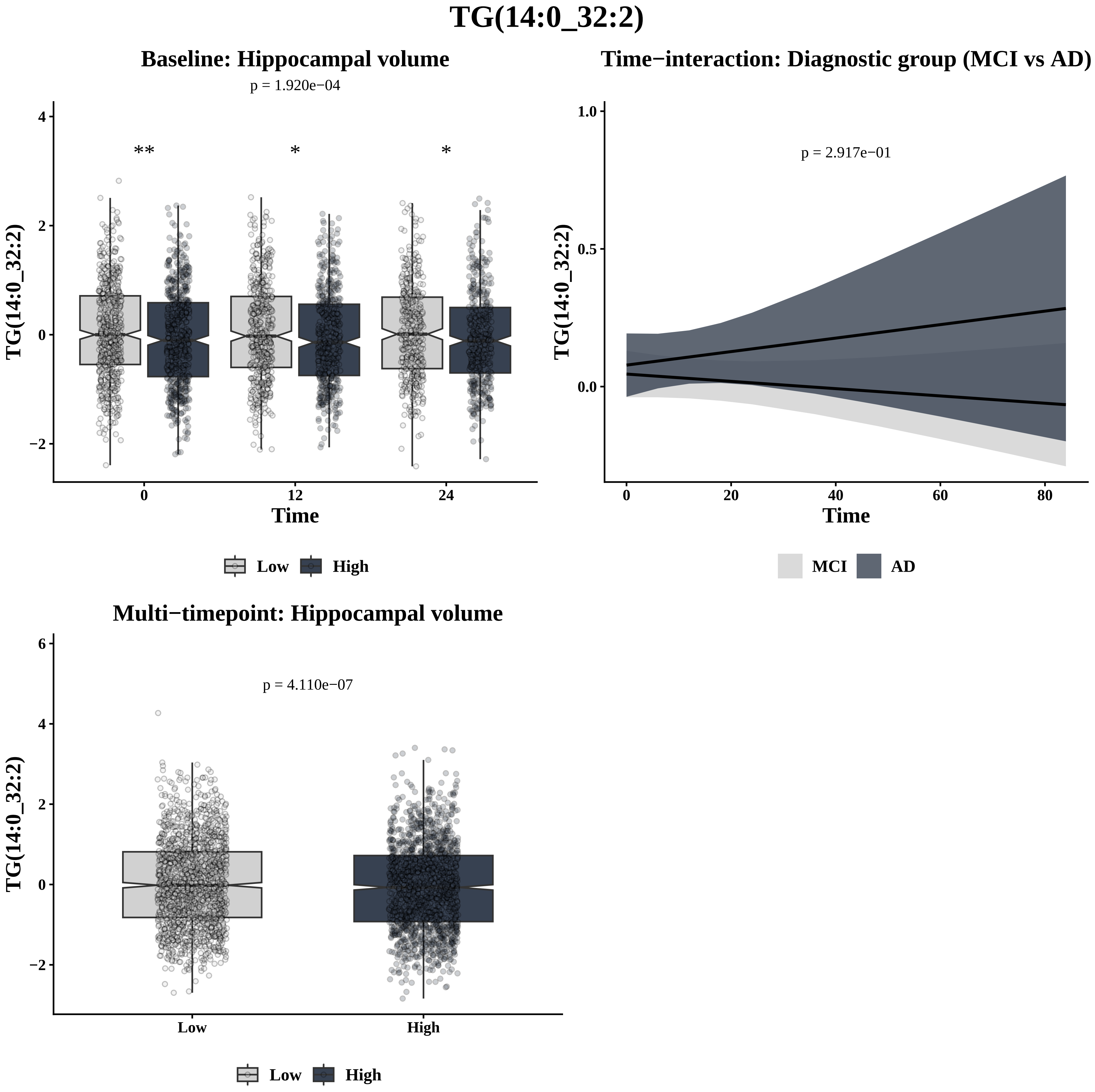

TG(14:0_32:2)
Baseline: Hippocampal volume
p = 1.920e−04
**
*
*
TG(14:0_32:2)
Time
0
12
24
Low
High
Time−interaction: Diagnostic group (MCI vs AD)
p = 2.917e−01
TG(14:0_32:2)
Time
0
20
40
60
80
MCI
AD
Multi−timepoint: Hippocampal volume
p = 4.110e−07
TG(14:0_32:2)
Low
High
Low
High

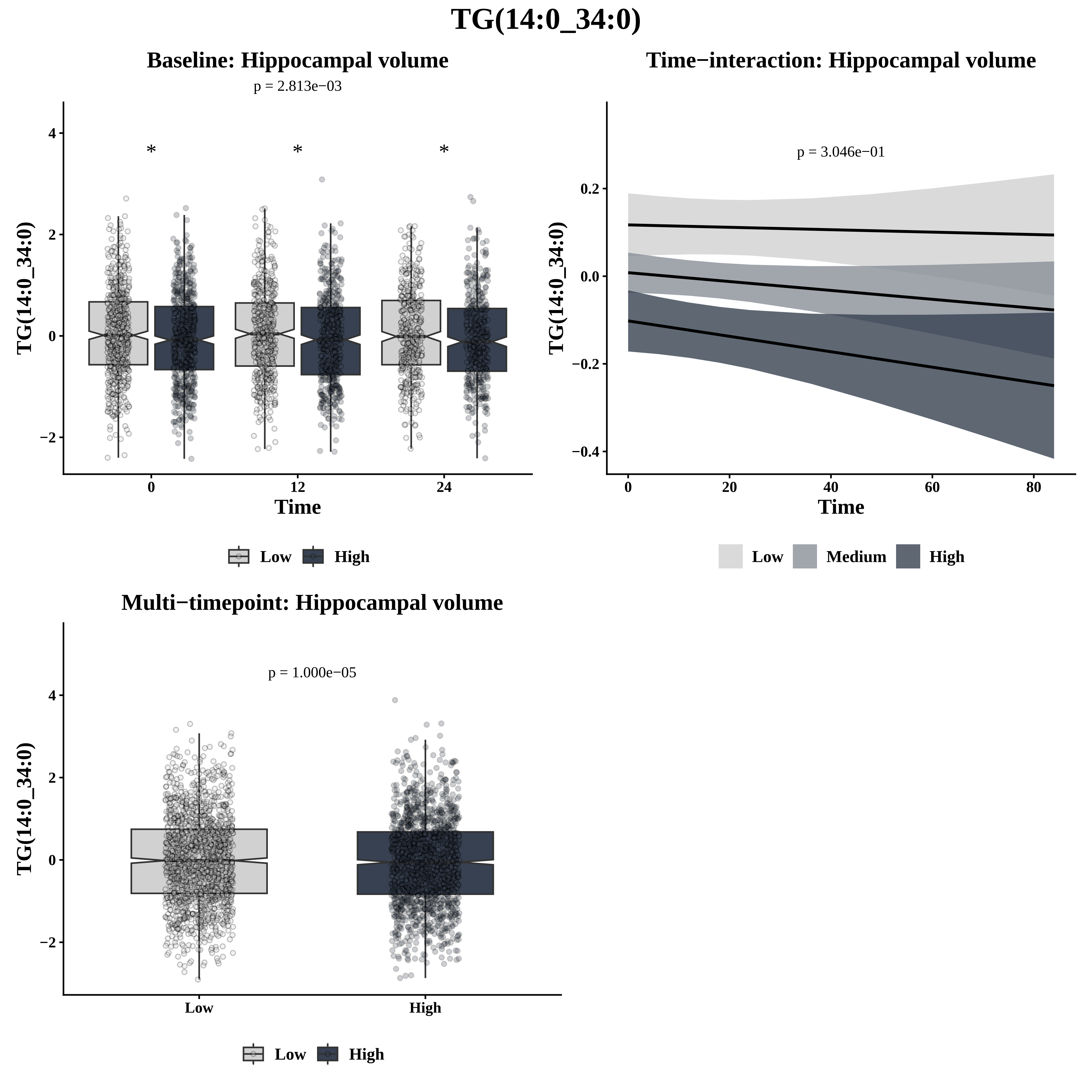

TG(14:0_34:0)
Baseline: Hippocampal volume
p = 2.813e−03
Time−interaction: Hippocampal volume
p = 3.046e−01
Multi−timepoint: Hippocampal volume
p = 1.000e−05
TG(14:0_34:0)
Time
Low
Medium
High

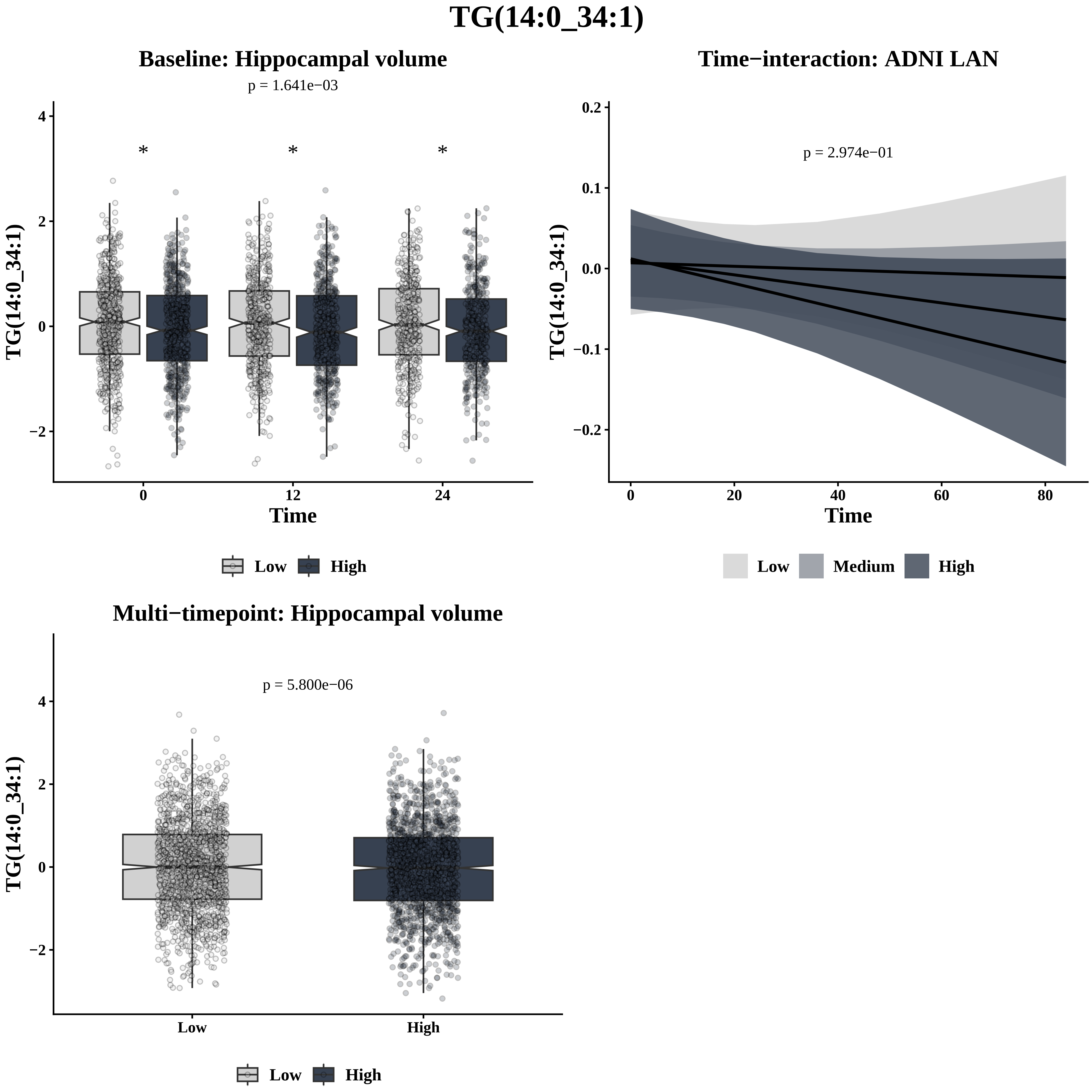
TG(14:0_34:1)
Baseline: Hippocampal volume
p = 1.641e−03
*
*
*
TG(14:0_34:1)
4
2
0
−2
0
12
24
Time
Low
High
Time−interaction: ADNI LAN
p = 2.974e−01
0.2
0.1
0.0
−0.1
−0.2
0
20
40
60
80
Time
Low
Medium
High
Multi−timepoint: Hippocampal volume
p = 5.800e−06
4
2
0
−2
Low
High
Low
High

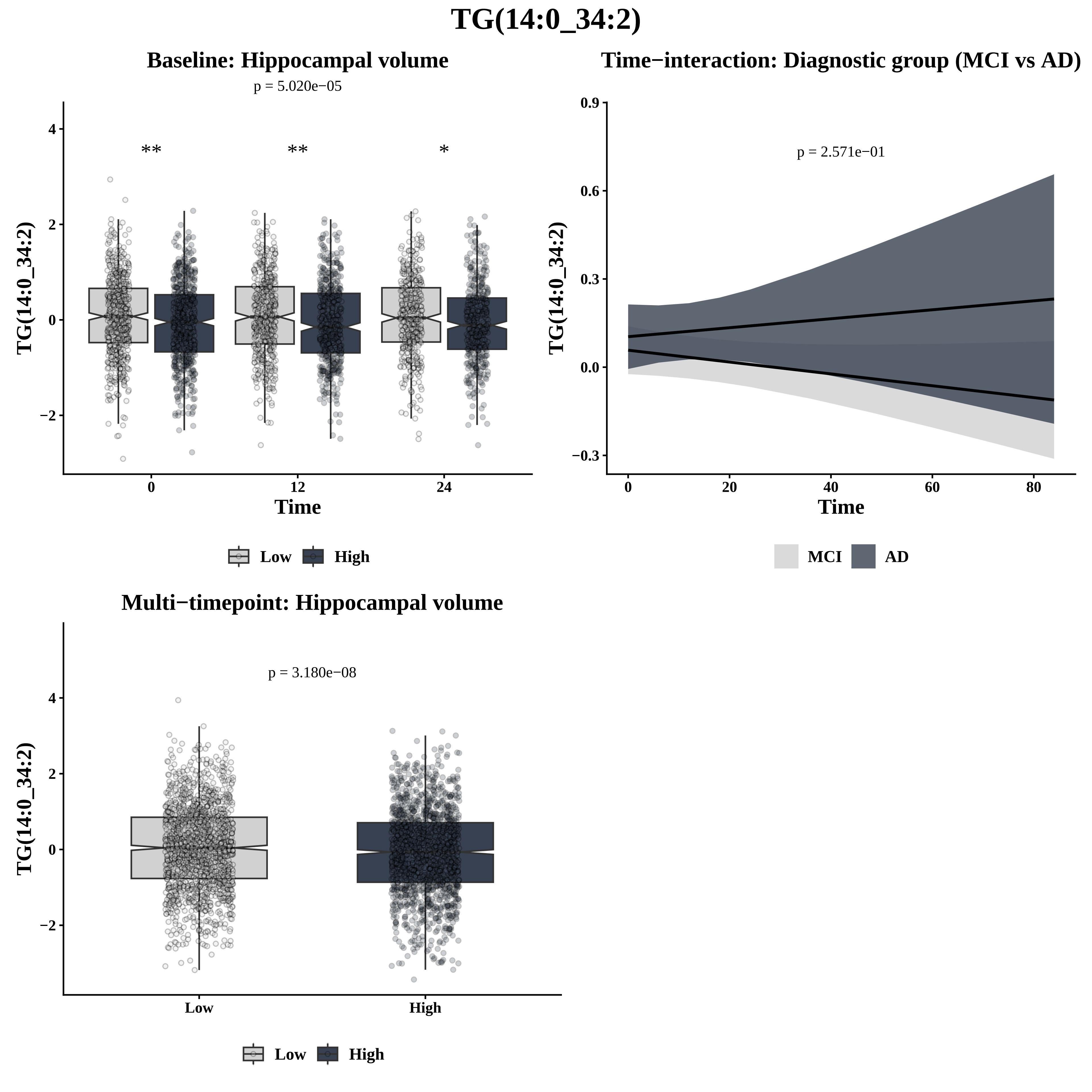

TG(14:0_34:2)
Baseline: Hippocampal volume
p = 5.020e−05
**
**
*
TG(14:0_34:2)
Time
0
12
24
Low
High
Time−interaction: Diagnostic group (MCI vs AD)
p = 2.571e−01
TG(14:0_34:2)
Time
MCI
AD
Multi−timepoint: Hippocampal volume
p = 3.180e−08
TG(14:0_34:2)
Low
High

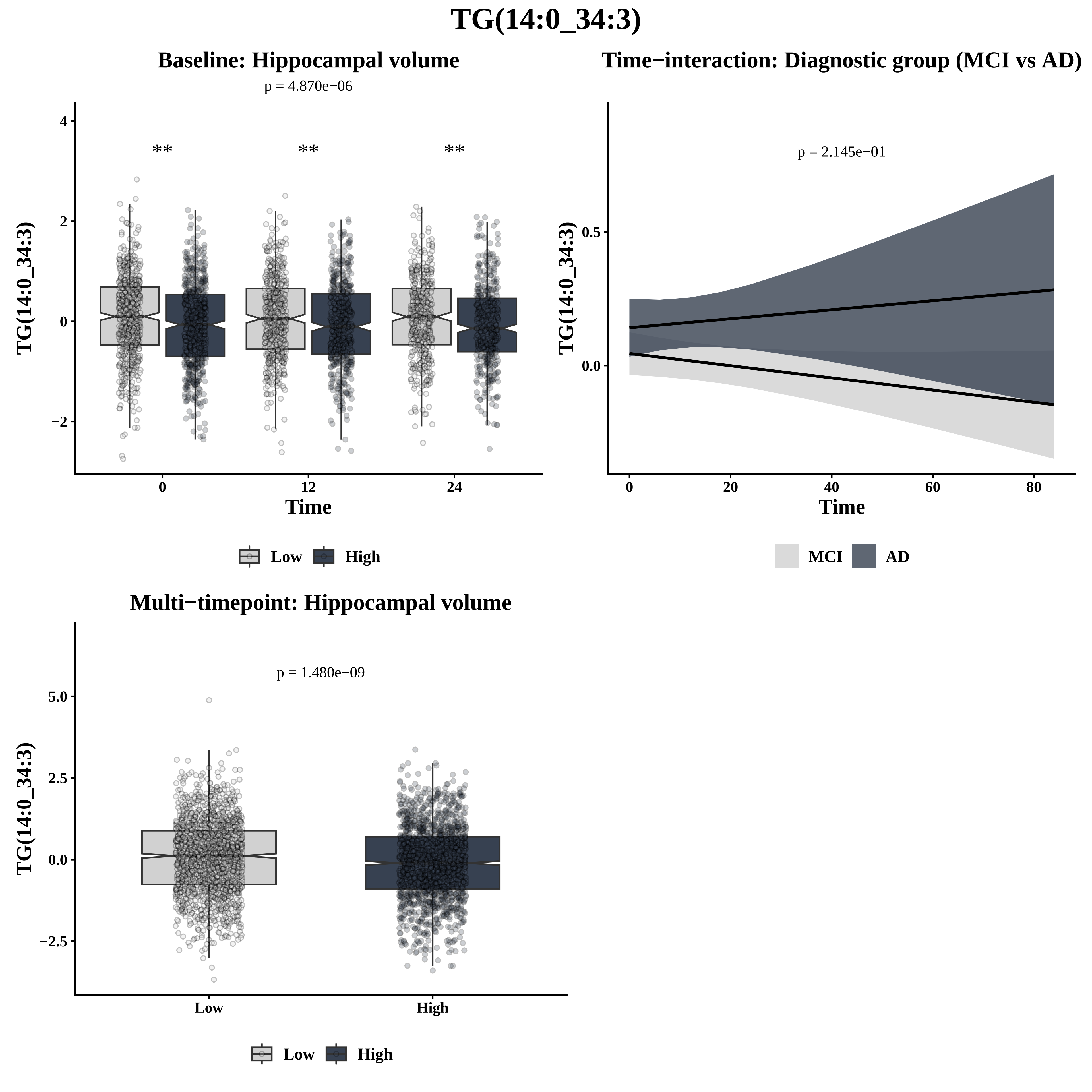

TG(14:0_34:3)
Baseline: Hippocampal volume
p = 4.870e−06
**
**
**
TG(14:0_34:3)
4
2
0
−2
0
12
24
Time
Low
High
Time−interaction: Diagnostic group (MCI vs AD)
p = 2.145e−01
TG(14:0_34:3)
0.5
0.0
0
20
40
60
80
Time
MCI
AD
Multi−timepoint: Hippocampal volume
p = 1.480e−09
TG(14:0_34:3)
5.0
2.5
0.0
−2.5
Low
High
Low
High

# TG(14:0_35:1)

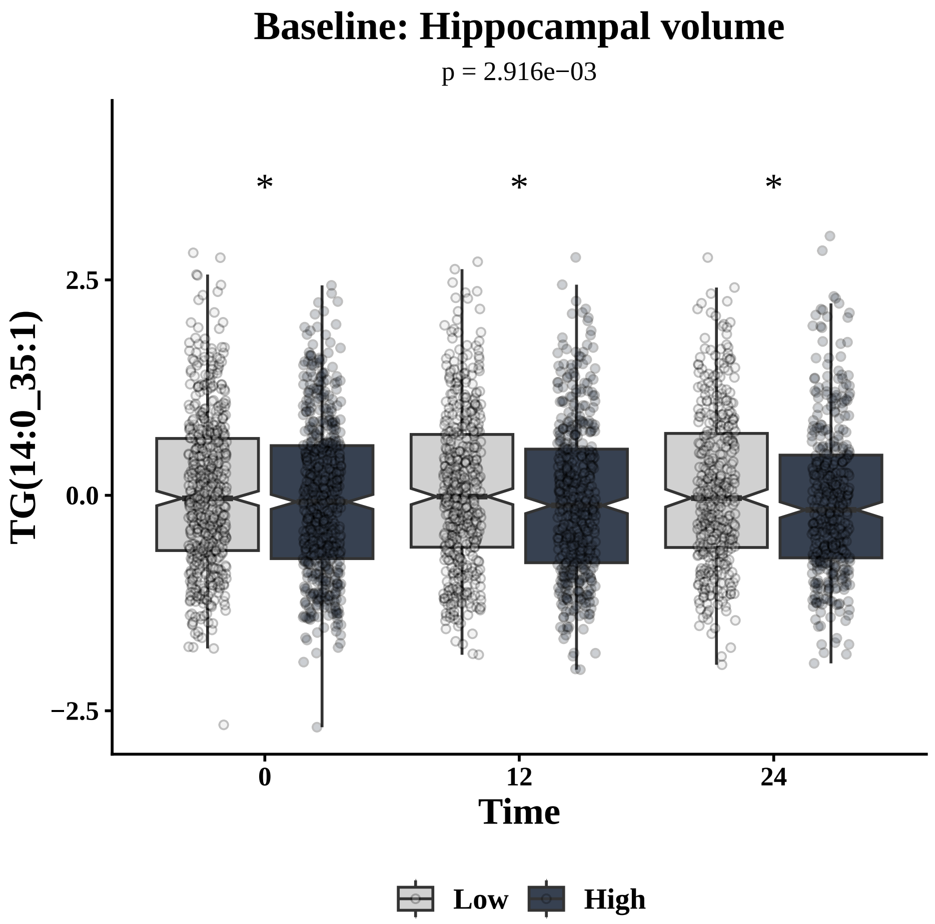
Baseline: Hippocampal volume
p = 2.916e−03
TG(14:0_35:1)
Time
Low
High

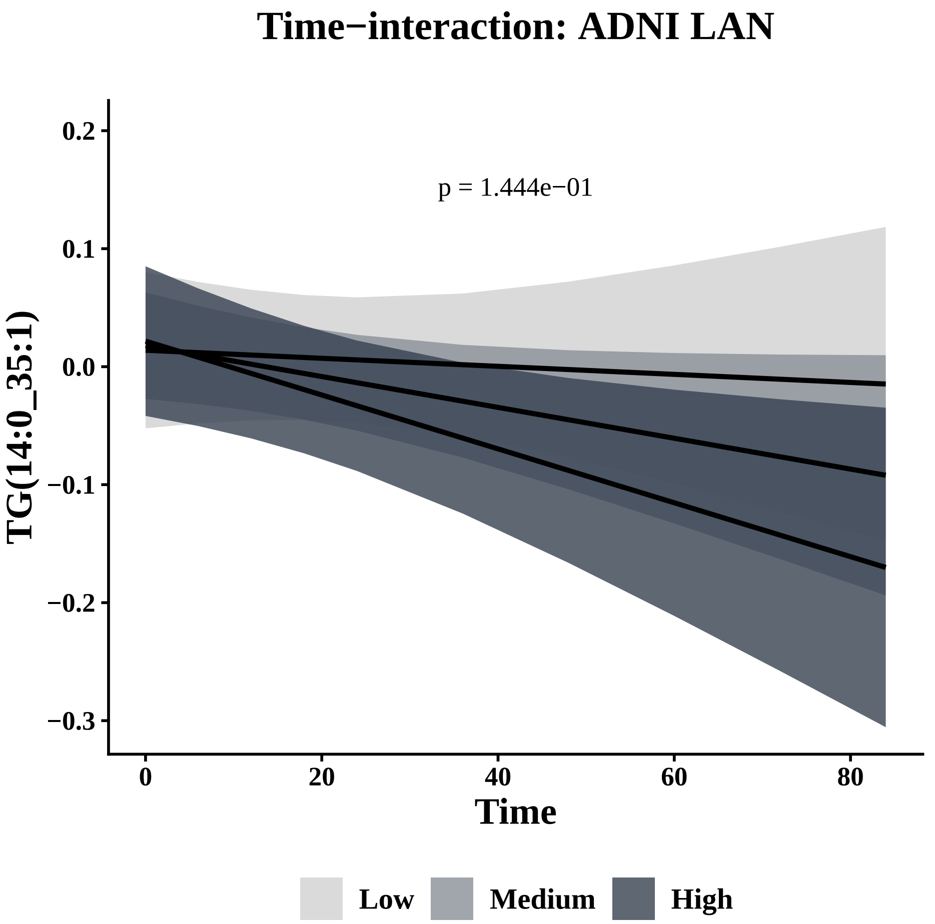
Time−interaction: ADNI LAN
p = 1.444e−01
TG(14:0_35:1)
Time
Low
Medium
High

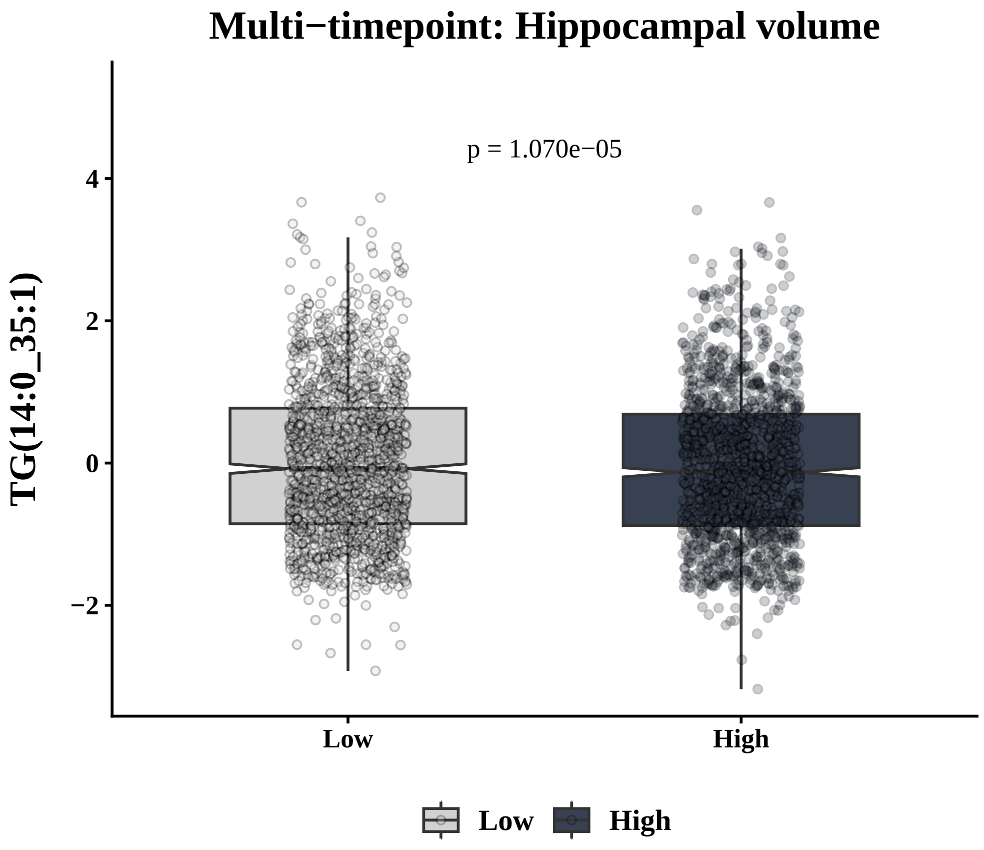
Multi−timepoint: Hippocampal volume
p = 1.070e−05
TG(14:0_35:1)
Low
High

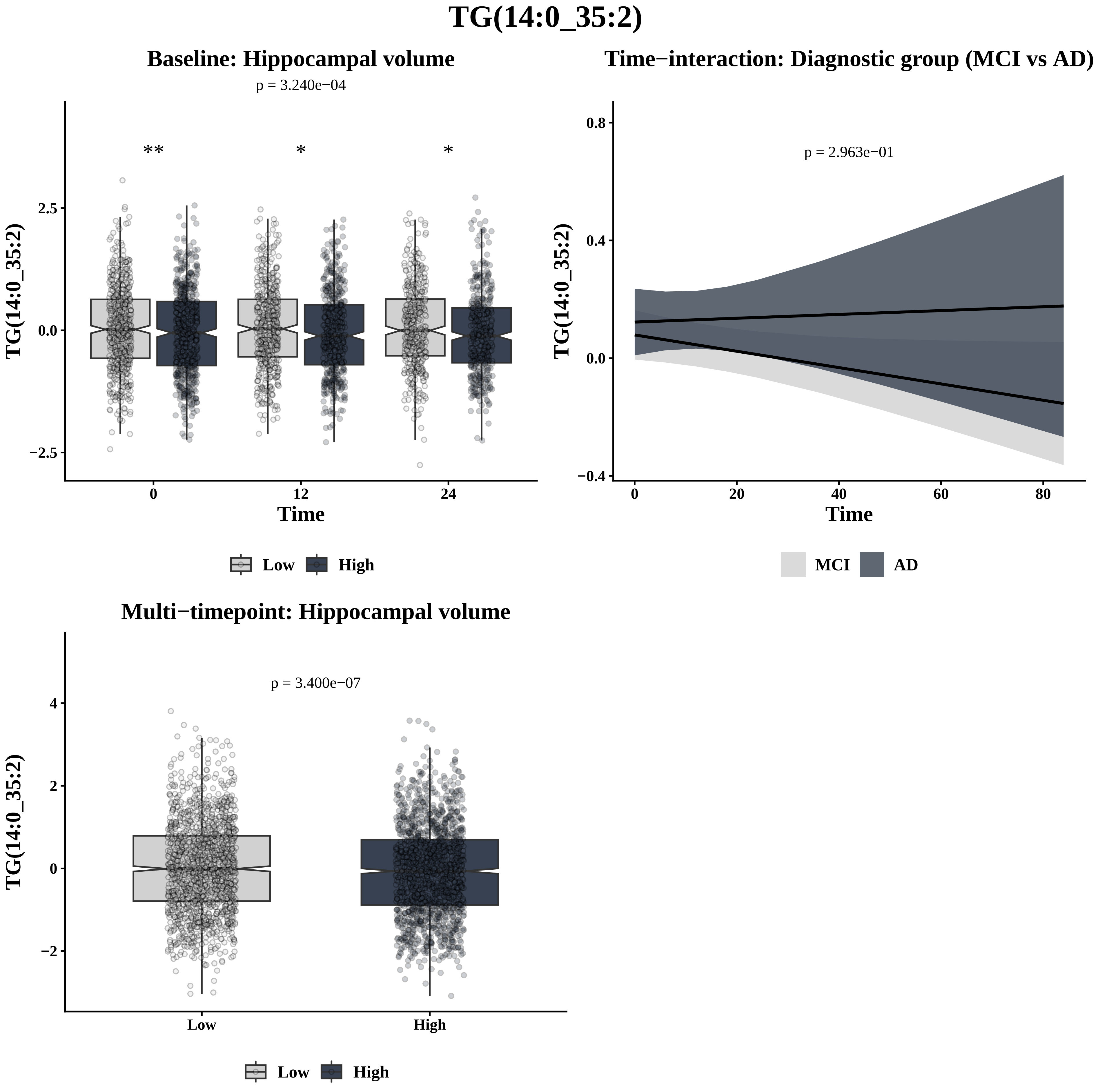

TG(14:0_35:2)
Baseline: Hippocampal volume
p = 3.240e−04
**
*
*
2.5
0.0
−2.5
TG(14:0_35:2)
0
12
24
Time
Low
High
Time−interaction: Diagnostic group (MCI vs AD)
p = 2.963e−01
0.8
0.4
0.0
−0.4
TG(14:0_35:2)
0
20
40
60
80
Time
MCI
AD
Multi−timepoint: Hippocampal volume
p = 3.400e−07
4
2
0
−2
TG(14:0_35:2)
Low
High
Low
High

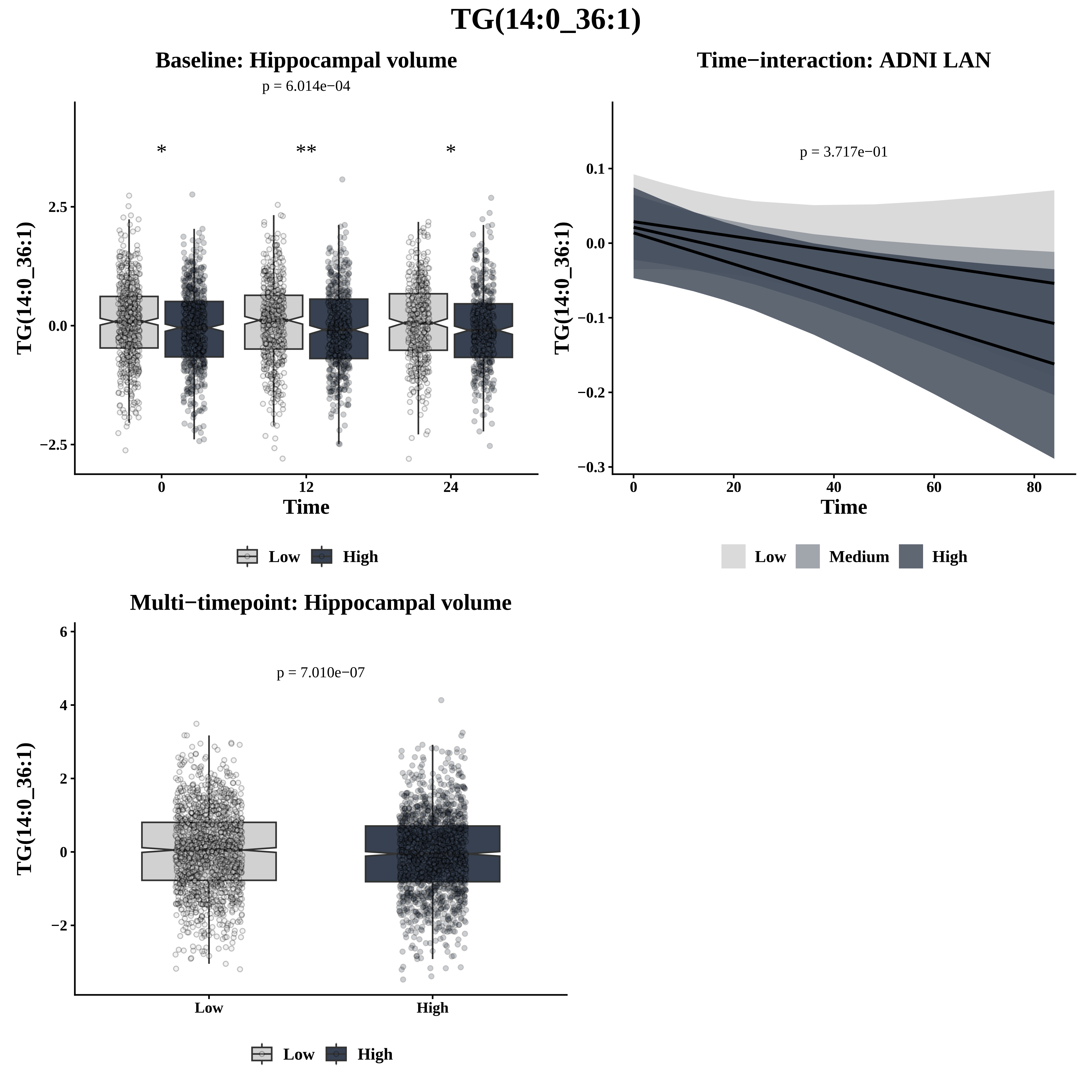
TG(14:0_36:1)
Baseline: Hippocampal volume
p = 6.014e−04
*
**
*
2.5
0.0
−2.5
TG(14:0_36:1)
0
12
24
Time
Low
High
Time−interaction: ADNI LAN
p = 3.717e−01
0.1
0.0
−0.1
−0.2
−0.3
TG(14:0_36:1)
0
20
40
60
80
Time
Low
Medium
High
Multi−timepoint: Hippocampal volume
p = 7.010e−07
6
4
2
0
−2
TG(14:0_36:1)
Low
High
Low
High

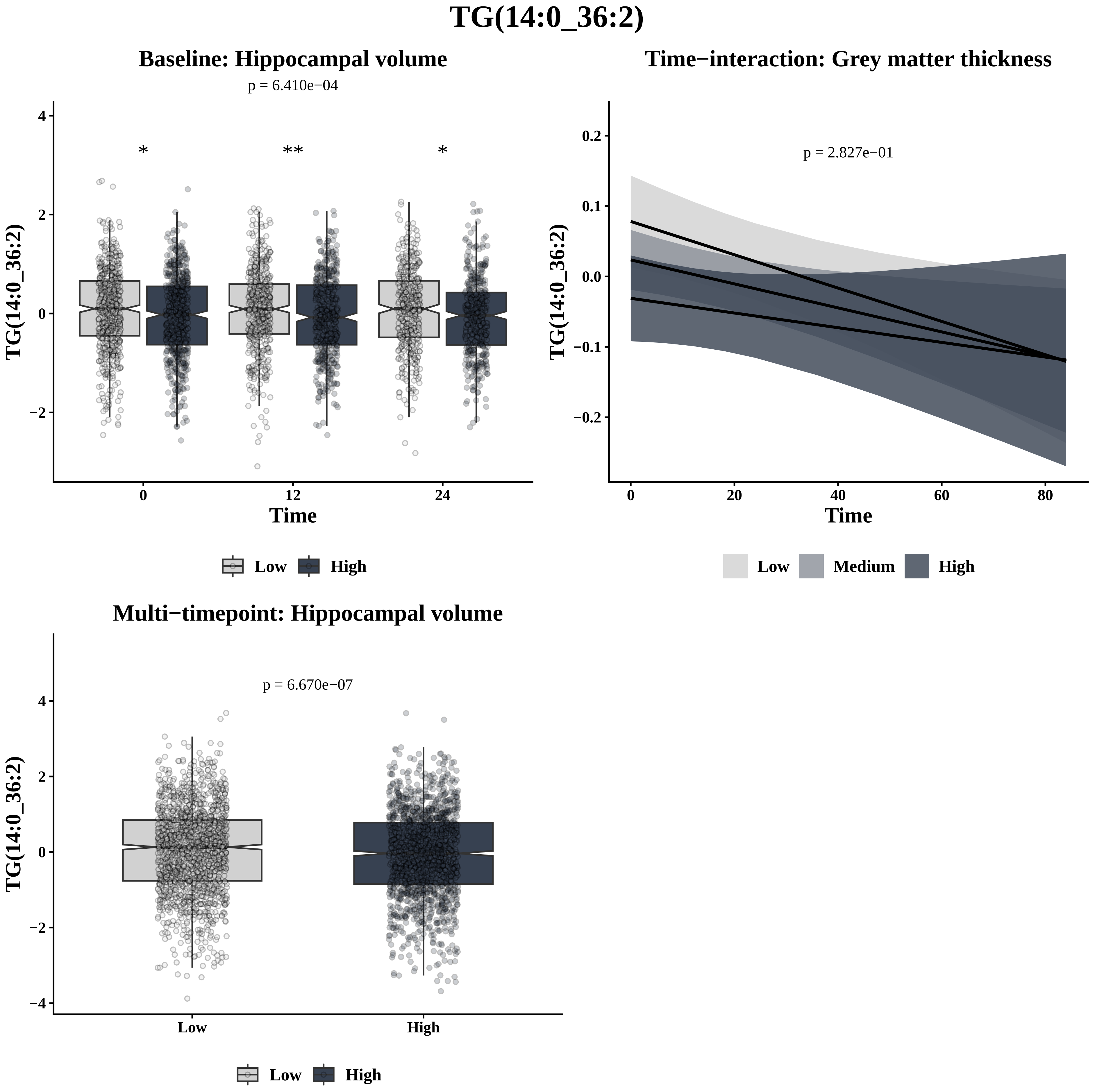
TG(14:0_36:2)
Baseline: Hippocampal volume
p = 6.410e−04
*
**
*
TG(14:0_36:2)
Time
0
12
24
4
2
0
−2
Low
High
Time−interaction: Grey matter thickness
p = 2.827e−01
TG(14:0_36:2)
Time
0.2
0.1
0.0
−0.1
−0.2
0
20
40
60
80
Low
Medium
High
Multi−timepoint: Hippocampal volume
p = 6.670e−07
TG(14:0_36:2)
4
2
0
−2
−4
Low
High
Low
High

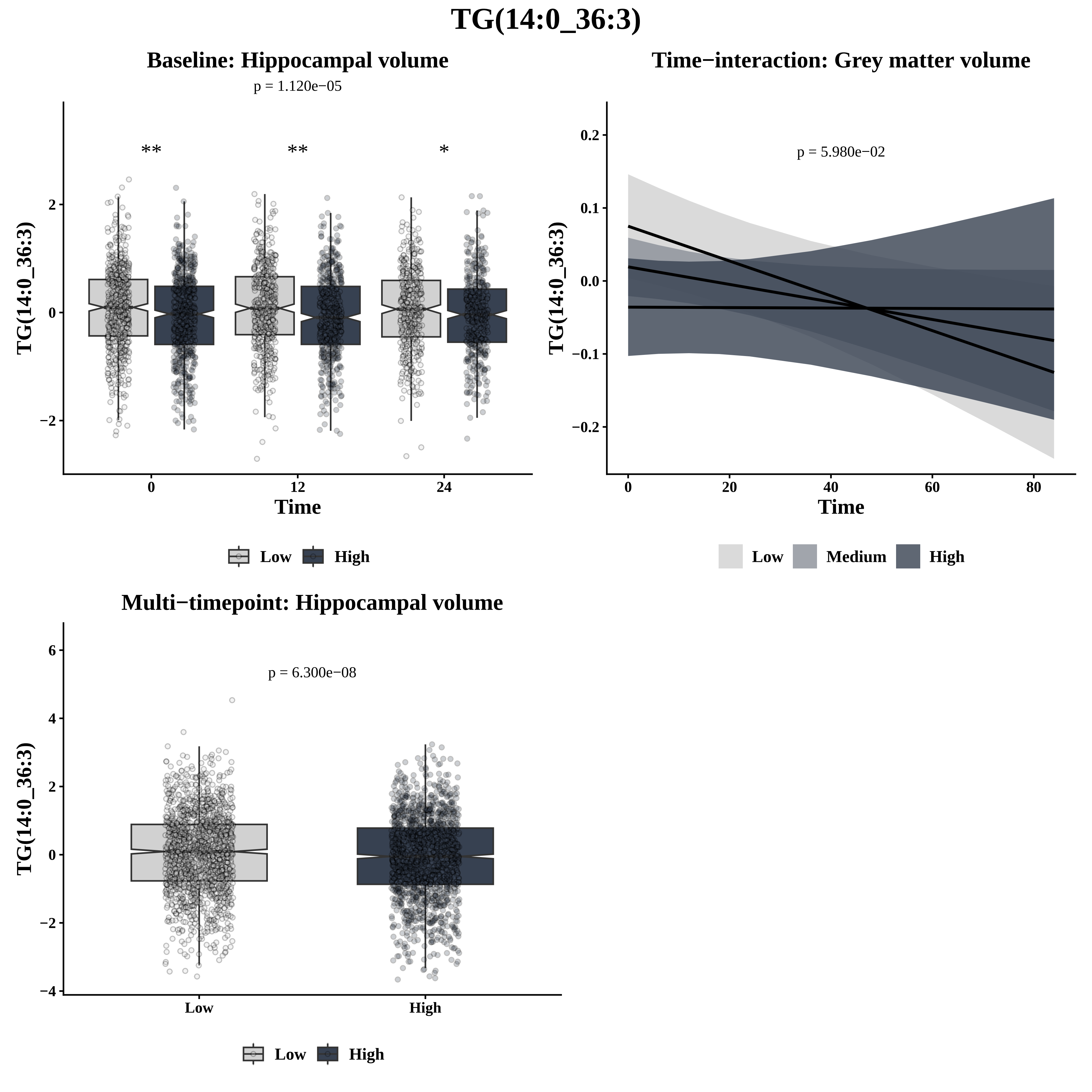

TG(14:0_36:3)
Baseline: Hippocampal volume
p = 1.120e−05
** ** *
TG(14:0_36:3)
2
0
−2
0
12
24
Time
Low
High
Time−interaction: Grey matter volume
p = 5.980e−02
0.2
0.1
0.0
−0.1
−0.2
0
20
40
60
80
Time
Low
Medium
High
Multi−timepoint: Hippocampal volume
p = 6.300e−08
6
4
2
0
−2
−4
Low
High

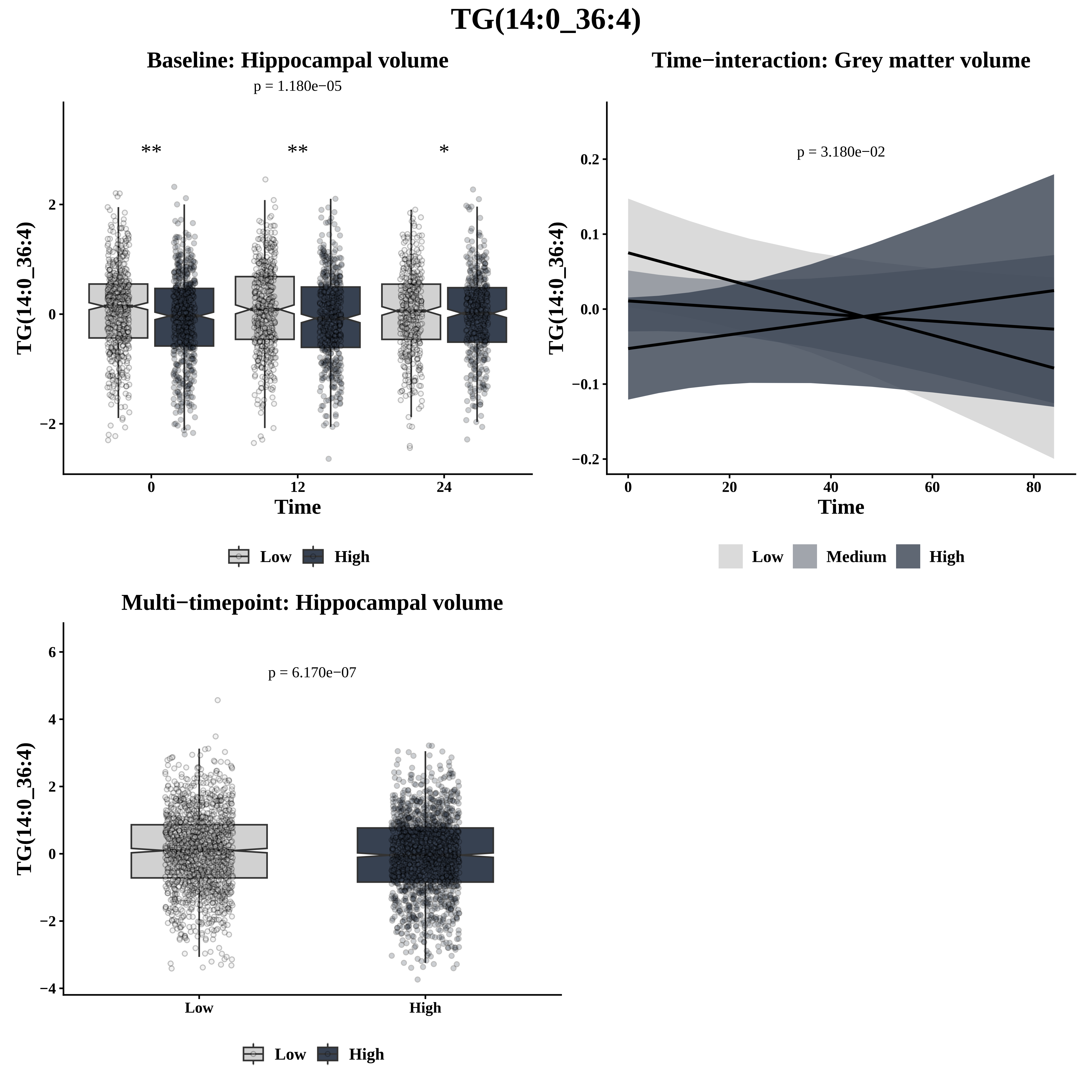

TG(14:0_36:4)
Baseline: Hippocampal volume
p = 1.180e−05
**
**
*
TG(14:0_36:4)
2
0
−2
0
12
24
Time
Low
High
Time−interaction: Grey matter volume
p = 3.180e−02
0.2
0.1
0.0
−0.1
−0.2
0
20
40
60
80
Time
Low
Medium
High
Multi−timepoint: Hippocampal volume
p = 6.170e−07
6
4
2
0
−2
−4
Low
High
Low
High

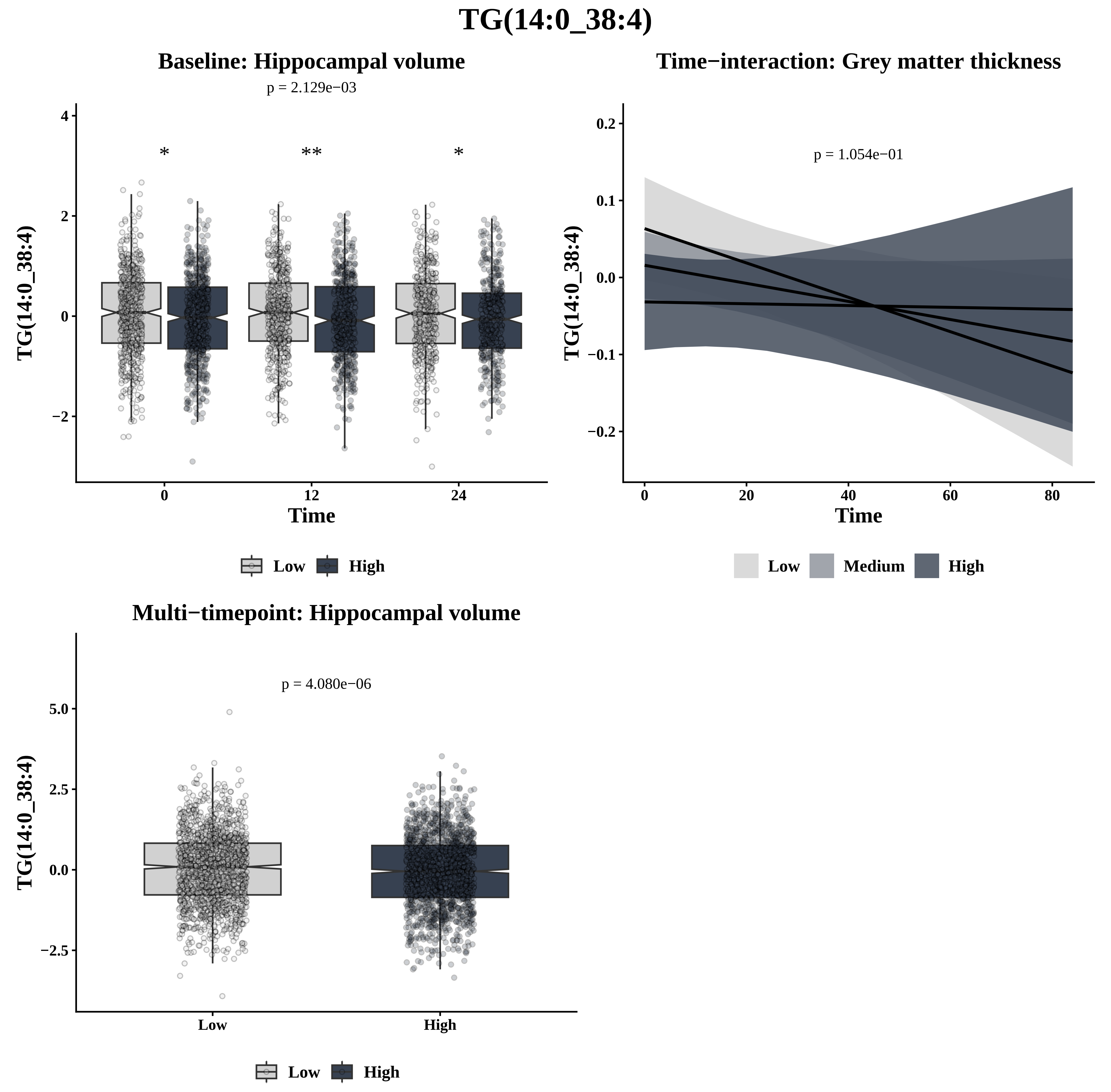

TG(14:0_38:4)
Baseline: Hippocampal volume
p = 2.129e−03
*
**
*
4
2
0
−2
TG(14:0_38:4)
0
12
24
Time
Low
High
Time−interaction: Grey matter thickness
p = 1.054e−01
0.2
0.1
0.0
−0.1
−0.2
TG(14:0_38:4)
0
20
40
60
80
Time
Low
Medium
High
Multi−timepoint: Hippocampal volume
p = 4.080e−06
5.0
2.5
0.0
−2.5
TG(14:0_38:4)
Low
High
Low
High

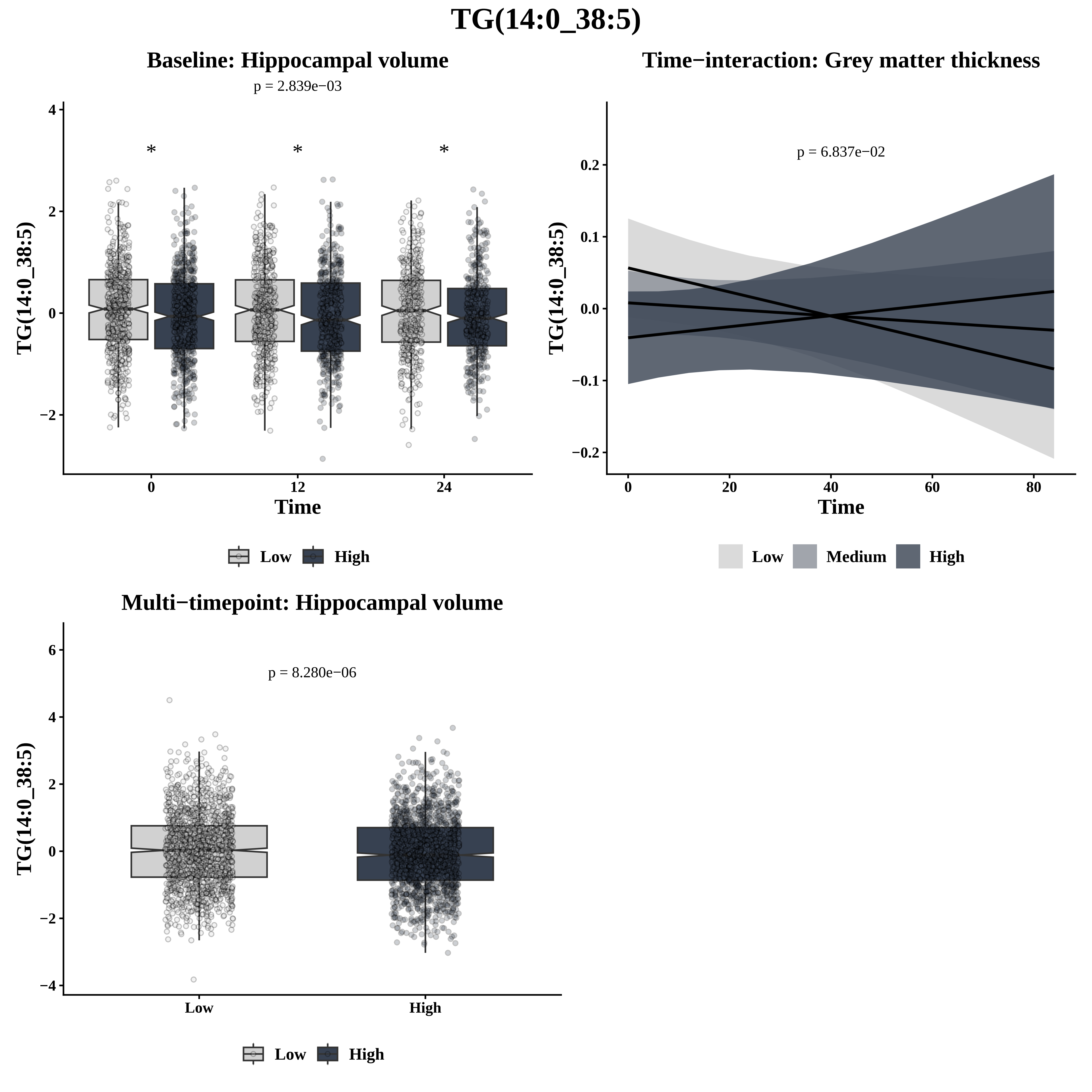

TG(14:0_38:5)
Baseline: Hippocampal volume
p = 2.839e−03
TG(14:0_38:5)
Time
Low
High
Time−interaction: Grey matter thickness
p = 6.837e−02
TG(14:0_38:5)
Time
Low
Medium
High
Multi−timepoint: Hippocampal volume
p = 8.280e−06
TG(14:0_38:5)
Low
High

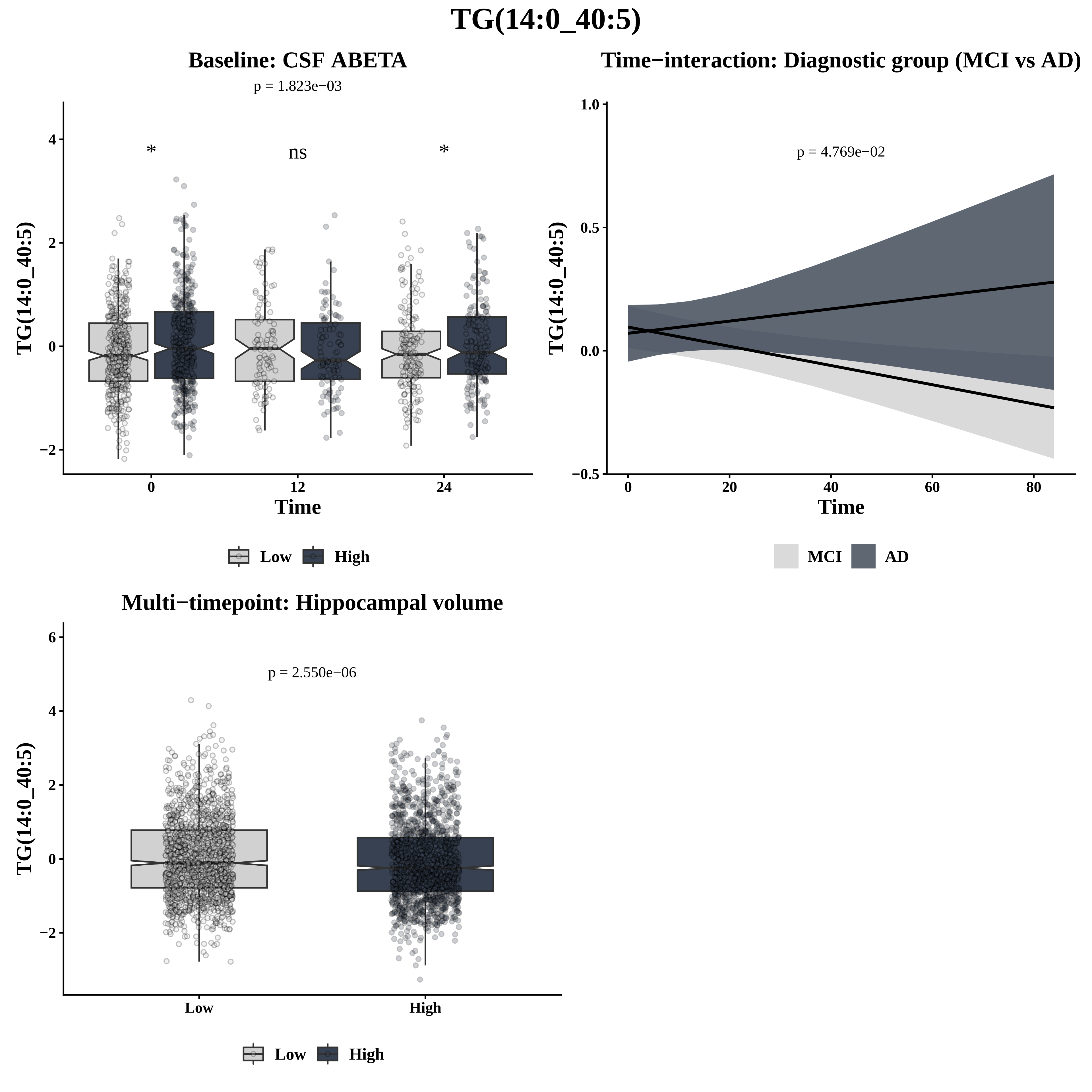
TG(14:0_40:5)
Baseline: CSF ABETA
p = 1.823e−03
*
ns
*
TG(14:0_40:5)
Time
Low
High
Time−interaction: Diagnostic group (MCI vs AD)
p = 4.769e−02
TG(14:0_40:5)
Time
MCI
AD
Multi−timepoint: Hippocampal volume
p = 2.550e−06
TG(14:0_40:5)
Low
High
Low
High

# TG(16:0_28:1)

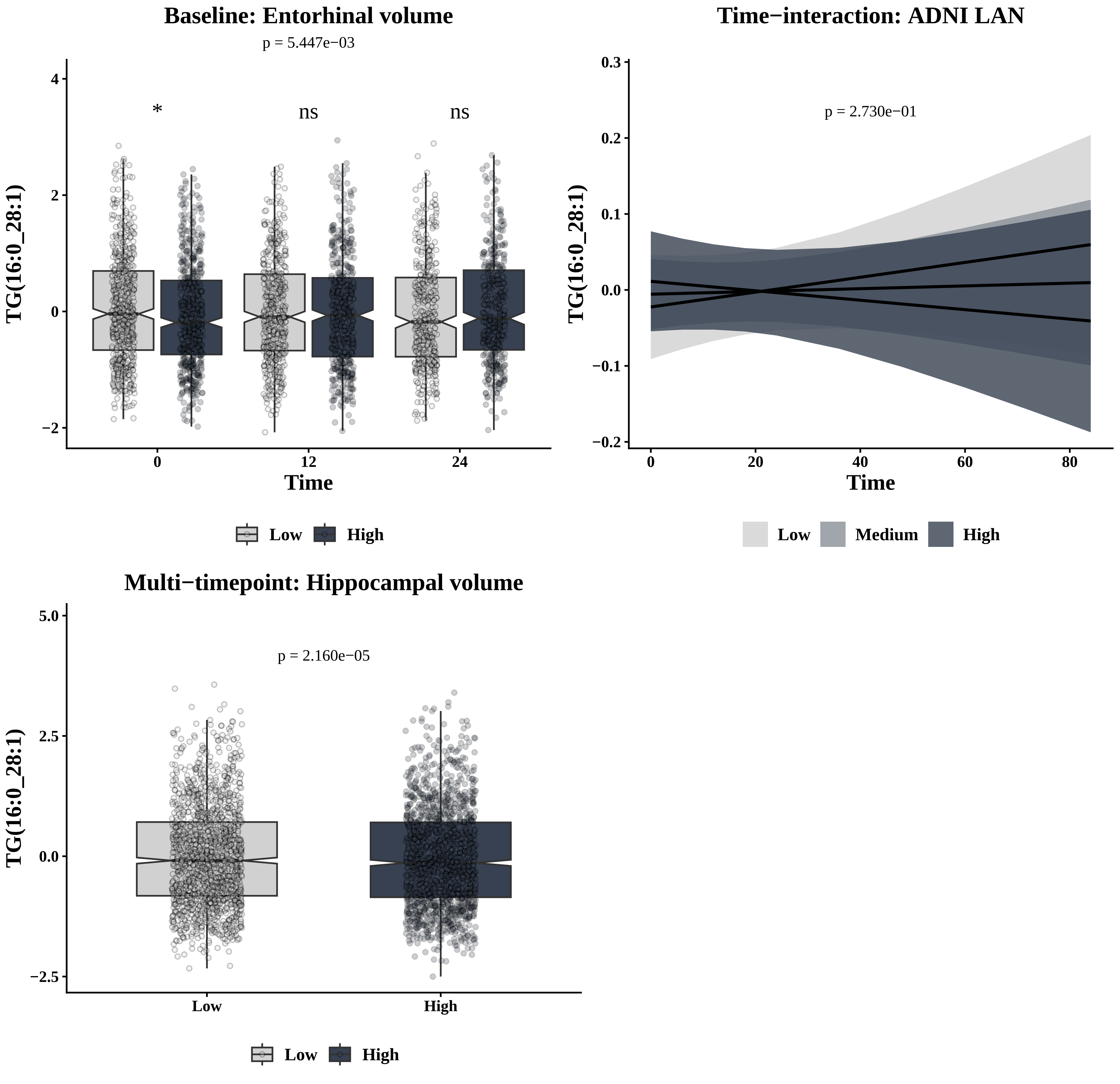

Baseline: Entorhinal volume
p = 5.447e−03
*
ns
ns
TG(16:0_28:1)
Time
0
12
24
4
2
0
−2
Low
High
Time−interaction: ADNI LAN
p = 2.730e−01
TG(16:0_28:1)
Time
0
20
40
60
80
0.3
0.2
0.1
0.0
−0.1
−0.2
Low
Medium
High
Multi−timepoint: Hippocampal volume
p = 2.160e−05
TG(16:0_28:1)
5.0
2.5
0.0
−2.5
Low
High
Low
High

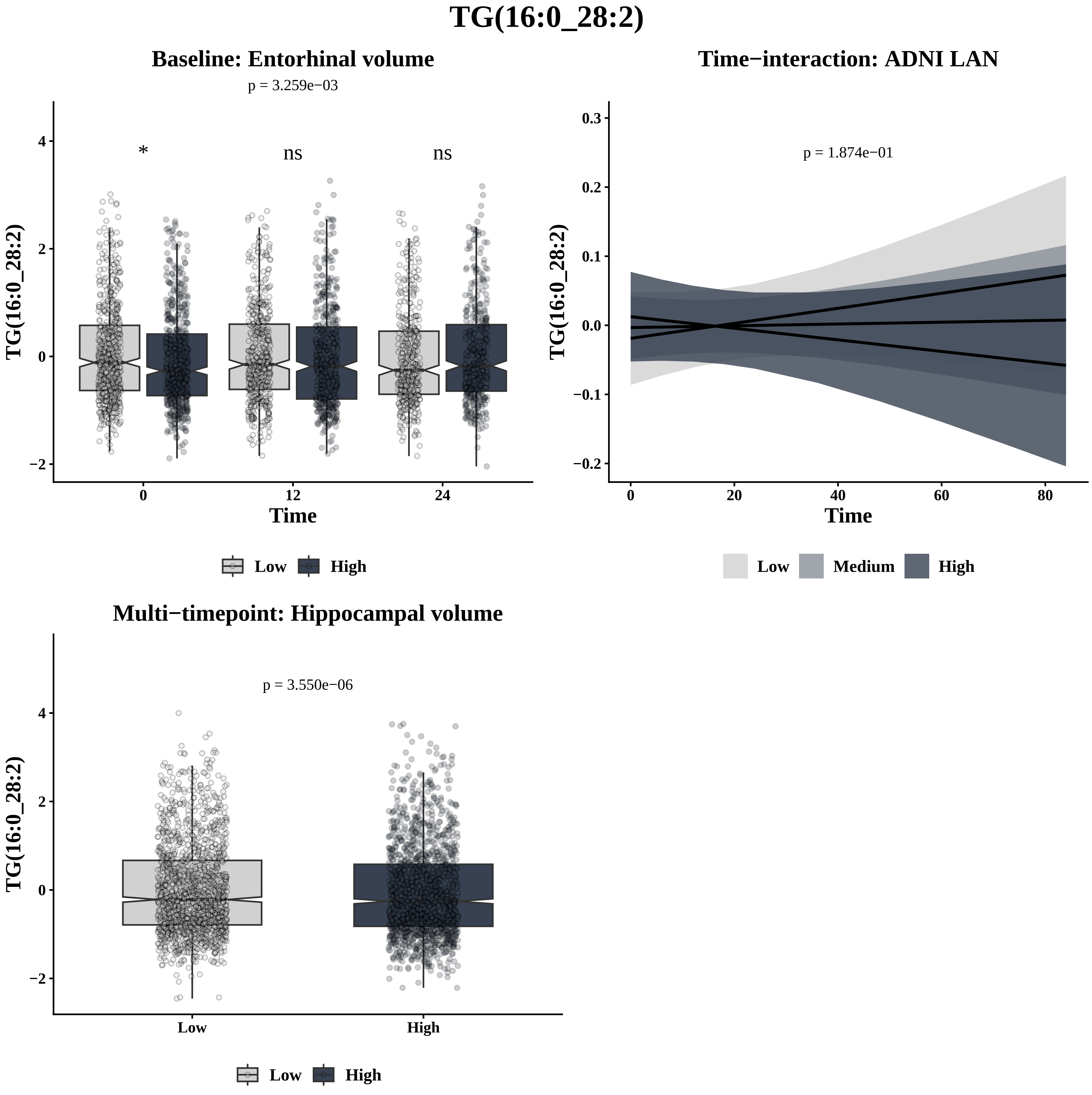
TG(16:0_28:2)
Baseline: Entorhinal volume
p = 3.259e−03
*
ns
ns
TG(16:0_28:2)
4
2
0
−2
0
12
24
Time
Low
High
Time−interaction: ADNI LAN
p = 1.874e−01
TG(16:0_28:2)
0.3
0.2
0.1
0.0
−0.1
−0.2
0
20
40
60
80
Time
Low
Medium
High
Multi−timepoint: Hippocampal volume
p = 3.550e−06
TG(16:0_28:2)
4
2
0
−2
Low
High
Low
High

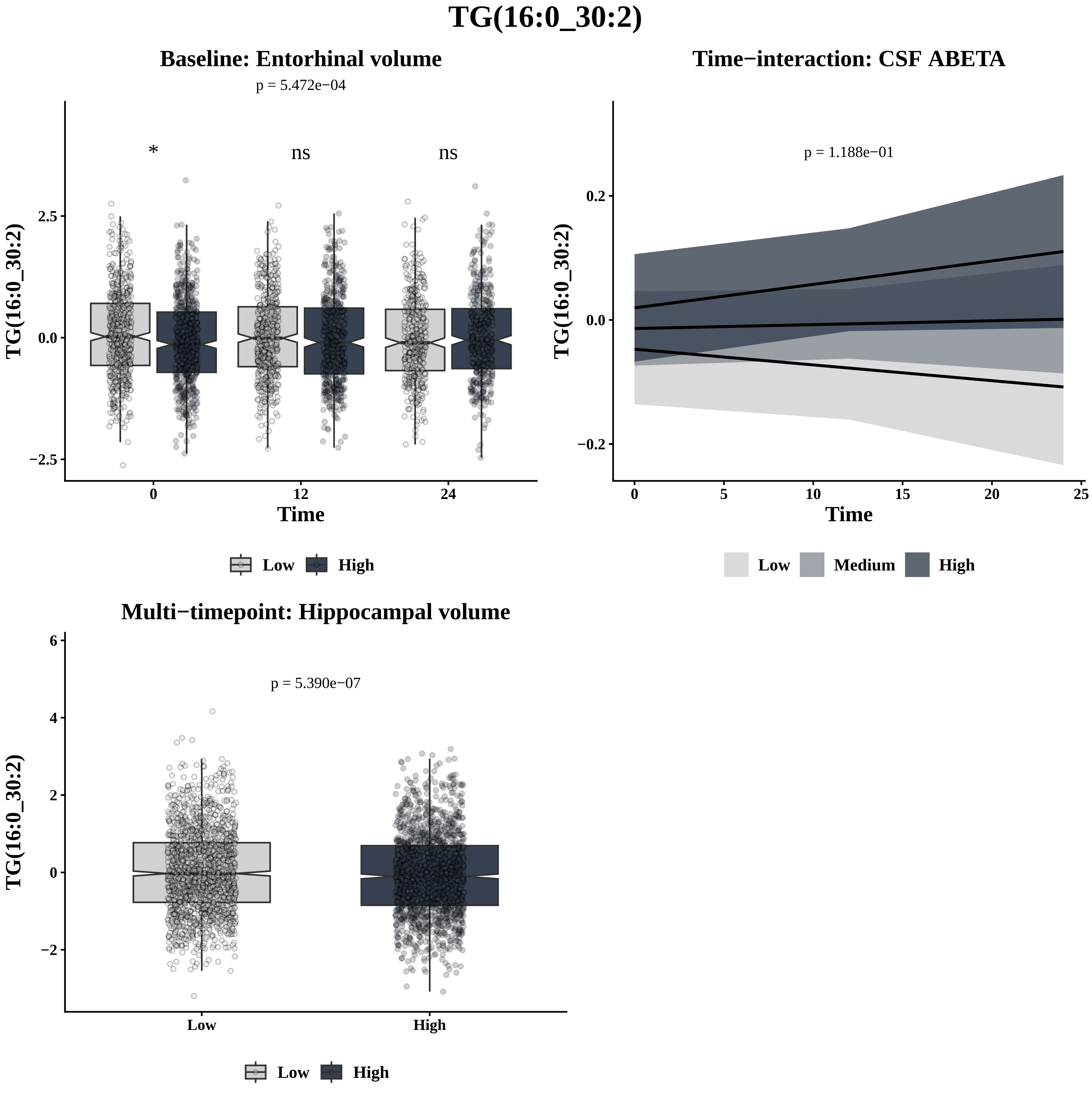

TG(16:0_30:2)
Baseline: Entorhinal volume
p = 5.472e−04
*
ns
ns
2.5
0.0
−2.5
TG(16:0_30:2)
0
12
24
Time
Low
High
Time−interaction: CSF ABETA
p = 1.188e−01
0.2
0.0
−0.2
TG(16:0_30:2)
0
5
10
15
20
25
Time
Low
Medium
High
Multi−timepoint: Hippocampal volume
p = 5.390e−07
6
4
2
0
−2
TG(16:0_30:2)
Low
High
Low
High

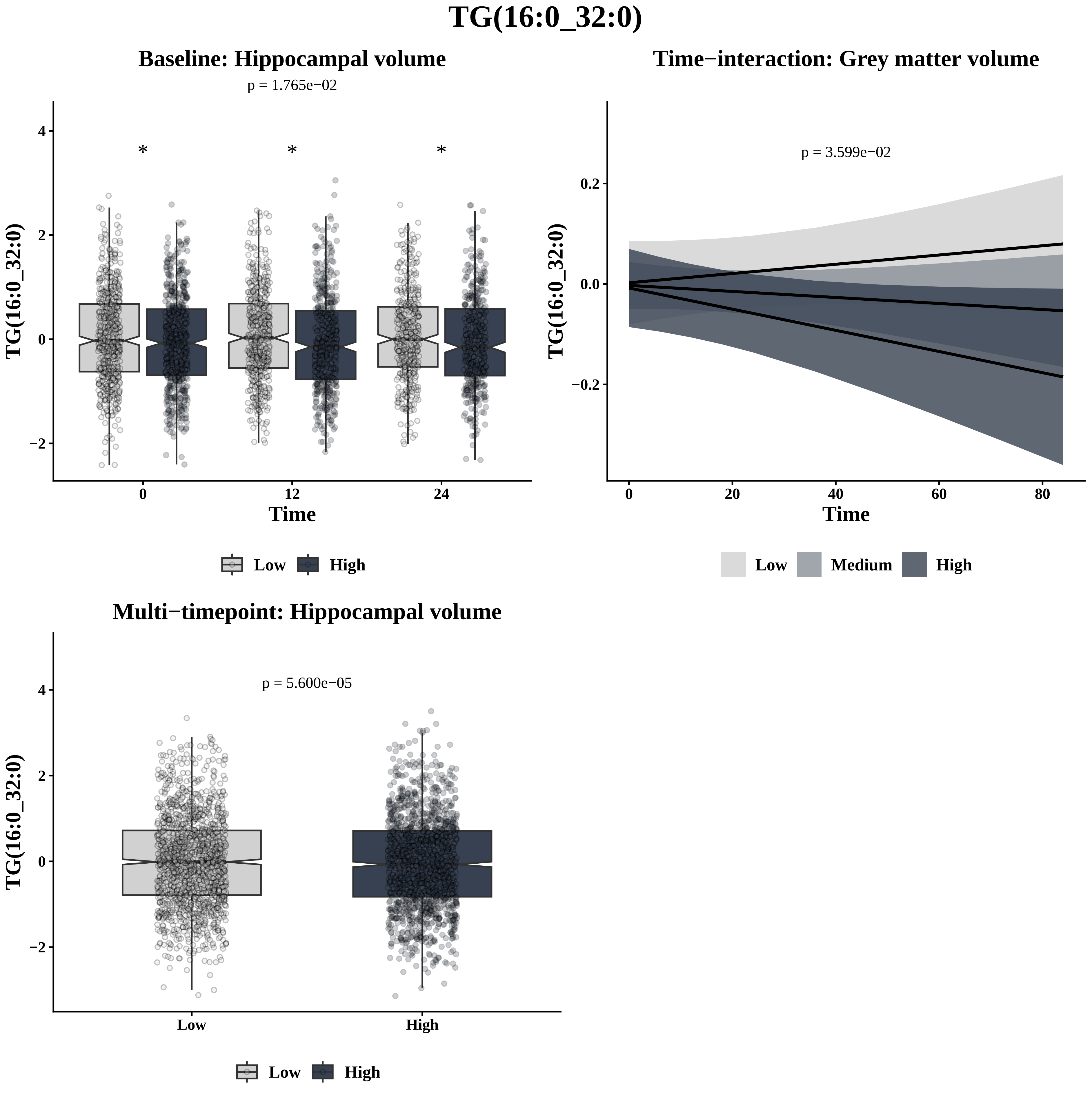
TG(16:0_32:0)
Baseline: Hippocampal volume
p = 1.765e−02
*
*
*
TG(16:0_32:0)
4
2
0
−2
0
12
24
Time
Low
High
Time−interaction: Grey matter volume
p = 3.599e−02
TG(16:0_32:0)
0.2
0.0
−0.2
0
20
40
60
80
Time
Low
Medium
High
Multi−timepoint: Hippocampal volume
p = 5.600e−05
TG(16:0_32:0)
4
2
0
−2
Low
High
Low
High

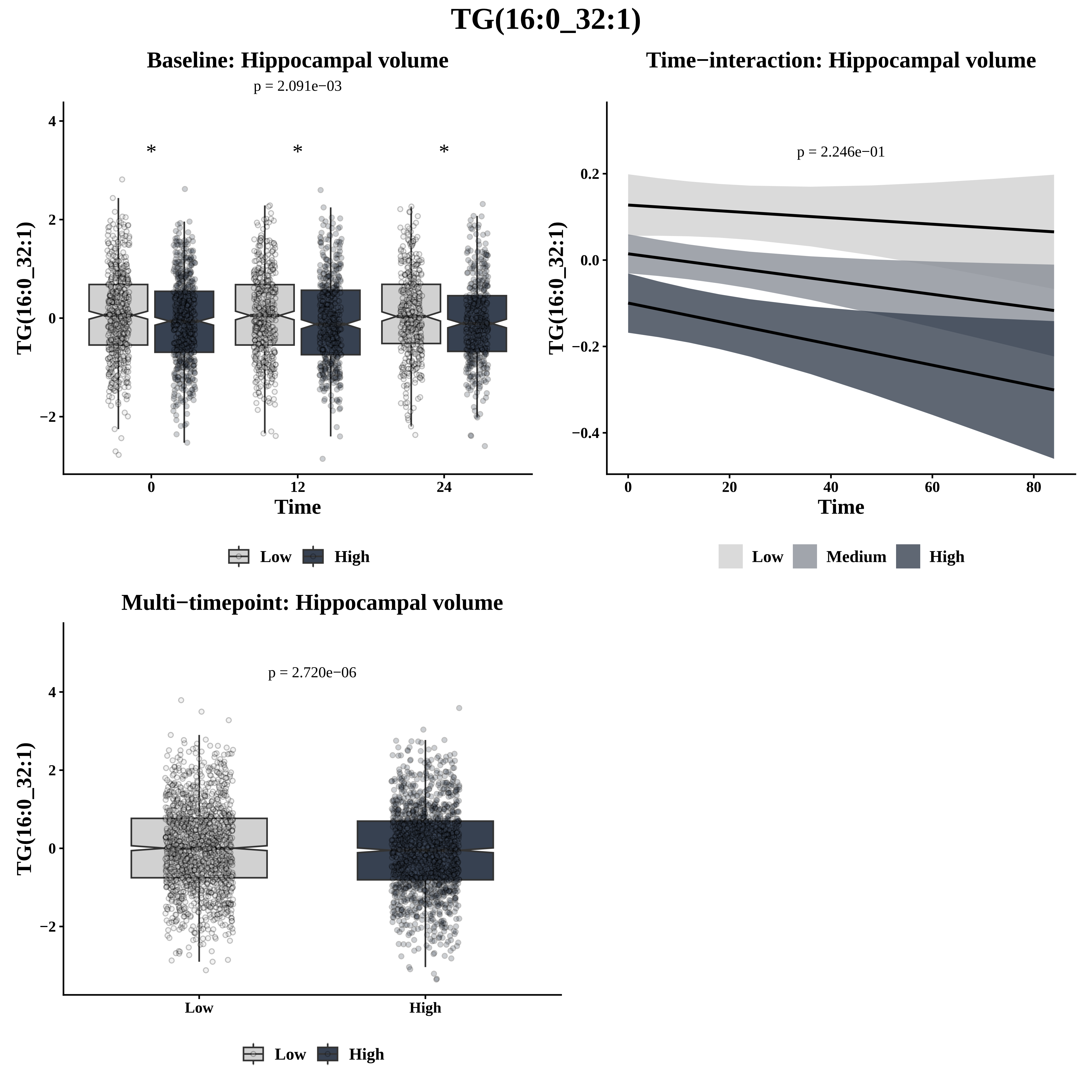

TG(16:0_32:1)
Baseline: Hippocampal volume
p = 2.091e−03
TG(16:0_32:1)
Time
Low
High
Time−interaction: Hippocampal volume
p = 2.246e−01
TG(16:0_32:1)
Time
Low
Medium
High
Multi−timepoint: Hippocampal volume
p = 2.720e−06
TG(16:0_32:1)
Low
High

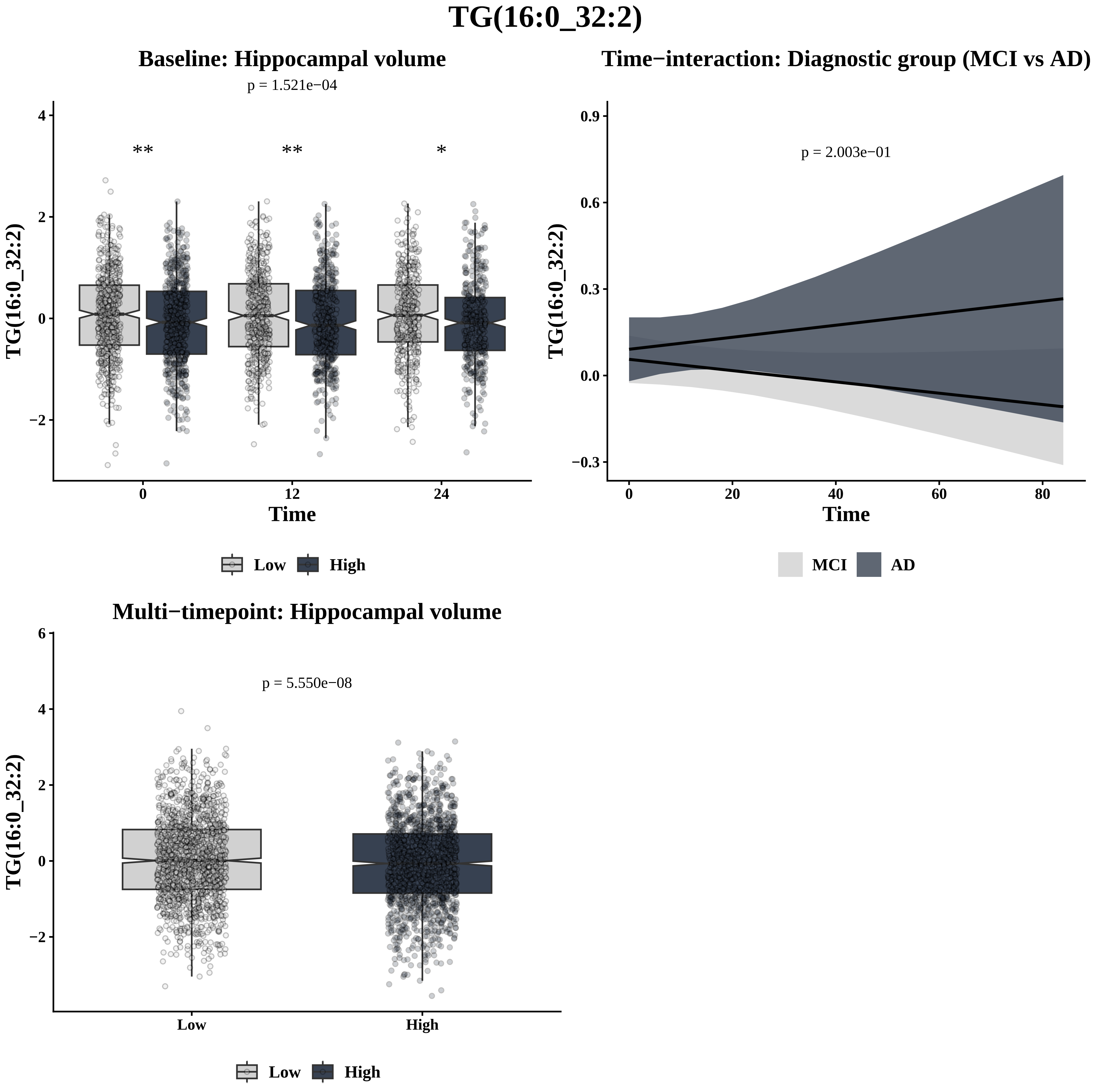

TG(16:0_32:2)
Baseline: Hippocampal volume
p = 1.521e−04
**
**
*
TG(16:0_32:2)
Time
0
12
24
4
2
0
−2
Low
High
Time−interaction: Diagnostic group (MCI vs AD)
p = 2.003e−01
TG(16:0_32:2)
0.9
0.6
0.3
0.0
−0.3
0
20
40
60
80
Time
MCI
AD
Multi−timepoint: Hippocampal volume
p = 5.550e−08
TG(16:0_32:2)
6
4
2
0
−2
Low
High
Low
High

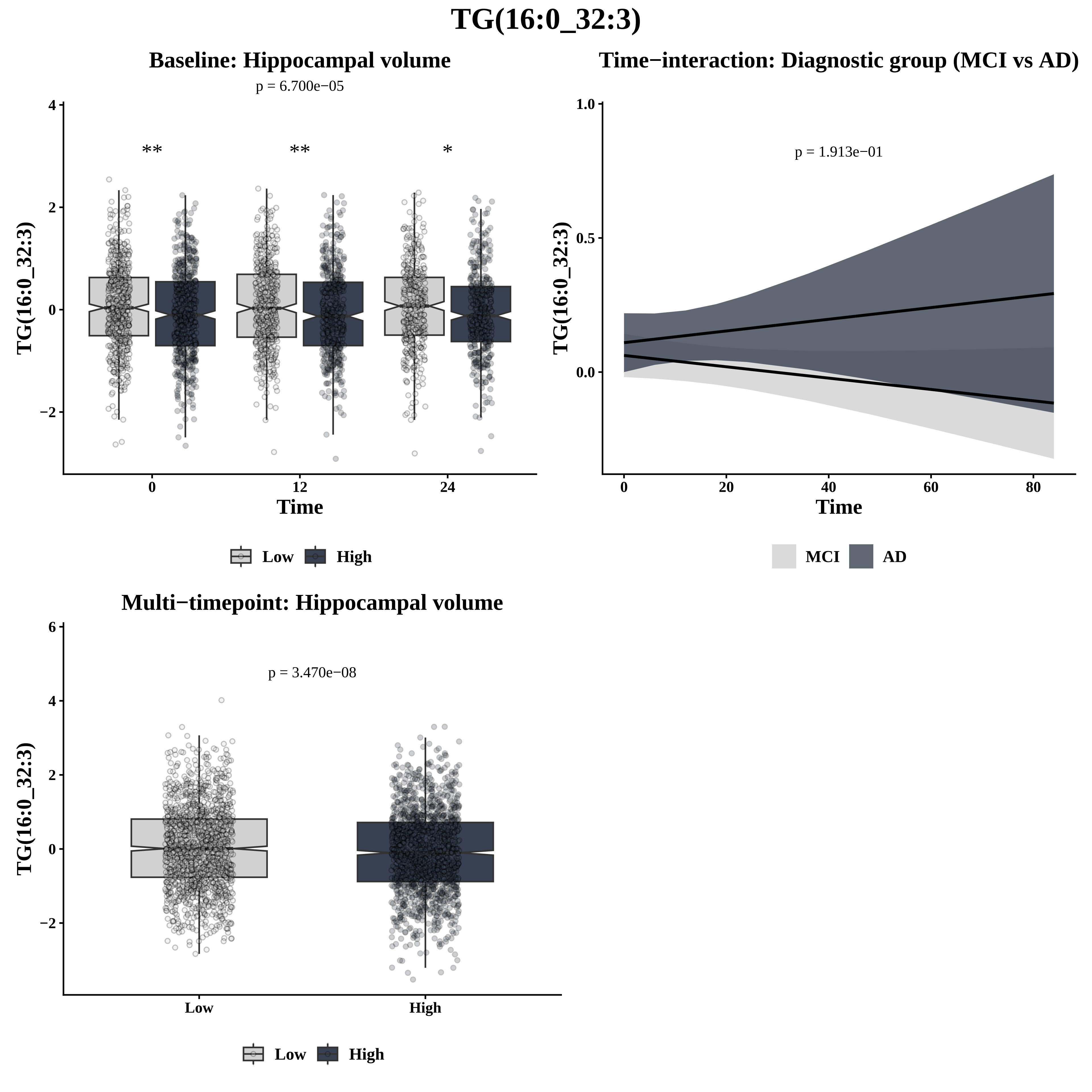
TG(16:0_32:3)
Baseline: Hippocampal volume
p = 6.700e−05
**
**
*
TG(16:0_32:3)
Time
0
12
24
4
2
0
−2
Low
High
Time−interaction: Diagnostic group (MCI vs AD)
p = 1.913e−01
1.0
0.5
0.0
TG(16:0_32:3)
Time
0
20
40
60
80
MCI
AD
Multi−timepoint: Hippocampal volume
p = 3.470e−08
6
4
2
0
−2
TG(16:0_32:3)
Low
High
Low
High

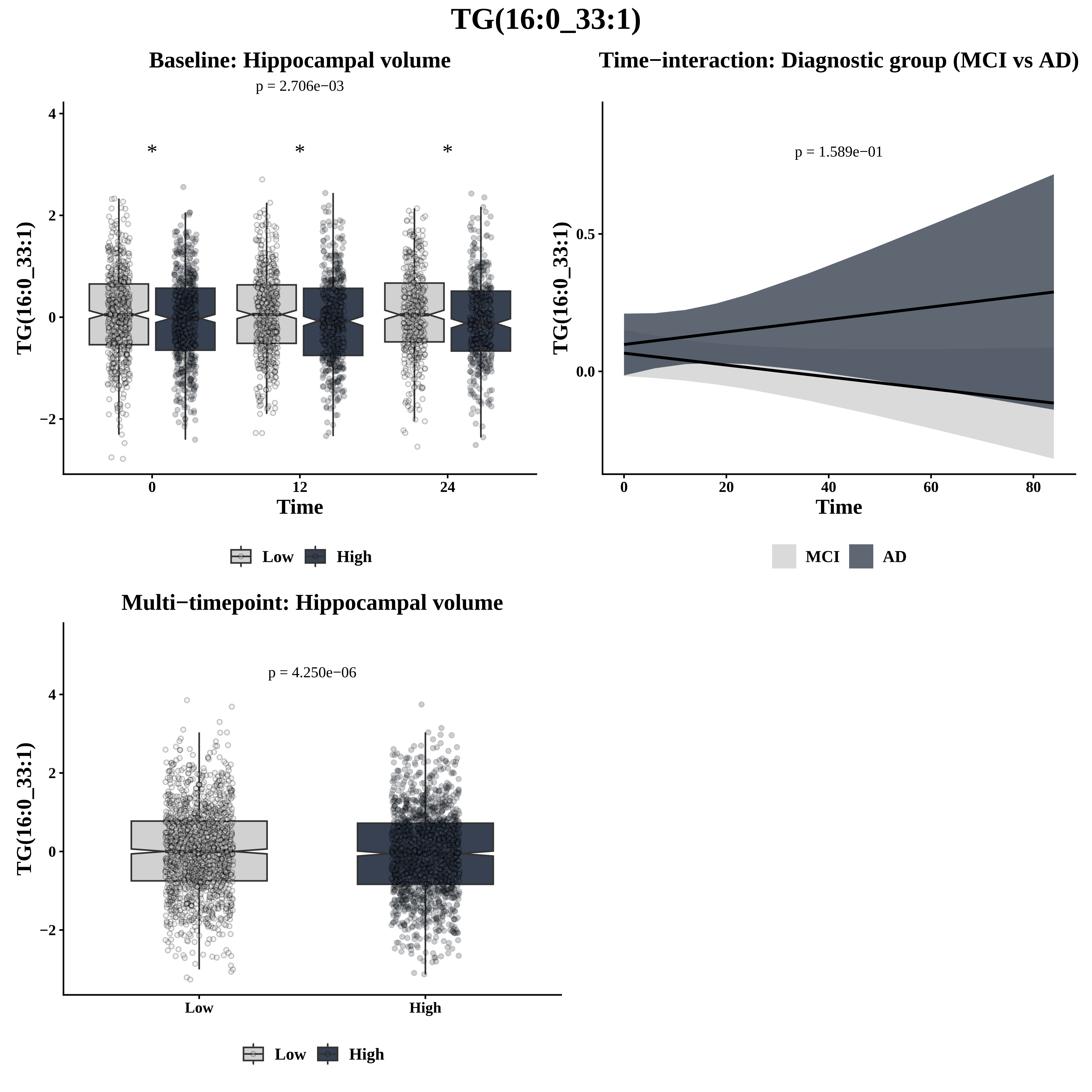

TG(16:0_33:1)
Baseline: Hippocampal volume
p = 2.706e−03
*
*
*
4
2
0
−2
TG(16:0_33:1)
0
12
24
Time
Low
High
Time−interaction: Diagnostic group (MCI vs AD)
p = 1.589e−01
0.5
0.0
TG(16:0_33:1)
0
20
40
60
80
Time
MCI
AD
Multi−timepoint: Hippocampal volume
p = 4.250e−06
4
2
0
−2
TG(16:0_33:1)
Low
High
Low
High

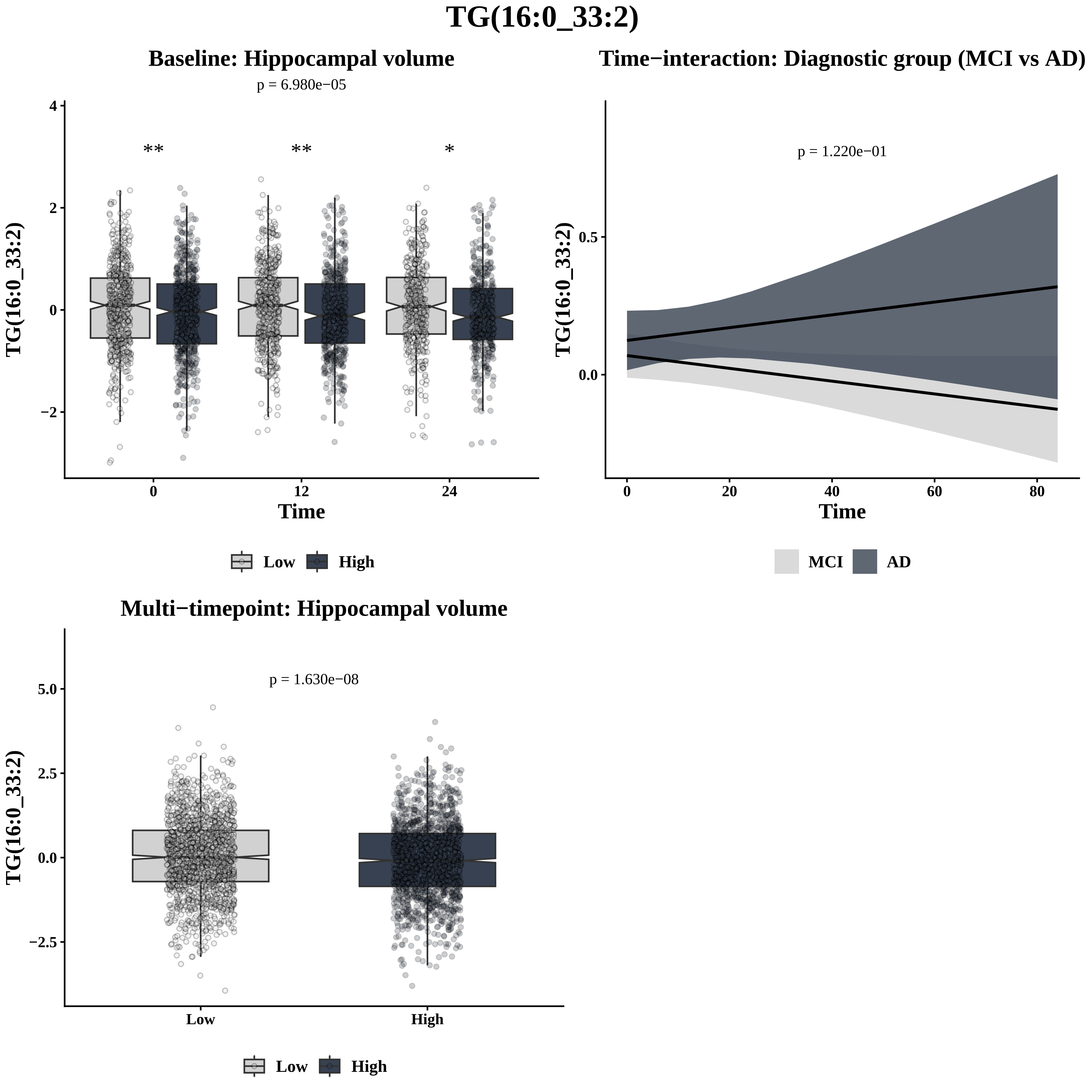
TG(16:0_33:2)
Baseline: Hippocampal volume
p = 6.980e−05
**
**
*
TG(16:0_33:2)
Time
0
12
24
Low
High
Time−interaction: Diagnostic group (MCI vs AD)
p = 1.220e−01
TG(16:0_33:2)
Time
MCI
AD
Multi−timepoint: Hippocampal volume
p = 1.630e−08
TG(16:0_33:2)
Low
High

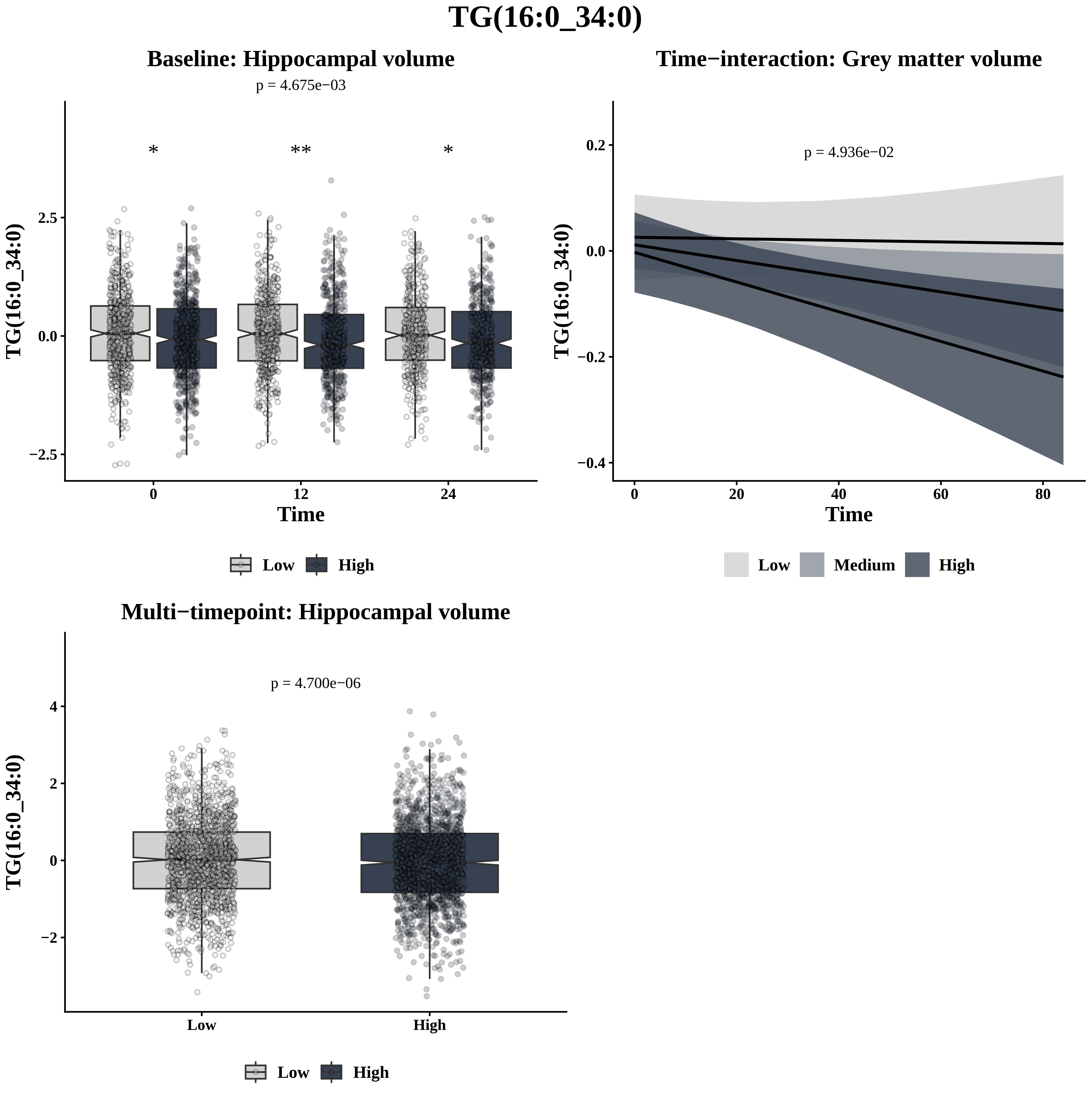
TG(16:0_34:0)
Baseline: Hippocampal volume
p = 4.675e−03
*
**
*
TG(16:0_34:0)
2.5
0.0
−2.5
0
12
24
Time
Low
High
Time−interaction: Grey matter volume
p = 4.936e−02
TG(16:0_34:0)
0.2
0.0
−0.2
−0.4
0
20
40
60
80
Time
Low
Medium
High
Multi−timepoint: Hippocampal volume
p = 4.700e−06
TG(16:0_34:0)
4
2
0
−2
Low
High
Low
High

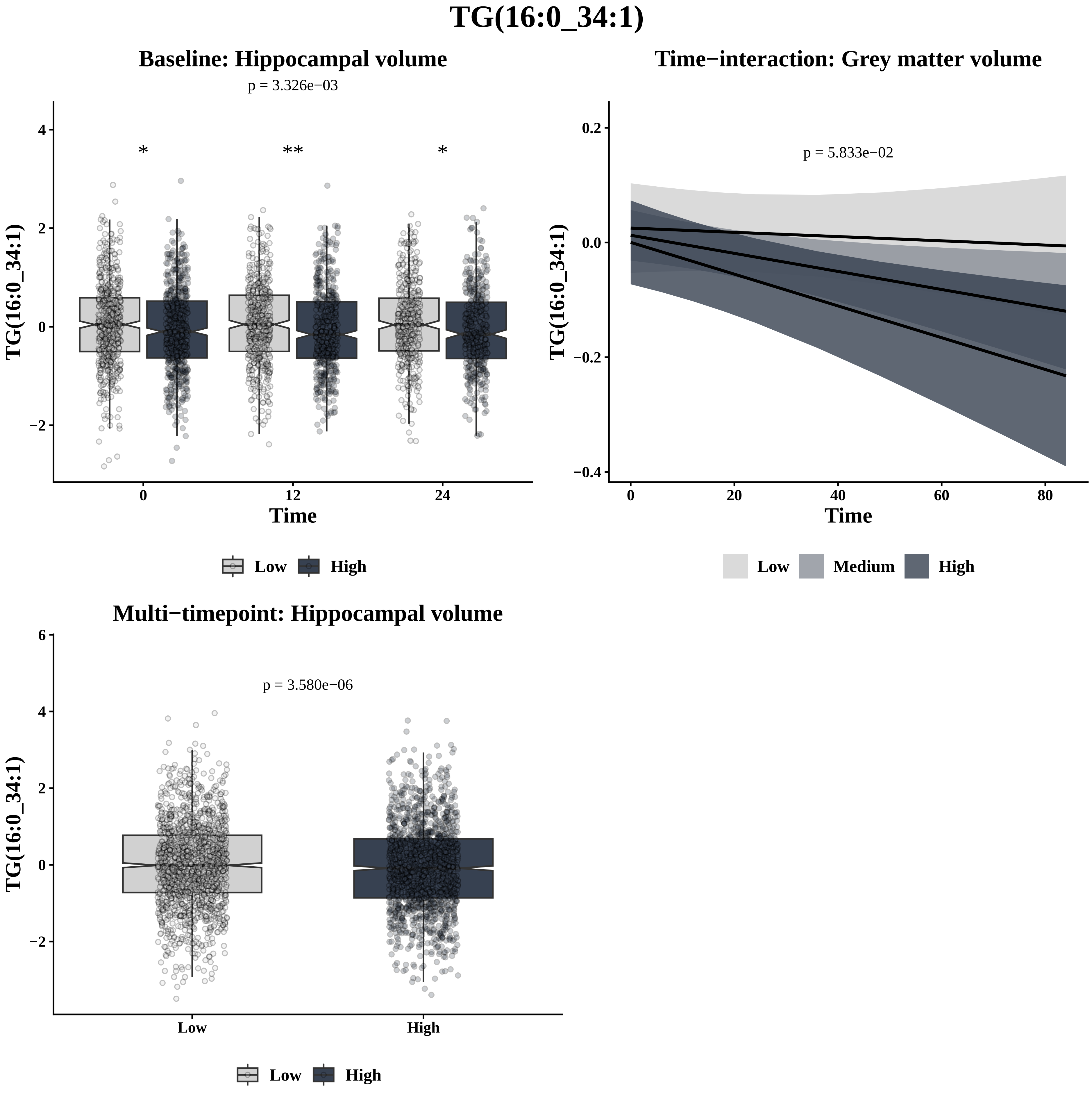
TG(16:0_34:1)
Baseline: Hippocampal volume
p = 3.326e−03
* ** *
TG(16:0_34:1)
Time
0 12 24
Low High
Time−interaction: Grey matter volume
p = 5.833e−02
TG(16:0_34:1)
Time
Low Medium High
Multi−timepoint: Hippocampal volume
p = 3.580e−06
TG(16:0_34:1)
Low High
Low High

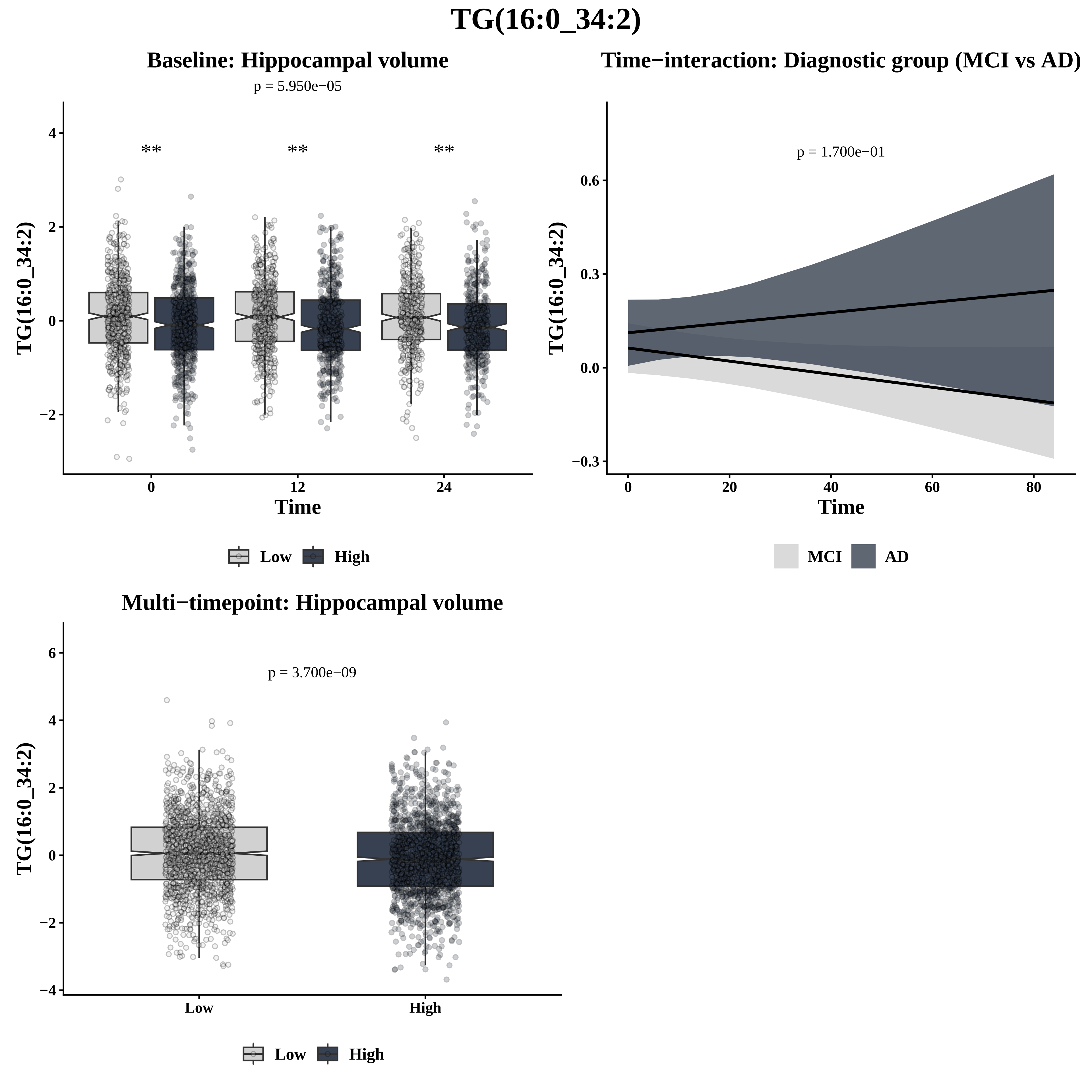

TG(16:0_34:2)
Baseline: Hippocampal volume
p = 5.950e−05
**
**
**
TG(16:0_34:2)
4
2
0
−2
0
12
24
Time
Low
High
Time−interaction: Diagnostic group (MCI vs AD)
p = 1.700e−01
TG(16:0_34:2)
0.6
0.3
0.0
−0.3
0
20
40
60
80
Time
MCI
AD
Multi−timepoint: Hippocampal volume
p = 3.700e−09
TG(16:0_34:2)
6
4
2
0
−2
−4
Low
High
Low
High

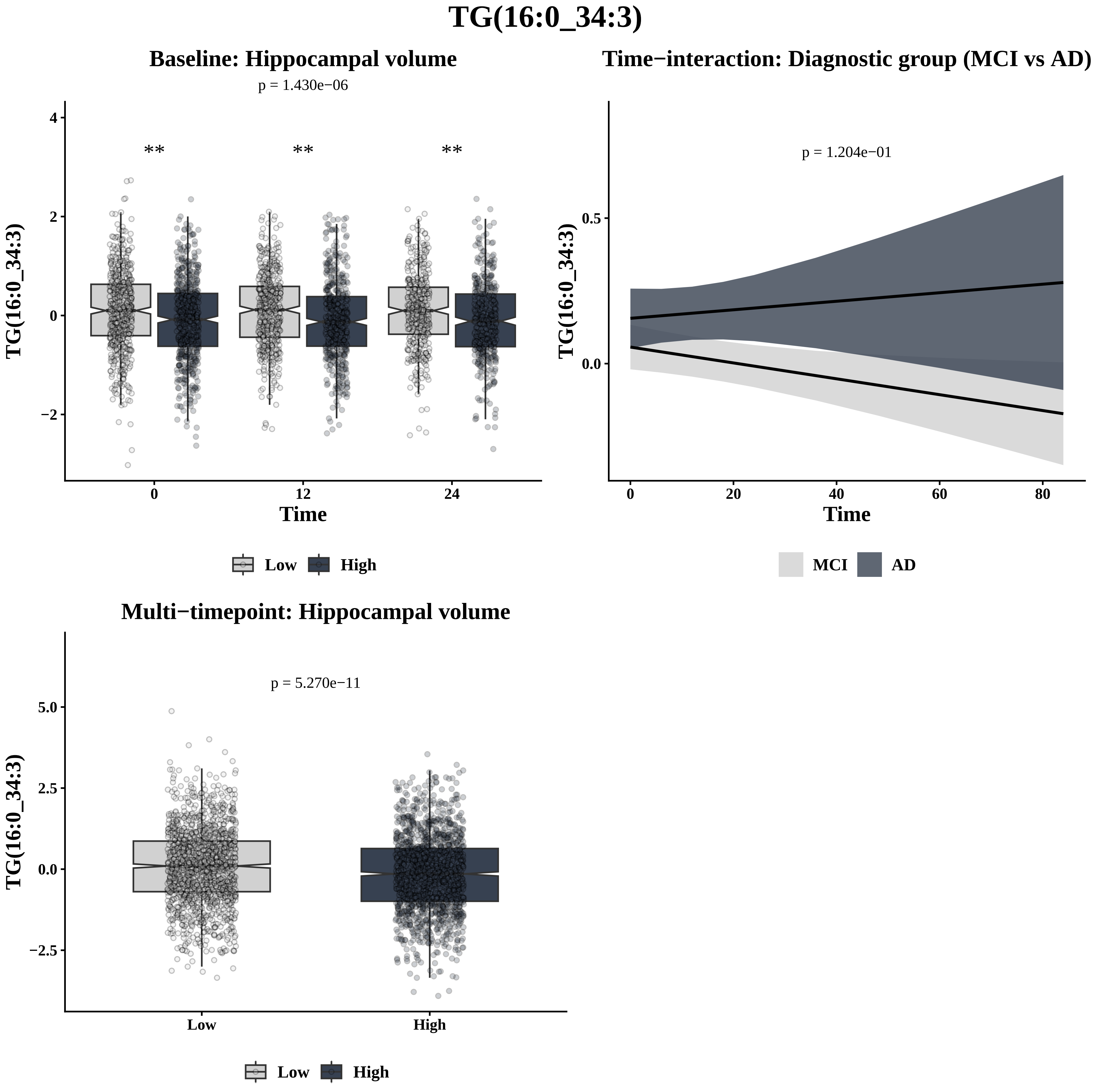

TG(16:0_34:3)
Baseline: Hippocampal volume
p = 1.430e−06
**
**
**
TG(16:0_34:3)
Time
4
2
0
−2
0
12
24
Low
High
Time−interaction: Diagnostic group (MCI vs AD)
p = 1.204e−01
TG(16:0_34:3)
0.5
0.0
0
20
40
60
80
Time
MCI
AD
Multi−timepoint: Hippocampal volume
p = 5.270e−11
TG(16:0_34:3)
5.0
2.5
0.0
−2.5
Low
High
Low
High

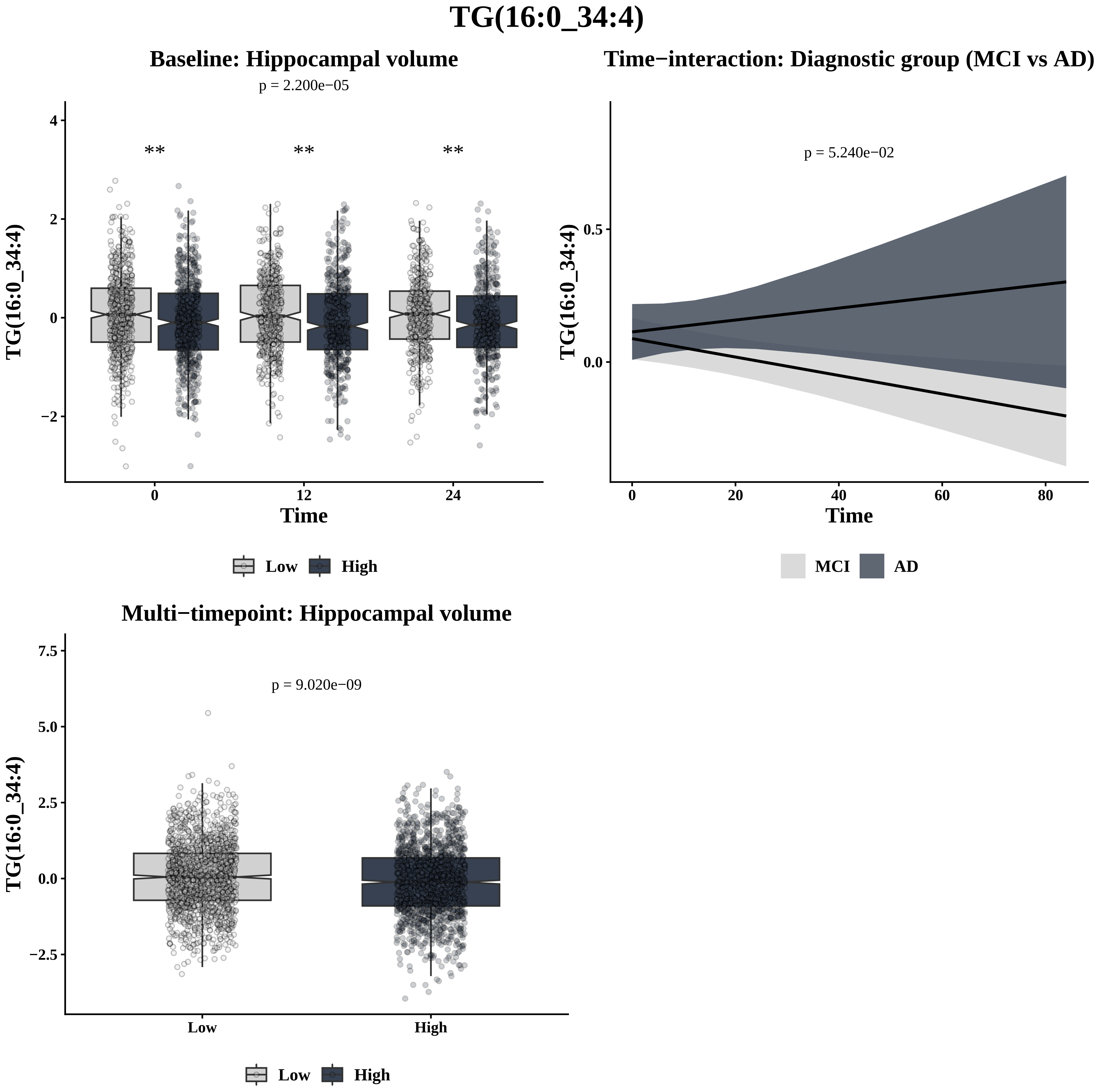
TG(16:0_34:4)
Baseline: Hippocampal volume
p = 2.200e−05
**
**
**
TG(16:0_34:4)
4
2
0
−2
0
12
24
Time
Low
High
Time−interaction: Diagnostic group (MCI vs AD)
p = 5.240e−02
TG(16:0_34:4)
0.5
0.0
0
20
40
60
80
Time
MCI
AD
Multi−timepoint: Hippocampal volume
p = 9.020e−09
TG(16:0_34:4)
7.5
5.0
2.5
0.0
−2.5
Low
High
Low
High

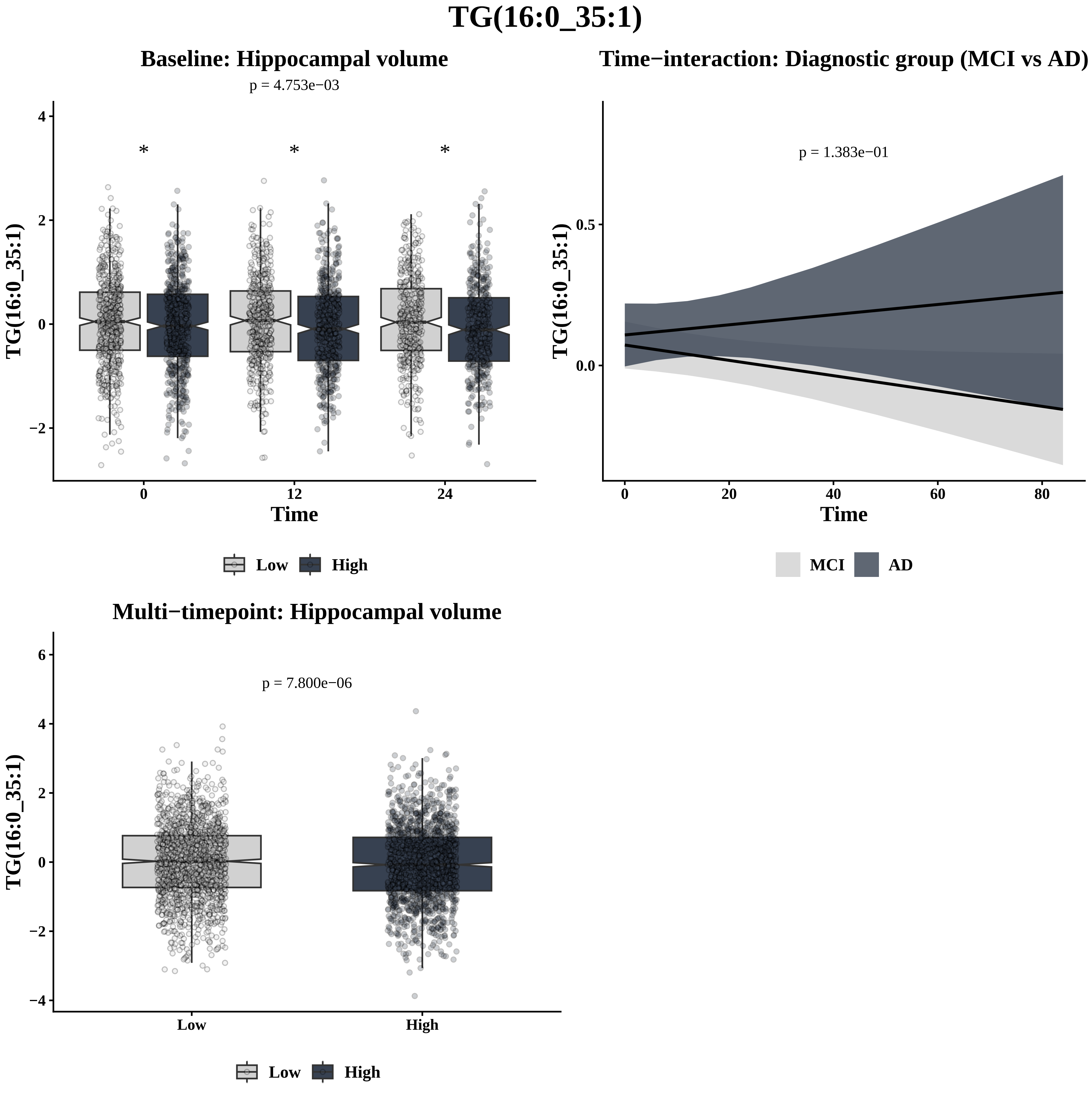
TG(16:0_35:1)
Baseline: Hippocampal volume
p = 4.753e−03
*
*
*
TG(16:0_35:1)
Time
0
12
24
4
2
0
−2
Low
High
Time−interaction: Diagnostic group (MCI vs AD)
p = 1.383e−01
TG(16:0_35:1)
0.5
0.0
0
20
40
60
80
Time
MCI
AD
Multi−timepoint: Hippocampal volume
p = 7.800e−06
TG(16:0_35:1)
6
4
2
0
−2
−4
Low
High
Low
High

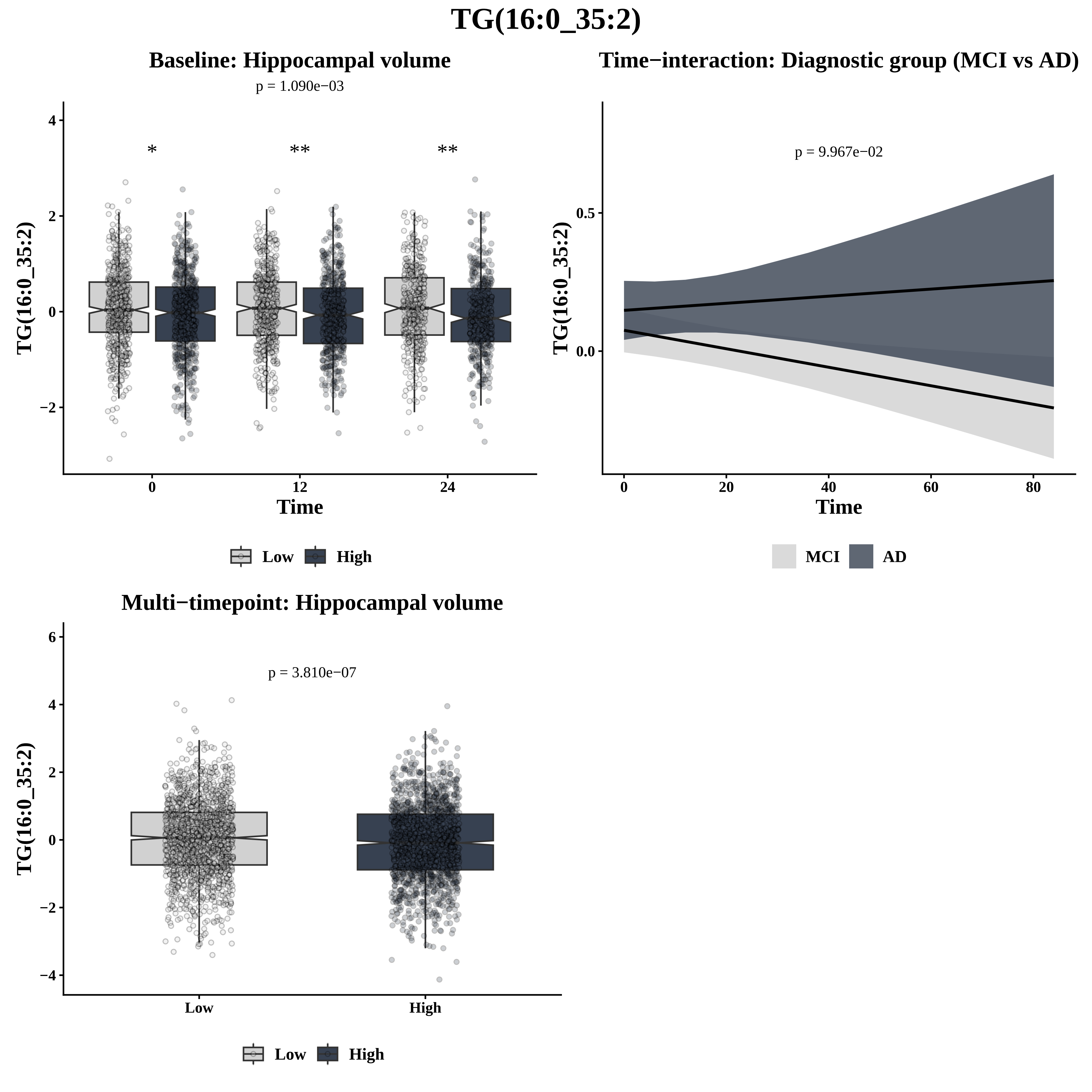

TG(16:0_35:2)
Baseline: Hippocampal volume
p = 1.090e−03
*
**
**
TG(16:0_35:2)
Time
Low
High
Time−interaction: Diagnostic group (MCI vs AD)
p = 9.967e−02
TG(16:0_35:2)
Time
MCI
AD
Multi−timepoint: Hippocampal volume
p = 3.810e−07
TG(16:0_35:2)
Low
High
Low
High

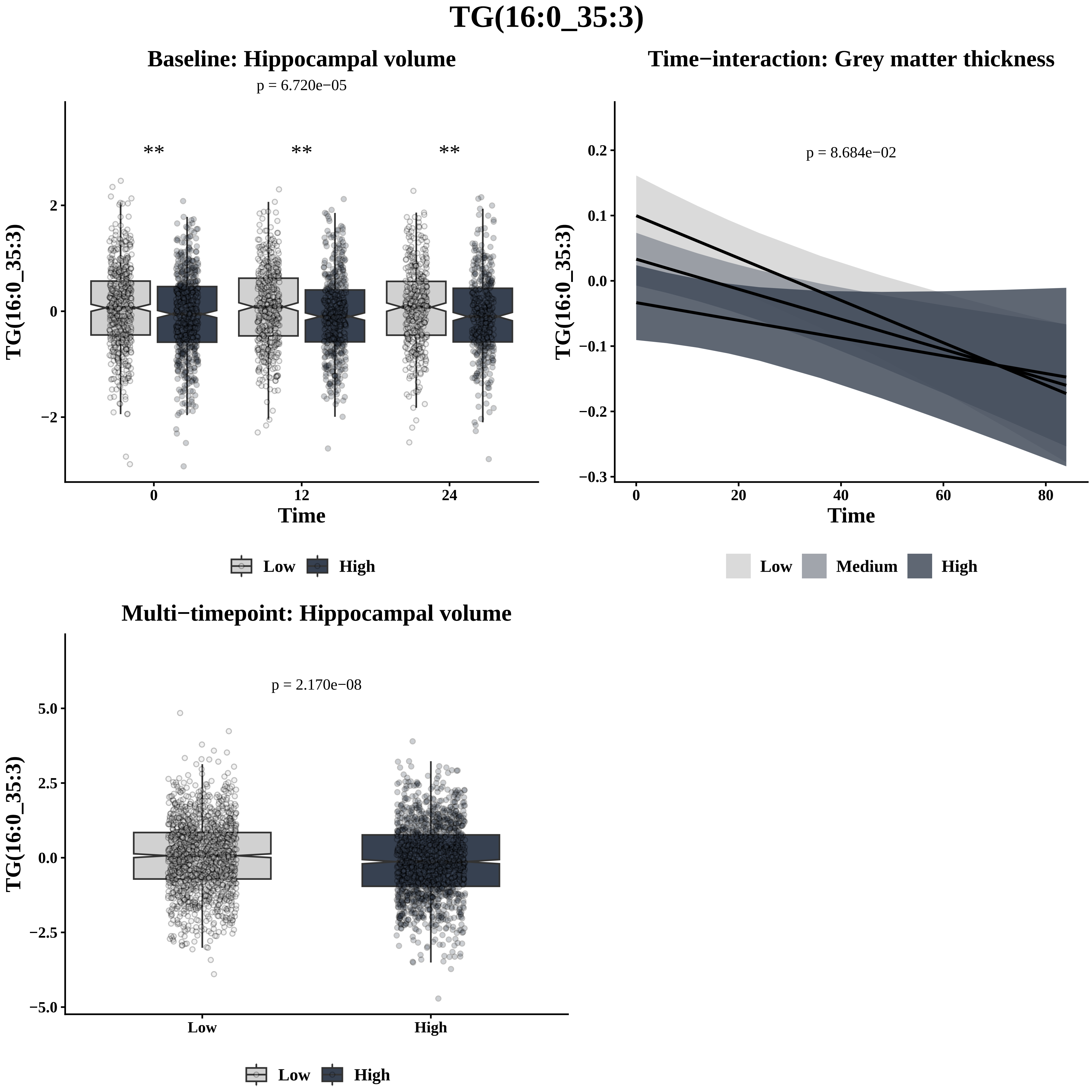
TG(16:0_35:3)
Baseline: Hippocampal volume
p = 6.720e−05
**
**
**
2
0
−2
0
12
24
Time
TG(16:0_35:3)
Low
High
Time−interaction: Grey matter thickness
p = 8.684e−02
0.2
0.1
0.0
−0.1
−0.2
−0.3
0
20
40
60
80
Time
TG(16:0_35:3)
Low
Medium
High
Multi−timepoint: Hippocampal volume
p = 2.170e−08
5.0
2.5
0.0
−2.5
−5.0
Low
High
TG(16:0_35:3)
Low
High

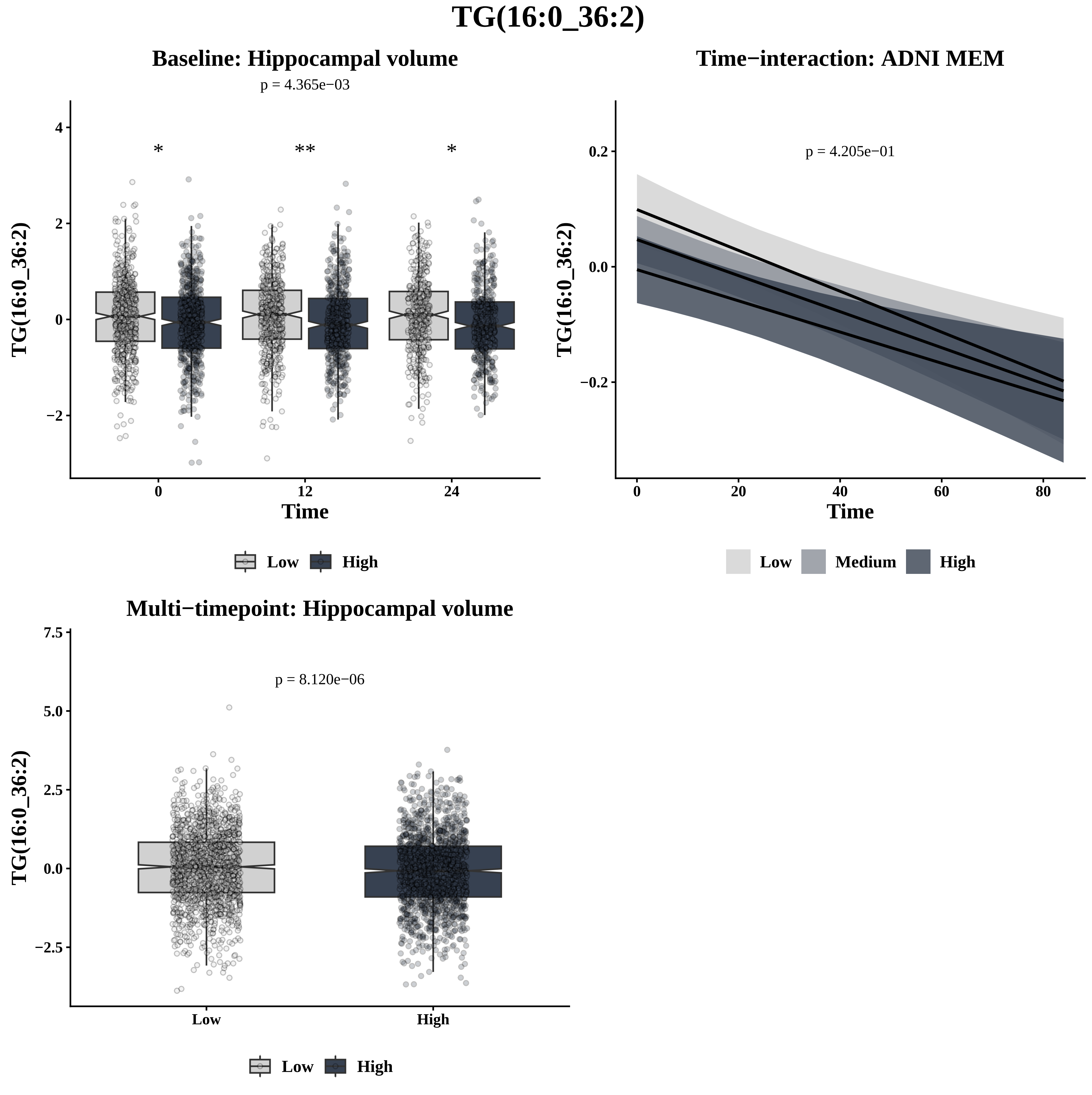
TG(16:0_36:2)
Baseline: Hippocampal volume
p = 4.365e−03
*
**
*
TG(16:0_36:2)
Time
0
12
24
Low
High
Time−interaction: ADNI MEM
p = 4.205e−01
TG(16:0_36:2)
Time
Low
Medium
High
Multi−timepoint: Hippocampal volume
p = 8.120e−06
TG(16:0_36:2)
Low
High

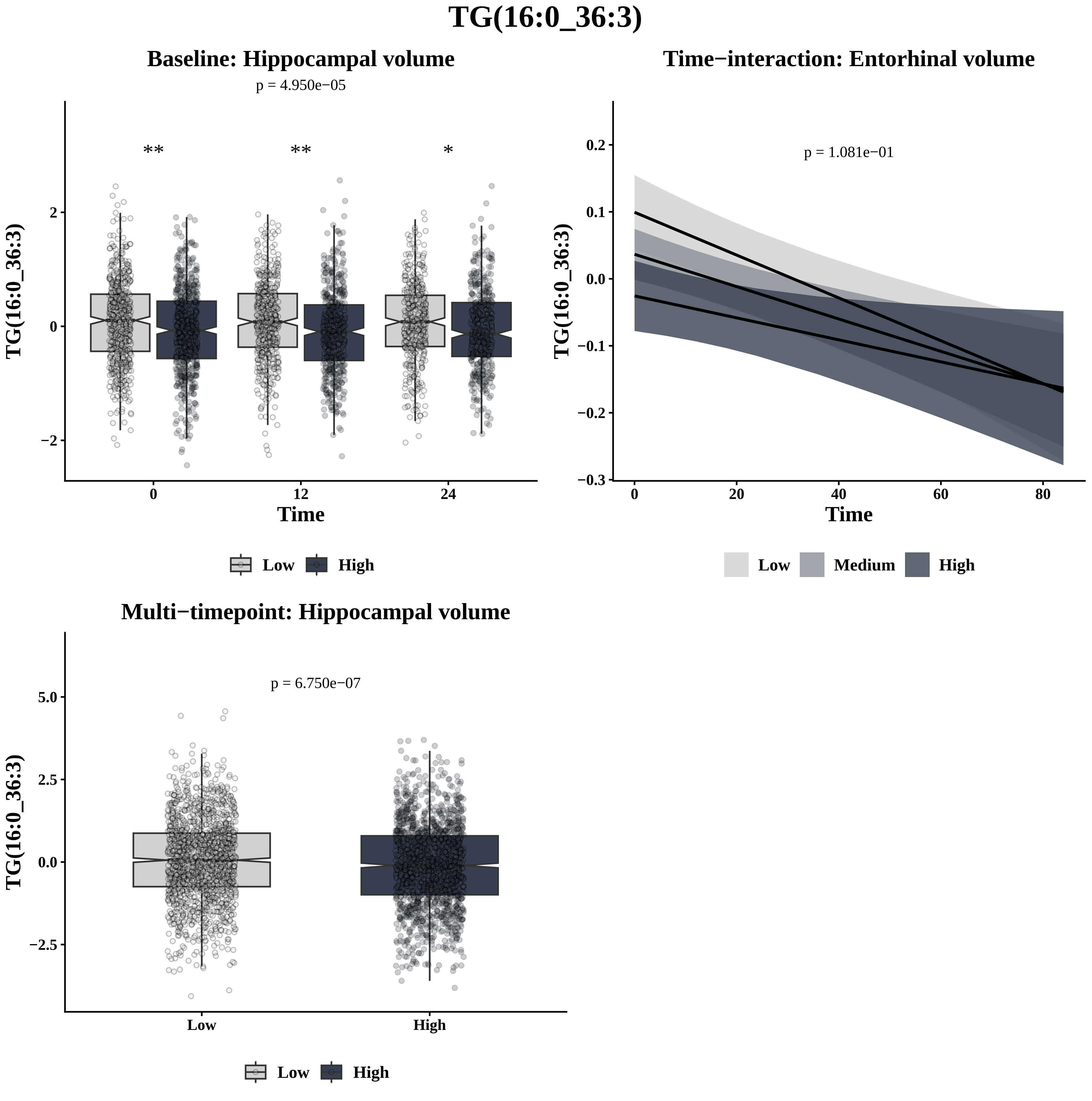

TG(16:0_36:3)
Baseline: Hippocampal volume
p = 4.950e−05
**
**
*
2
0
−2
0
12
24
Time
TG(16:0_36:3)
Low
High
Time−interaction: Entorhinal volume
p = 1.081e−01
0.2
0.1
0.0
−0.1
−0.2
−0.3
0
20
40
60
80
Time
TG(16:0_36:3)
Low
Medium
High
Multi−timepoint: Hippocampal volume
p = 6.750e−07
5.0
2.5
0.0
−2.5
Low
High
TG(16:0_36:3)
Low
High

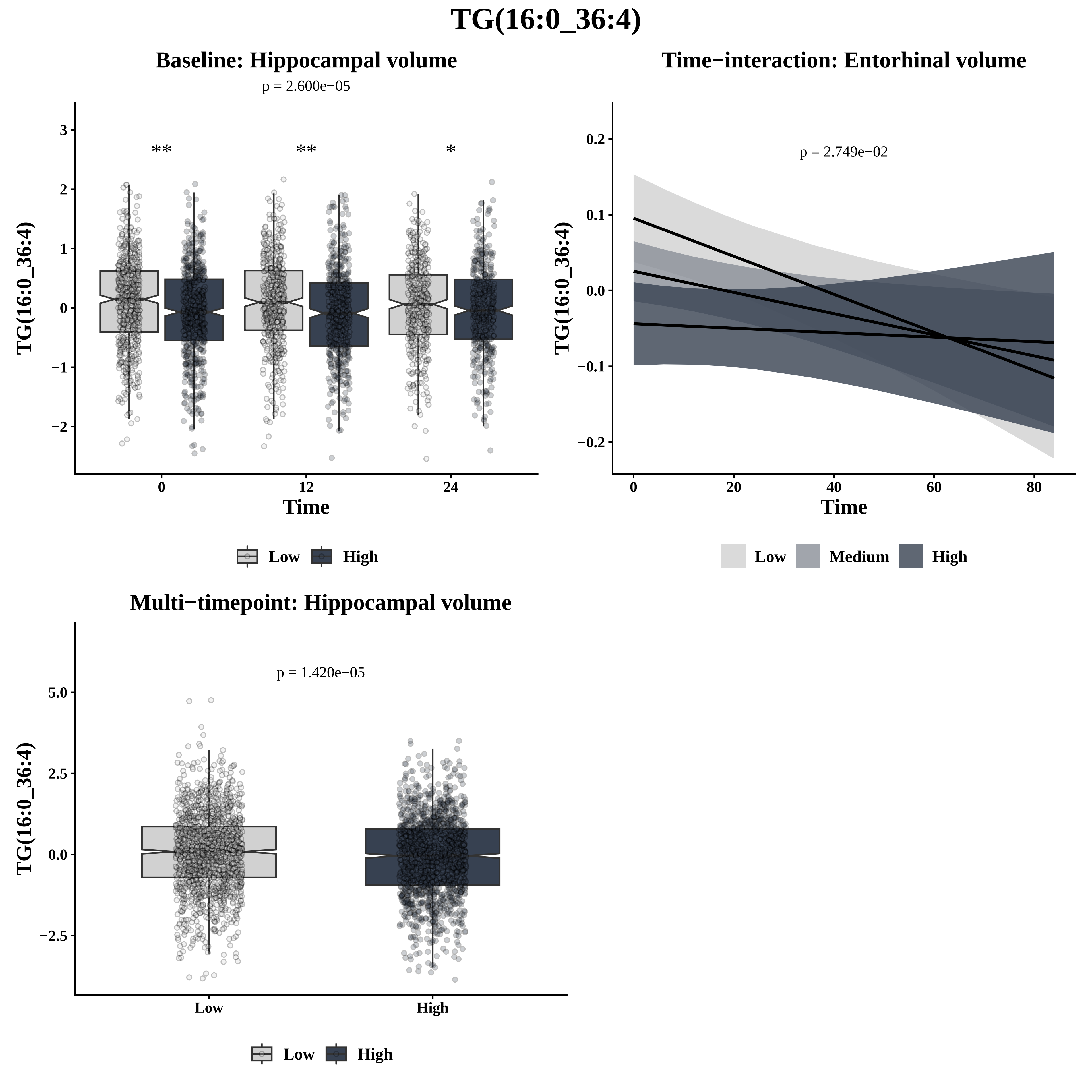

TG(16:0_36:4)
Baseline: Hippocampal volume
p = 2.600e−05
**
**
*
3
2
1
0
−1
−2
TG(16:0_36:4)
0
12
24
Time
Low
High
Time−interaction: Entorhinal volume
p = 2.749e−02
0.2
0.1
0.0
−0.1
−0.2
TG(16:0_36:4)
0
20
40
60
80
Time
Low
Medium
High
Multi−timepoint: Hippocampal volume
p = 1.420e−05
5.0
2.5
0.0
−2.5
TG(16:0_36:4)
Low
High
Low
High

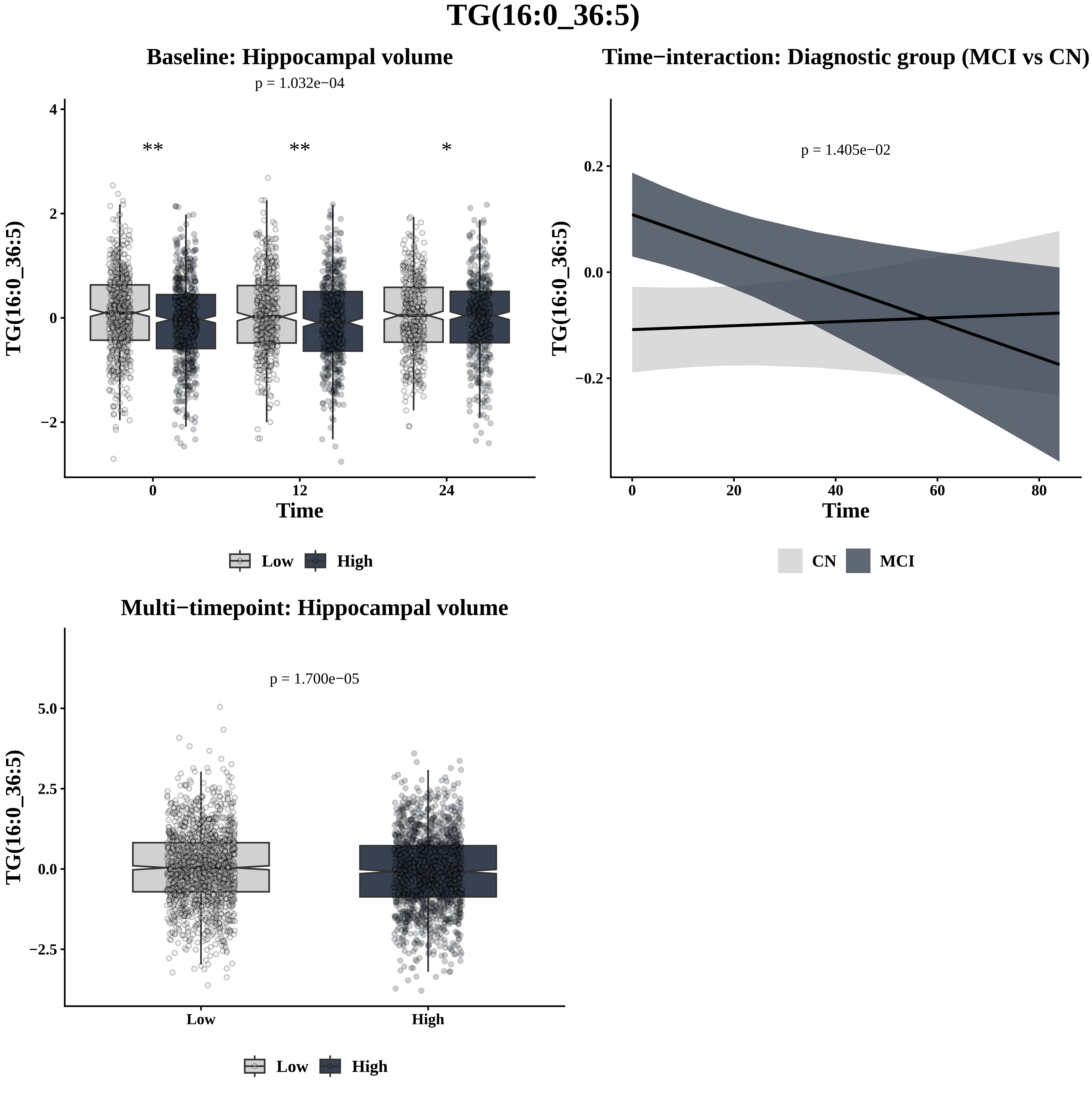
TG(16:0_36:5)
Baseline: Hippocampal volume
p = 1.032e−04
**
**
*
4
2
0
−2
TG(16:0_36:5)
0
12
24
Time
Low
High
Time−interaction: Diagnostic group (MCI vs CN)
p = 1.405e−02
0.2
0.0
−0.2
TG(16:0_36:5)
0
20
40
60
80
Time
CN
MCI
Multi−timepoint: Hippocampal volume
p = 1.700e−05
5.0
2.5
0.0
−2.5
TG(16:0_36:5)
Low
High
Low
High

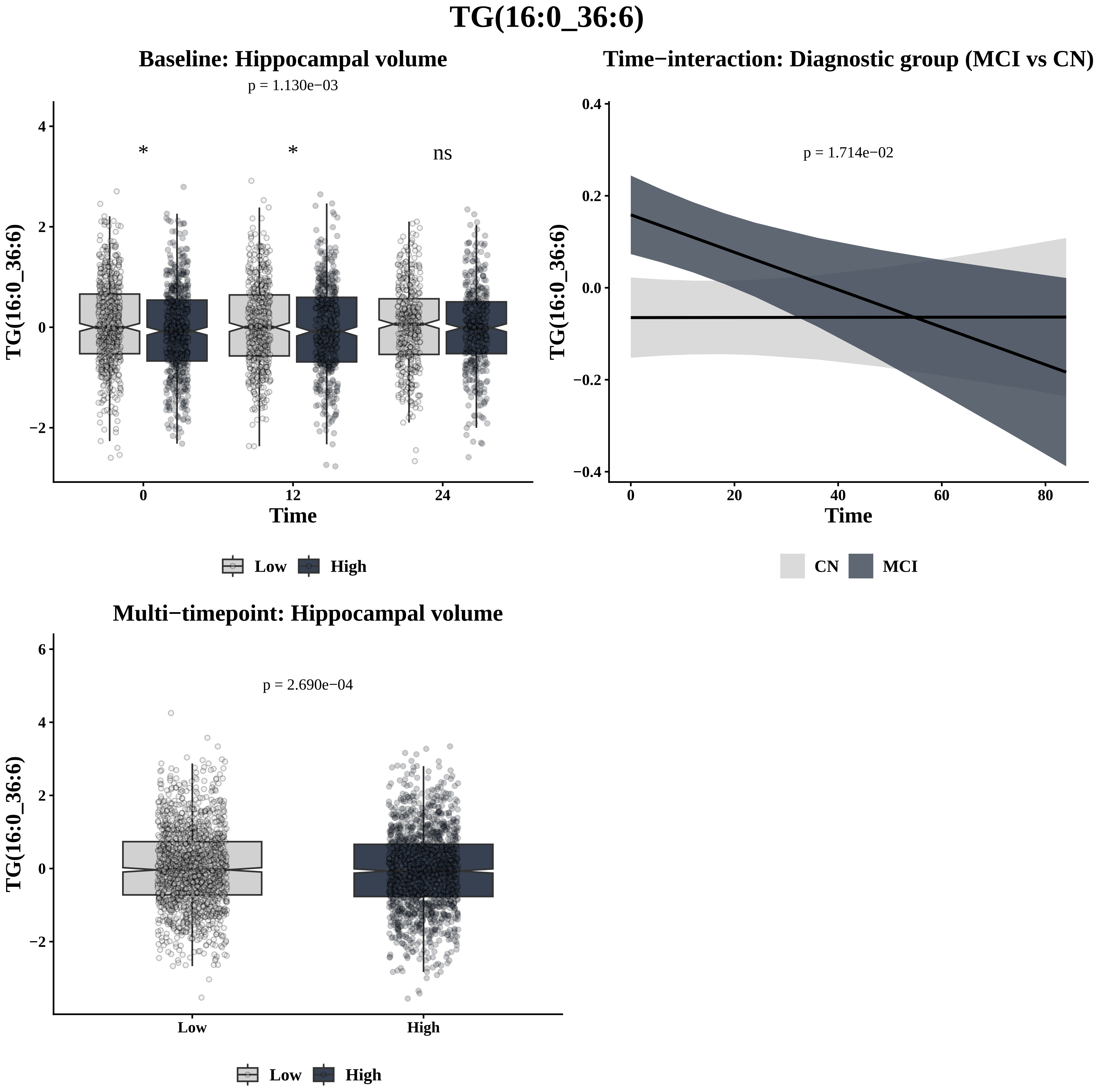
TG(16:0_36:6)
Baseline: Hippocampal volume
p = 1.130e−03
*
*
ns
TG(16:0_36:6)
Time
Low
High
Time−interaction: Diagnostic group (MCI vs CN)
p = 1.714e−02
TG(16:0_36:6)
Time
CN
MCI
Multi−timepoint: Hippocampal volume
p = 2.690e−04
TG(16:0_36:6)
Low
High
Low
High

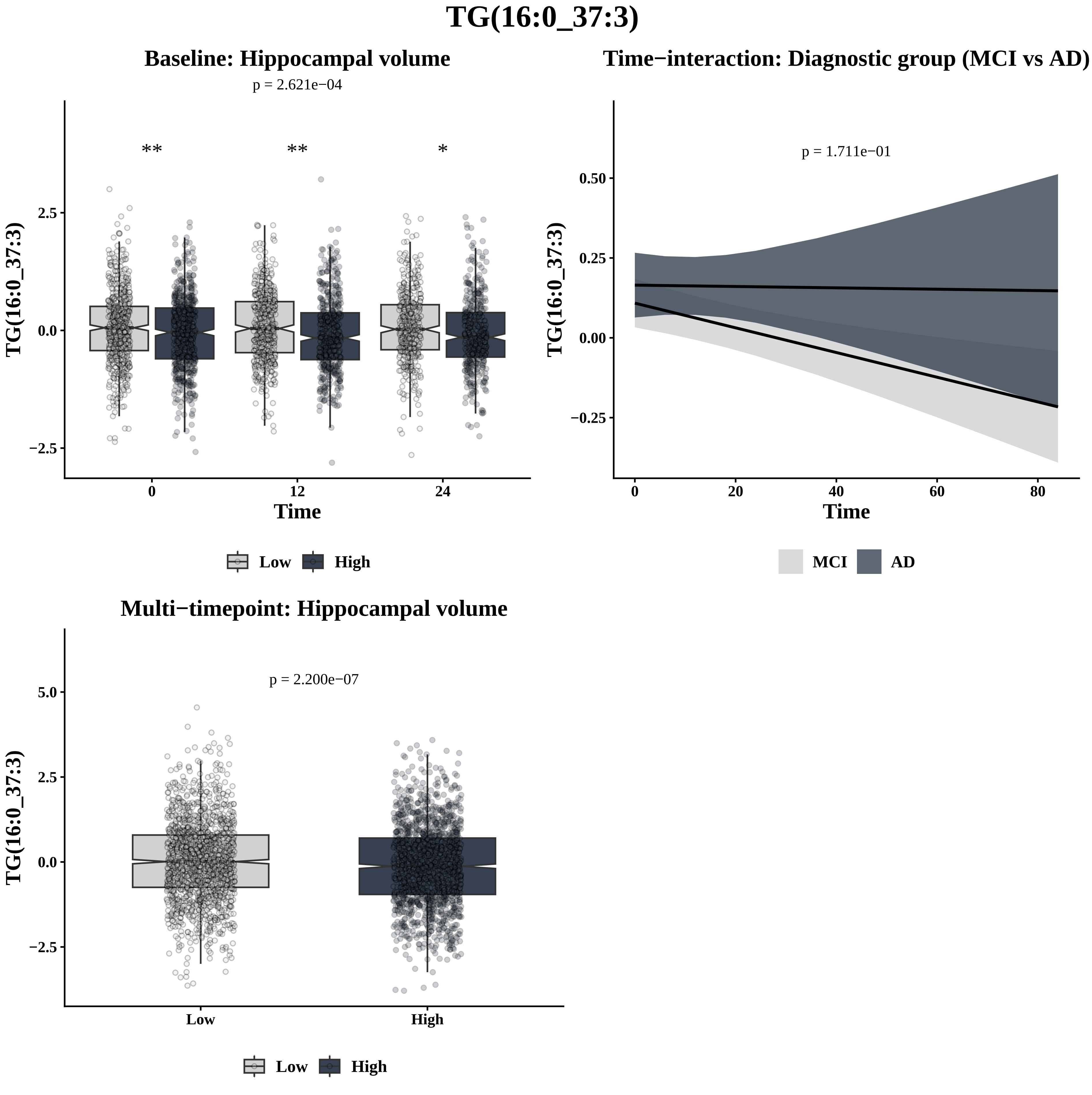
TG(16:0_37:3)
Baseline: Hippocampal volume
p = 2.621e−04
**
**
*
TG(16:0_37:3)
2.5
0.0
−2.5
0
12
24
Time
Low
High
Time−interaction: Diagnostic group (MCI vs AD)
p = 1.711e−01
TG(16:0_37:3)
0.50
0.25
0.00
−0.25
0
20
40
60
80
Time
MCI
AD
Multi−timepoint: Hippocampal volume
p = 2.200e−07
TG(16:0_37:3)
5.0
2.5
0.0
−2.5
Low
High
Low
High

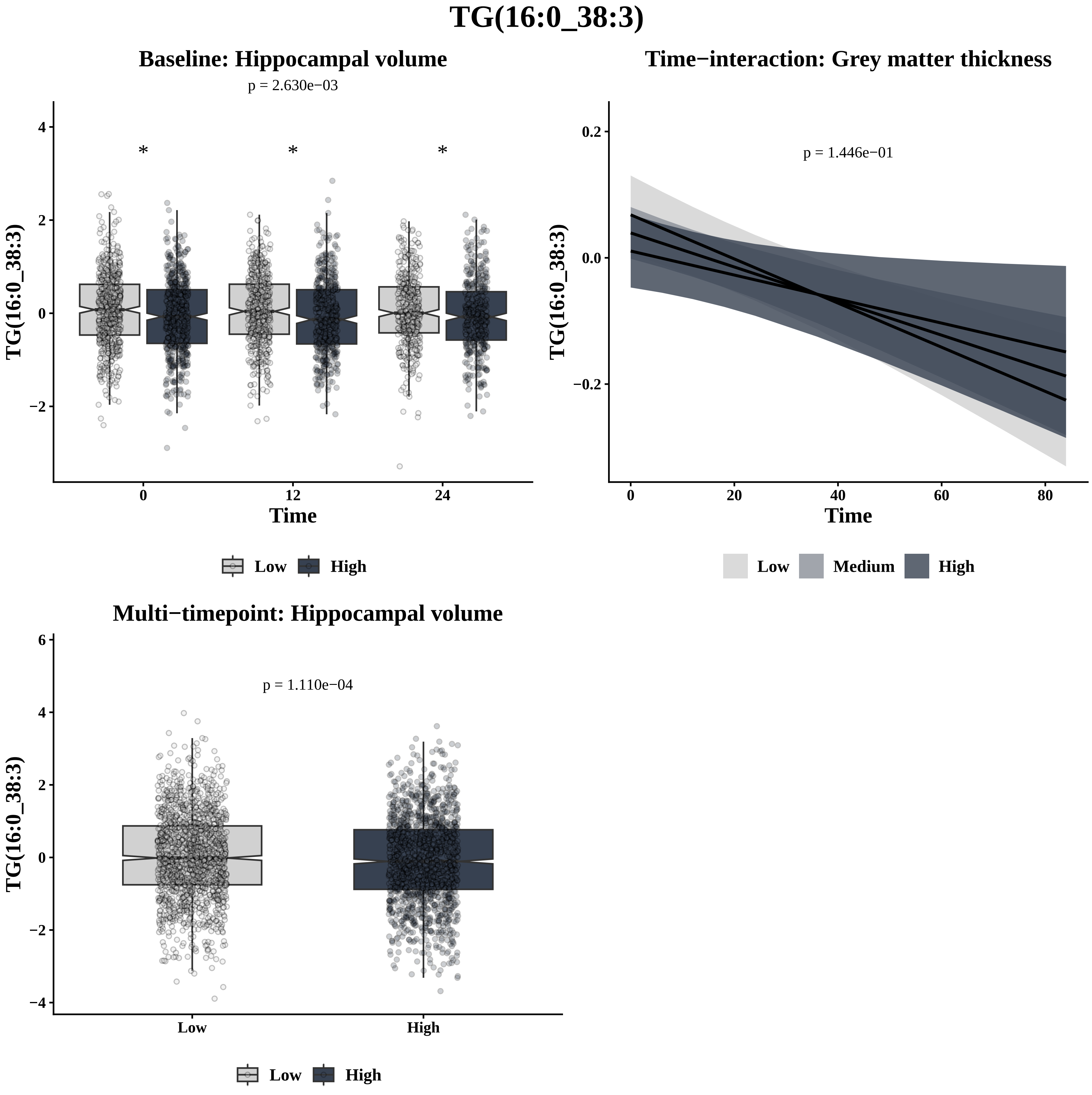

TG(16:0_38:3)
Baseline: Hippocampal volume
p = 2.630e−03
TG(16:0_38:3)
Time
Low
High
Time−interaction: Grey matter thickness
p = 1.446e−01
TG(16:0_38:3)
Time
Low
Medium
High
Multi−timepoint: Hippocampal volume
p = 1.110e−04
TG(16:0_38:3)
Low
High

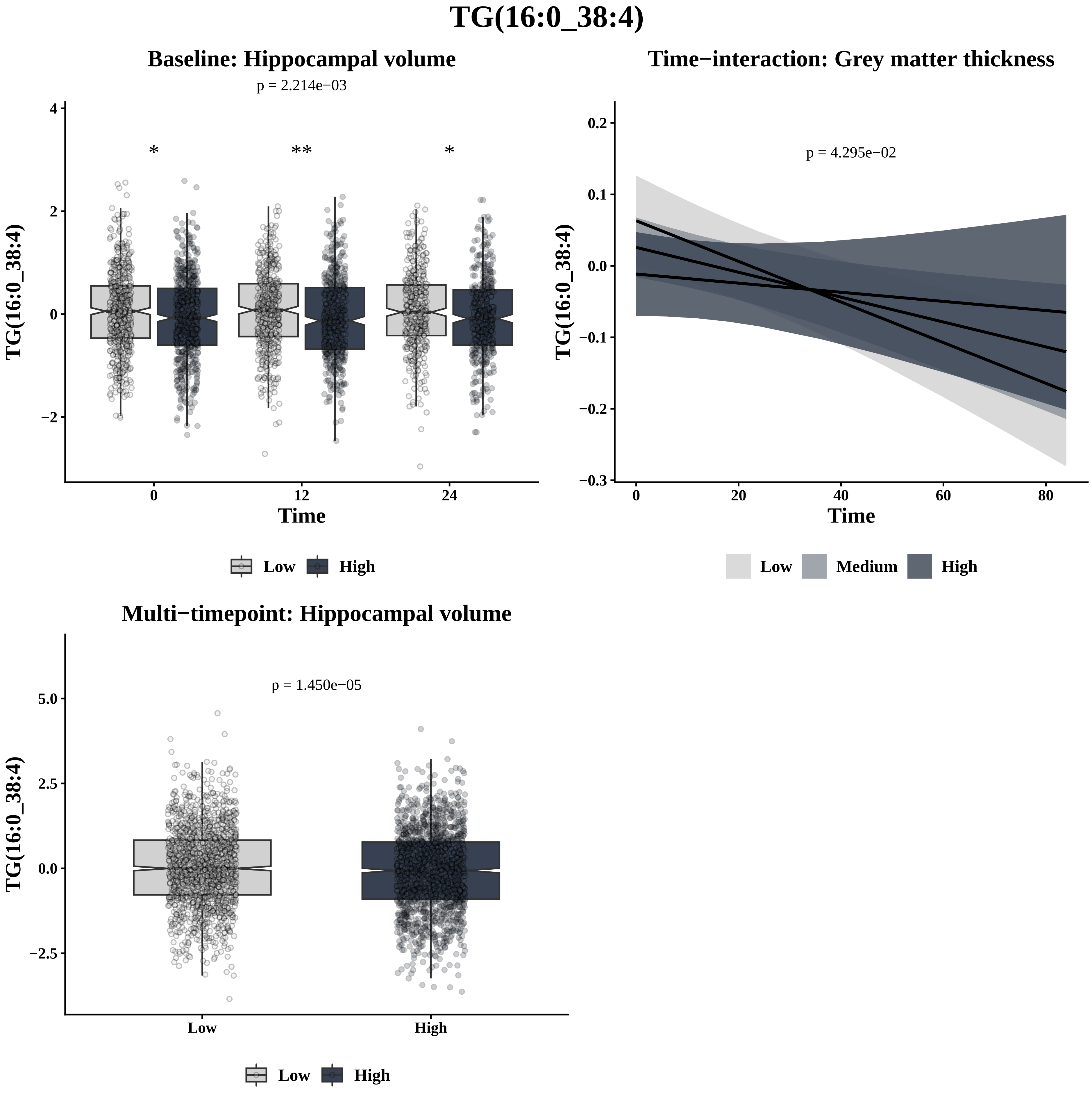
TG(16:0_38:4)
Baseline: Hippocampal volume
p = 2.214e−03
*
**
*
TG(16:0_38:4)
Time
0
12
24
Low
High
Time−interaction: Grey matter thickness
p = 4.295e−02
TG(16:0_38:4)
Time
Low
Medium
High
Multi−timepoint: Hippocampal volume
p = 1.450e−05
TG(16:0_38:4)
Low
High
Low
High

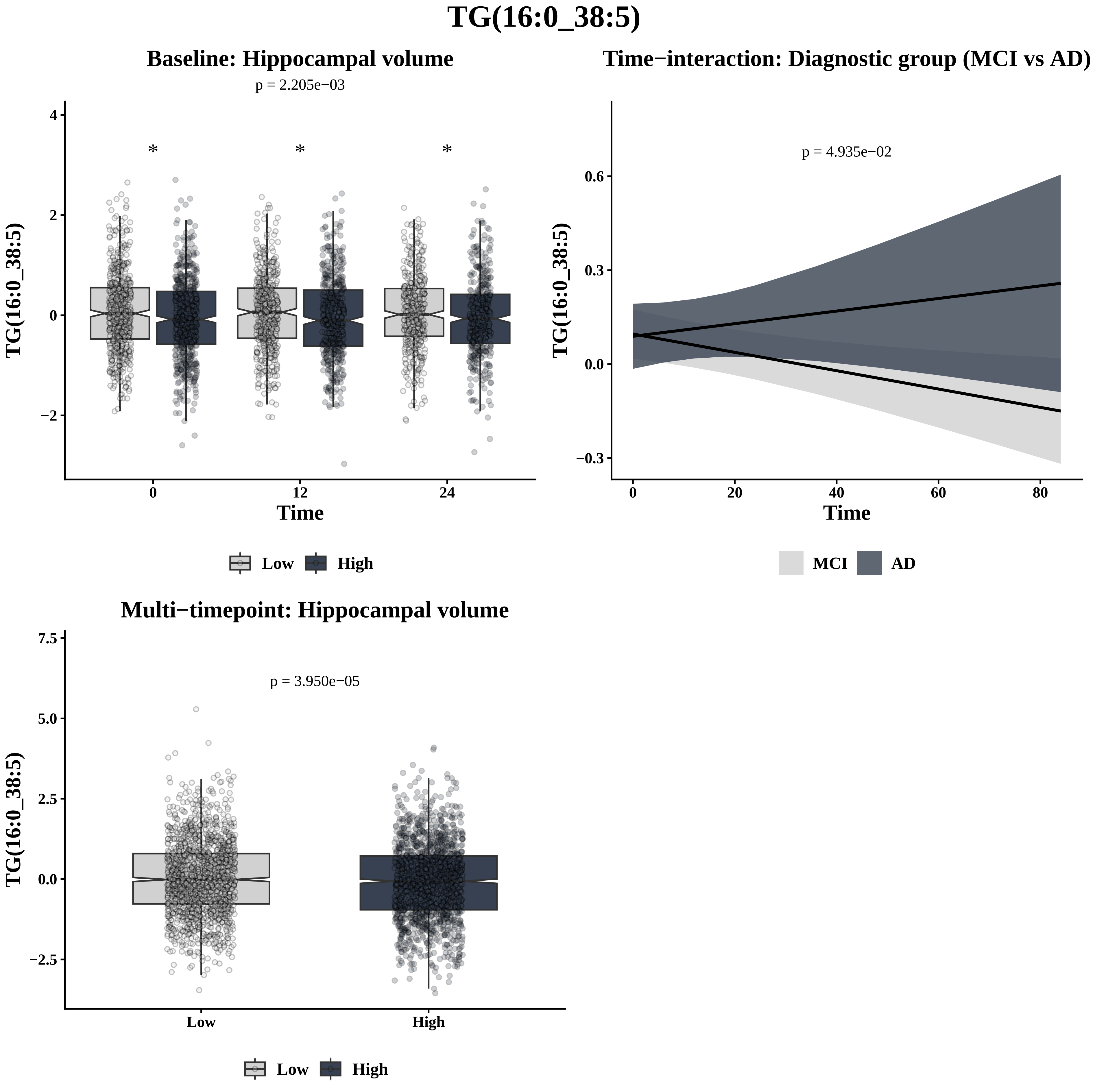

TG(16:0_38:5)
Baseline: Hippocampal volume
p = 2.205e−03
TG(16:0_38:5)
Time
Low
High
Time−interaction: Diagnostic group (MCI vs AD)
p = 4.935e−02
Time
MCI
AD
Multi−timepoint: Hippocampal volume
p = 3.950e−05
Low
High

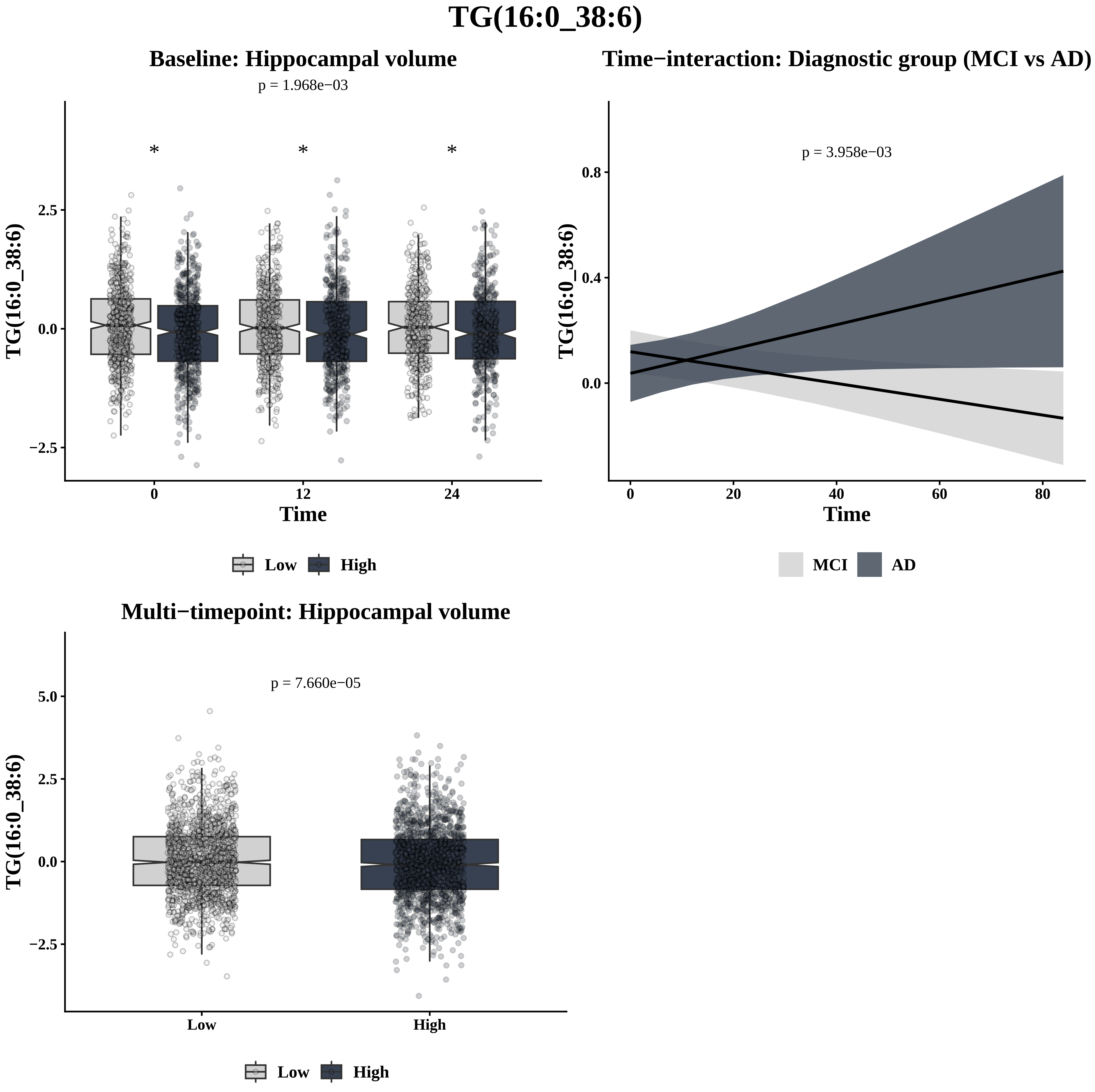
TG(16:0_38:6)
Baseline: Hippocampal volume
p = 1.968e−03
*
*
*
2.5
0.0
−2.5
TG(16:0_38:6)
0
12
24
Time
Low
High
Time−interaction: Diagnostic group (MCI vs AD)
p = 3.958e−03
0.8
0.4
0.0
TG(16:0_38:6)
0
20
40
60
80
Time
MCI
AD
Multi−timepoint: Hippocampal volume
p = 7.660e−05
5.0
2.5
0.0
−2.5
TG(16:0_38:6)
Low
High
Low
High

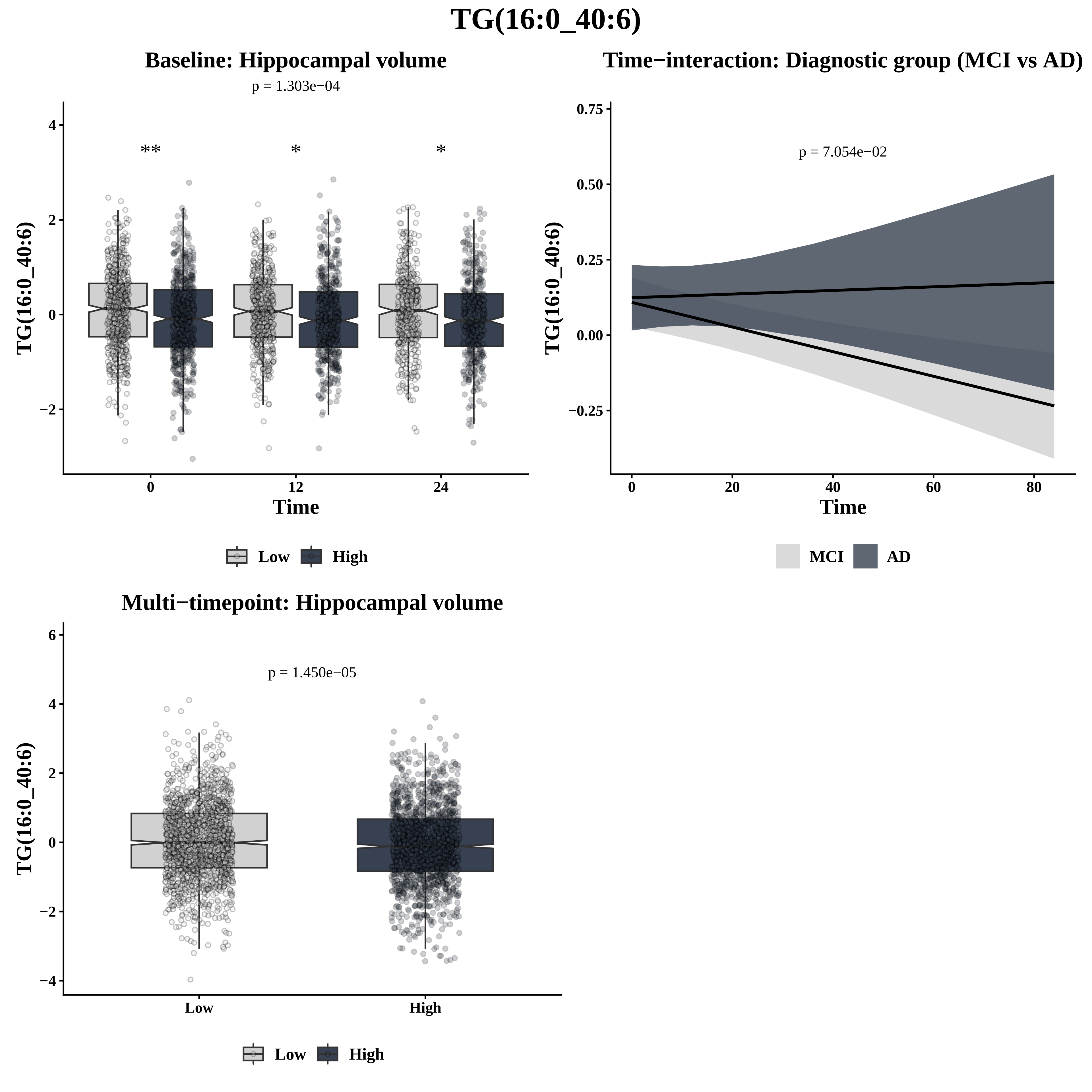

TG(16:0_40:6)
Baseline: Hippocampal volume
p = 1.303e−04
**
*
*
TG(16:0_40:6)
Time
0
12
24
Low
High
Time−interaction: Diagnostic group (MCI vs AD)
p = 7.054e−02
TG(16:0_40:6)
Time
0.75
0.50
0.25
0.00
−0.25
0
20
40
60
80
MCI
AD
Multi−timepoint: Hippocampal volume
p = 1.450e−05
TG(16:0_40:6)
6
4
2
0
−2
−4
Low
High
Low
High

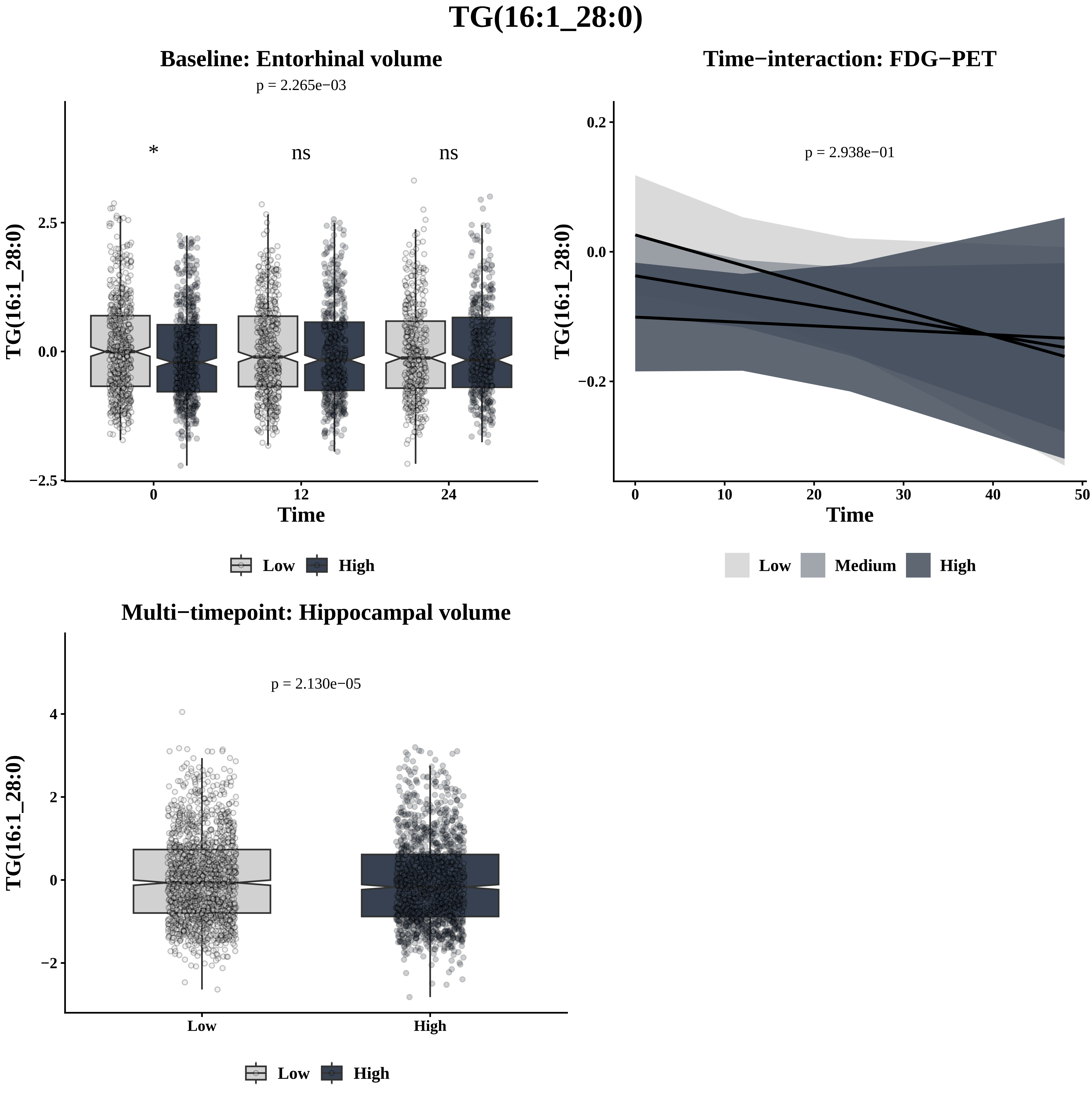

TG(16:1_28:0)
Baseline: Entorhinal volume
p = 2.265e−03
*
ns
ns
TG(16:1_28:0)
2.5
0.0
−2.5
0
12
24
Time
Low
High
Time−interaction: FDG−PET
p = 2.938e−01
0.2
0.0
−0.2
0
10
20
30
40
50
Time
Low
Medium
High
Multi−timepoint: Hippocampal volume
p = 2.130e−05
4
2
0
−2
Low
High

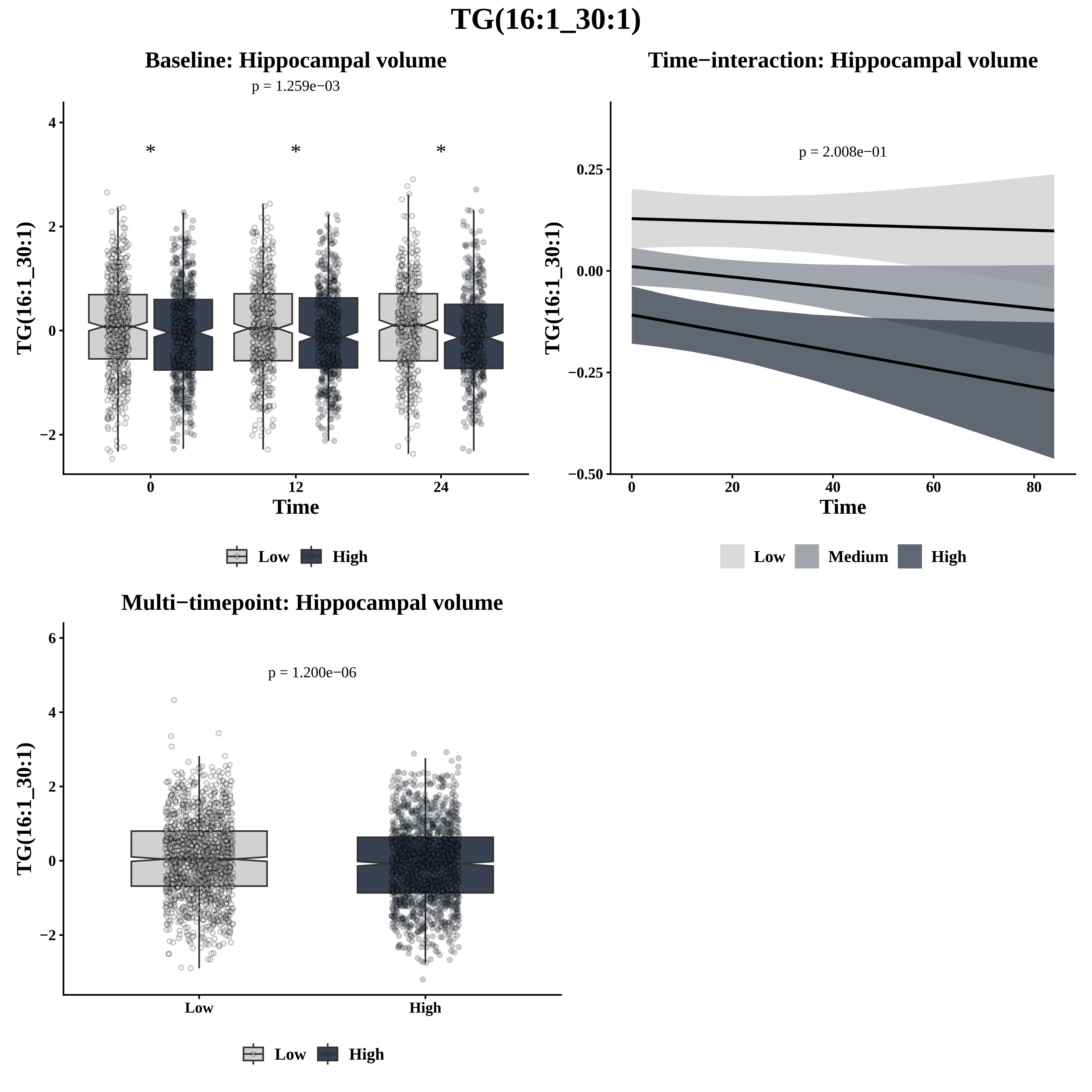

TG(16:1_30:1)
Baseline: Hippocampal volume
p = 1.259e−03
*
*
*
4
2
0
−2
0
12
24
Time
TG(16:1_30:1)
Low
High
Time−interaction: Hippocampal volume
p = 2.008e−01
0.25
0.00
−0.25
−0.50
0
20
40
60
80
Time
TG(16:1_30:1)
Low
Medium
High
Multi−timepoint: Hippocampal volume
p = 1.200e−06
6
4
2
0
−2
Low
High
TG(16:1_30:1)
Low
High

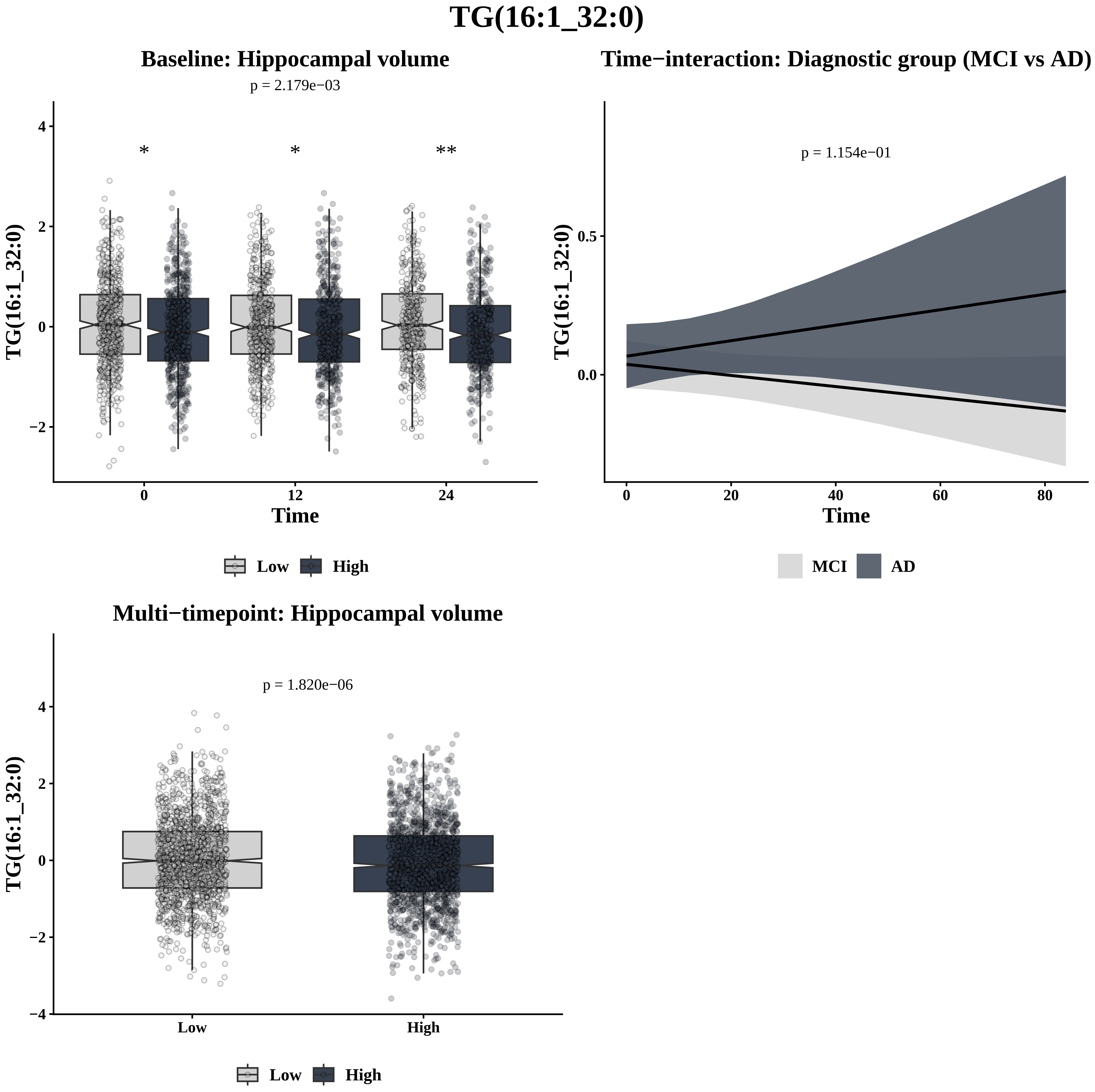
TG(16:1_32:0)
Baseline: Hippocampal volume
p = 2.179e−03
*
*
**
TG(16:1_32:0)
4
2
0
−2
0
12
24
Time
Low
High
Time−interaction: Diagnostic group (MCI vs AD)
p = 1.154e−01
TG(16:1_32:0)
0.5
0.0
0
20
40
60
80
Time
MCI
AD
Multi−timepoint: Hippocampal volume
p = 1.820e−06
TG(16:1_32:0)
4
2
0
−2
−4
Low
High
Low
High

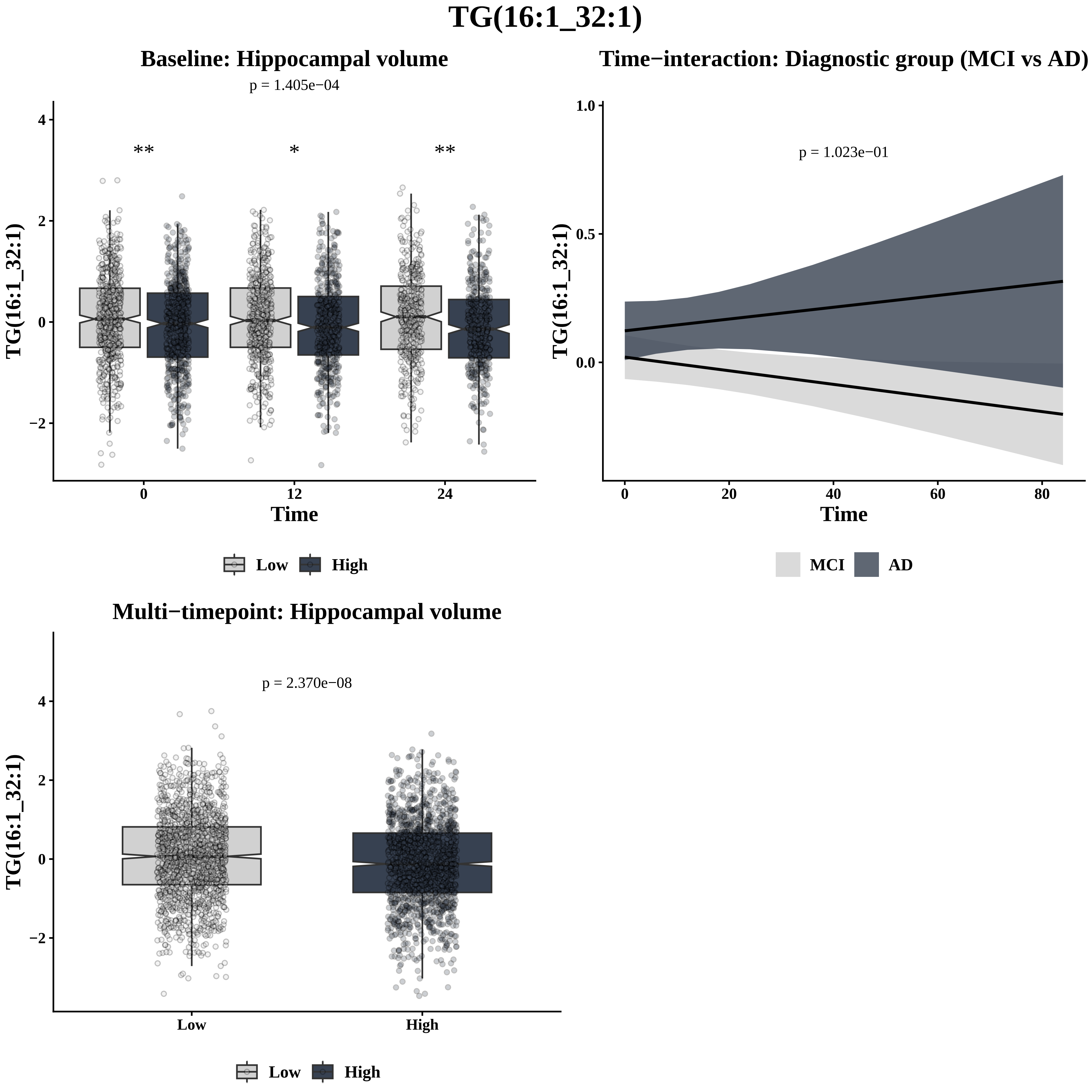

TG(16:1_32:1)
Baseline: Hippocampal volume
p = 1.405e−04
**
*
**
TG(16:1_32:1)
Time
0
12
24
4
2
0
−2
Low
High
Time−interaction: Diagnostic group (MCI vs AD)
p = 1.023e−01
TG(16:1_32:1)
1.0
0.5
0.0
0
20
40
60
80
Time
MCI
AD
Multi−timepoint: Hippocampal volume
p = 2.370e−08
TG(16:1_32:1)
4
2
0
−2
Low
High
Low
High

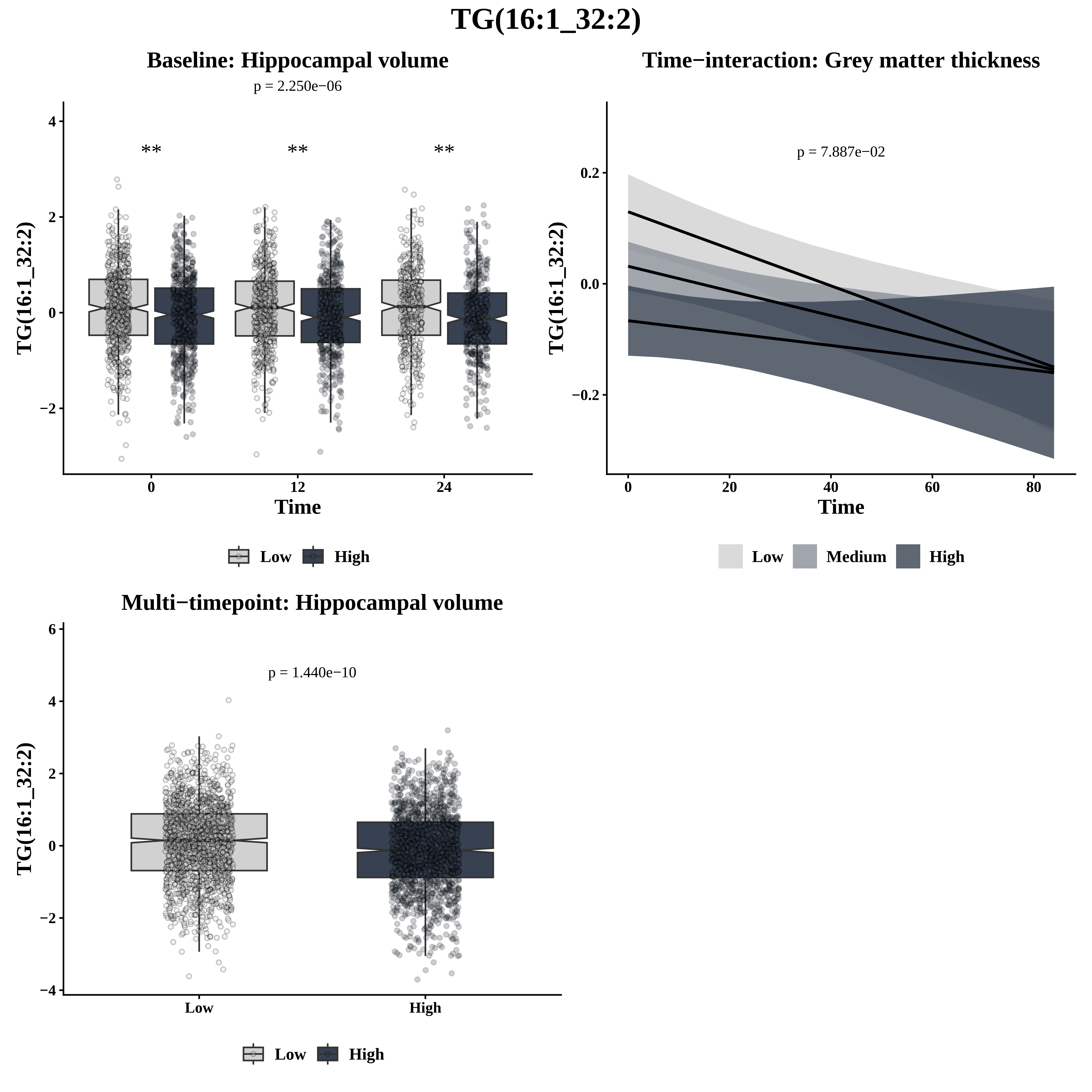
TG(16:1_32:2)
Baseline: Hippocampal volume
p = 2.250e−06
**
**
**
TG(16:1_32:2)
Time
0
12
24
Low
High
Time−interaction: Grey matter thickness
p = 7.887e−02
TG(16:1_32:2)
Time
0
20
40
60
80
0.2
0.0
−0.2
Low
Medium
High
Multi−timepoint: Hippocampal volume
p = 1.440e−10
TG(16:1_32:2)
Low
High
Low
High

# TG(16:1_33:1)

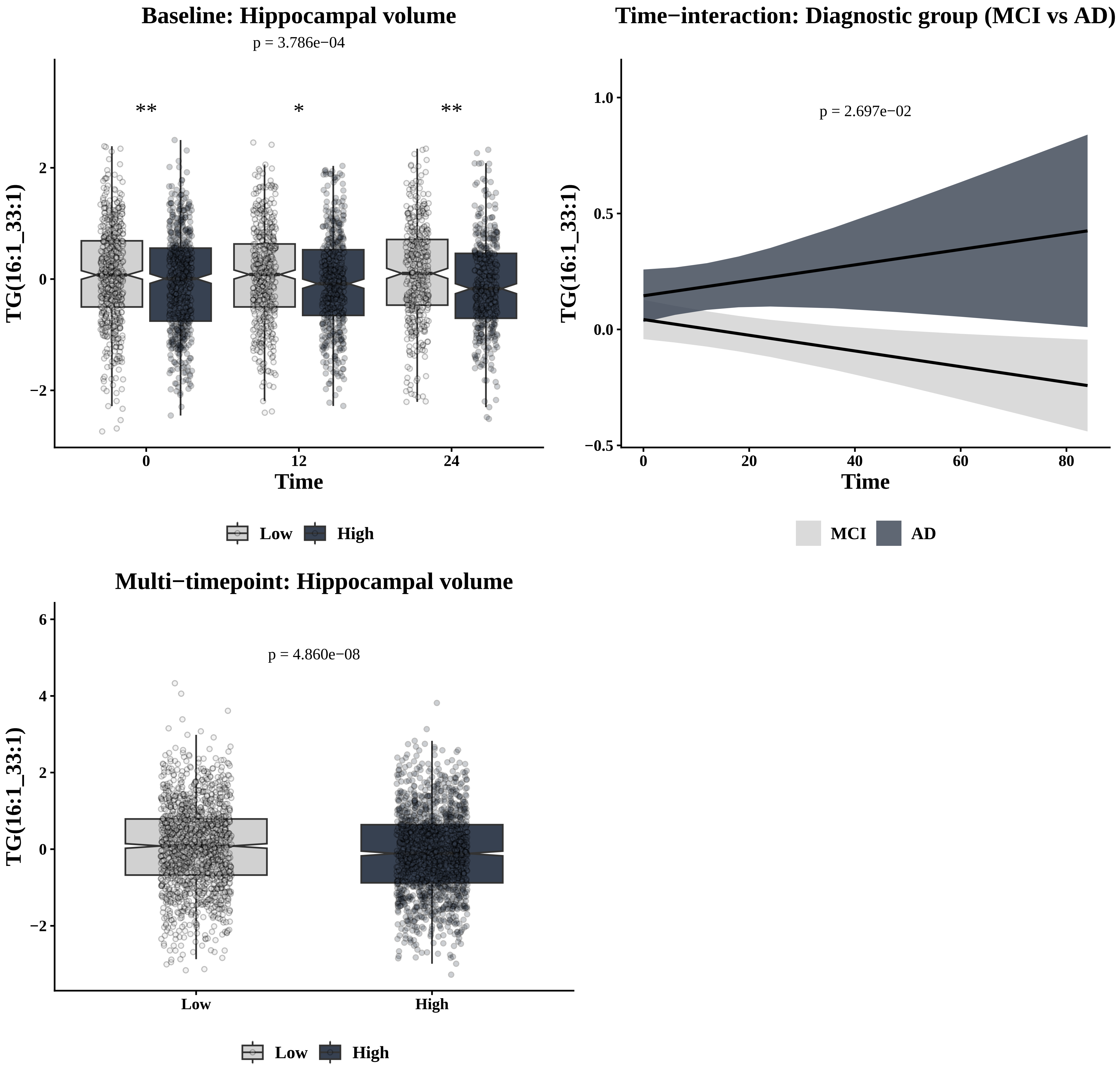

Baseline: Hippocampal volume
p = 3.786e−04
**
*
**
TG(16:1_33:1)
2
0
−2
0
12
24
Time
Low
High
Time−interaction: Diagnostic group (MCI vs AD)
p = 2.697e−02
1.0
0.5
0.0
−0.5
0
20
40
60
80
Time
MCI
AD
Multi−timepoint: Hippocampal volume
p = 4.860e−08
6
4
2
0
−2
Low
High

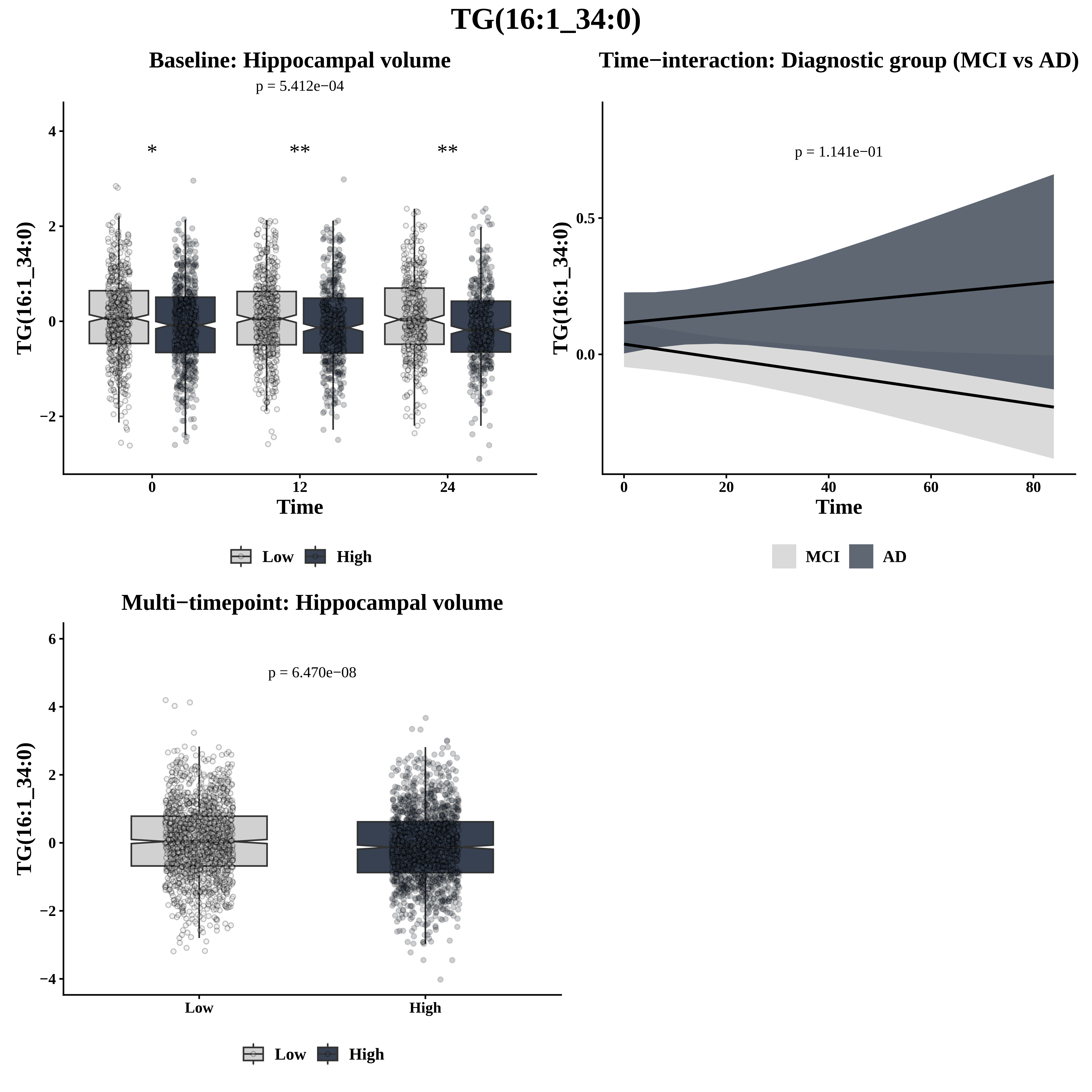
TG(16:1_34:0)
Baseline: Hippocampal volume
p = 5.412e−04
*
**
**
TG(16:1_34:0)
Time
0
12
24
Low
High
Time−interaction: Diagnostic group (MCI vs AD)
p = 1.141e−01
TG(16:1_34:0)
Time
MCI
AD
Multi−timepoint: Hippocampal volume
p = 6.470e−08
TG(16:1_34:0)
Low
High
Low
High

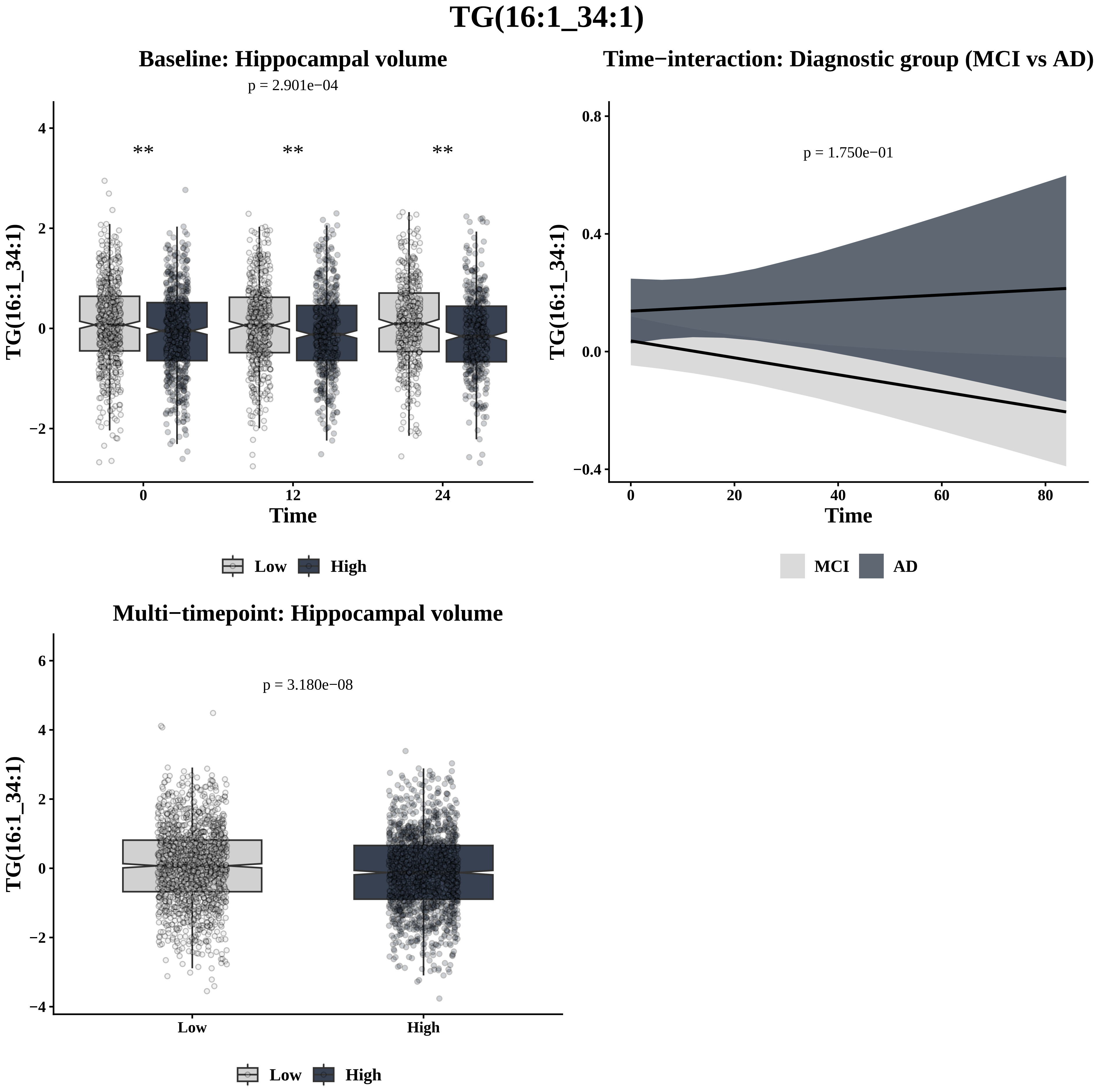
TG(16:1_34:1)
Baseline: Hippocampal volume
p = 2.901e−04
**
**
**
TG(16:1_34:1)
Time
0
12
24
Low
High
Time−interaction: Diagnostic group (MCI vs AD)
p = 1.750e−01
TG(16:1_34:1)
Time
MCI
AD
Multi−timepoint: Hippocampal volume
p = 3.180e−08
TG(16:1_34:1)
Low
High
Low
High

# TG(16:1_34:2)

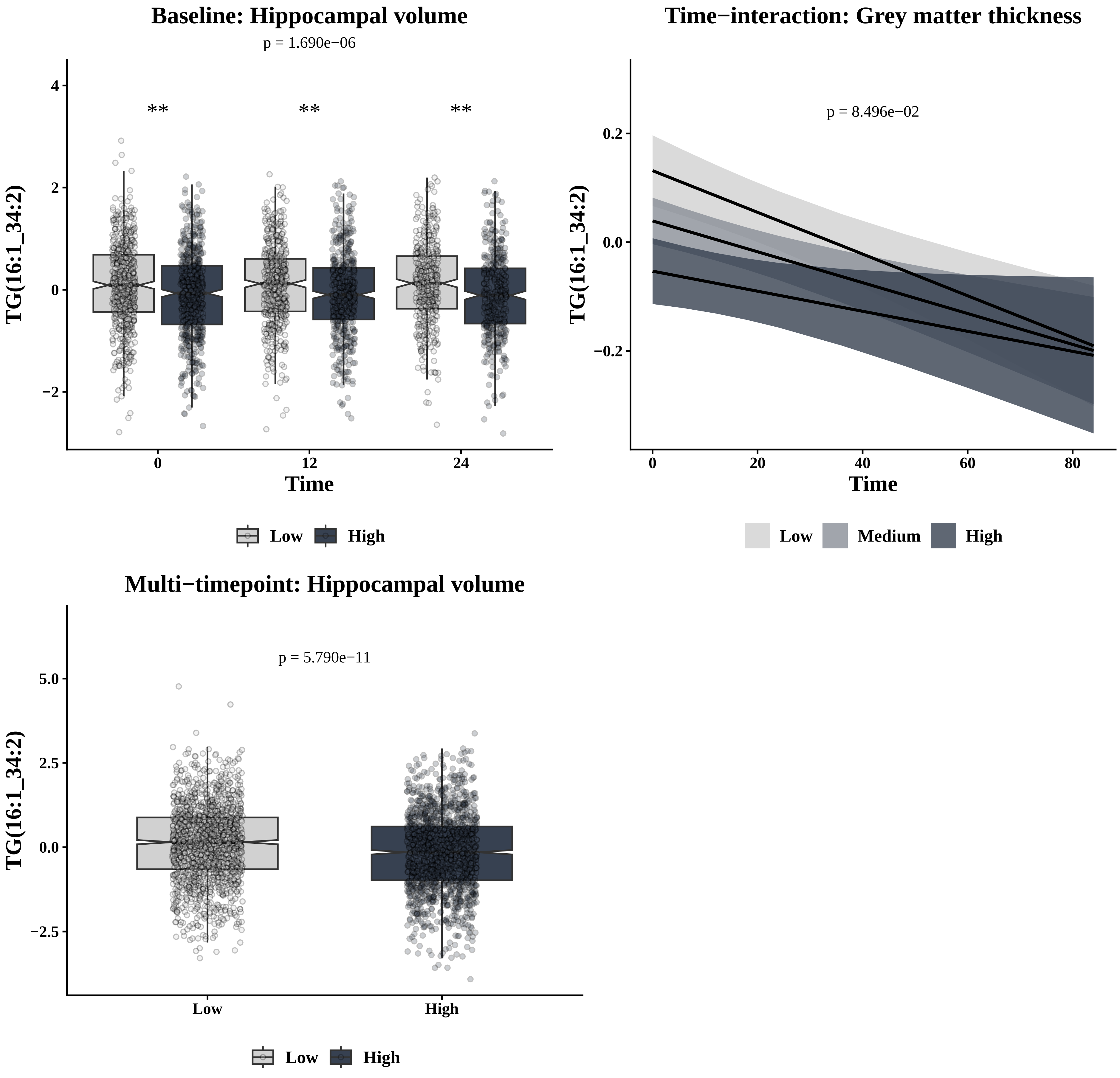

Baseline: Hippocampal volume
p = 1.690e−06
**
**
**
TG(16:1_34:2)
Time
0
12
24
Low
High
Time−interaction: Grey matter thickness
p = 8.496e−02
TG(16:1_34:2)
Time
0
20
40
60
80
Low
Medium
High
Multi−timepoint: Hippocampal volume
p = 5.790e−11
TG(16:1_34:2)
Low
High

# TG(16:1_34:3)

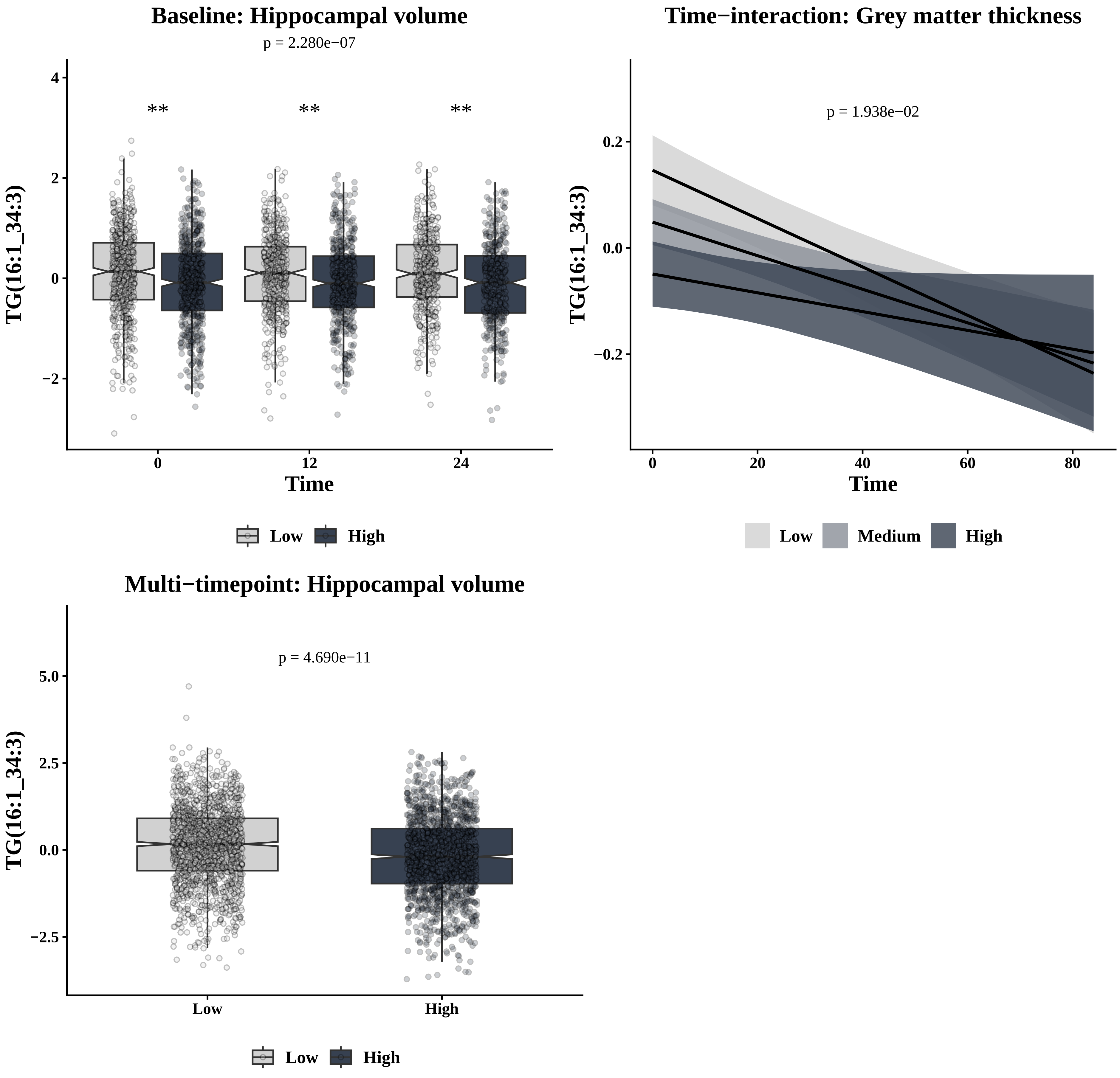

Baseline: Hippocampal volume
p = 2.280e−07
TG(16:1_34:3)
Time
Low
High
Time−interaction: Grey matter thickness
p = 1.938e−02
TG(16:1_34:3)
Time
Low
Medium
High
Multi−timepoint: Hippocampal volume
p = 4.690e−11
TG(16:1_34:3)
Low
High

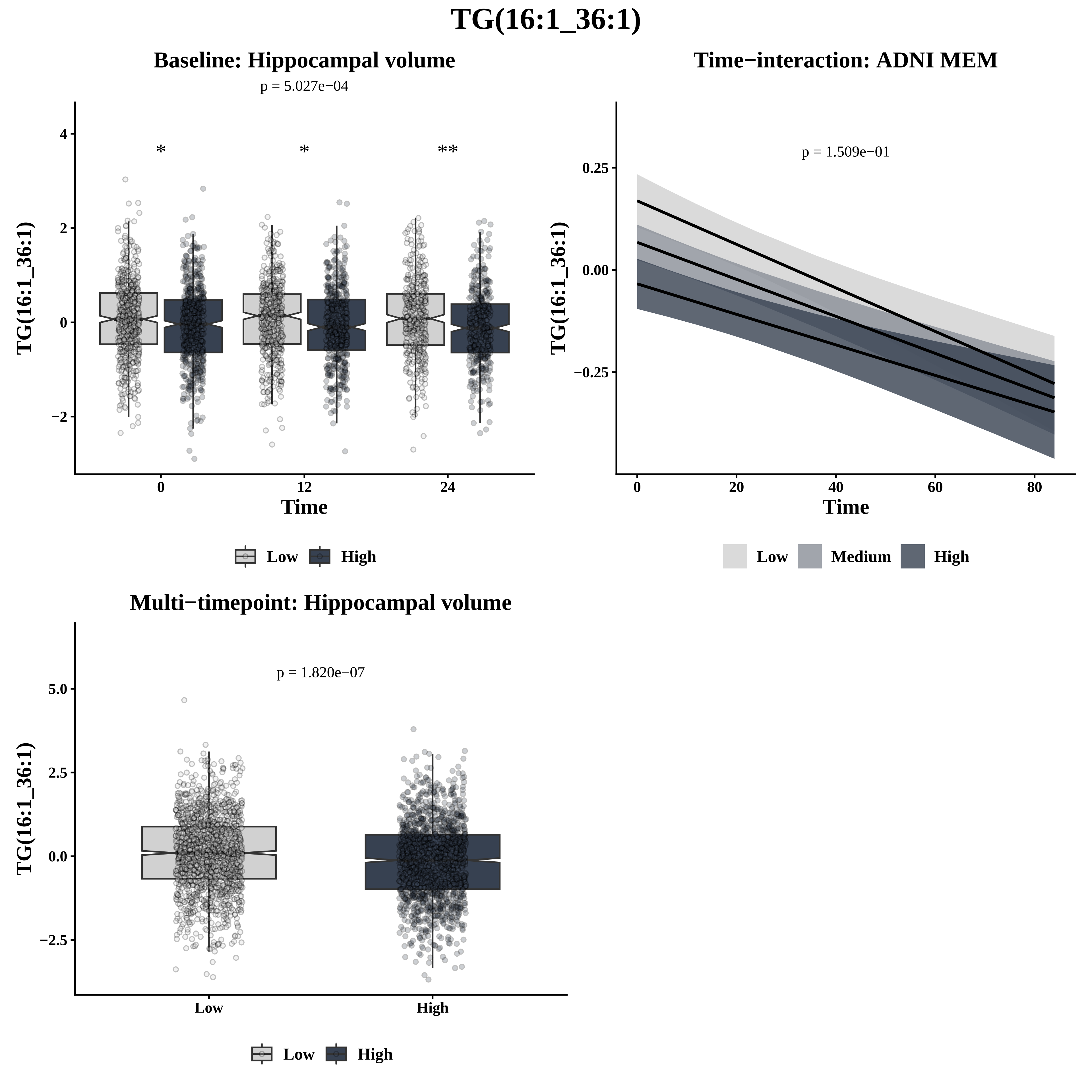
TG(16:1_36:1)
Baseline: Hippocampal volume
p = 5.027e−04
*
*
**
4
2
0
−2
TG(16:1_36:1)
0
12
24
Time
Low
High
Time−interaction: ADNI MEM
p = 1.509e−01
0.25
0.00
−0.25
TG(16:1_36:1)
0
20
40
60
80
Time
Low
Medium
High
Multi−timepoint: Hippocampal volume
p = 1.820e−07
5.0
2.5
0.0
−2.5
TG(16:1_36:1)
Low
High
Low
High

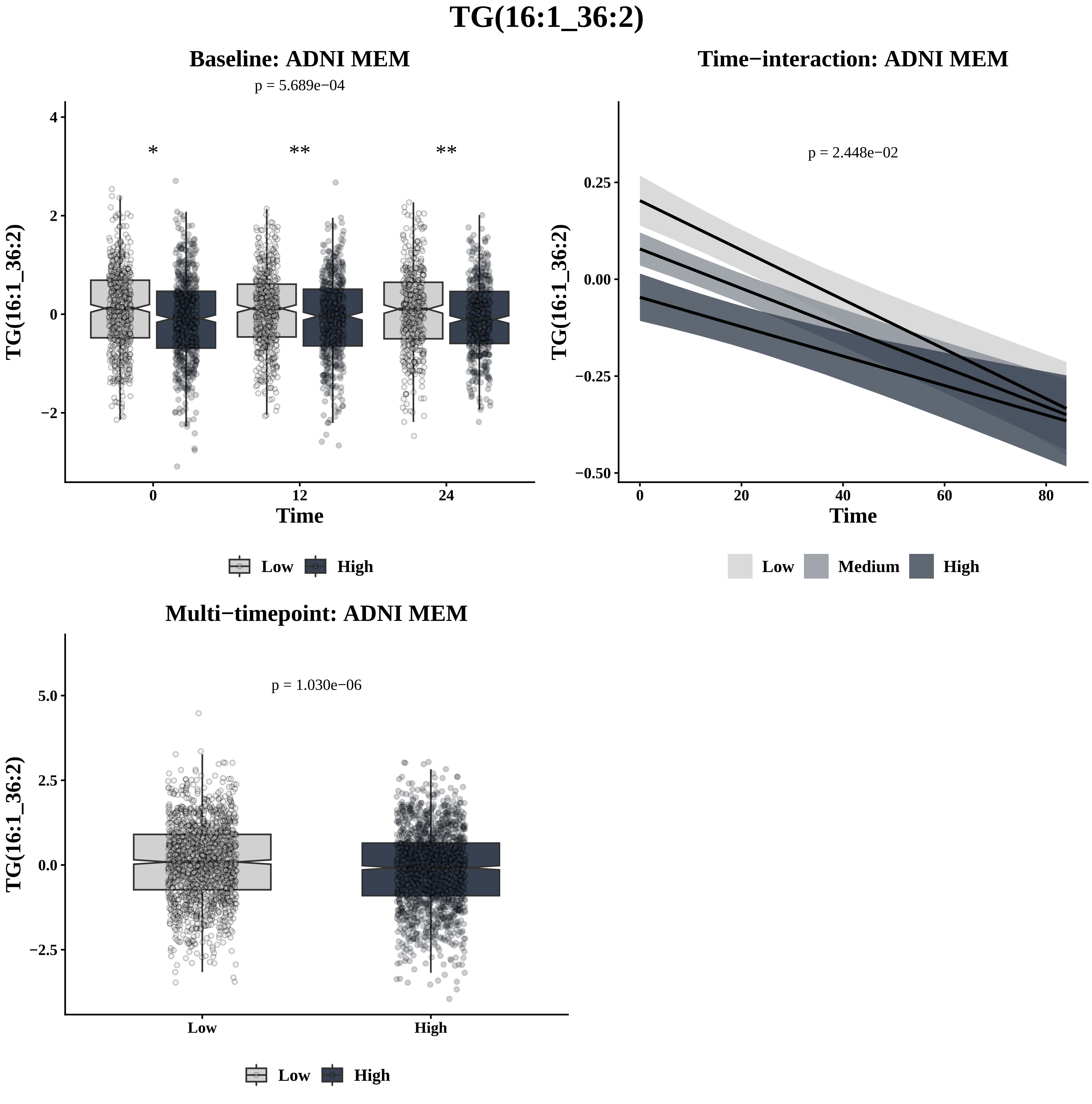

TG(16:1_36:2)
Baseline: ADNI MEM
p = 5.689e−04
*
**
**
TG(16:1_36:2)
Time
Low
High
Time−interaction: ADNI MEM
p = 2.448e−02
TG(16:1_36:2)
Time
Low
Medium
High
Multi−timepoint: ADNI MEM
p = 1.030e−06
TG(16:1_36:2)
Low
High
Low
High

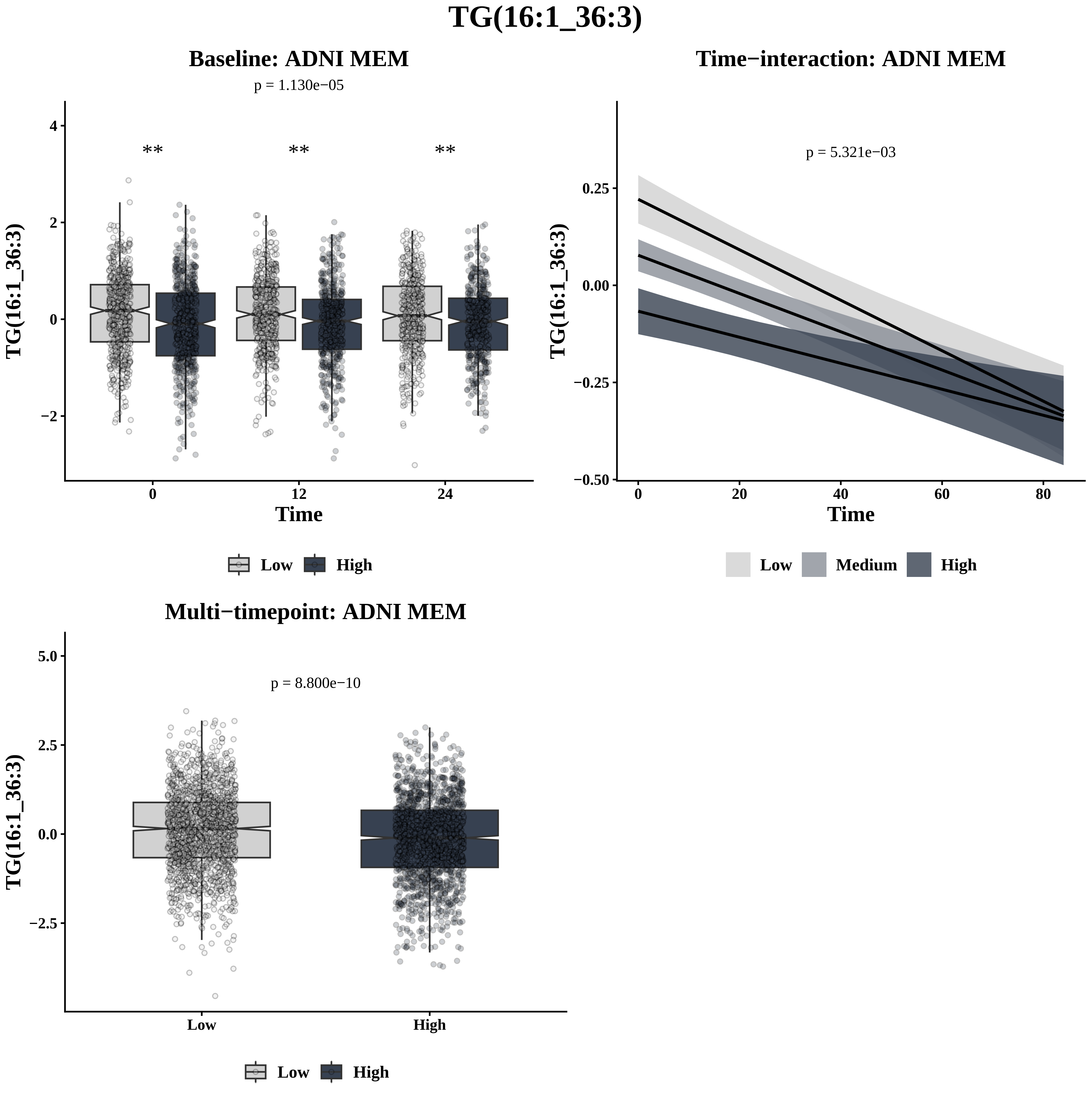

TG(16:1_36:3)
Baseline: ADNI MEM
p = 1.130e−05
**
**
**
TG(16:1_36:3)
4
2
0
−2
0
12
24
Time
Low
High
Time−interaction: ADNI MEM
p = 5.321e−03
TG(16:1_36:3)
0.25
0.00
−0.25
−0.50
0
20
40
60
80
Time
Low
Medium
High
Multi−timepoint: ADNI MEM
p = 8.800e−10
TG(16:1_36:3)
5.0
2.5
0.0
−2.5
Low
High
Low
High

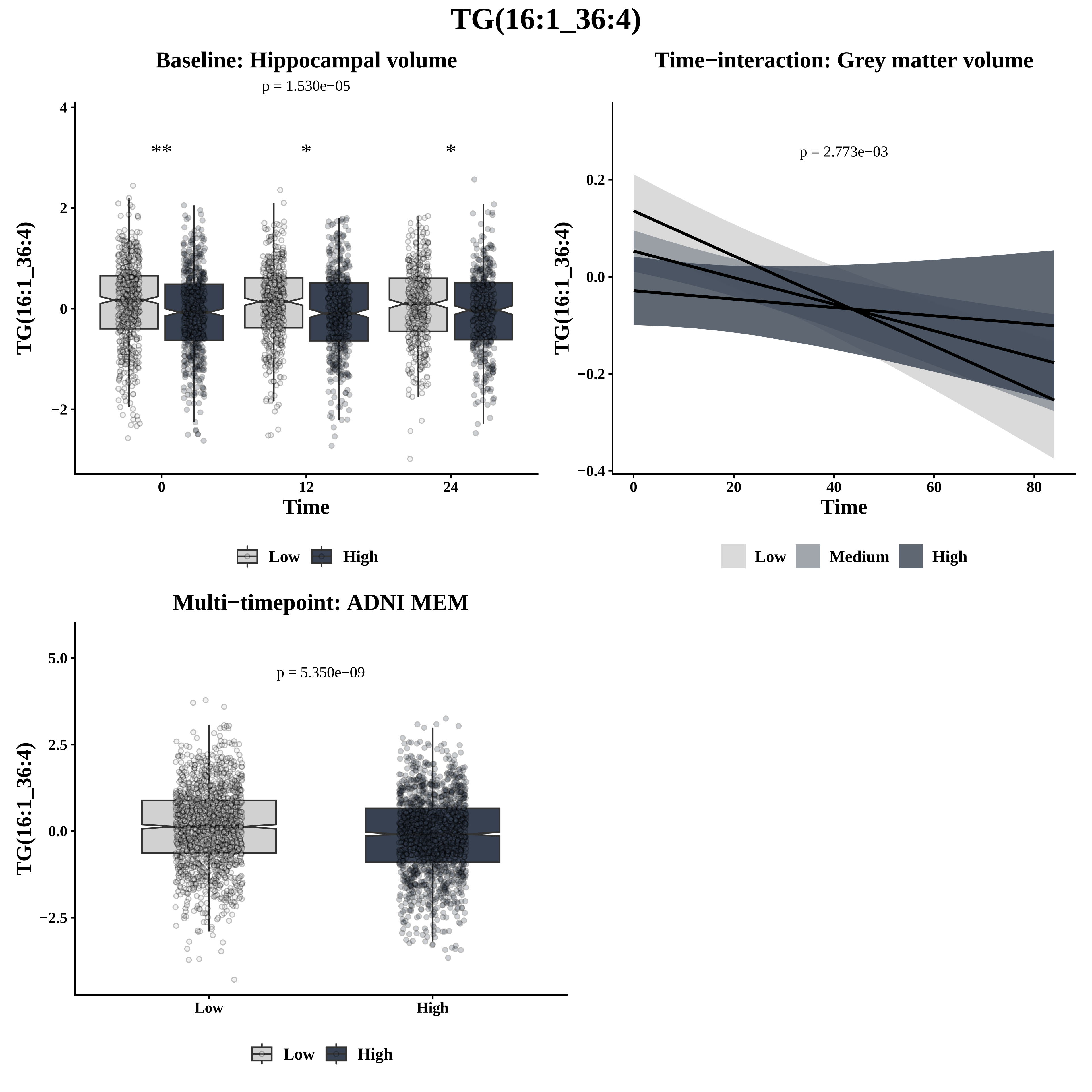

TG(16:1_36:4)
Baseline: Hippocampal volume
p = 1.530e−05
**
*
*
TG(16:1_36:4)
4
2
0
−2
0
12
24
Time
Low
High
Time−interaction: Grey matter volume
p = 2.773e−03
TG(16:1_36:4)
0.2
0.0
−0.2
−0.4
0
20
40
60
80
Time
Low
Medium
High
Multi−timepoint: ADNI MEM
p = 5.350e−09
TG(16:1_36:4)
5.0
2.5
0.0
−2.5
Low
High
Low
High

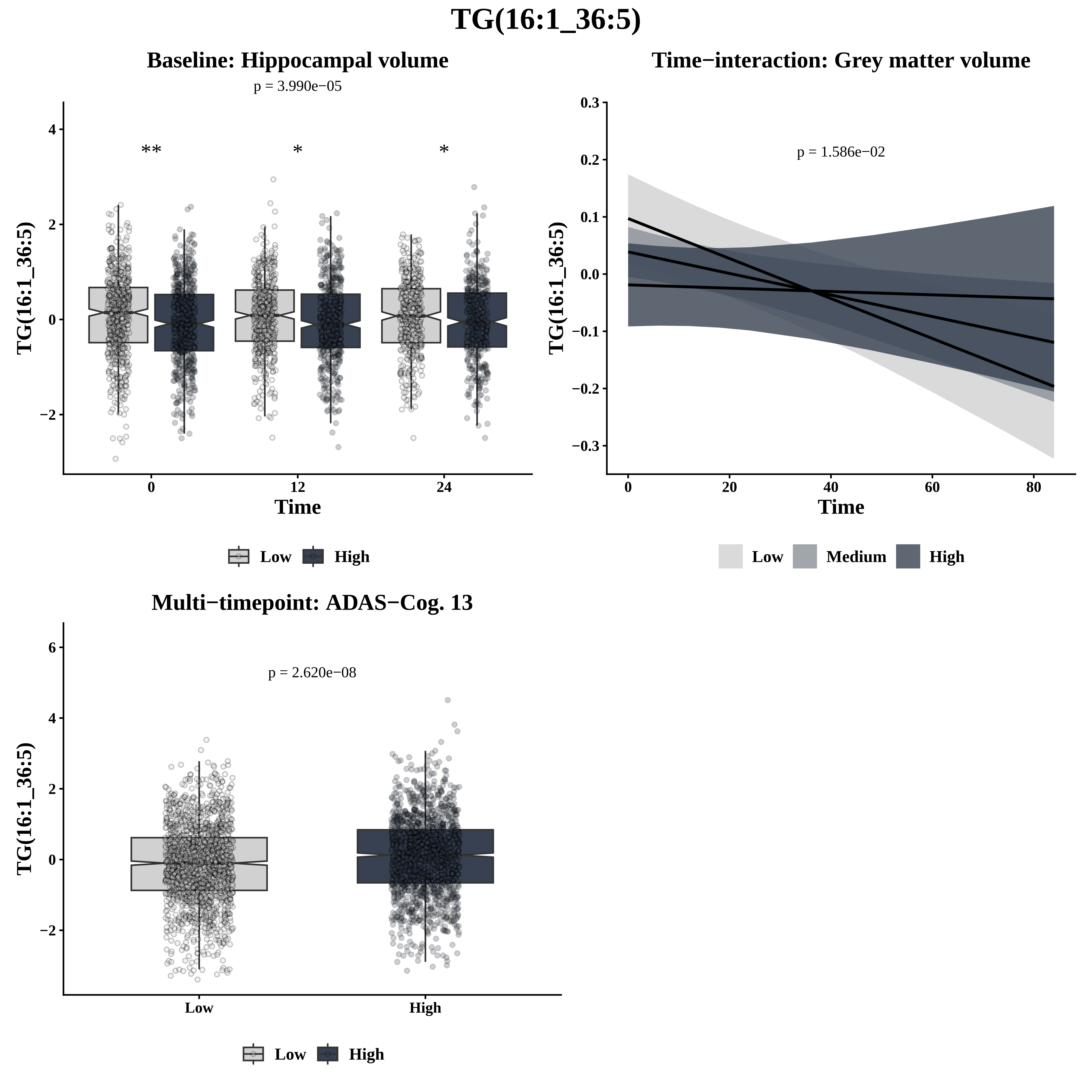
TG(16:1_36:5)
Baseline: Hippocampal volume
p = 3.990e−05
**
*
*
TG(16:1_36:5)
Time
Low
High
Time−interaction: Grey matter volume
p = 1.586e−02
TG(16:1_36:5)
Time
Low
Medium
High
Multi−timepoint: ADAS−Cog. 13
p = 2.620e−08
TG(16:1_36:5)
Low
High
Low
High

# TG(16:1_38:3)

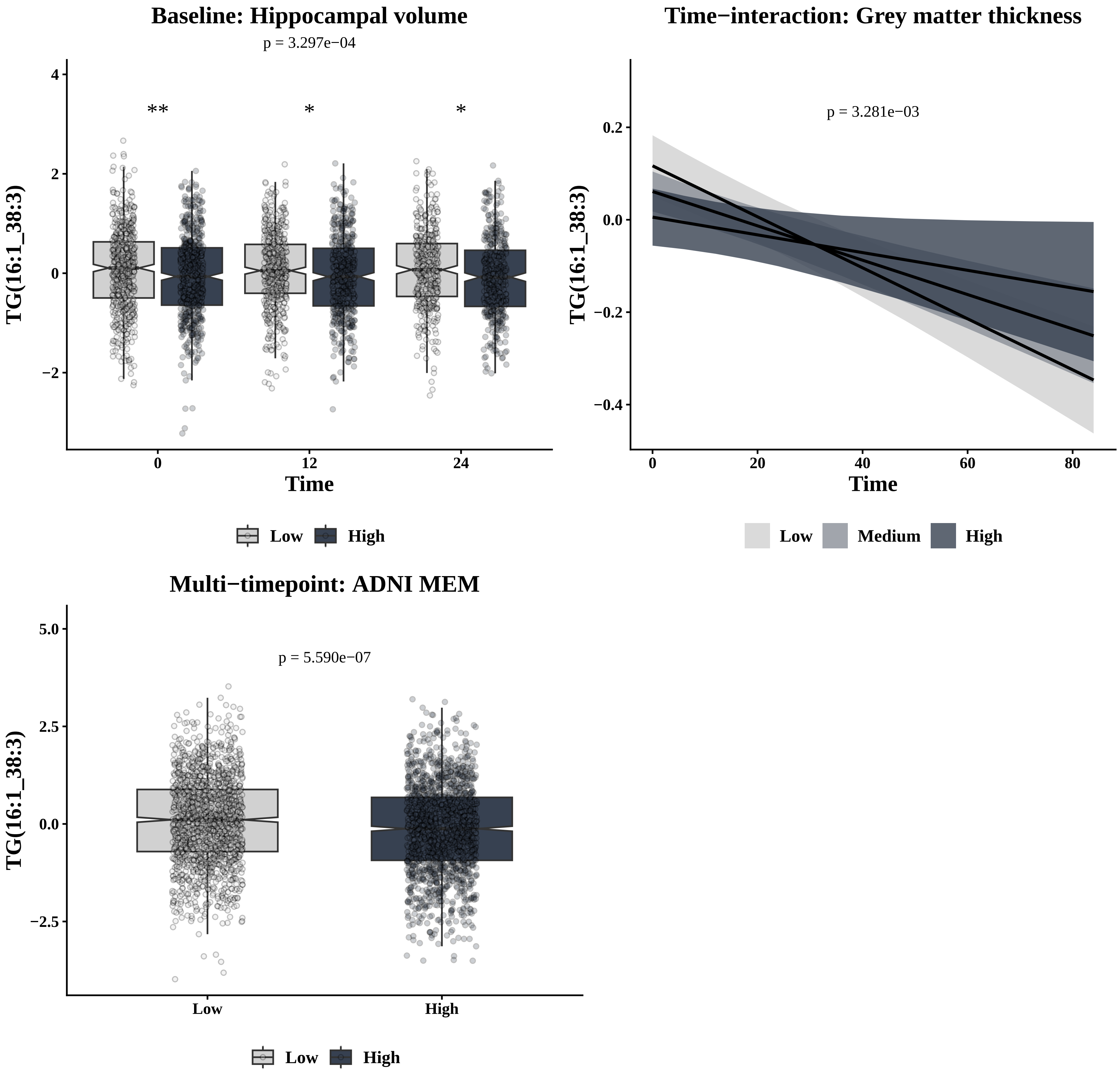
Baseline: Hippocampal volume
p = 3.297e−04
**
*
*
TG(16:1_38:3)
Time
0
12
24
Low
High
Time−interaction: Grey matter thickness
p = 3.281e−03
TG(16:1_38:3)
Time
Low
Medium
High
Multi−timepoint: ADNI MEM
p = 5.590e−07
TG(16:1_38:3)
Low
High
Low
High

# TG(16:1_38:4)

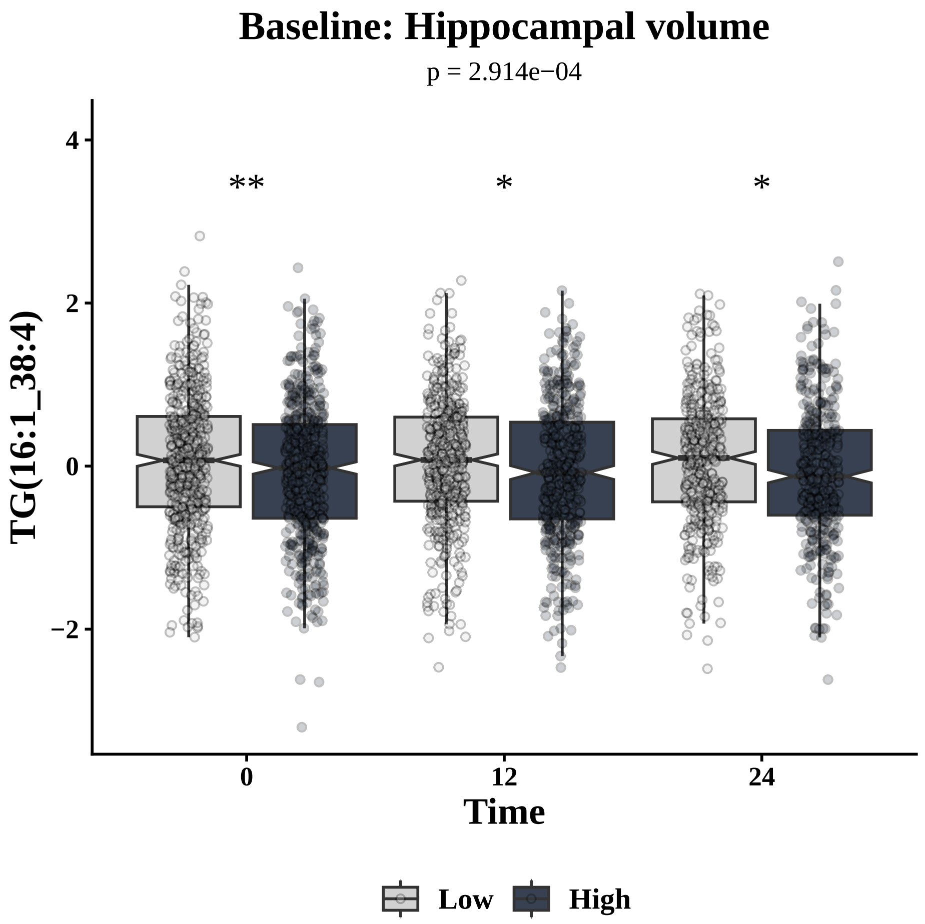

Baseline: Hippocampal volume
p = 2.914e−04
**
*
*
TG(16:1_38:4)
Time
Low
High


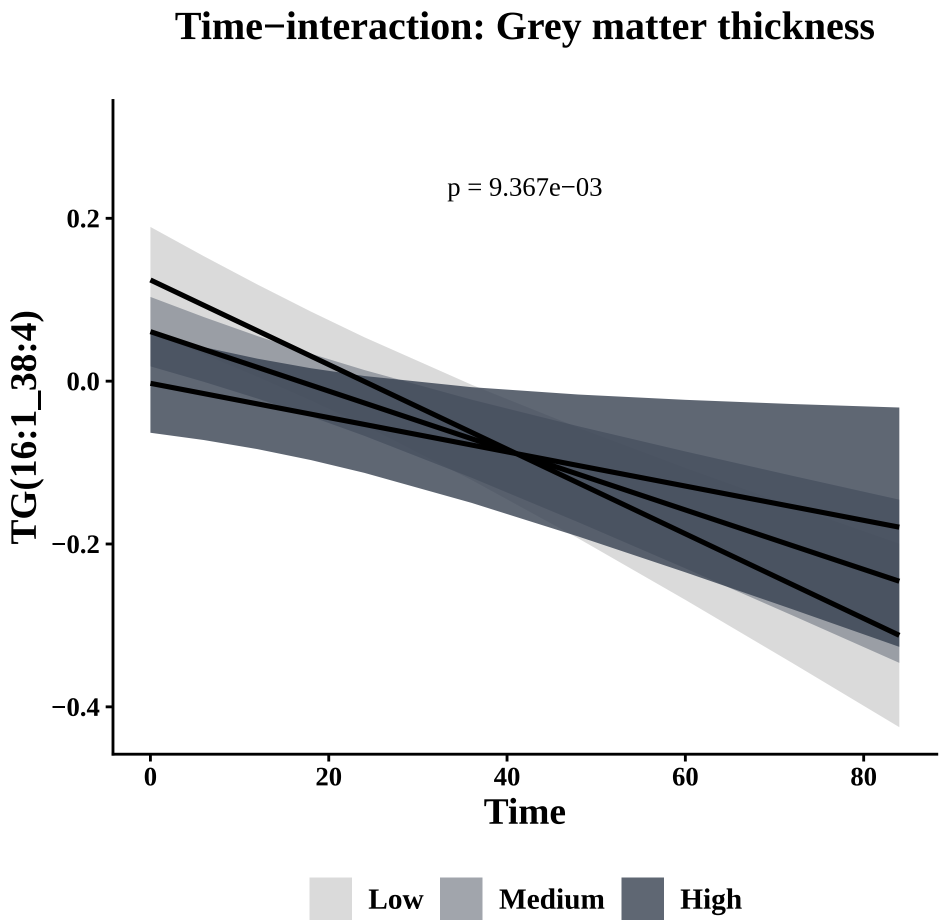

Time−interaction: Grey matter thickness
p = 9.367e−03
TG(16:1_38:4)
Time
Low
Medium
High


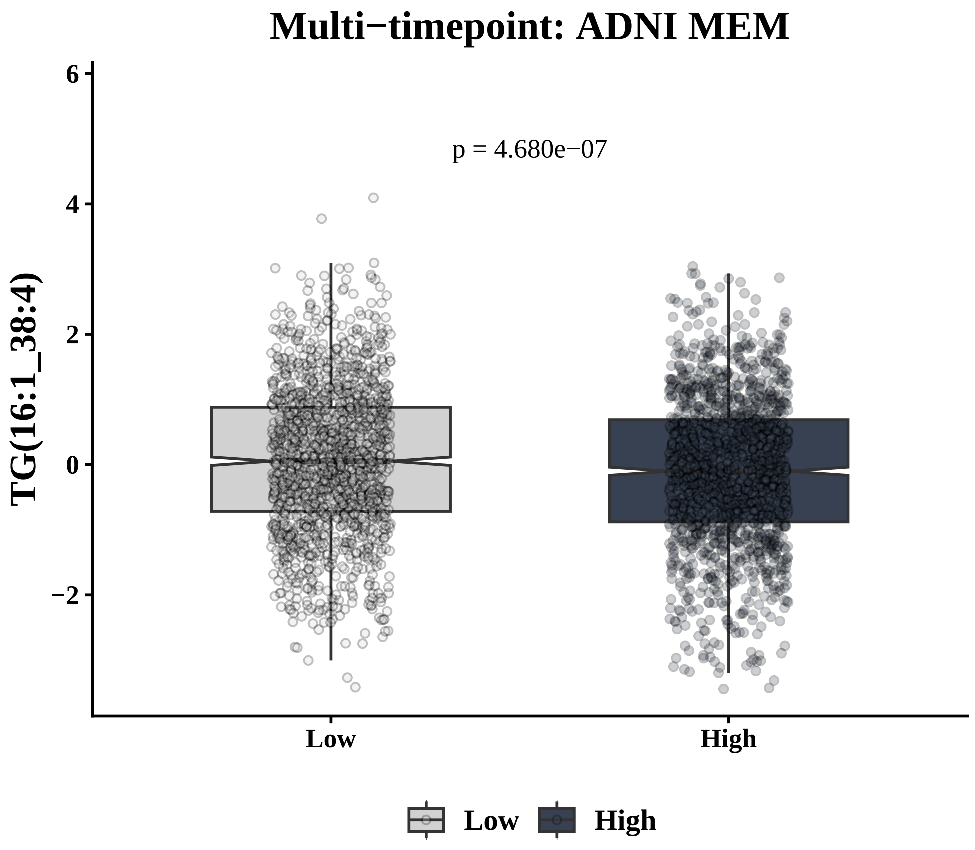

Multi−timepoint: ADNI MEM
p = 4.680e−07
TG(16:1_38:4)
Low
High

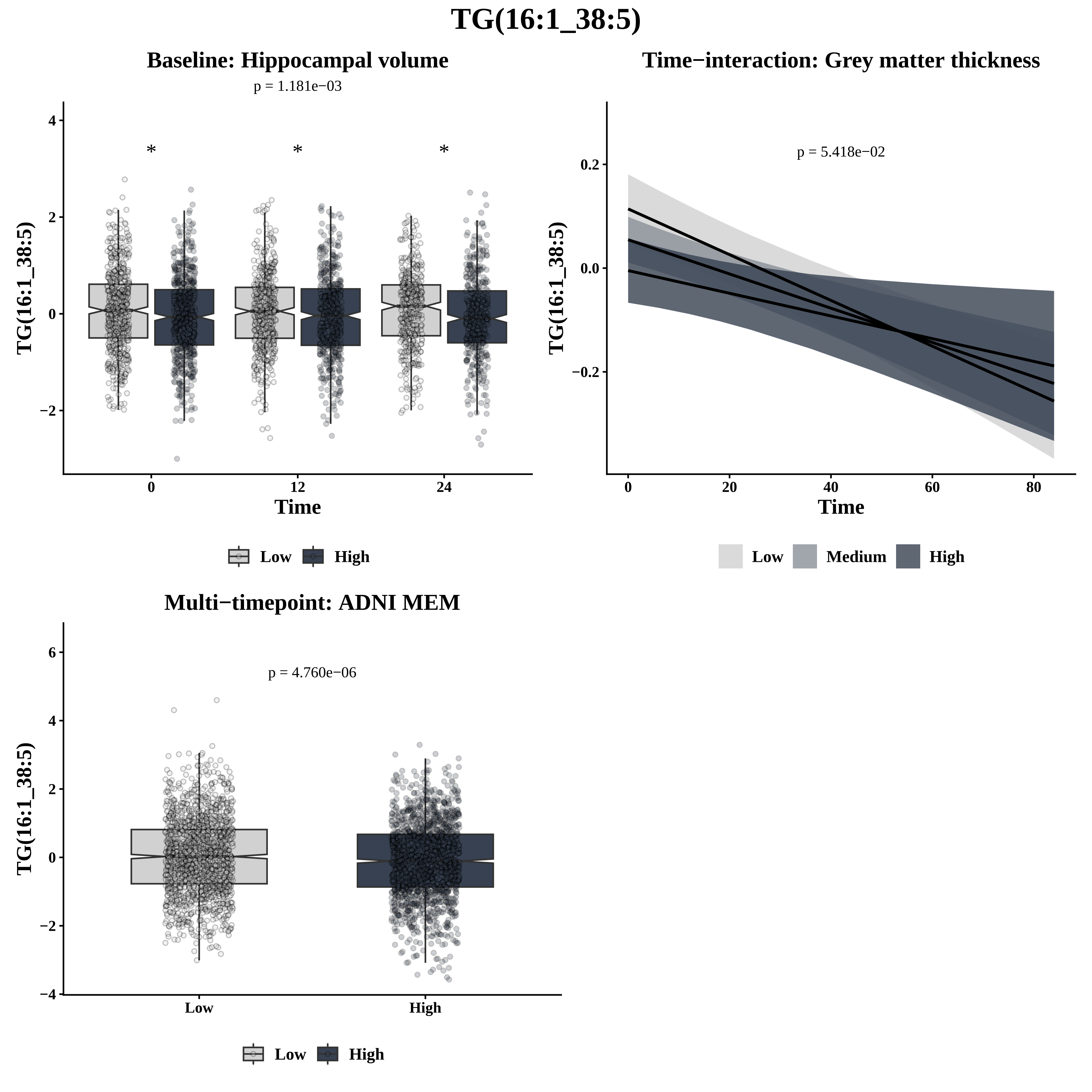
TG(16:1_38:5)
Baseline: Hippocampal volume
p = 1.181e−03
TG(16:1_38:5)
Time
Low
High
Time−interaction: Grey matter thickness
p = 5.418e−02
TG(16:1_38:5)
Time
Low
Medium
High
Multi−timepoint: ADNI MEM
p = 4.760e−06
TG(16:1_38:5)
Low
High

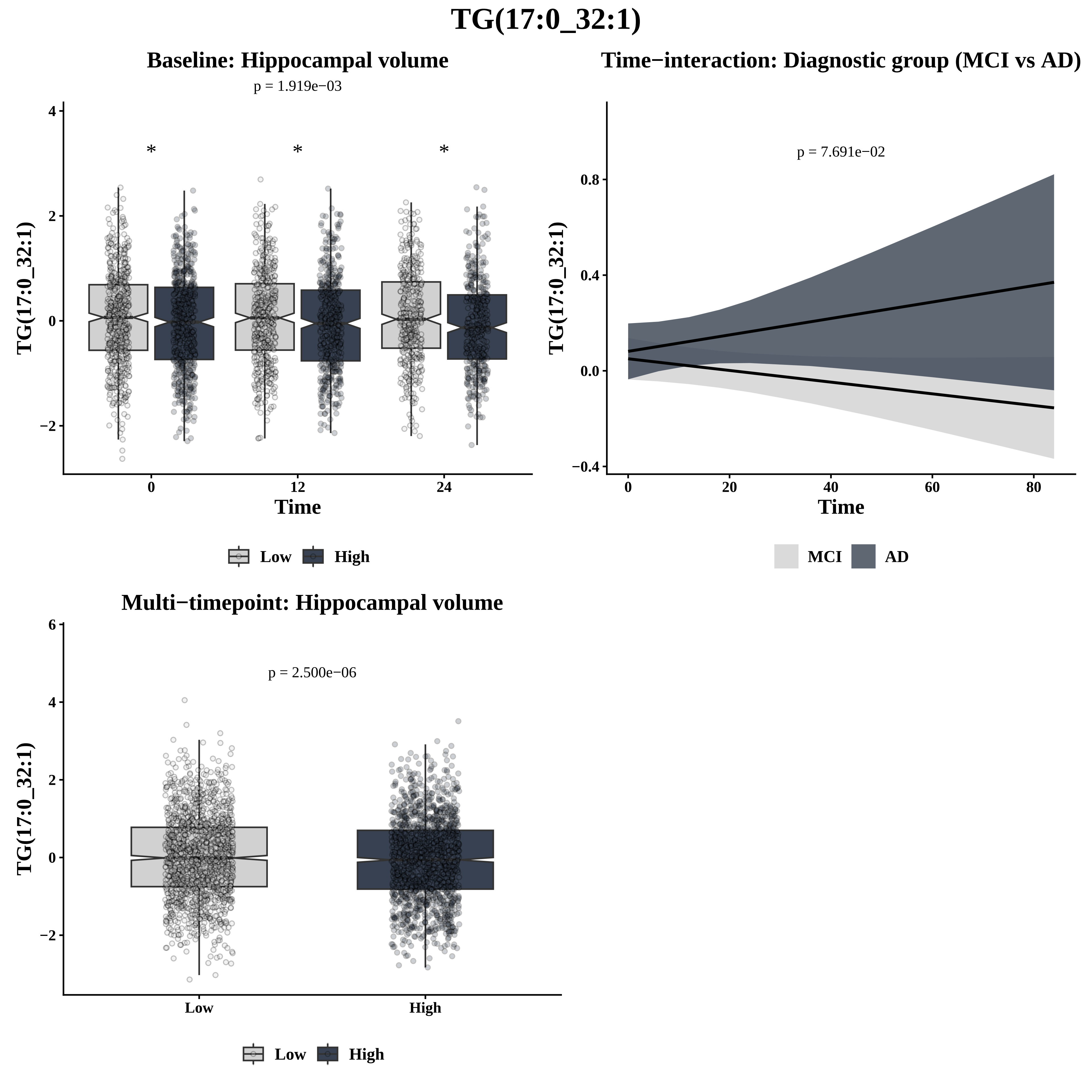

TG(17:0_32:1)
Baseline: Hippocampal volume
p = 1.919e−03
*
*
*
TG(17:0_32:1)
Time
0
12
24
4
2
0
−2
Low
High
Time−interaction: Diagnostic group (MCI vs AD)
p = 7.691e−02
TG(17:0_32:1)
0.8
0.4
0.0
−0.4
0
20
40
60
80
Time
MCI
AD
Multi−timepoint: Hippocampal volume
p = 2.500e−06
TG(17:0_32:1)
6
4
2
0
−2
Low
High
Low
High

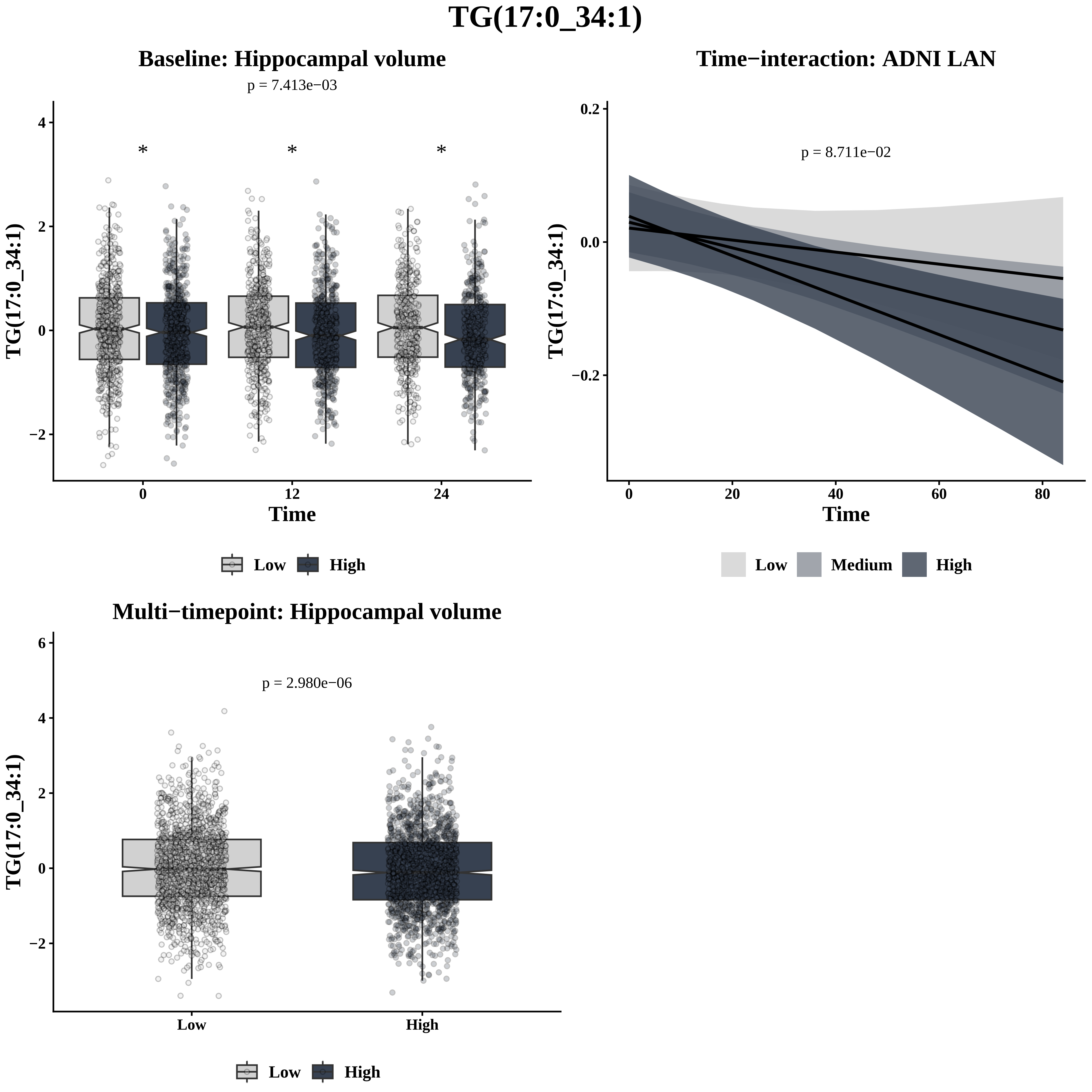
TG(17:0_34:1)
Baseline: Hippocampal volume
p = 7.413e−03
*
*
*
TG(17:0_34:1)
Time
0
12
24
Low
High
Time−interaction: ADNI LAN
p = 8.711e−02
TG(17:0_34:1)
Time
Low
Medium
High
Multi−timepoint: Hippocampal volume
p = 2.980e−06
TG(17:0_34:1)
Low
High
Low
High

# TG(17:0_34:2)

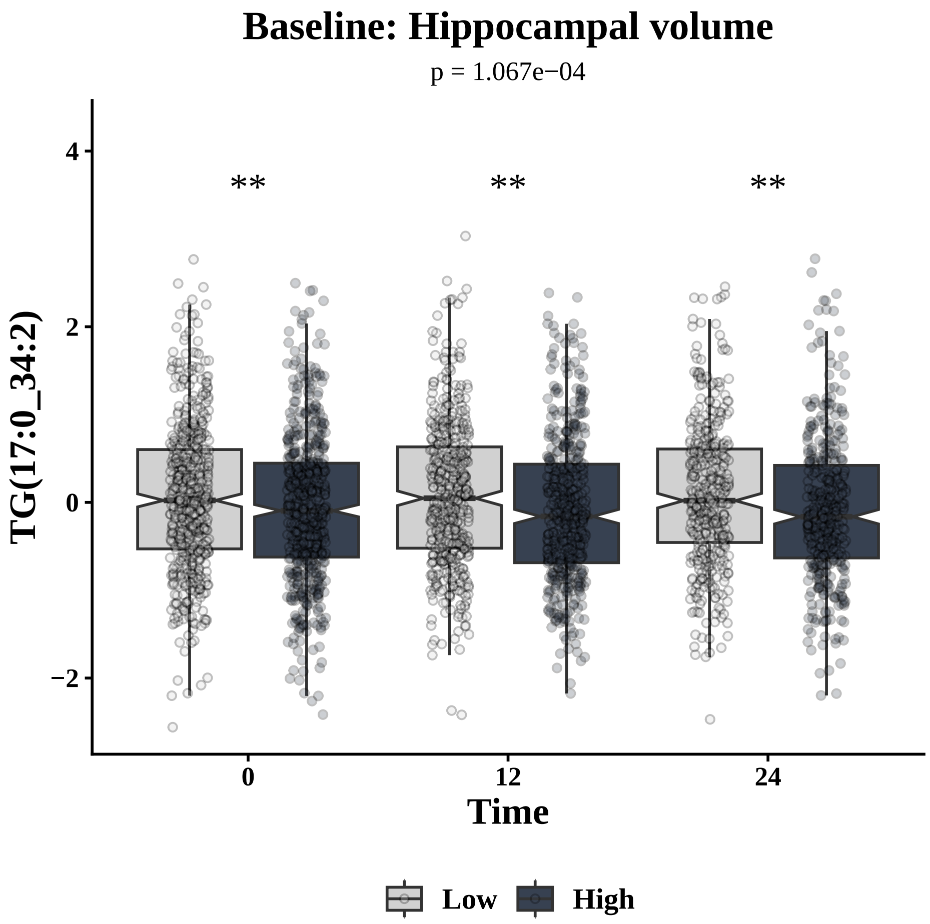

Baseline: Hippocampal volume
p = 1.067e−04
**
**
**
TG(17:0_34:2)
4
2
0
−2
0
12
24
Time
Low
High


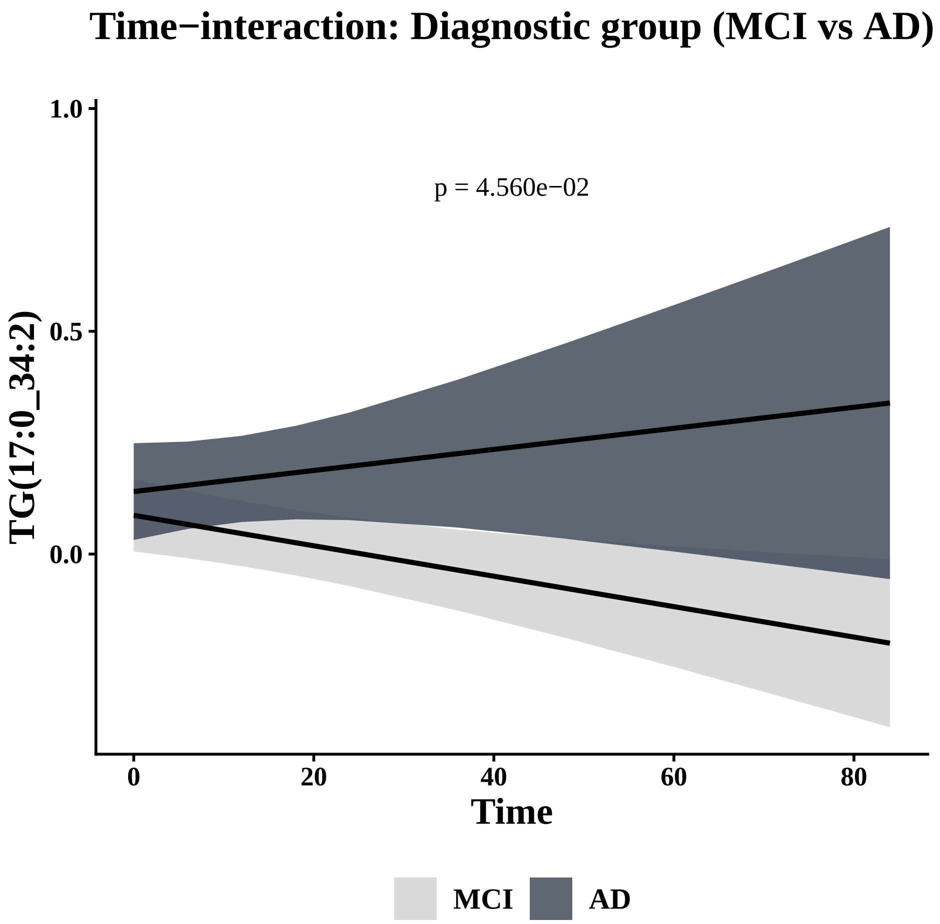

Time−interaction: Diagnostic group (MCI vs AD)
p = 4.560e−02
TG(17:0_34:2)
1.0
0.5
0.0
0
20
40
60
80
Time
MCI
AD


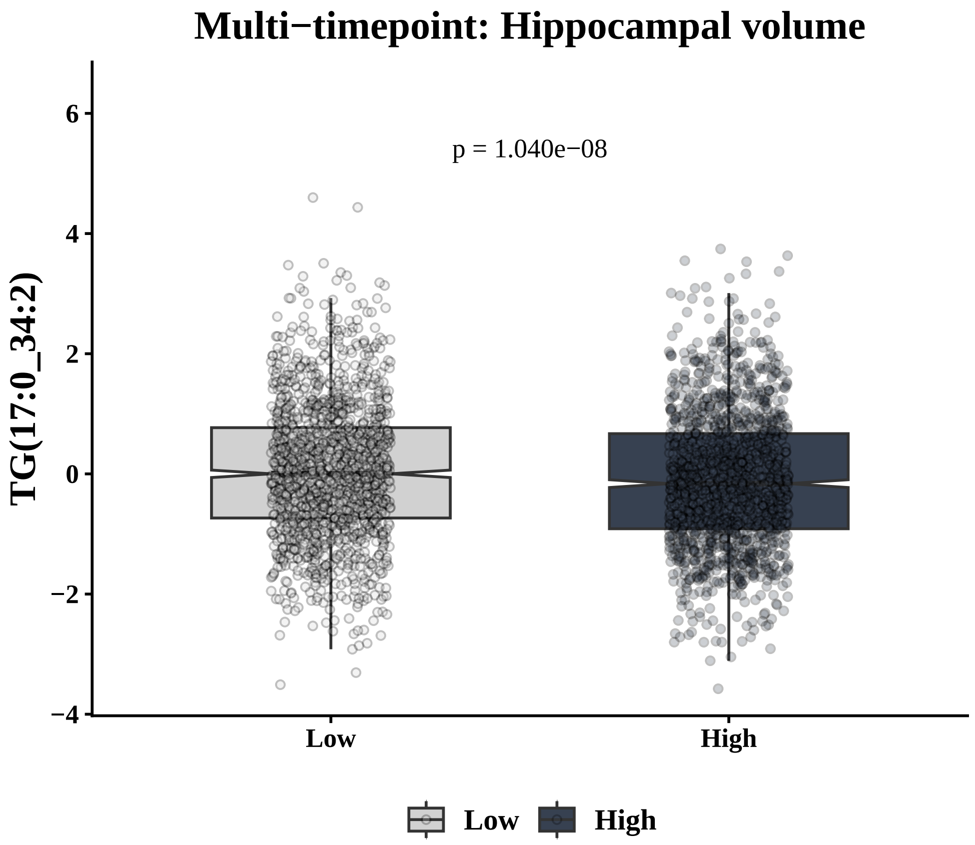

Multi−timepoint: Hippocampal volume
p = 1.040e−08
TG(17:0_34:2)
6
4
2
0
−2
−4
Low
High
Low
High

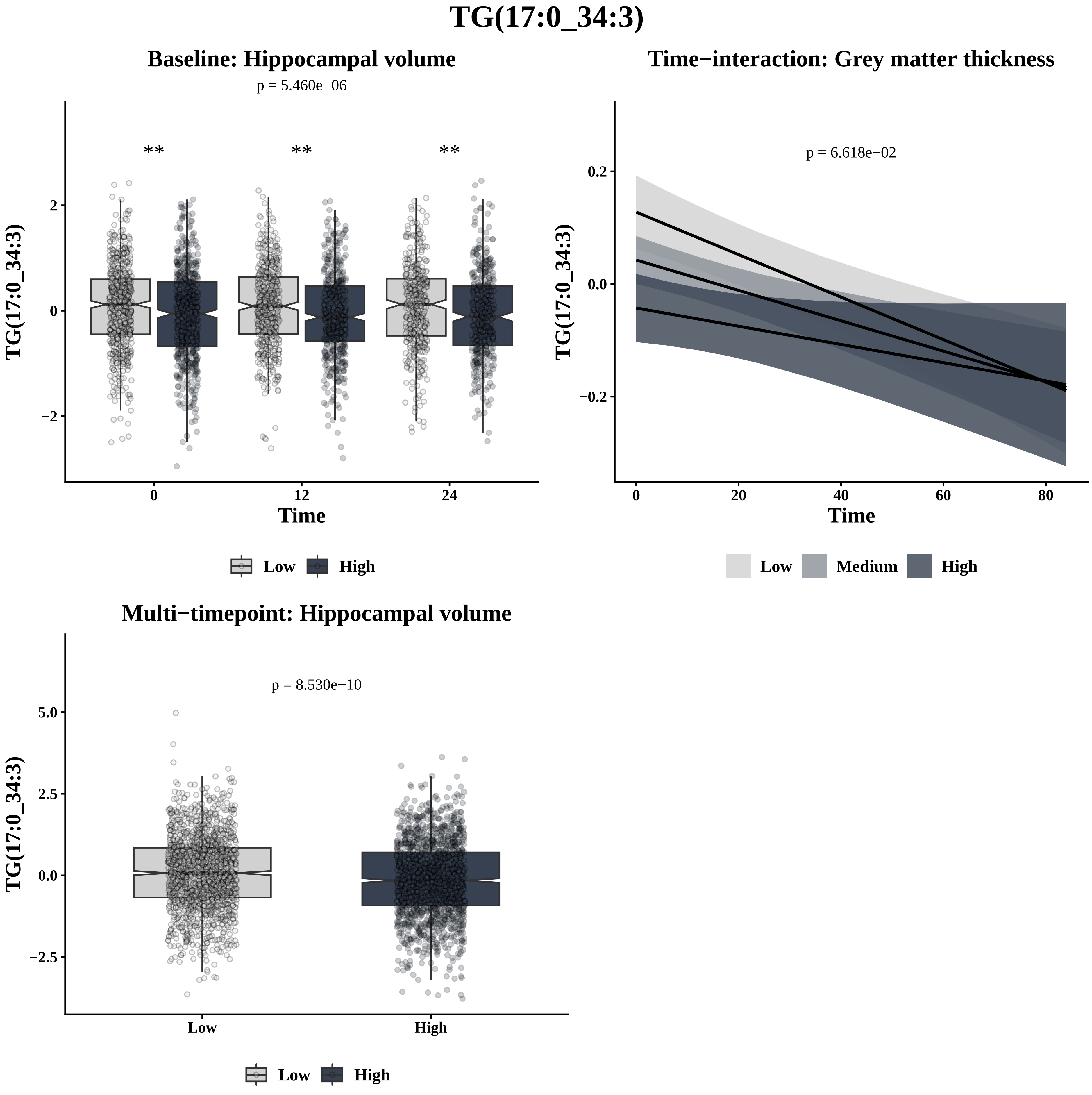

TG(17:0_34:3)
Baseline: Hippocampal volume
p = 5.460e−06
**
**
**
TG(17:0_34:3)
Time
0
12
24
Low
High
Time−interaction: Grey matter thickness
p = 6.618e−02
TG(17:0_34:3)
Time
Low
Medium
High
Multi−timepoint: Hippocampal volume
p = 8.530e−10
TG(17:0_34:3)
Low
High
Low
High

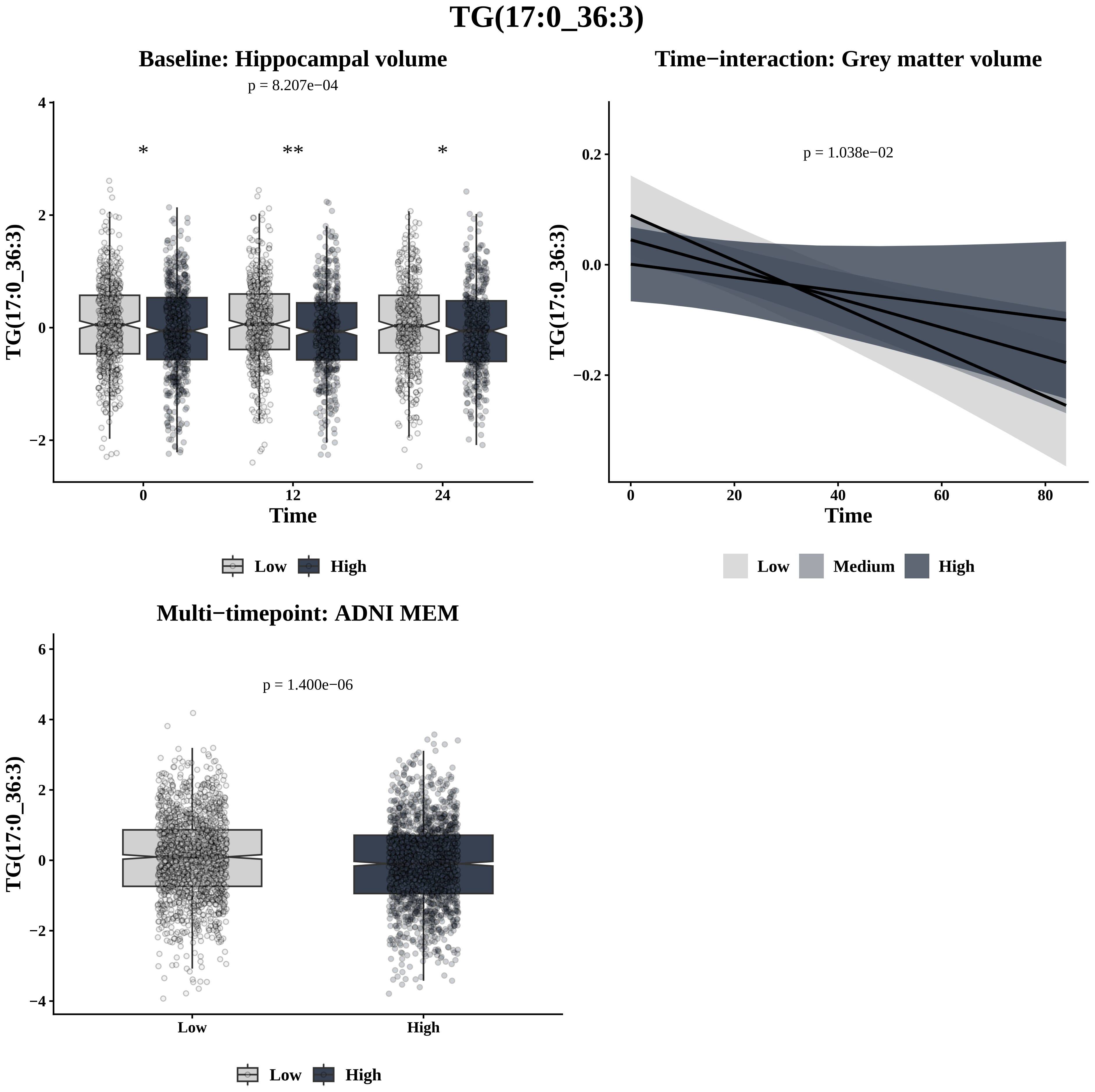

TG(17:0_36:3)
Baseline: Hippocampal volume
p = 8.207e−04
*
**
*
TG(17:0_36:3)
Time
0
12
24
Low
High
Time−interaction: Grey matter volume
p = 1.038e−02
TG(17:0_36:3)
Time
Low
Medium
High
Multi−timepoint: ADNI MEM
p = 1.400e−06
TG(17:0_36:3)
Low
High

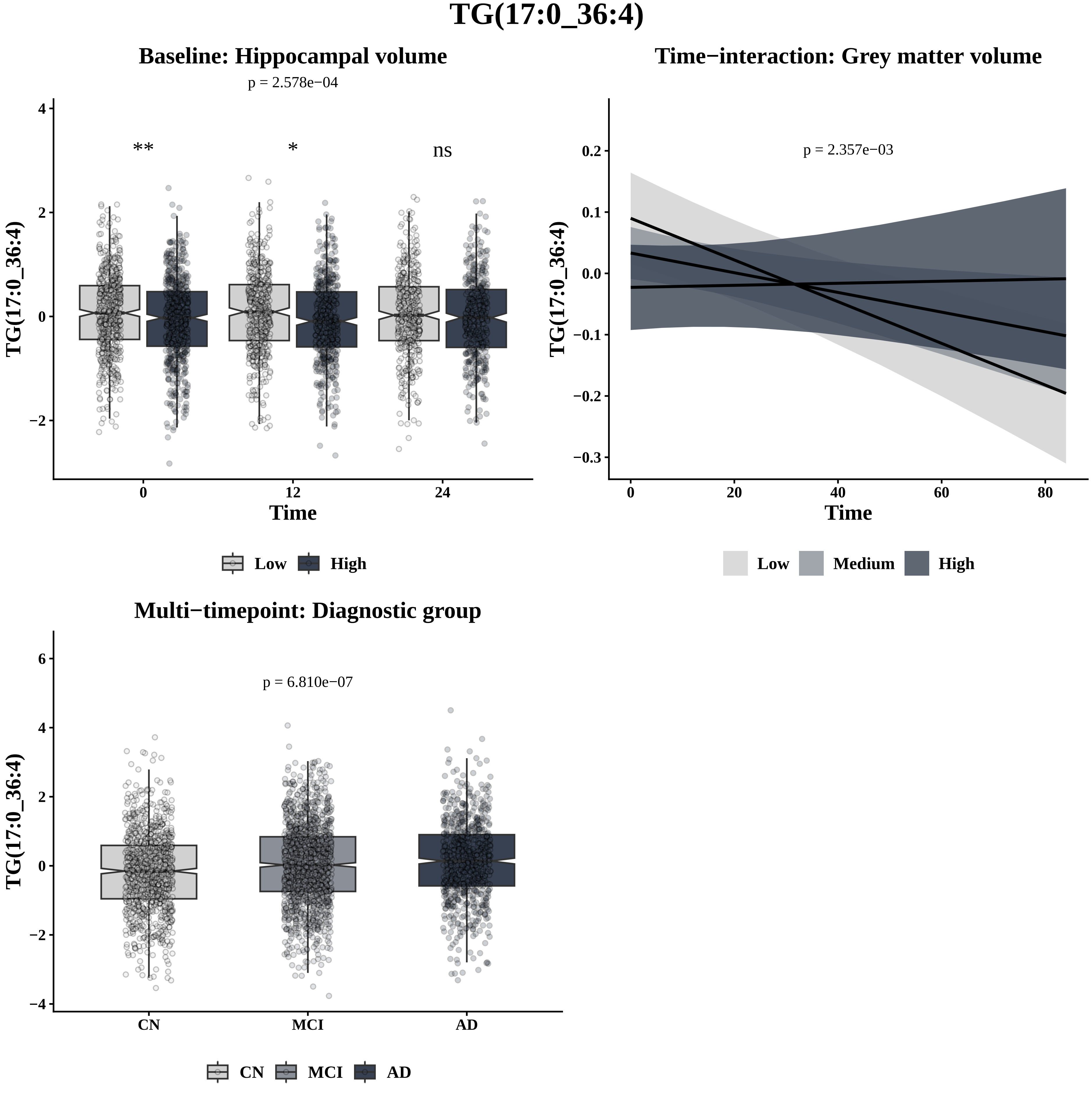
TG(17:0_36:4)
Baseline: Hippocampal volume
p = 2.578e−04
**
*
ns
TG(17:0_36:4)
Time
0
12
24
Low
High
Time−interaction: Grey matter volume
p = 2.357e−03
TG(17:0_36:4)
Time
Low
Medium
High
Multi−timepoint: Diagnostic group
p = 6.810e−07
TG(17:0_36:4)
CN
MCI
AD

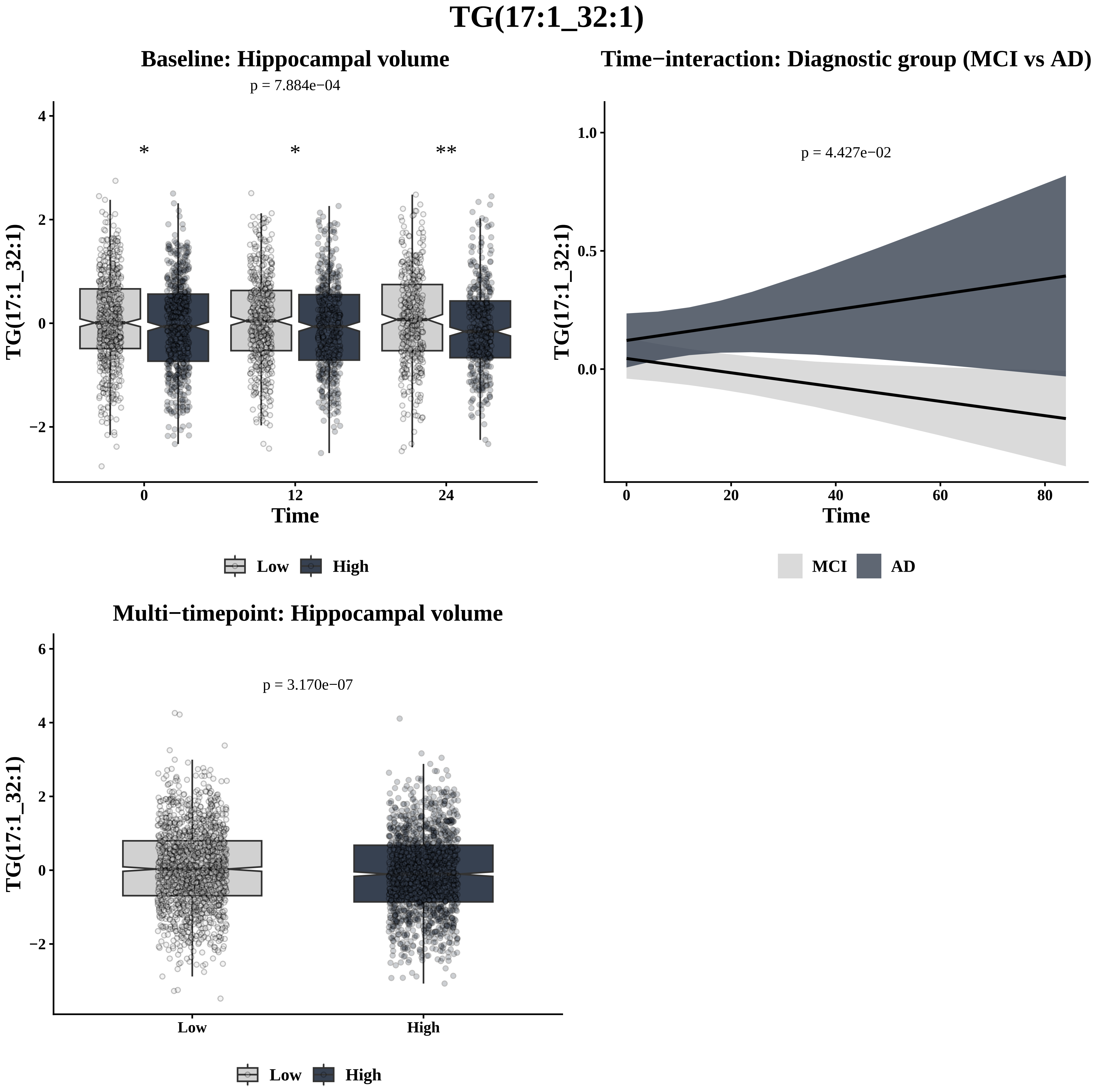

TG(17:1_32:1)
Baseline: Hippocampal volume
p = 7.884e−04
*
*
**
TG(17:1_32:1)
Time
Low
High
Time−interaction: Diagnostic group (MCI vs AD)
p = 4.427e−02
TG(17:1_32:1)
Time
MCI
AD
Multi−timepoint: Hippocampal volume
p = 3.170e−07
TG(17:1_32:1)
Low
High

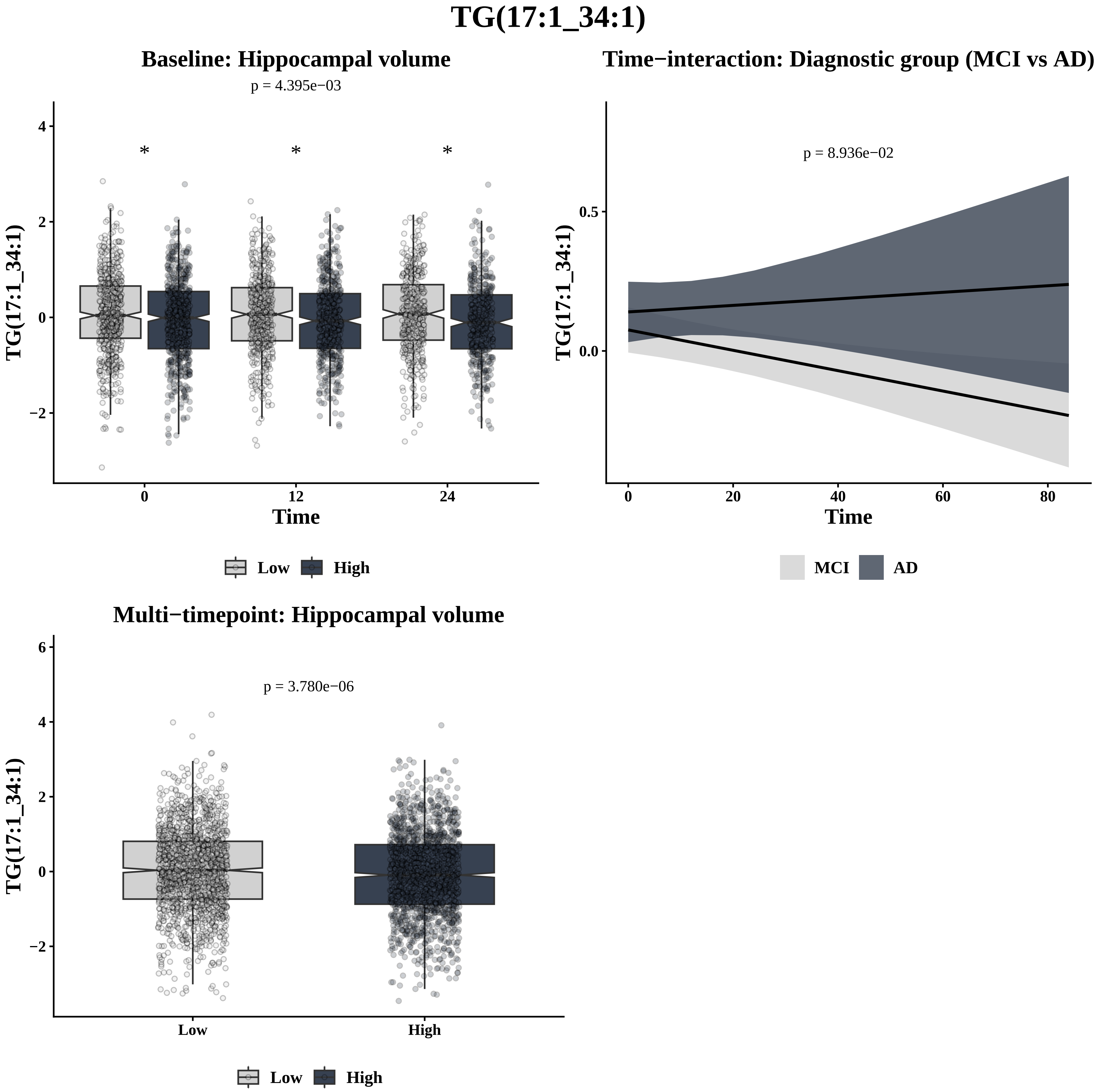
TG(17:1_34:1)
Baseline: Hippocampal volume
p = 4.395e−03
*
*
*
TG(17:1_34:1)
4
2
0
−2
0
12
24
Time
Low
High
Time−interaction: Diagnostic group (MCI vs AD)
p = 8.936e−02
TG(17:1_34:1)
0.5
0.0
0
20
40
60
80
Time
MCI
AD
Multi−timepoint: Hippocampal volume
p = 3.780e−06
TG(17:1_34:1)
6
4
2
0
−2
Low
High
Low
High

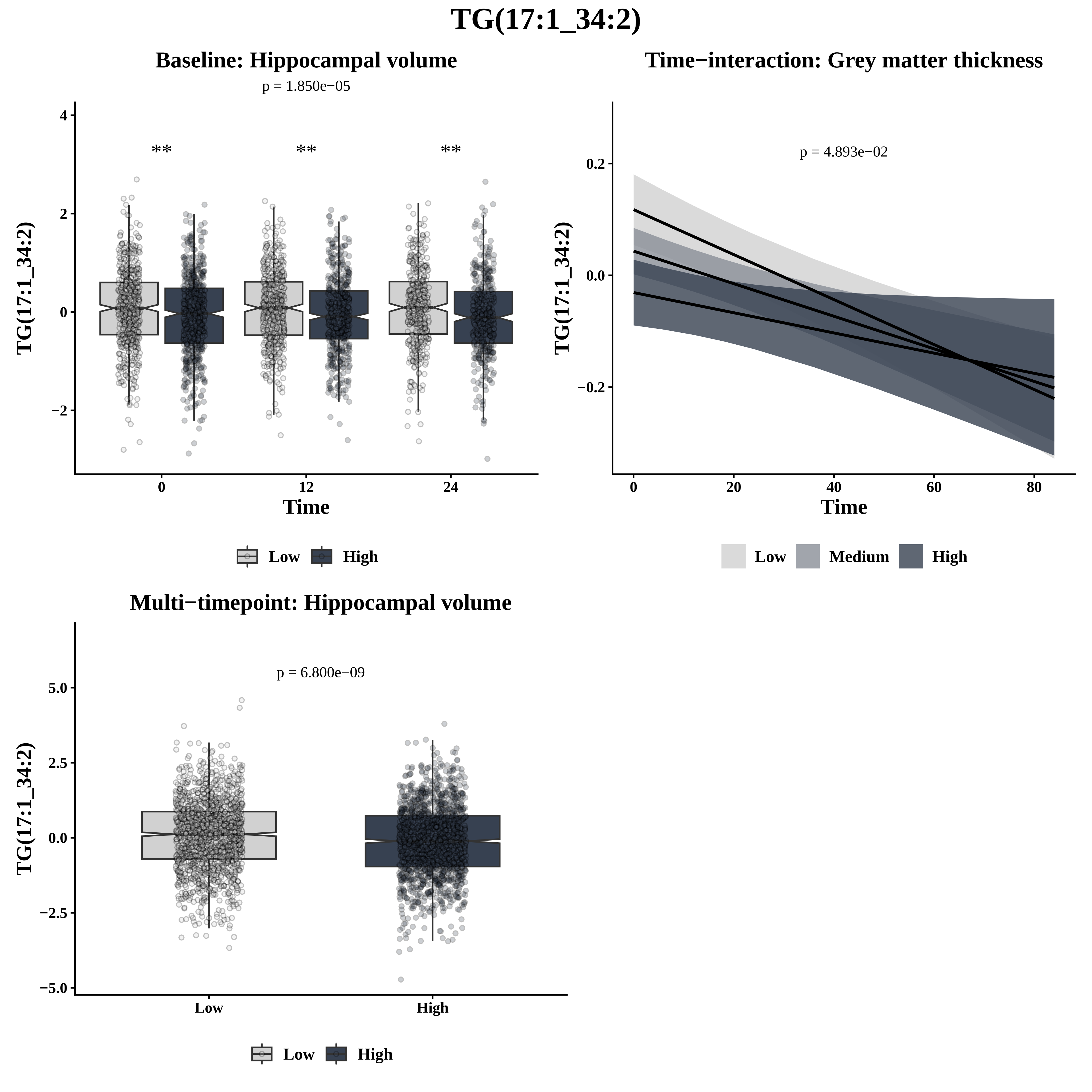

TG(17:1_34:2)
Baseline: Hippocampal volume
p = 1.850e−05
**
**
**
TG(17:1_34:2)
Time
0
12
24
4
2
0
−2
Low
High
Time−interaction: Grey matter thickness
p = 4.893e−02
TG(17:1_34:2)
0.2
0.0
−0.2
0
20
40
60
80
Time
Low
Medium
High
Multi−timepoint: Hippocampal volume
p = 6.800e−09
TG(17:1_34:2)
5.0
2.5
0.0
−2.5
−5.0
Low
High
Low
High

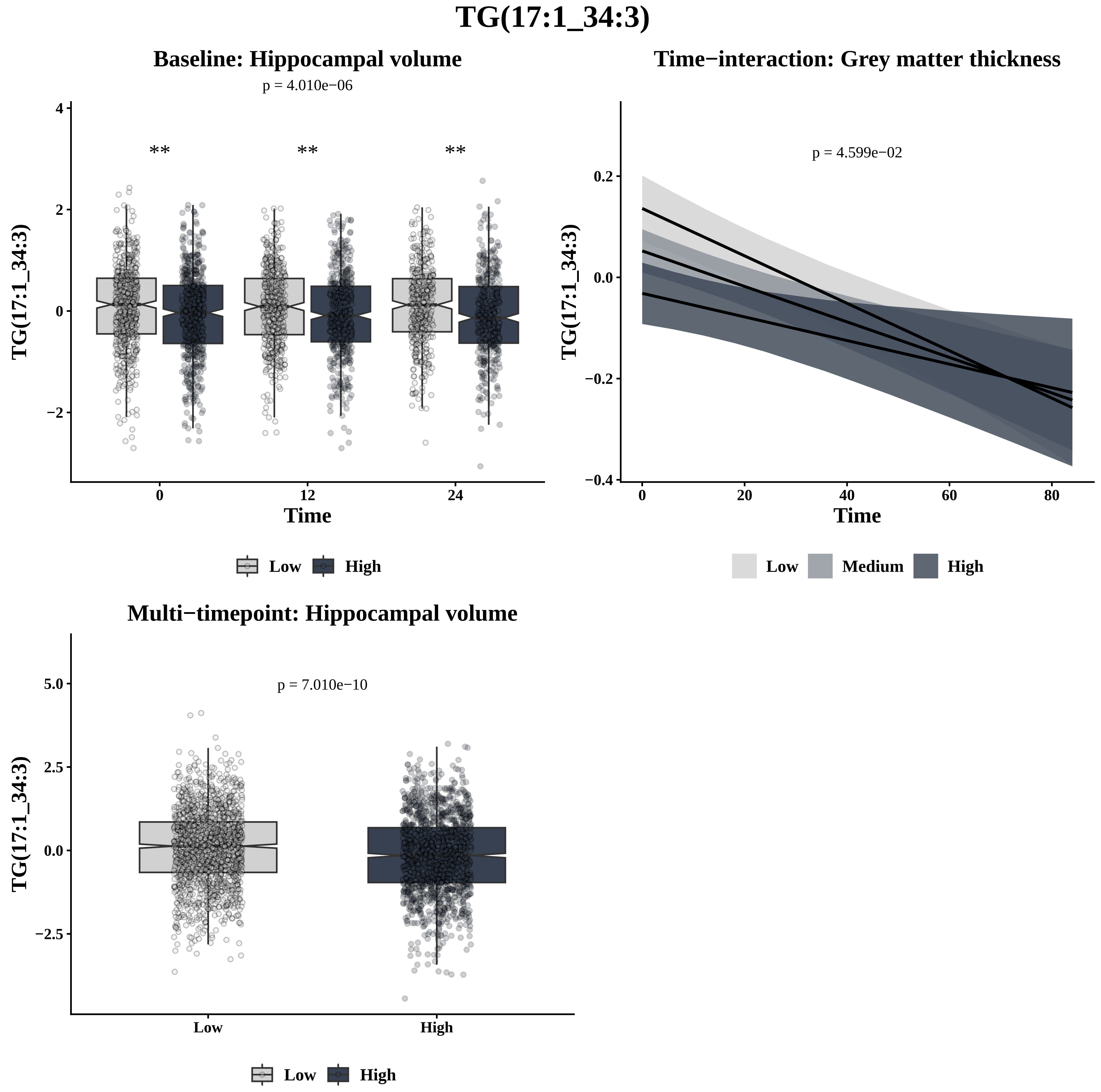
TG(17:1_34:3)
Baseline: Hippocampal volume
p = 4.010e−06
**
**
**
TG(17:1_34:3)
Time
0
12
24
4
2
0
−2
Low
High
Time−interaction: Grey matter thickness
p = 4.599e−02
TG(17:1_34:3)
Time
0.2
0.0
−0.2
−0.4
0
20
40
60
80
Low
Medium
High
Multi−timepoint: Hippocampal volume
p = 7.010e−10
TG(17:1_34:3)
5.0
2.5
0.0
−2.5
Low
High
Low
High

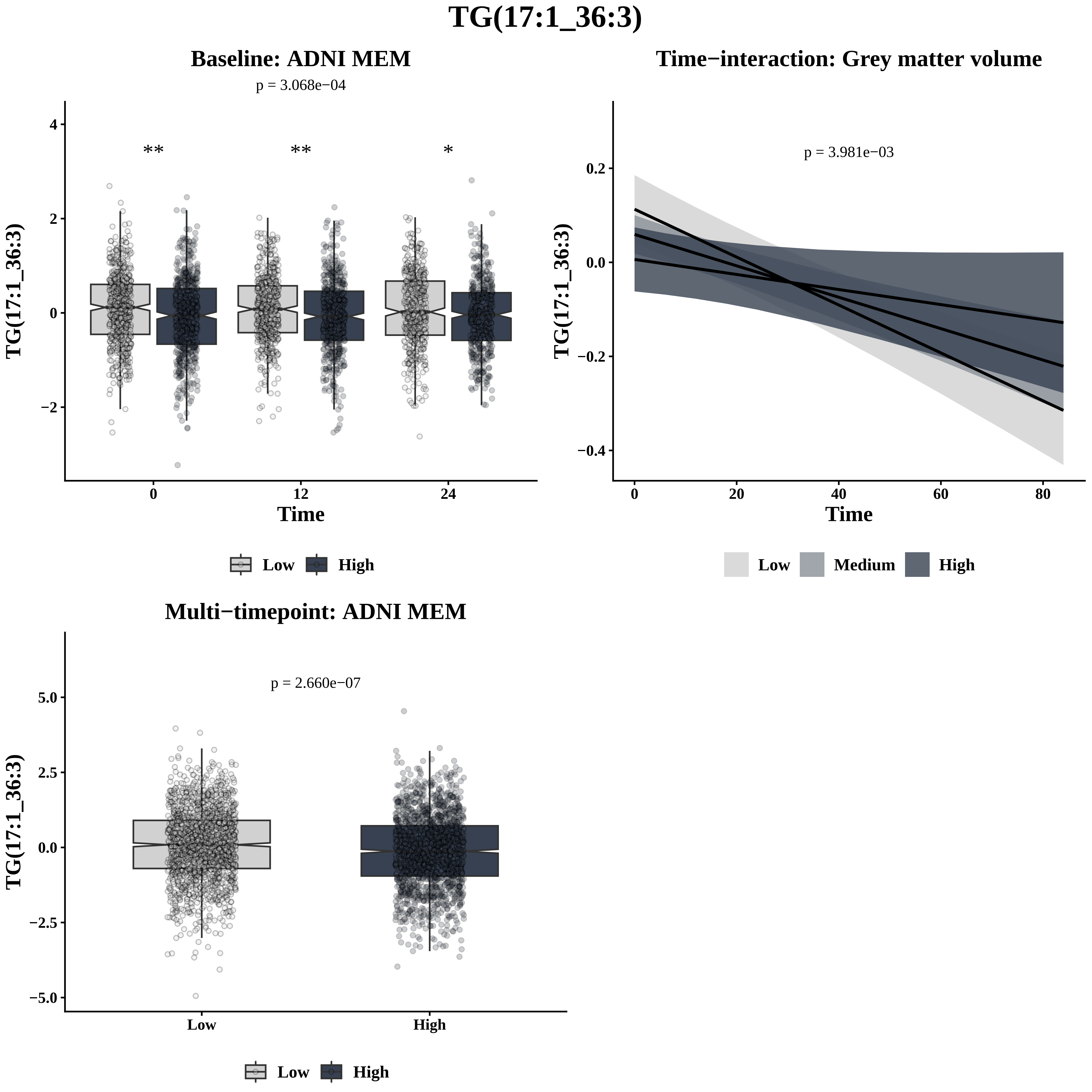
TG(17:1_36:3)
Baseline: ADNI MEM
p = 3.068e−04
**
**
*
TG(17:1_36:3)
Time
0
12
24
4
2
0
−2
Low
High
Time−interaction: Grey matter volume
p = 3.981e−03
TG(17:1_36:3)
Time
0.2
0.0
−0.2
−0.4
0
20
40
60
80
Low
Medium
High
Multi−timepoint: ADNI MEM
p = 2.660e−07
TG(17:1_36:3)
5.0
2.5
0.0
−2.5
−5.0
Low
High
Low
High

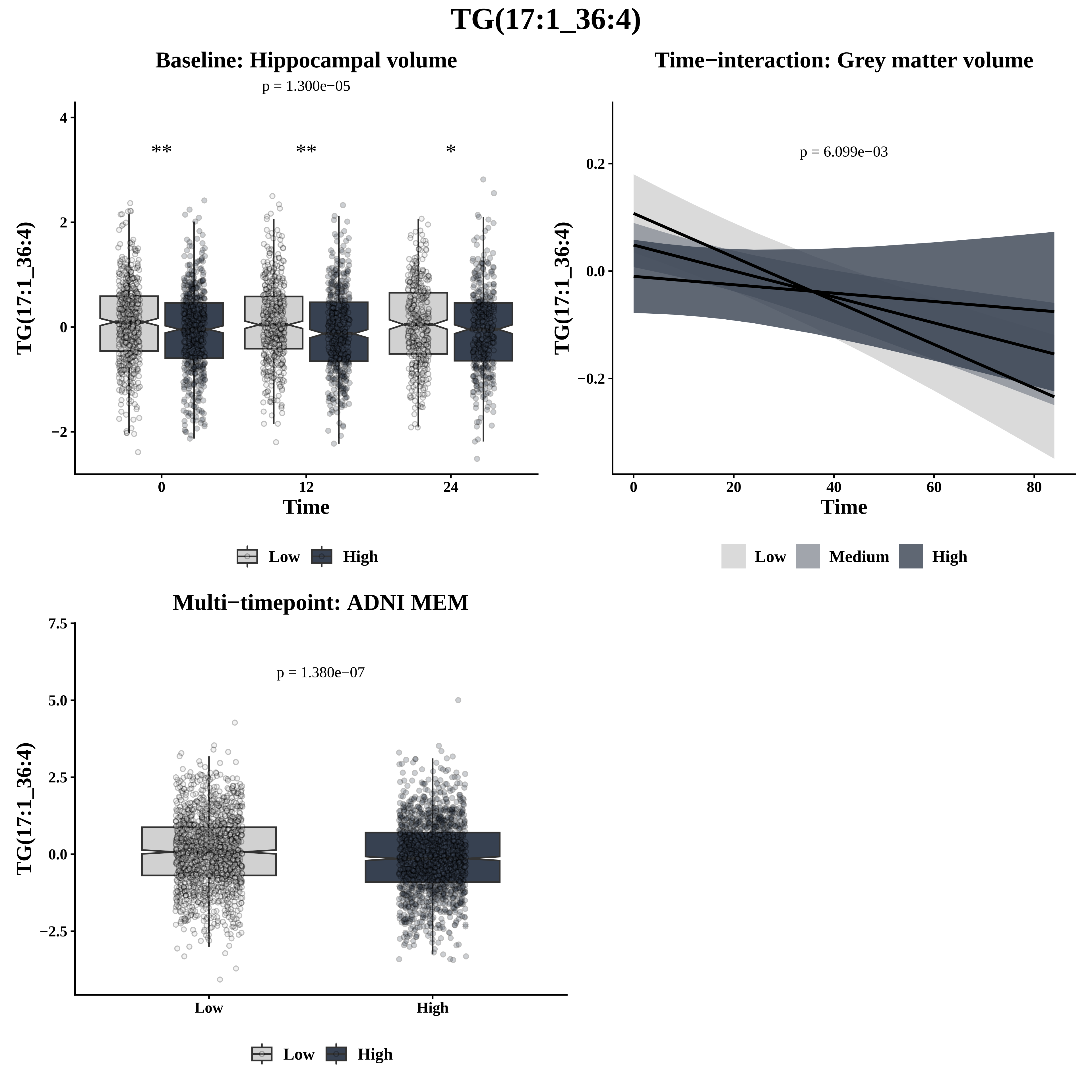

TG(17:1_36:4)
Baseline: Hippocampal volume
p = 1.300e−05
**
**
*
TG(17:1_36:4)
Time
Low
High
Time−interaction: Grey matter volume
p = 6.099e−03
TG(17:1_36:4)
Time
Low
Medium
High
Multi−timepoint: ADNI MEM
p = 1.380e−07
TG(17:1_36:4)
Low
High
Low
High

# TG(17:1_36:5)

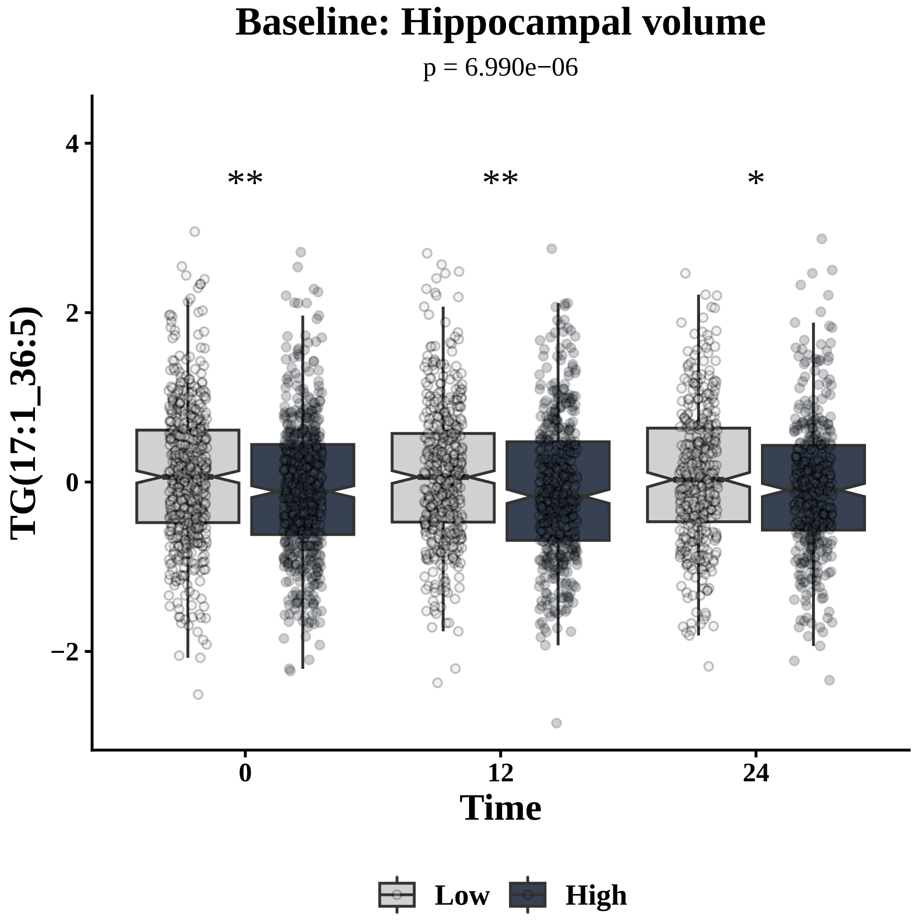

Baseline: Hippocampal volume
p = 6.990e−06
**
**
*
TG(17:1_36:5)
Time
0
12
24
Low
High


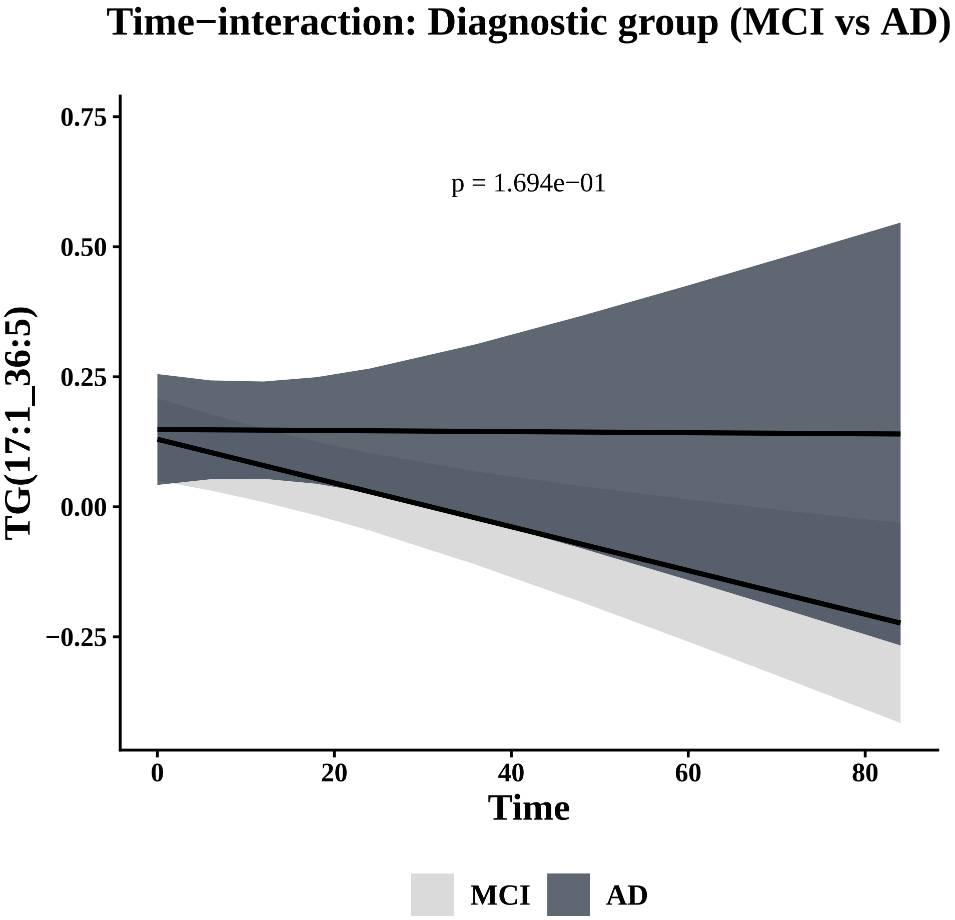

Time−interaction: Diagnostic group (MCI vs AD)
p = 1.694e−01
TG(17:1_36:5)
Time
MCI
AD


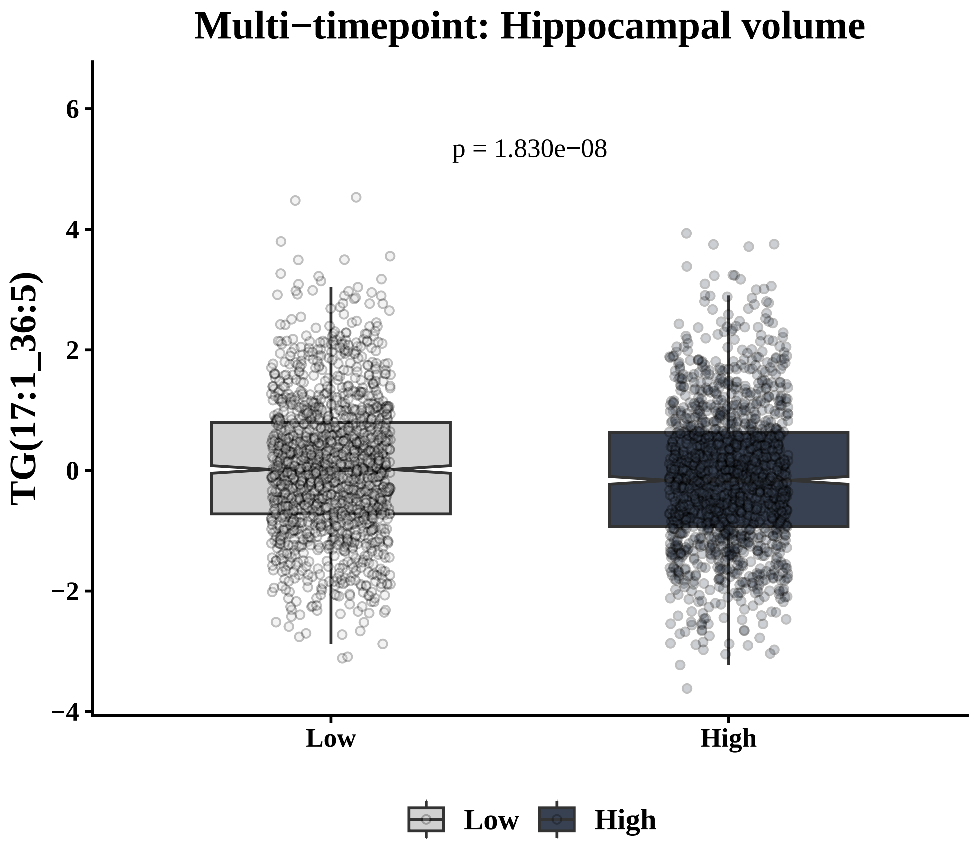

Multi−timepoint: Hippocampal volume
p = 1.830e−08
TG(17:1_36:5)
Low
High

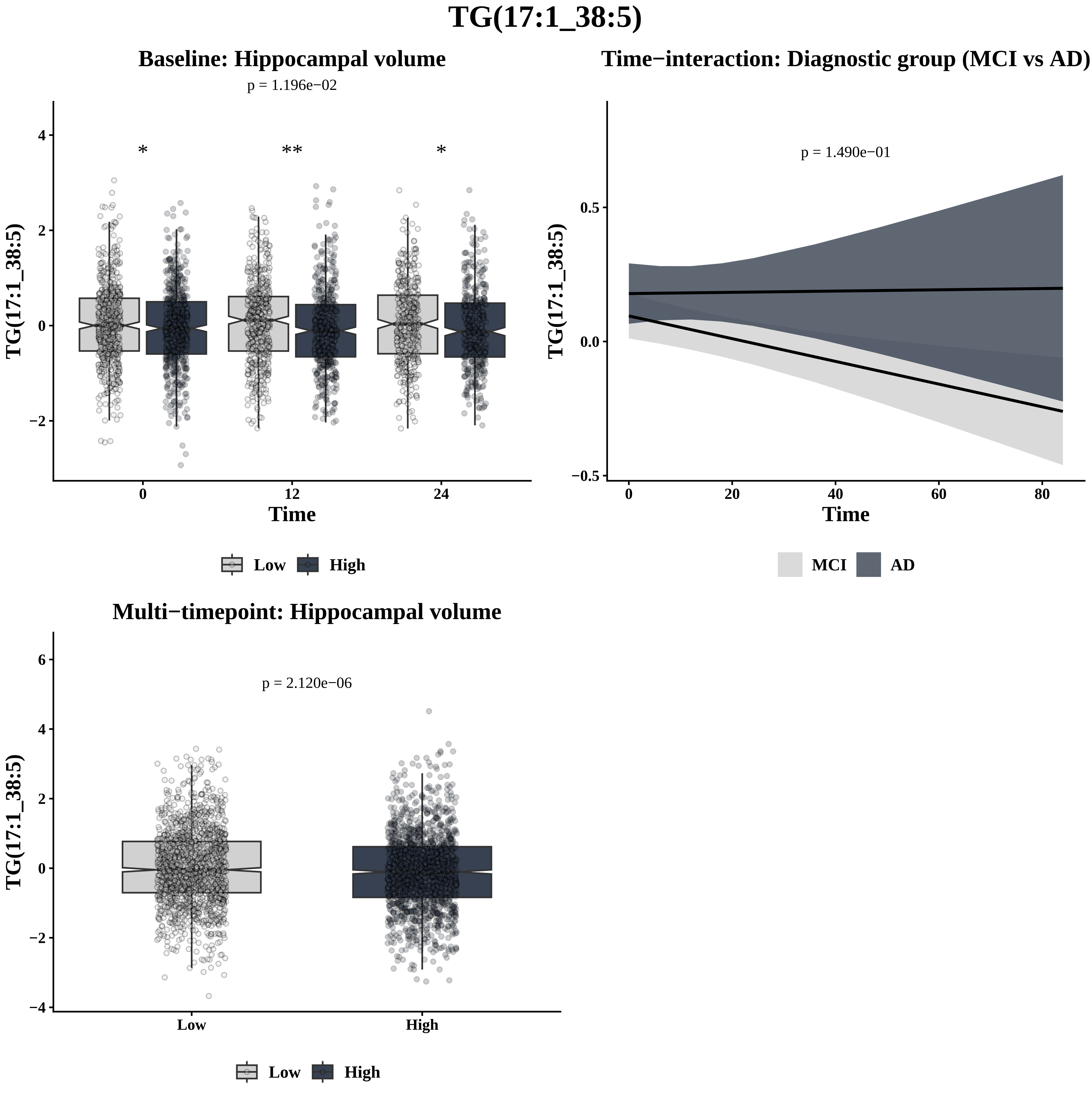
TG(17:1_38:5)
Baseline: Hippocampal volume
p = 1.196e−02
*
**
*
TG(17:1_38:5)
Time
Low
High
Time−interaction: Diagnostic group (MCI vs AD)
p = 1.490e−01
TG(17:1_38:5)
Time
MCI
AD
Multi−timepoint: Hippocampal volume
p = 2.120e−06
TG(17:1_38:5)
Low
High
Low
High

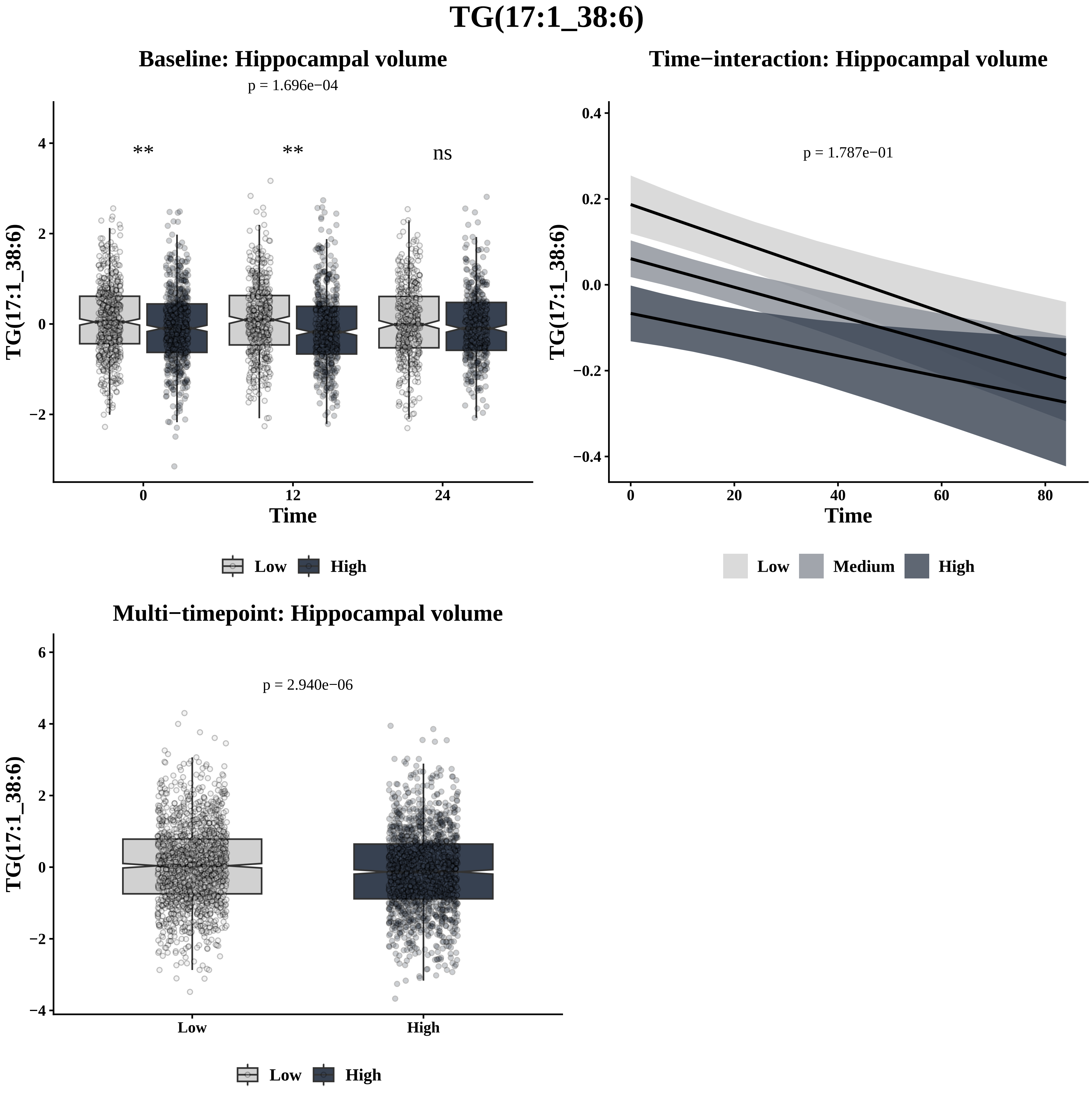

TG(17:1_38:6)
Baseline: Hippocampal volume
p = 1.696e−04
**
**
ns
TG(17:1_38:6)
Time
0
12
24
Low
High
Time−interaction: Hippocampal volume
p = 1.787e−01
TG(17:1_38:6)
Time
0
20
40
60
80
Low
Medium
High
Multi−timepoint: Hippocampal volume
p = 2.940e−06
TG(17:1_38:6)
Low
High
Low
High

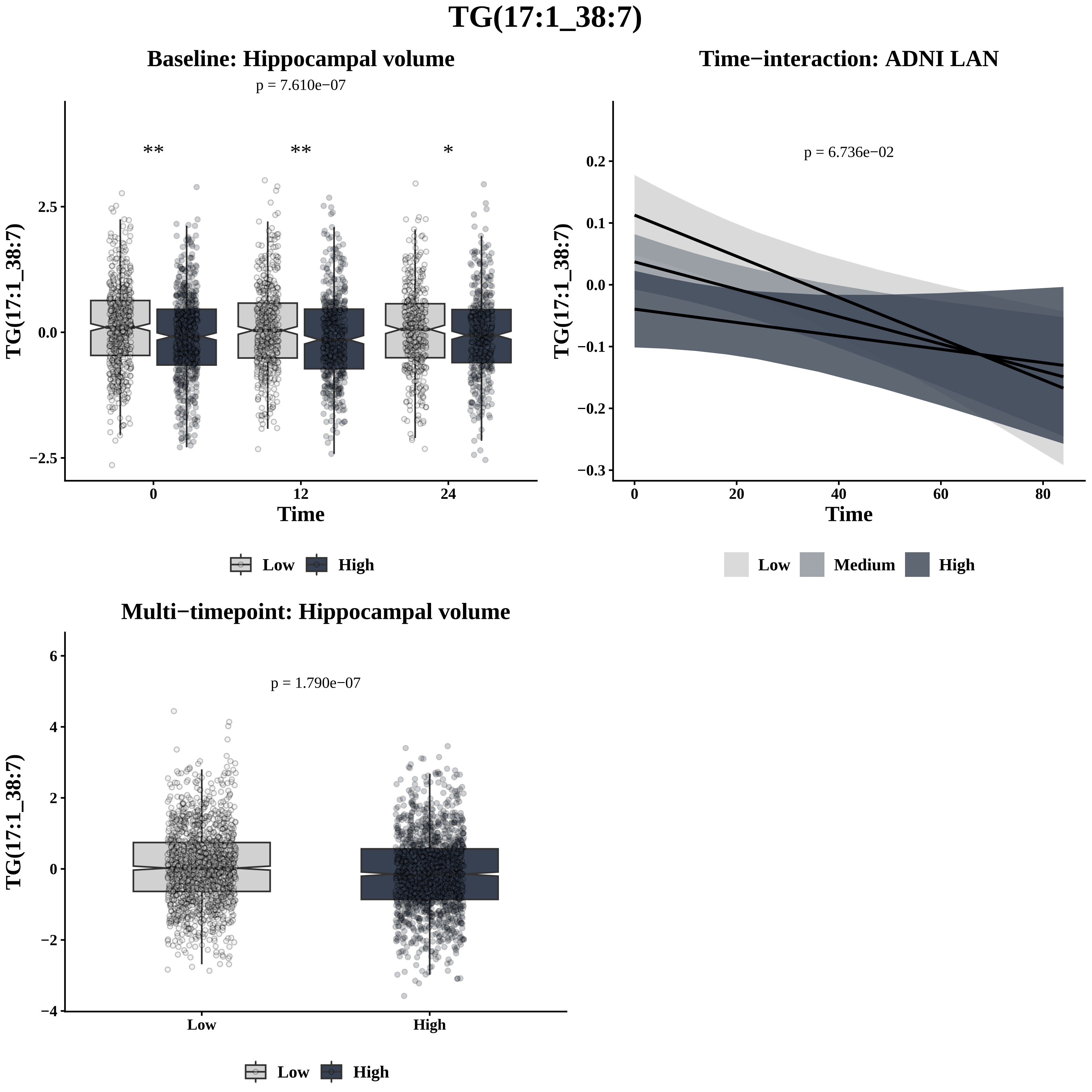

TG(17:1_38:7)
Baseline: Hippocampal volume
p = 7.610e−07
**
**
*
TG(17:1_38:7)
2.5
0.0
−2.5
0
12
24
Time
Low
High
Time−interaction: ADNI LAN
p = 6.736e−02
TG(17:1_38:7)
0.2
0.1
0.0
−0.1
−0.2
−0.3
0
20
40
60
80
Time
Low
Medium
High
Multi−timepoint: Hippocampal volume
p = 1.790e−07
TG(17:1_38:7)
6
4
2
0
−2
−4
Low
High
Low
High

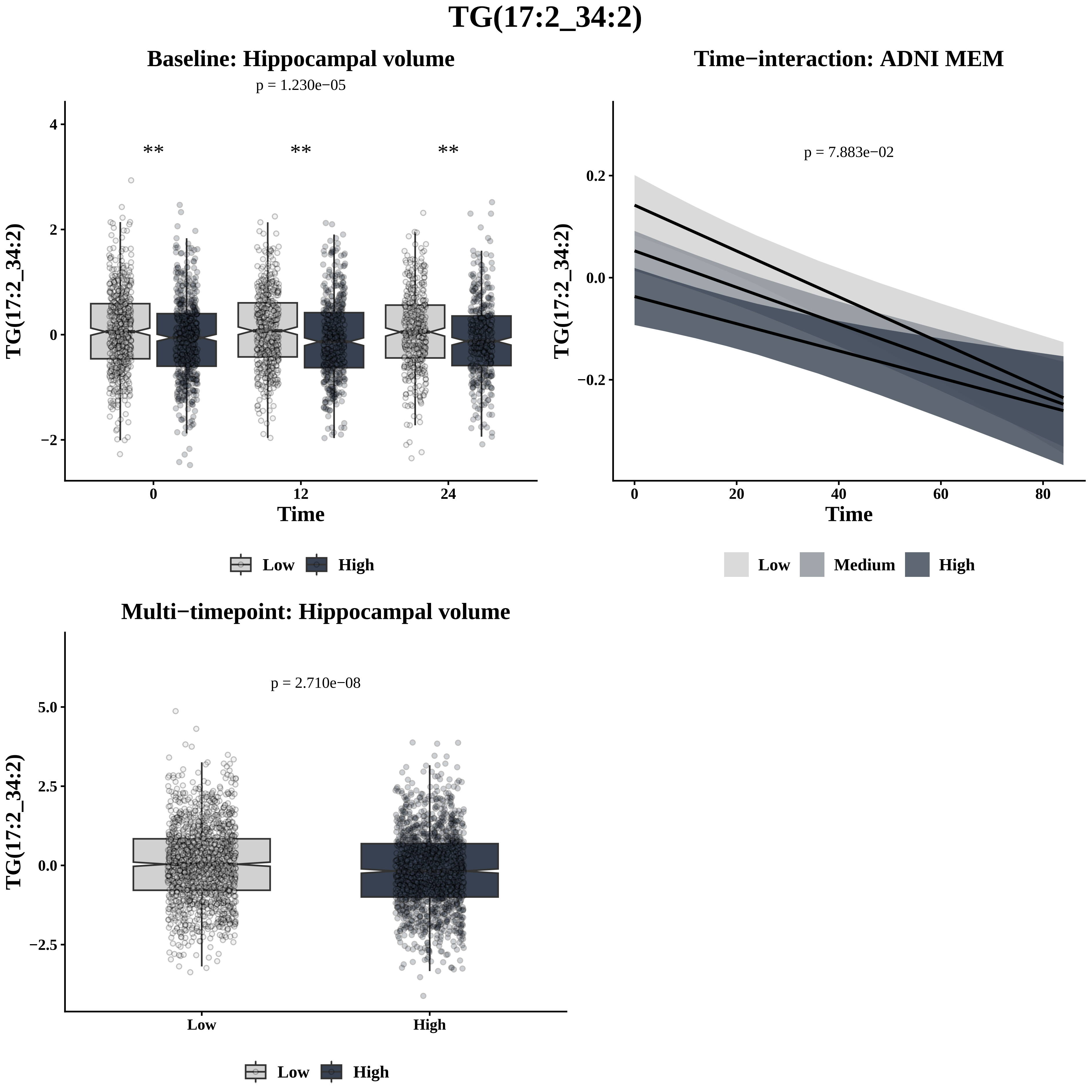
TG(17:2_34:2)
Baseline: Hippocampal volume
p = 1.230e−05
**
**
**
TG(17:2_34:2)
Time
0
12
24
4
2
0
−2
Low
High
Time−interaction: ADNI MEM
p = 7.883e−02
0.2
0.0
−0.2
0
20
40
60
80
Low
Medium
High
Multi−timepoint: Hippocampal volume
p = 2.710e−08
5.0
2.5
0.0
−2.5
Low
High

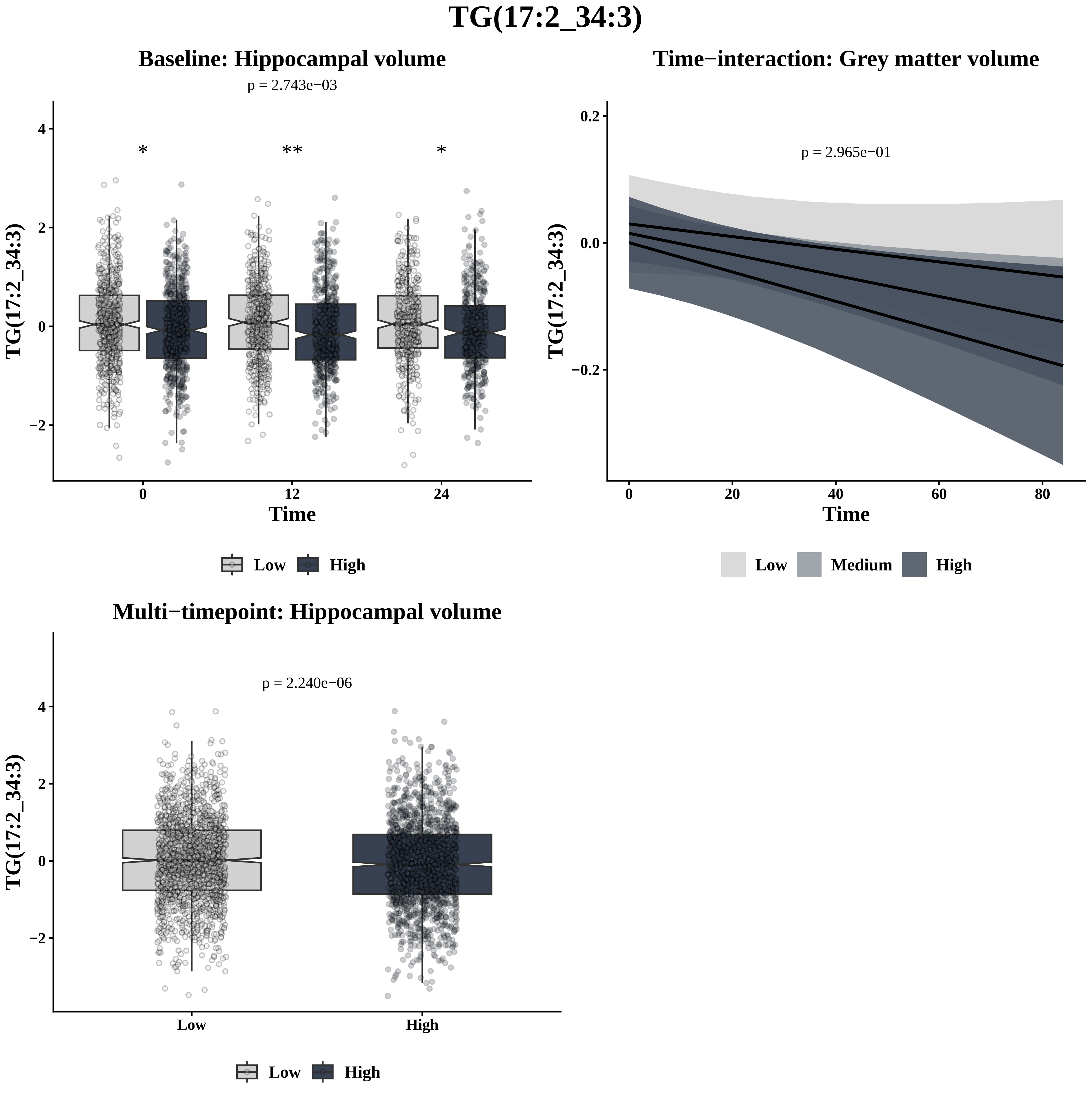

TG(17:2_34:3)
Baseline: Hippocampal volume
p = 2.743e−03
*
**
*
TG(17:2_34:3)
Time
Low
High
Time−interaction: Grey matter volume
p = 2.965e−01
TG(17:2_34:3)
Time
Low
Medium
High
Multi−timepoint: Hippocampal volume
p = 2.240e−06
TG(17:2_34:3)
Low
High

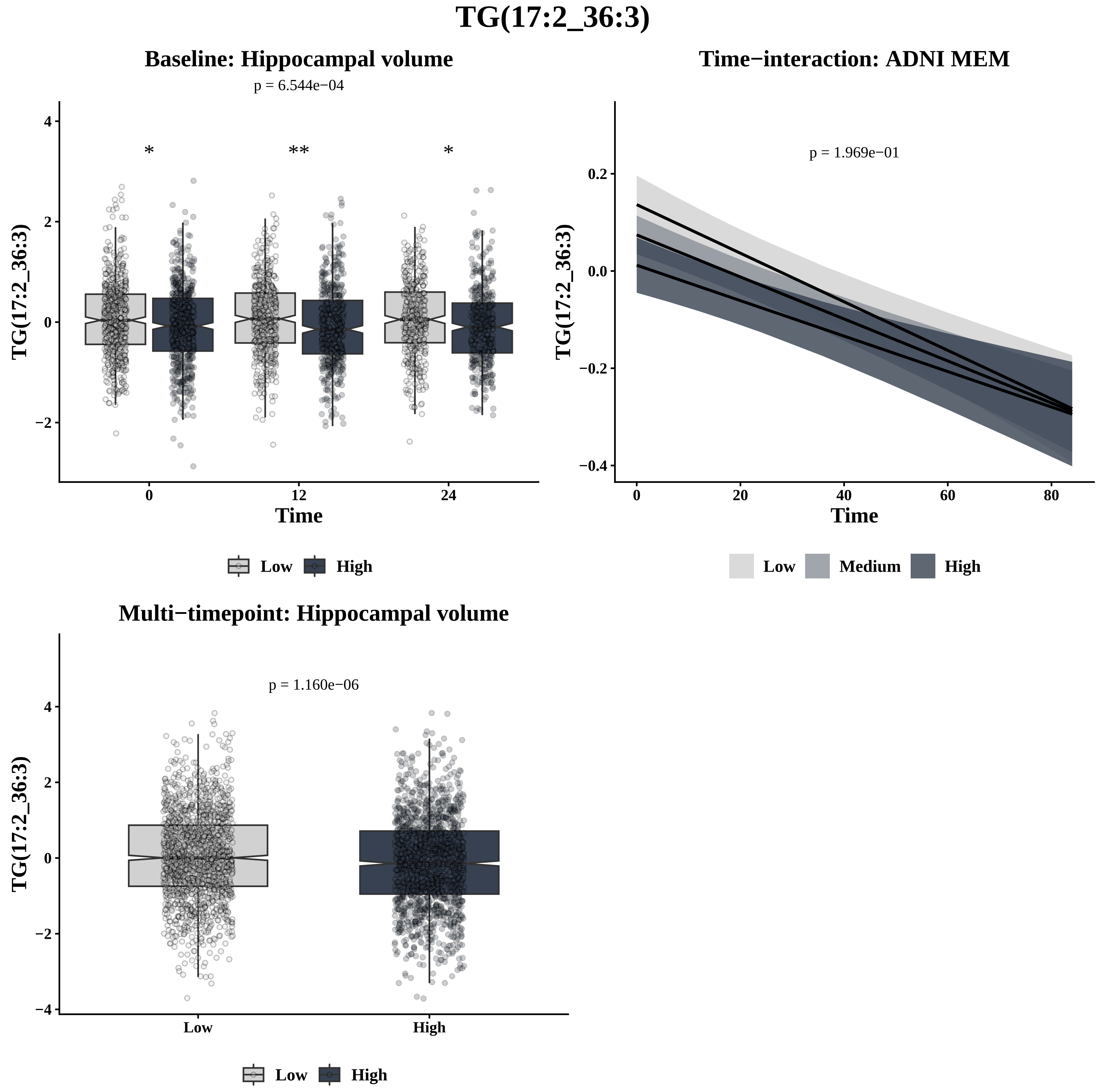
TG(17:2_36:3)
Baseline: Hippocampal volume
p = 6.544e−04
*
**
*
TG(17:2_36:3)
4
2
0
−2
0
12
24
Time
Low
High
Time−interaction: ADNI MEM
p = 1.969e−01
TG(17:2_36:3)
0.2
0.0
−0.2
−0.4
0
20
40
60
80
Time
Low
Medium
High
Multi−timepoint: Hippocampal volume
p = 1.160e−06
TG(17:2_36:3)
4
2
0
−2
−4
Low
High
Low
High

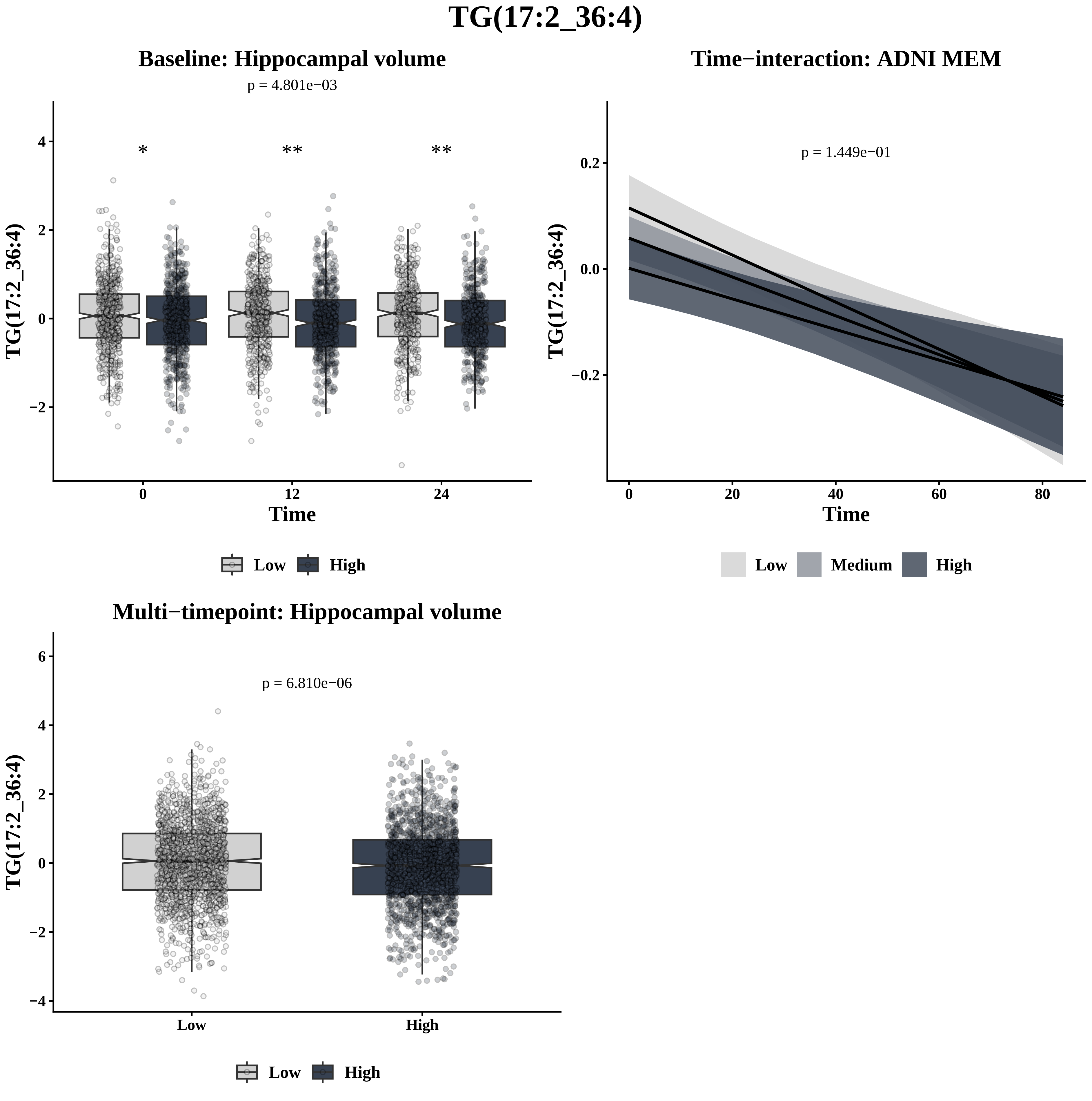

TG(17:2_36:4)
Baseline: Hippocampal volume
p = 4.801e−03
*
**
**
TG(17:2_36:4)
Time
0
12
24
Low
High
Time−interaction: ADNI MEM
p = 1.449e−01
TG(17:2_36:4)
Time
Low
Medium
High
Multi−timepoint: Hippocampal volume
p = 6.810e−06
TG(17:2_36:4)
Low
High
Low
High

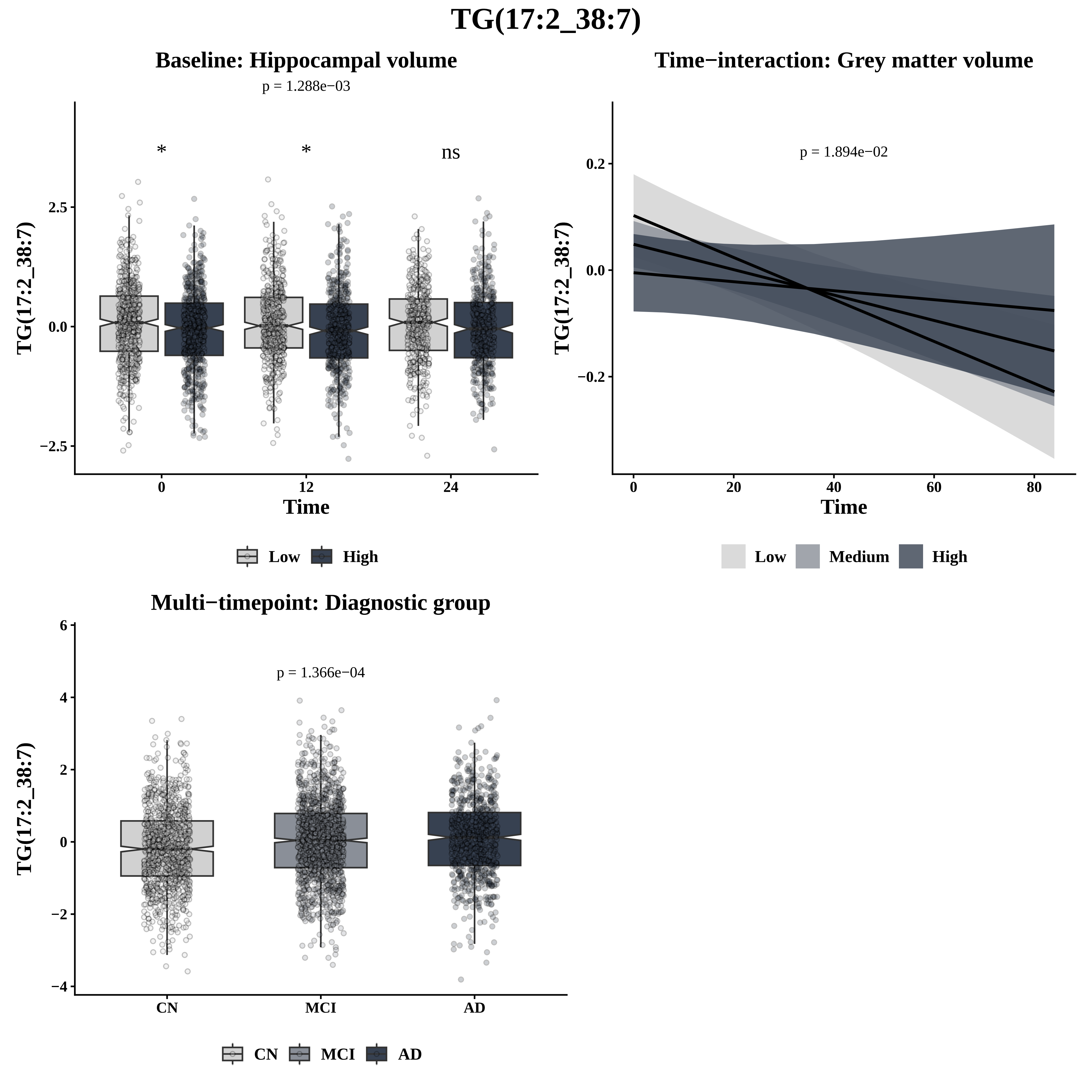
TG(17:2_38:7)
Baseline: Hippocampal volume
p = 1.288e−03
*
*
ns
2.5
0.0
−2.5
TG(17:2_38:7)
0
12
24
Time
Low
High
Time−interaction: Grey matter volume
p = 1.894e−02
0.2
0.0
−0.2
TG(17:2_38:7)
0
20
40
60
80
Time
Low
Medium
High
Multi−timepoint: Diagnostic group
p = 1.366e−04
6
4
2
0
−2
−4
TG(17:2_38:7)
CN
MCI
AD
CN
MCI
AD

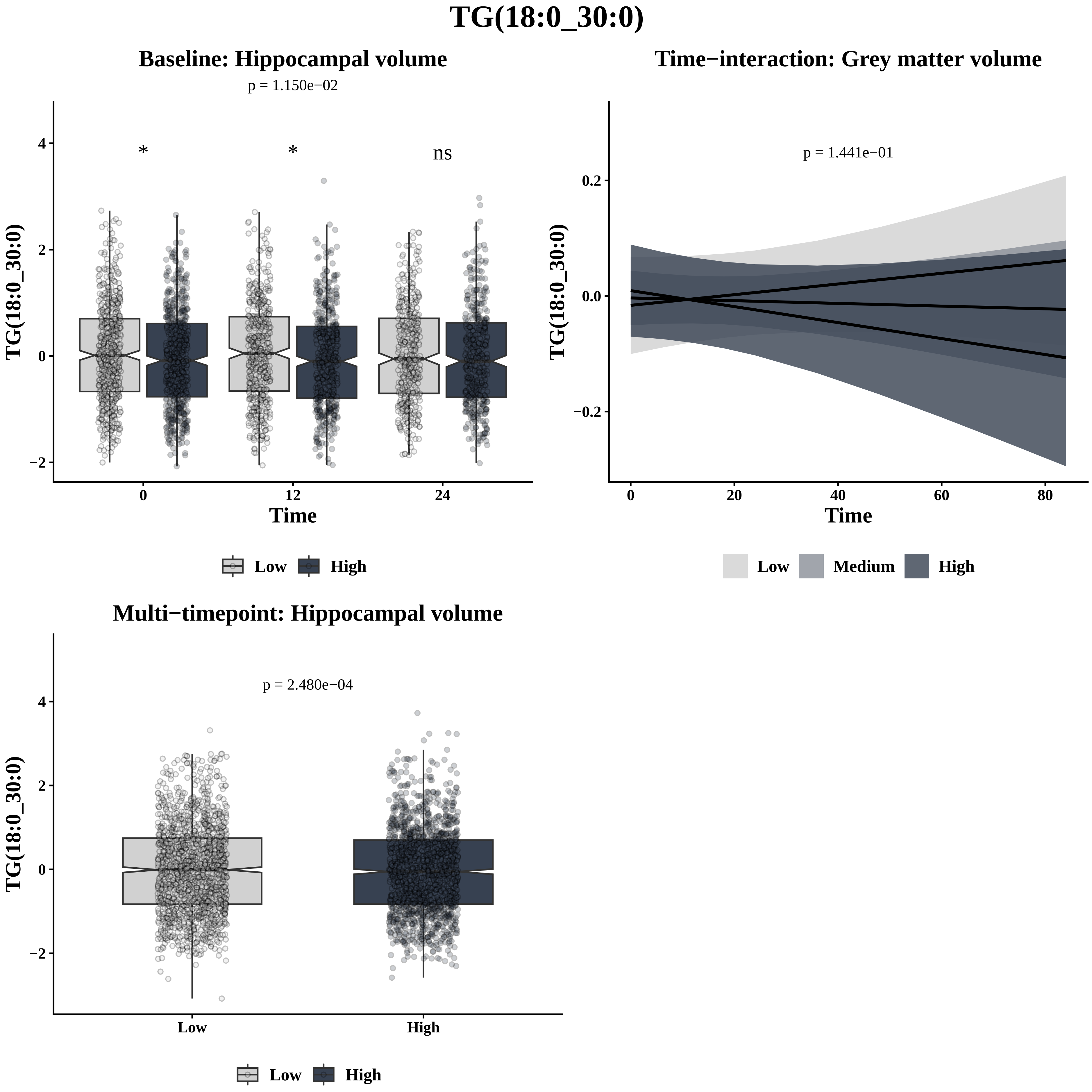
TG(18:0_30:0)
Baseline: Hippocampal volume
p = 1.150e−02
*
*
ns
4
2
0
−2
TG(18:0_30:0)
0
12
24
Time
Low
High
Time−interaction: Grey matter volume
p = 1.441e−01
0.2
0.0
−0.2
TG(18:0_30:0)
0
20
40
60
80
Time
Low
Medium
High
Multi−timepoint: Hippocampal volume
p = 2.480e−04
4
2
0
−2
TG(18:0_30:0)
Low
High
Low
High

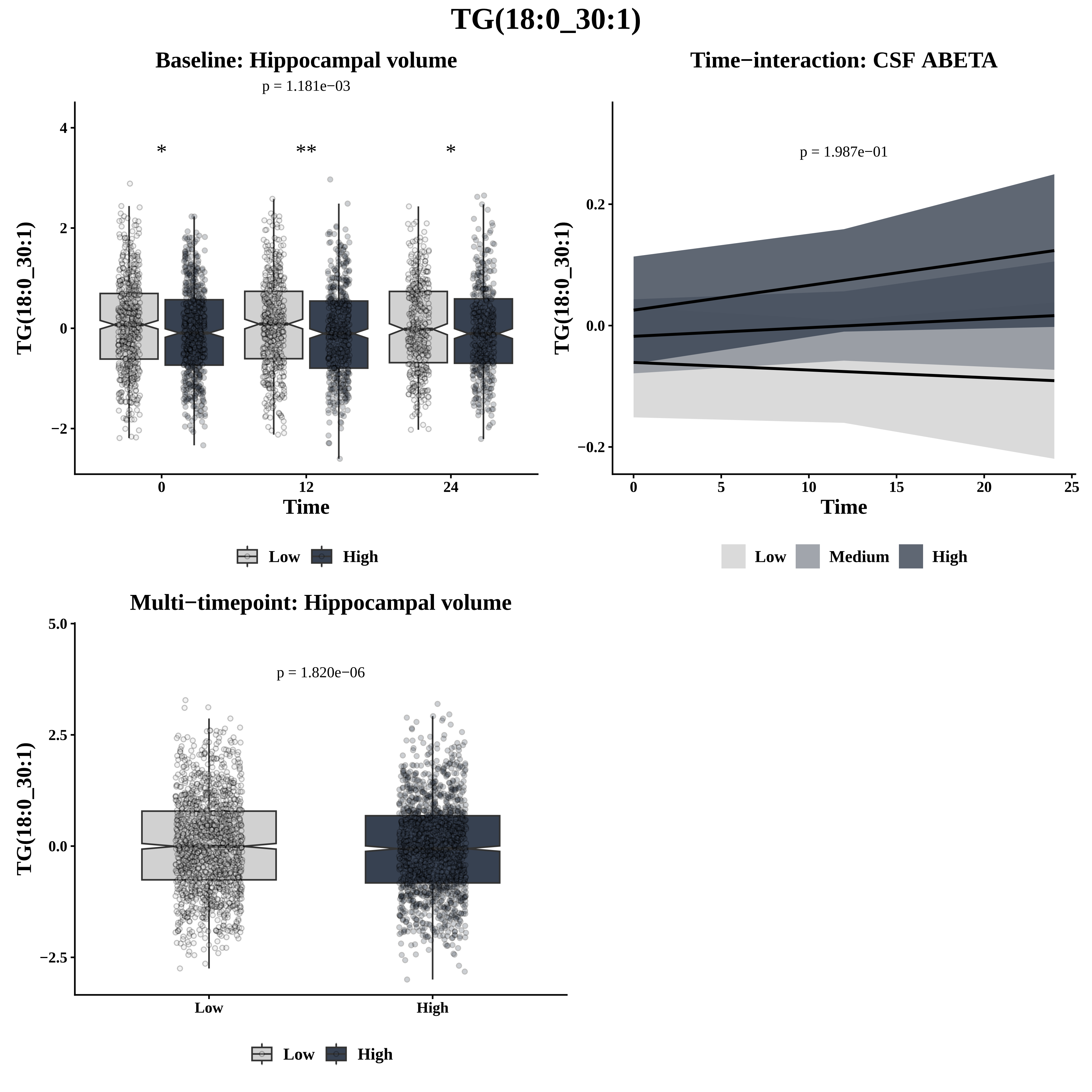

TG(18:0_30:1)
Baseline: Hippocampal volume
p = 1.181e−03
*
**
*
TG(18:0_30:1)
Time
Low
High
Time−interaction: CSF ABETA
p = 1.987e−01
TG(18:0_30:1)
Time
Low
Medium
High
Multi−timepoint: Hippocampal volume
p = 1.820e−06
TG(18:0_30:1)
Low
High

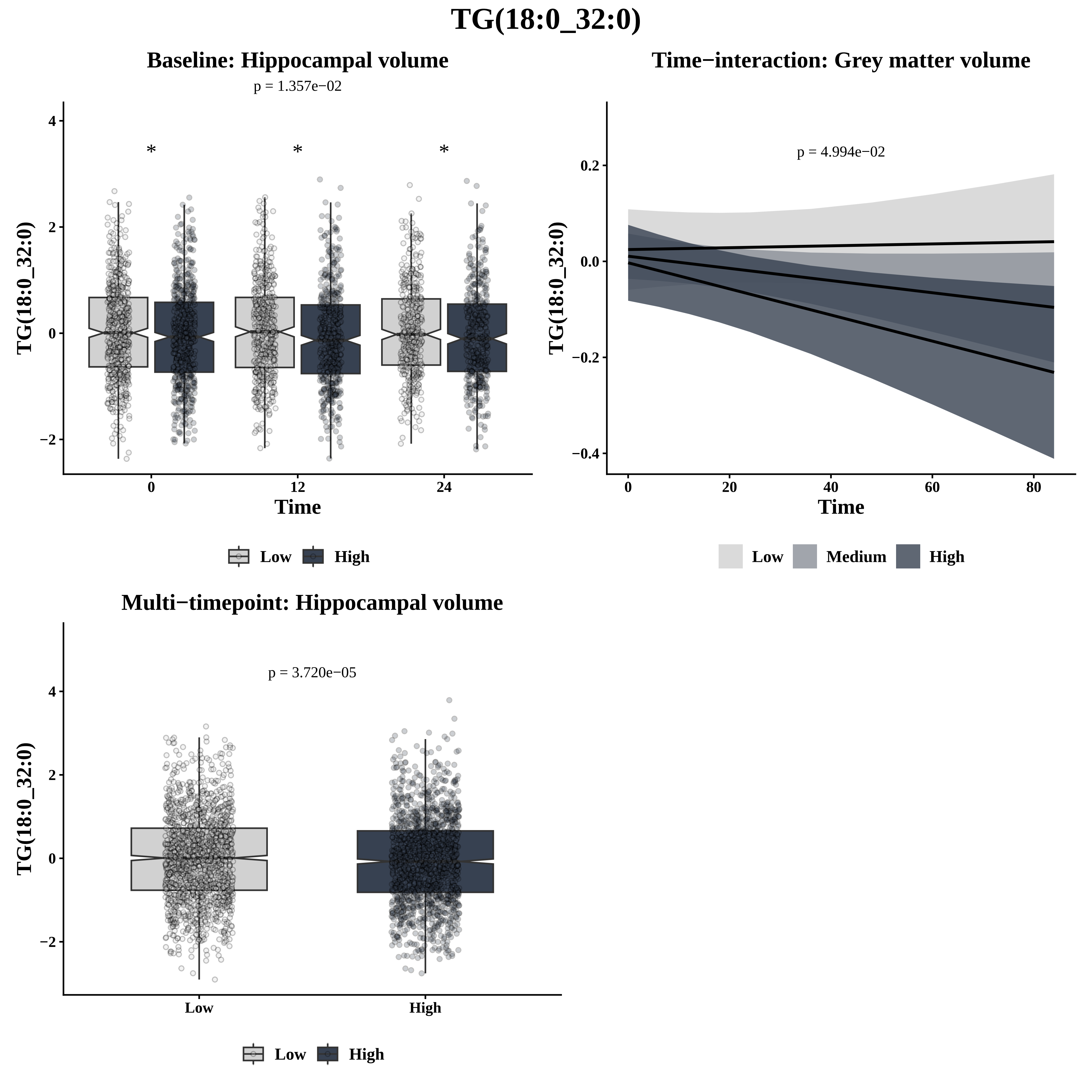
TG(18:0_32:0)
Baseline: Hippocampal volume
p = 1.357e−02
TG(18:0_32:0)
Time
Low
High
Time−interaction: Grey matter volume
p = 4.994e−02
TG(18:0_32:0)
Time
Low
Medium
High
Multi−timepoint: Hippocampal volume
p = 3.720e−05
TG(18:0_32:0)
Low
High

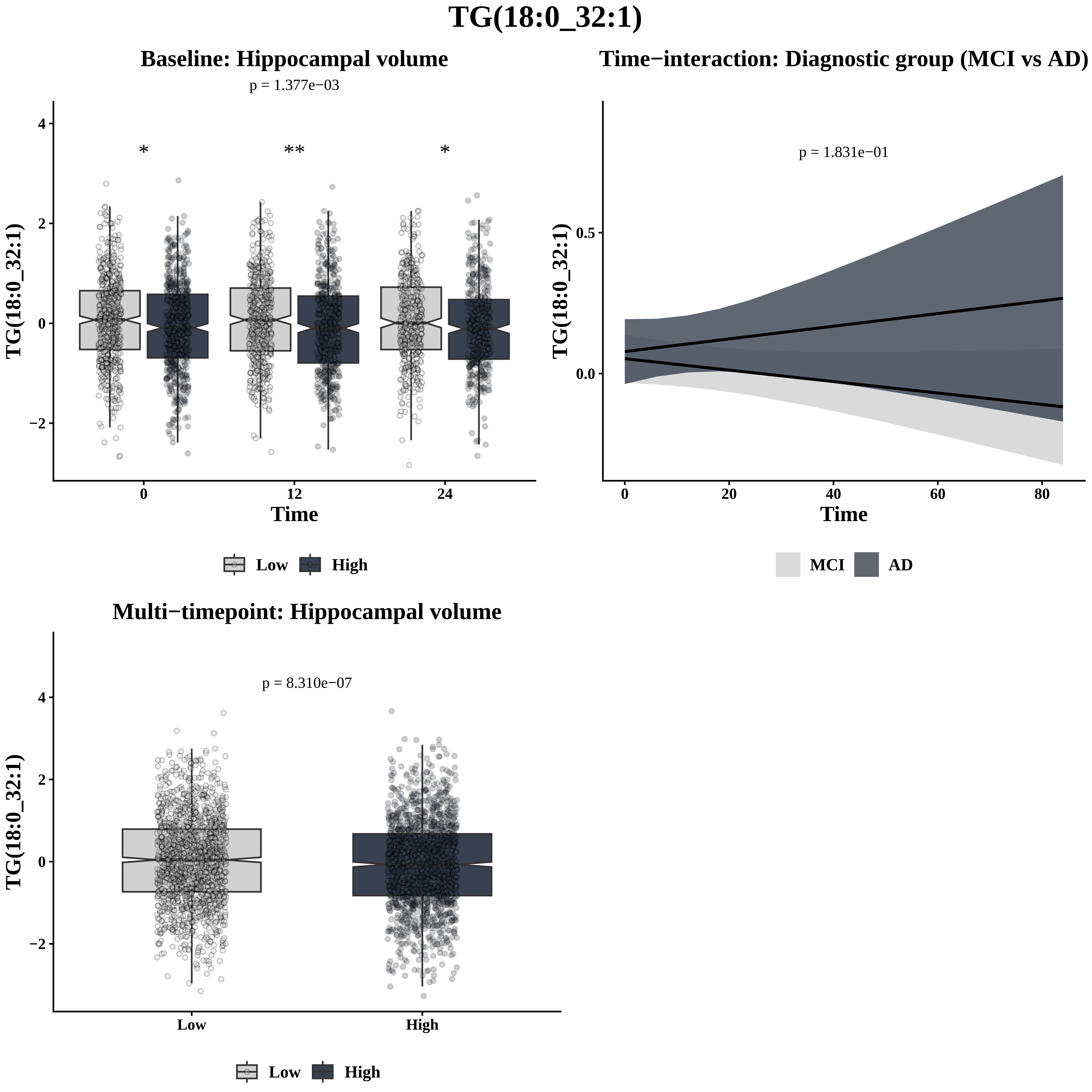
TG(18:0_32:1)
Baseline: Hippocampal volume
p = 1.377e−03
*
**
*
TG(18:0_32:1)
Time
0
12
24
4
2
0
−2
Low
High
Time−interaction: Diagnostic group (MCI vs AD)
p = 1.831e−01
TG(18:0_32:1)
0.5
0.0
0
20
40
60
80
Time
MCI
AD
Multi−timepoint: Hippocampal volume
p = 8.310e−07
TG(18:0_32:1)
4
2
0
−2
Low
High
Low
High

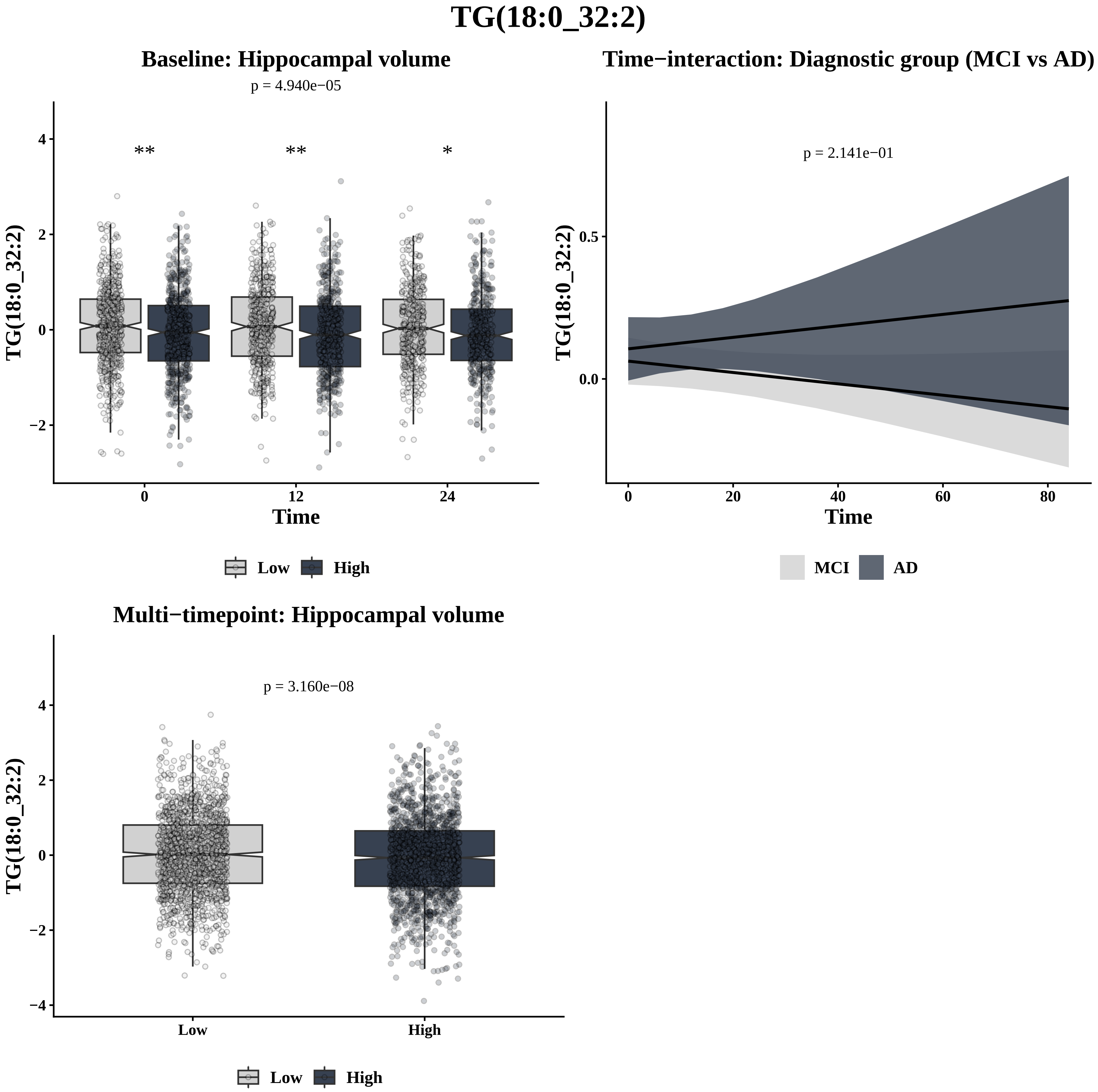

TG(18:0_32:2)
Baseline: Hippocampal volume
p = 4.940e−05
**
**
*
TG(18:0_32:2)
Time
0
12
24
Low
High
Time−interaction: Diagnostic group (MCI vs AD)
p = 2.141e−01
0.5
0.0
0
20
40
60
80
MCI
AD
Multi−timepoint: Hippocampal volume
p = 3.160e−08
4
2
0
−2
−4
Low
High

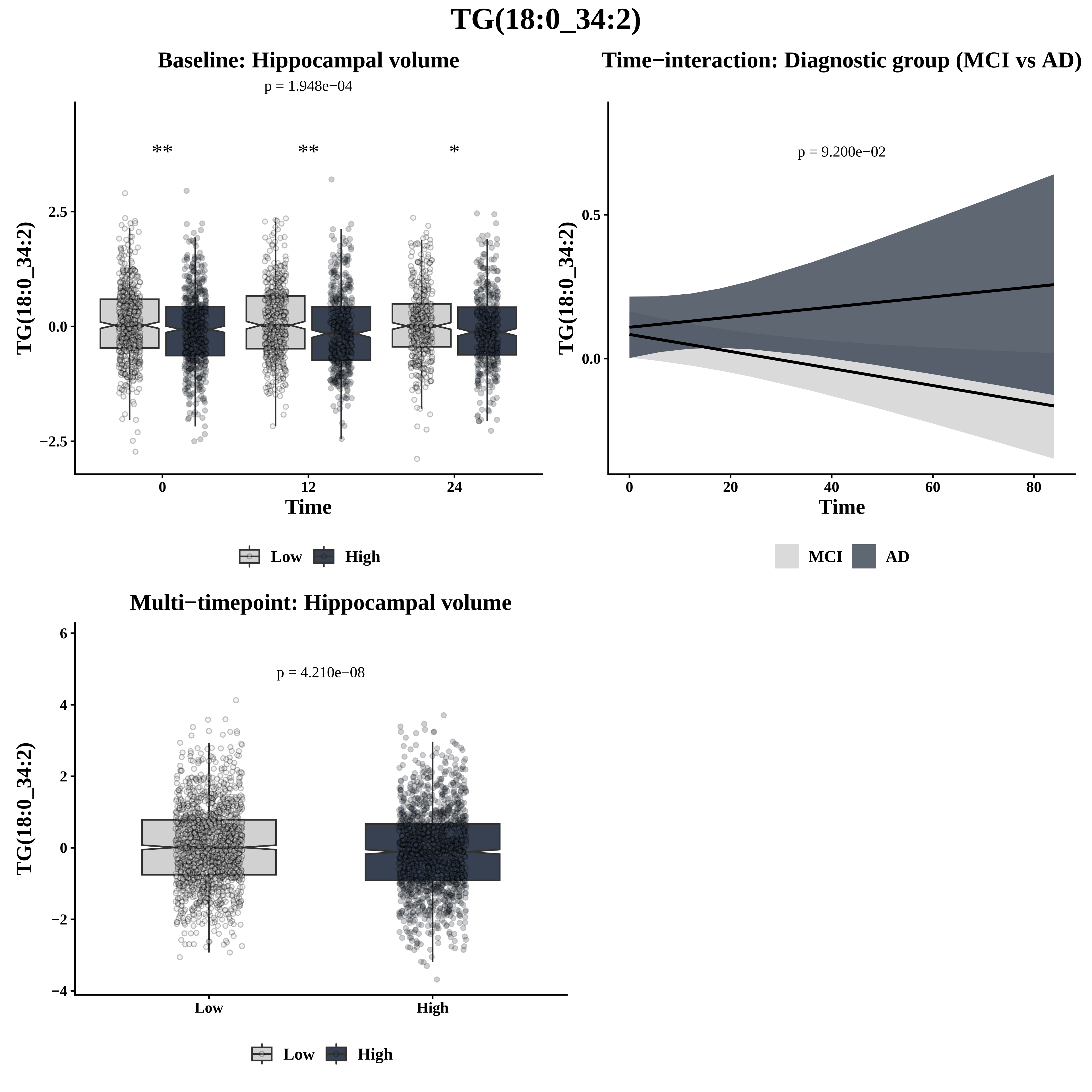

TG(18:0_34:2)
Baseline: Hippocampal volume
p = 1.948e−04
**
**
*
2.5
0.0
−2.5
TG(18:0_34:2)
0
12
24
Time
Low
High
Time−interaction: Diagnostic group (MCI vs AD)
p = 9.200e−02
0.5
0.0
TG(18:0_34:2)
0
20
40
60
80
Time
MCI
AD
Multi−timepoint: Hippocampal volume
p = 4.210e−08
6
4
2
0
−2
−4
TG(18:0_34:2)
Low
High
Low
High

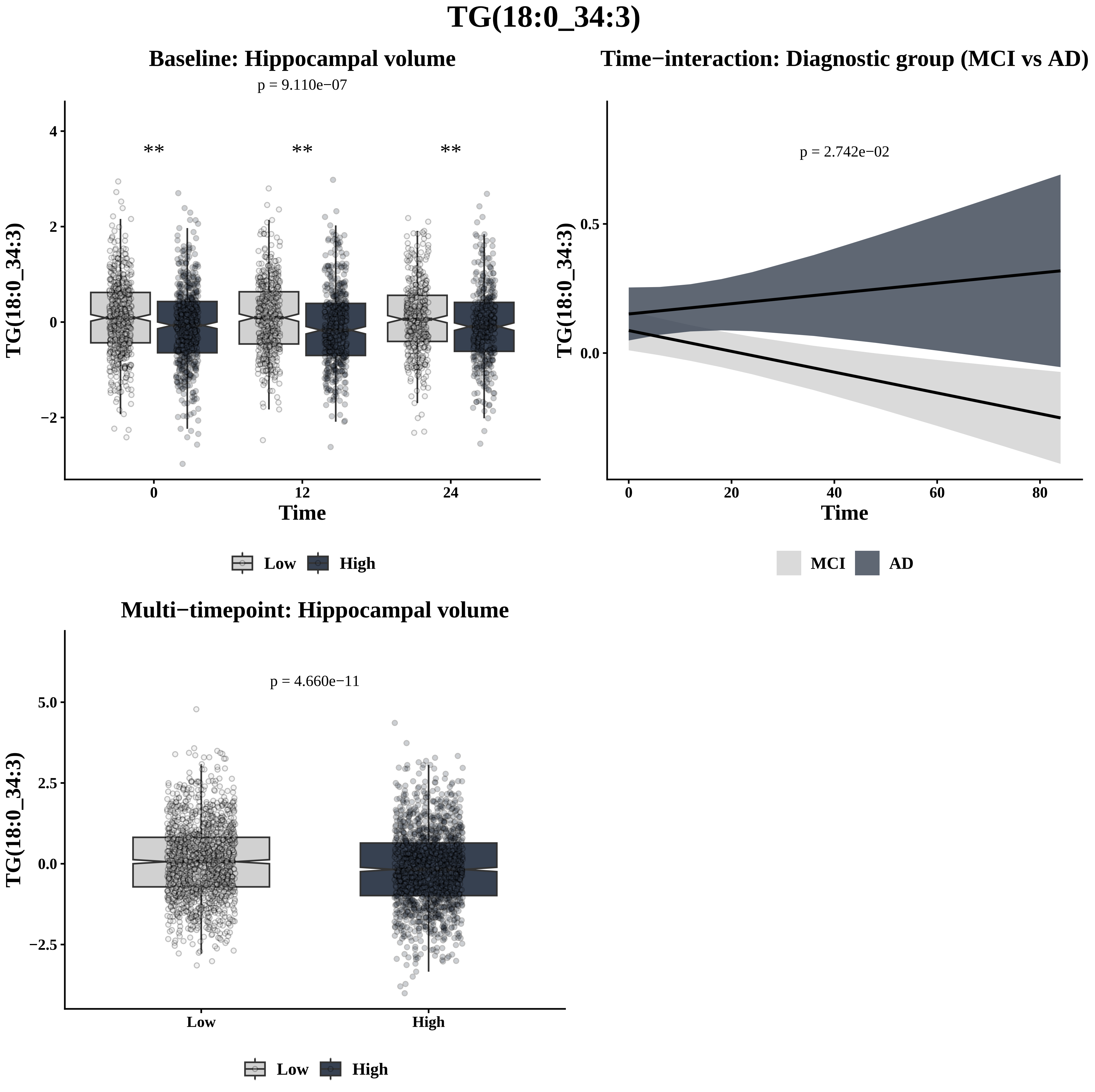
TG(18:0_34:3)
Baseline: Hippocampal volume
p = 9.110e−07
**
**
**
4
2
0
−2
TG(18:0_34:3)
0
12
24
Time
Low
High
Time−interaction: Diagnostic group (MCI vs AD)
p = 2.742e−02
0.5
0.0
TG(18:0_34:3)
0
20
40
60
80
Time
MCI
AD
Multi−timepoint: Hippocampal volume
p = 4.660e−11
5.0
2.5
0.0
−2.5
TG(18:0_34:3)
Low
High
Low
High

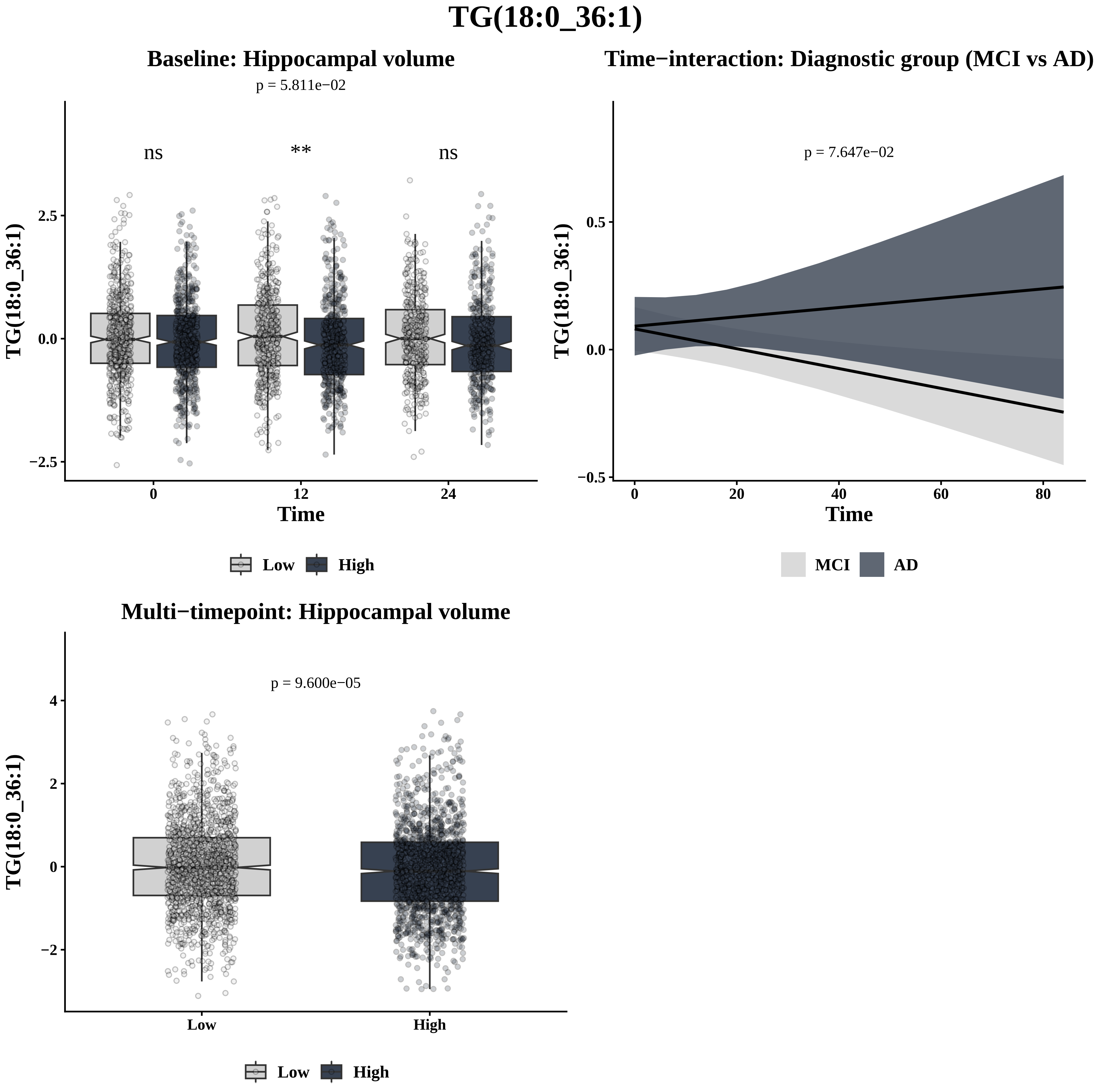
TG(18:0_36:1)
Baseline: Hippocampal volume
p = 5.811e−02
ns
**
ns
TG(18:0_36:1)
2.5
0.0
−2.5
0
12
24
Time
Low
High
Time−interaction: Diagnostic group (MCI vs AD)
p = 7.647e−02
TG(18:0_36:1)
0.5
0.0
−0.5
0
20
40
60
80
Time
MCI
AD
Multi−timepoint: Hippocampal volume
p = 9.600e−05
TG(18:0_36:1)
4
2
0
−2
Low
High
Low
High

# TG(18:0_36:2)

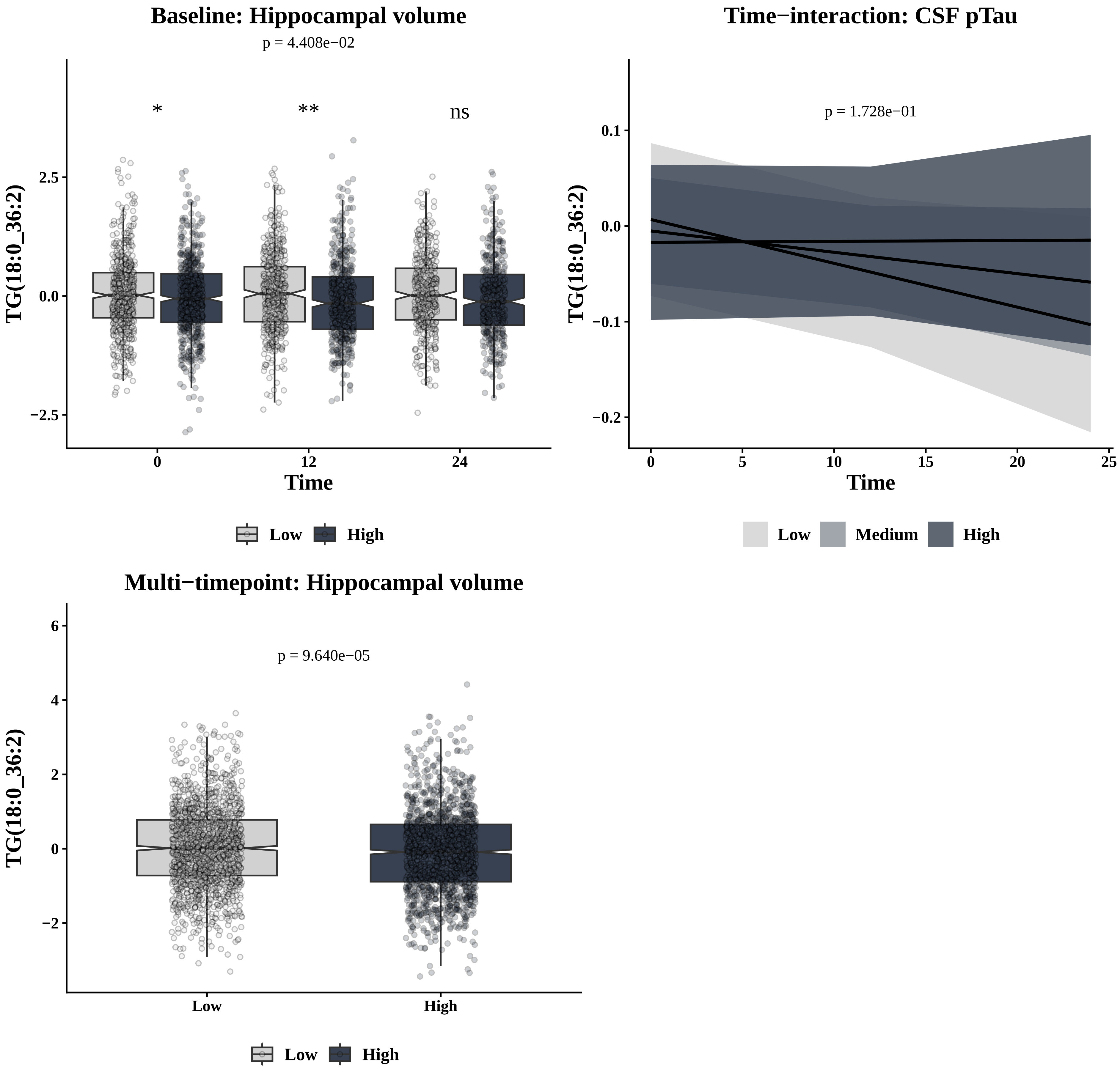

Baseline: Hippocampal volume
p = 4.408e−02
*
**
ns
TG(18:0_36:2)
2.5
0.0
−2.5
0
12
24
Time
Low
High
Time−interaction: CSF pTau
p = 1.728e−01
TG(18:0_36:2)
0.1
0.0
−0.1
−0.2
0
5
10
15
20
25
Time
Low
Medium
High
Multi−timepoint: Hippocampal volume
p = 9.640e−05
TG(18:0_36:2)
6
4
2
0
−2
Low
High
Low
High

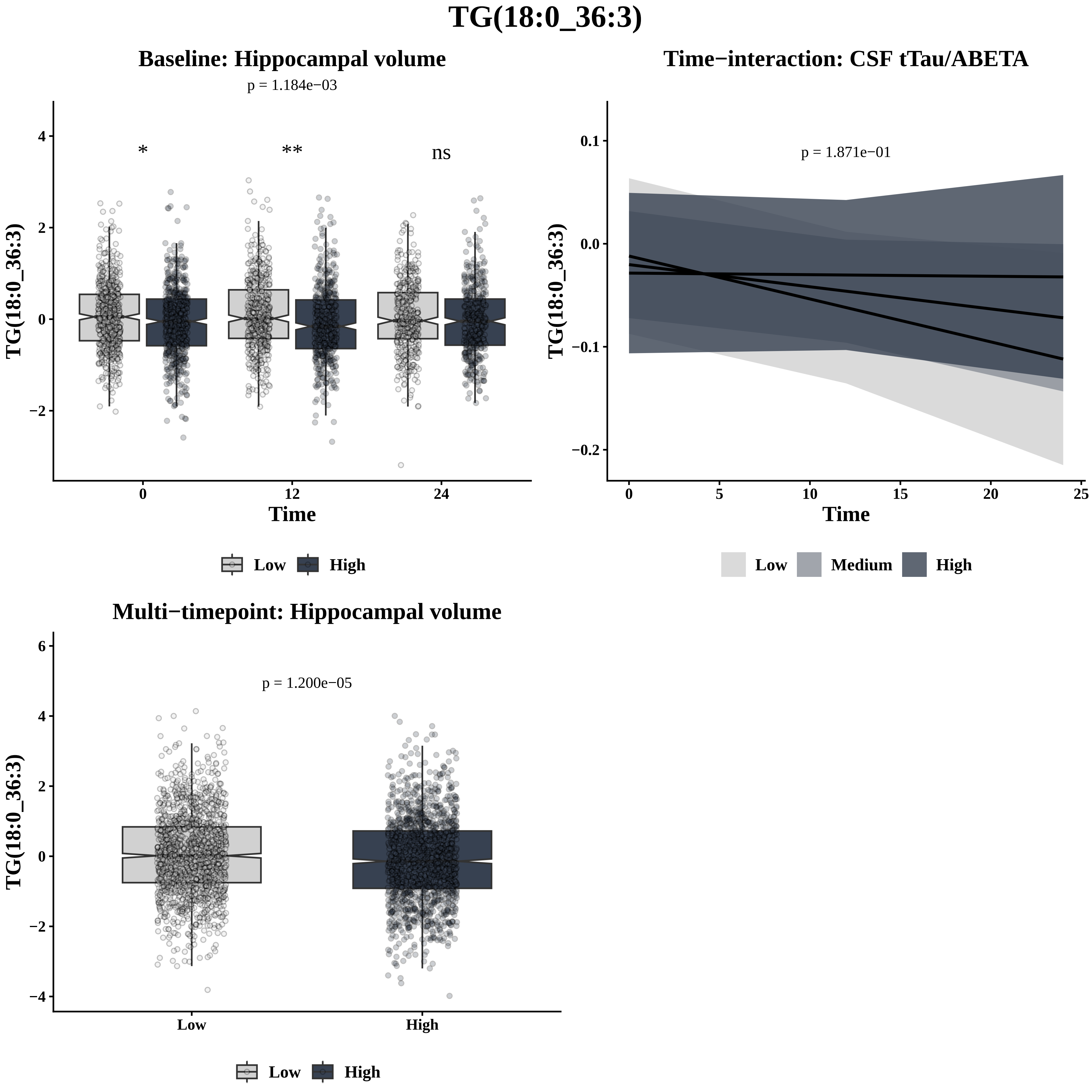
TG(18:0_36:3)
Baseline: Hippocampal volume
p = 1.184e−03
*
**
ns
TG(18:0_36:3)
Time
0
12
24
4
2
0
−2
Low
High
Time−interaction: CSF tTau/ABETA
p = 1.871e−01
TG(18:0_36:3)
0.1
0.0
−0.1
−0.2
0
5
10
15
20
25
Time
Low
Medium
High
Multi−timepoint: Hippocampal volume
p = 1.200e−05
TG(18:0_36:3)
6
4
2
0
−2
−4
Low
High
Low
High

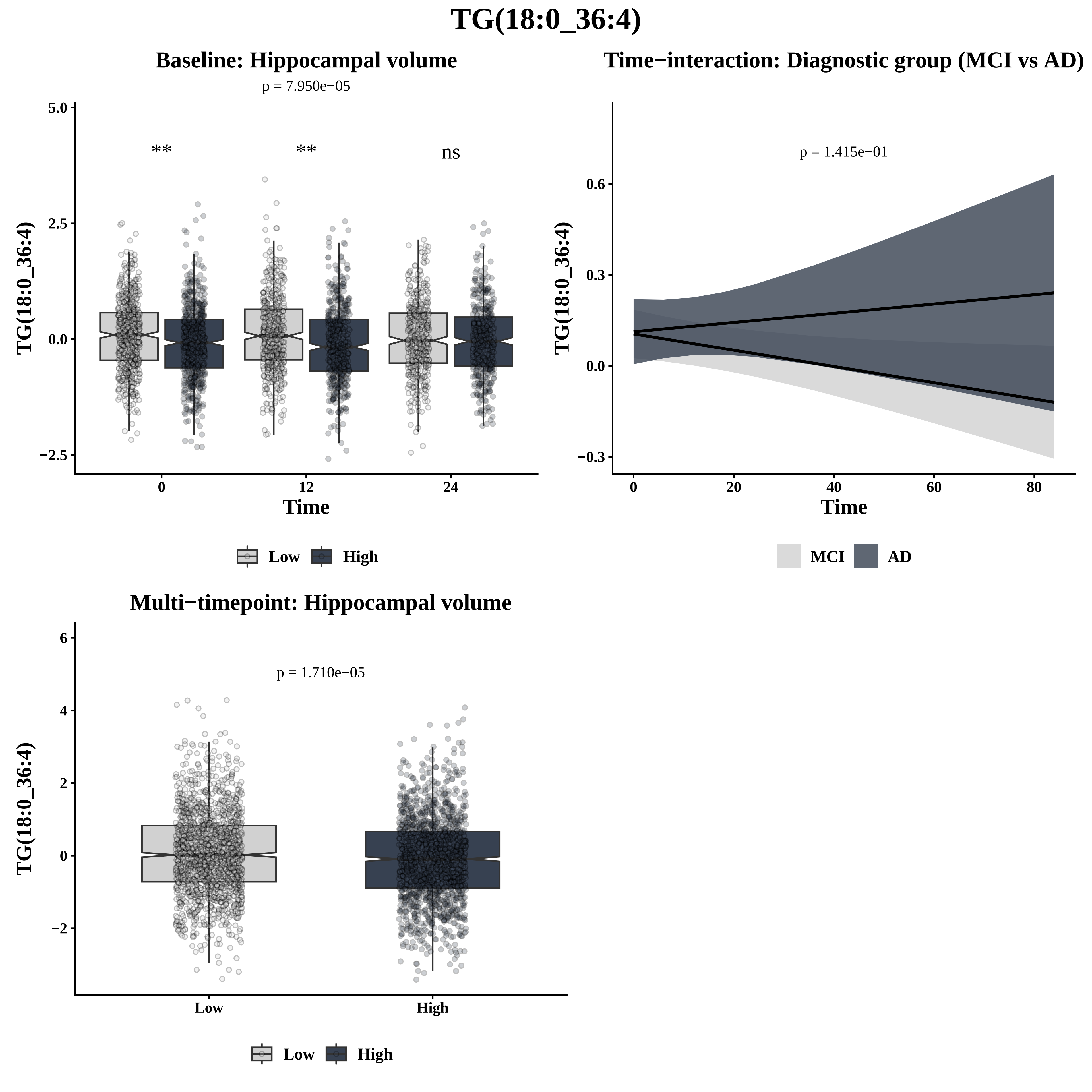
TG(18:0_36:4)
Baseline: Hippocampal volume
p = 7.950e−05
**
**
ns
TG(18:0_36:4)
5.0
2.5
0.0
−2.5
0
12
24
Time
Low
High
Time−interaction: Diagnostic group (MCI vs AD)
p = 1.415e−01
TG(18:0_36:4)
0.6
0.3
0.0
−0.3
0
20
40
60
80
Time
MCI
AD
Multi−timepoint: Hippocampal volume
p = 1.710e−05
TG(18:0_36:4)
6
4
2
0
−2
Low
High
Low
High

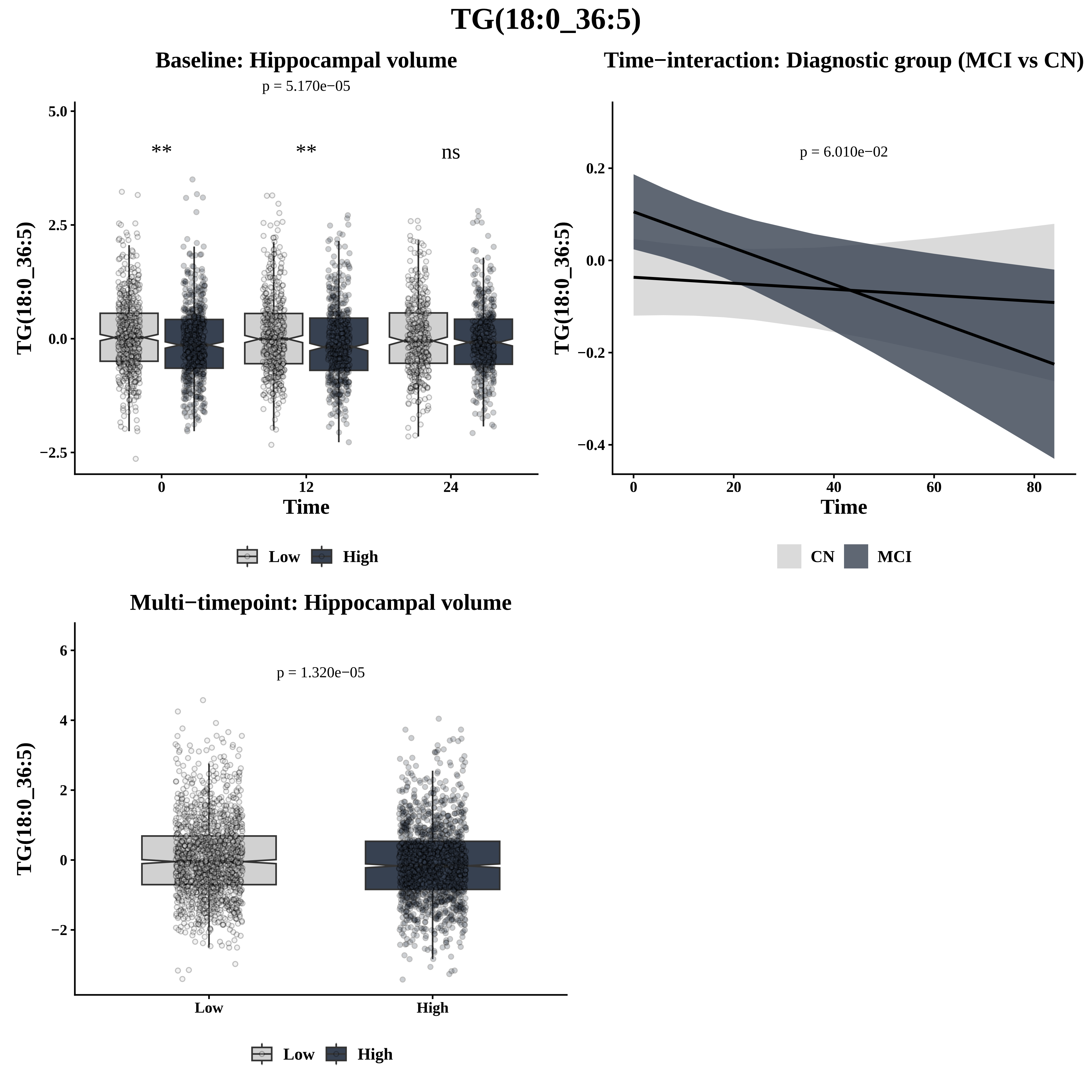

TG(18:0_36:5)
Baseline: Hippocampal volume
p = 5.170e−05
**
**
ns
5.0
2.5
0.0
−2.5
0
12
24
Time
TG(18:0_36:5)
Low
High
Time−interaction: Diagnostic group (MCI vs CN)
p = 6.010e−02
0.2
0.0
−0.2
−0.4
0
20
40
60
80
Time
TG(18:0_36:5)
CN
MCI
Multi−timepoint: Hippocampal volume
p = 1.320e−05
6
4
2
0
−2
Low
High
TG(18:0_36:5)
Low
High

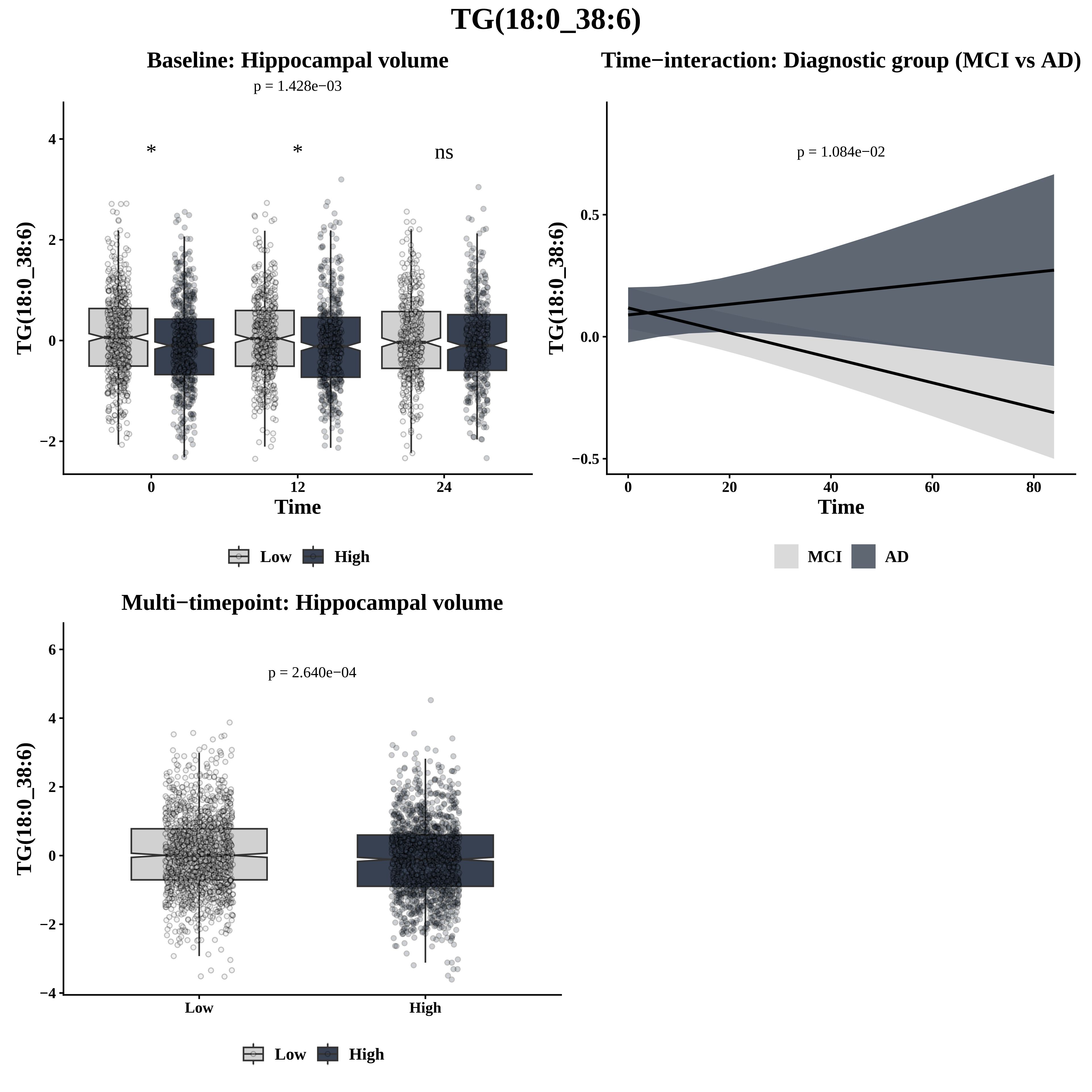
TG(18:0_38:6)
Baseline: Hippocampal volume
p = 1.428e−03
*
*
ns
TG(18:0_38:6)
Time
0
12
24
4
2
0
−2
Low
High
Time−interaction: Diagnostic group (MCI vs AD)
p = 1.084e−02
TG(18:0_38:6)
0.5
0.0
−0.5
0
20
40
60
80
Time
MCI
AD
Multi−timepoint: Hippocampal volume
p = 2.640e−04
TG(18:0_38:6)
6
4
2
0
−2
−4
Low
High
Low
High

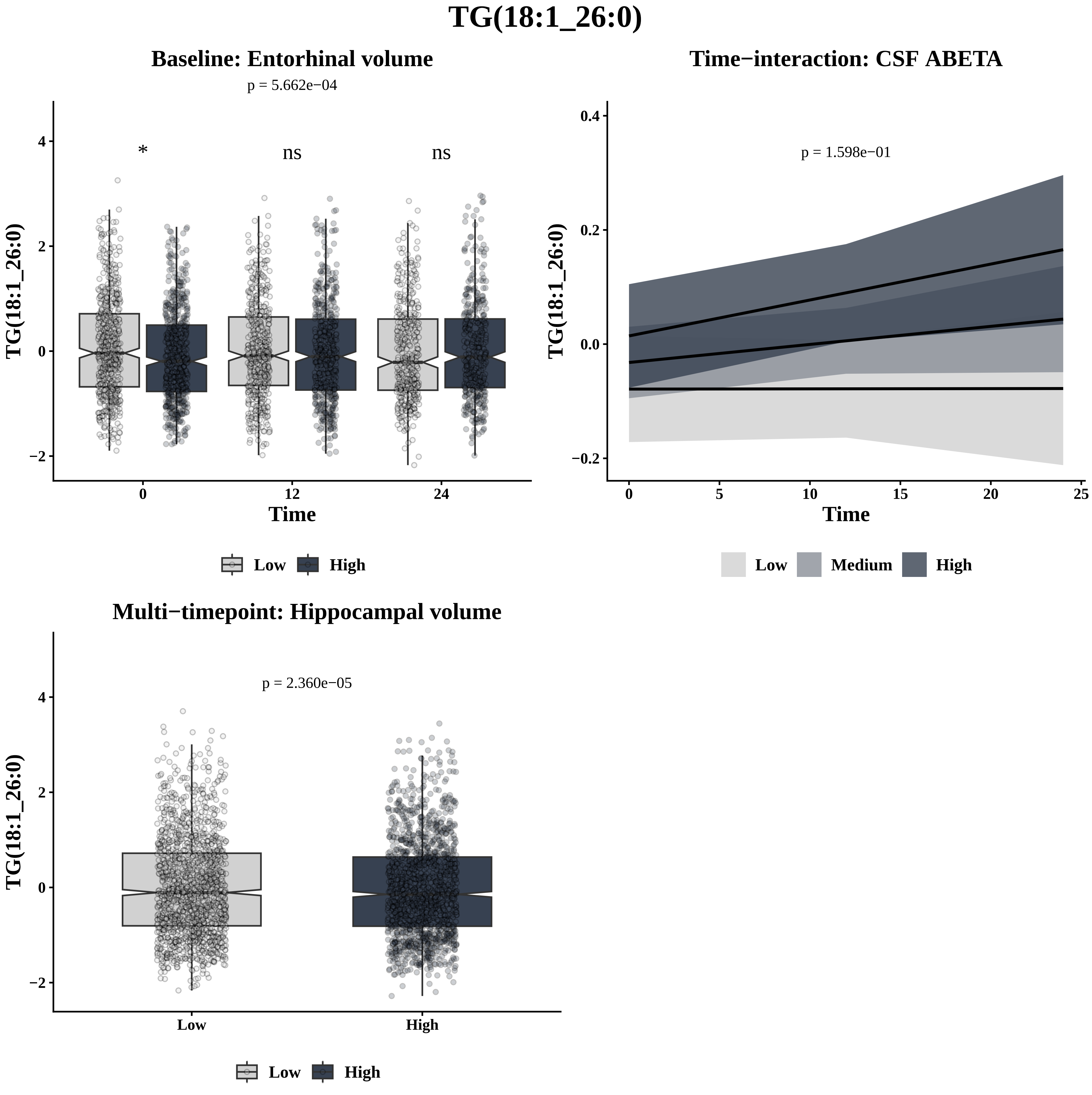

TG(18:1_26:0)
Baseline: Entorhinal volume
p = 5.662e−04
*
ns
ns
TG(18:1_26:0)
Time
Low
High
Time−interaction: CSF ABETA
p = 1.598e−01
Low
Medium
High
Multi−timepoint: Hippocampal volume
p = 2.360e−05

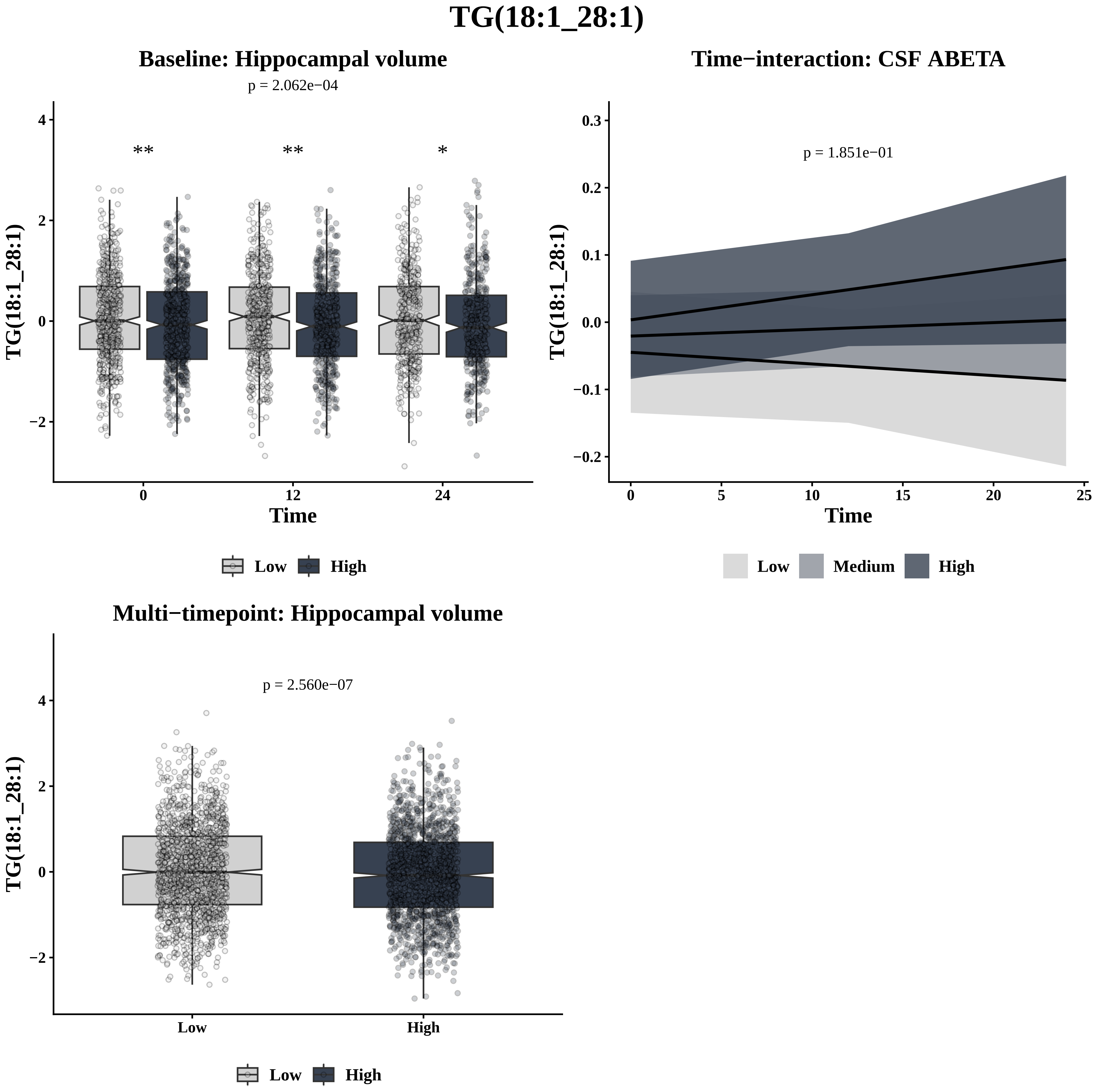

TG(18:1_28:1)
Baseline: Hippocampal volume
p = 2.062e−04
**
**
*
TG(18:1_28:1)
Time
0
12
24
Low
High
Time−interaction: CSF ABETA
p = 1.851e−01
TG(18:1_28:1)
Time
Low
Medium
High
Multi−timepoint: Hippocampal volume
p = 2.560e−07
TG(18:1_28:1)
Low
High
Low
High

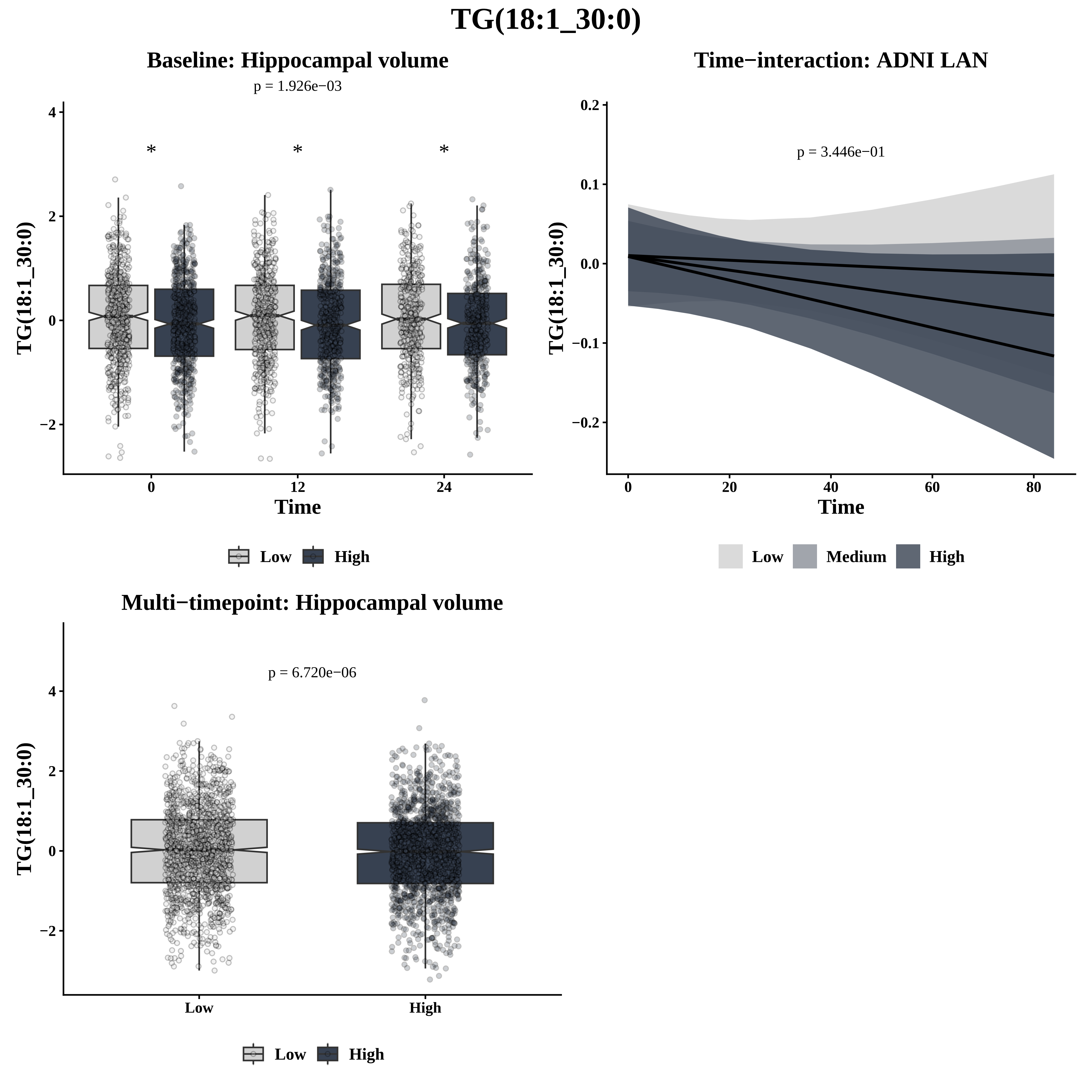

TG(18:1_30:0)
Baseline: Hippocampal volume
p = 1.926e−03
*
*
*
TG(18:1_30:0)
4
2
0
−2
0
12
24
Time
Low
High
Time−interaction: ADNI LAN
p = 3.446e−01
TG(18:1_30:0)
0.2
0.1
0.0
−0.1
−0.2
0
20
40
60
80
Time
Low
Medium
High
Multi−timepoint: Hippocampal volume
p = 6.720e−06
TG(18:1_30:0)
4
2
0
−2
Low
High
Low
High

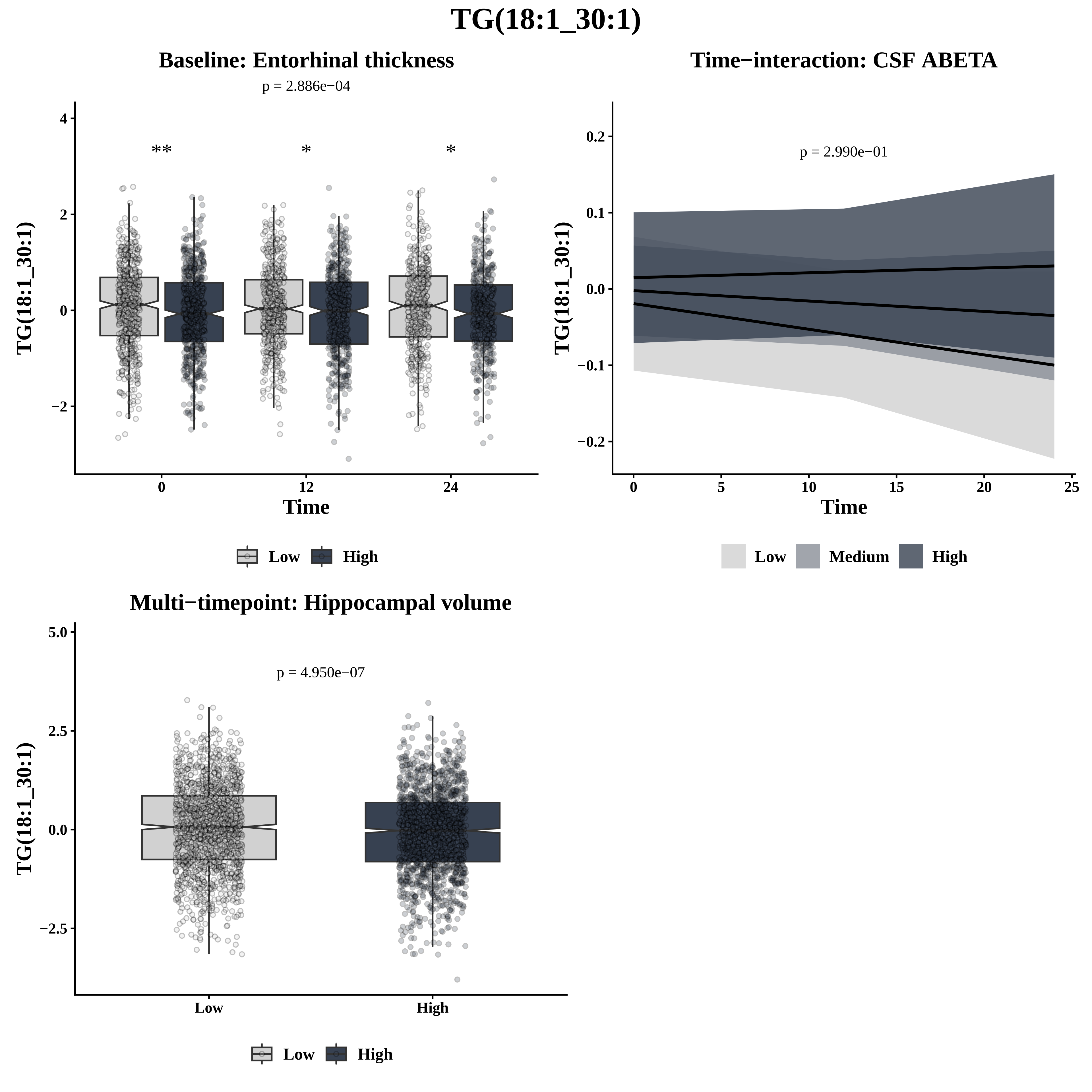

TG(18:1_30:1)
Baseline: Entorhinal thickness
p = 2.886e−04
**
*
*
TG(18:1_30:1)
Time
0
12
24
4
2
0
−2
Low
High
Time−interaction: CSF ABETA
p = 2.990e−01
0.2
0.1
0.0
−0.1
−0.2
0
5
10
15
20
25
Time
Low
Medium
High
Multi−timepoint: Hippocampal volume
p = 4.950e−07
5.0
2.5
0.0
−2.5
Low
High

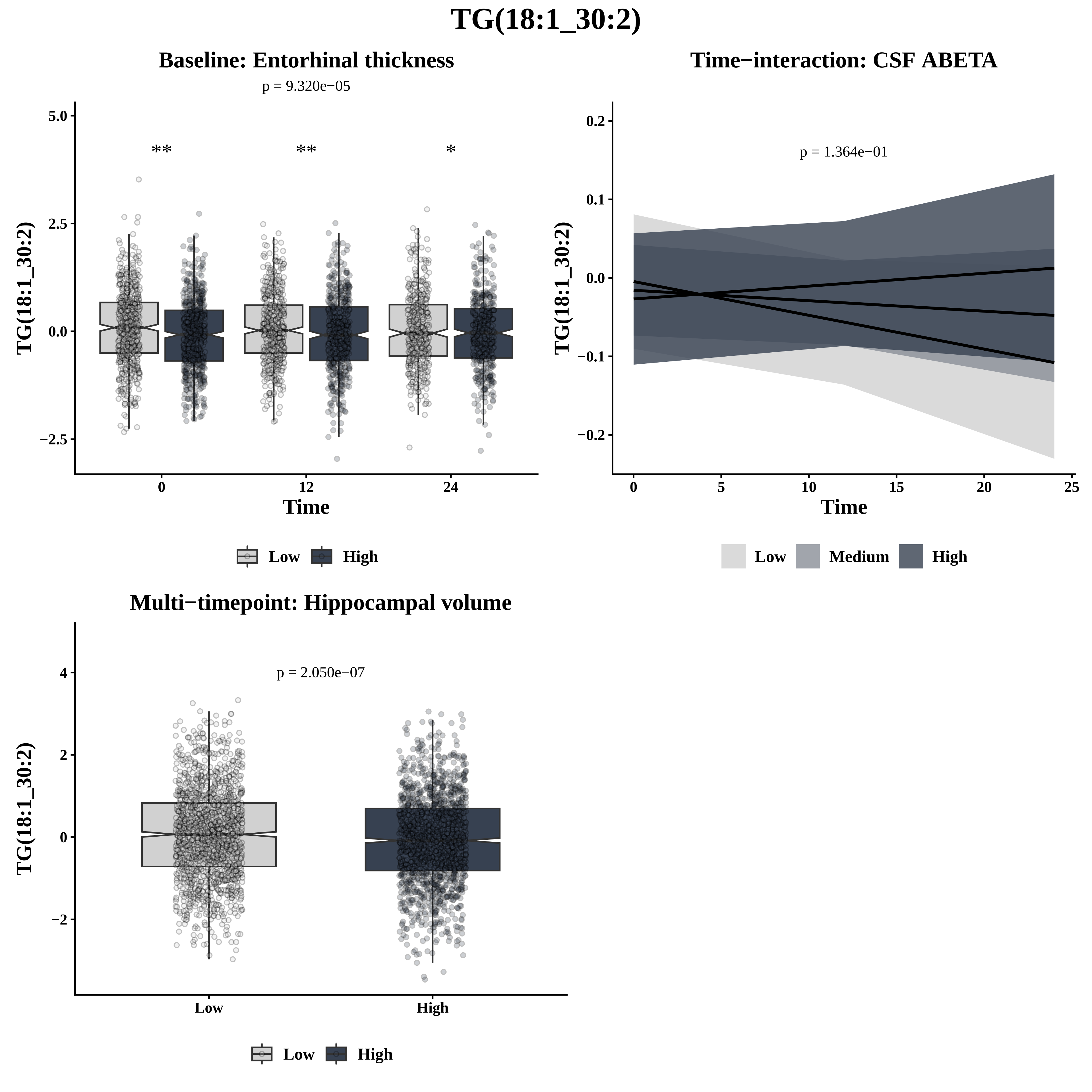

TG(18:1_30:2)
Baseline: Entorhinal thickness
p = 9.320e−05
5.0
2.5
0.0
−2.5
**
**
*
0
12
24
Time
TG(18:1_30:2)
Low
High
Time−interaction: CSF ABETA
p = 1.364e−01
0.2
0.1
0.0
−0.1
−0.2
0
5
10
15
20
25
Time
TG(18:1_30:2)
Low
Medium
High
Multi−timepoint: Hippocampal volume
p = 2.050e−07
4
2
0
−2
Low
High
TG(18:1_30:2)
Low
High

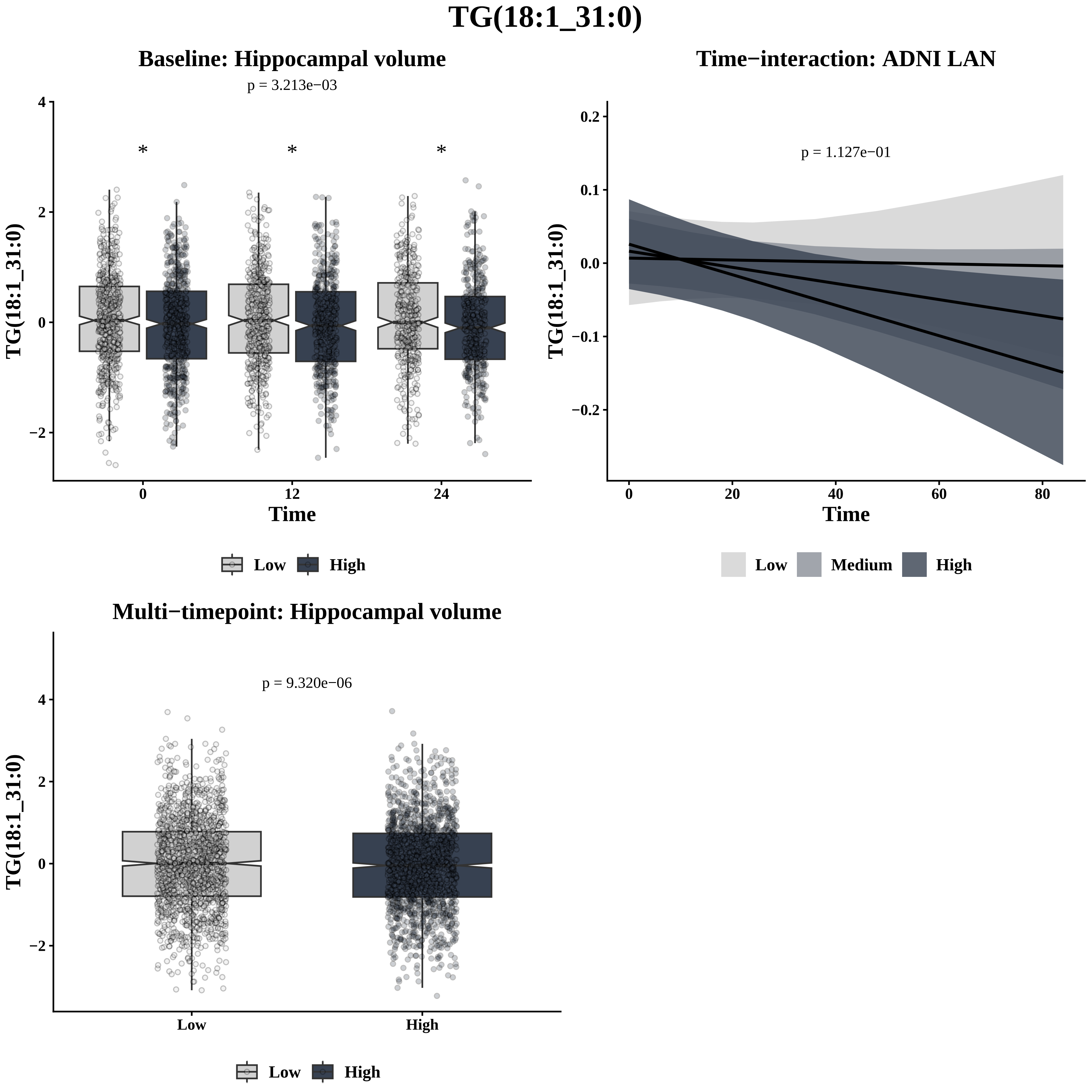
TG(18:1_31:0)
Baseline: Hippocampal volume
p = 3.213e−03
TG(18:1_31:0)
Time
0
12
24
Low
High
Time−interaction: ADNI LAN
p = 1.127e−01
TG(18:1_31:0)
Time
Low
Medium
High
Multi−timepoint: Hippocampal volume
p = 9.320e−06
TG(18:1_31:0)
Low
High

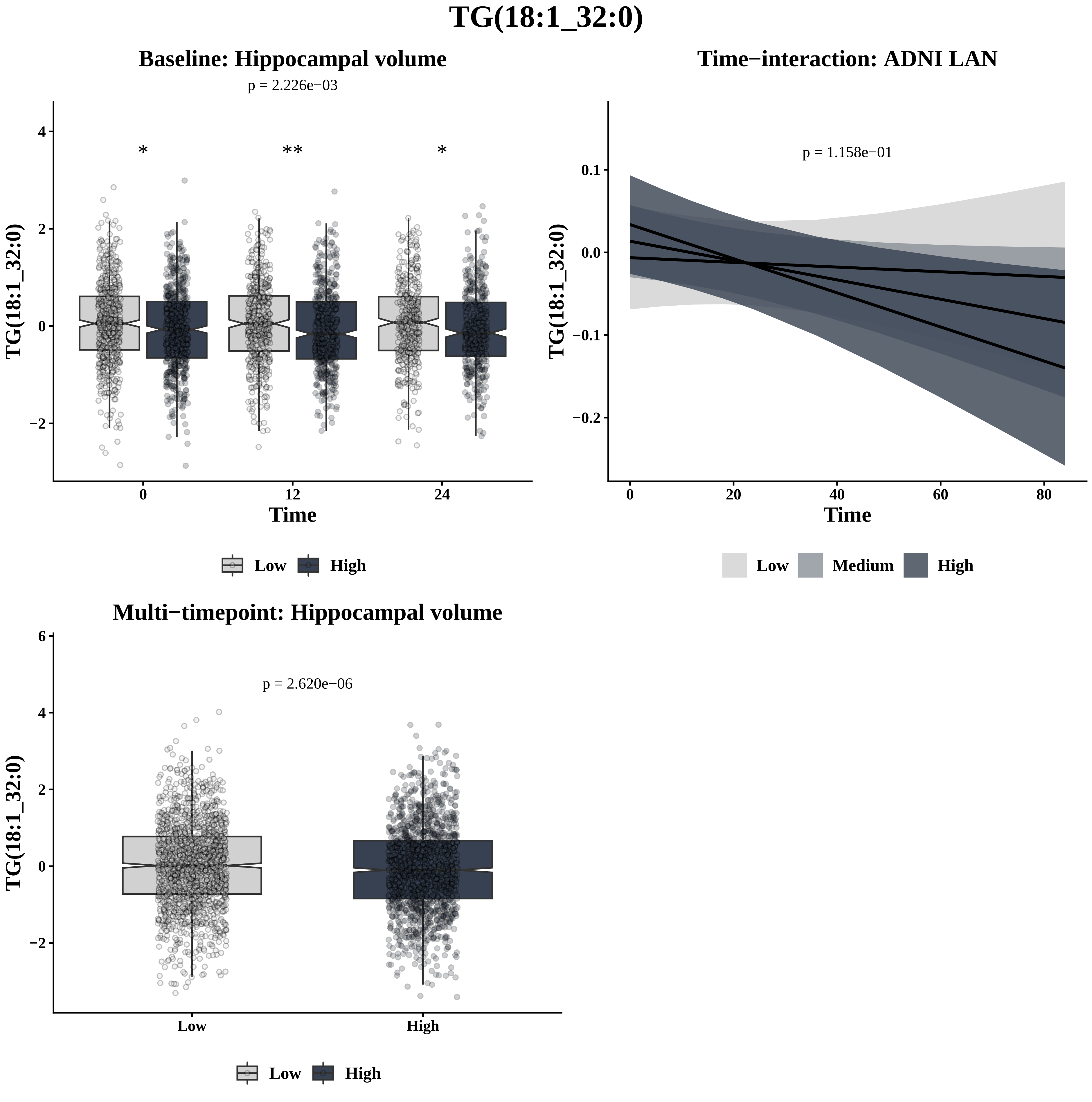

TG(18:1_32:0)
Baseline: Hippocampal volume
p = 2.226e−03
*
**
*
TG(18:1_32:0)
Time
0
12
24
Low
High
Time−interaction: ADNI LAN
p = 1.158e−01
TG(18:1_32:0)
Time
Low
Medium
High
Multi−timepoint: Hippocampal volume
p = 2.620e−06
TG(18:1_32:0)
Low
High

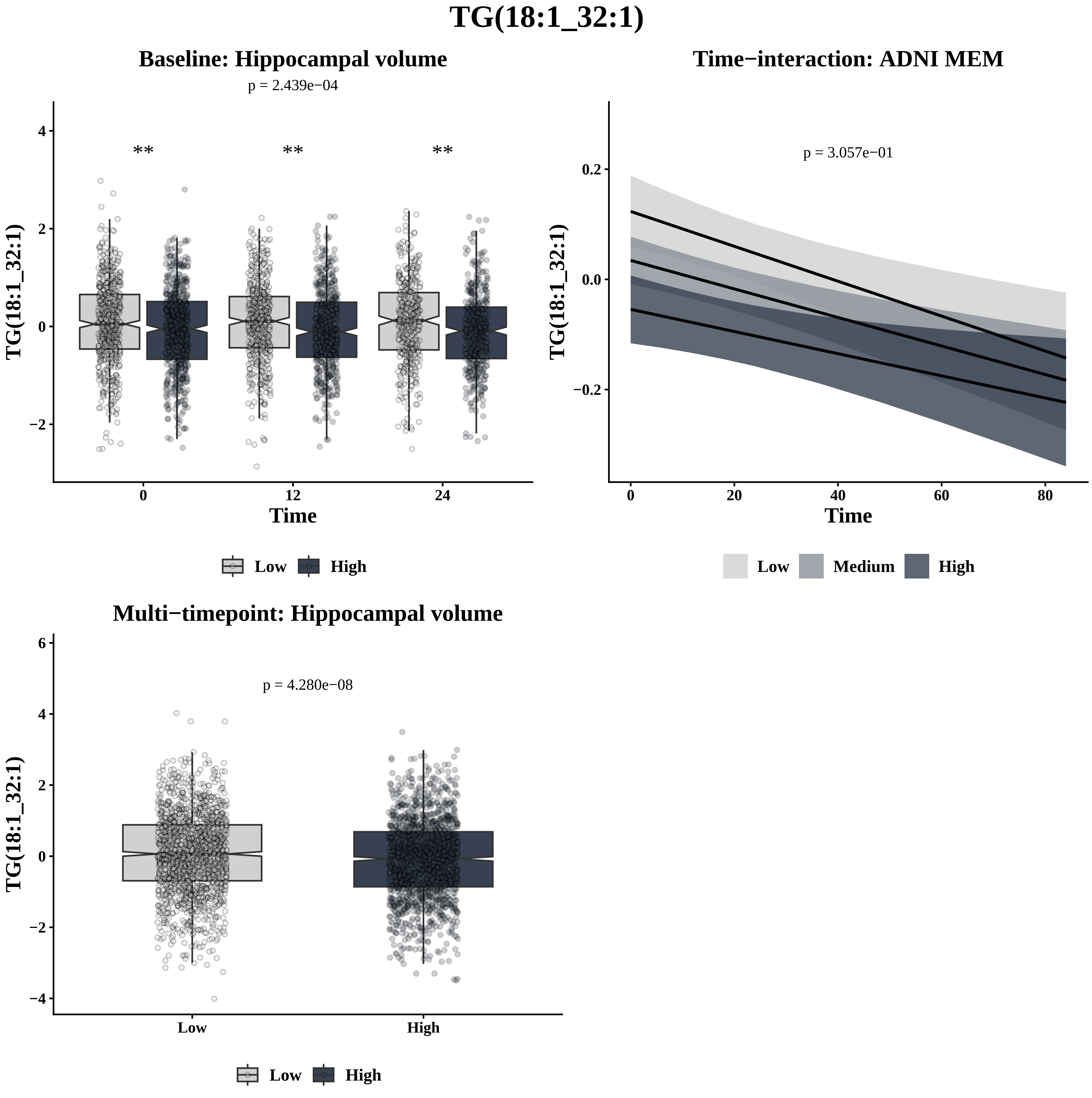
TG(18:1_32:1)
Baseline: Hippocampal volume
p = 2.439e−04
**
**
**
TG(18:1_32:1)
4
2
0
−2
0
12
24
Time
Low
High
Time−interaction: ADNI MEM
p = 3.057e−01
TG(18:1_32:1)
0.2
0.0
−0.2
0
20
40
60
80
Time
Low
Medium
High
Multi−timepoint: Hippocampal volume
p = 4.280e−08
TG(18:1_32:1)
6
4
2
0
−2
−4
Low
High
Low
High

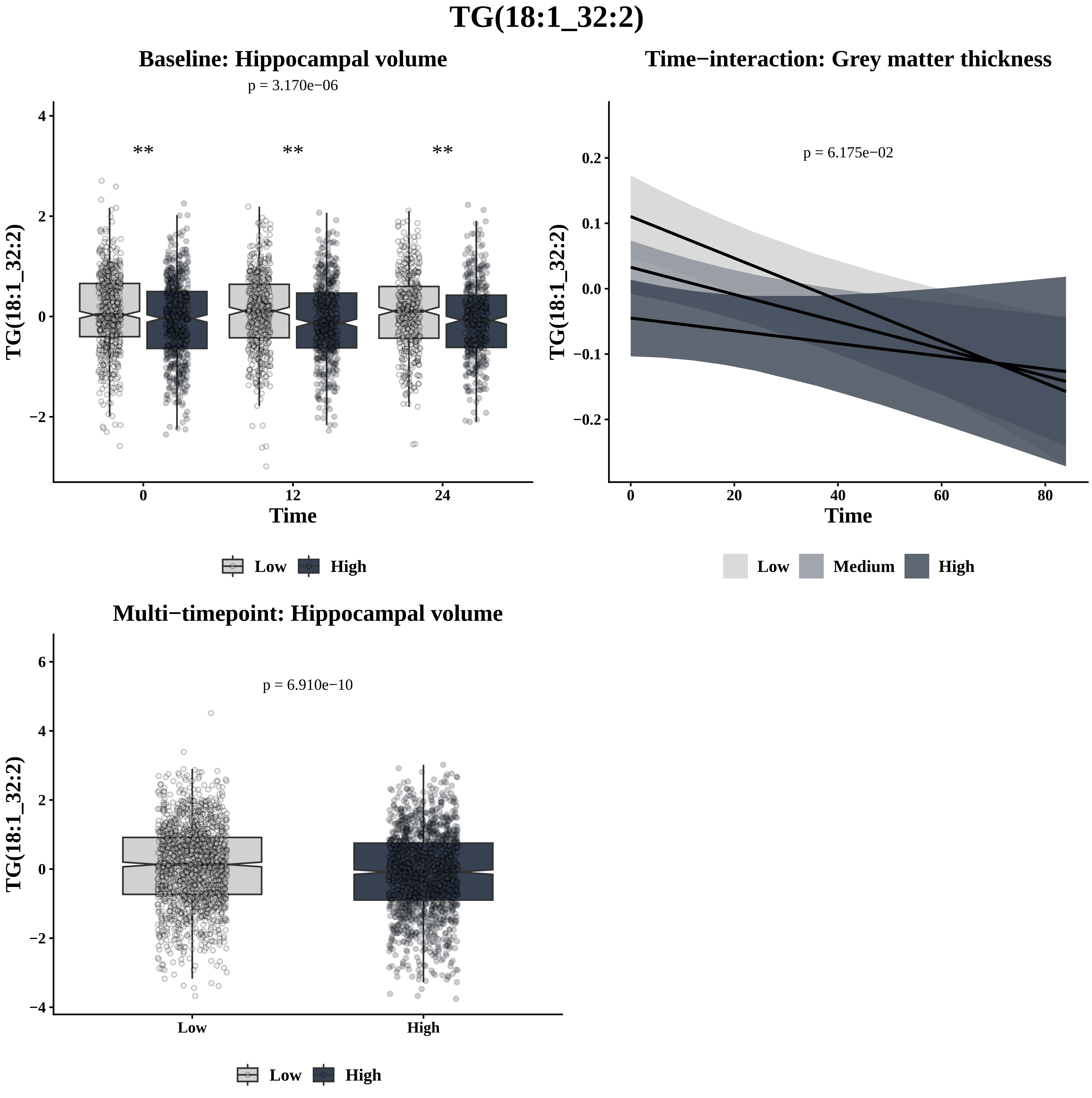

TG(18:1_32:2)
Baseline: Hippocampal volume
p = 3.170e−06
**
**
**
TG(18:1_32:2)
Time
0
12
24
Low
High
Time−interaction: Grey matter thickness
p = 6.175e−02
TG(18:1_32:2)
Time
0
20
40
60
80
Low
Medium
High
Multi−timepoint: Hippocampal volume
p = 6.910e−10
TG(18:1_32:2)
Low
High

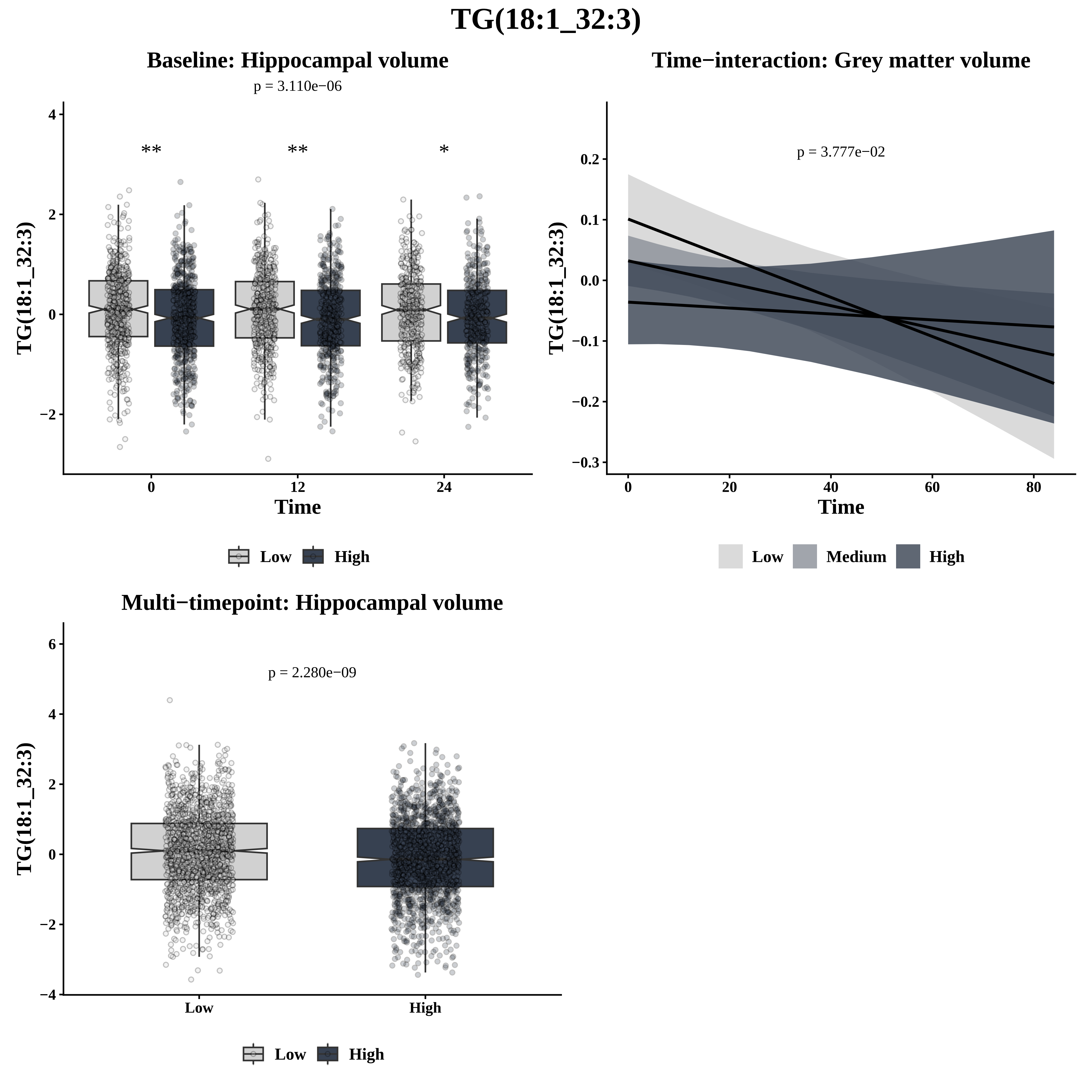
TG(18:1_32:3)
Baseline: Hippocampal volume
p = 3.110e−06
**
**
*
TG(18:1_32:3)
Time
0
12
24
Low
High
Time−interaction: Grey matter volume
p = 3.777e−02
TG(18:1_32:3)
Time
Low
Medium
High
Multi−timepoint: Hippocampal volume
p = 2.280e−09
TG(18:1_32:3)
Low
High
Low
High

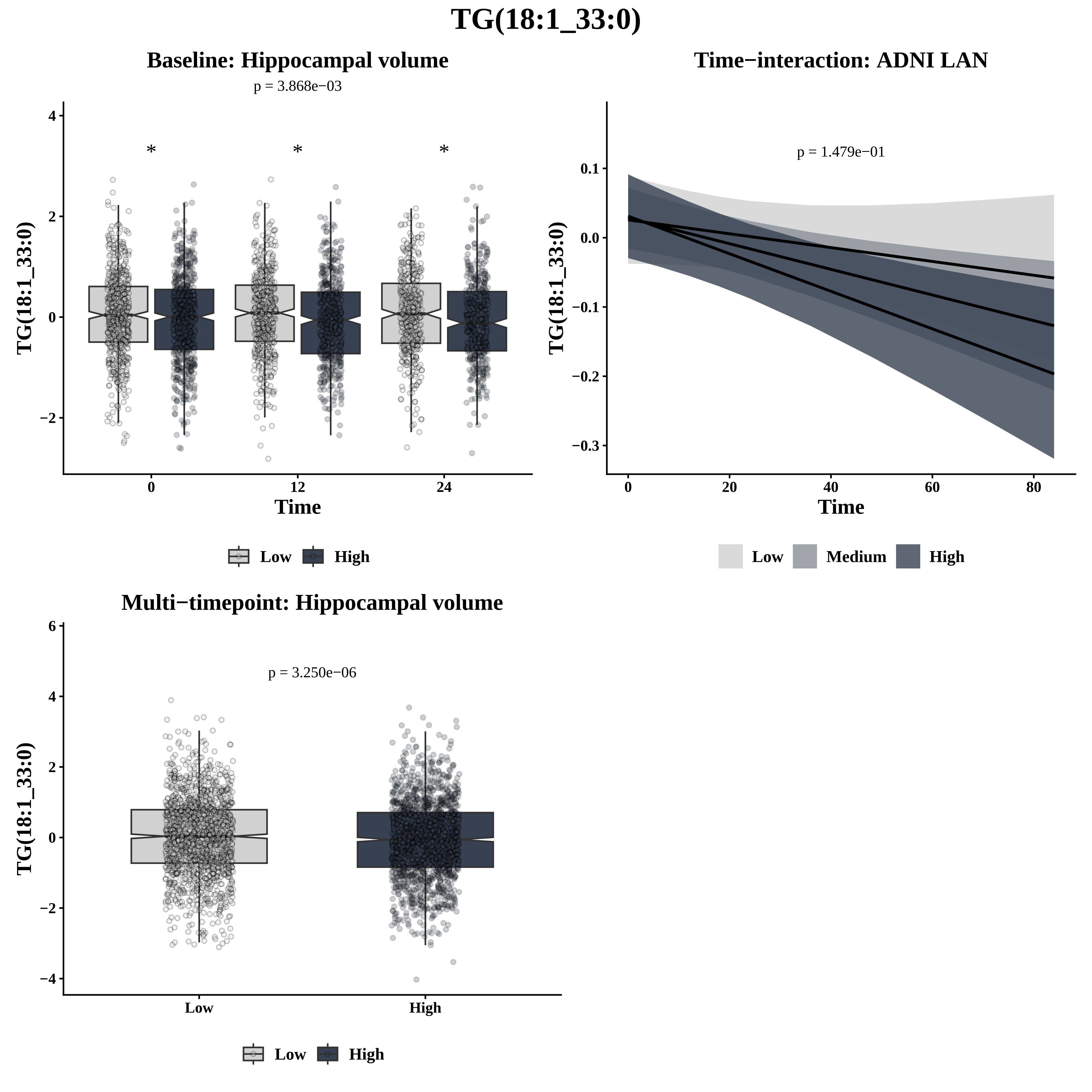
TG(18:1_33:0)
Baseline: Hippocampal volume
p = 3.868e−03
TG(18:1_33:0)
Time
Low
High
Time−interaction: ADNI LAN
p = 1.479e−01
TG(18:1_33:0)
Time
Low
Medium
High
Multi−timepoint: Hippocampal volume
p = 3.250e−06
TG(18:1_33:0)
Low
High

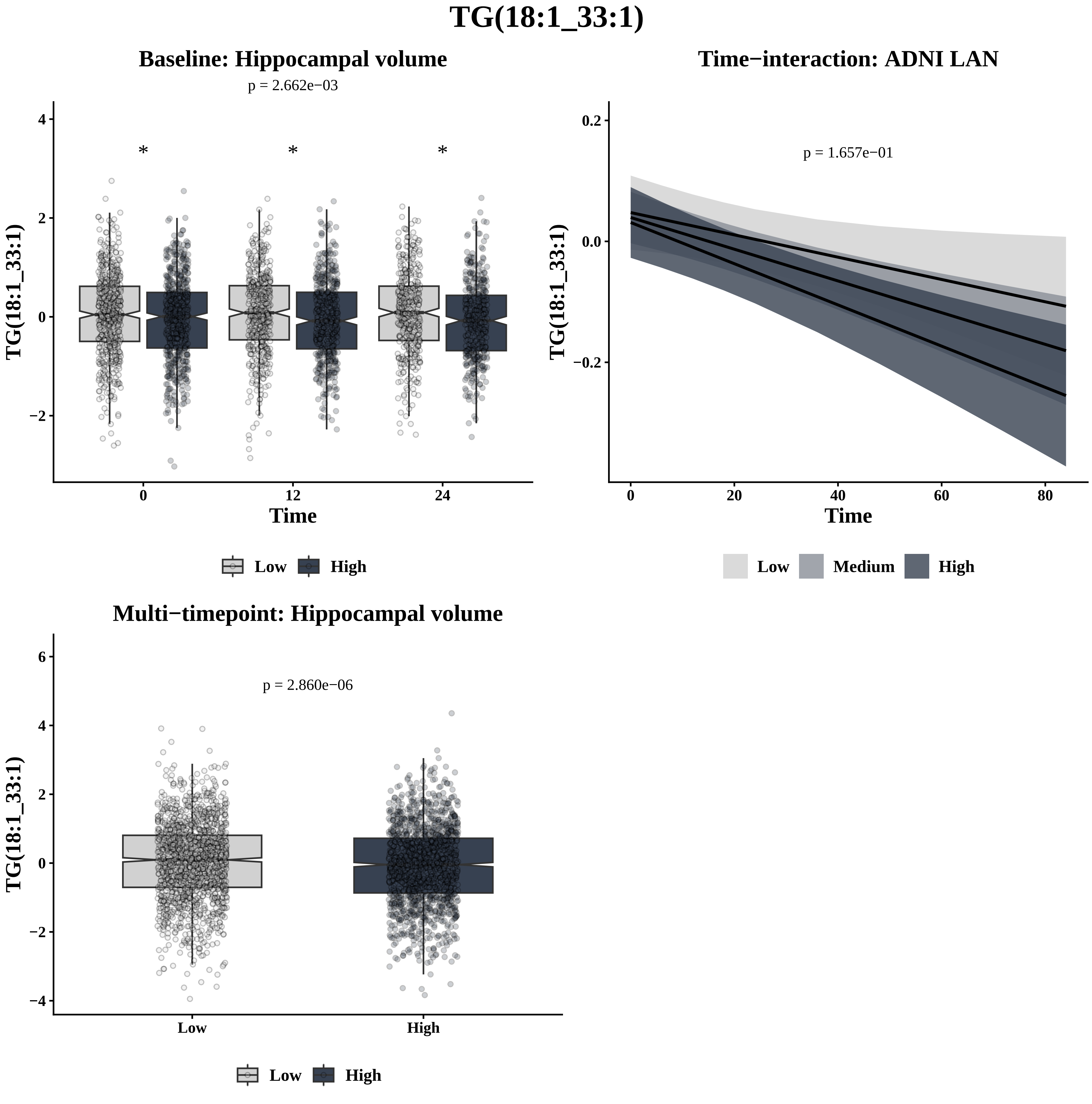

TG(18:1_33:1)
Baseline: Hippocampal volume
p = 2.662e−03
*
*
*
TG(18:1_33:1)
Time
0
12
24
4
2
0
−2
Low
High
Time−interaction: ADNI LAN
p = 1.657e−01
0.2
0.0
−0.2
0
20
40
60
80
Time
Low
Medium
High
Multi−timepoint: Hippocampal volume
p = 2.860e−06
6
4
2
0
−2
−4
Low
High
Low
High

# TG(18:1_33:2)

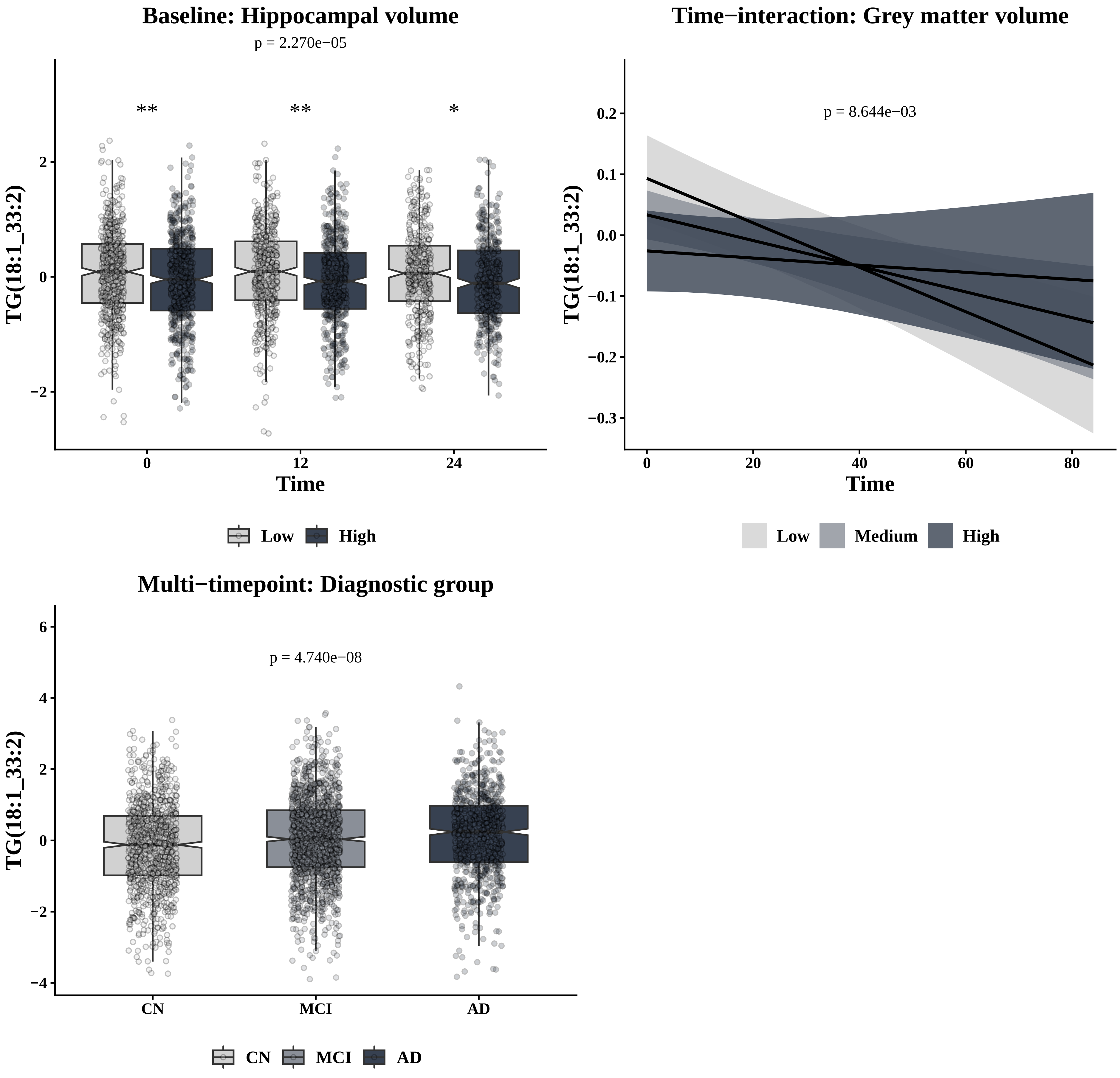

Baseline: Hippocampal volume
p = 2.270e−05
**
**
*
TG(18:1_33:2)
2
0
−2
0
12
24
Time
Low
High
Time−interaction: Grey matter volume
p = 8.644e−03
TG(18:1_33:2)
0.2
0.1
0.0
−0.1
−0.2
−0.3
0
20
40
60
80
Time
Low
Medium
High
Multi−timepoint: Diagnostic group
p = 4.740e−08
TG(18:1_33:2)
6
4
2
0
−2
−4
CN
MCI
AD
CN
MCI
AD

# TG(18:1_33:3)

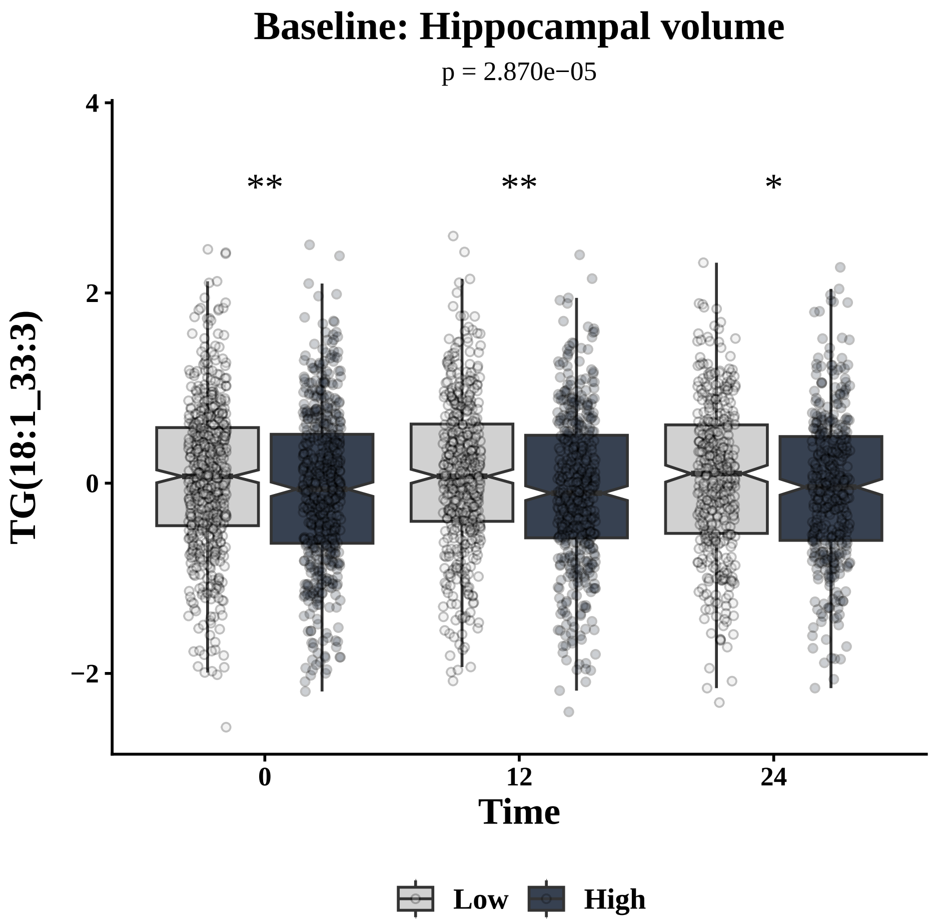

Baseline: Hippocampal volume
p = 2.870e−05
**
**
*
TG(18:1_33:3)
Time
0
12
24
Low
High


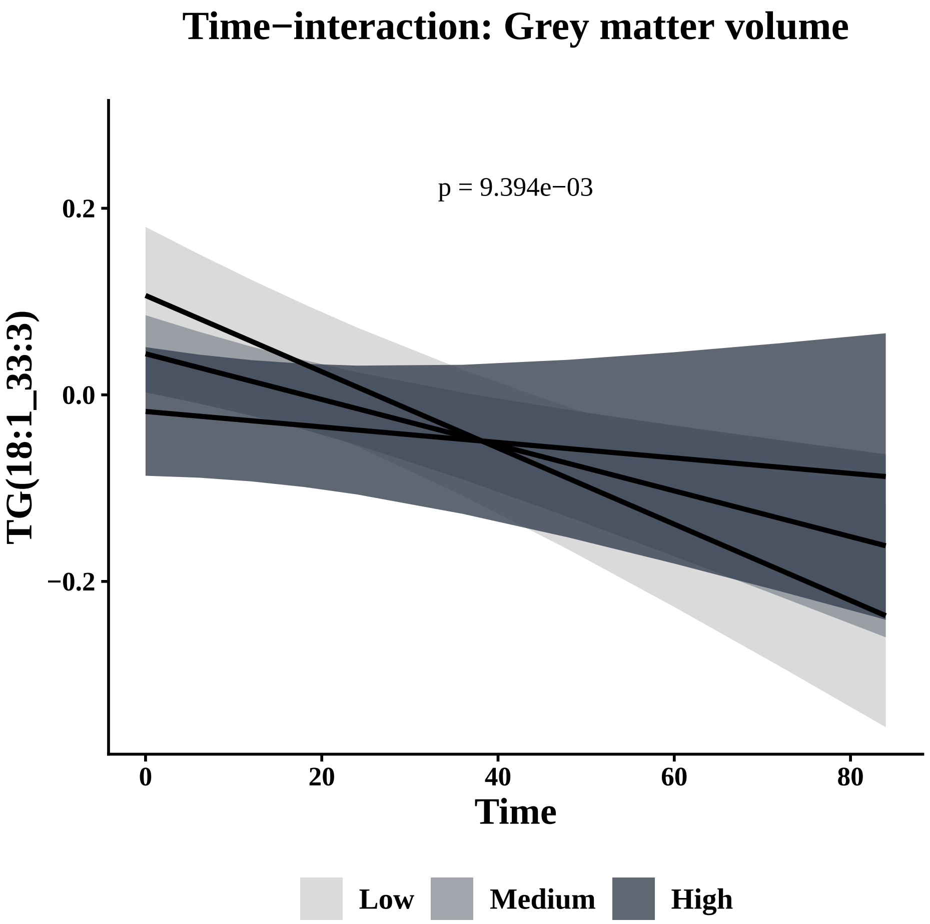

Time−interaction: Grey matter volume
p = 9.394e−03
TG(18:1_33:3)
Time
Low
Medium
High


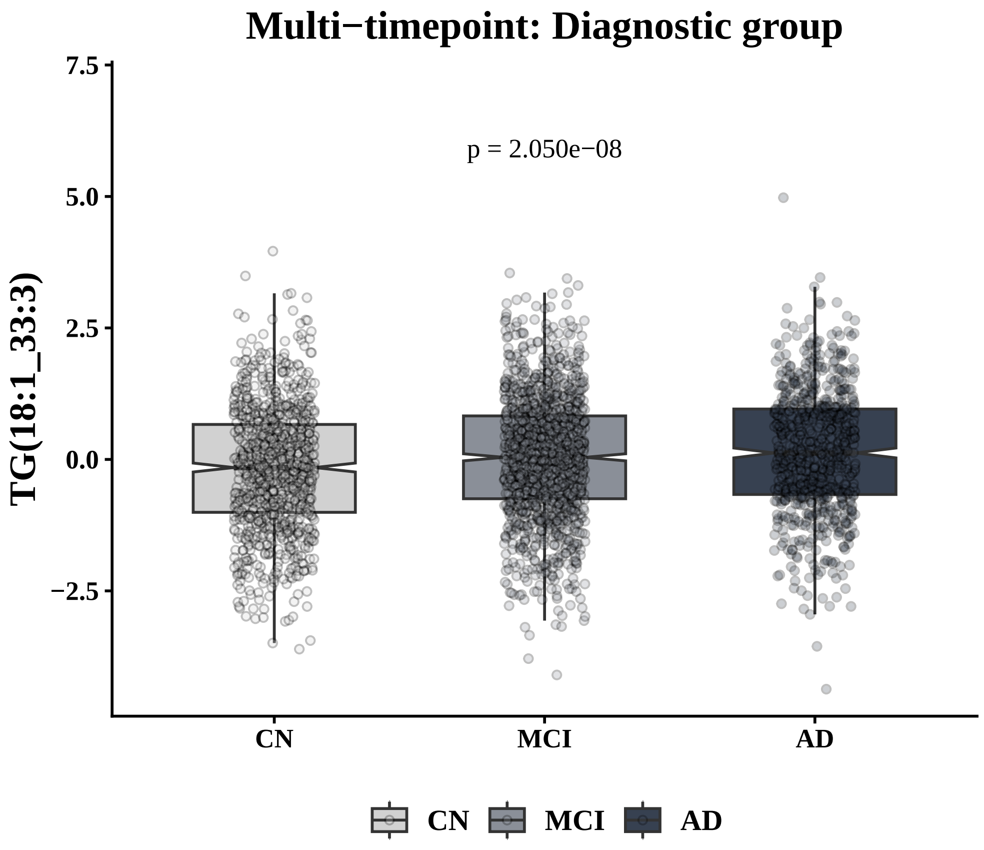

Multi−timepoint: Diagnostic group
p = 2.050e−08
TG(18:1_33:3)
CN
MCI
AD

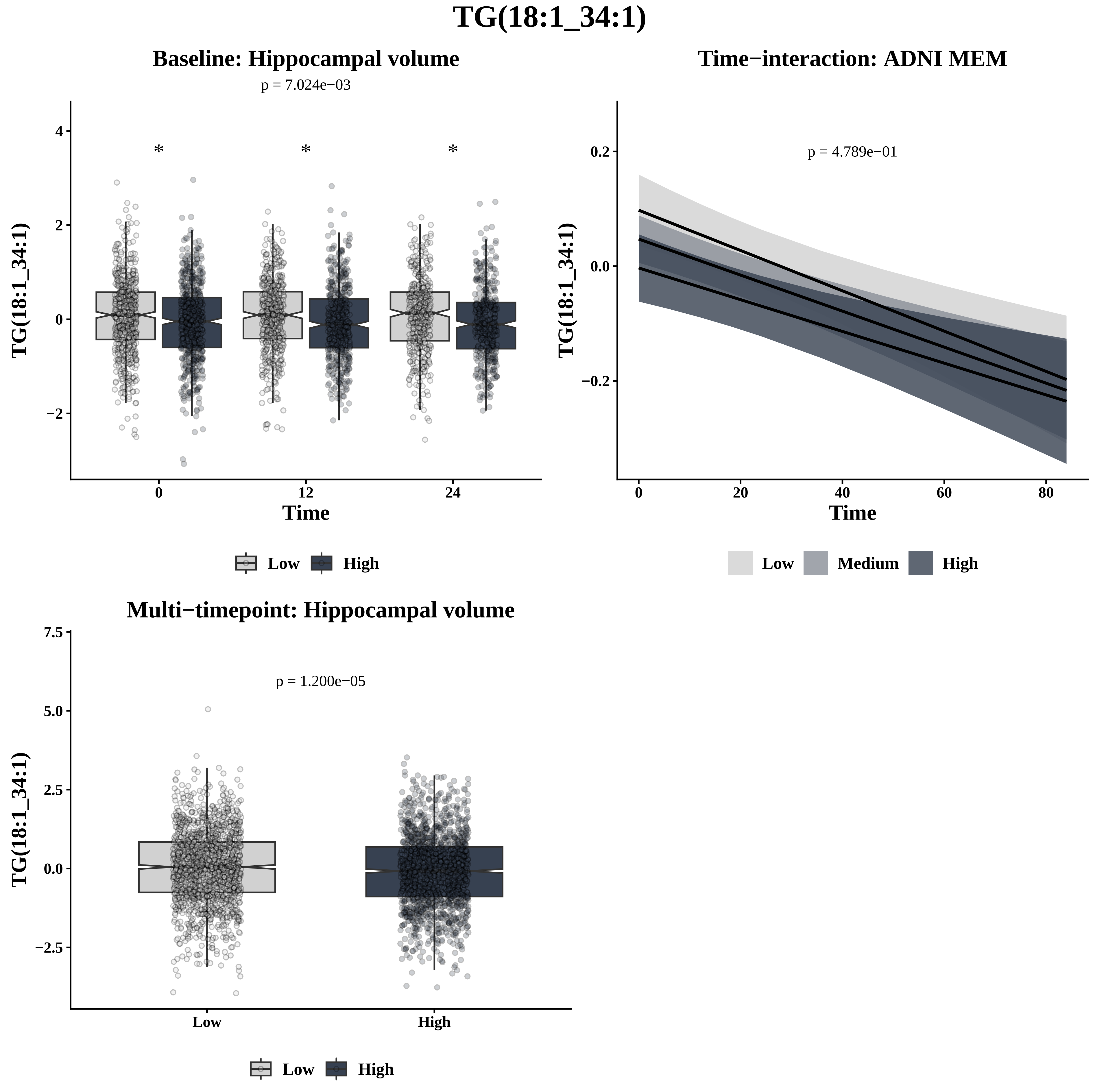
TG(18:1_34:1)
Baseline: Hippocampal volume
p = 7.024e−03
*
*
*
4
2
0
−2
TG(18:1_34:1)
0
12
24
Time
Low
High
Time−interaction: ADNI MEM
p = 4.789e−01
0.2
0.0
−0.2
TG(18:1_34:1)
0
20
40
60
80
Time
Low
Medium
High
Multi−timepoint: Hippocampal volume
p = 1.200e−05
7.5
5.0
2.5
0.0
−2.5
TG(18:1_34:1)
Low
High
Low
High

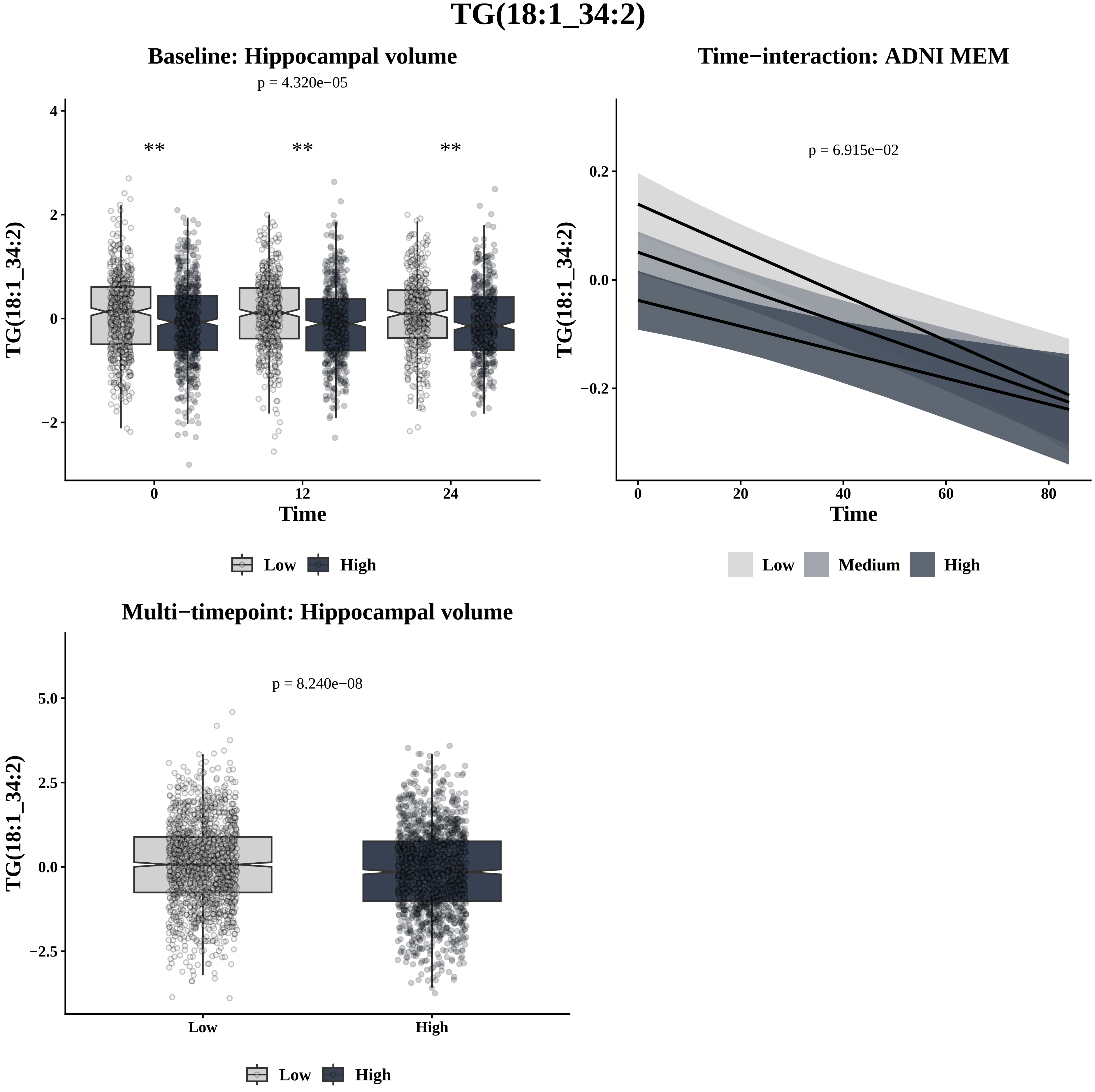
TG(18:1_34:2)
Baseline: Hippocampal volume
p = 4.320e−05
**
**
**
TG(18:1_34:2)
4
2
0
−2
0
12
24
Time
Low
High
Time−interaction: ADNI MEM
p = 6.915e−02
0.2
0.0
−0.2
0
20
40
60
80
Time
Low
Medium
High
Multi−timepoint: Hippocampal volume
p = 8.240e−08
5.0
2.5
0.0
−2.5
Low
High

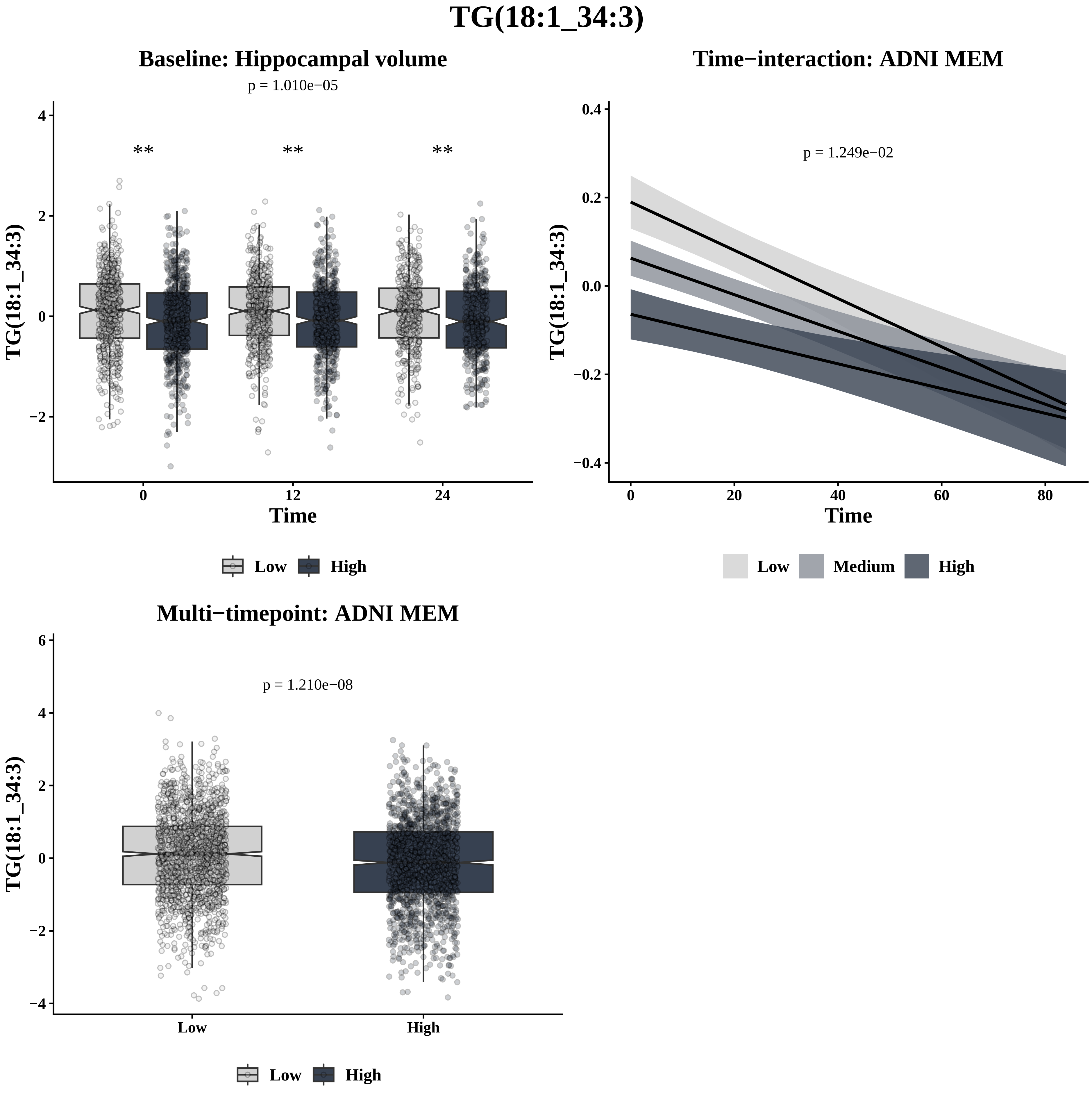
TG(18:1_34:3)
Baseline: Hippocampal volume
p = 1.010e−05
**
**
**
TG(18:1_34:3)
Time
0
12
24
4
2
0
−2
Low
High
Time−interaction: ADNI MEM
p = 1.249e−02
TG(18:1_34:3)
Time
0.4
0.2
0.0
−0.2
−0.4
0
20
40
60
80
Low
Medium
High
Multi−timepoint: ADNI MEM
p = 1.210e−08
TG(18:1_34:3)
6
4
2
0
−2
−4
Low
High
Low
High

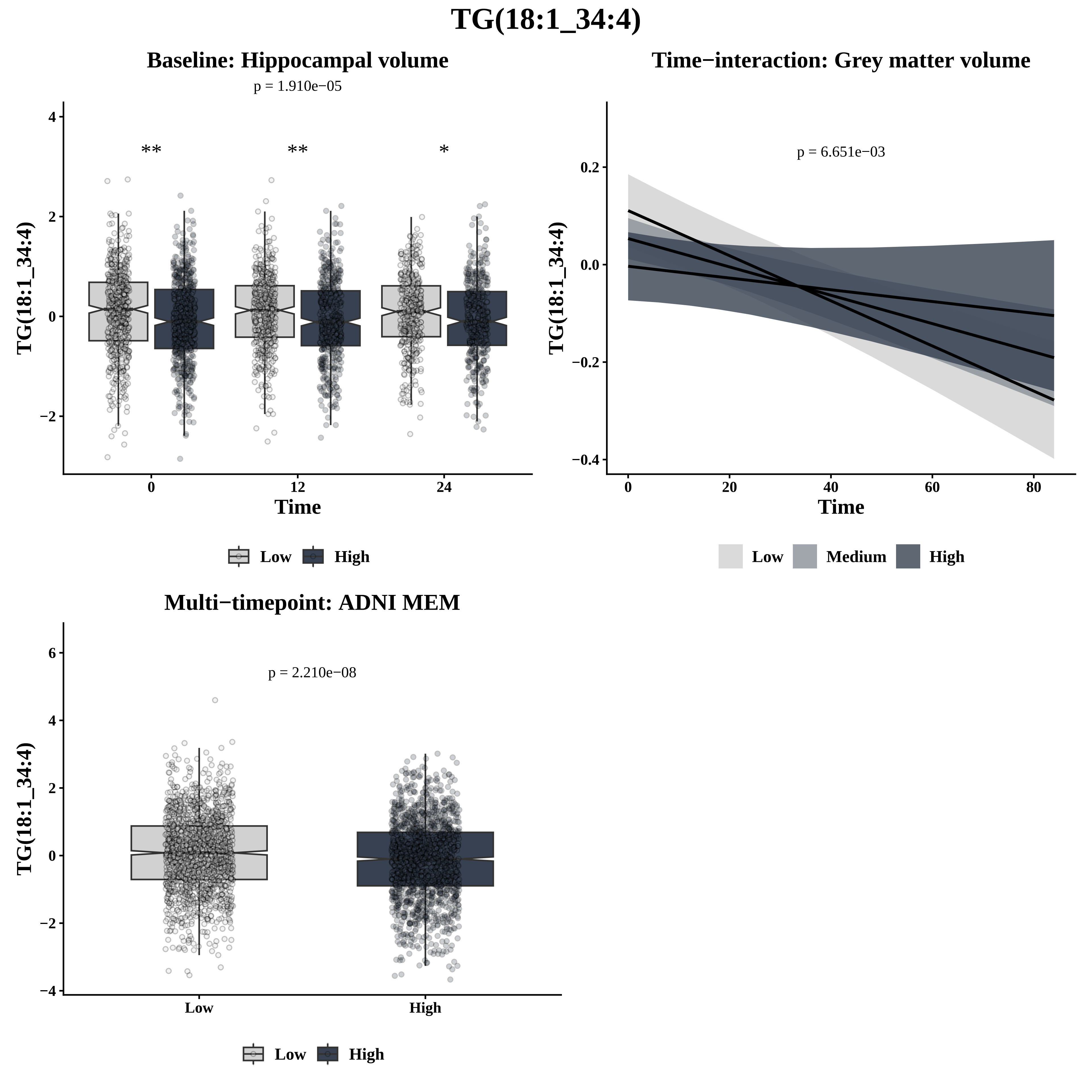

TG(18:1_34:4)
Baseline: Hippocampal volume
p = 1.910e−05
**
**
*
TG(18:1_34:4)
Time
0
12
24
Low
High
Time−interaction: Grey matter volume
p = 6.651e−03
TG(18:1_34:4)
Time
Low
Medium
High
Multi−timepoint: ADNI MEM
p = 2.210e−08
TG(18:1_34:4)
Low
High

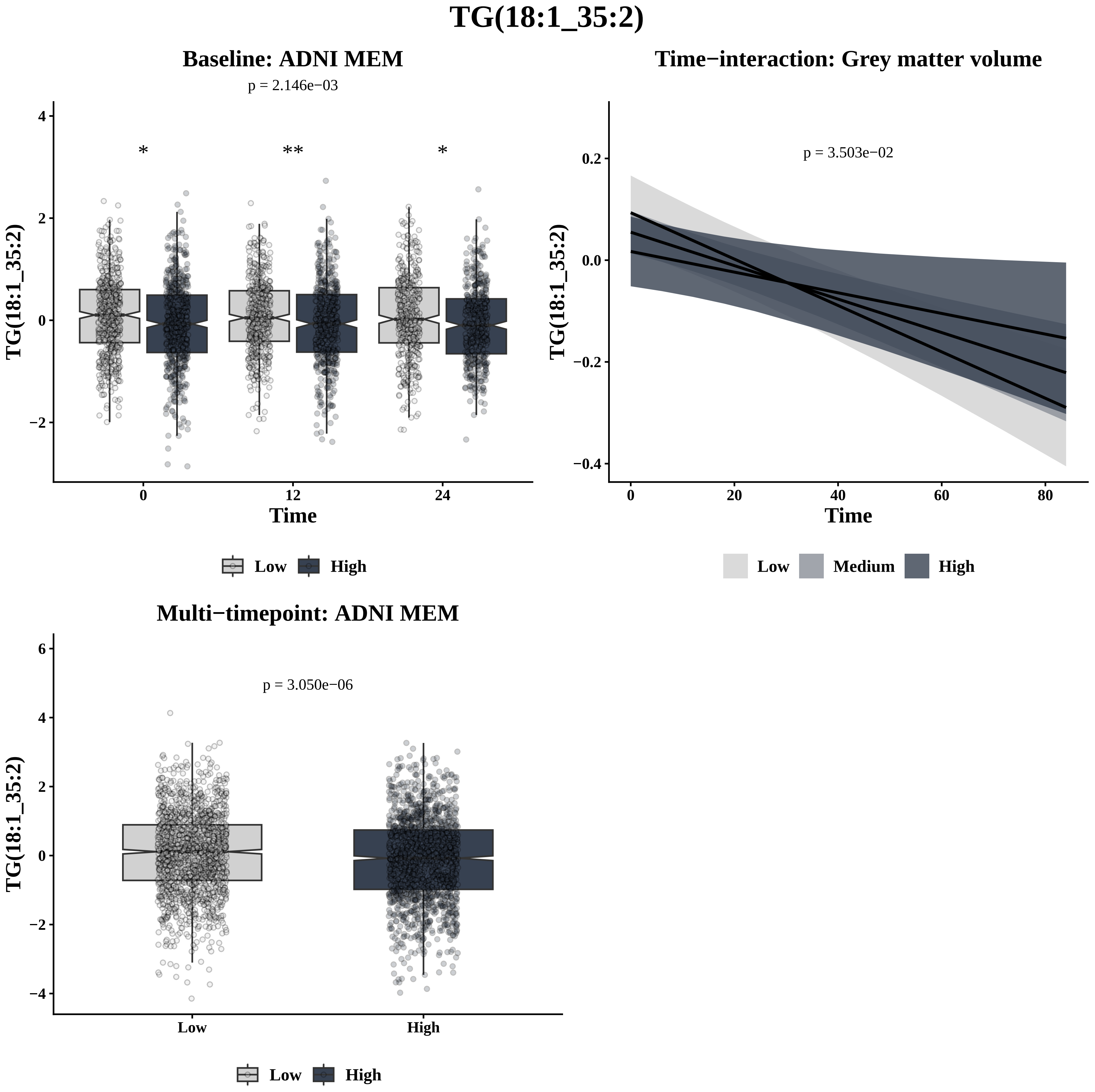
TG(18:1_35:2)
Baseline: ADNI MEM
p = 2.146e−03
*
**
*
4
2
0
−2
TG(18:1_35:2)
0
12
24
Time
Low
High
Time−interaction: Grey matter volume
p = 3.503e−02
0.2
0.0
−0.2
−0.4
TG(18:1_35:2)
0
20
40
60
80
Time
Low
Medium
High
Multi−timepoint: ADNI MEM
p = 3.050e−06
6
4
2
0
−2
−4
TG(18:1_35:2)
Low
High
Low
High

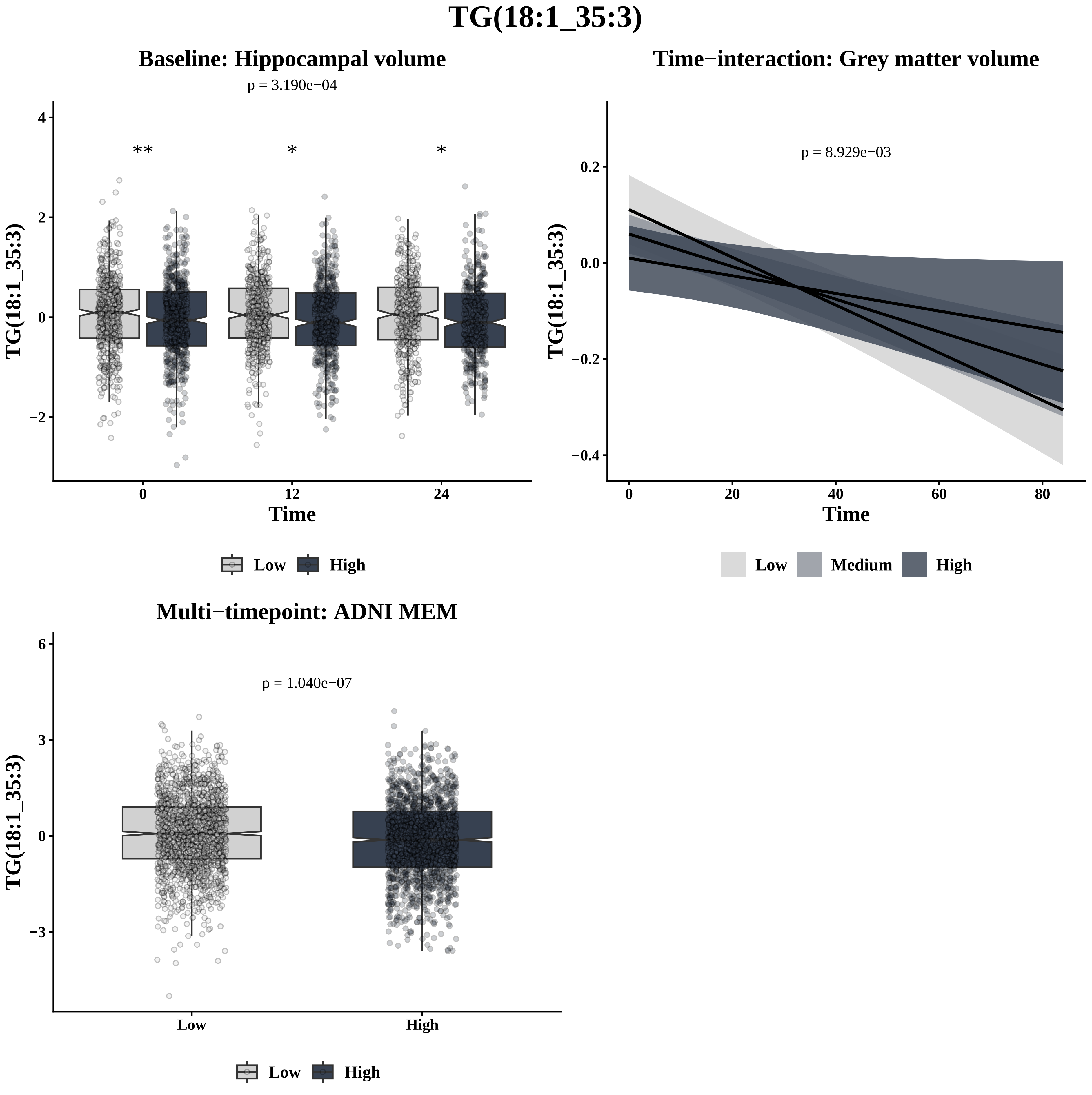

TG(18:1_35:3)
Baseline: Hippocampal volume
p = 3.190e−04
**
*
*
4
2
0
−2
0
12
24
Time
TG(18:1_35:3)
Low
High
Time−interaction: Grey matter volume
p = 8.929e−03
0.2
0.0
−0.2
−0.4
0
20
40
60
80
Time
TG(18:1_35:3)
Low
Medium
High
Multi−timepoint: ADNI MEM
p = 1.040e−07
6
3
0
−3
Low
High
TG(18:1_35:3)
Low
High

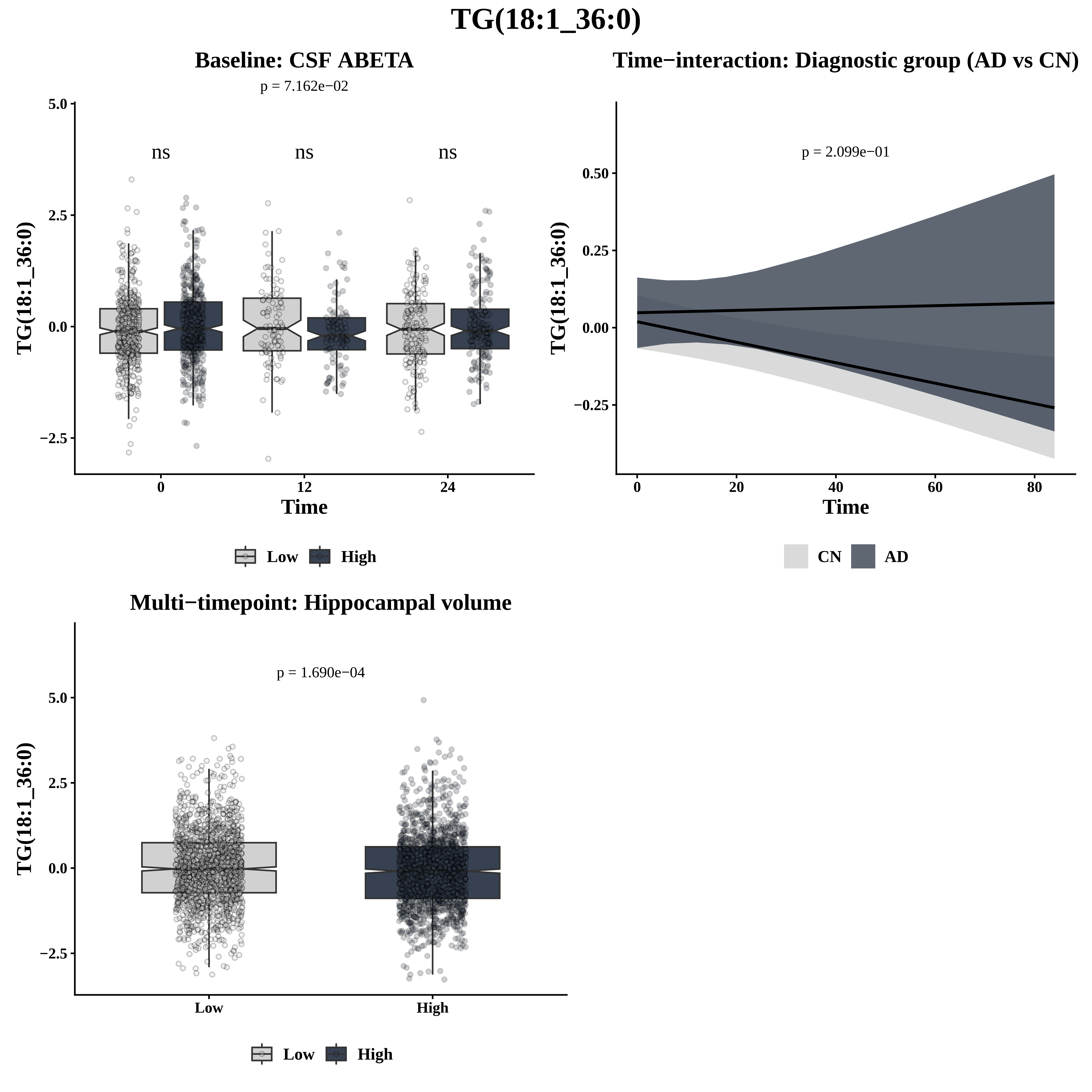
TG(18:1_36:0)
Baseline: CSF ABETA
p = 7.162e−02
ns
ns
ns
TG(18:1_36:0)
Time
Low
High
Time−interaction: Diagnostic group (AD vs CN)
p = 2.099e−01
TG(18:1_36:0)
Time
CN
AD
Multi−timepoint: Hippocampal volume
p = 1.690e−04
TG(18:1_36:0)
Low
High

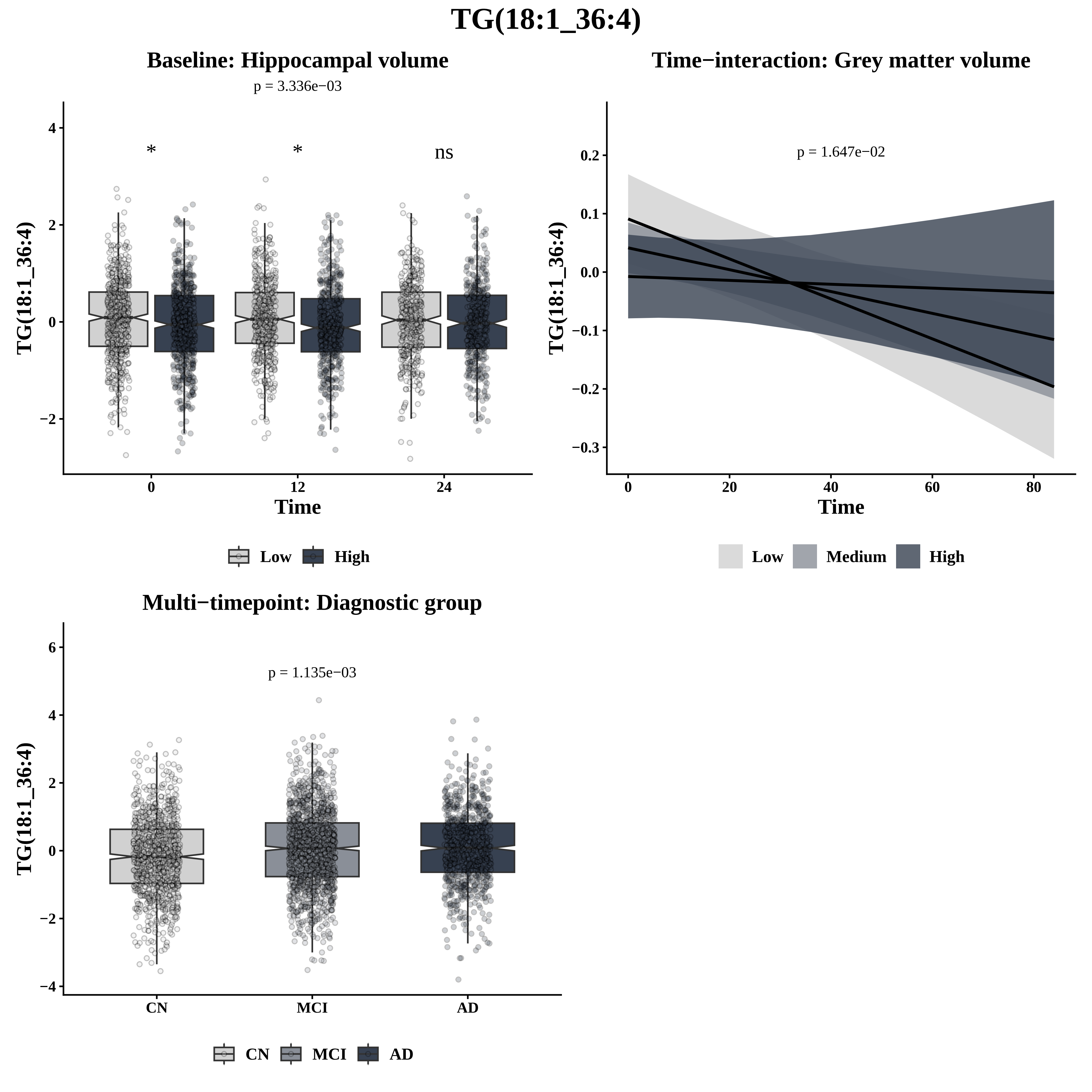

TG(18:1_36:4)
Baseline: Hippocampal volume
p = 3.336e−03
*
*
ns
TG(18:1_36:4)
Time
0
12
24
Low
High
Time−interaction: Grey matter volume
p = 1.647e−02
TG(18:1_36:4)
Time
Low
Medium
High
Multi−timepoint: Diagnostic group
p = 1.135e−03
TG(18:1_36:4)
CN
MCI
AD

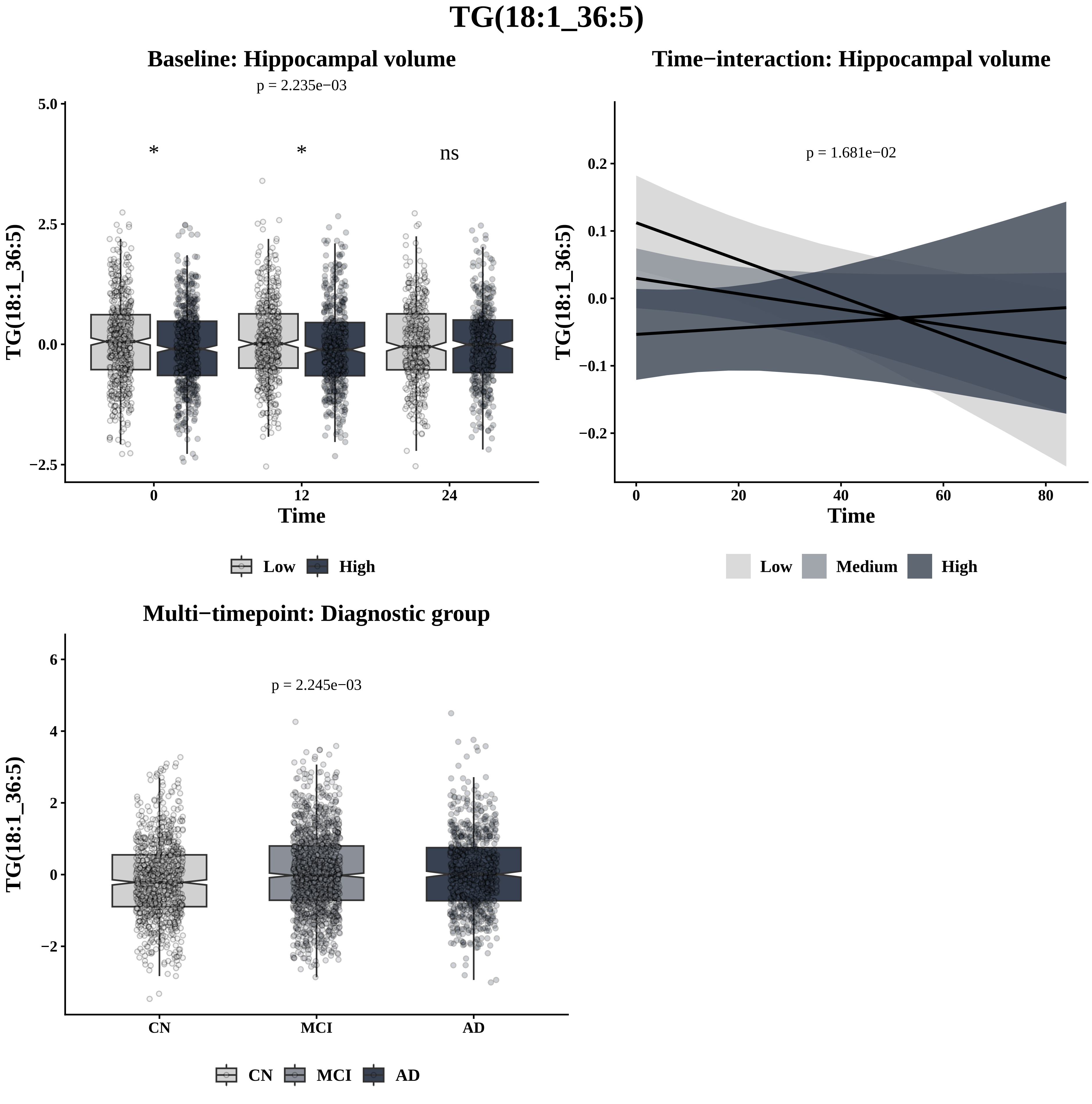

TG(18:1_36:5)
Baseline: Hippocampal volume
p = 2.235e−03
*
*
ns
5.0
2.5
0.0
−2.5
TG(18:1_36:5)
0
12
24
Time
Low
High
Time−interaction: Hippocampal volume
p = 1.681e−02
0.2
0.1
0.0
−0.1
−0.2
TG(18:1_36:5)
0
20
40
60
80
Time
Low
Medium
High
Multi−timepoint: Diagnostic group
p = 2.245e−03
6
4
2
0
−2
TG(18:1_36:5)
CN
MCI
AD
CN
MCI
AD

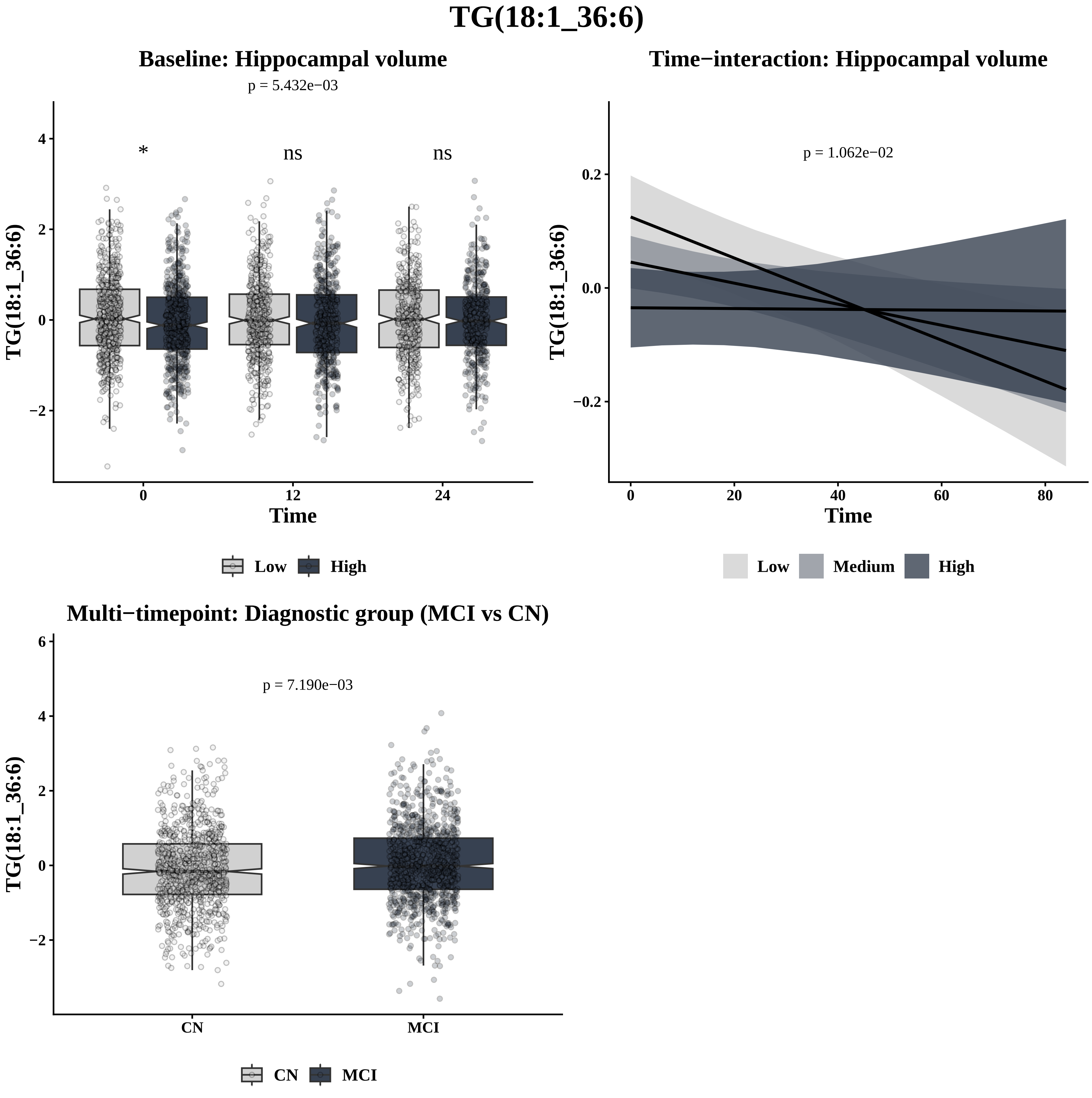

TG(18:1_36:6)
Baseline: Hippocampal volume
p = 5.432e−03
*
ns
ns
TG(18:1_36:6)
Time
0
12
24
Low
High
Time−interaction: Hippocampal volume
p = 1.062e−02
TG(18:1_36:6)
Time
Low
Medium
High
Multi−timepoint: Diagnostic group (MCI vs CN)
p = 7.190e−03
TG(18:1_36:6)
CN
MCI

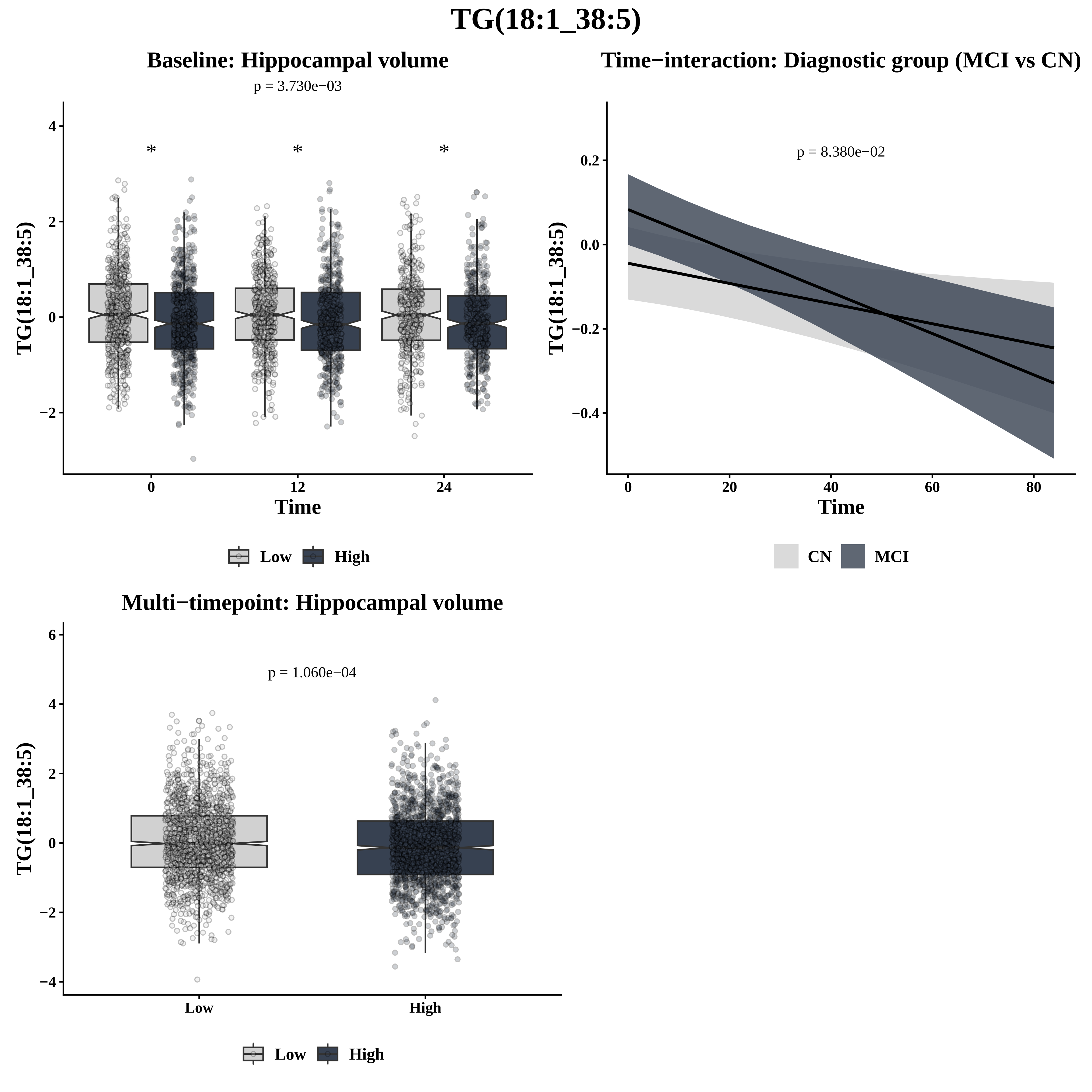

TG(18:1_38:5)
Baseline: Hippocampal volume
p = 3.730e−03
*
*
*
TG(18:1_38:5)
Time
0
12
24
Low
High
Time−interaction: Diagnostic group (MCI vs CN)
p = 8.380e−02
TG(18:1_38:5)
Time
CN
MCI
Multi−timepoint: Hippocampal volume
p = 1.060e−04
TG(18:1_38:5)
Low
High

# TG(18:2_28:0)

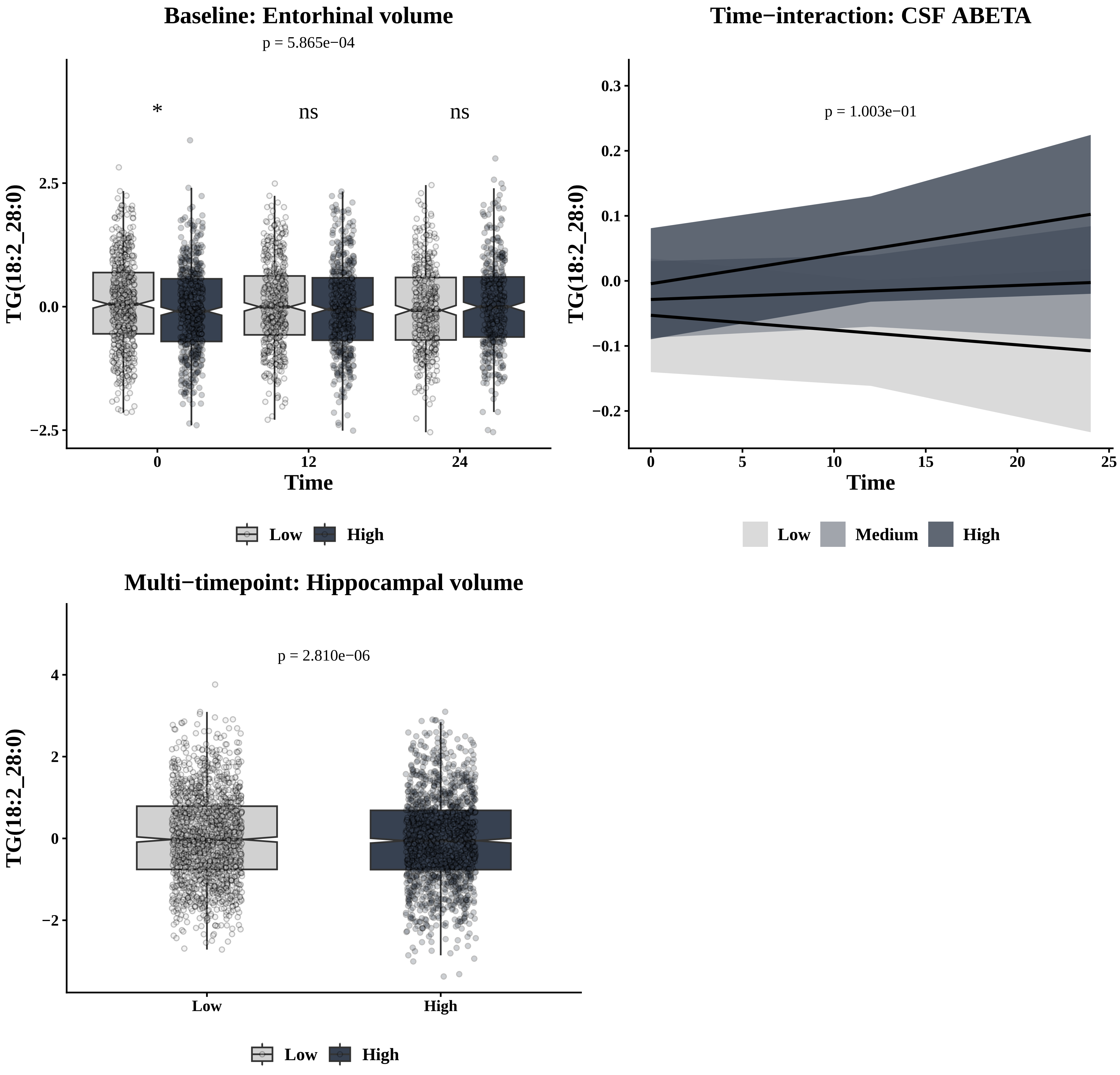

Baseline: Entorhinal volume
p = 5.865e−04
*
ns
ns
TG(18:2_28:0)
2.5
0.0
−2.5
0
12
24
Time
Low
High
Time−interaction: CSF ABETA
p = 1.003e−01
TG(18:2_28:0)
0.3
0.2
0.1
0.0
−0.1
−0.2
0
5
10
15
20
25
Time
Low
Medium
High
Multi−timepoint: Hippocampal volume
p = 2.810e−06
TG(18:2_28:0)
4
2
0
−2
Low
High
Low
High

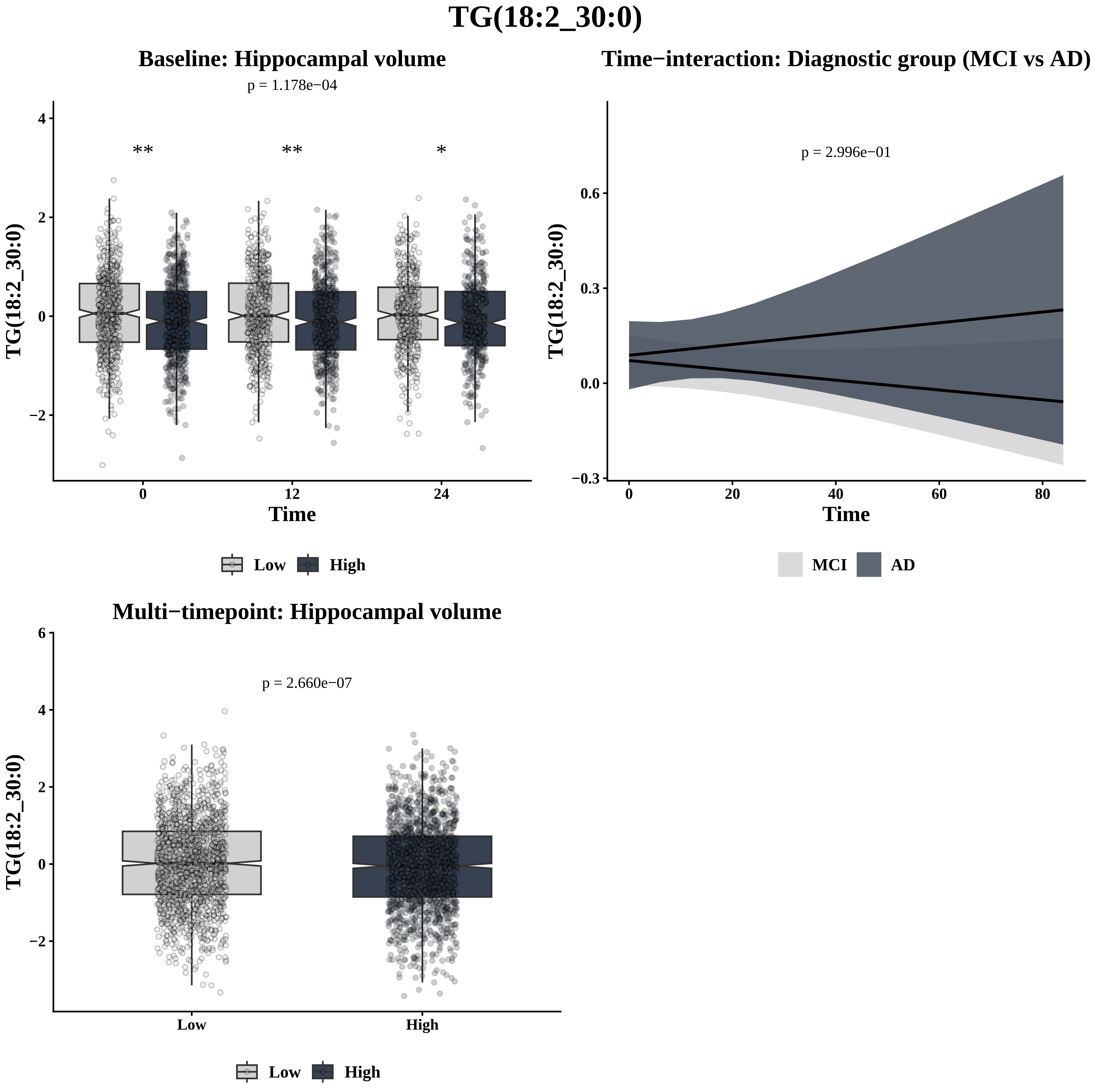
TG(18:2_30:0)
Baseline: Hippocampal volume
p = 1.178e−04
**
**
*
TG(18:2_30:0)
4
2
0
−2
0
12
24
Time
Low
High
Time−interaction: Diagnostic group (MCI vs AD)
p = 2.996e−01
TG(18:2_30:0)
0.6
0.3
0.0
−0.3
0
20
40
60
80
Time
MCI
AD
Multi−timepoint: Hippocampal volume
p = 2.660e−07
TG(18:2_30:0)
6
4
2
0
−2
Low
High
Low
High

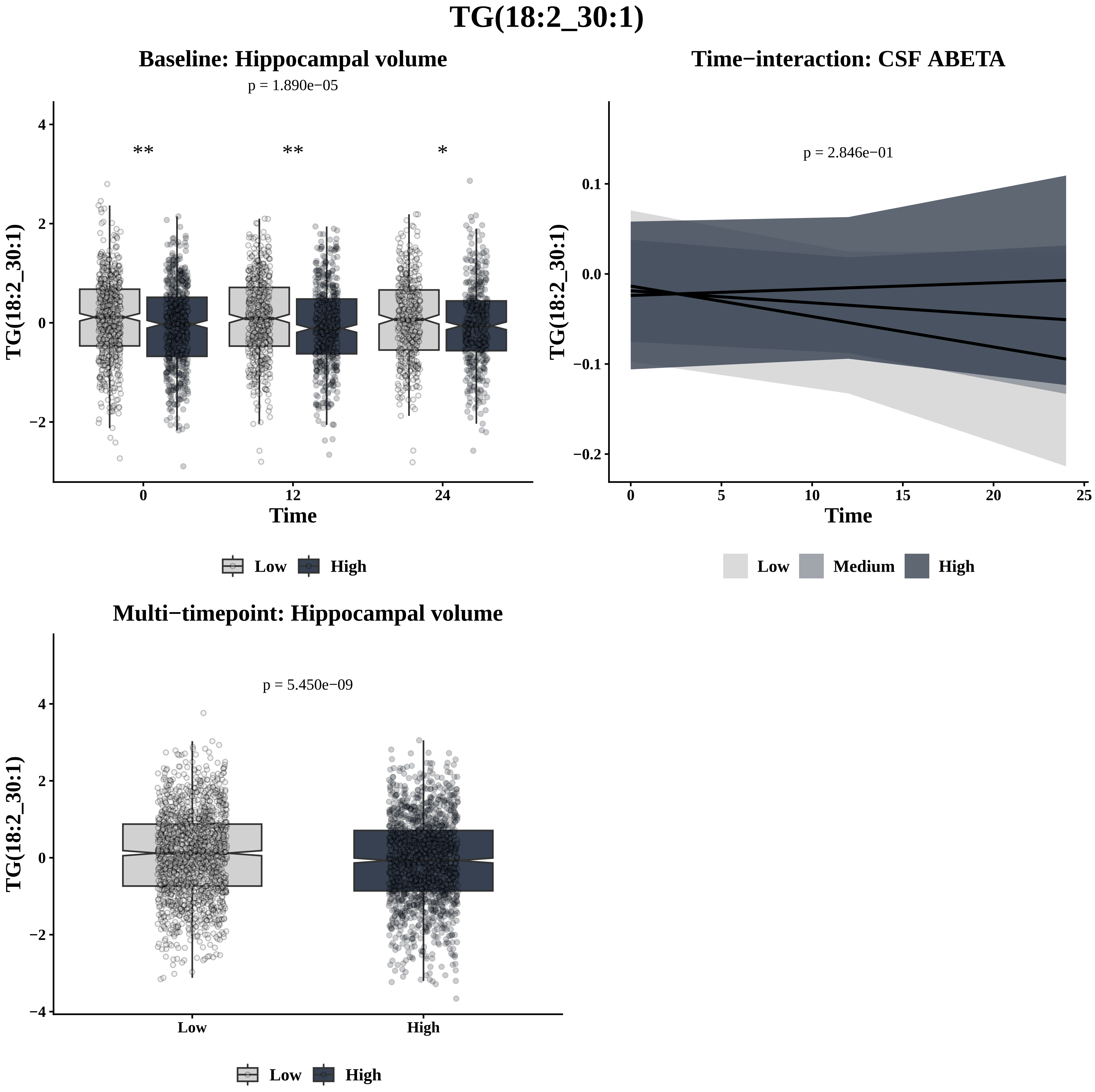

TG(18:2_30:1)
Baseline: Hippocampal volume
p = 1.890e−05
**
**
*
TG(18:2_30:1)
Time
Low
High
Time−interaction: CSF ABETA
p = 2.846e−01
TG(18:2_30:1)
Time
Low
Medium
High
Multi−timepoint: Hippocampal volume
p = 5.450e−09
TG(18:2_30:1)
Low
High
Low
High

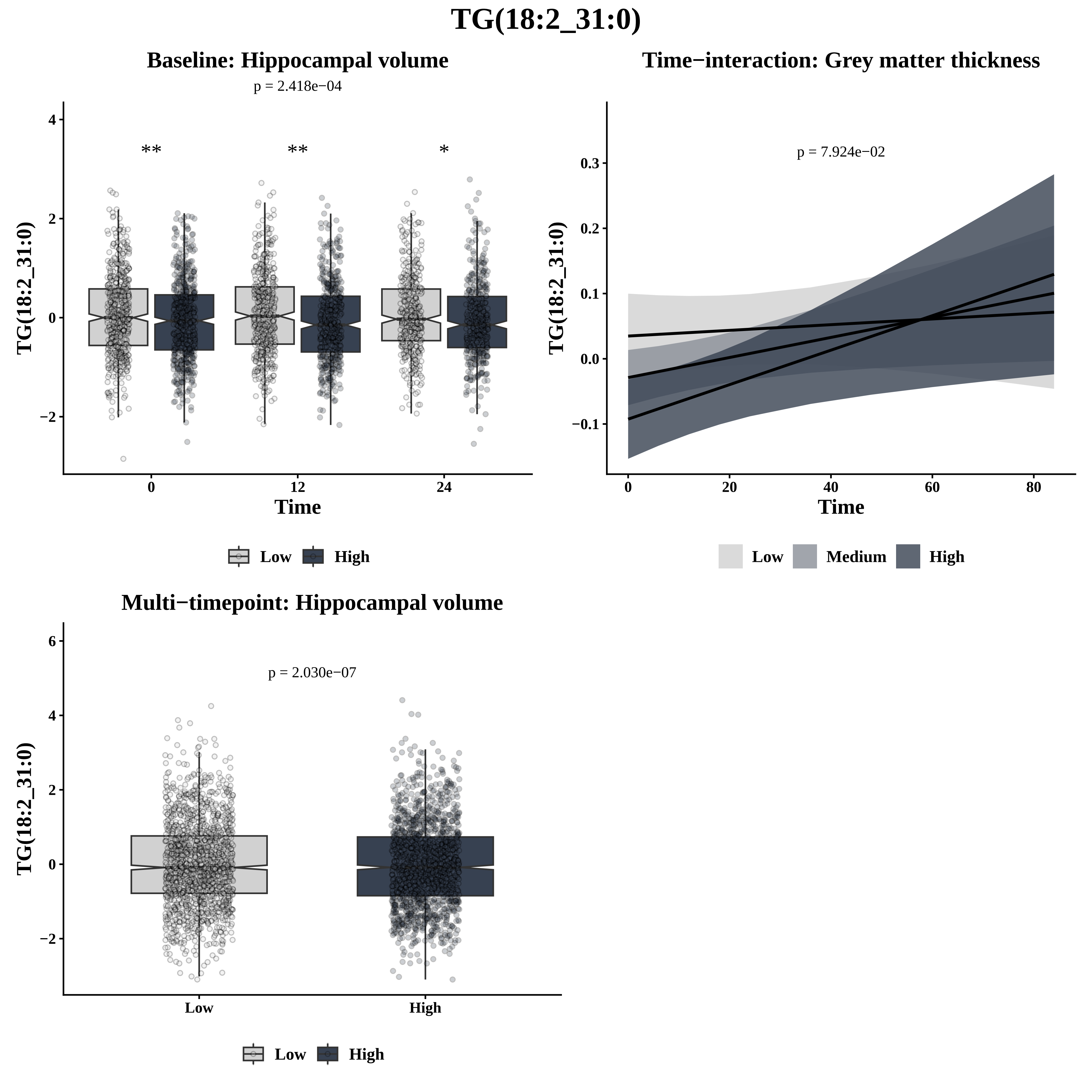

TG(18:2_31:0)
Baseline: Hippocampal volume
p = 2.418e−04
**
**
*
TG(18:2_31:0)
Time
Low
High
Time−interaction: Grey matter thickness
p = 7.924e−02
TG(18:2_31:0)
Time
Low
Medium
High
Multi−timepoint: Hippocampal volume
p = 2.030e−07
TG(18:2_31:0)
Low
High

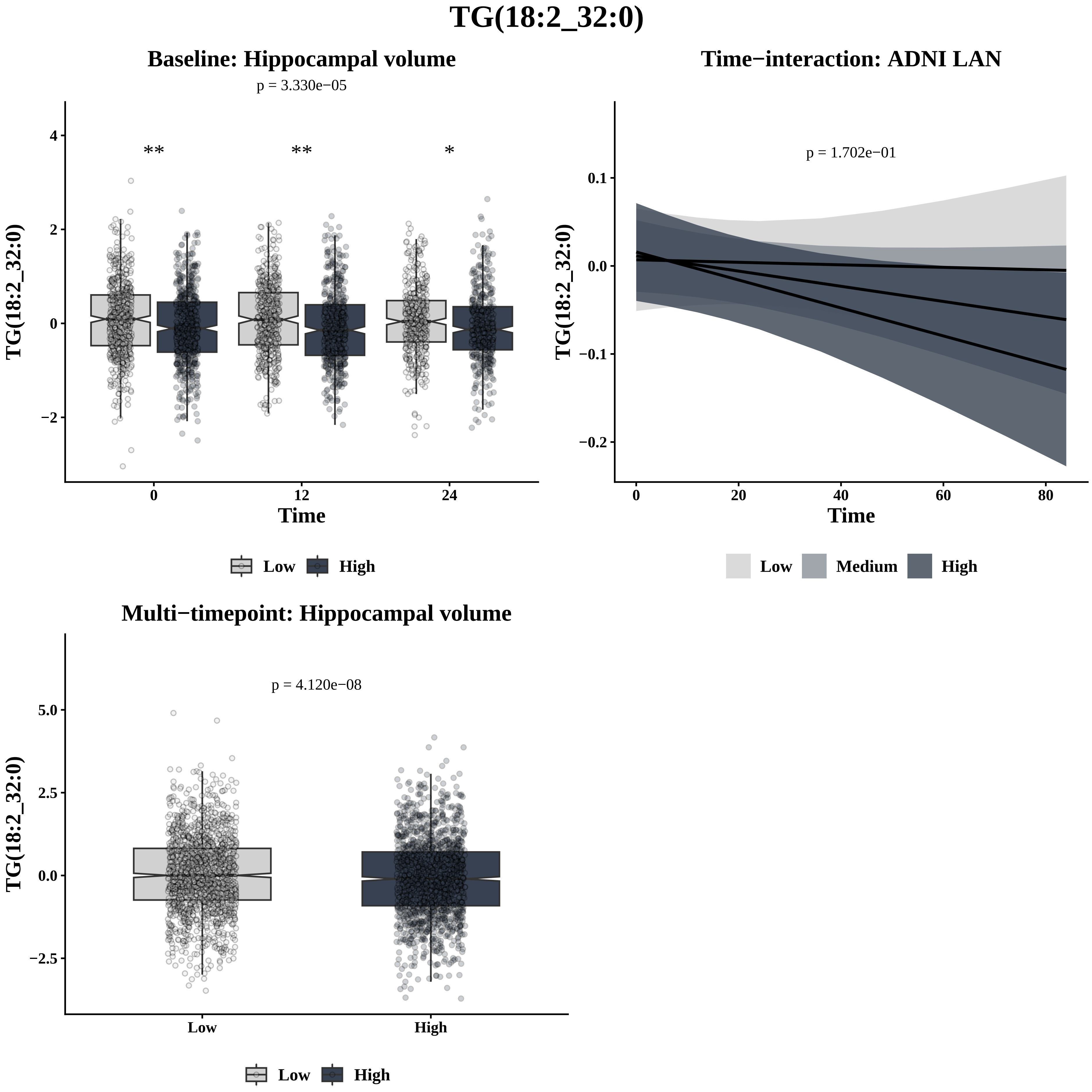

TG(18:2_32:0)
Baseline: Hippocampal volume
p = 3.330e−05
**
**
*
4
2
0
−2
0
12
24
Time
TG(18:2_32:0)
Low
High
Time−interaction: ADNI LAN
p = 1.702e−01
0.1
0.0
−0.1
−0.2
0
20
40
60
80
Time
TG(18:2_32:0)
Low
Medium
High
Multi−timepoint: Hippocampal volume
p = 4.120e−08
5.0
2.5
0.0
−2.5
Low
High
TG(18:2_32:0)
Low
High

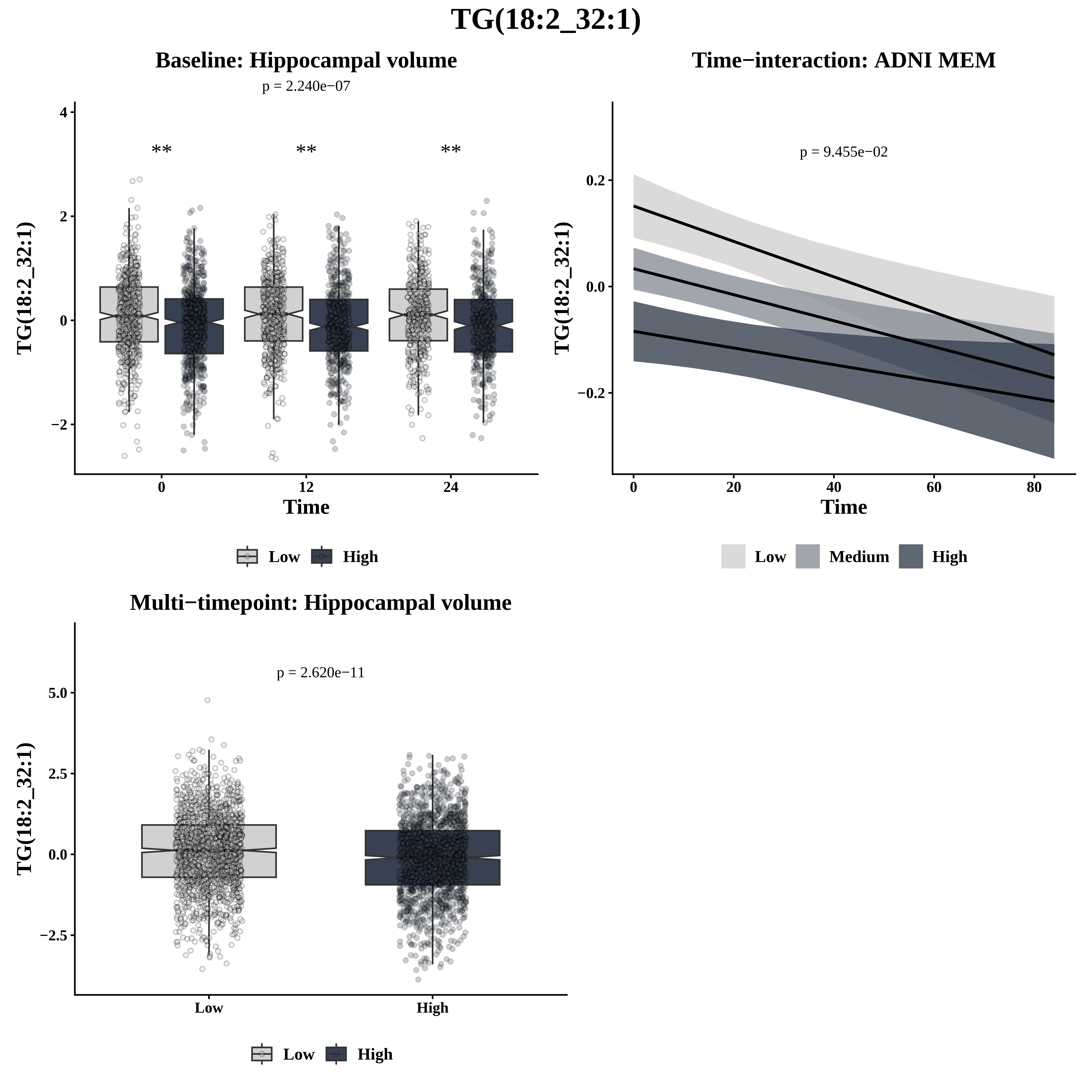

TG(18:2_32:1)
Baseline: Hippocampal volume
p = 2.240e−07
**
**
**
TG(18:2_32:1)
Time
4
2
0
−2
0
12
24
Low
High
Time−interaction: ADNI MEM
p = 9.455e−02
TG(18:2_32:1)
Time
0.2
0.0
−0.2
0
20
40
60
80
Low
Medium
High
Multi−timepoint: Hippocampal volume
p = 2.620e−11
TG(18:2_32:1)
5.0
2.5
0.0
−2.5
Low
High
Low
High

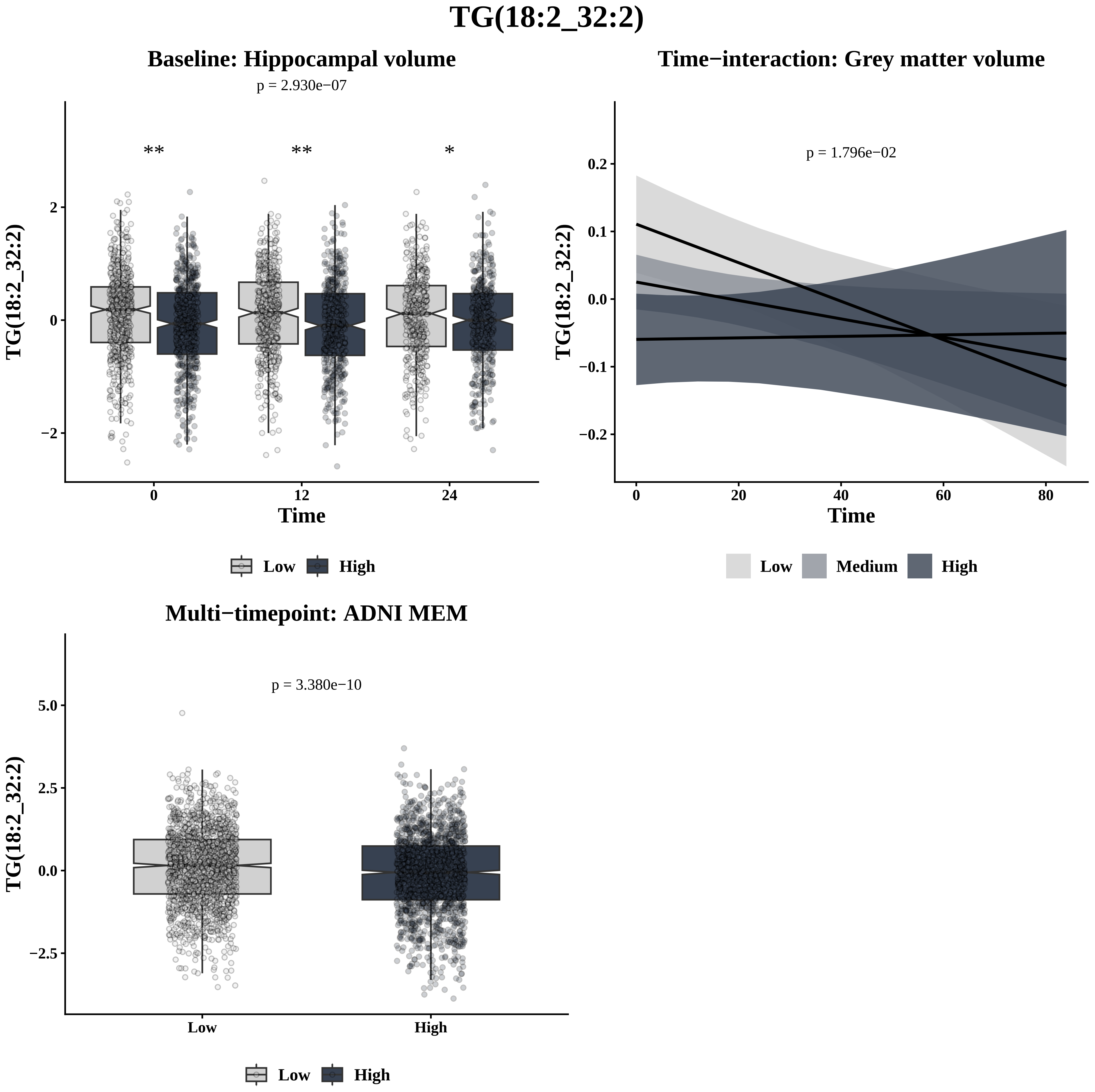

TG(18:2_32:2)
Baseline: Hippocampal volume
p = 2.930e−07
** ** *
TG(18:2_32:2)
2
0
−2
0 12 24
Time
Low High
Time−interaction: Grey matter volume
p = 1.796e−02
TG(18:2_32:2)
0.2
0.1
0.0
−0.1
−0.2
0 20 40 60 80
Time
Low Medium High
Multi−timepoint: ADNI MEM
p = 3.380e−10
TG(18:2_32:2)
5.0
2.5
0.0
−2.5
Low High
Low High

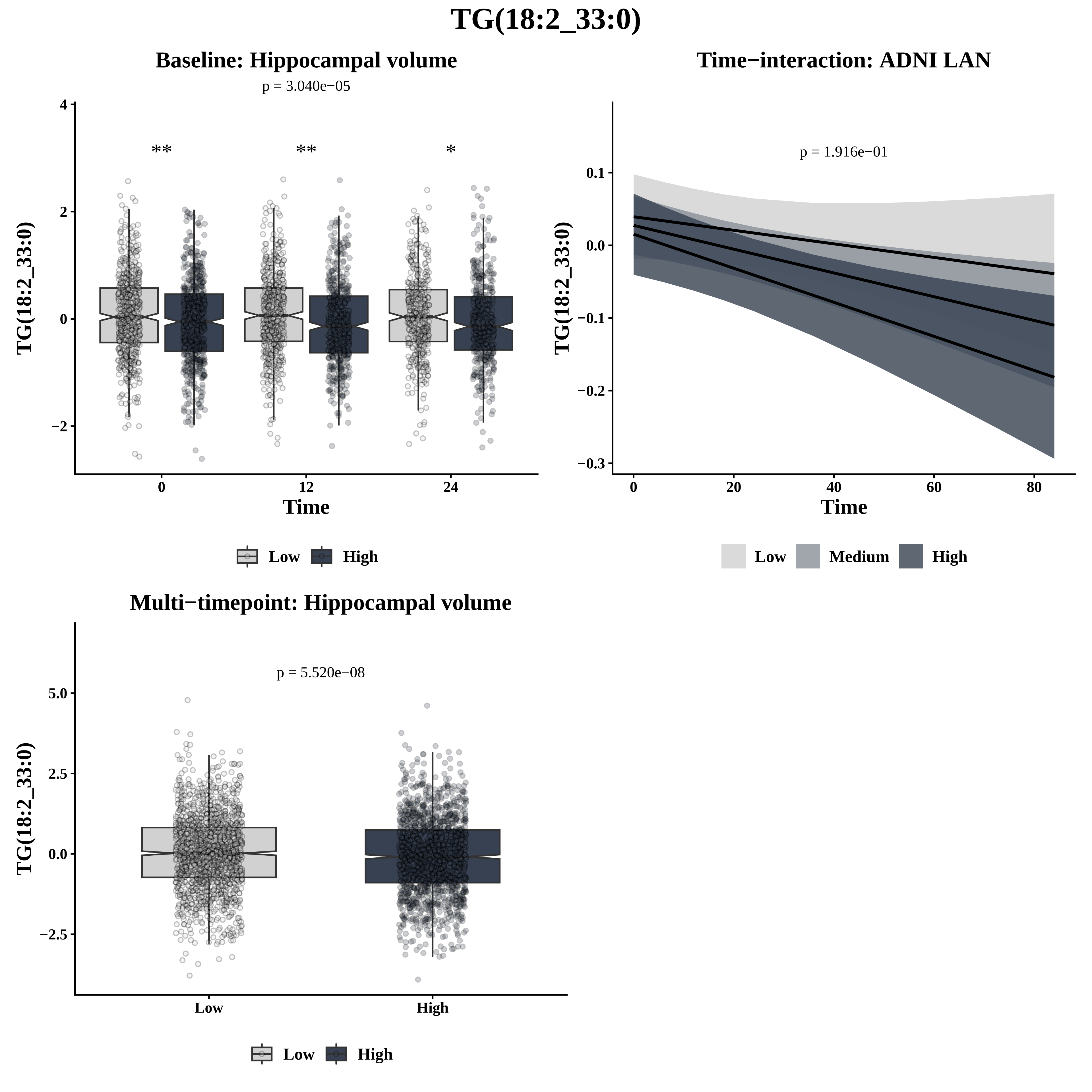

TG(18:2_33:0)
Baseline: Hippocampal volume
p = 3.040e−05
**
**
*
TG(18:2_33:0)
Time
0
12
24
4
2
0
−2
Low
High
Time−interaction: ADNI LAN
p = 1.916e−01
TG(18:2_33:0)
Time
0.1
0.0
−0.1
−0.2
−0.3
0
20
40
60
80
Low
Medium
High
Multi−timepoint: Hippocampal volume
p = 5.520e−08
TG(18:2_33:0)
5.0
2.5
0.0
−2.5
Low
High
Low
High

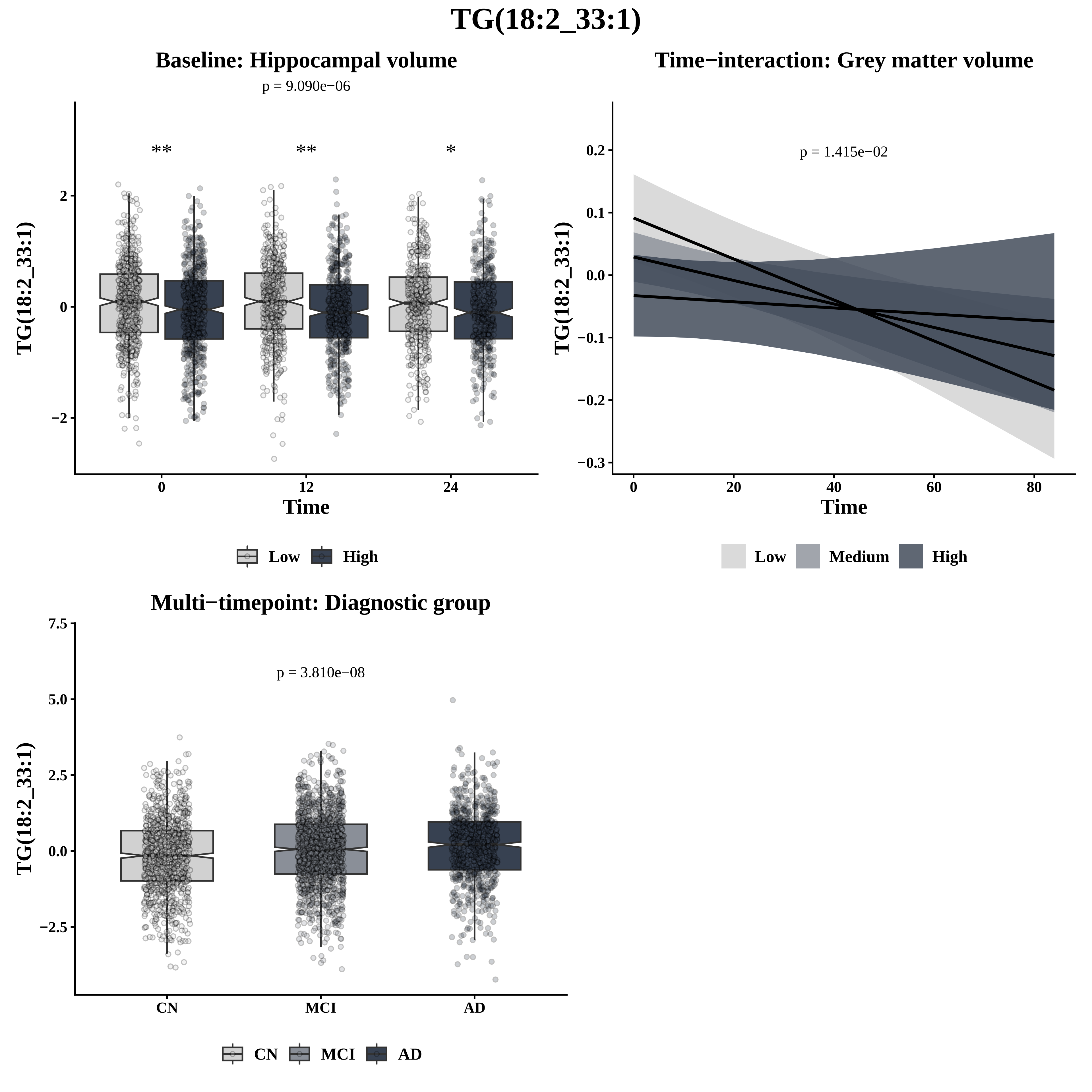
TG(18:2_33:1)
Baseline: Hippocampal volume
p = 9.090e−06
**
**
*
2
0
−2
TG(18:2_33:1)
0
12
24
Time
Low
High
Time−interaction: Grey matter volume
p = 1.415e−02
0.2
0.1
0.0
−0.1
−0.2
−0.3
TG(18:2_33:1)
0
20
40
60
80
Time
Low
Medium
High
Multi−timepoint: Diagnostic group
p = 3.810e−08
7.5
5.0
2.5
0.0
−2.5
TG(18:2_33:1)
CN
MCI
AD
CN
MCI
AD

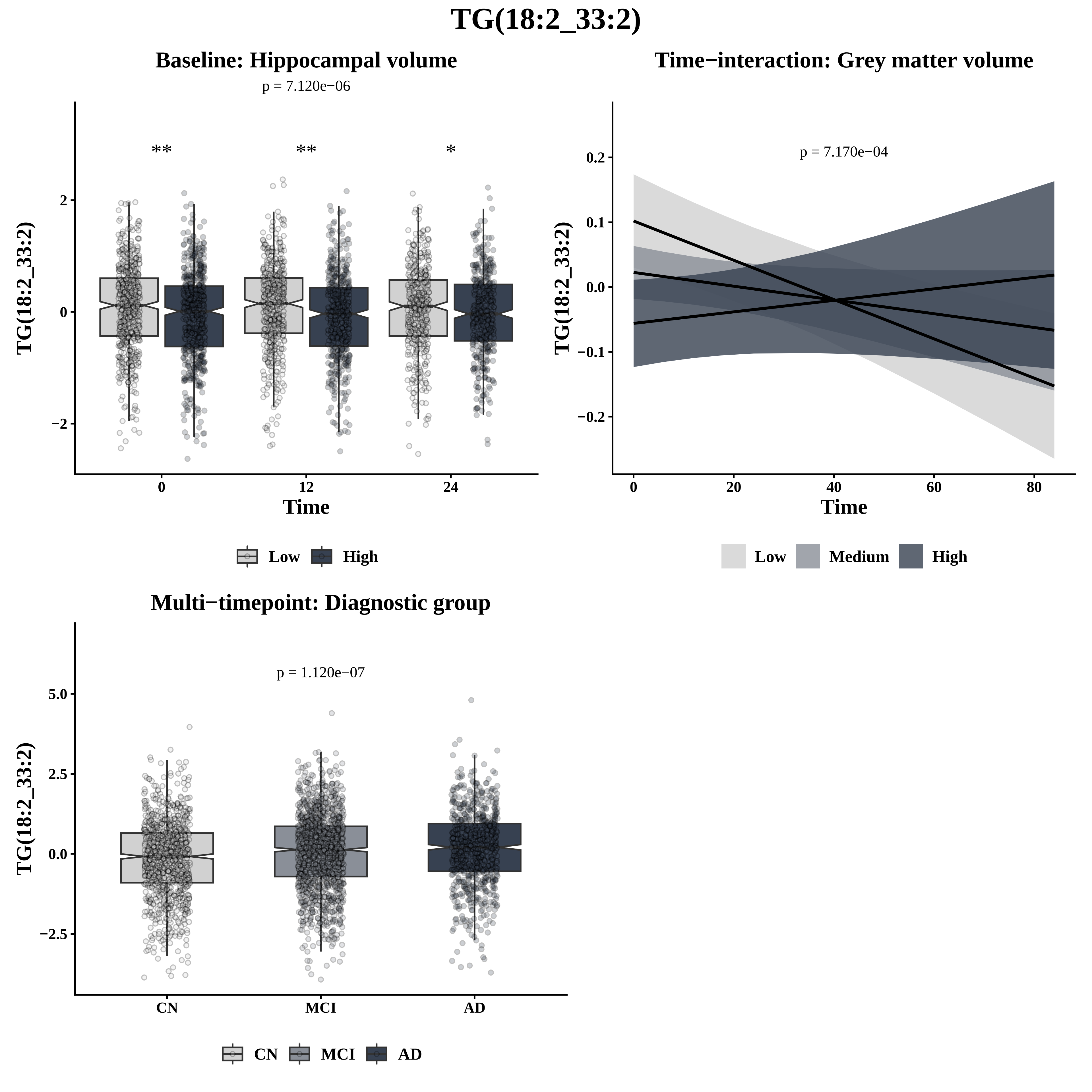
TG(18:2_33:2)
Baseline: Hippocampal volume
p = 7.120e−06
**
**
*
TG(18:2_33:2)
Time
Low
High
Time−interaction: Grey matter volume
p = 7.170e−04
TG(18:2_33:2)
Time
Low
Medium
High
Multi−timepoint: Diagnostic group
p = 1.120e−07
TG(18:2_33:2)
CN
MCI
AD

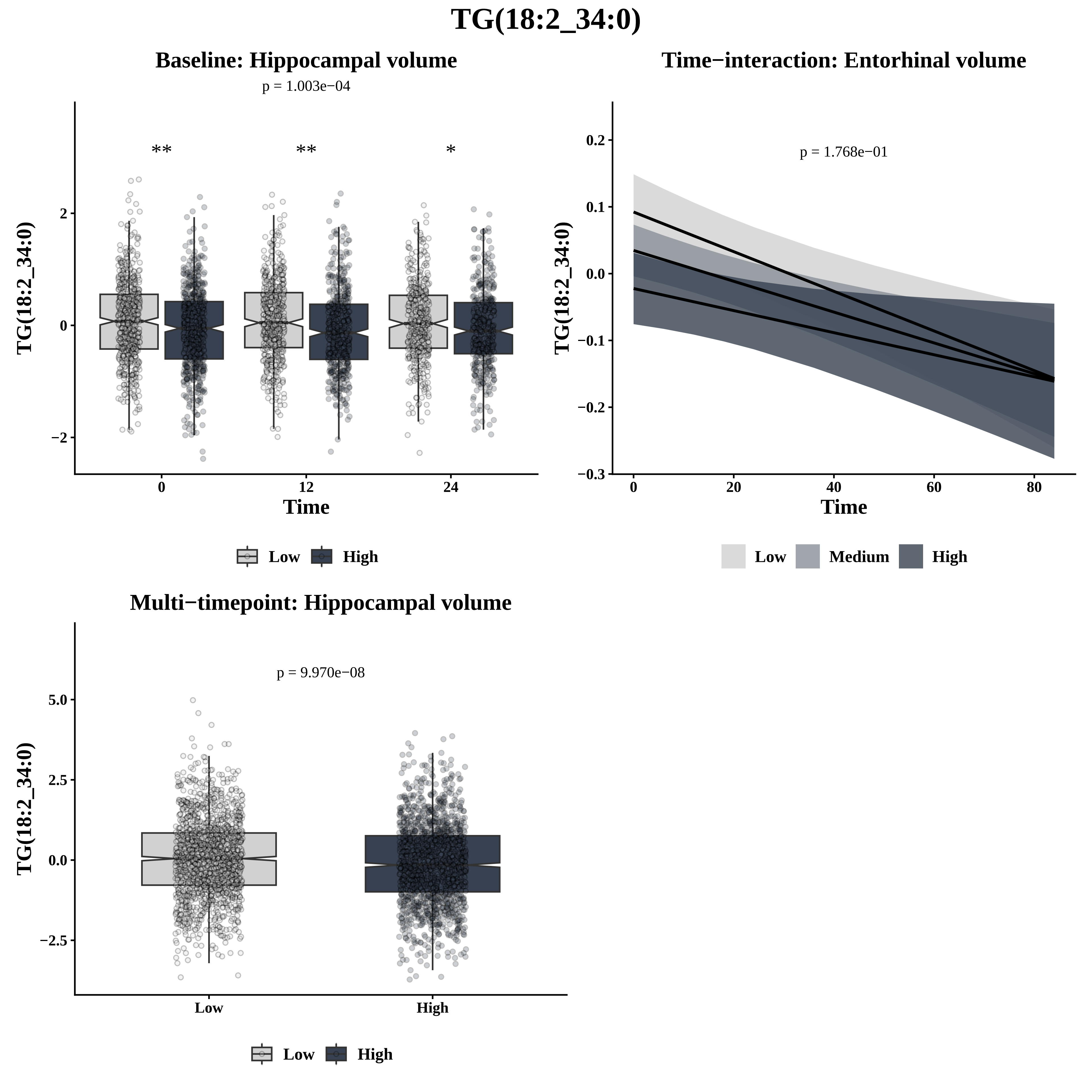

TG(18:2_34:0)
Baseline: Hippocampal volume
p = 1.003e−04
**
**
*
TG(18:2_34:0)
2
0
−2
0
12
24
Time
Low
High
Time−interaction: Entorhinal volume
p = 1.768e−01
TG(18:2_34:0)
0.2
0.1
0.0
−0.1
−0.2
−0.3
0
20
40
60
80
Time
Low
Medium
High
Multi−timepoint: Hippocampal volume
p = 9.970e−08
TG(18:2_34:0)
5.0
2.5
0.0
−2.5
Low
High
Low
High

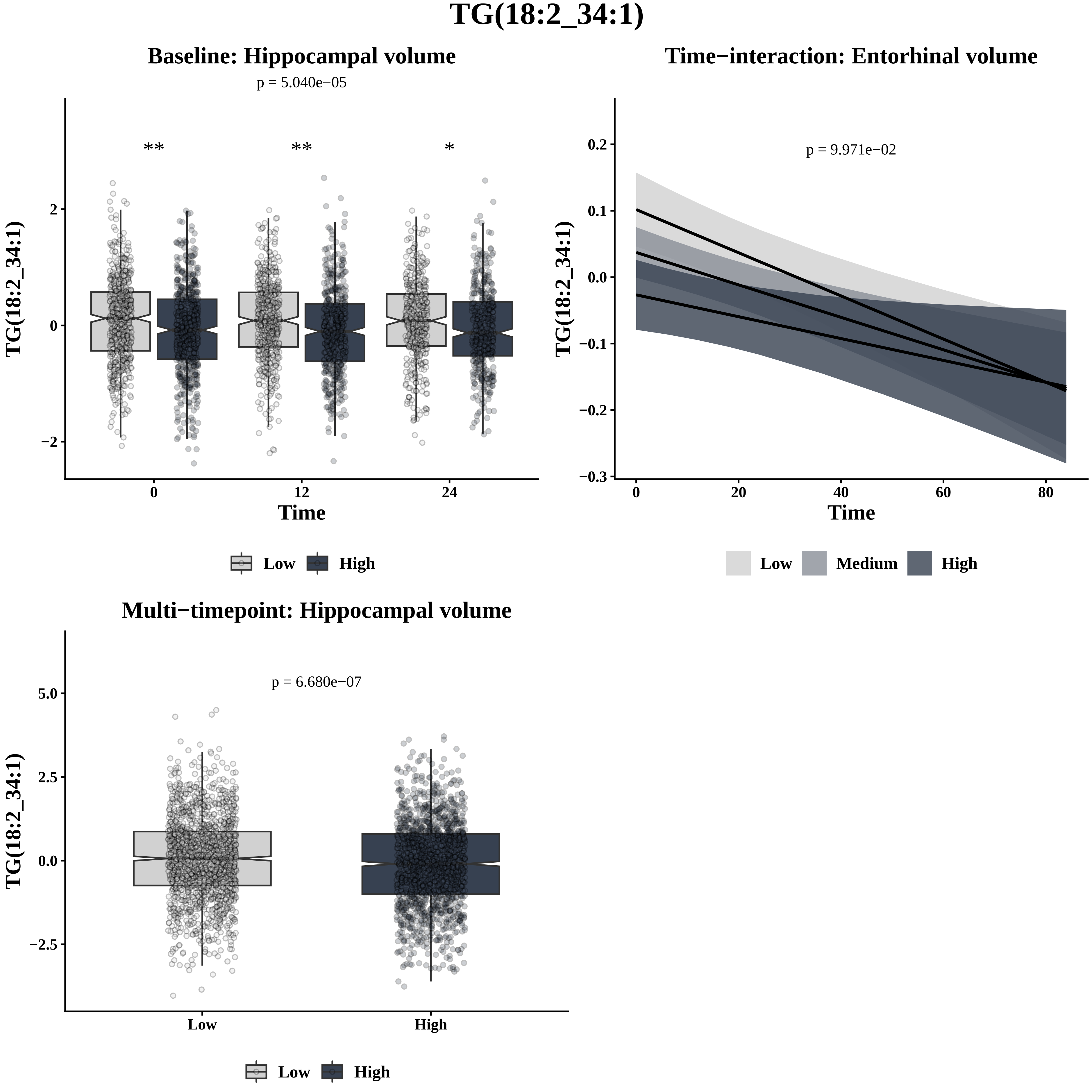
TG(18:2_34:1)
Baseline: Hippocampal volume
p = 5.040e−05
**
**
*
TG(18:2_34:1)
2
0
−2
0
12
24
Time
Low
High
Time−interaction: Entorhinal volume
p = 9.971e−02
TG(18:2_34:1)
0.2
0.1
0.0
−0.1
−0.2
−0.3
0
20
40
60
80
Time
Low
Medium
High
Multi−timepoint: Hippocampal volume
p = 6.680e−07
TG(18:2_34:1)
5.0
2.5
0.0
−2.5
Low
High
Low
High

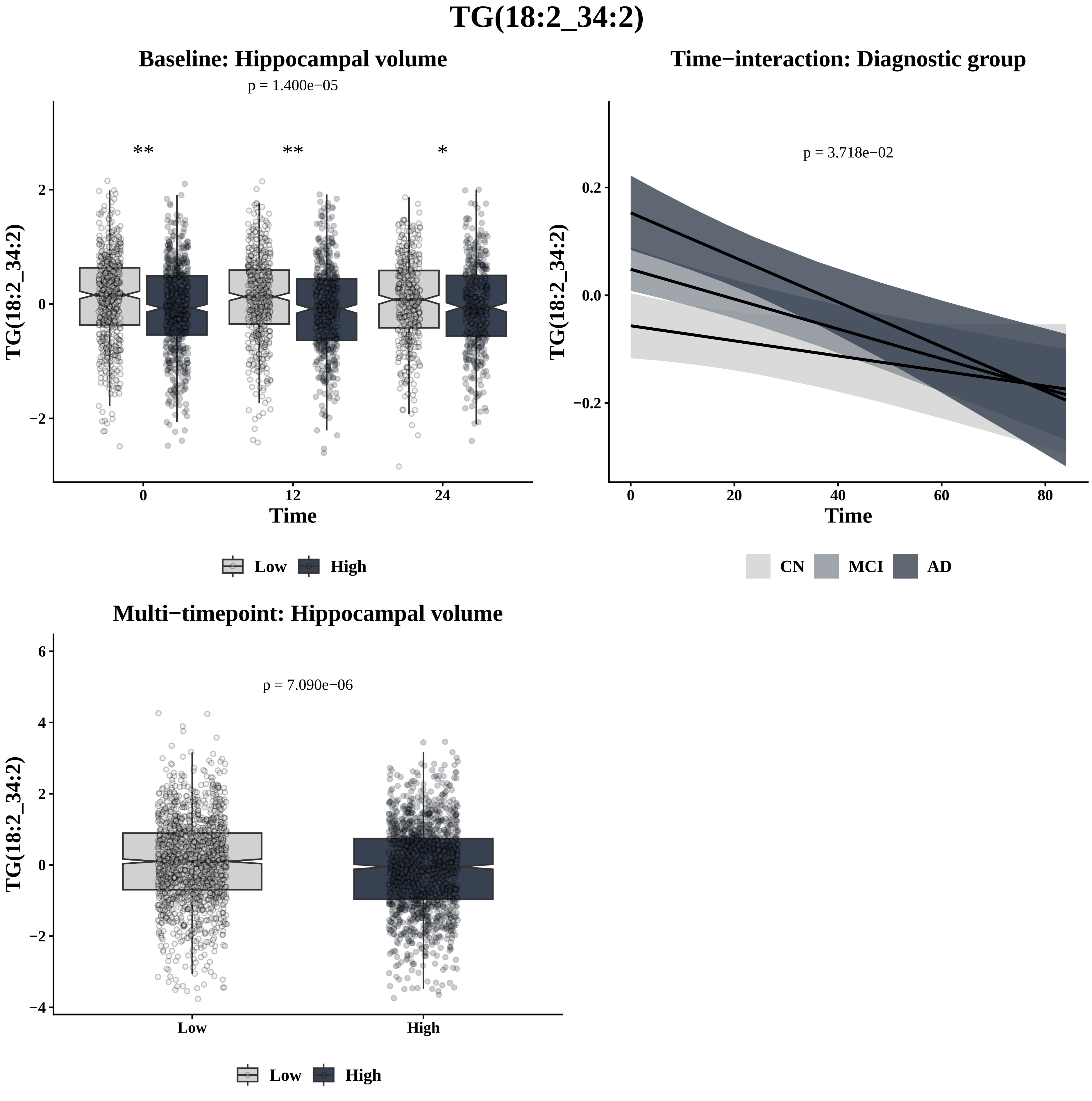
TG(18:2_34:2)
Baseline: Hippocampal volume
p = 1.400e−05
**
**
*
TG(18:2_34:2)
Time
0
12
24
Low
High
Time−interaction: Diagnostic group
p = 3.718e−02
TG(18:2_34:2)
Time
CN
MCI
AD
Multi−timepoint: Hippocampal volume
p = 7.090e−06
TG(18:2_34:2)
Low
High

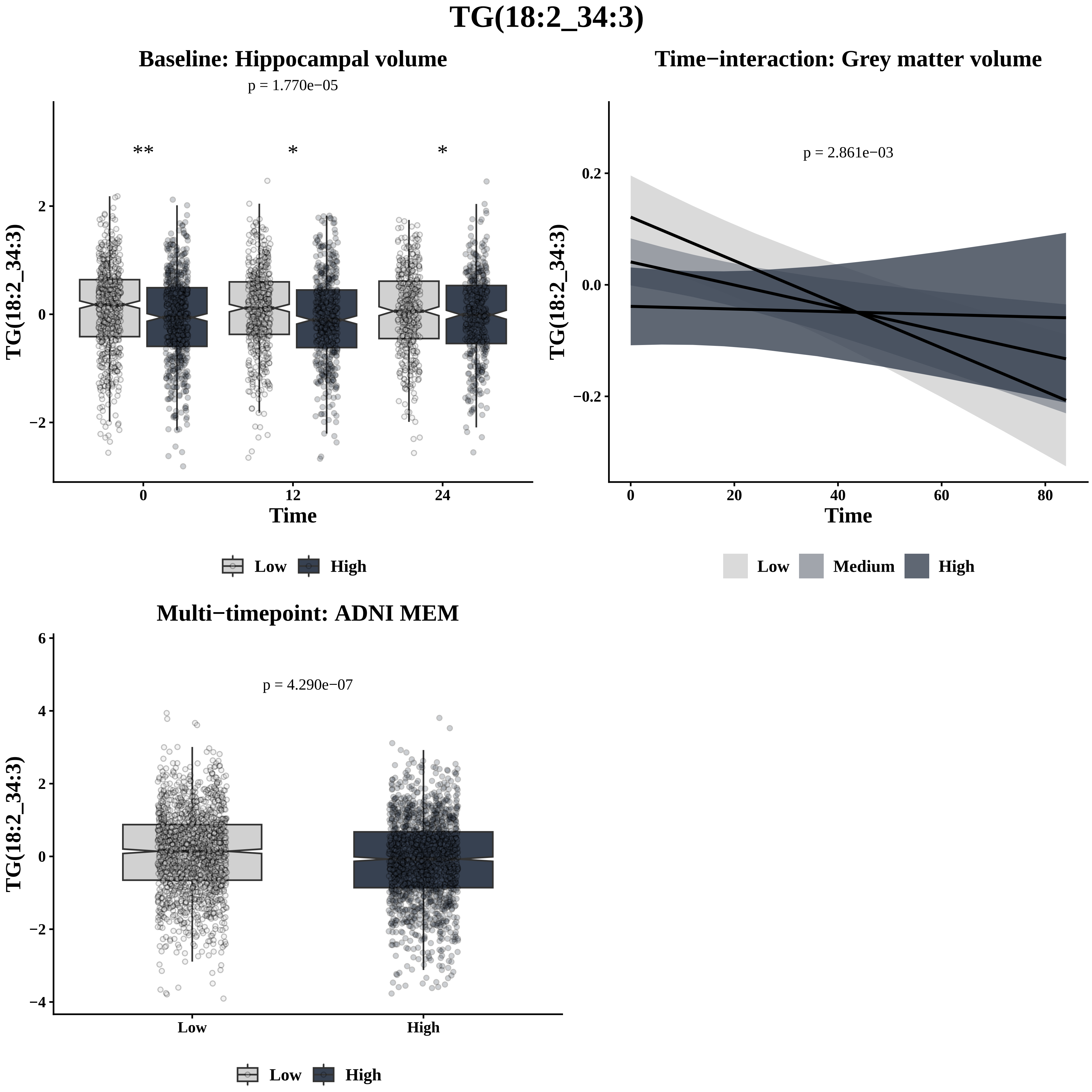
TG(18:2_34:3)
Baseline: Hippocampal volume
p = 1.770e−05
**
*
*
TG(18:2_34:3)
2
0
−2
0
12
24
Time
Low
High
Time−interaction: Grey matter volume
p = 2.861e−03
TG(18:2_34:3)
0.2
0.0
−0.2
0
20
40
60
80
Time
Low
Medium
High
Multi−timepoint: ADNI MEM
p = 4.290e−07
TG(18:2_34:3)
6
4
2
0
−2
−4
Low
High
Low
High

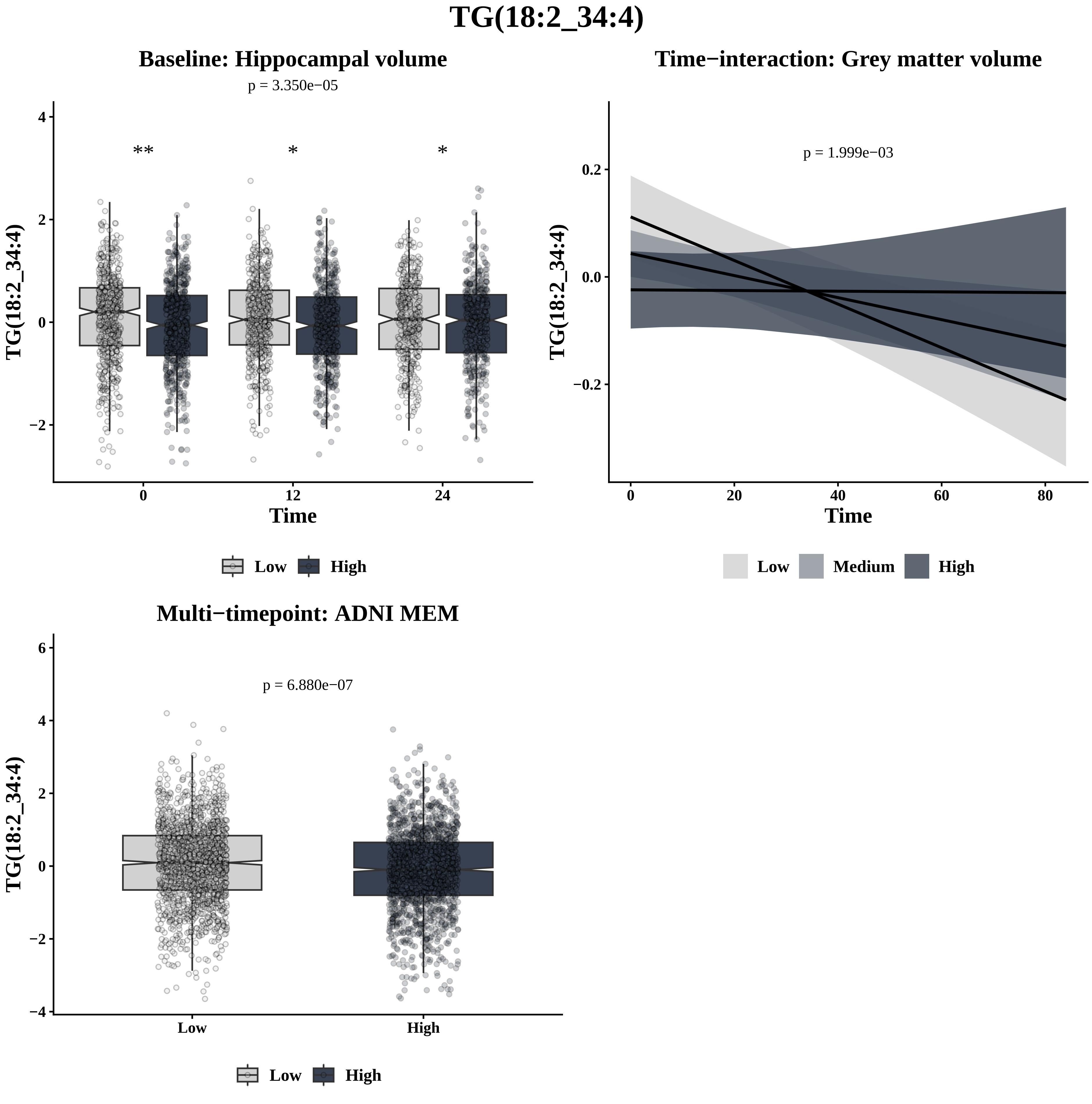
TG(18:2_34:4)
Baseline: Hippocampal volume
p = 3.350e−05
**
*
*
TG(18:2_34:4)
4
2
0
−2
0
12
24
Time
Low
High
Time−interaction: Grey matter volume
p = 1.999e−03
TG(18:2_34:4)
0.2
0.0
−0.2
0
20
40
60
80
Time
Low
Medium
High
Multi−timepoint: ADNI MEM
p = 6.880e−07
TG(18:2_34:4)
6
4
2
0
−2
−4
Low
High
Low
High

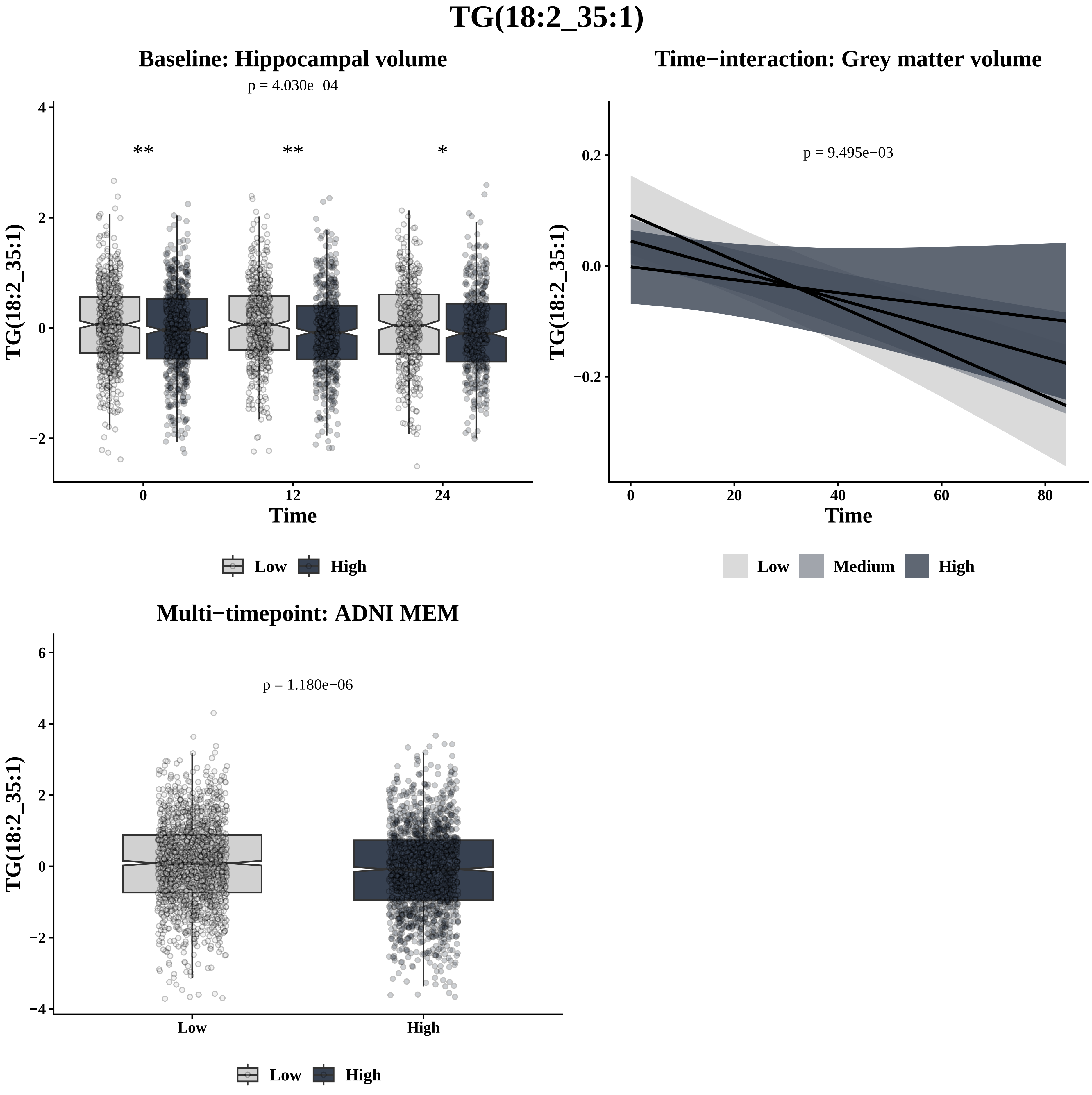

TG(18:2_35:1)
Baseline: Hippocampal volume
p = 4.030e−04
**
**
*
TG(18:2_35:1)
Time
0
12
24
Low
High
Time−interaction: Grey matter volume
p = 9.495e−03
TG(18:2_35:1)
Time
0
20
40
60
80
Low
Medium
High
Multi−timepoint: ADNI MEM
p = 1.180e−06
TG(18:2_35:1)
Low
High
Low
High

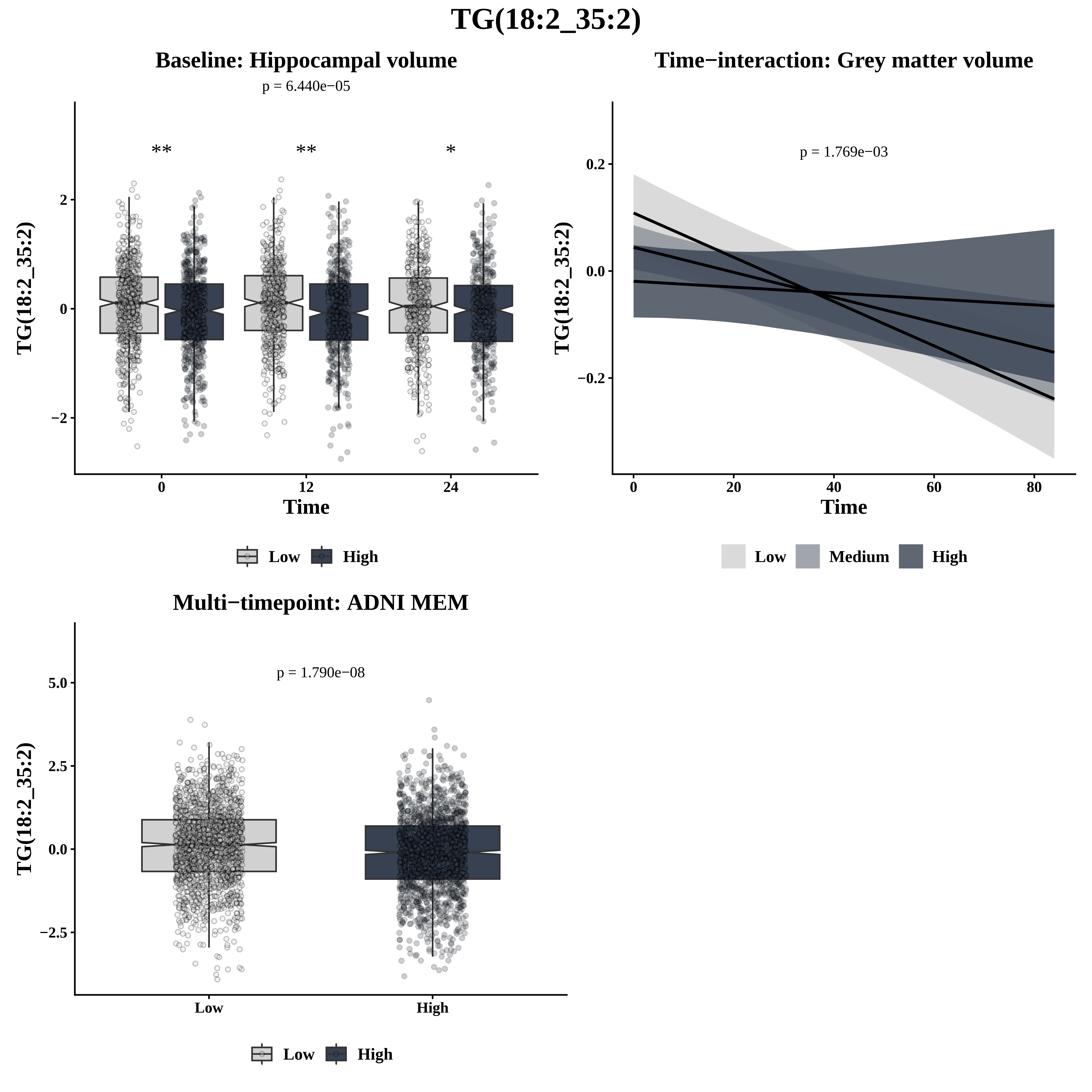
TG(18:2_35:2)
Baseline: Hippocampal volume
p = 6.440e−05
**
**
*
2
0
−2
TG(18:2_35:2)
0
12
24
Time
Low
High
Time−interaction: Grey matter volume
p = 1.769e−03
0.2
0.0
−0.2
TG(18:2_35:2)
0
20
40
60
80
Time
Low
Medium
High
Multi−timepoint: ADNI MEM
p = 1.790e−08
5.0
2.5
0.0
−2.5
TG(18:2_35:2)
Low
High
Low
High

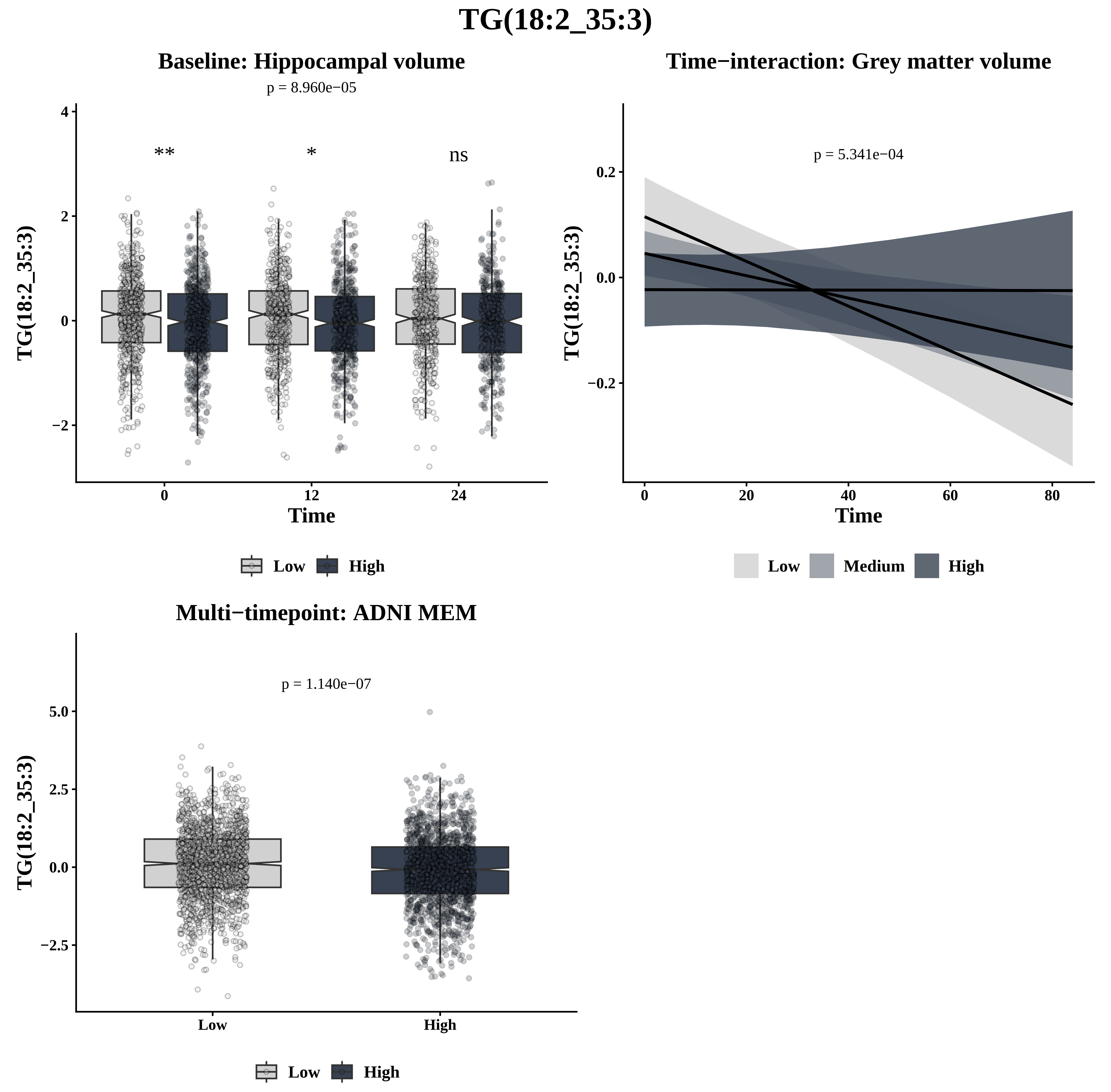

TG(18:2_35:3)
Baseline: Hippocampal volume
p = 8.960e−05
**
*
ns
TG(18:2_35:3)
Time
0
12
24
Low
High
Time−interaction: Grey matter volume
p = 5.341e−04
TG(18:2_35:3)
Time
Low
Medium
High
Multi−timepoint: ADNI MEM
p = 1.140e−07
TG(18:2_35:3)
Low
High

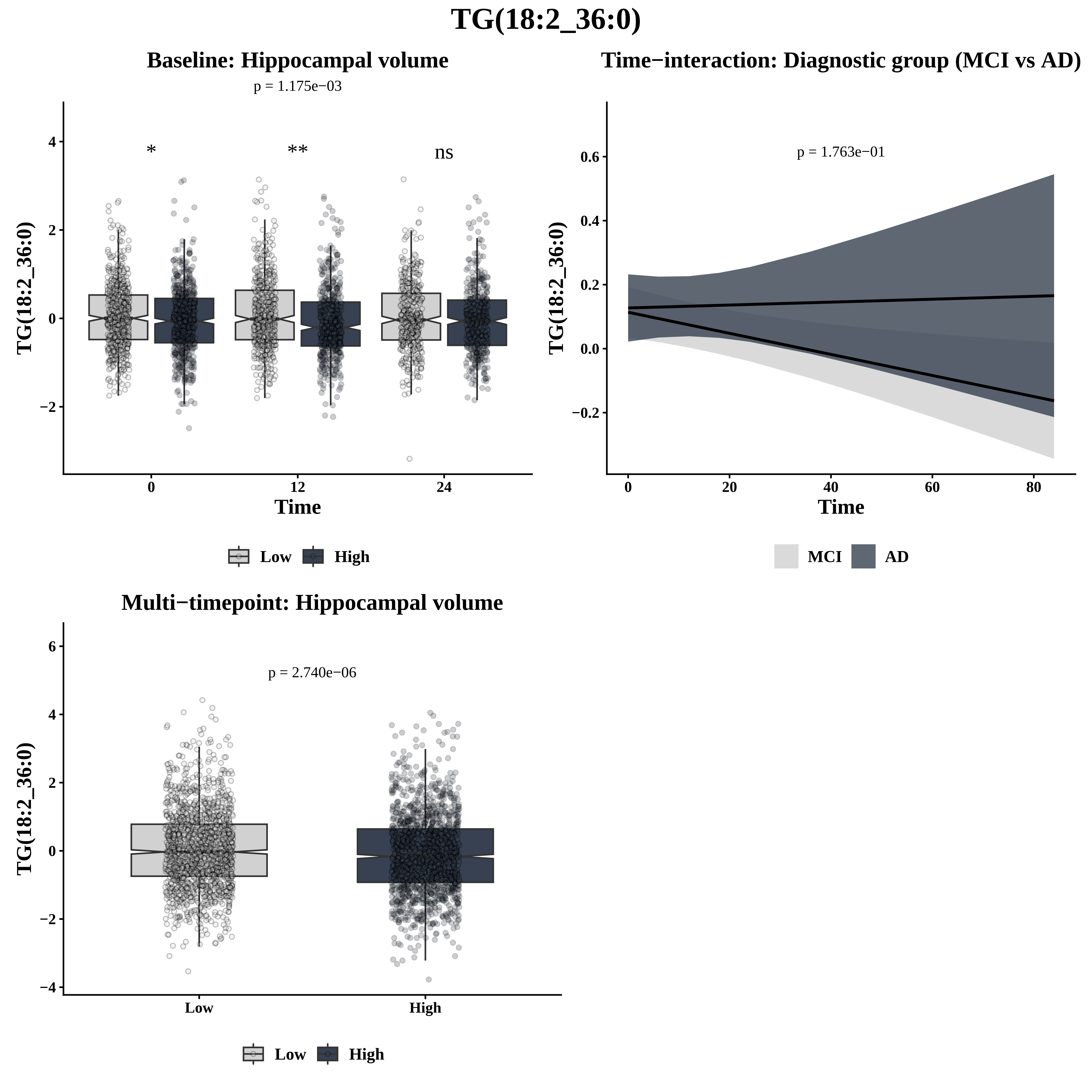

TG(18:2_36:0)
Baseline: Hippocampal volume
p = 1.175e−03
*
**
ns
4
2
0
−2
TG(18:2_36:0)
0
12
24
Time
Low
High
Time−interaction: Diagnostic group (MCI vs AD)
p = 1.763e−01
0.6
0.4
0.2
0.0
−0.2
TG(18:2_36:0)
0
20
40
60
80
Time
MCI
AD
Multi−timepoint: Hippocampal volume
p = 2.740e−06
6
4
2
0
−2
−4
TG(18:2_36:0)
Low
High
Low
High

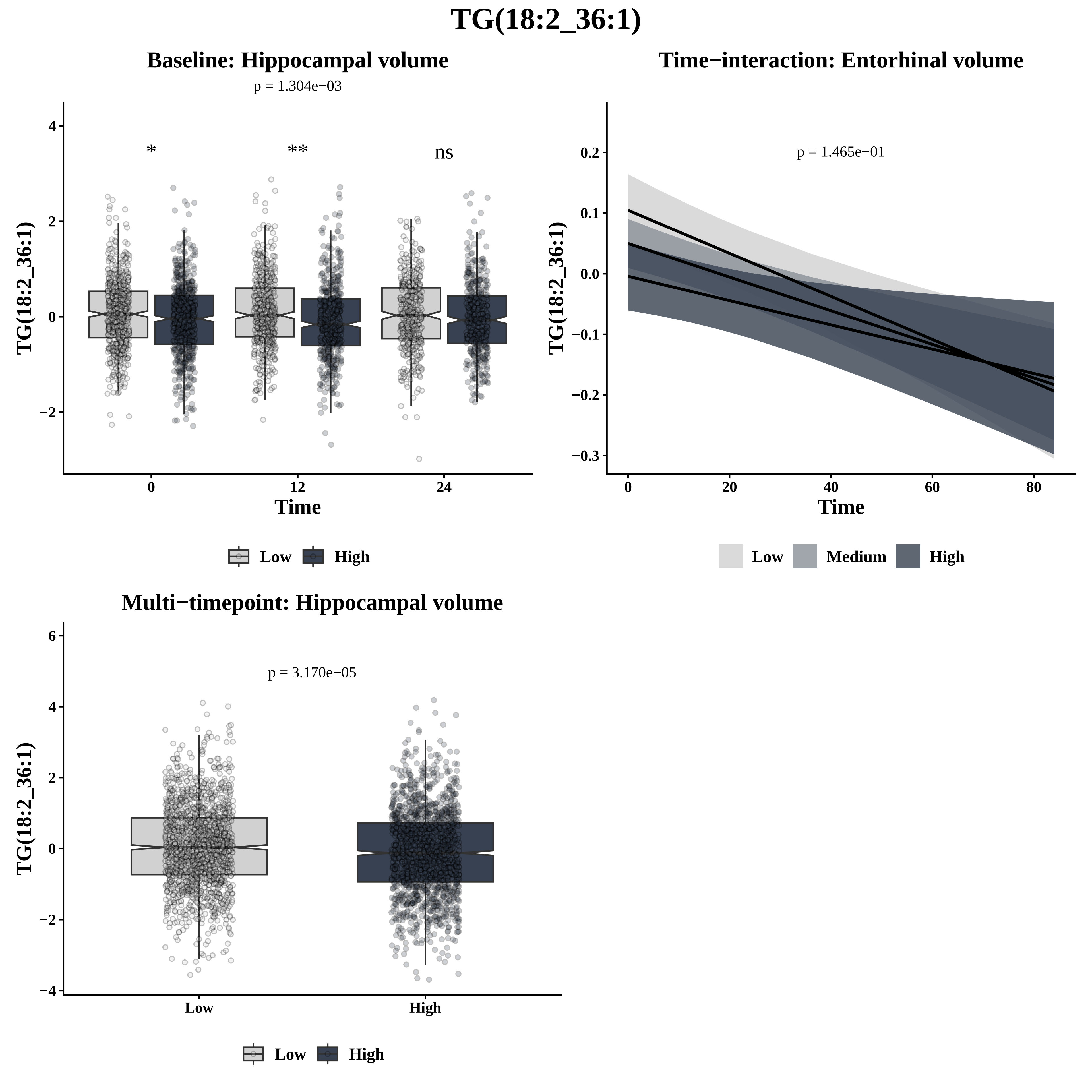

TG(18:2_36:1)
Baseline: Hippocampal volume
p = 1.304e−03
*
**
ns
TG(18:2_36:1)
4
2
0
−2
0
12
24
Time
Low
High
Time−interaction: Entorhinal volume
p = 1.465e−01
TG(18:2_36:1)
0.2
0.1
0.0
−0.1
−0.2
−0.3
0
20
40
60
80
Time
Low
Medium
High
Multi−timepoint: Hippocampal volume
p = 3.170e−05
TG(18:2_36:1)
6
4
2
0
−2
−4
Low
High
Low
High

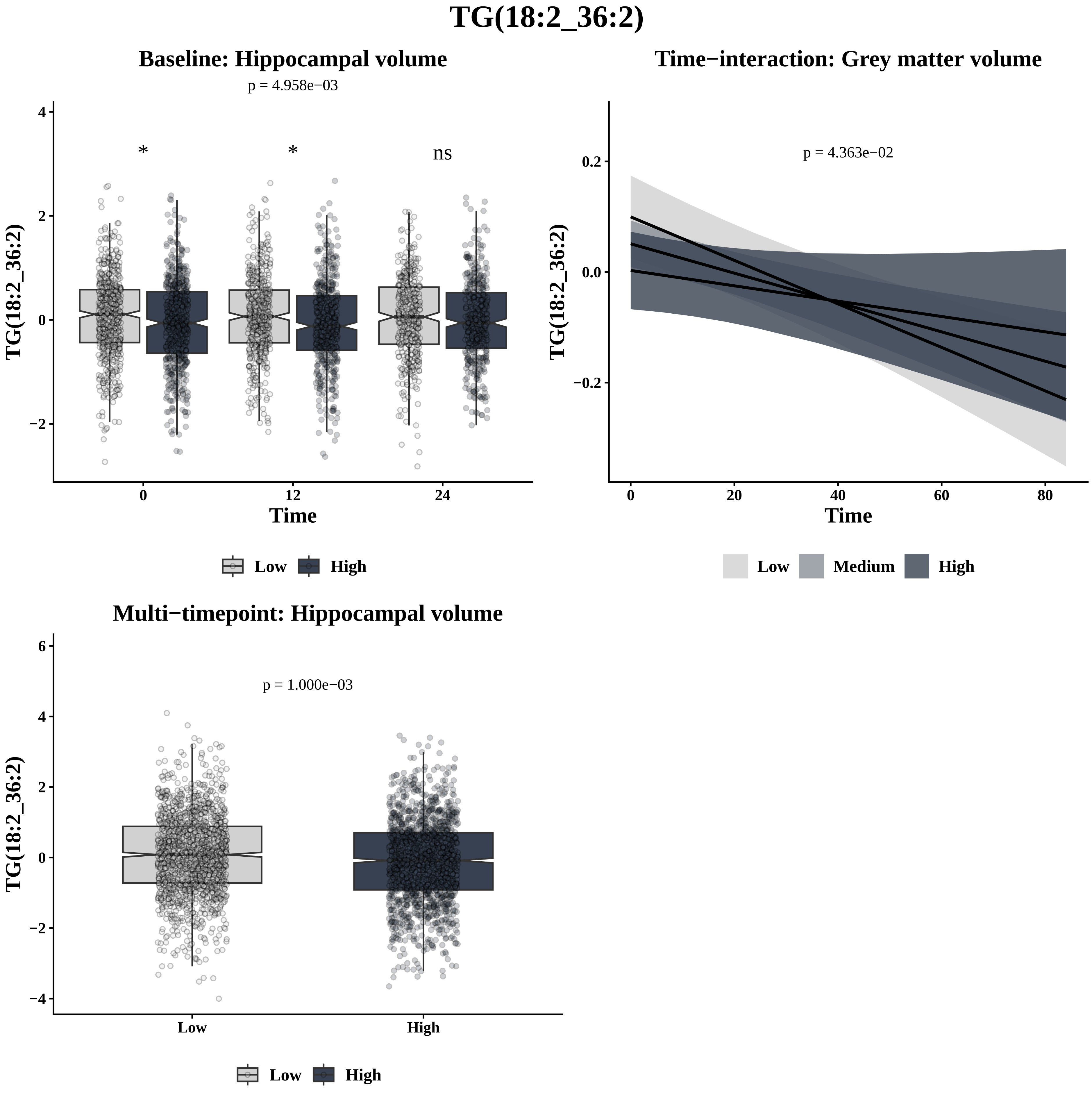
TG(18:2_36:2)
Baseline: Hippocampal volume
p = 4.958e−03
*
*
ns
TG(18:2_36:2)
Time
0
12
24
Low
High
Time−interaction: Grey matter volume
p = 4.363e−02
TG(18:2_36:2)
Time
Low
Medium
High
Multi−timepoint: Hippocampal volume
p = 1.000e−03
TG(18:2_36:2)
Low
High
Low
High

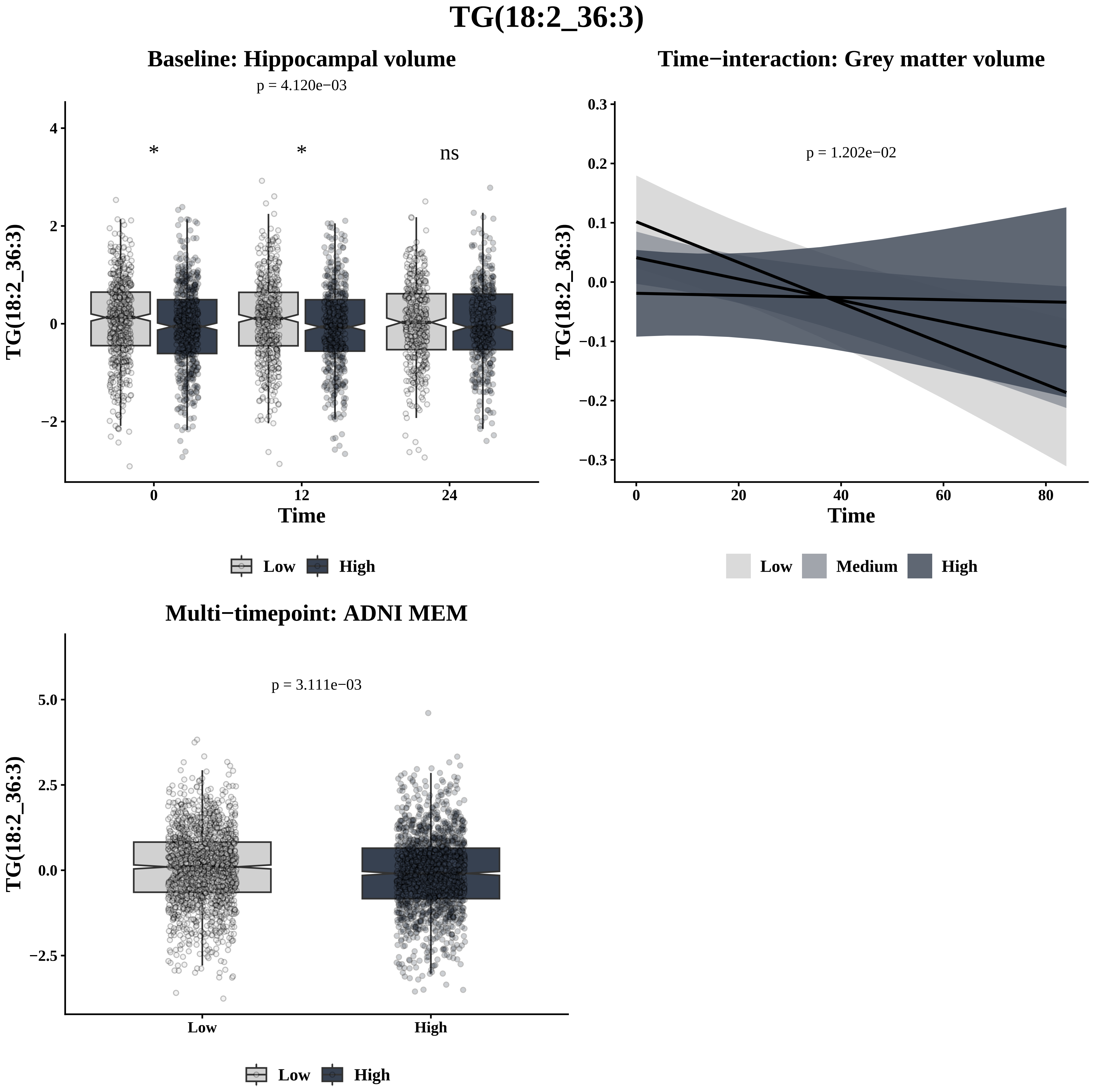

TG(18:2_36:3)
Baseline: Hippocampal volume
p = 4.120e−03
*
*
ns
TG(18:2_36:3)
Time
0
12
24
Low
High
Time−interaction: Grey matter volume
p = 1.202e−02
TG(18:2_36:3)
Time
Low
Medium
High
Multi−timepoint: ADNI MEM
p = 3.111e−03
TG(18:2_36:3)
Low
High

# TG(18:2_36:4)

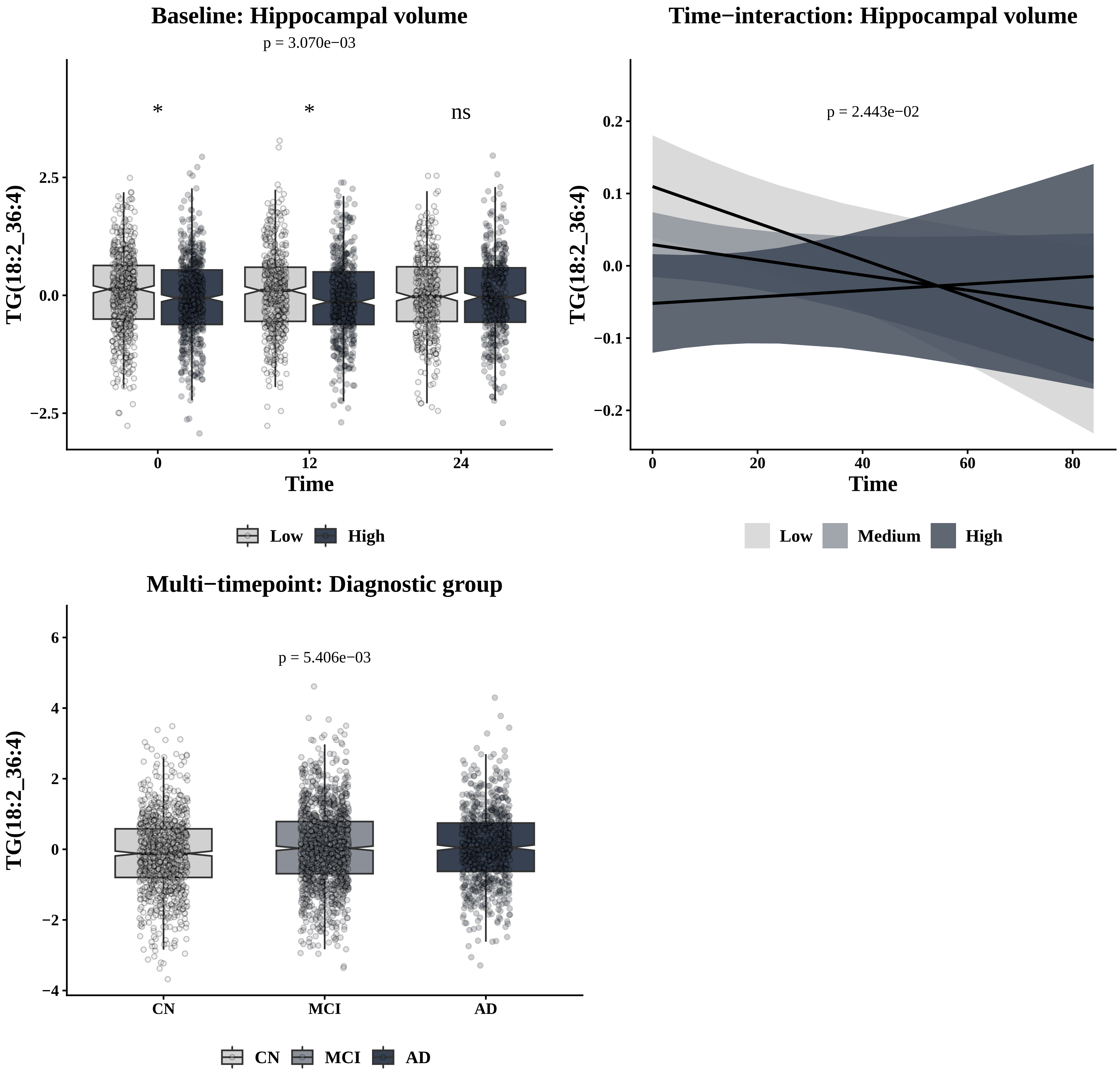

Baseline: Hippocampal volume
p = 3.070e−03
*
*
ns
TG(18:2_36:4)
2.5
0.0
−2.5
0
12
24
Time
Low
High
Time−interaction: Hippocampal volume
p = 2.443e−02
0.2
0.1
0.0
−0.1
−0.2
0
20
40
60
80
Time
Low
Medium
High
Multi−timepoint: Diagnostic group
p = 5.406e−03
6
4
2
0
−2
−4
CN
MCI
AD
CN
MCI
AD

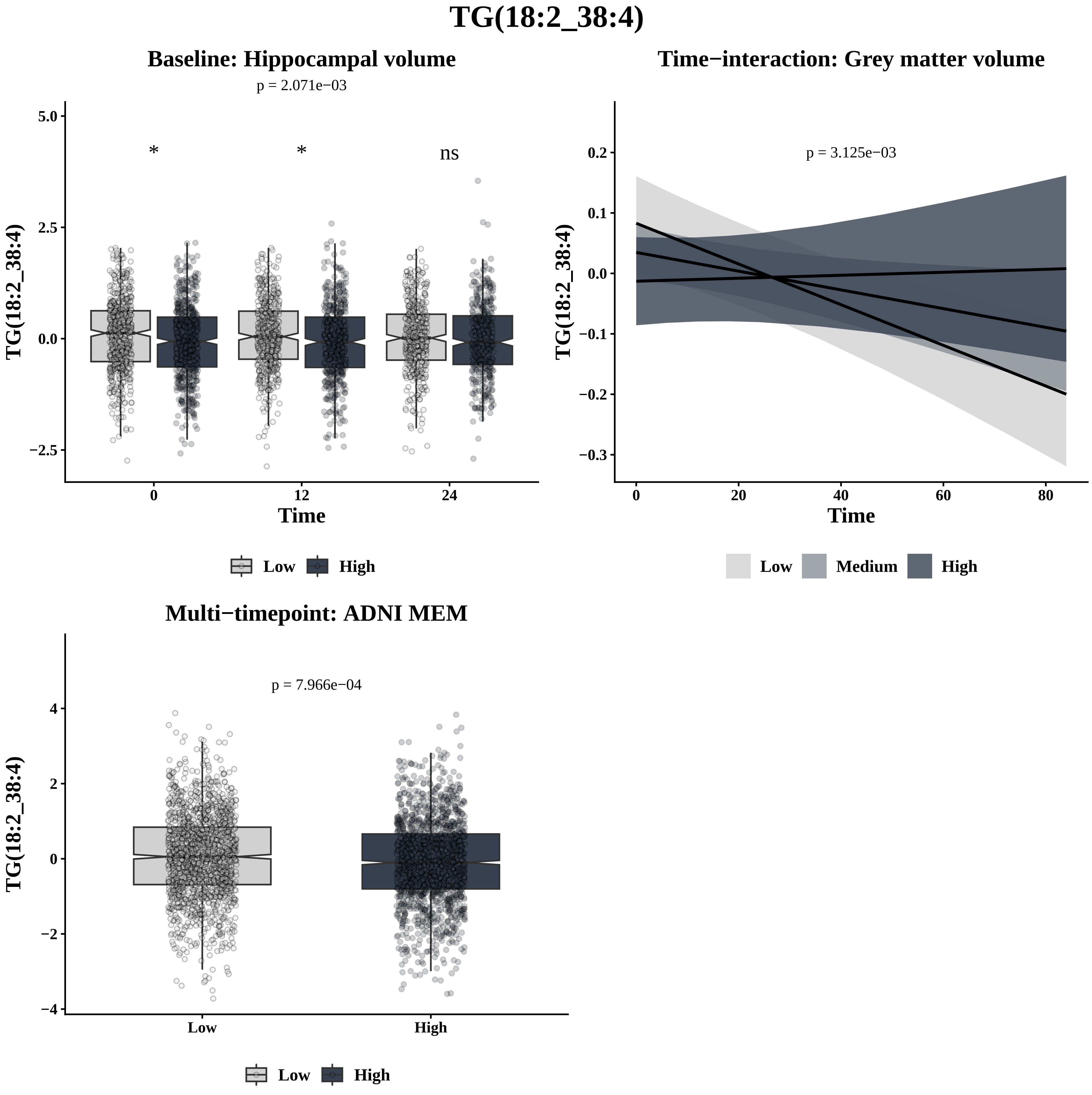

TG(18:2_38:4)
Baseline: Hippocampal volume
p = 2.071e−03
*
*
ns
5.0
2.5
0.0
−2.5
TG(18:2_38:4)
0
12
24
Time
Low
High
Time−interaction: Grey matter volume
p = 3.125e−03
0.2
0.1
0.0
−0.1
−0.2
−0.3
TG(18:2_38:4)
0
20
40
60
80
Time
Low
Medium
High
Multi−timepoint: ADNI MEM
p = 7.966e−04
4
2
0
−2
−4
TG(18:2_38:4)
Low
High
Low
High

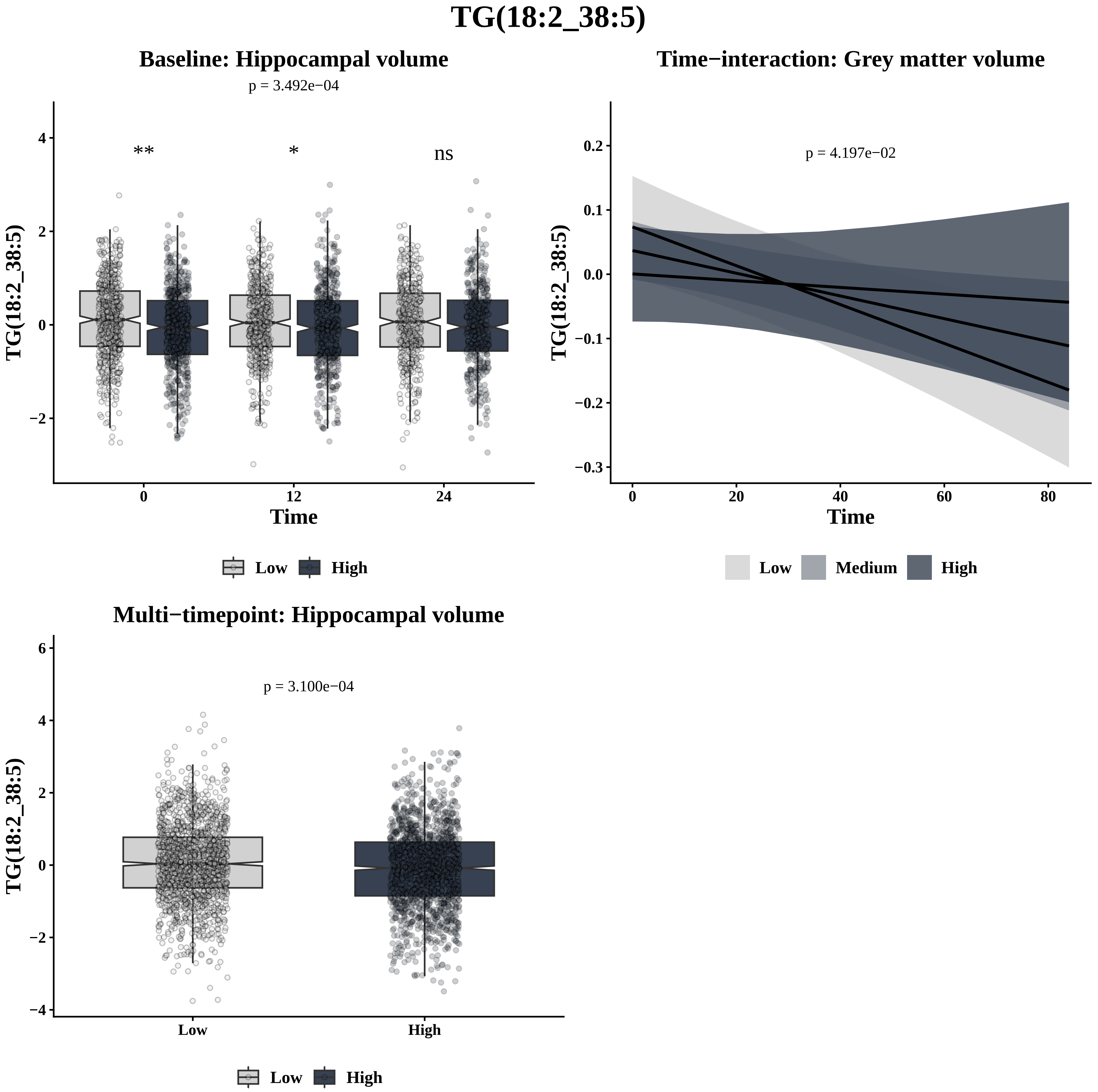
TG(18:2_38:5)
Baseline: Hippocampal volume
p = 3.492e−04
**
*
ns
TG(18:2_38:5)
4
2
0
−2
0
12
24
Time
Low
High
Time−interaction: Grey matter volume
p = 4.197e−02
TG(18:2_38:5)
0.2
0.1
0.0
−0.1
−0.2
−0.3
0
20
40
60
80
Time
Low
Medium
High
Multi−timepoint: Hippocampal volume
p = 3.100e−04
TG(18:2_38:5)
6
4
2
0
−2
−4
Low
High
Low
High

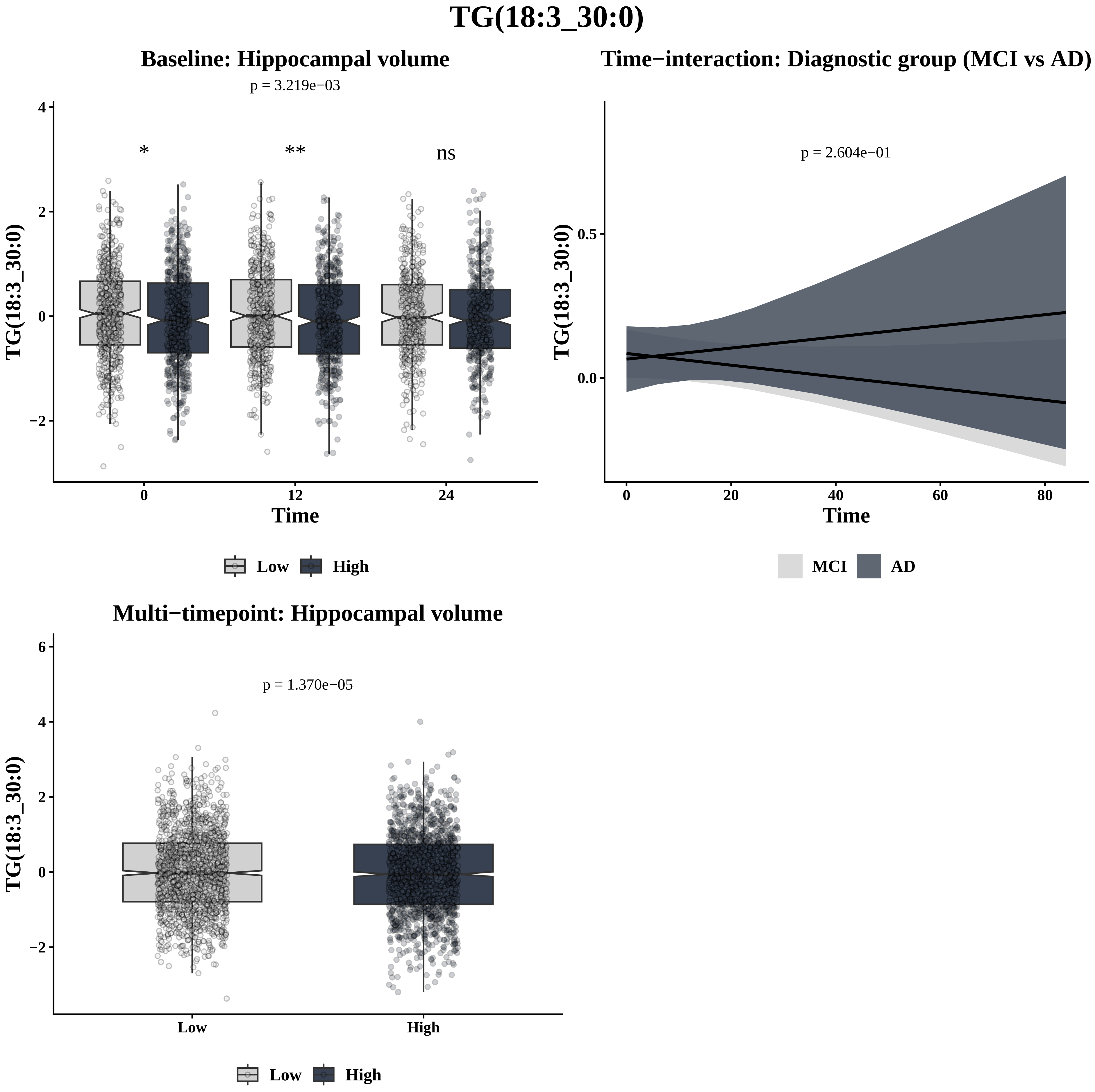

TG(18:3_30:0)
Baseline: Hippocampal volume
p = 3.219e−03
*
**
ns
TG(18:3_30:0)
Time
Low
High
Time−interaction: Diagnostic group (MCI vs AD)
p = 2.604e−01
TG(18:3_30:0)
Time
MCI
AD
Multi−timepoint: Hippocampal volume
p = 1.370e−05
TG(18:3_30:0)
Low
High

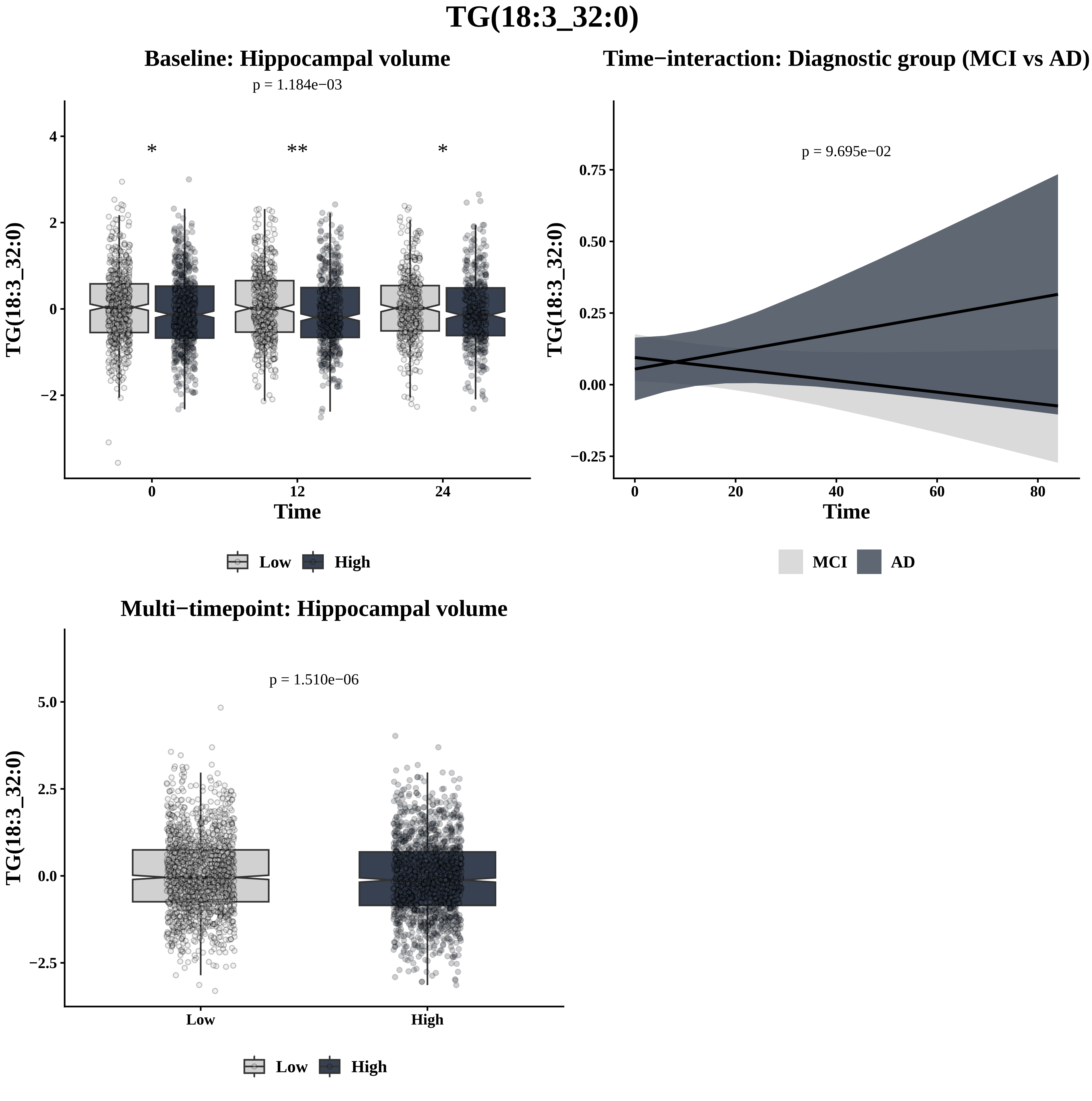

TG(18:3_32:0)
Baseline: Hippocampal volume
p = 1.184e−03
*
**
*
TG(18:3_32:0)
Time
0
12
24
Low
High
Time−interaction: Diagnostic group (MCI vs AD)
p = 9.695e−02
TG(18:3_32:0)
Time
MCI
AD
Multi−timepoint: Hippocampal volume
p = 1.510e−06
TG(18:3_32:0)
Low
High

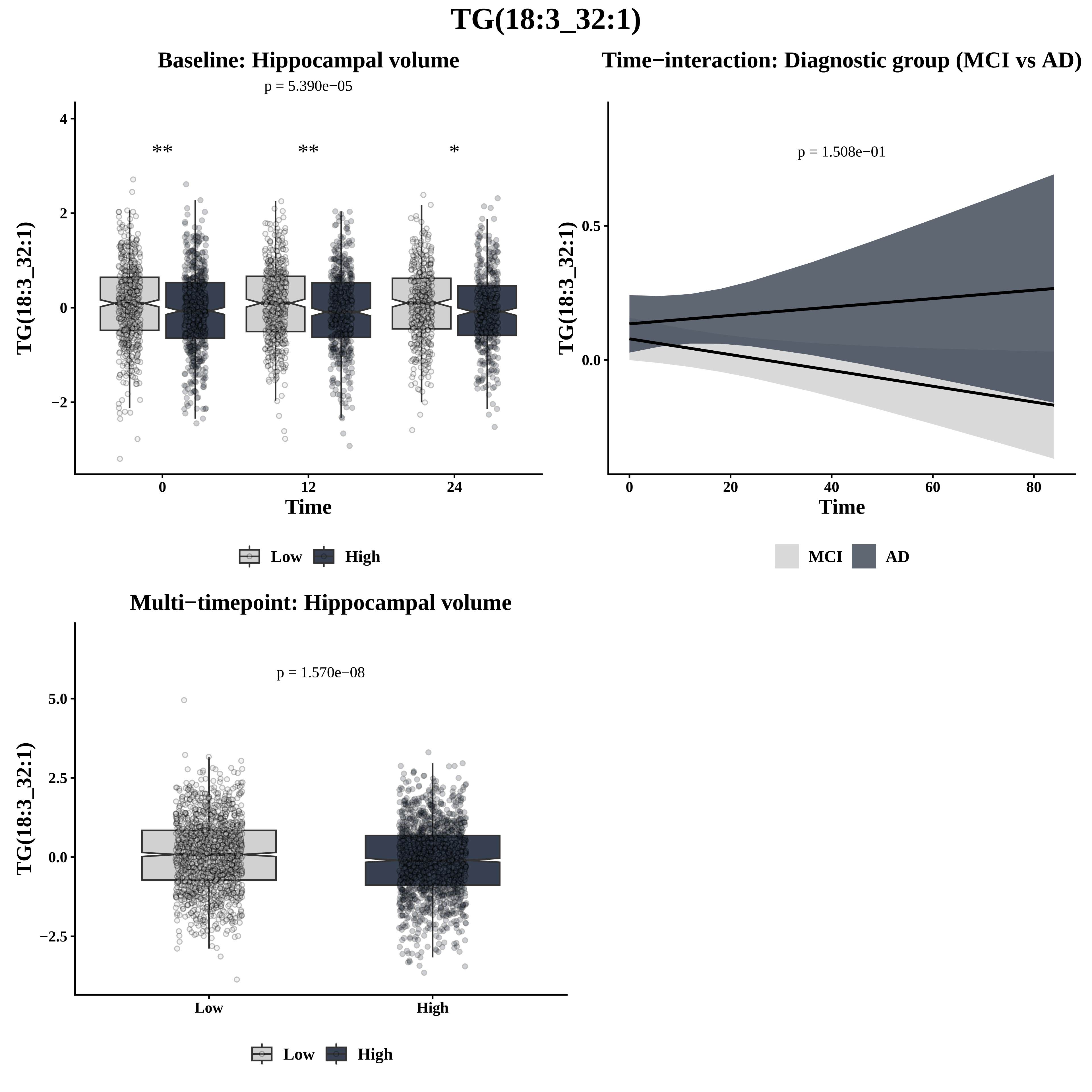

TG(18:3_32:1)
Baseline: Hippocampal volume
p = 5.390e−05
**
**
*
TG(18:3_32:1)
Time
0
12
24
Low
High
Time−interaction: Diagnostic group (MCI vs AD)
p = 1.508e−01
TG(18:3_32:1)
Time
MCI
AD
Multi−timepoint: Hippocampal volume
p = 1.570e−08
TG(18:3_32:1)
Low
High

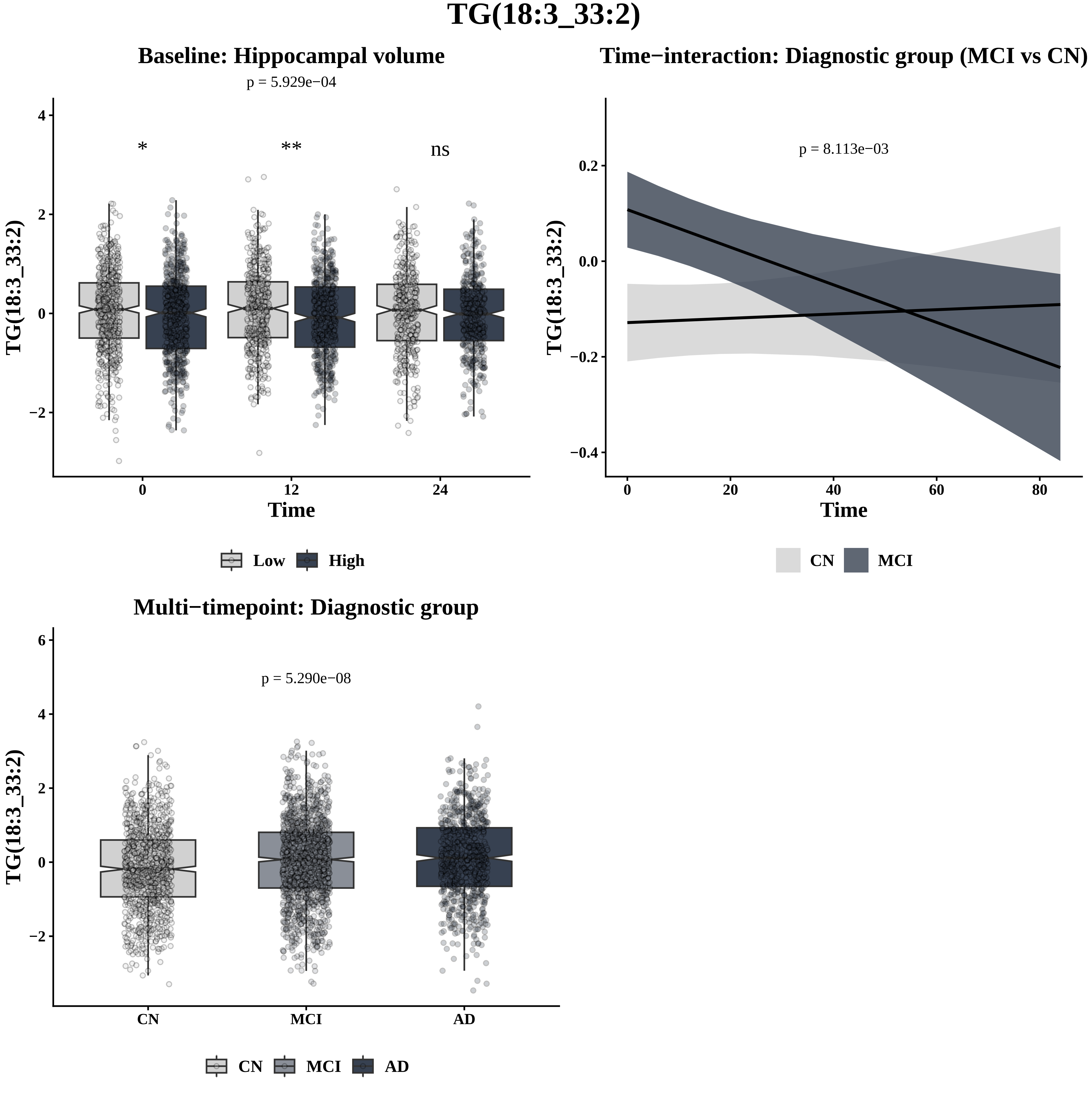

TG(18:3_33:2)
Baseline: Hippocampal volume
p = 5.929e−04
*
**
ns
TG(18:3_33:2)
Time
Low
High
Time−interaction: Diagnostic group (MCI vs CN)
p = 8.113e−03
TG(18:3_33:2)
Time
CN
MCI
Multi−timepoint: Diagnostic group
p = 5.290e−08
TG(18:3_33:2)
CN
MCI
AD

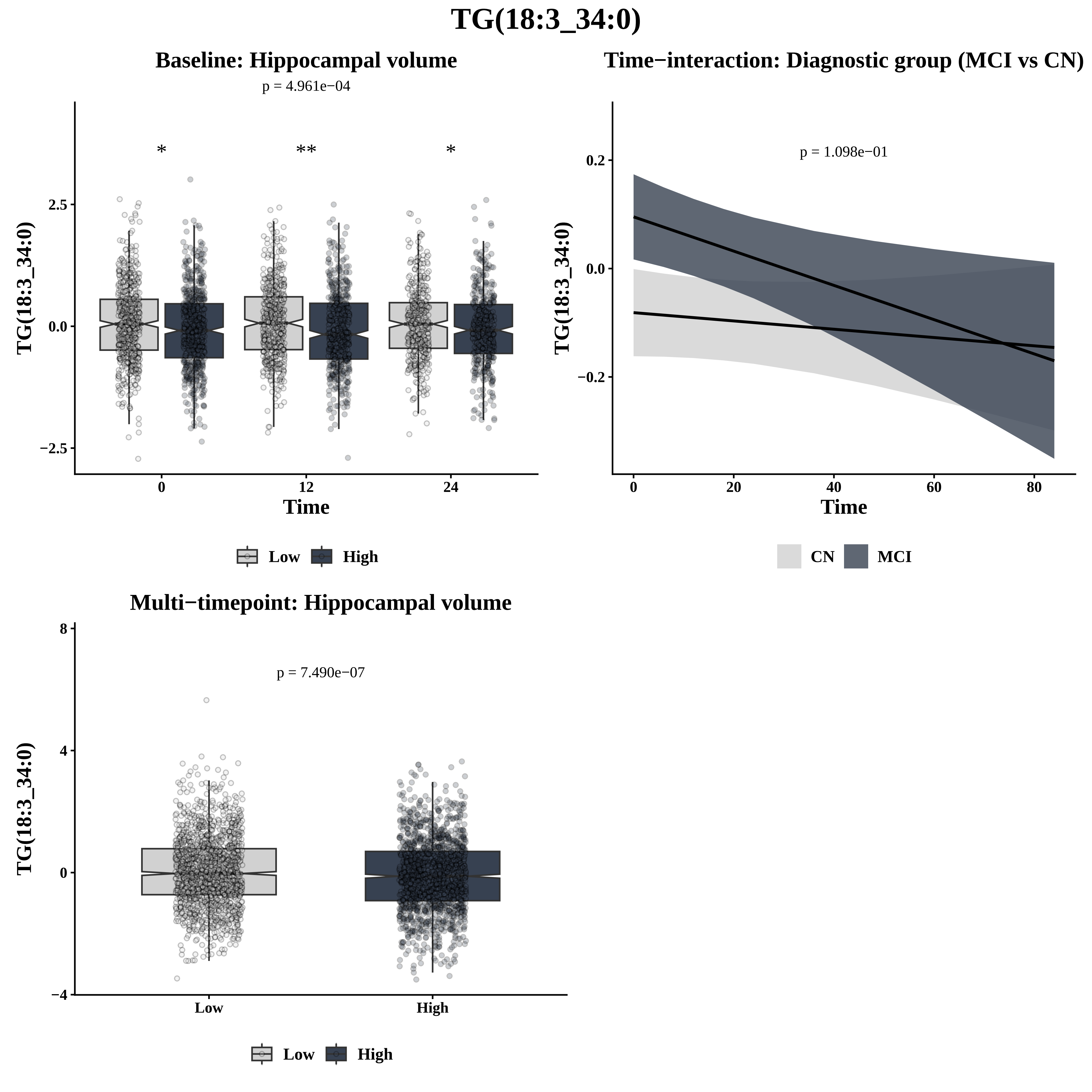

TG(18:3_34:0)
Baseline: Hippocampal volume
p = 4.961e−04
*
**
*
2.5
0.0
−2.5
0
12
24
Time
TG(18:3_34:0)
Low
High
Time−interaction: Diagnostic group (MCI vs CN)
p = 1.098e−01
0.2
0.0
−0.2
0
20
40
60
80
Time
TG(18:3_34:0)
CN
MCI
Multi−timepoint: Hippocampal volume
p = 7.490e−07
8
4
0
−4
Low
High
TG(18:3_34:0)
Low
High

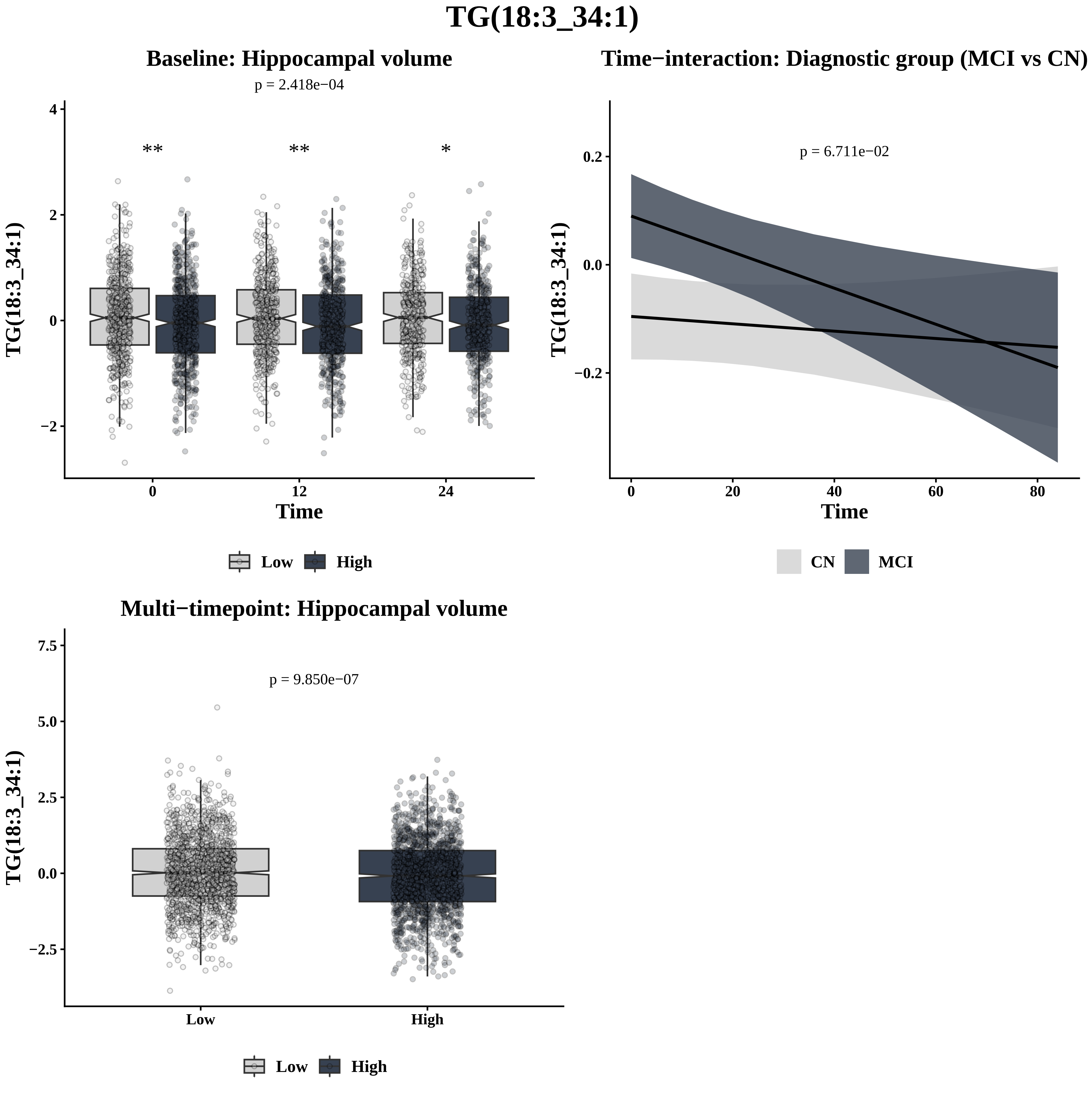
TG(18:3_34:1)
Baseline: Hippocampal volume
p = 2.418e−04
**
**
*
TG(18:3_34:1)
Time
Low
High
Time−interaction: Diagnostic group (MCI vs CN)
p = 6.711e−02
TG(18:3_34:1)
Time
CN
MCI
Multi−timepoint: Hippocampal volume
p = 9.850e−07
TG(18:3_34:1)
Low
High
Low
High

# TG(18:3_34:2)

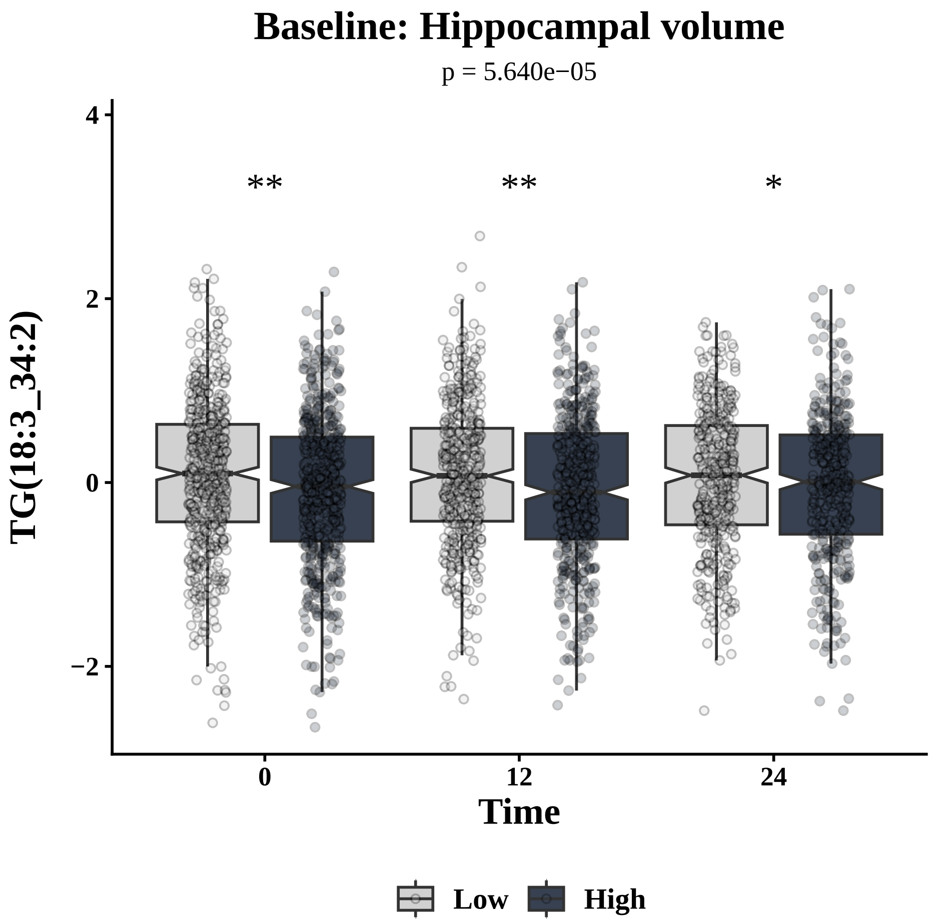

Baseline: Hippocampal volume
p = 5.640e−05
TG(18:3_34:2)
**
**
*
4
2
0
−2
0
12
24
Time
Low
High


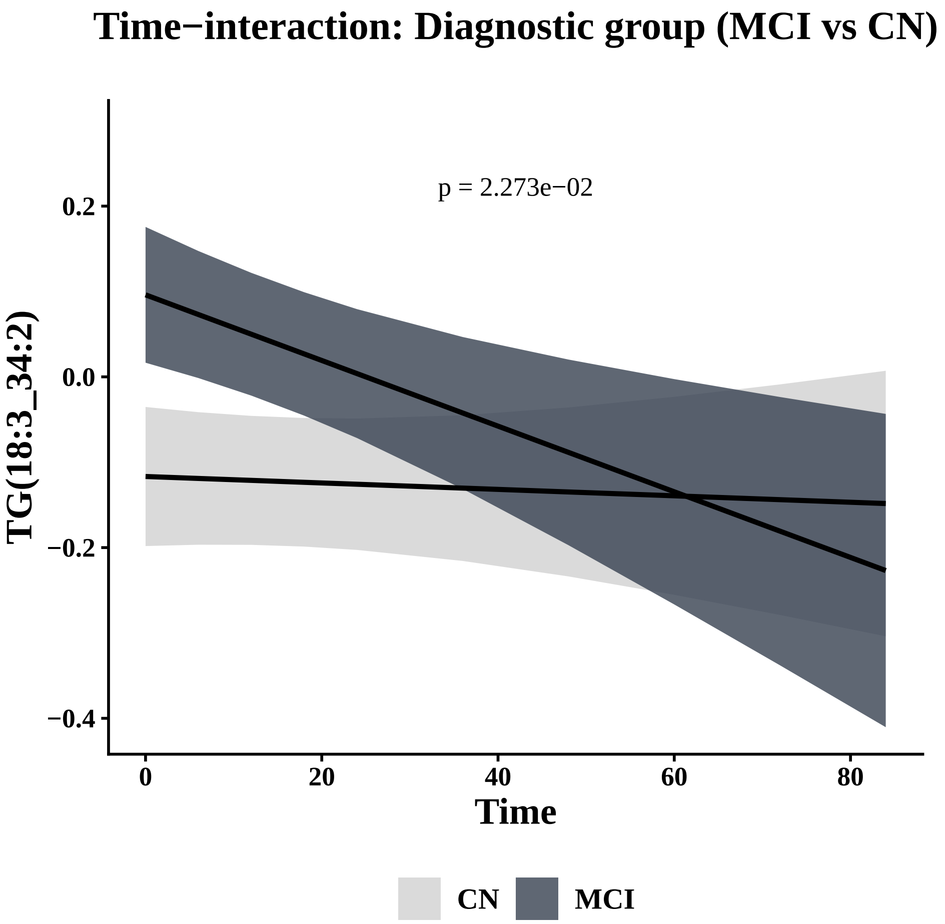

Time−interaction: Diagnostic group (MCI vs CN)
p = 2.273e−02
TG(18:3_34:2)
0.2
0.0
−0.2
−0.4
0
20
40
60
80
Time
CN
MCI


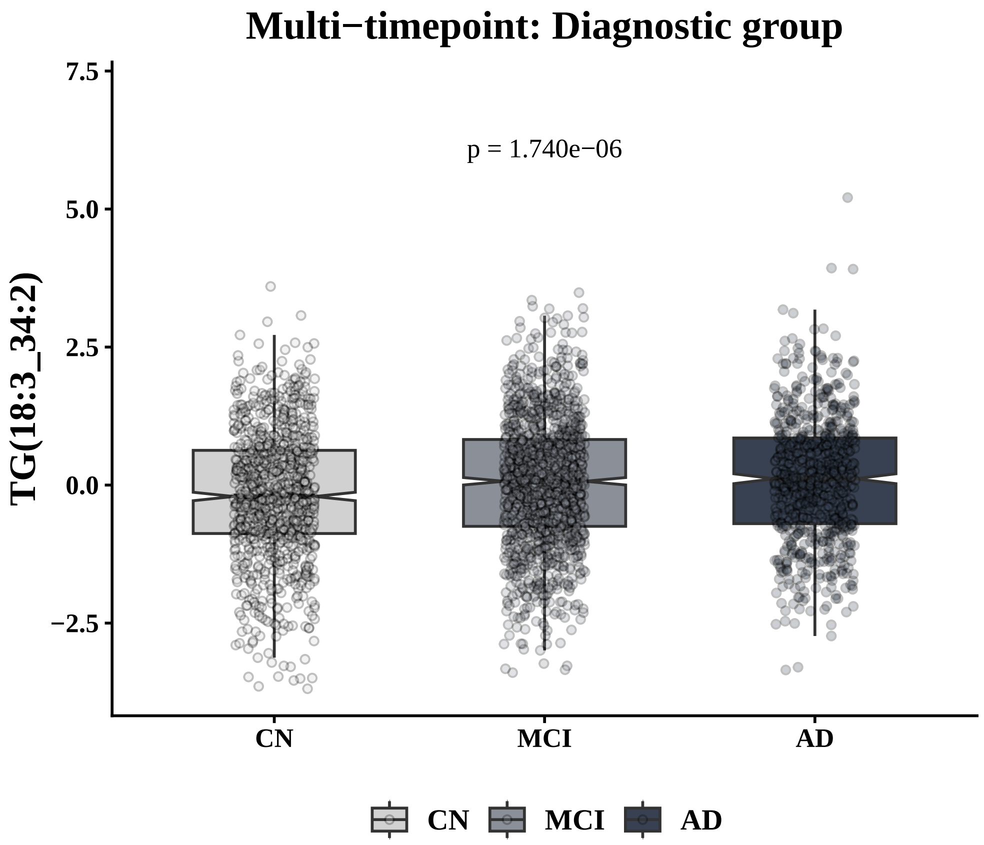

Multi−timepoint: Diagnostic group
p = 1.740e−06
TG(18:3_34:2)
7.5
5.0
2.5
0.0
−2.5
CN
MCI
AD
CN
MCI
AD

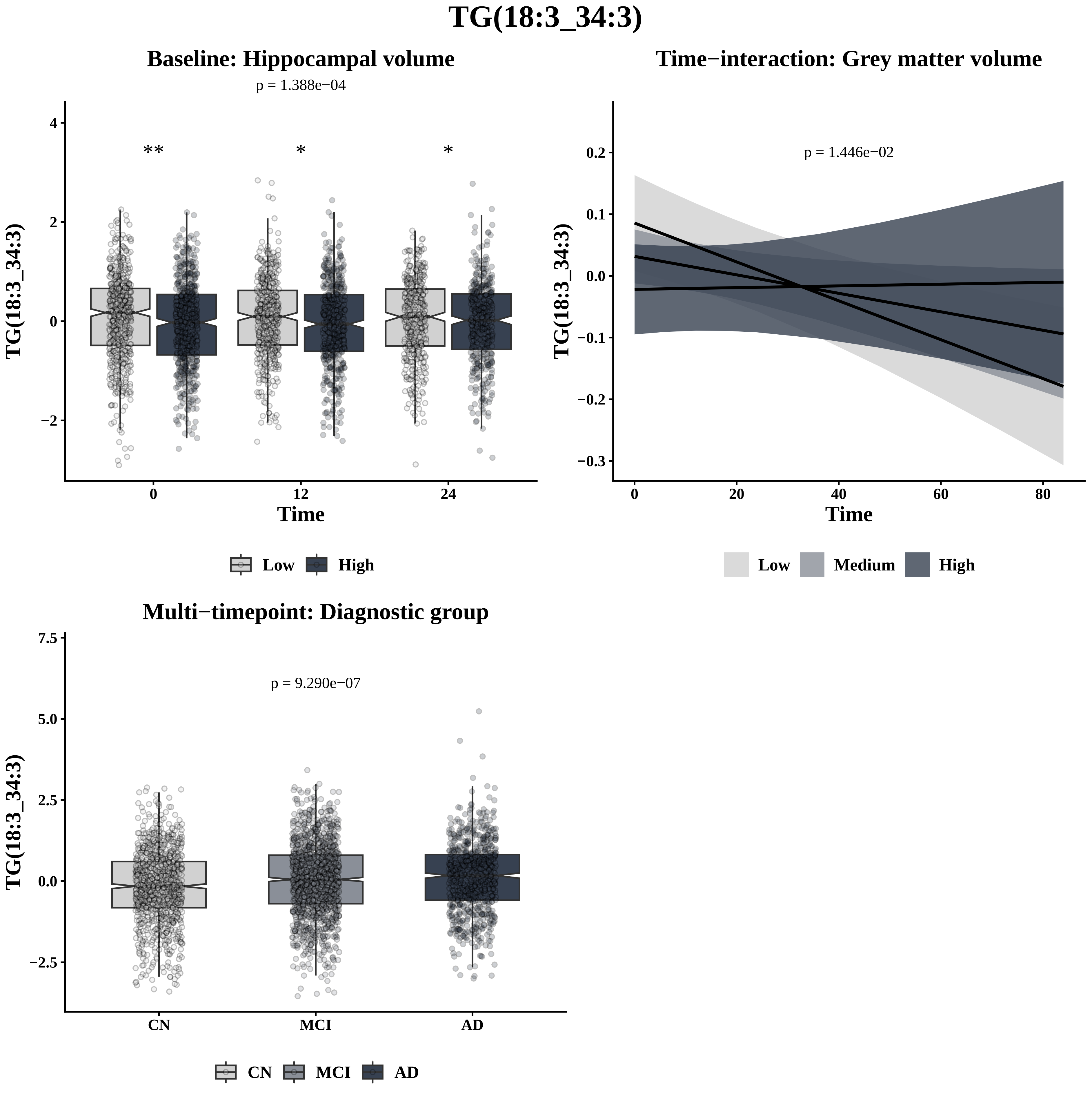

TG(18:3_34:3)
Baseline: Hippocampal volume
p = 1.388e−04
**
*
*
TG(18:3_34:3)
Time
Low
High
Time−interaction: Grey matter volume
p = 1.446e−02
TG(18:3_34:3)
Time
Low
Medium
High
Multi−timepoint: Diagnostic group
p = 9.290e−07
TG(18:3_34:3)
CN
MCI
AD

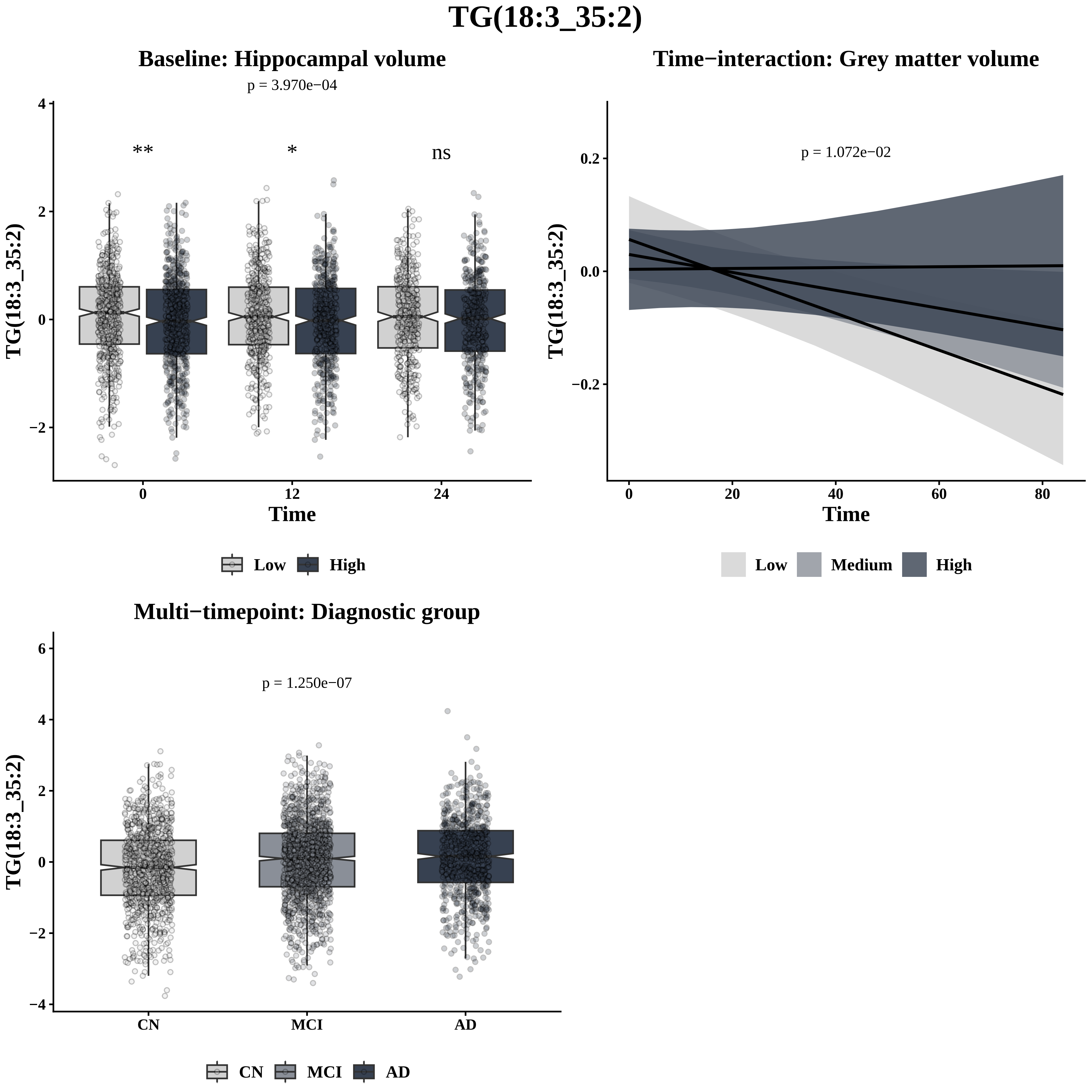

TG(18:3_35:2)
Baseline: Hippocampal volume
p = 3.970e−04
**
*
ns
TG(18:3_35:2)
Time
0
12
24
Low
High
Time−interaction: Grey matter volume
p = 1.072e−02
TG(18:3_35:2)
Time
Low
Medium
High
Multi−timepoint: Diagnostic group
p = 1.250e−07
TG(18:3_35:2)
CN
MCI
AD

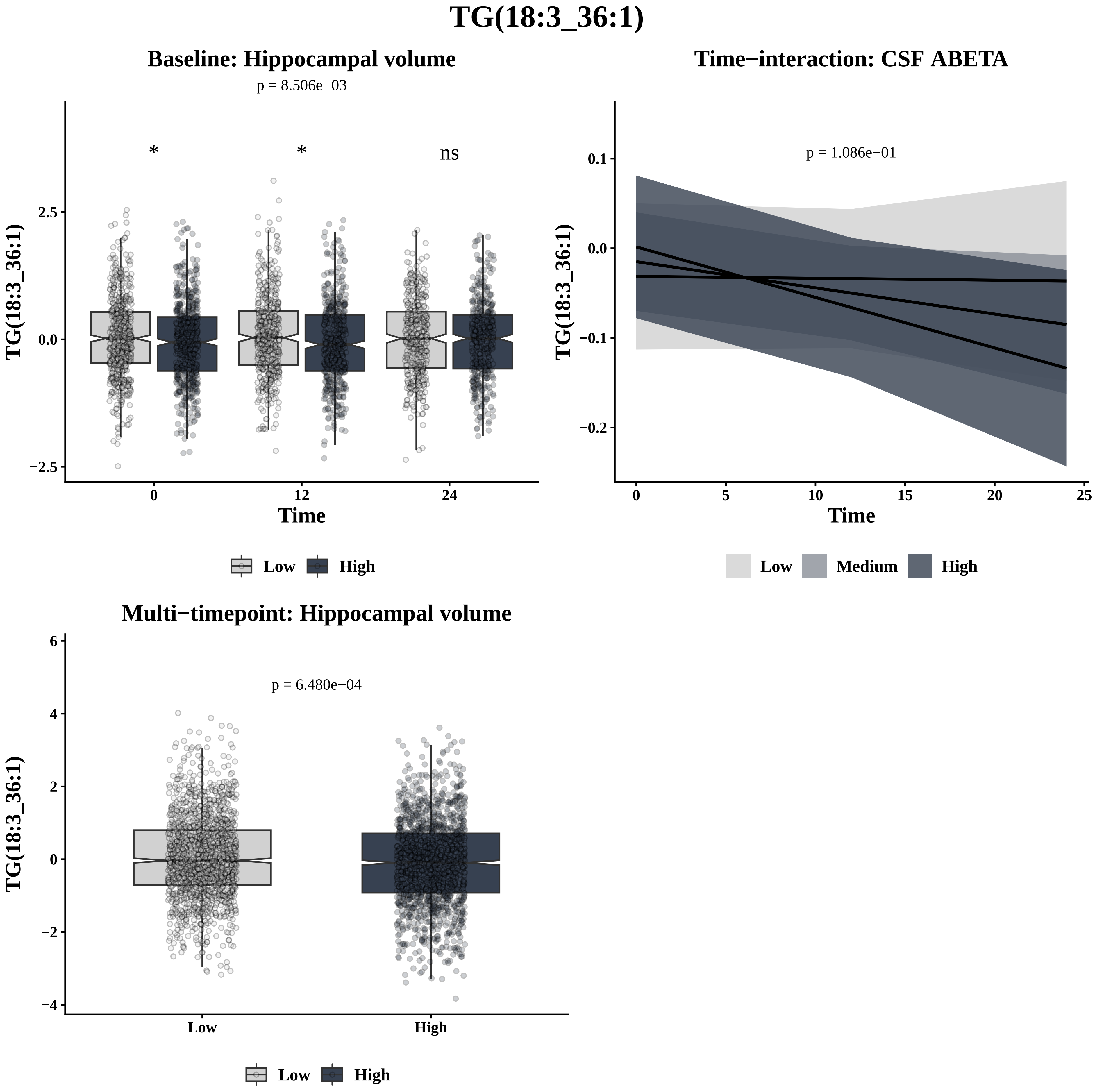
TG(18:3_36:1)
Baseline: Hippocampal volume
p = 8.506e−03
*
*
ns
TG(18:3_36:1)
Time
Low
High
Time−interaction: CSF ABETA
p = 1.086e−01
TG(18:3_36:1)
Time
Low
Medium
High
Multi−timepoint: Hippocampal volume
p = 6.480e−04
TG(18:3_36:1)
Low
High
Low
High

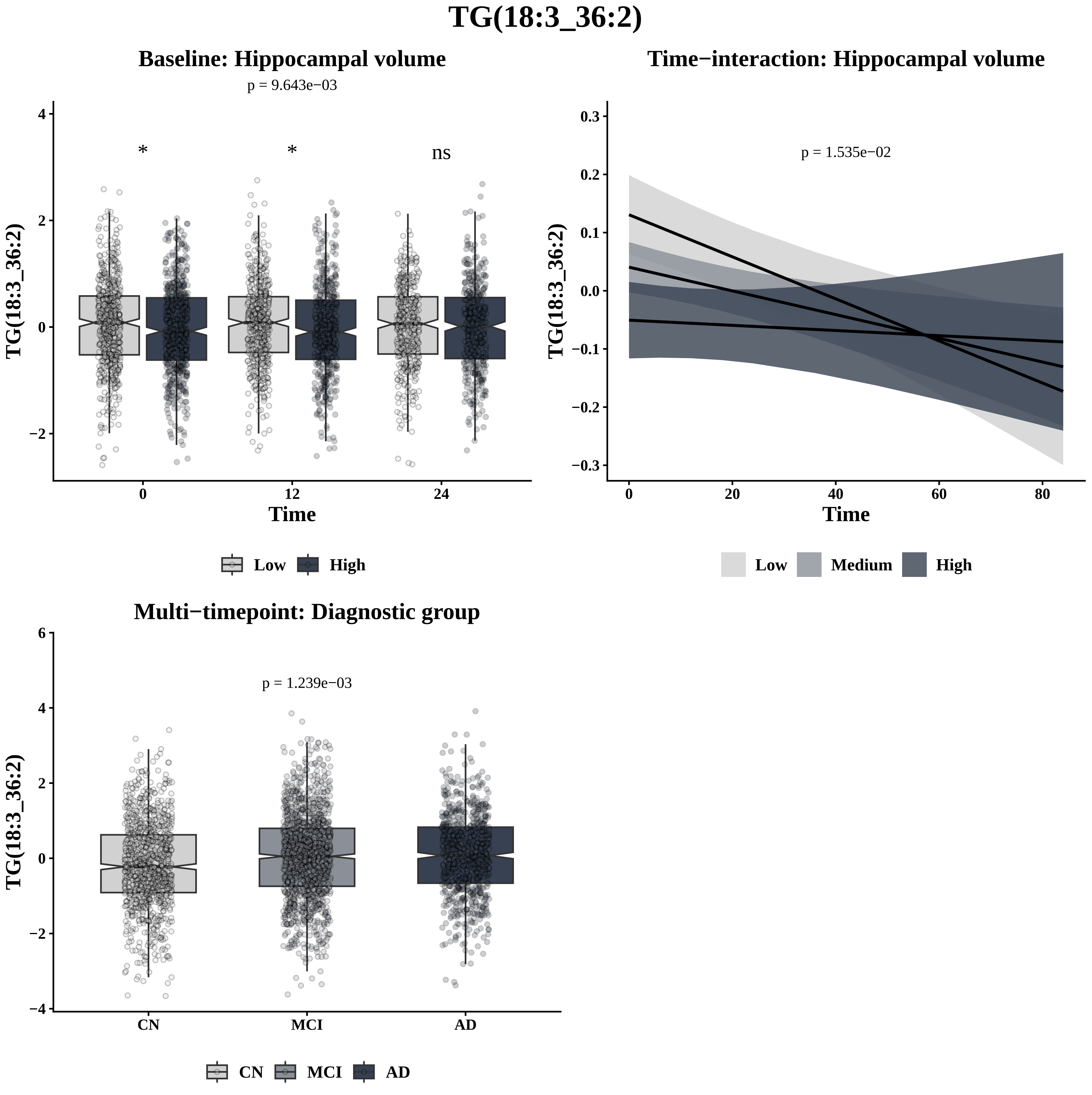
TG(18:3_36:2)
Baseline: Hippocampal volume
p = 9.643e−03
*
*
ns
TG(18:3_36:2)
4
2
0
−2
0
12
24
Time
Low
High
Time−interaction: Hippocampal volume
p = 1.535e−02
TG(18:3_36:2)
0.3
0.2
0.1
0.0
−0.1
−0.2
−0.3
0
20
40
60
80
Time
Low
Medium
High
Multi−timepoint: Diagnostic group
p = 1.239e−03
TG(18:3_36:2)
6
4
2
0
−2
−4
CN
MCI
AD
CN
MCI
AD

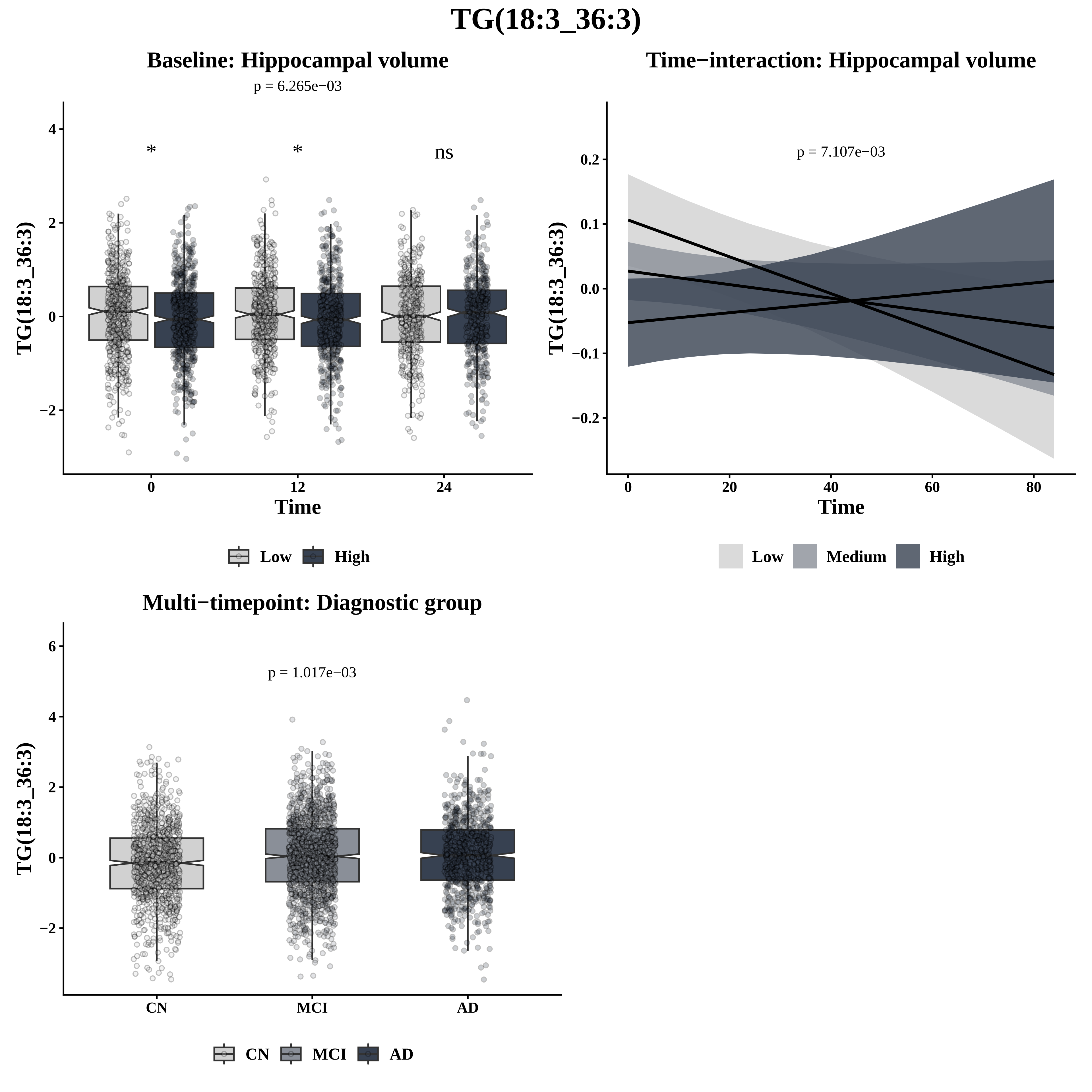

TG(18:3_36:3)
Baseline: Hippocampal volume
p = 6.265e−03
*
*
ns
TG(18:3_36:3)
Time
0
12
24
Low
High
Time−interaction: Hippocampal volume
p = 7.107e−03
TG(18:3_36:3)
Time
Low
Medium
High
Multi−timepoint: Diagnostic group
p = 1.017e−03
TG(18:3_36:3)
CN
MCI
AD

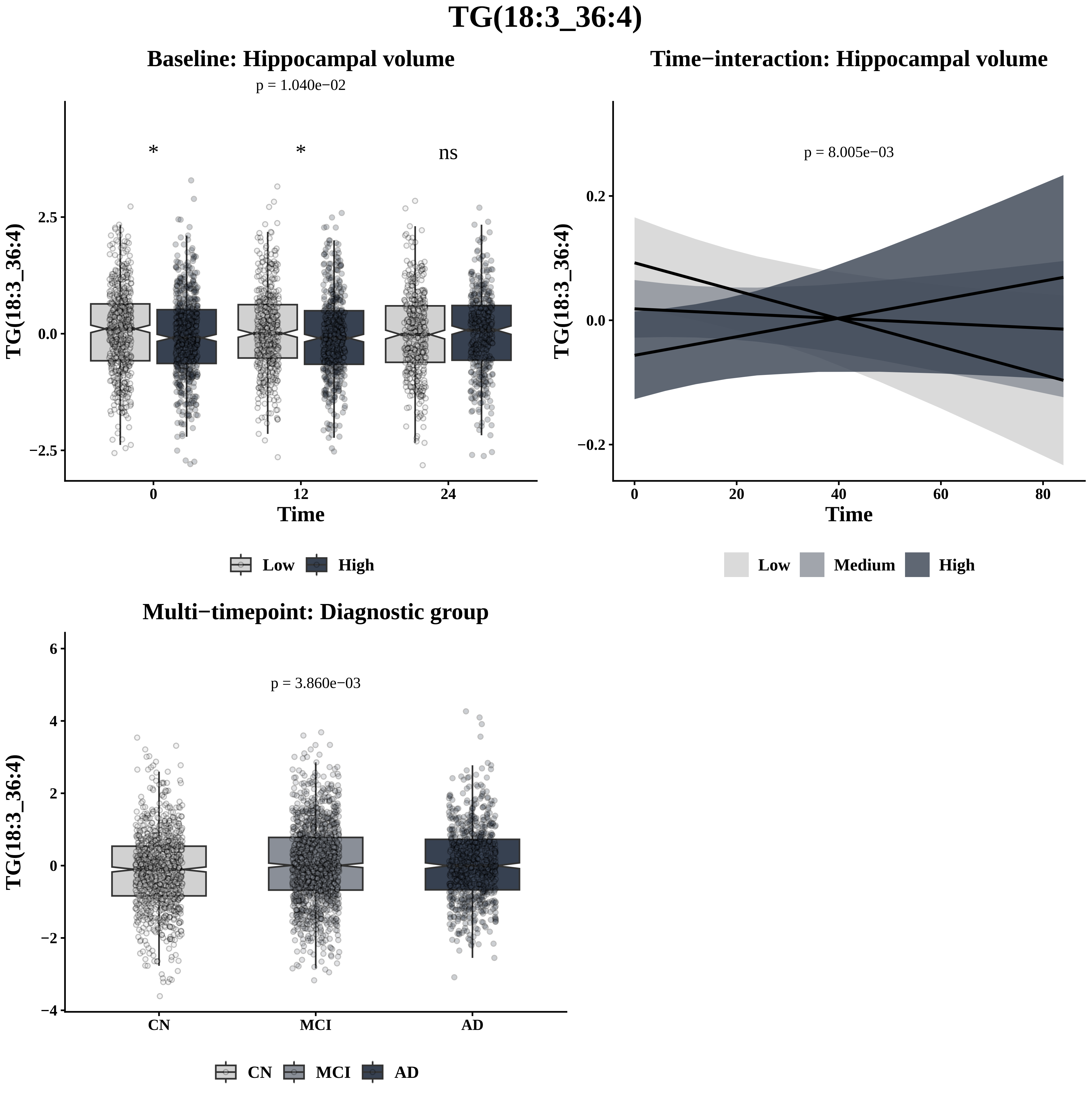

TG(18:3_36:4)
Baseline: Hippocampal volume
p = 1.040e−02
*
*
ns
2.5
0.0
−2.5
TG(18:3_36:4)
0
12
24
Time
Low
High
Time−interaction: Hippocampal volume
p = 8.005e−03
0.2
0.0
−0.2
TG(18:3_36:4)
0
20
40
60
80
Time
Low
Medium
High
Multi−timepoint: Diagnostic group
p = 3.860e−03
6
4
2
0
−2
−4
TG(18:3_36:4)
CN
MCI
AD
CN
MCI
AD

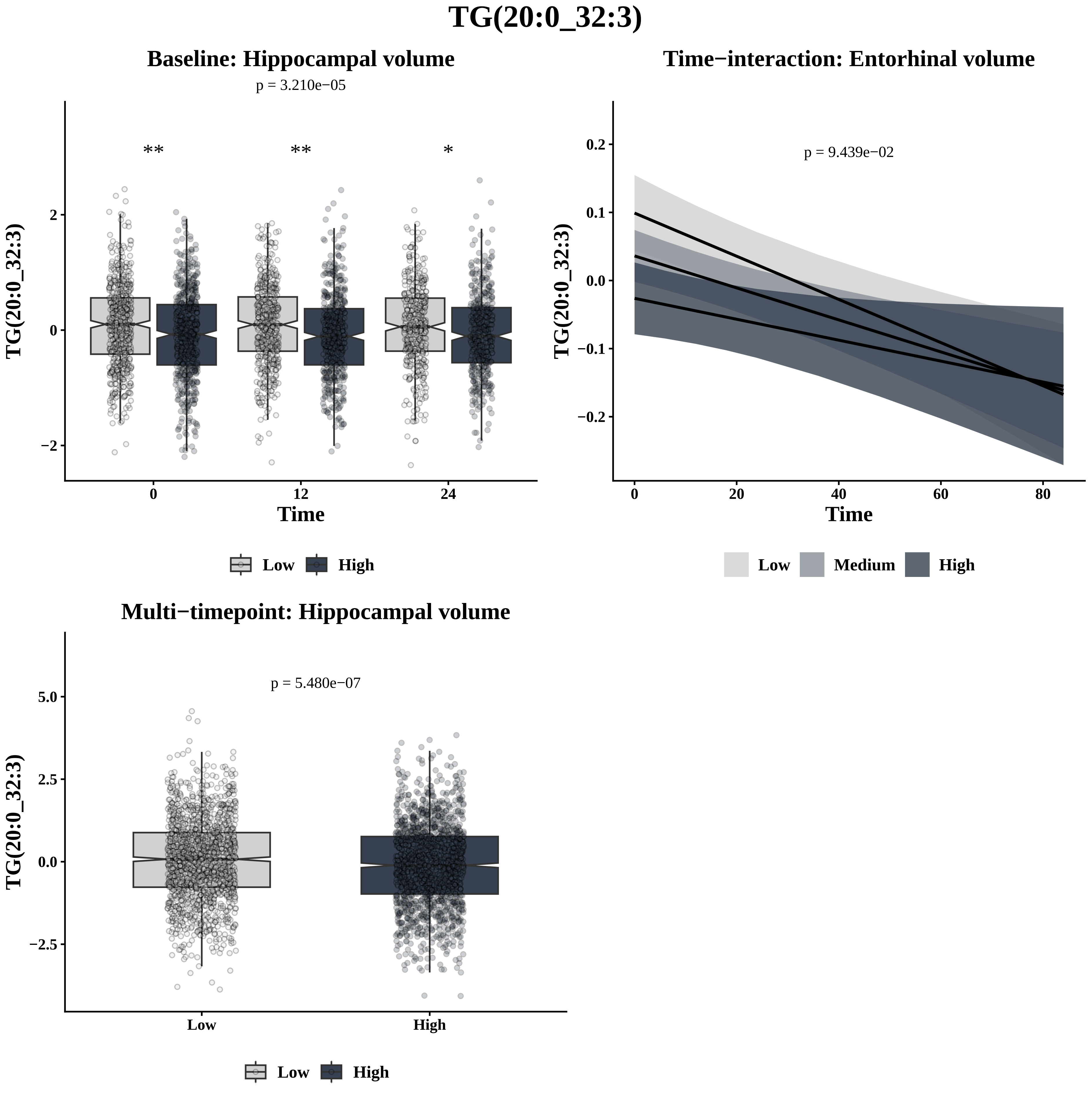

TG(20:0_32:3)
Baseline: Hippocampal volume
p = 3.210e−05
**
**
*
TG(20:0_32:3)
2
0
−2
0
12
24
Time
Low
High
Time−interaction: Entorhinal volume
p = 9.439e−02
0.2
0.1
0.0
−0.1
−0.2
0
20
40
60
80
Time
Low
Medium
High
Multi−timepoint: Hippocampal volume
p = 5.480e−07
5.0
2.5
0.0
−2.5
Low
High
Low
High

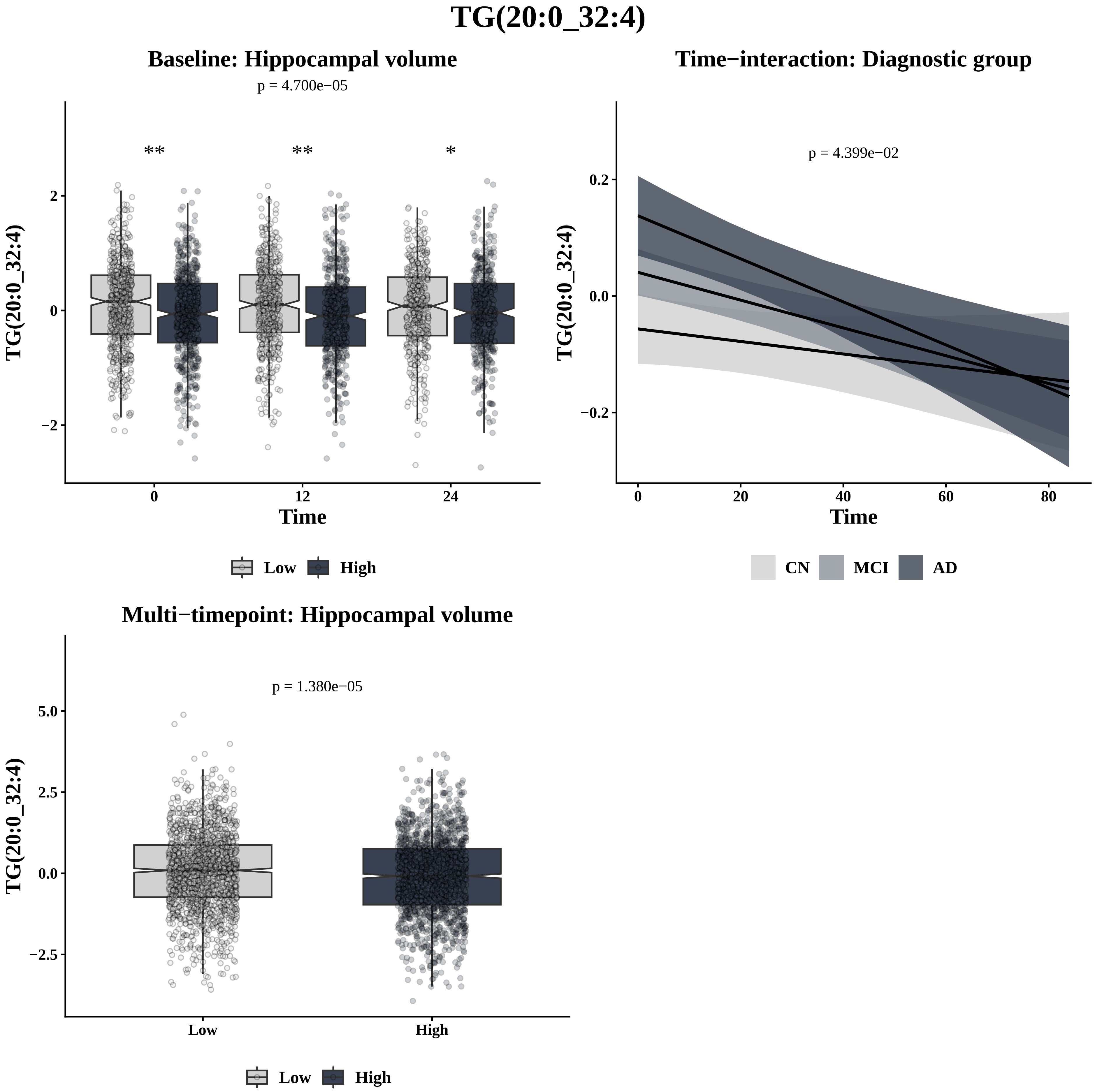
TG(20:0_32:4)
Baseline: Hippocampal volume
p = 4.700e−05
**
**
*
TG(20:0_32:4)
Time
0
12
24
2
0
−2
Low
High
Time−interaction: Diagnostic group
p = 4.399e−02
TG(20:0_32:4)
0.2
0.0
−0.2
0
20
40
60
80
Time
CN
MCI
AD
Multi−timepoint: Hippocampal volume
p = 1.380e−05
TG(20:0_32:4)
5.0
2.5
0.0
−2.5
Low
High
Low
High

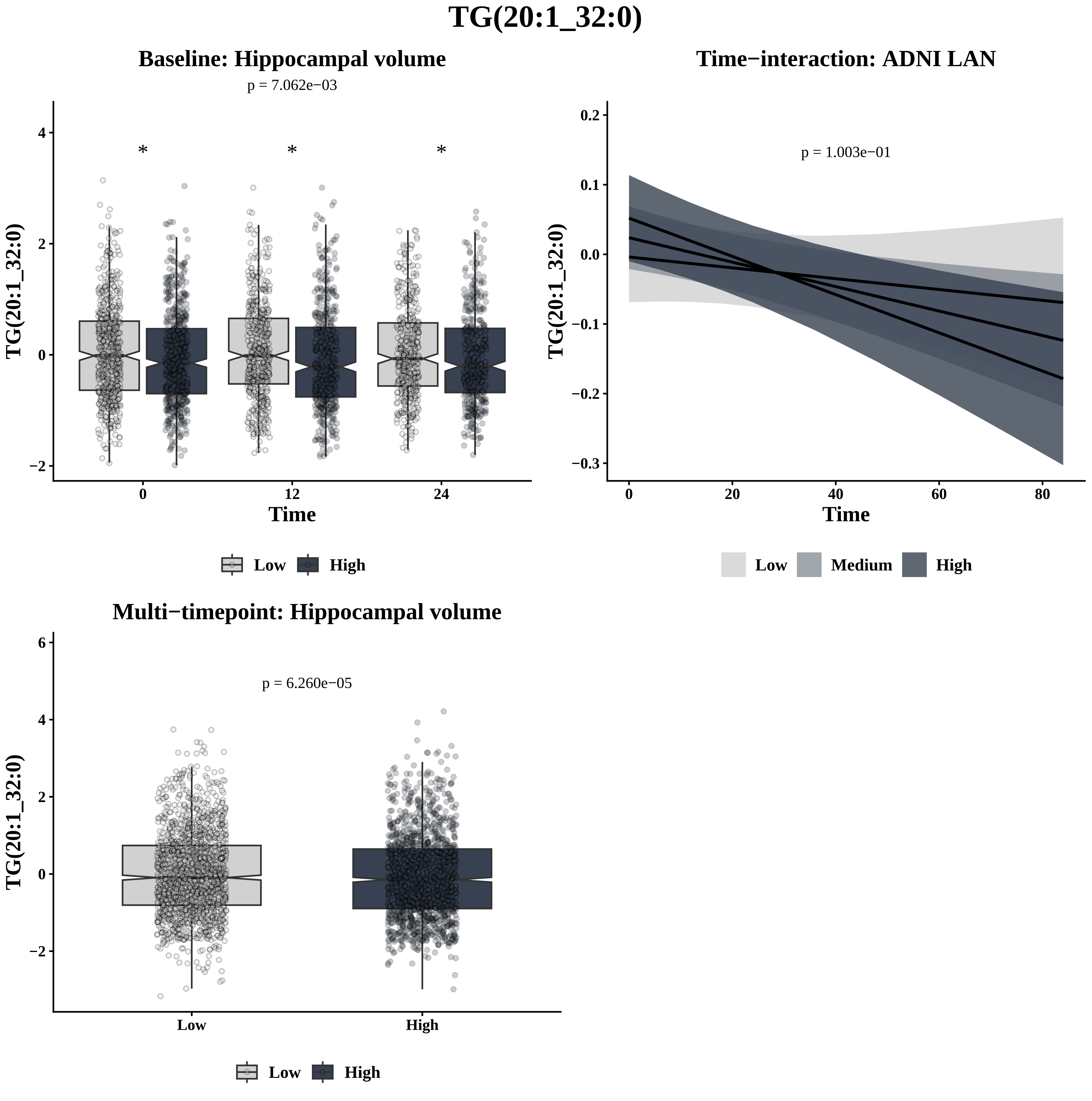
TG(20:1_32:0)
Baseline: Hippocampal volume
p = 7.062e−03
*
*
*
TG(20:1_32:0)
Time
0
12
24
4
2
0
−2
Low
High
Time−interaction: ADNI LAN
p = 1.003e−01
0.2
0.1
0.0
−0.1
−0.2
−0.3
0
20
40
60
80
Time
Low
Medium
High
Multi−timepoint: Hippocampal volume
p = 6.260e−05
6
4
2
0
−2
Low
High

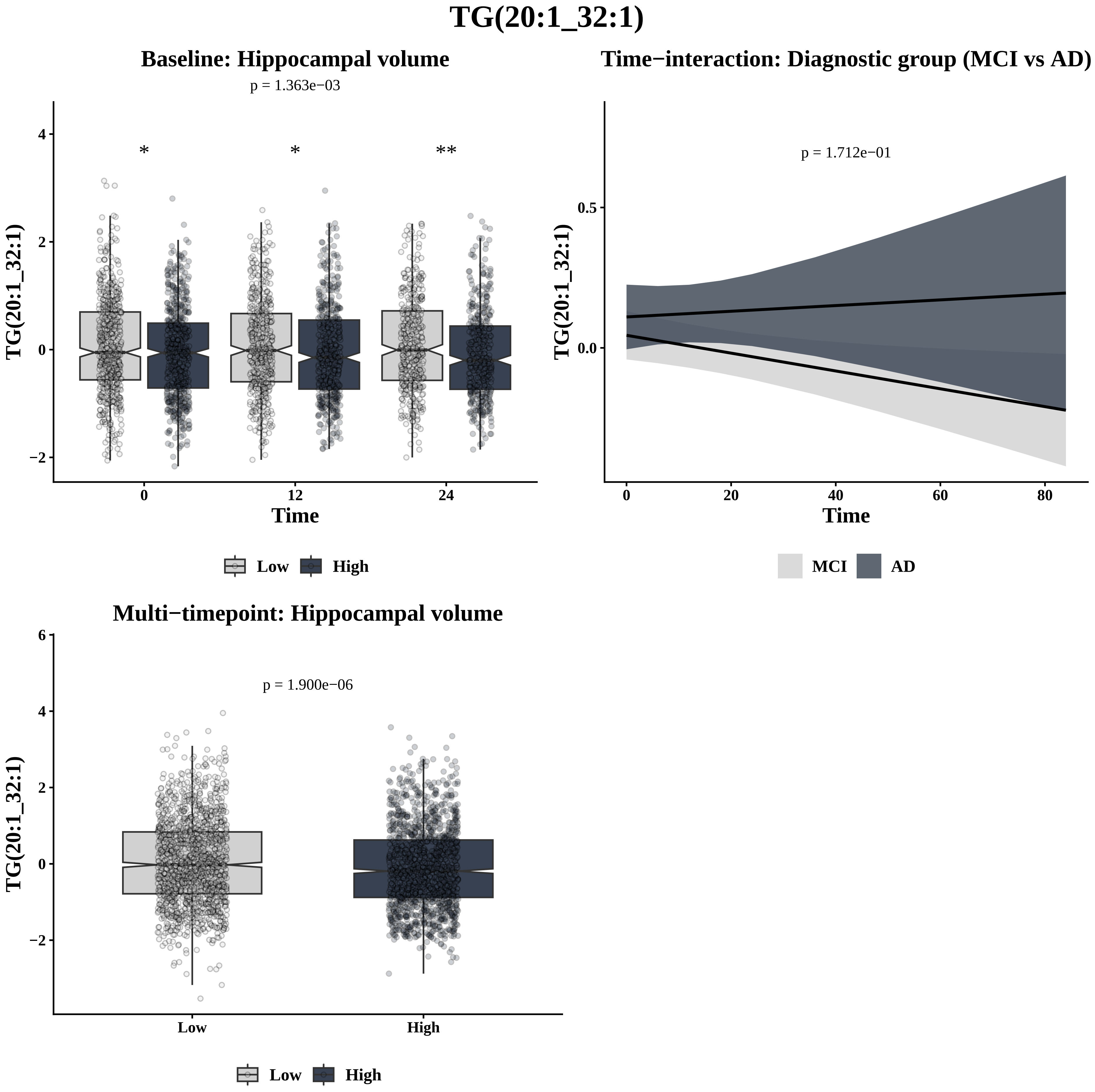
TG(20:1_32:1)
Baseline: Hippocampal volume
p = 1.363e−03
*
*
**
TG(20:1_32:1)
Time
4
2
0
−2
0
12
24
Low
High
Time−interaction: Diagnostic group (MCI vs AD)
p = 1.712e−01
TG(20:1_32:1)
0.5
0.0
0
20
40
60
80
Time
MCI
AD
Multi−timepoint: Hippocampal volume
p = 1.900e−06
TG(20:1_32:1)
6
4
2
0
−2
Low
High
Low
High

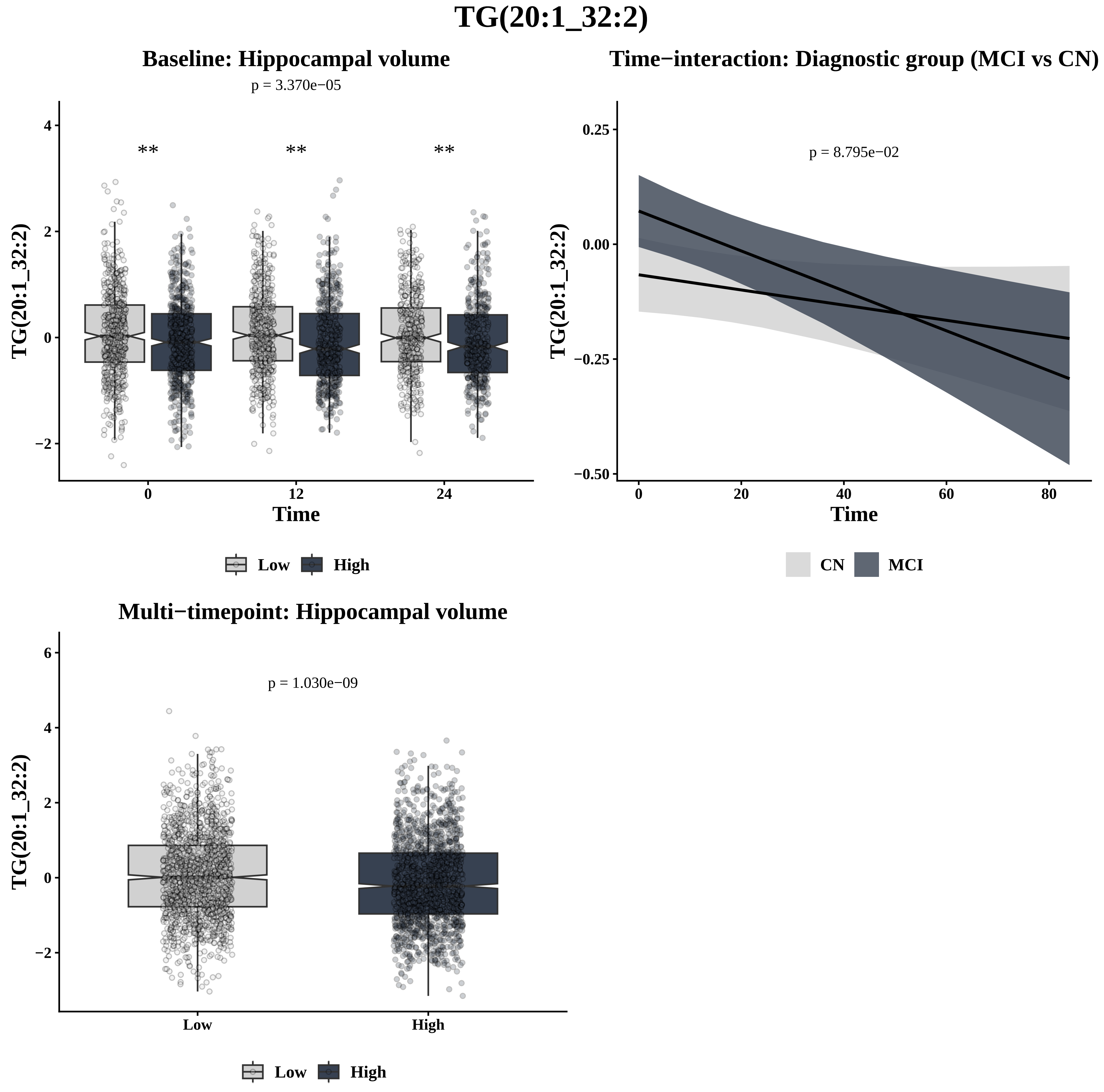

TG(20:1_32:2)
Baseline: Hippocampal volume
p = 3.370e−05
**
**
**
TG(20:1_32:2)
4
2
0
−2
0
12
24
Time
Low
High
Time−interaction: Diagnostic group (MCI vs CN)
p = 8.795e−02
TG(20:1_32:2)
0.25
0.00
−0.25
−0.50
0
20
40
60
80
Time
CN
MCI
Multi−timepoint: Hippocampal volume
p = 1.030e−09
TG(20:1_32:2)
6
4
2
0
−2
Low
High
Low
High

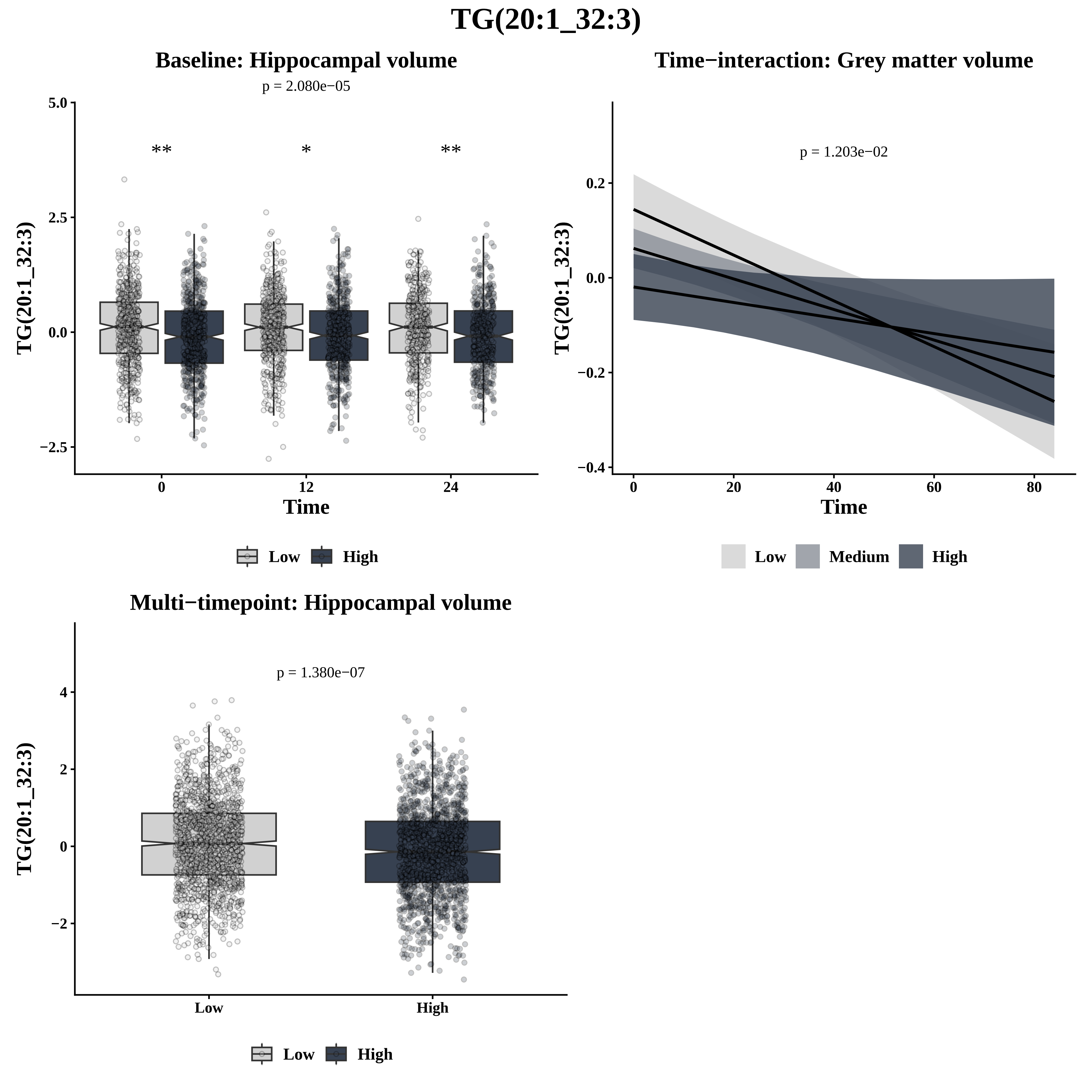
TG(20:1_32:3)
Baseline: Hippocampal volume
p = 2.080e−05
**
*
**
5.0
2.5
0.0
−2.5
TG(20:1_32:3)
0
12
24
Time
Low
High
Time−interaction: Grey matter volume
p = 1.203e−02
0.2
0.0
−0.2
−0.4
TG(20:1_32:3)
0
20
40
60
80
Time
Low
Medium
High
Multi−timepoint: Hippocampal volume
p = 1.380e−07
4
2
0
−2
TG(20:1_32:3)
Low
High
Low
High

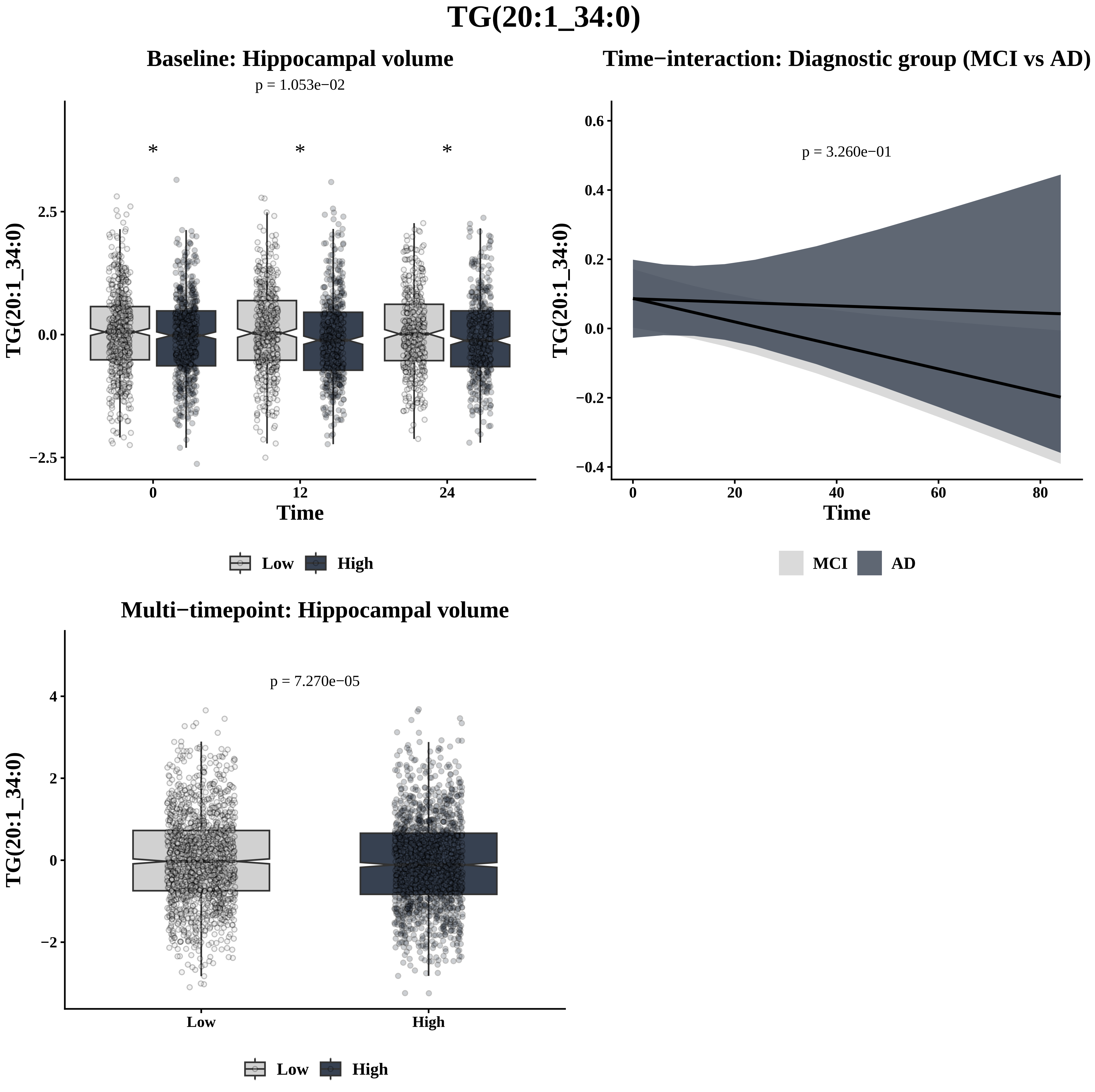

TG(20:1_34:0)
Baseline: Hippocampal volume
p = 1.053e−02
*
*
*
2.5
0.0
−2.5
TG(20:1_34:0)
0
12
24
Time
Low
High
Time−interaction: Diagnostic group (MCI vs AD)
p = 3.260e−01
0.6
0.4
0.2
0.0
−0.2
−0.4
TG(20:1_34:0)
0
20
40
60
80
Time
MCI
AD
Multi−timepoint: Hippocampal volume
p = 7.270e−05
4
2
0
−2
TG(20:1_34:0)
Low
High
Low
High

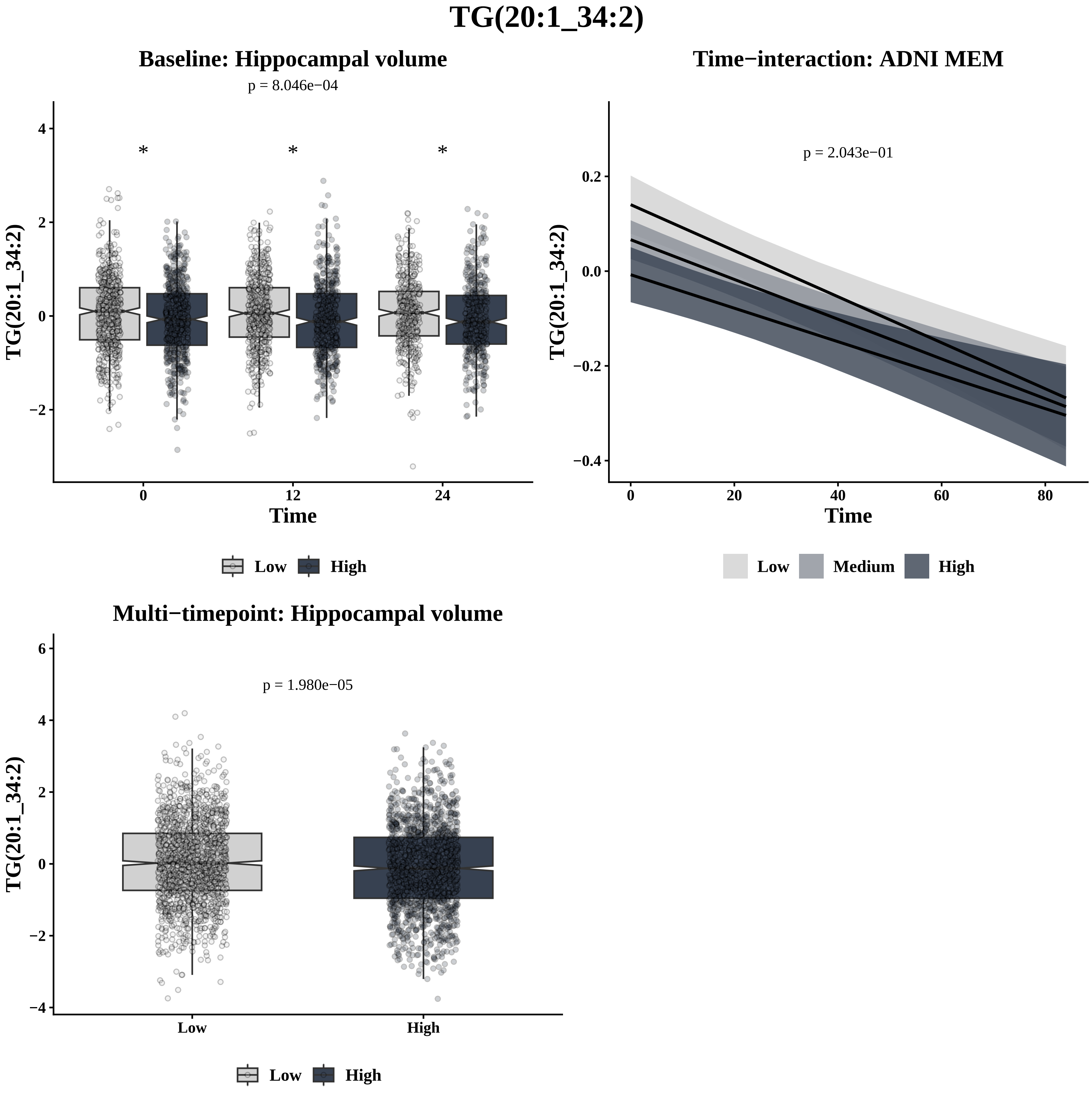
TG(20:1_34:2)
Baseline: Hippocampal volume
p = 8.046e−04
TG(20:1_34:2)
Time
Low
High
Time−interaction: ADNI MEM
p = 2.043e−01
TG(20:1_34:2)
Time
Low
Medium
High
Multi−timepoint: Hippocampal volume
p = 1.980e−05
TG(20:1_34:2)
Low
High

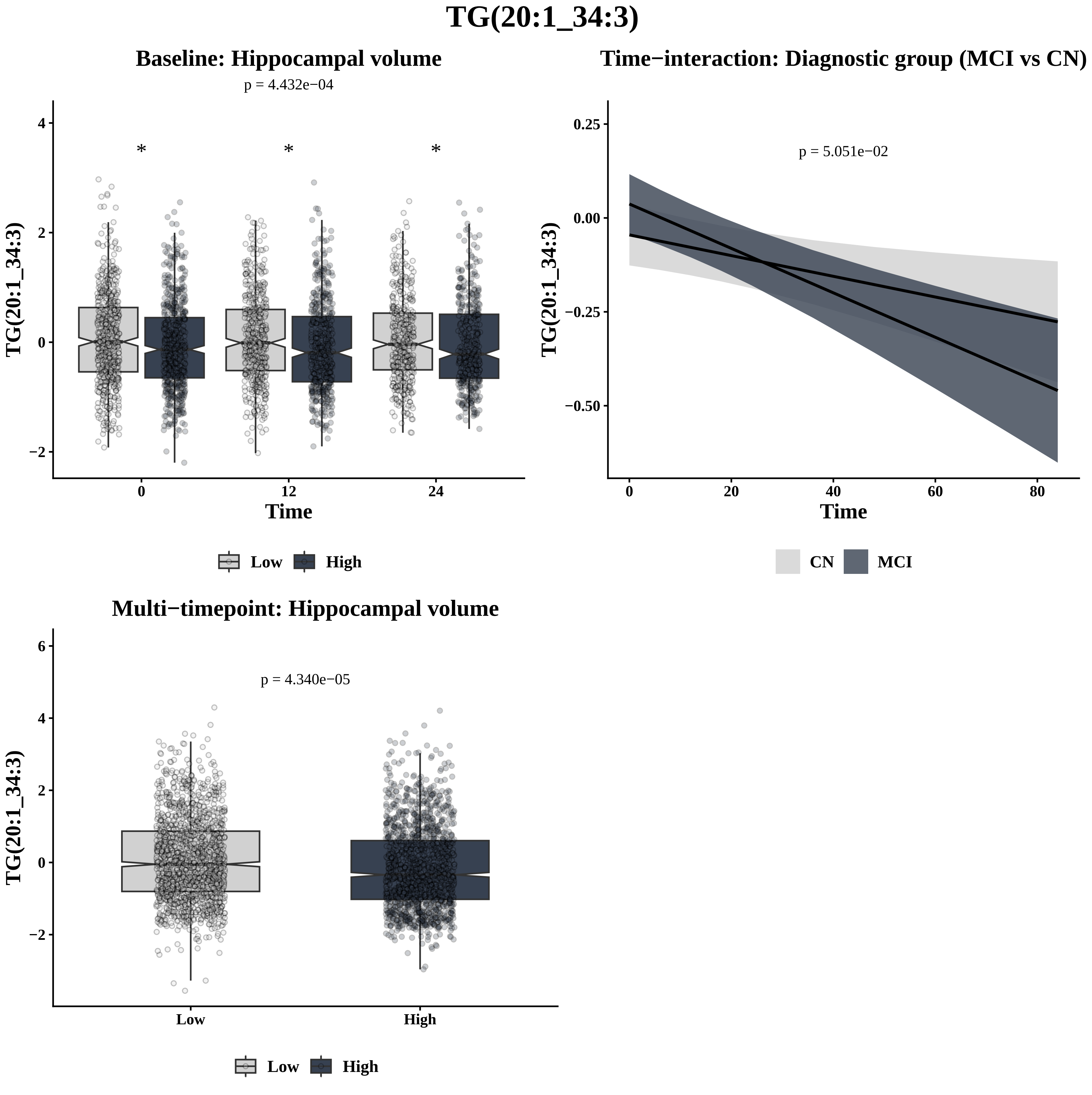

TG(20:1_34:3)
Baseline: Hippocampal volume
p = 4.432e−04
*
*
*
TG(20:1_34:3)
Time
0
12
24
4
2
0
−2
Low
High
Time−interaction: Diagnostic group (MCI vs CN)
p = 5.051e−02
TG(20:1_34:3)
0.25
0.00
−0.25
−0.50
0
20
40
60
80
Time
CN
MCI
Multi−timepoint: Hippocampal volume
p = 4.340e−05
TG(20:1_34:3)
6
4
2
0
−2
Low
High
Low
High

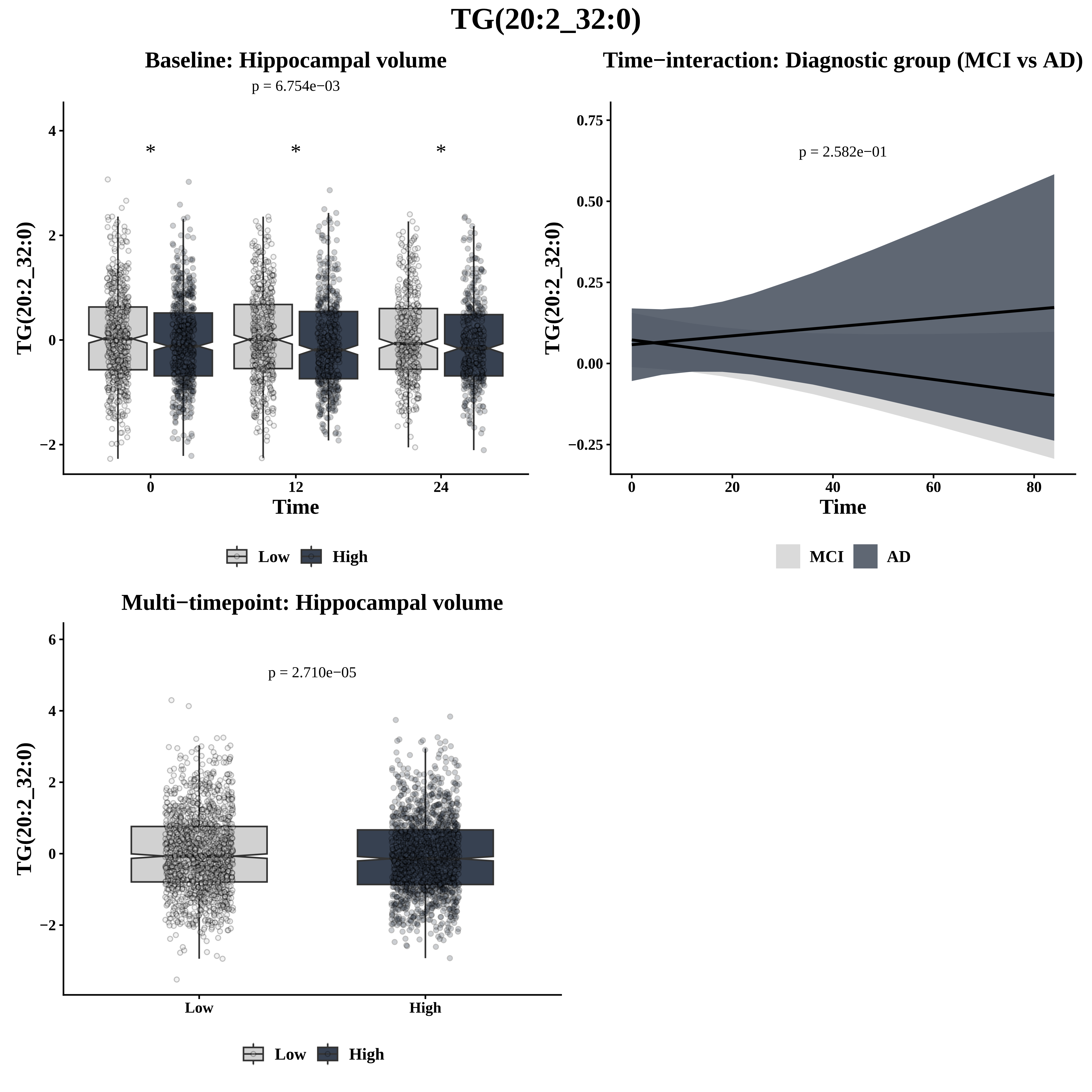

TG(20:2_32:0)
Baseline: Hippocampal volume
p = 6.754e−03
*
*
*
TG(20:2_32:0)
4
2
0
−2
0
12
24
Time
Low
High
Time−interaction: Diagnostic group (MCI vs AD)
p = 2.582e−01
TG(20:2_32:0)
0.75
0.50
0.25
0.00
−0.25
0
20
40
60
80
Time
MCI
AD
Multi−timepoint: Hippocampal volume
p = 2.710e−05
TG(20:2_32:0)
6
4
2
0
−2
Low
High
Low
High

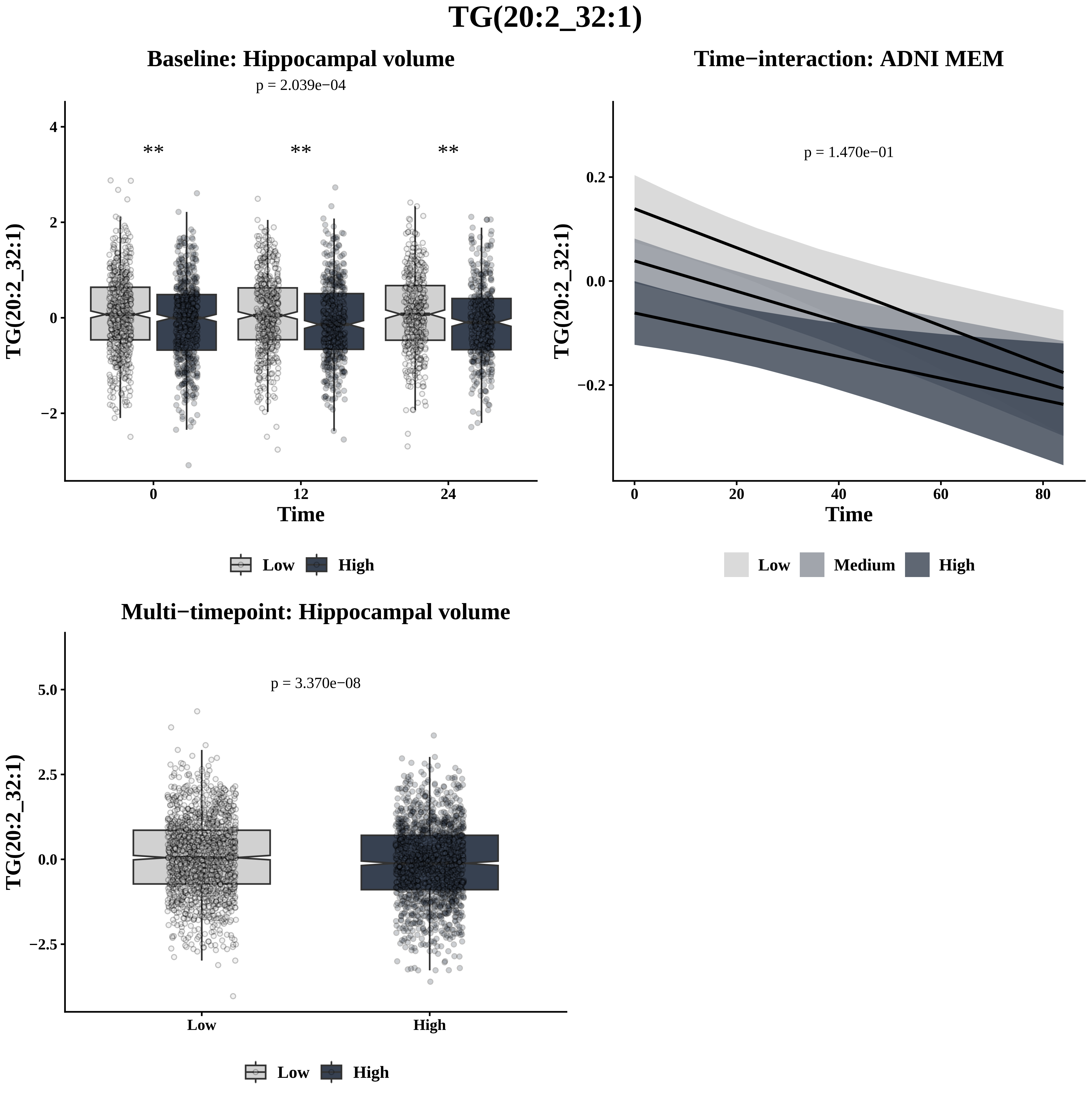
TG(20:2_32:1)
Baseline: Hippocampal volume
p = 2.039e−04
**
**
**
TG(20:2_32:1)
4
2
0
−2
0
12
24
Time
Low
High
Time−interaction: ADNI MEM
p = 1.470e−01
TG(20:2_32:1)
0.2
0.0
−0.2
0
20
40
60
80
Time
Low
Medium
High
Multi−timepoint: Hippocampal volume
p = 3.370e−08
TG(20:2_32:1)
5.0
2.5
0.0
−2.5
Low
High
Low
High

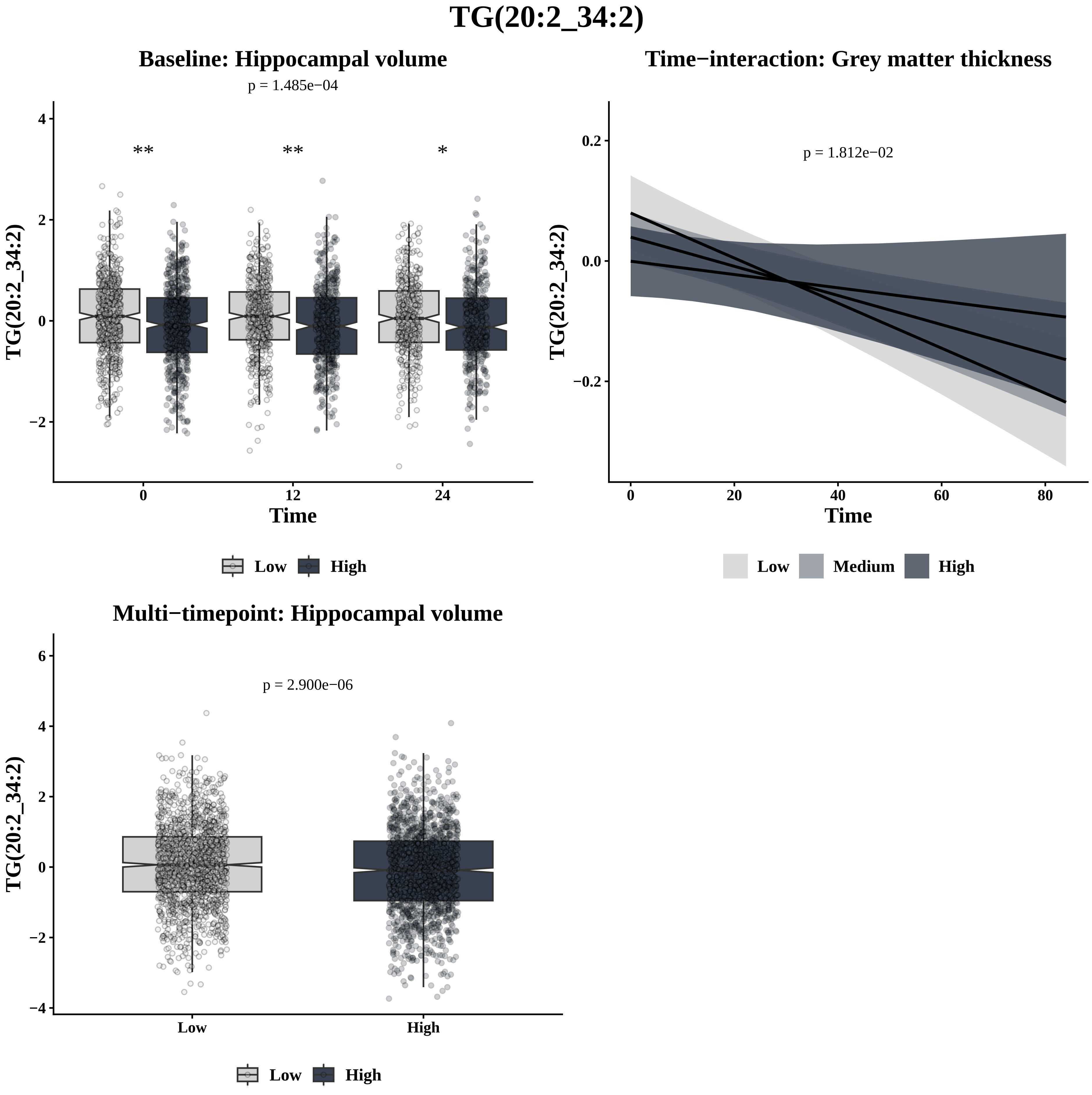
TG(20:2_34:2)
Baseline: Hippocampal volume
p = 1.485e−04
** ** *
TG(20:2_34:2)
Time
0 12 24
Low High
Time−interaction: Grey matter thickness
p = 1.812e−02
TG(20:2_34:2)
Time
0 20 40 60 80
Low Medium High
Multi−timepoint: Hippocampal volume
p = 2.900e−06
TG(20:2_34:2)
Low High
Low High

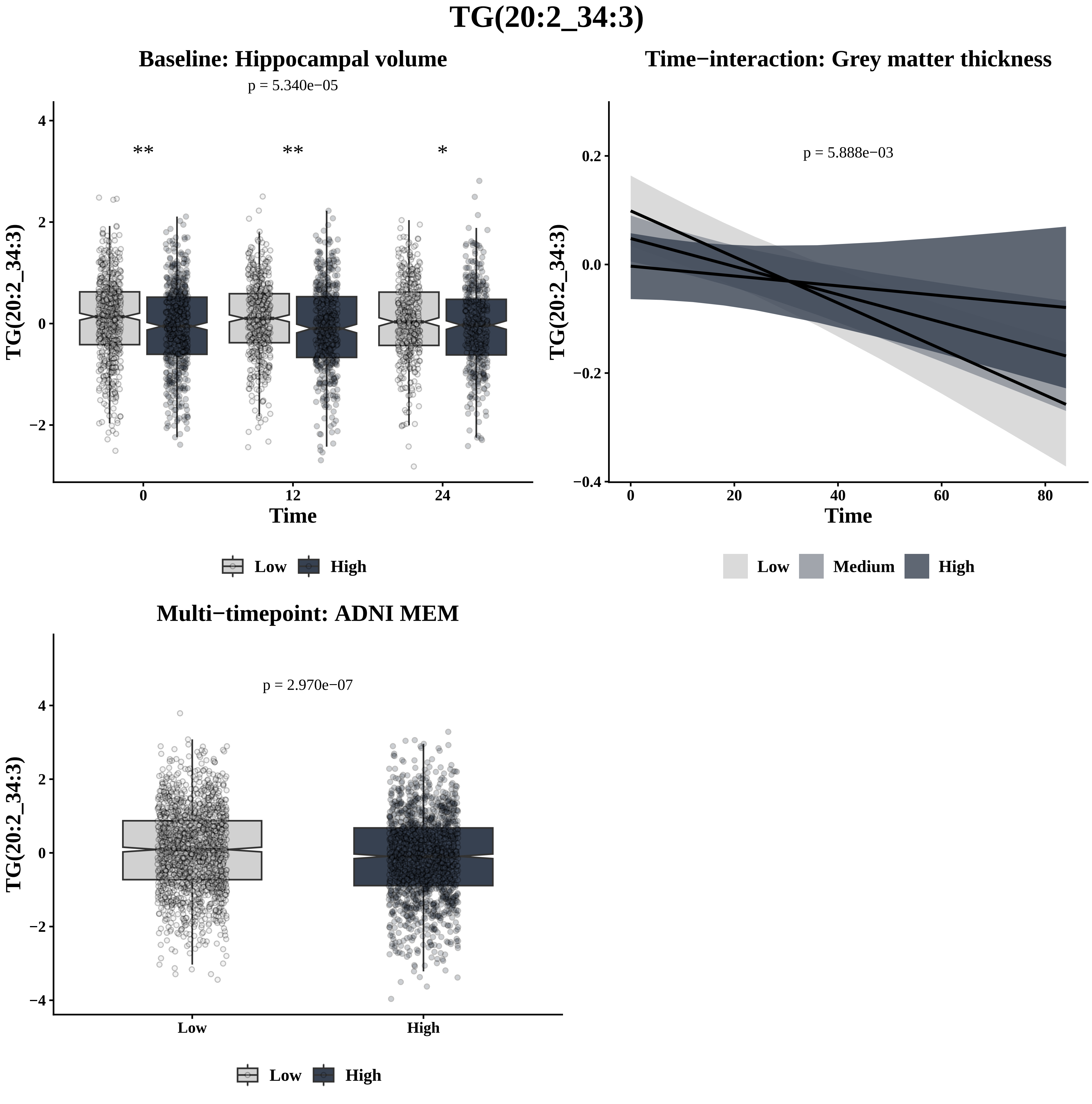
TG(20:2_34:3)
Baseline: Hippocampal volume
p = 5.340e−05
**
**
*
TG(20:2_34:3)
Time
0
12
24
Low
High
Time−interaction: Grey matter thickness
p = 5.888e−03
TG(20:2_34:3)
Time
Low
Medium
High
Multi−timepoint: ADNI MEM
p = 2.970e−07
TG(20:2_34:3)
Low
High
Low
High

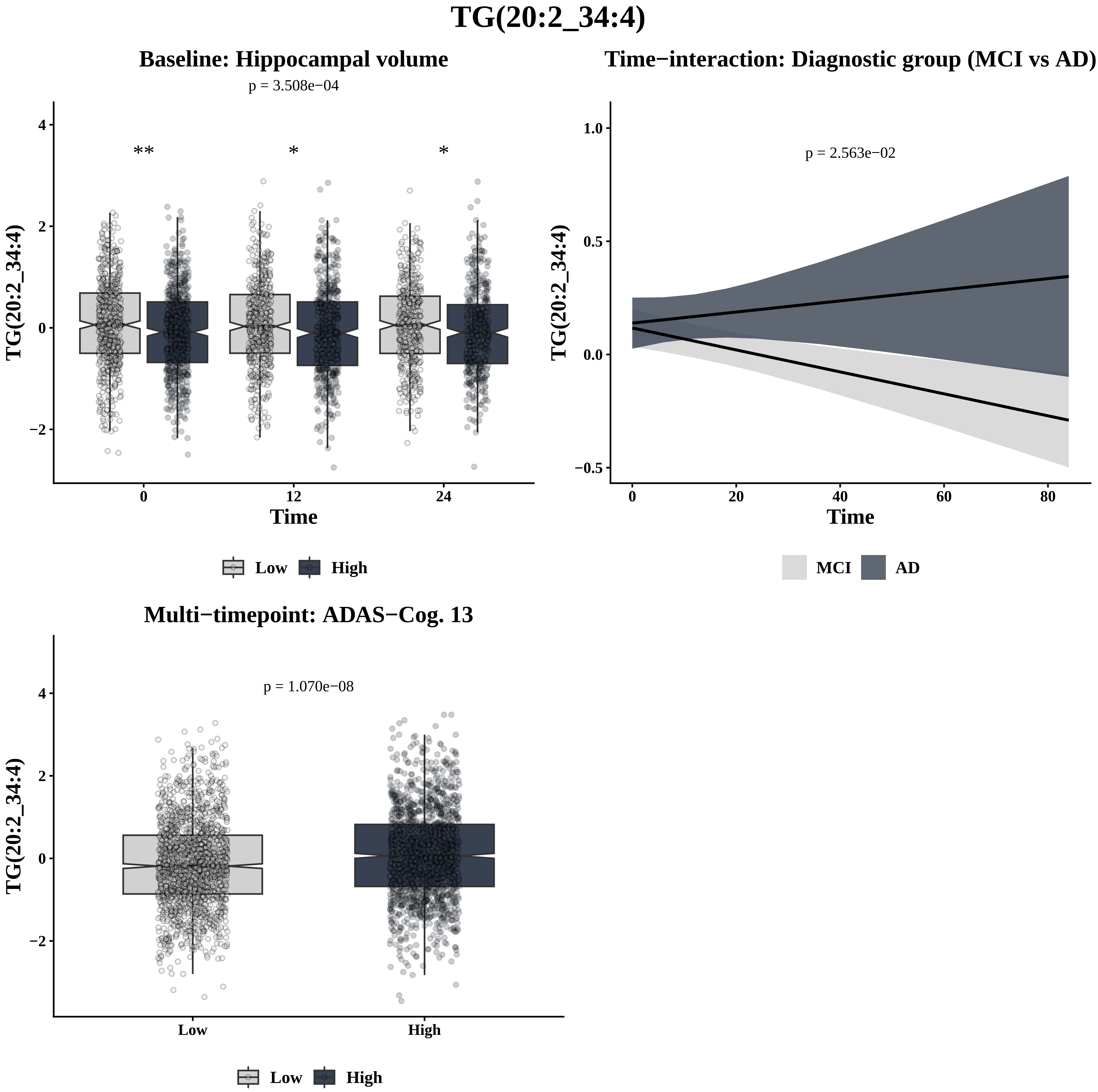

TG(20:2_34:4)
Baseline: Hippocampal volume
p = 3.508e−04
**
*
*
TG(20:2_34:4)
Time
0
12
24
Low
High
Time−interaction: Diagnostic group (MCI vs AD)
p = 2.563e−02
TG(20:2_34:4)
Time
MCI
AD
Multi−timepoint: ADAS−Cog. 13
p = 1.070e−08
TG(20:2_34:4)
Low
High

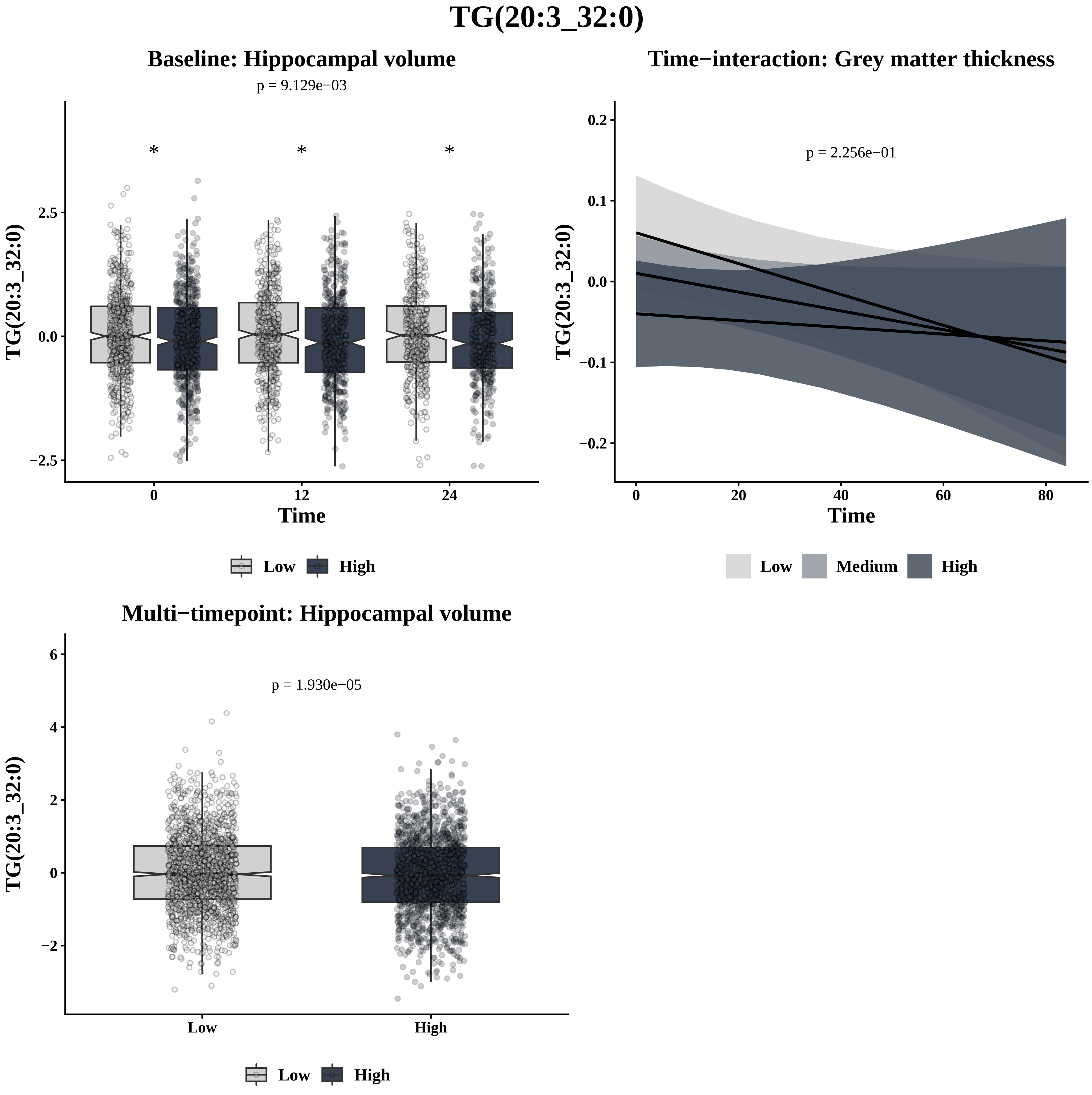

TG(20:3_32:0)
Baseline: Hippocampal volume
p = 9.129e−03
*
*
*
2.5
0.0
−2.5
TG(20:3_32:0)
0
12
24
Time
Low
High
Time−interaction: Grey matter thickness
p = 2.256e−01
0.2
0.1
0.0
−0.1
−0.2
TG(20:3_32:0)
0
20
40
60
80
Time
Low
Medium
High
Multi−timepoint: Hippocampal volume
p = 1.930e−05
6
4
2
0
−2
TG(20:3_32:0)
Low
High
Low
High

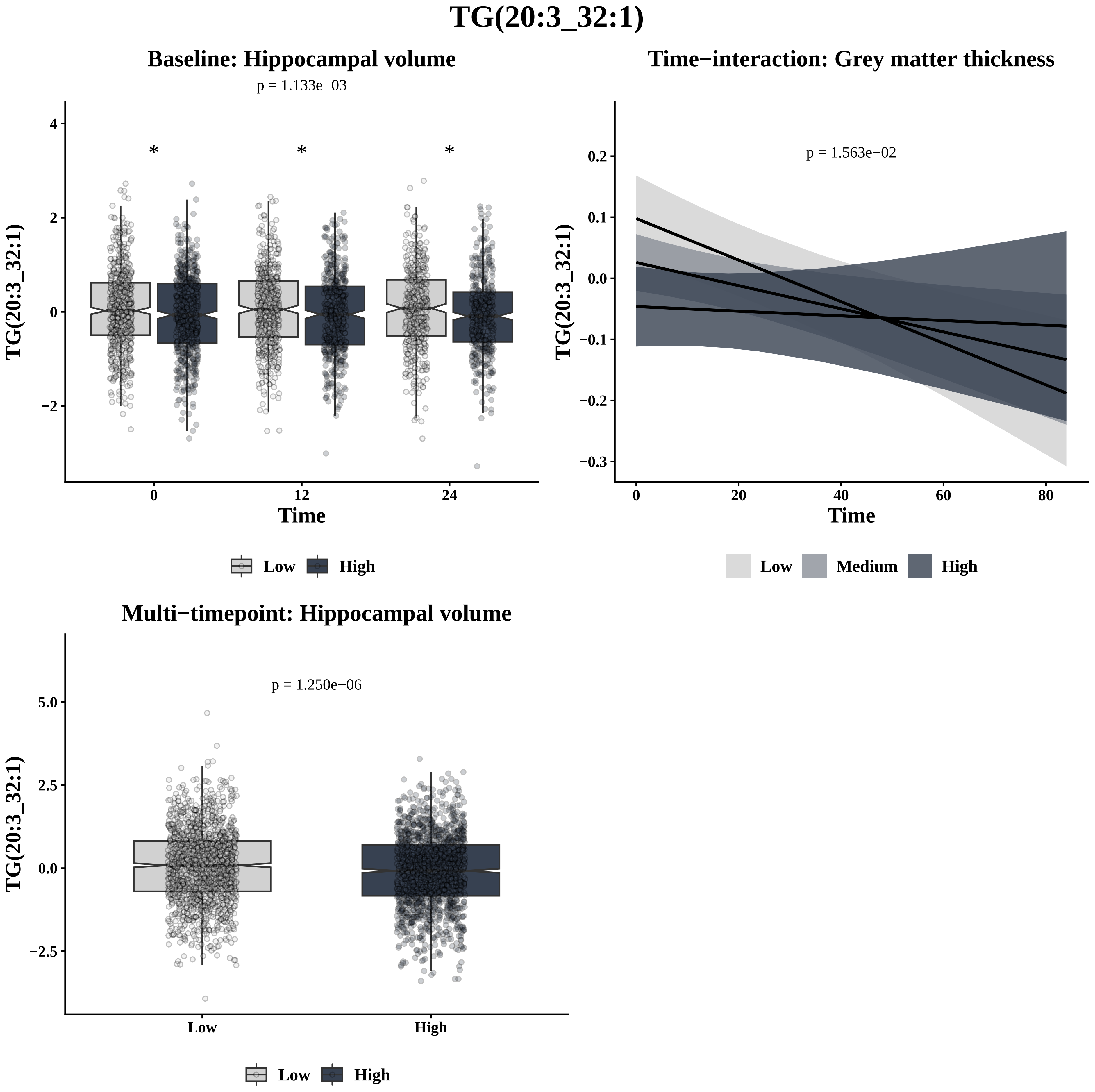

TG(20:3_32:1)
Baseline: Hippocampal volume
p = 1.133e−03
*
*
*
4
2
0
−2
TG(20:3_32:1)
0
12
24
Time
Low
High
Time−interaction: Grey matter thickness
p = 1.563e−02
0.2
0.1
0.0
−0.1
−0.2
−0.3
TG(20:3_32:1)
0
20
40
60
80
Time
Low
Medium
High
Multi−timepoint: Hippocampal volume
p = 1.250e−06
5.0
2.5
0.0
−2.5
TG(20:3_32:1)
Low
High
Low
High

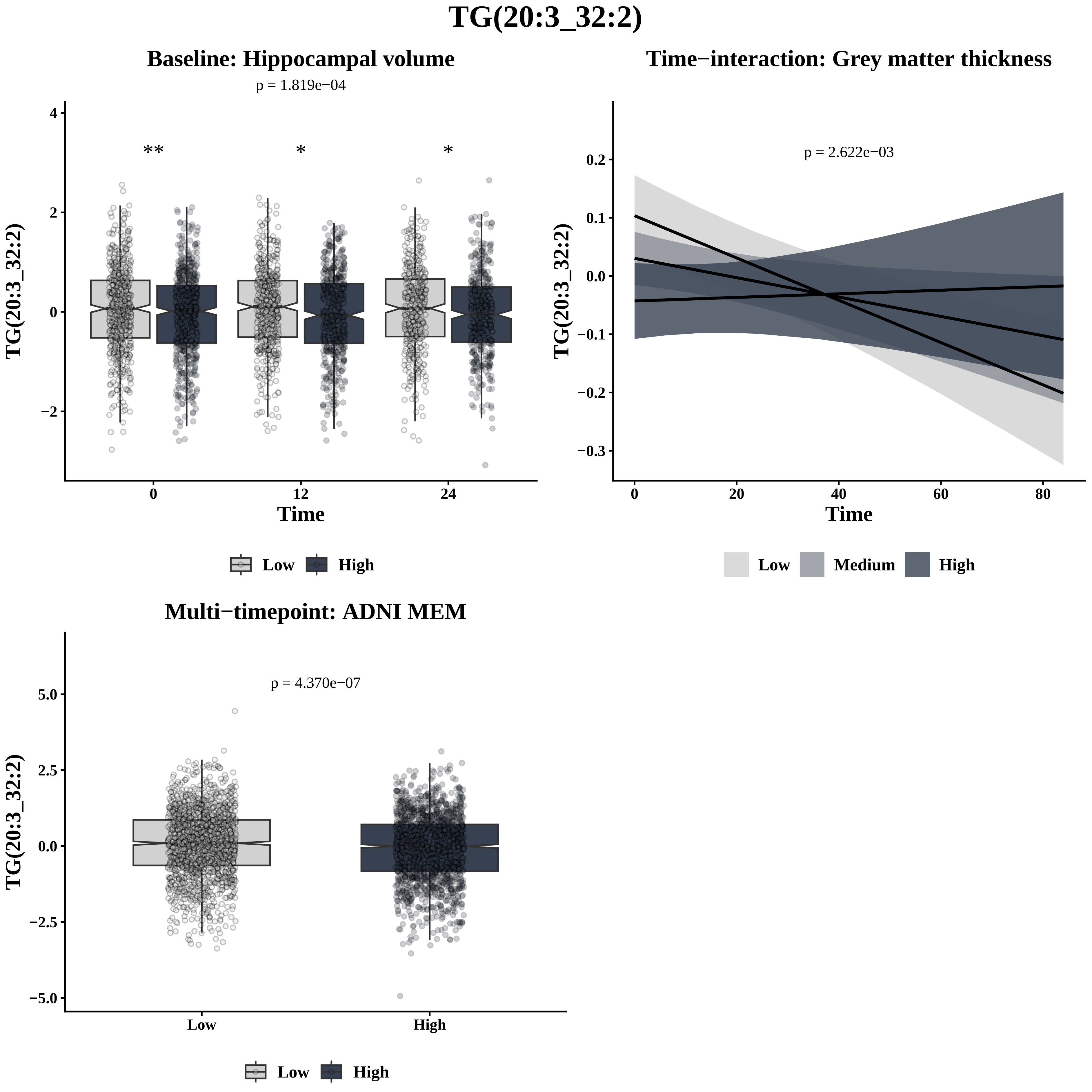

TG(20:3_32:2)
Baseline: Hippocampal volume
p = 1.819e−04
**
*
*
TG(20:3_32:2)
Time
Low
High
Time−interaction: Grey matter thickness
p = 2.622e−03
TG(20:3_32:2)
Time
Low
Medium
High
Multi−timepoint: ADNI MEM
p = 4.370e−07
TG(20:3_32:2)
Low
High

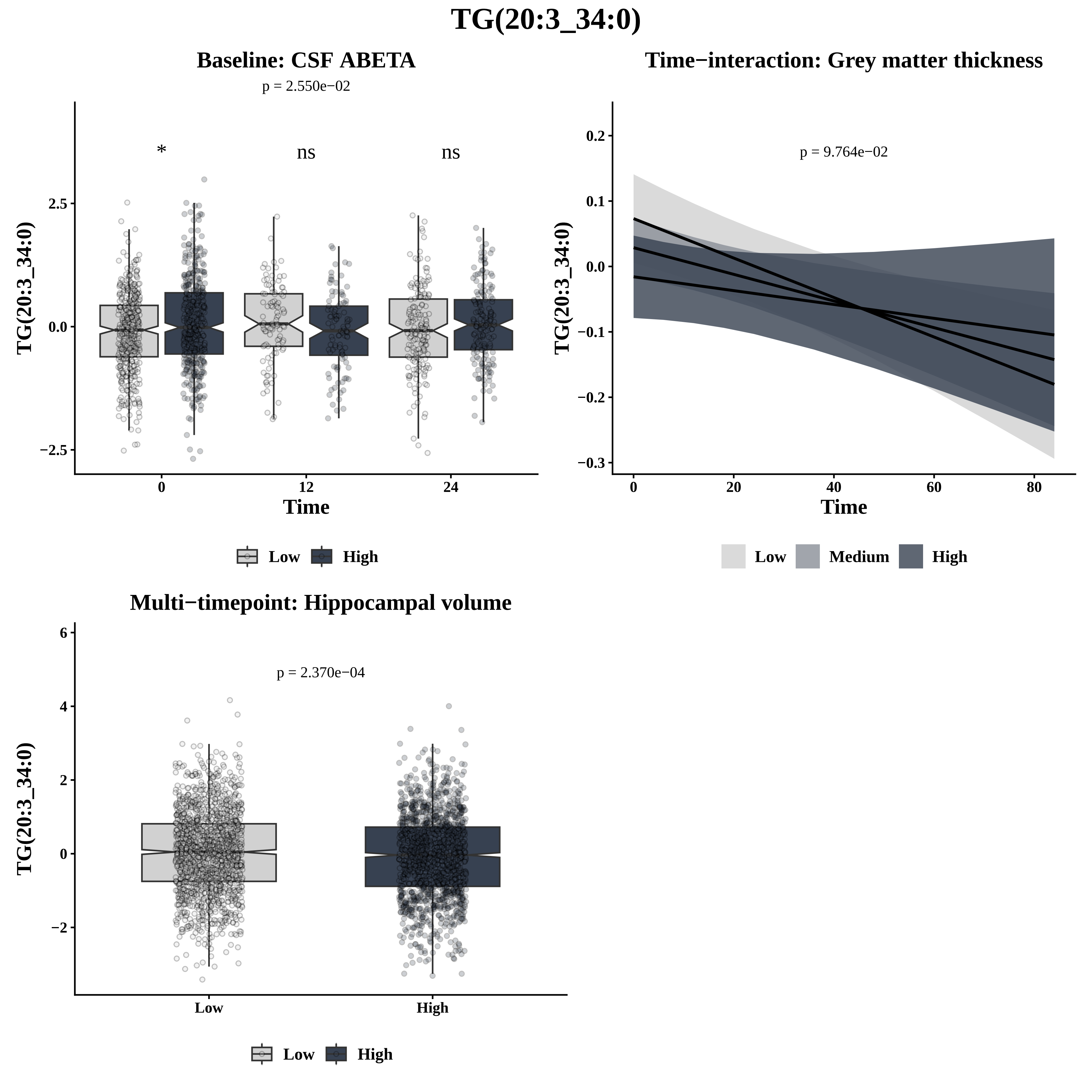
TG(20:3_34:0)
Baseline: CSF ABETA
p = 2.550e−02
*
ns
ns
TG(20:3_34:0)
2.5
0.0
−2.5
0
12
24
Time
Low
High
Time−interaction: Grey matter thickness
p = 9.764e−02
TG(20:3_34:0)
0.2
0.1
0.0
−0.1
−0.2
−0.3
0
20
40
60
80
Time
Low
Medium
High
Multi−timepoint: Hippocampal volume
p = 2.370e−04
TG(20:3_34:0)
6
4
2
0
−2
Low
High
Low
High

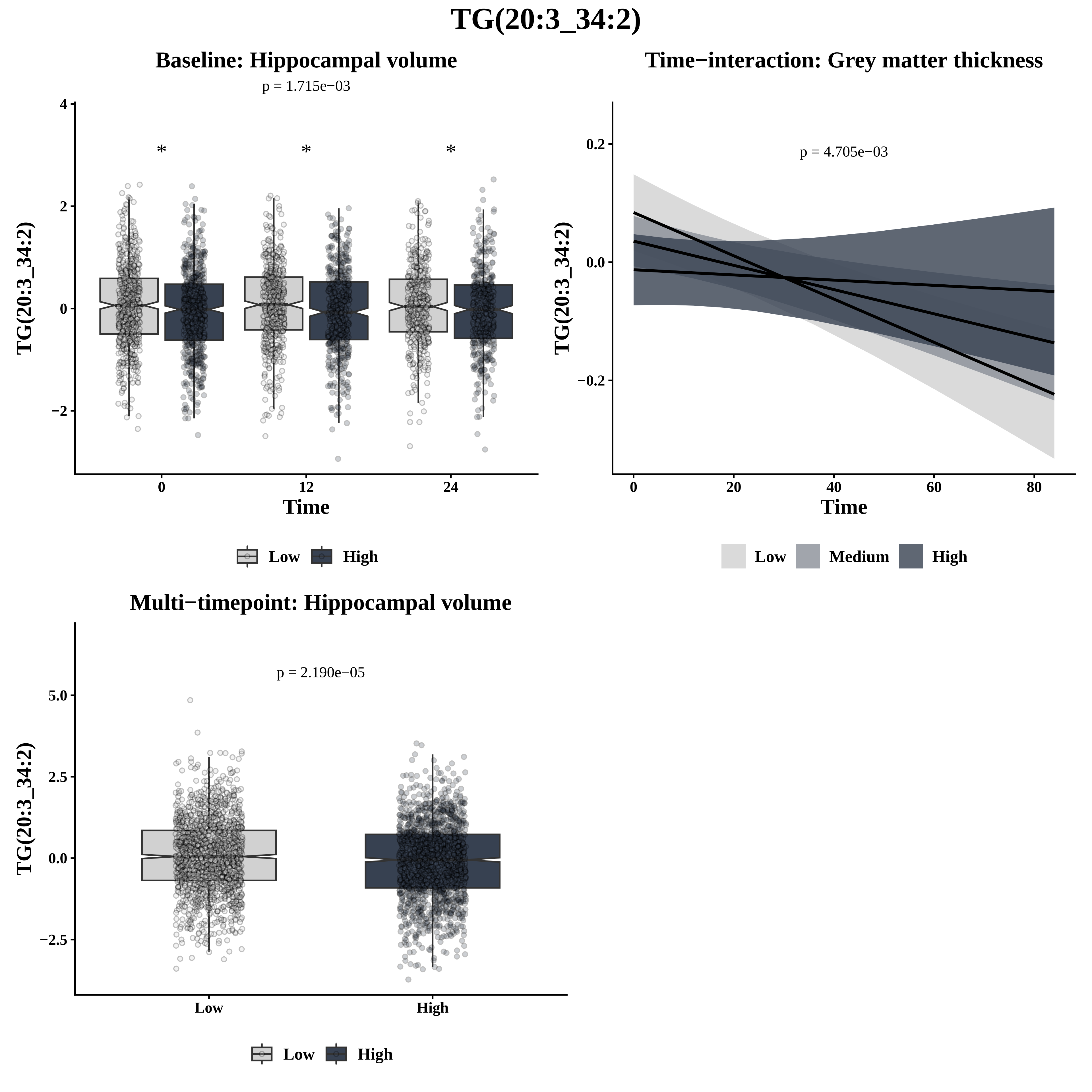

TG(20:3_34:2)
Baseline: Hippocampal volume
p = 1.715e−03
*
*
*
TG(20:3_34:2)
Time
0
12
24
Low
High
Time−interaction: Grey matter thickness
p = 4.705e−03
TG(20:3_34:2)
Time
0
20
40
60
80
Low
Medium
High
Multi−timepoint: Hippocampal volume
p = 2.190e−05
TG(20:3_34:2)
Low
High
Low
High

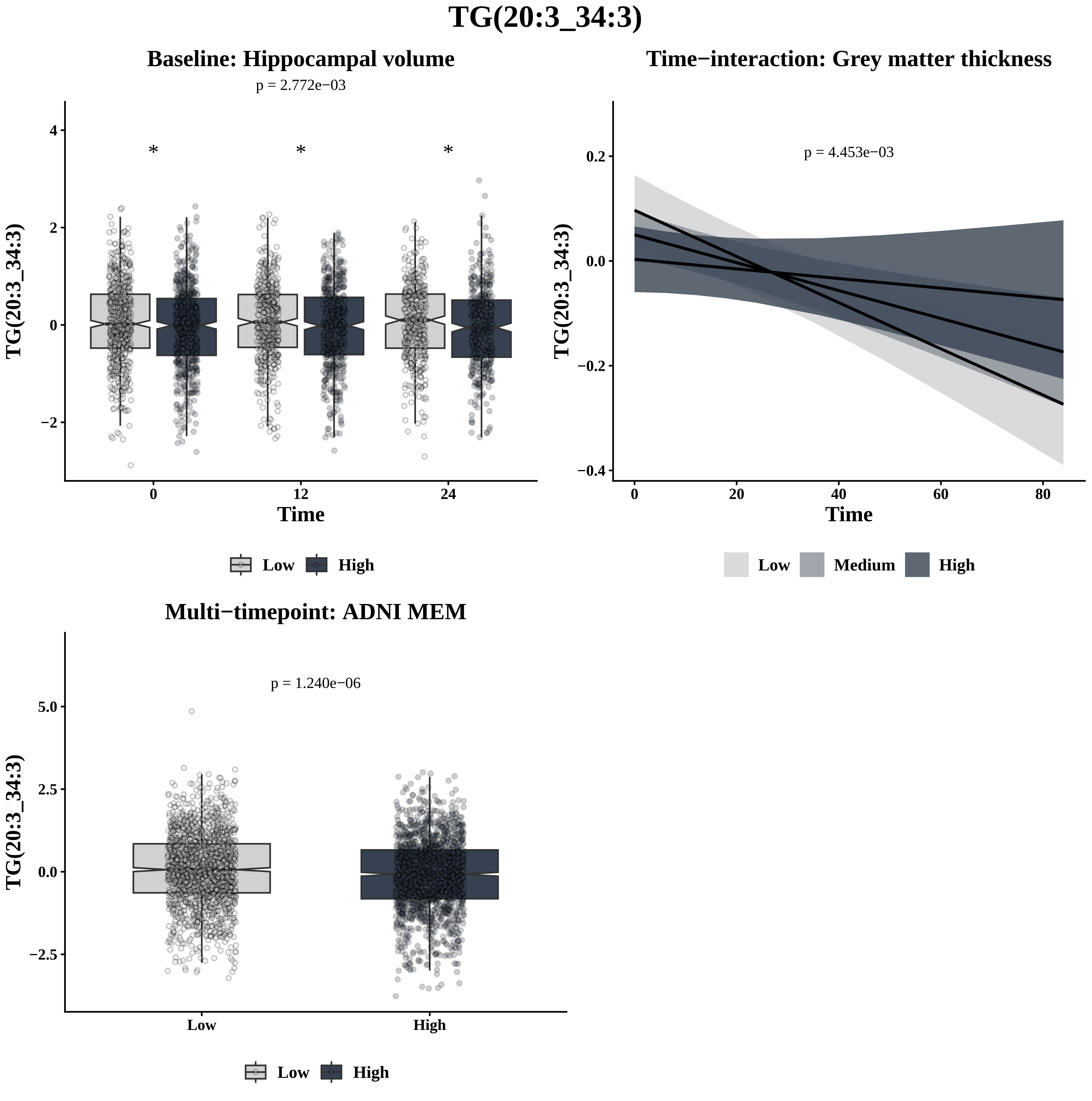

TG(20:3_34:3)
Baseline: Hippocampal volume
p = 2.772e−03
TG(20:3_34:3)
Time
Low
High
Time−interaction: Grey matter thickness
p = 4.453e−03
TG(20:3_34:3)
Time
Low
Medium
High
Multi−timepoint: ADNI MEM
p = 1.240e−06
TG(20:3_34:3)
Low
High

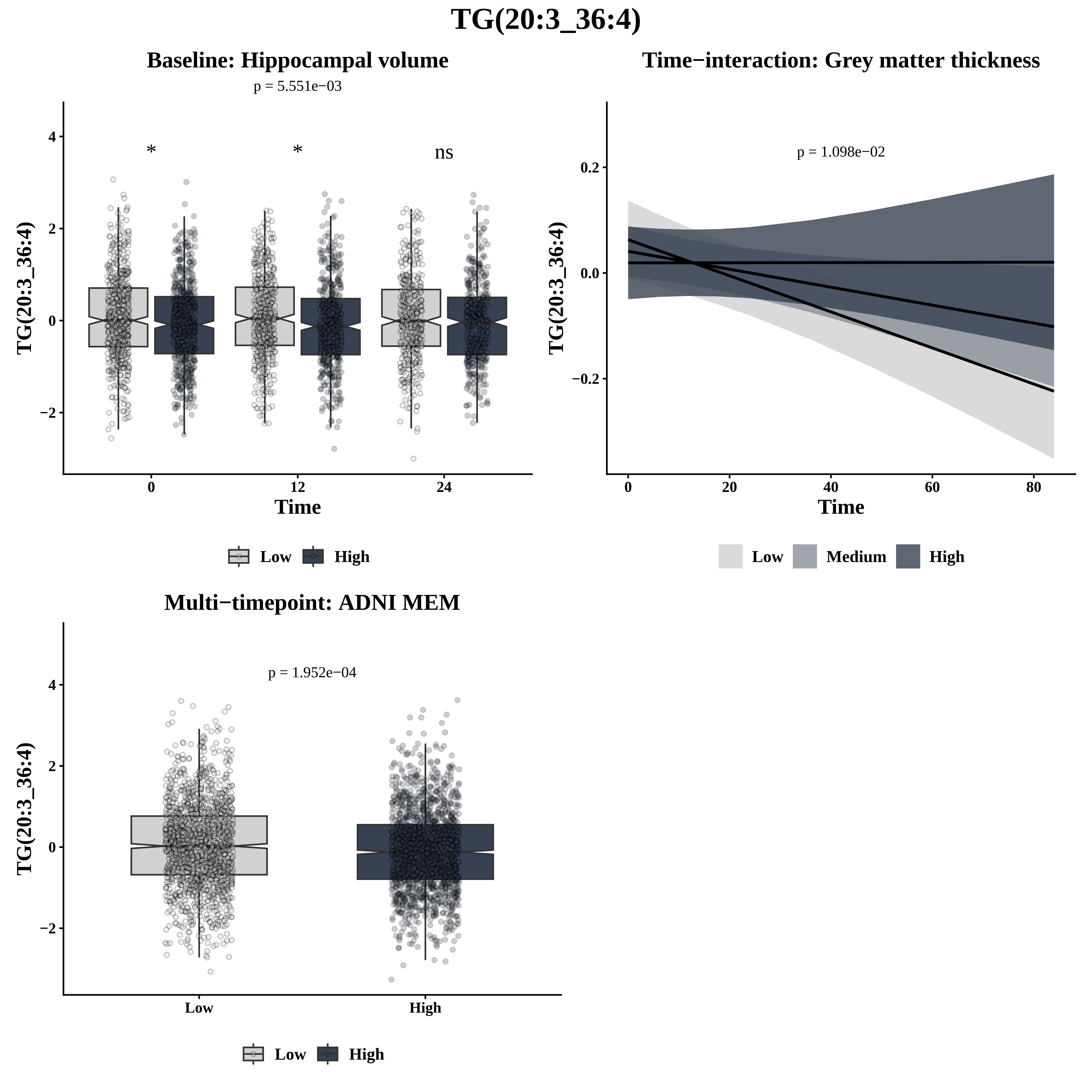
TG(20:3_36:4)
Baseline: Hippocampal volume
p = 5.551e−03
*
*
ns
TG(20:3_36:4)
Time
Low
High
Time−interaction: Grey matter thickness
p = 1.098e−02
TG(20:3_36:4)
Time
Low
Medium
High
Multi−timepoint: ADNI MEM
p = 1.952e−04
TG(20:3_36:4)
Low
High
Low
High

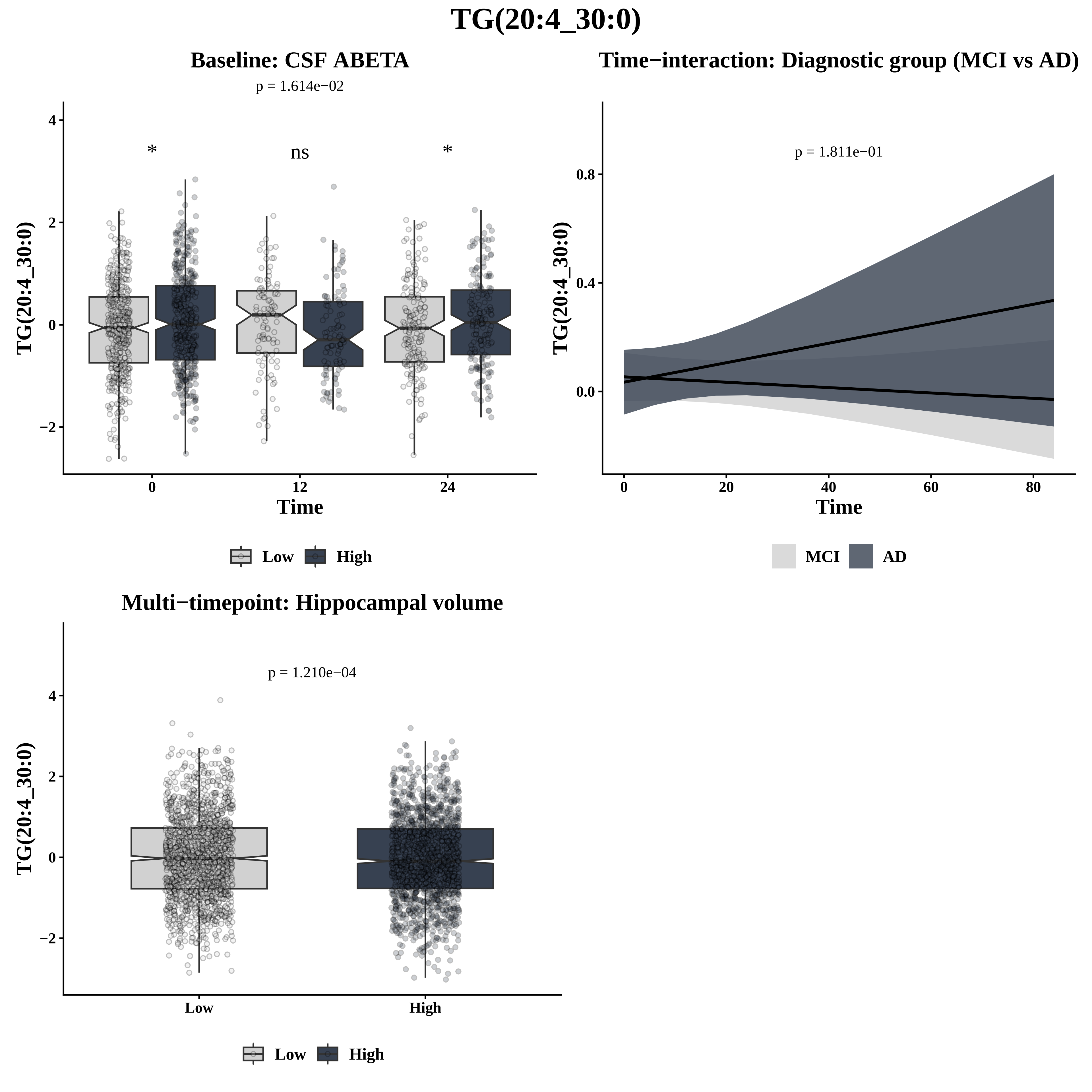

TG(20:4_30:0)
Baseline: CSF ABETA
p = 1.614e−02
*
ns
*
TG(20:4_30:0)
Time
0
12
24
Low
High
Time−interaction: Diagnostic group (MCI vs AD)
p = 1.811e−01
TG(20:4_30:0)
Time
MCI
AD
Multi−timepoint: Hippocampal volume
p = 1.210e−04
TG(20:4_30:0)
Low
High

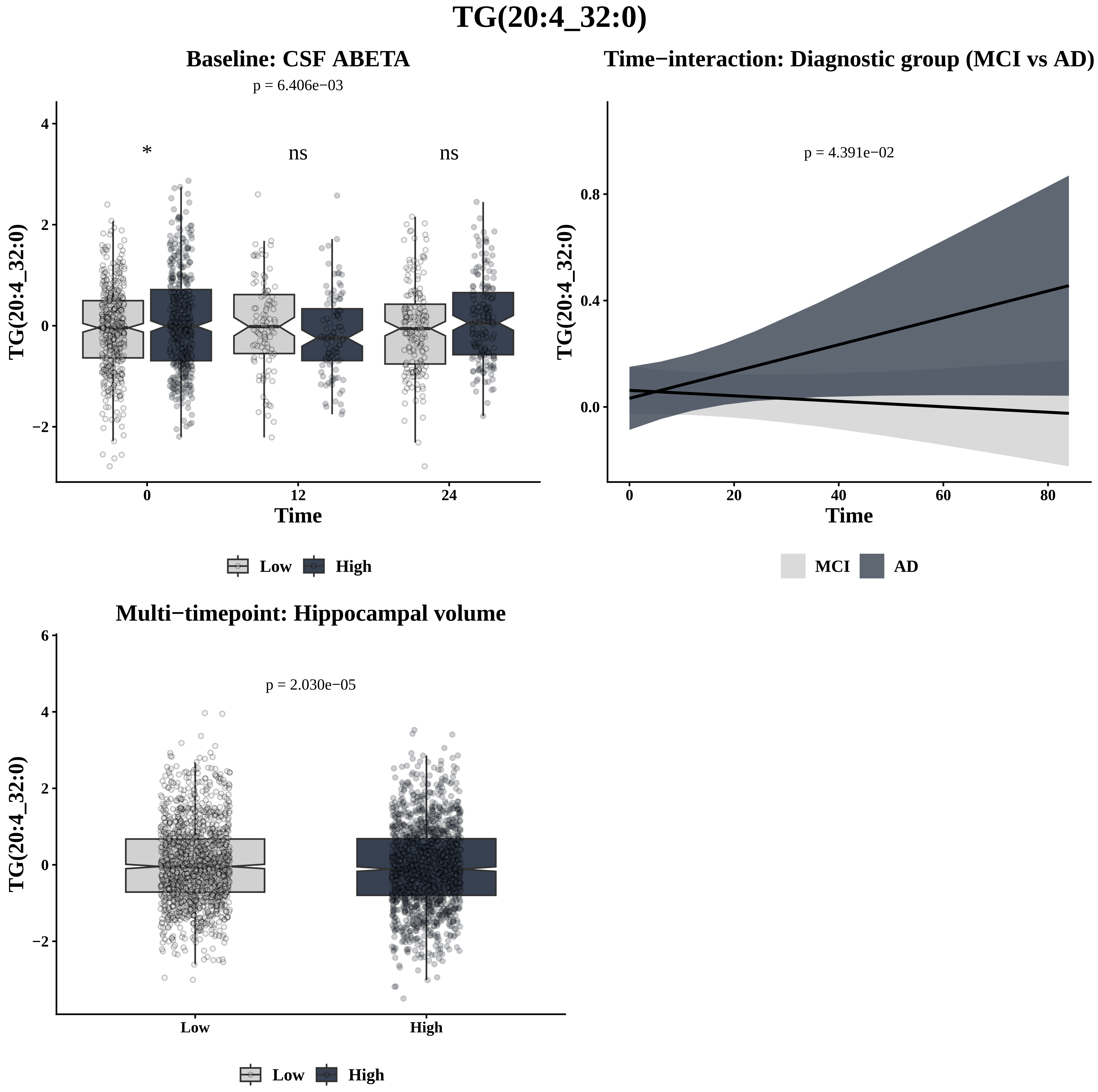

TG(20:4_32:0)
Baseline: CSF ABETA
p = 6.406e−03
*
ns
ns
TG(20:4_32:0)
4
2
0
−2
0
12
24
Time
Low
High
Time−interaction: Diagnostic group (MCI vs AD)
p = 4.391e−02
TG(20:4_32:0)
0.8
0.4
0.0
0
20
40
60
80
Time
MCI
AD
Multi−timepoint: Hippocampal volume
p = 2.030e−05
TG(20:4_32:0)
6
4
2
0
−2
Low
High
Low
High

# TG(20:4_32:1)

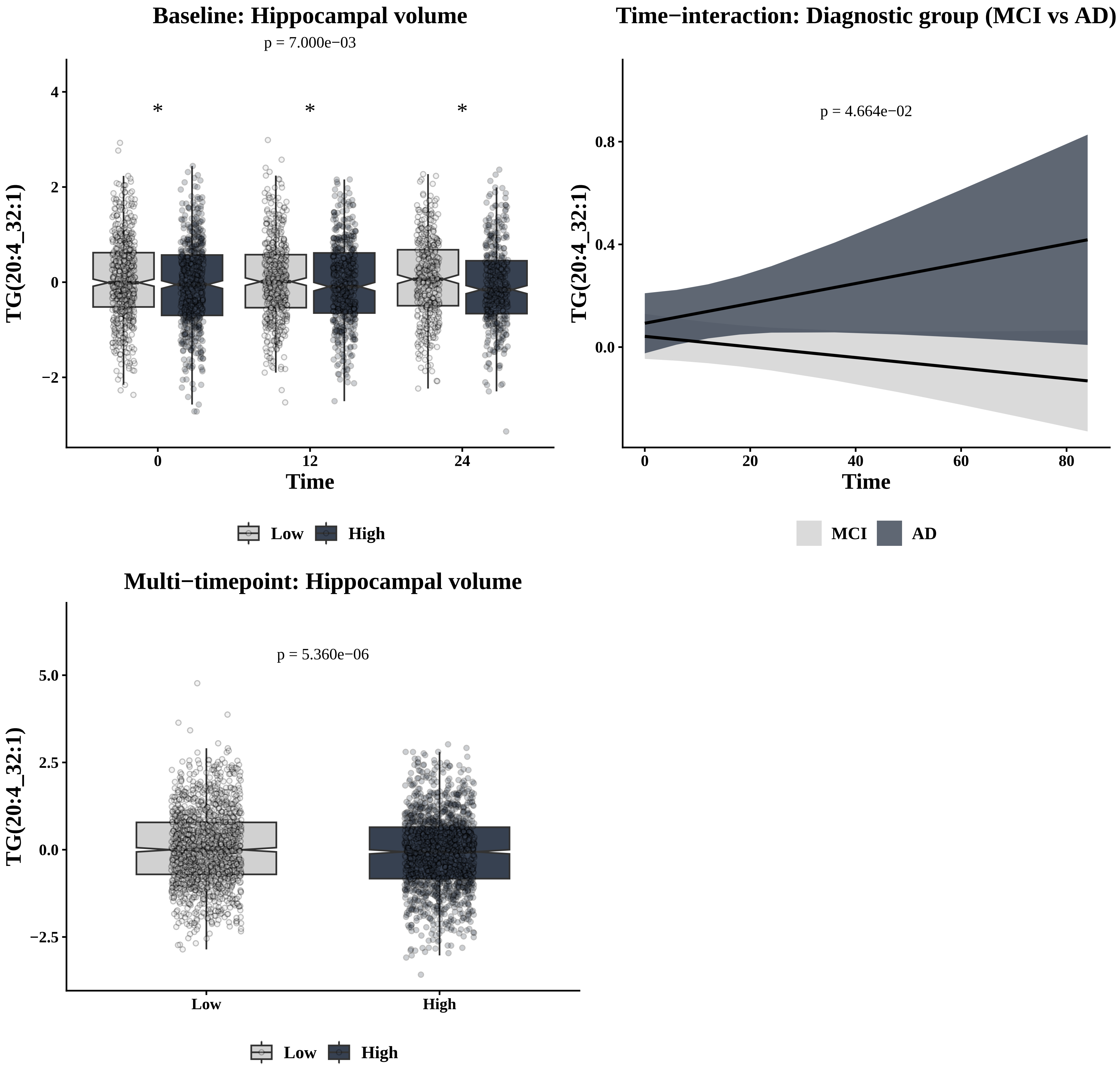
Baseline: Hippocampal volume
p = 7.000e−03
*
*
*
4
2
0
−2
0
12
24
Time
TG(20:4_32:1)
Low
High
Time−interaction: Diagnostic group (MCI vs AD)
p = 4.664e−02
0.8
0.4
0.0
0
20
40
60
80
Time
TG(20:4_32:1)
MCI
AD
Multi−timepoint: Hippocampal volume
p = 5.360e−06
5.0
2.5
0.0
−2.5
Low
High
TG(20:4_32:1)
Low
High

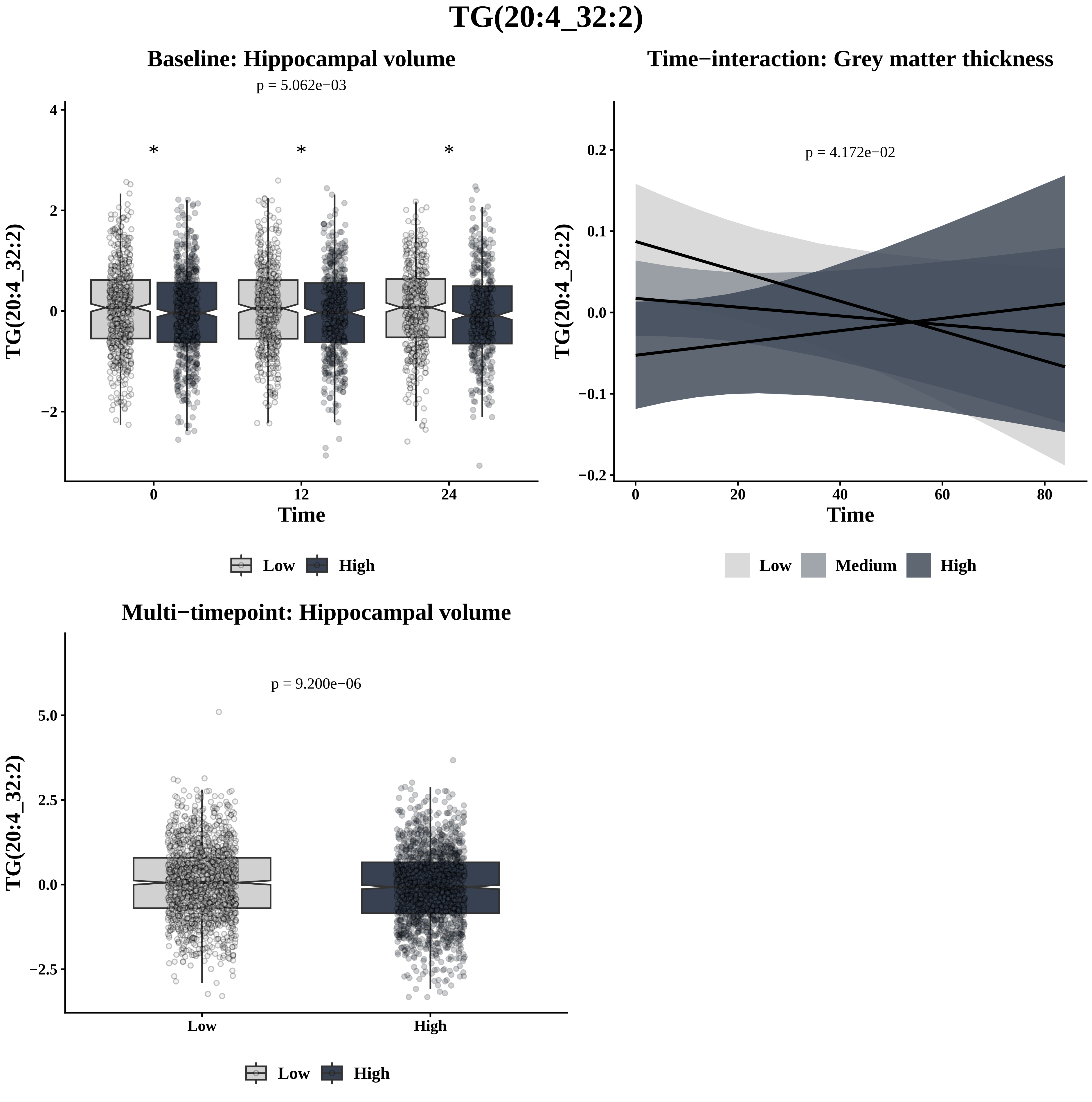
TG(20:4_32:2)
Baseline: Hippocampal volume
p = 5.062e−03
*
*
*
TG(20:4_32:2)
4
2
0
−2
0
12
24
Time
Low
High
Time−interaction: Grey matter thickness
p = 4.172e−02
TG(20:4_32:2)
0.2
0.1
0.0
−0.1
−0.2
0
20
40
60
80
Time
Low
Medium
High
Multi−timepoint: Hippocampal volume
p = 9.200e−06
TG(20:4_32:2)
5.0
2.5
0.0
−2.5
Low
High
Low
High

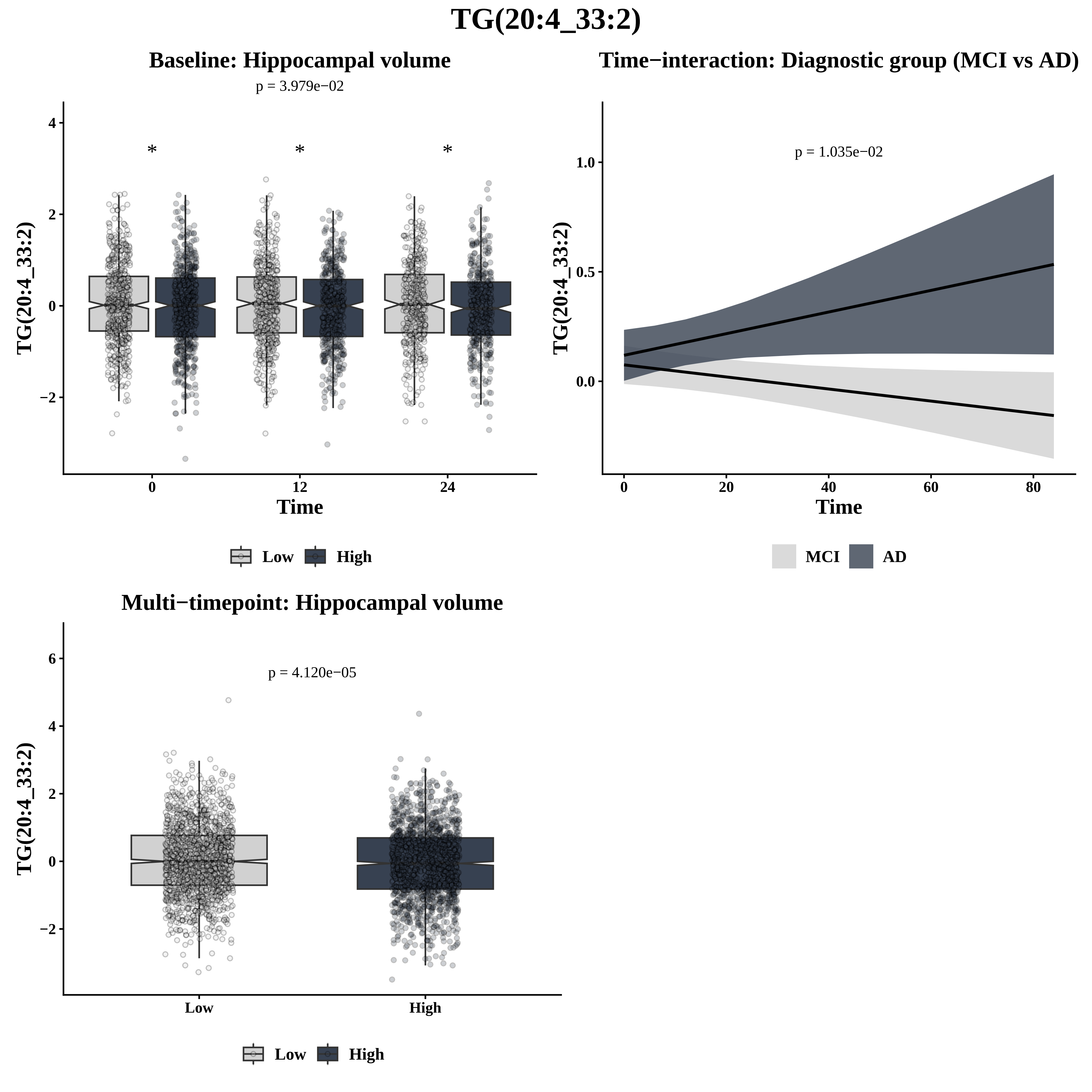

TG(20:4_33:2)
Baseline: Hippocampal volume
p = 3.979e−02
*
*
*
TG(20:4_33:2)
4
2
0
−2
0
12
24
Time
Low
High
Time−interaction: Diagnostic group (MCI vs AD)
p = 1.035e−02
TG(20:4_33:2)
1.0
0.5
0.0
0
20
40
60
80
Time
MCI
AD
Multi−timepoint: Hippocampal volume
p = 4.120e−05
TG(20:4_33:2)
6
4
2
0
−2
Low
High
Low
High

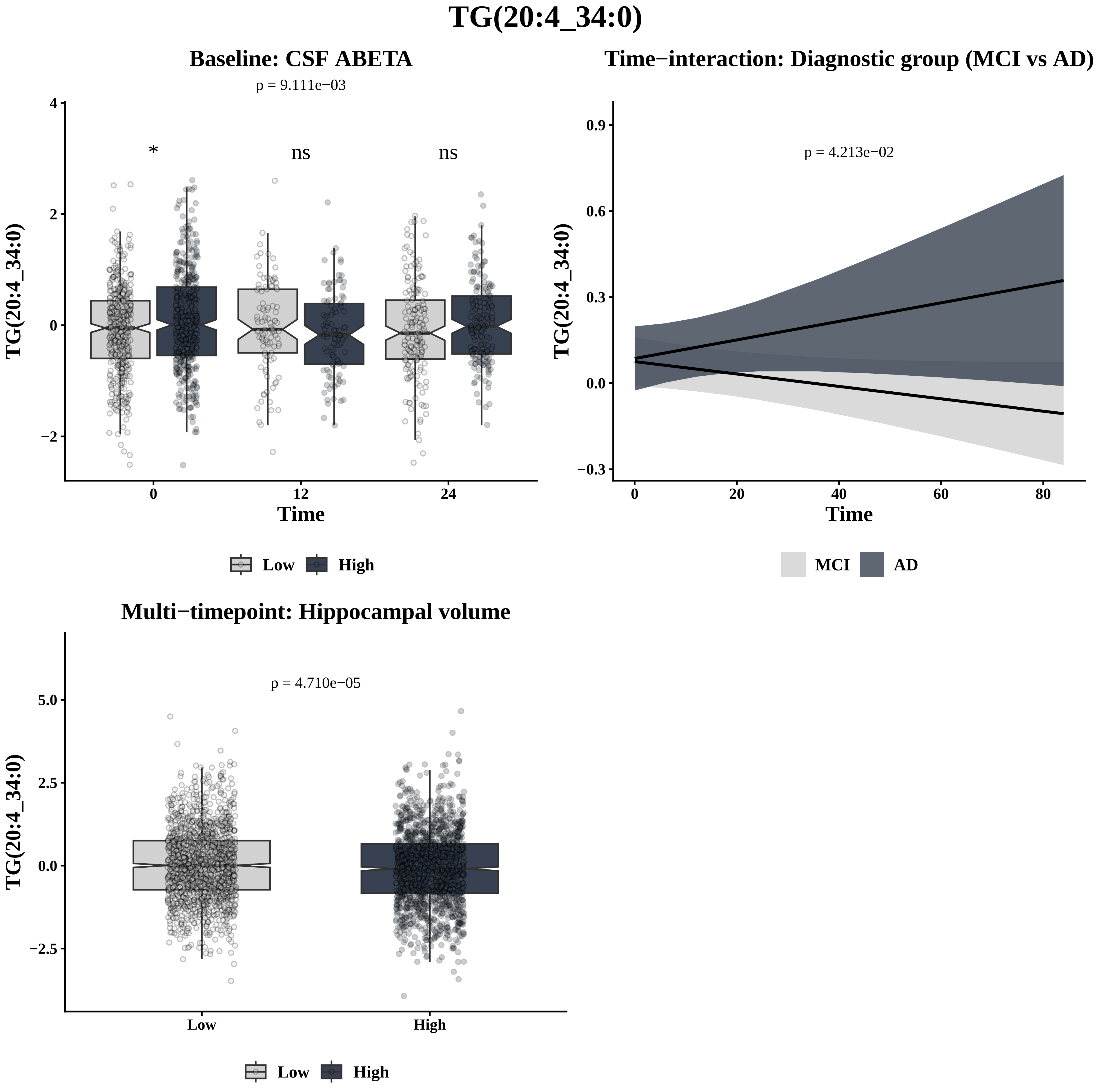

TG(20:4_34:0)
Baseline: CSF ABETA
p = 9.111e−03
*
ns
ns
TG(20:4_34:0)
Time
0
12
24
4
2
0
−2
Low
High
Time−interaction: Diagnostic group (MCI vs AD)
p = 4.213e−02
TG(20:4_34:0)
Time
0.9
0.6
0.3
0.0
−0.3
0
20
40
60
80
MCI
AD
Multi−timepoint: Hippocampal volume
p = 4.710e−05
TG(20:4_34:0)
5.0
2.5
0.0
−2.5
Low
High
Low
High

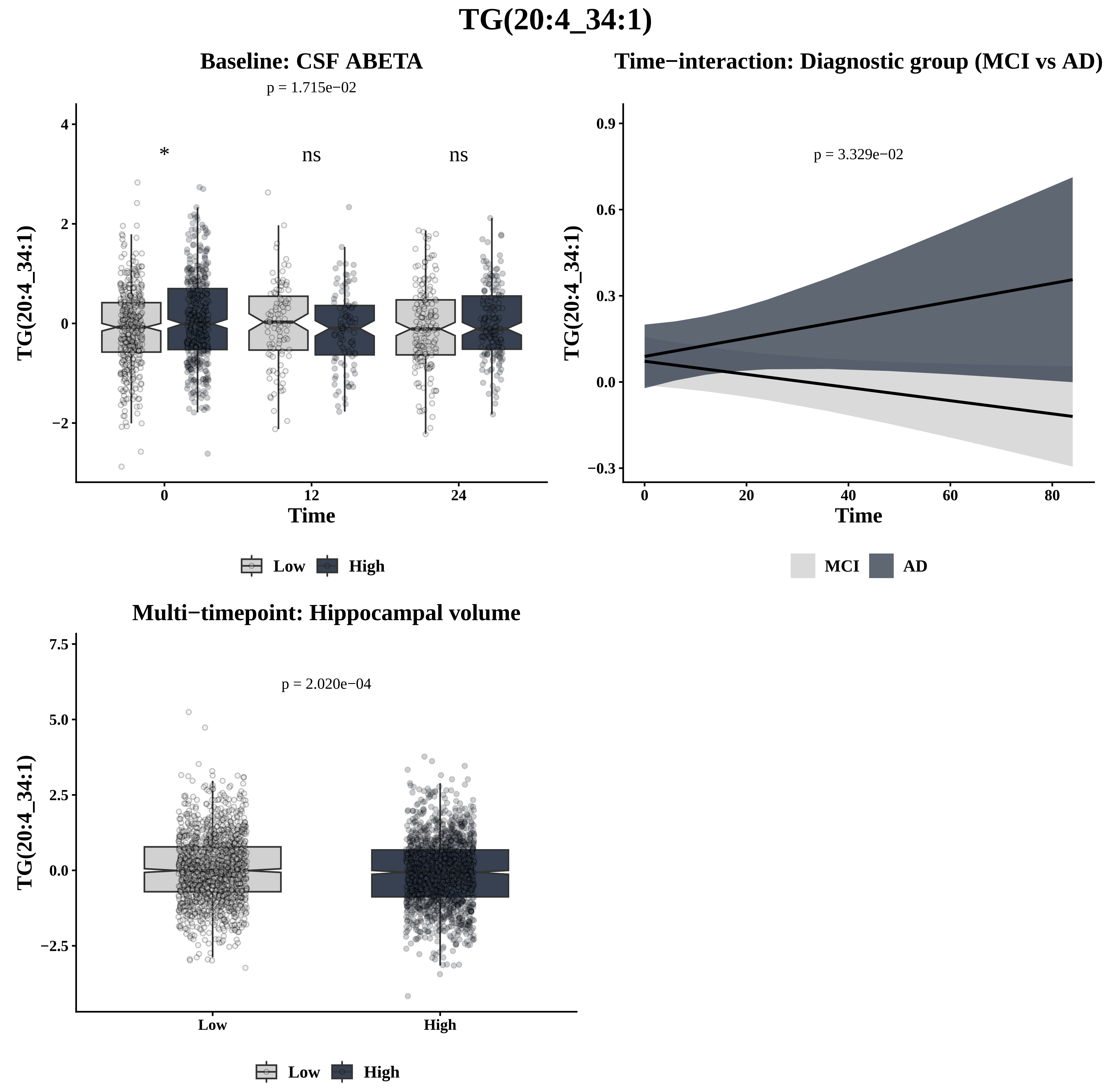

TG(20:4_34:1)
Baseline: CSF ABETA
p = 1.715e−02
*
ns
ns
TG(20:4_34:1)
Time
0
12
24
4
2
0
−2
Low
High
Time−interaction: Diagnostic group (MCI vs AD)
p = 3.329e−02
TG(20:4_34:1)
Time
0.9
0.6
0.3
0.0
−0.3
0
20
40
60
80
MCI
AD
Multi−timepoint: Hippocampal volume
p = 2.020e−04
TG(20:4_34:1)
7.5
5.0
2.5
0.0
−2.5
Low
High
Low
High

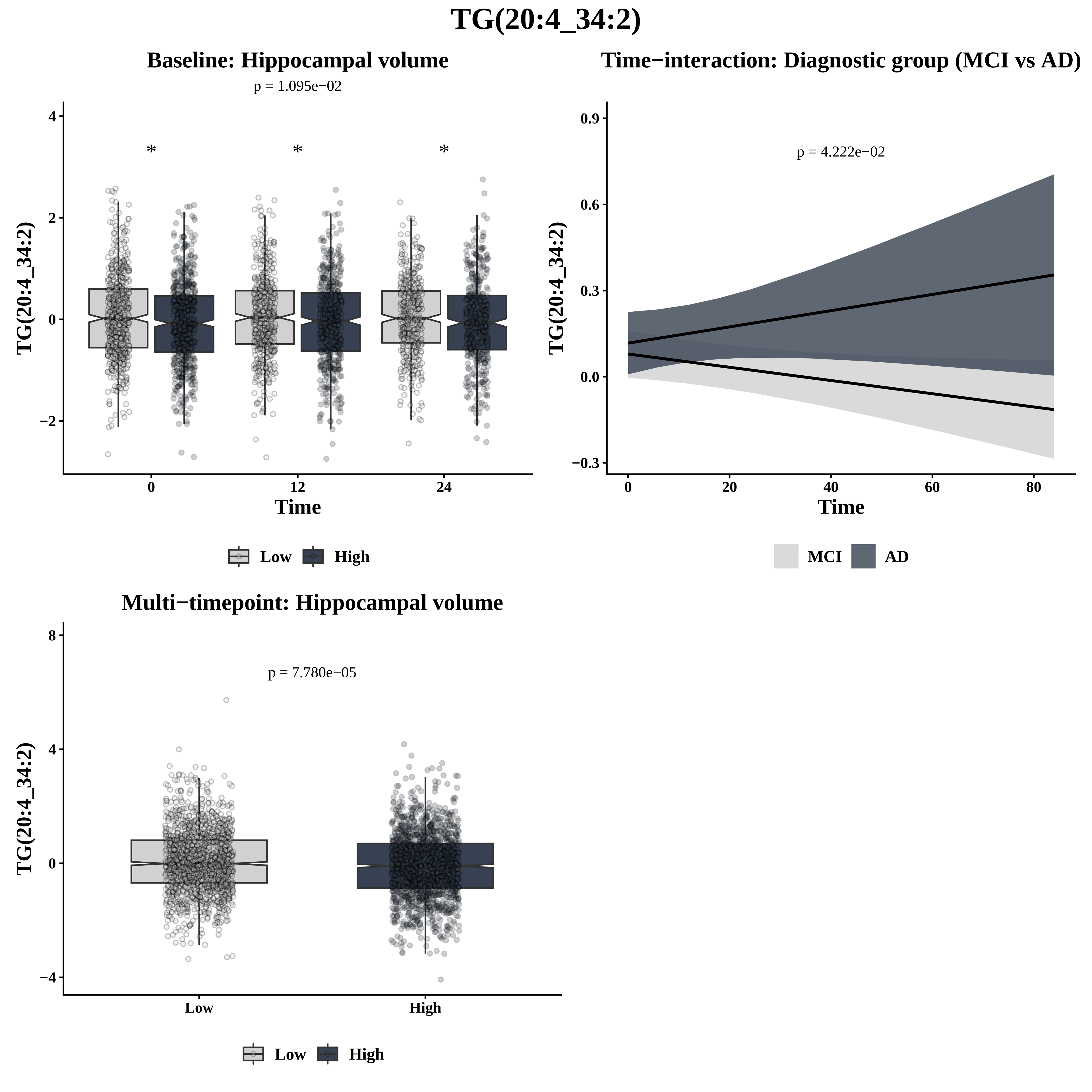
TG(20:4_34:2)
Baseline: Hippocampal volume
p = 1.095e−02
*
*
*
4
2
0
−2
TG(20:4_34:2)
0
12
24
Time
Low
High
Time−interaction: Diagnostic group (MCI vs AD)
p = 4.222e−02
0.9
0.6
0.3
0.0
−0.3
TG(20:4_34:2)
0
20
40
60
80
Time
MCI
AD
Multi−timepoint: Hippocampal volume
p = 7.780e−05
8
4
0
−4
TG(20:4_34:2)
Low
High
Low
High

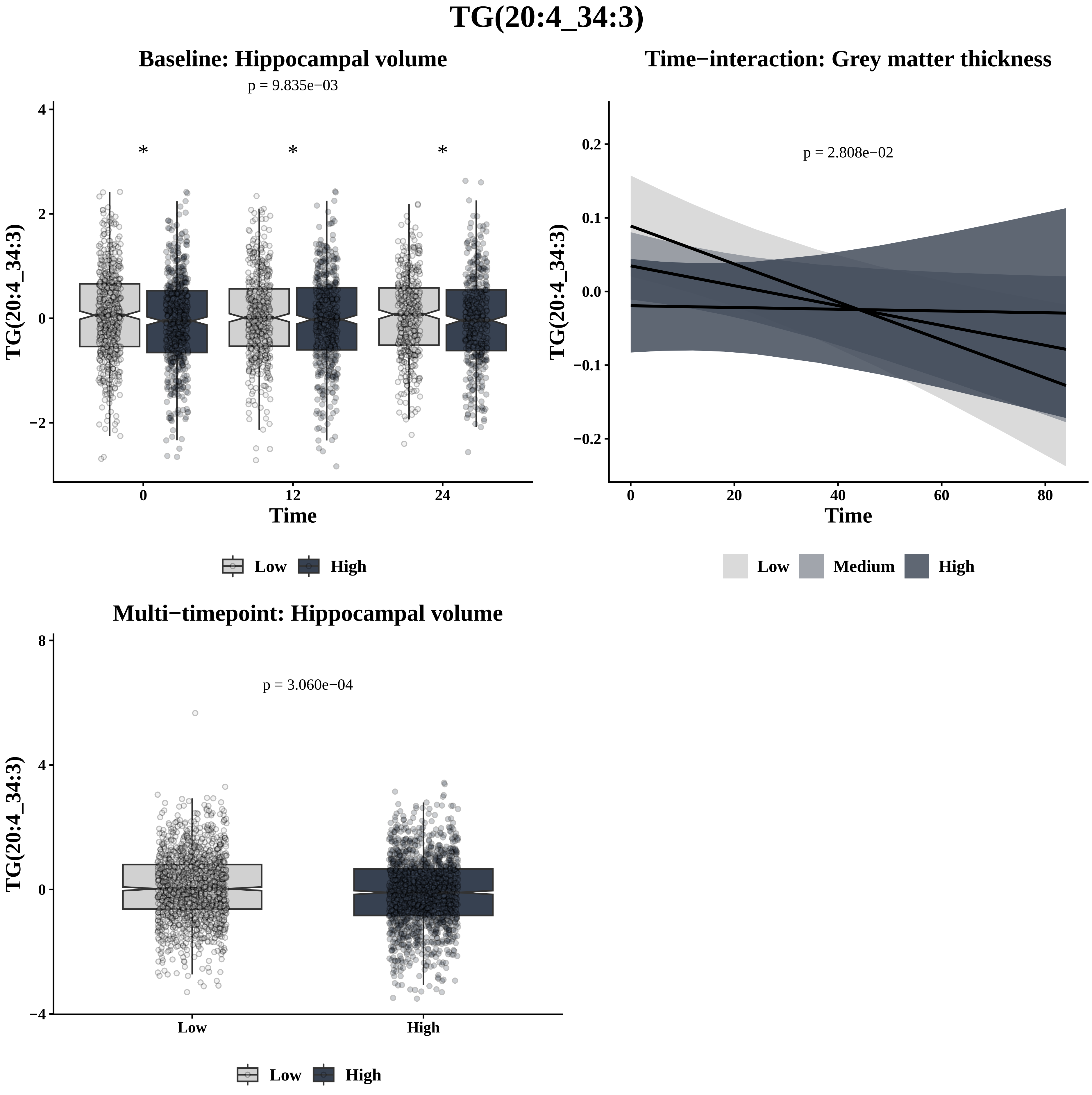

TG(20:4_34:3)
Baseline: Hippocampal volume
p = 9.835e−03
*
*
*
TG(20:4_34:3)
4
2
0
−2
0
12
24
Time
Low
High
Time−interaction: Grey matter thickness
p = 2.808e−02
TG(20:4_34:3)
0.2
0.1
0.0
−0.1
−0.2
0
20
40
60
80
Time
Low
Medium
High
Multi−timepoint: Hippocampal volume
p = 3.060e−04
TG(20:4_34:3)
8
4
0
−4
Low
High
Low
High

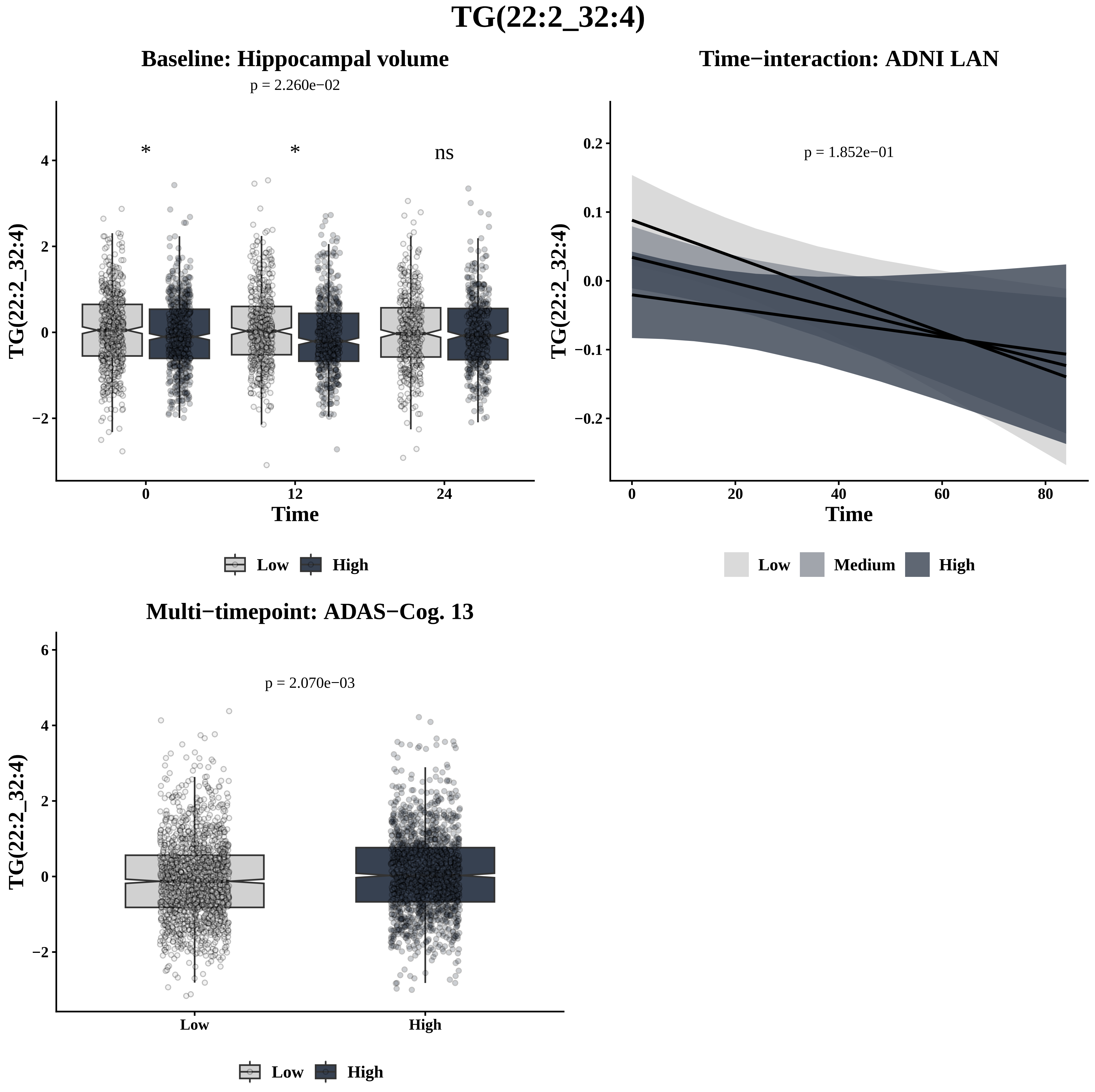
TG(22:2_32:4)
Baseline: Hippocampal volume
p = 2.260e−02
*
*
ns
TG(22:2_32:4)
4
2
0
−2
0
12
24
Time
Low
High
Time−interaction: ADNI LAN
p = 1.852e−01
TG(22:2_32:4)
0.2
0.1
0.0
−0.1
−0.2
0
20
40
60
80
Time
Low
Medium
High
Multi−timepoint: ADAS−Cog. 13
p = 2.070e−03
TG(22:2_32:4)
6
4
2
0
−2
Low
High
Low
High

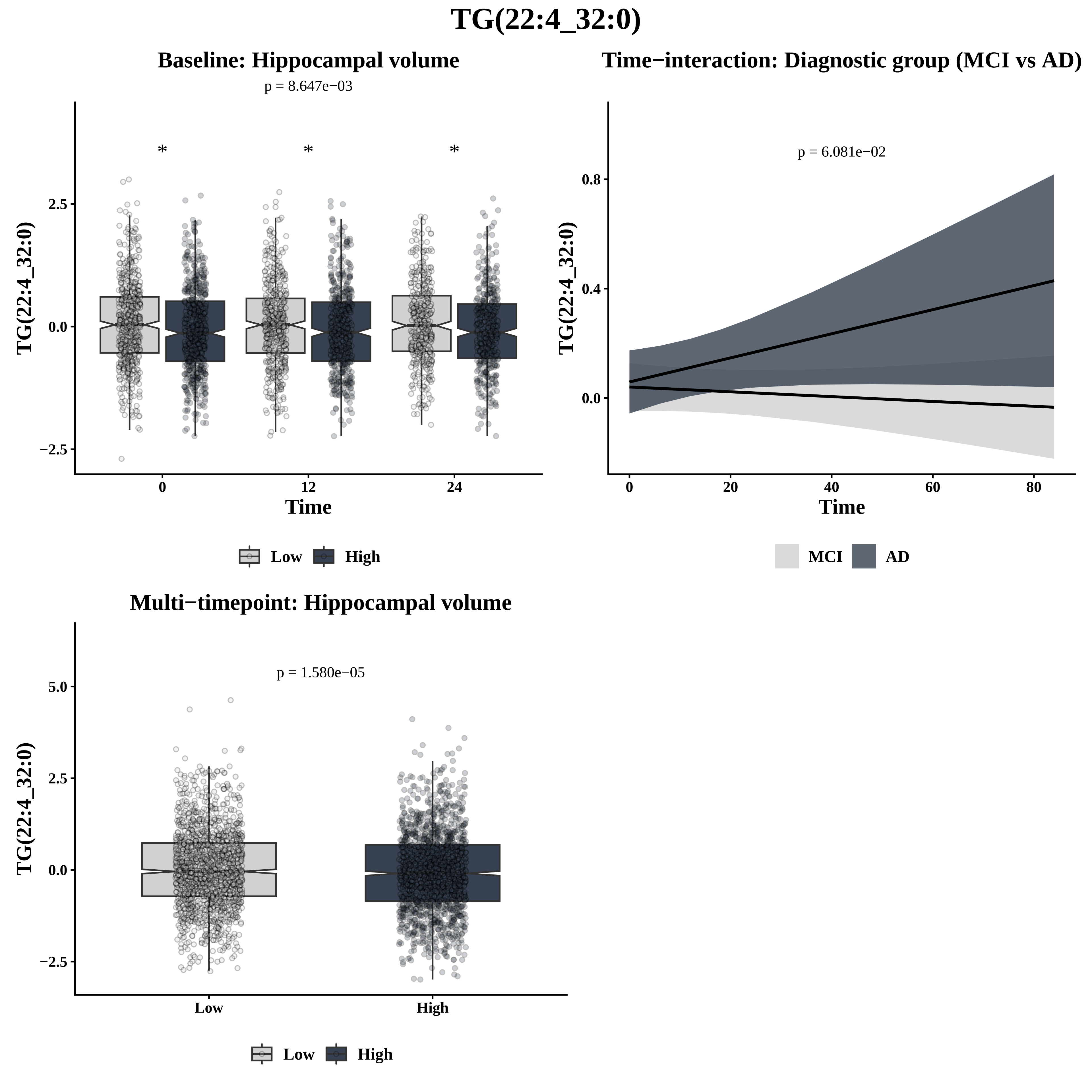

TG(22:4_32:0)
Baseline: Hippocampal volume
p = 8.647e−03
*
*
*
2.5
0.0
−2.5
TG(22:4_32:0)
0
12
24
Time
Low
High
Time−interaction: Diagnostic group (MCI vs AD)
p = 6.081e−02
0.8
0.4
0.0
TG(22:4_32:0)
0
20
40
60
80
Time
MCI
AD
Multi−timepoint: Hippocampal volume
p = 1.580e−05
5.0
2.5
0.0
−2.5
TG(22:4_32:0)
Low
High
Low
High

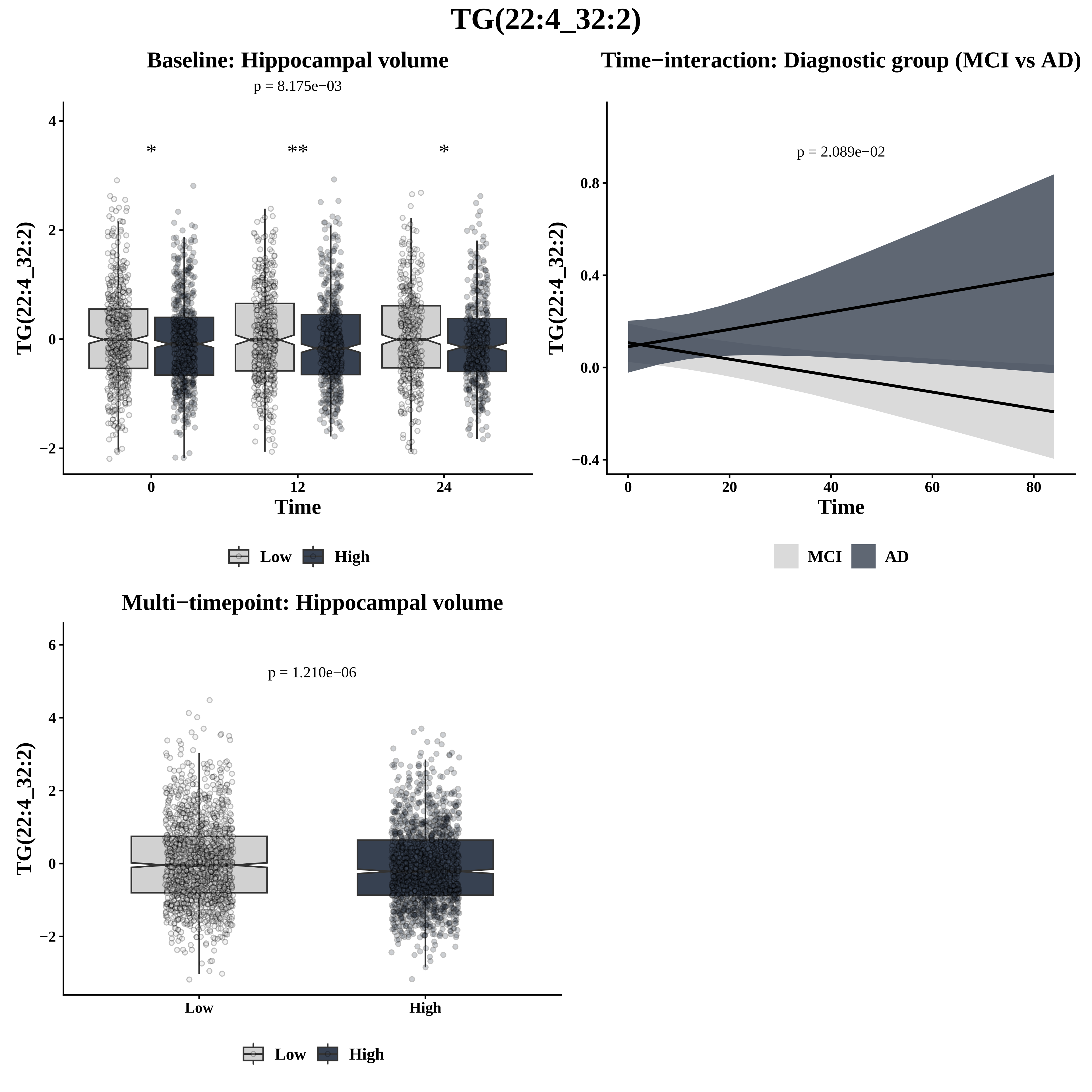

TG(22:4_32:2)
Baseline: Hippocampal volume
p = 8.175e−03
*
**
*
TG(22:4_32:2)
Time
0
12
24
Low
High
Time−interaction: Diagnostic group (MCI vs AD)
p = 2.089e−02
TG(22:4_32:2)
Time
MCI
AD
Multi−timepoint: Hippocampal volume
p = 1.210e−06
TG(22:4_32:2)
Low
High

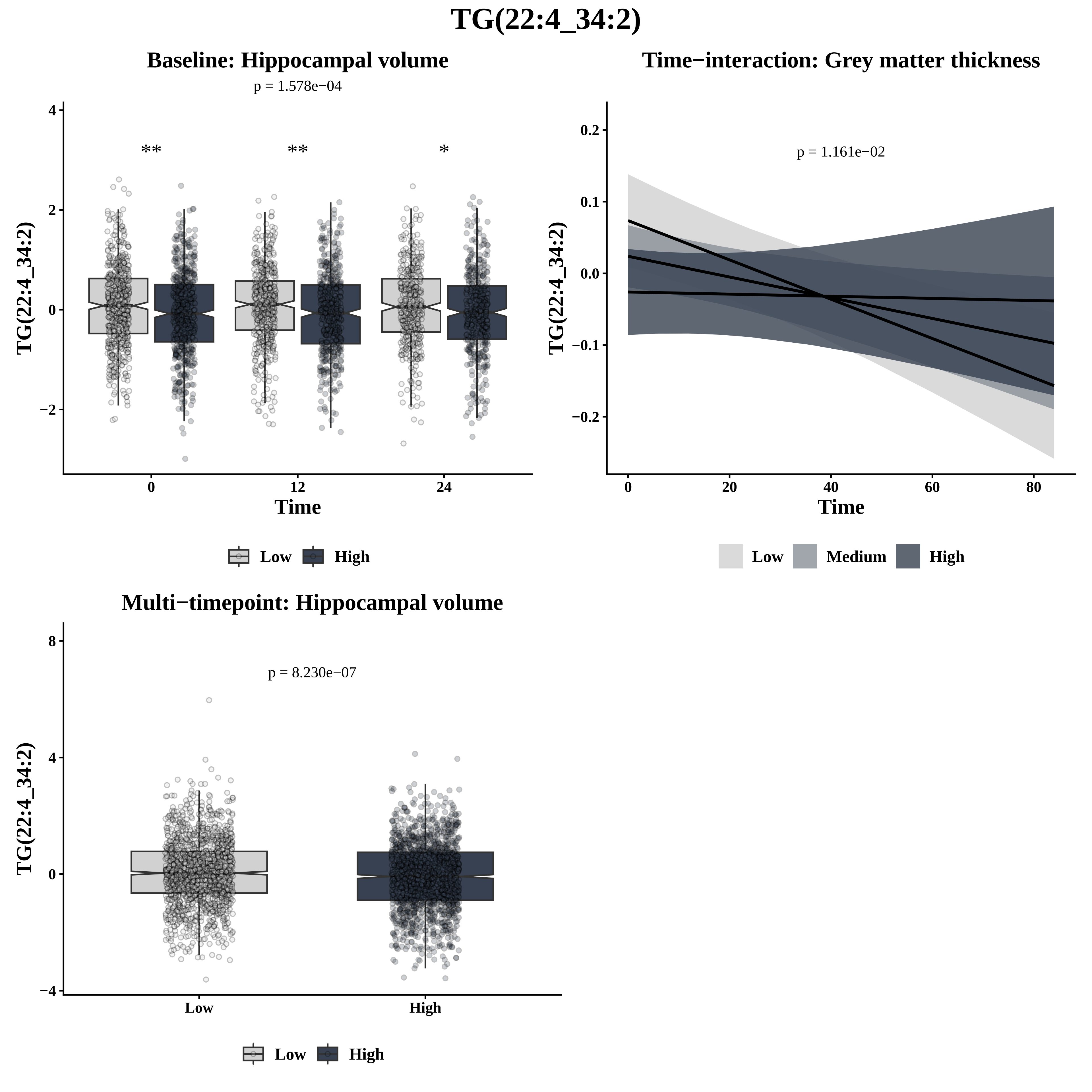
TG(22:4_34:2)
Baseline: Hippocampal volume
p = 1.578e−04
**
**
*
TG(22:4_34:2)
4
2
0
−2
0
12
24
Time
Low
High
Time−interaction: Grey matter thickness
p = 1.161e−02
TG(22:4_34:2)
0.2
0.1
0.0
−0.1
−0.2
0
20
40
60
80
Time
Low
Medium
High
Multi−timepoint: Hippocampal volume
p = 8.230e−07
TG(22:4_34:2)
8
4
0
−4
Low
High
Low
High

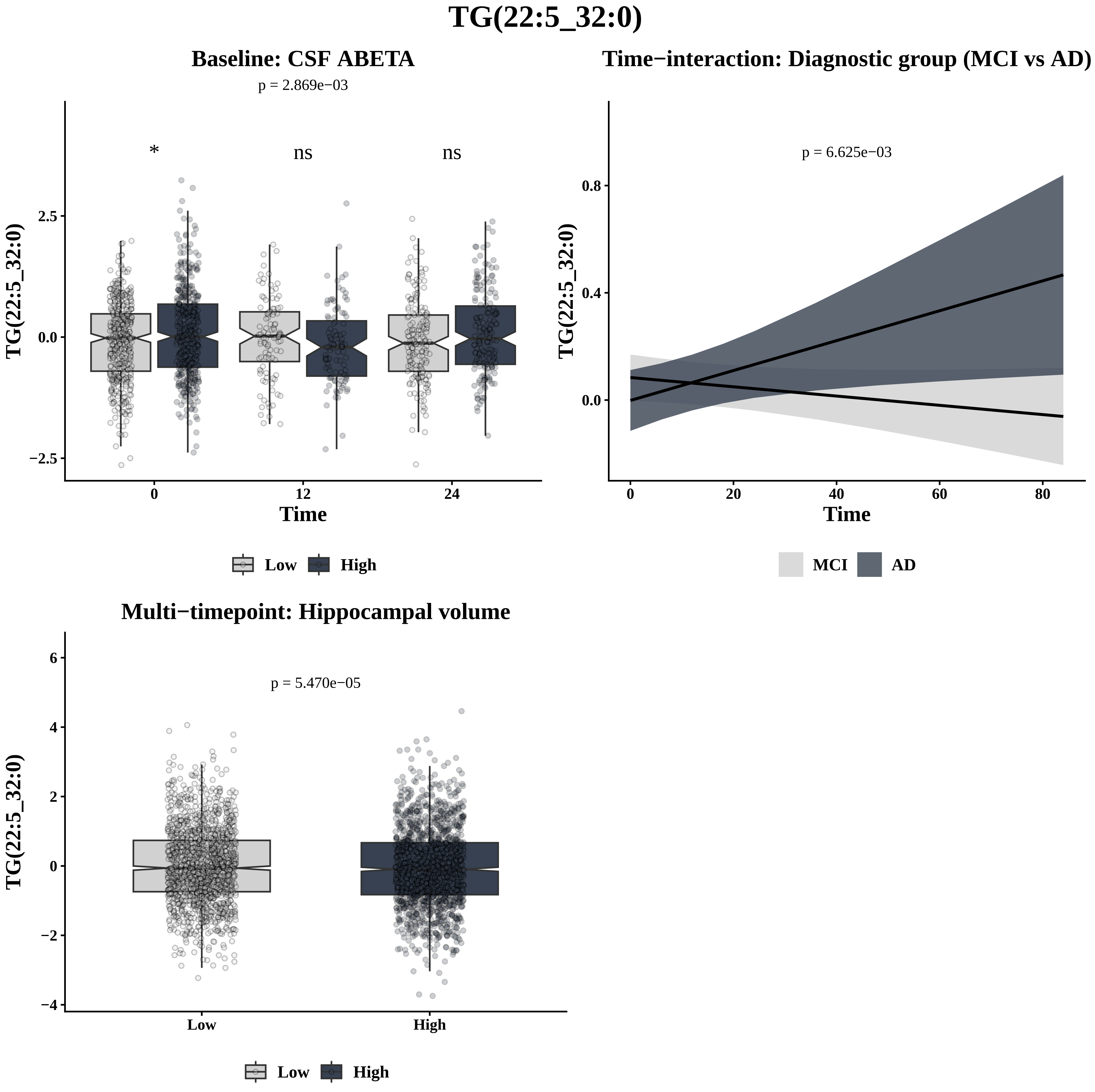
TG(22:5_32:0)
Baseline: CSF ABETA
p = 2.869e−03
*
ns
ns
2.5
0.0
−2.5
TG(22:5_32:0)
0
12
24
Time
Low
High
Time−interaction: Diagnostic group (MCI vs AD)
p = 6.625e−03
0.8
0.4
0.0
TG(22:5_32:0)
0
20
40
60
80
Time
MCI
AD
Multi−timepoint: Hippocampal volume
p = 5.470e−05
6
4
2
0
−2
−4
TG(22:5_32:0)
Low
High
Low
High

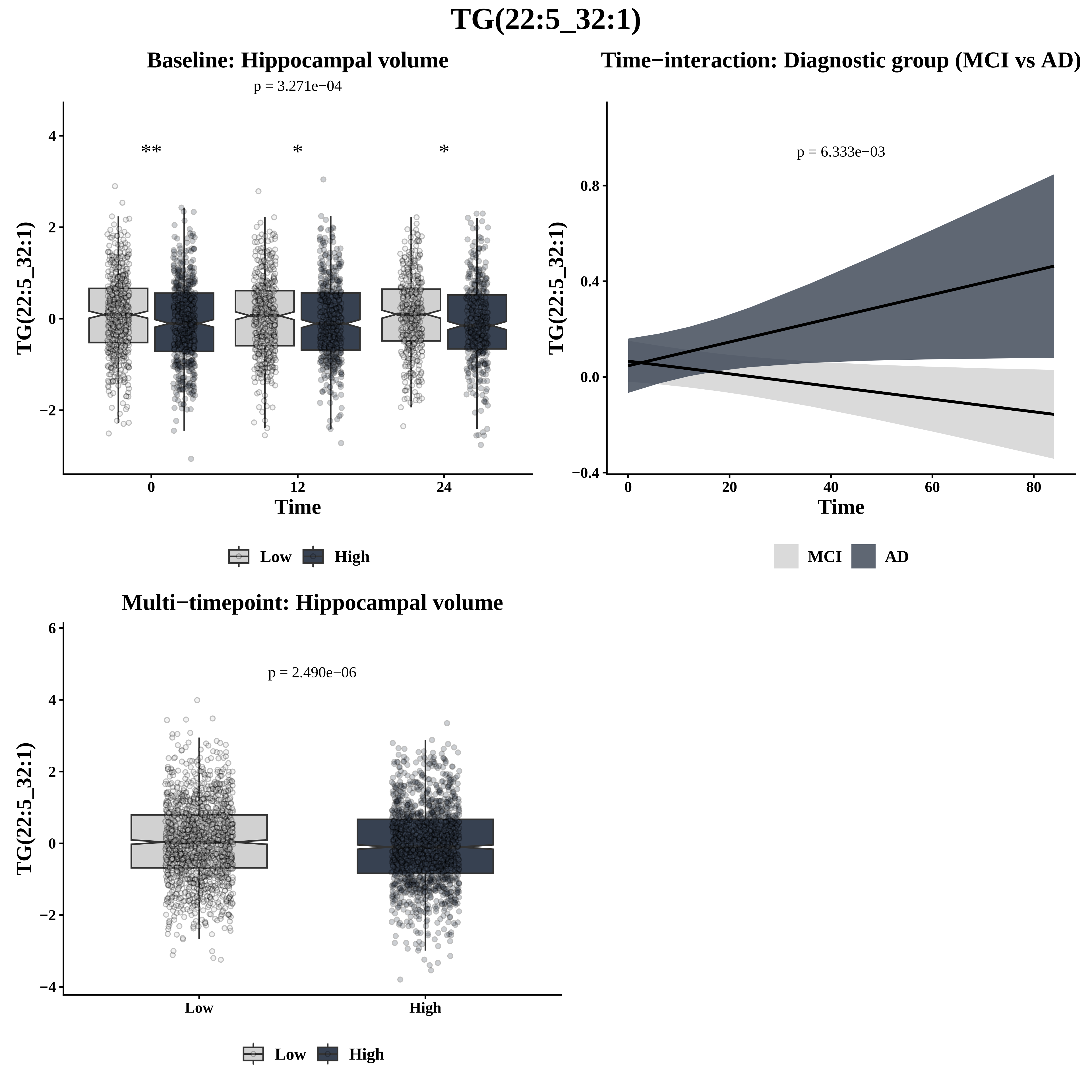
TG(22:5_32:1)
Baseline: Hippocampal volume
p = 3.271e−04
**
*
*
TG(22:5_32:1)
Time
0
12
24
Low
High
Time−interaction: Diagnostic group (MCI vs AD)
p = 6.333e−03
TG(22:5_32:1)
Time
MCI
AD
Multi−timepoint: Hippocampal volume
p = 2.490e−06
TG(22:5_32:1)
Low
High

# TG(22:5_34:1)

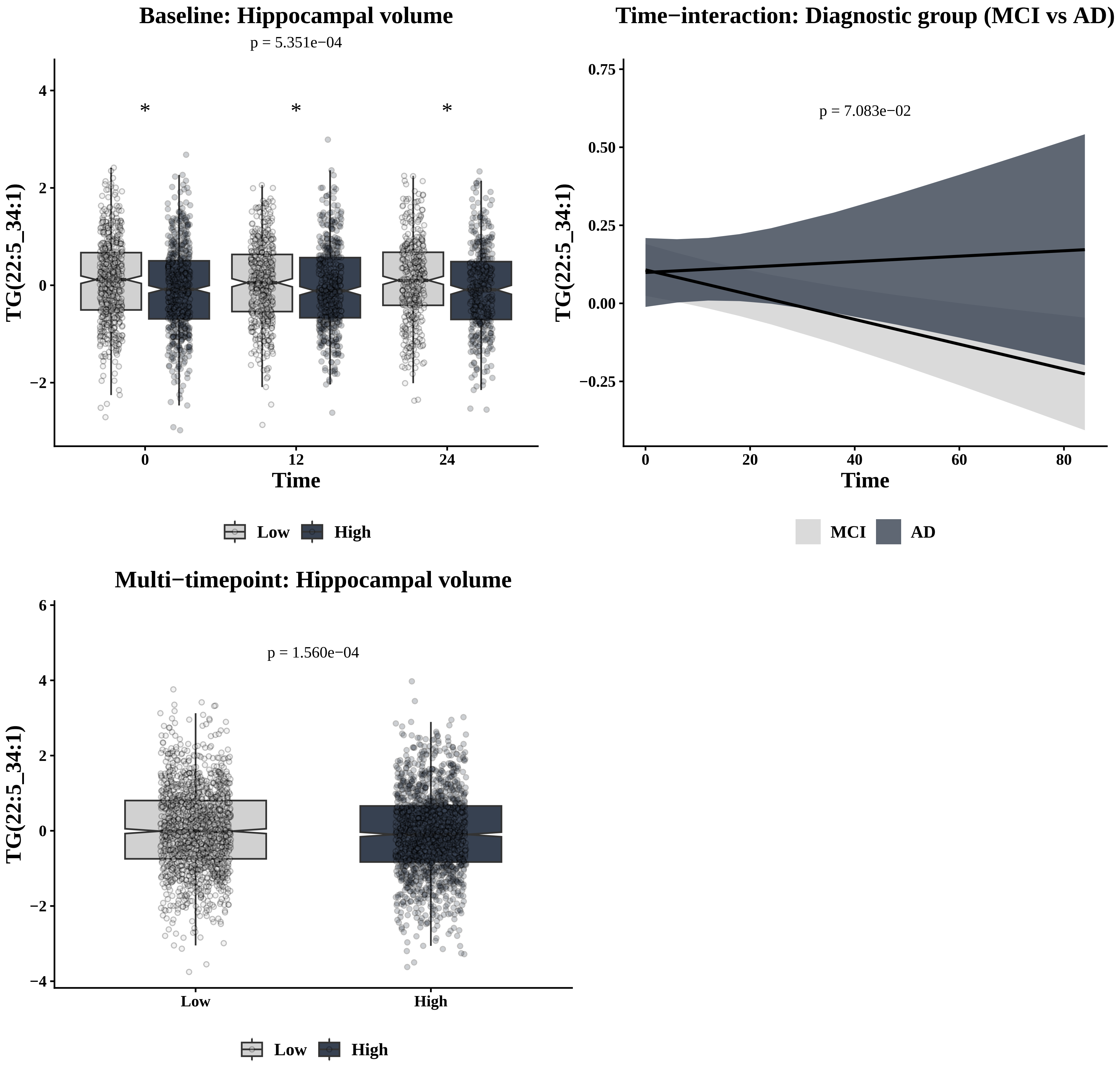
Baseline: Hippocampal volume
p = 5.351e−04
TG(22:5_34:1)
Time
0
12
24
Low
High
Time−interaction: Diagnostic group (MCI vs AD)
p = 7.083e−02
TG(22:5_34:1)
Time
MCI
AD
Multi−timepoint: Hippocampal volume
p = 1.560e−04
TG(22:5_34:1)
Low
High

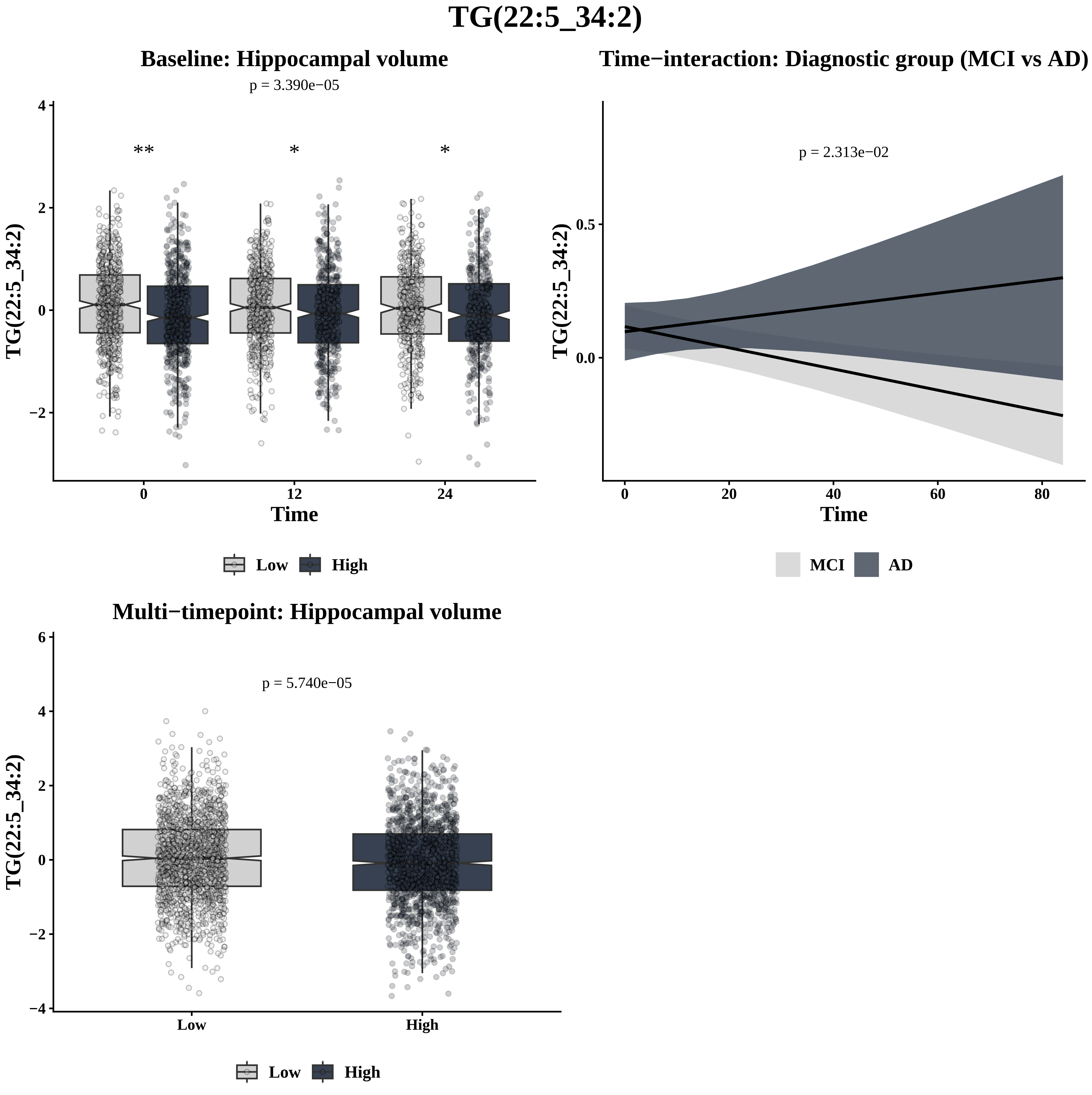
TG(22:5_34:2)
Baseline: Hippocampal volume
p = 3.390e−05
**
*
*
TG(22:5_34:2)
Time
0
12
24
Low
High
Time−interaction: Diagnostic group (MCI vs AD)
p = 2.313e−02
TG(22:5_34:2)
Time
MCI
AD
Multi−timepoint: Hippocampal volume
p = 5.740e−05
TG(22:5_34:2)
Low
High

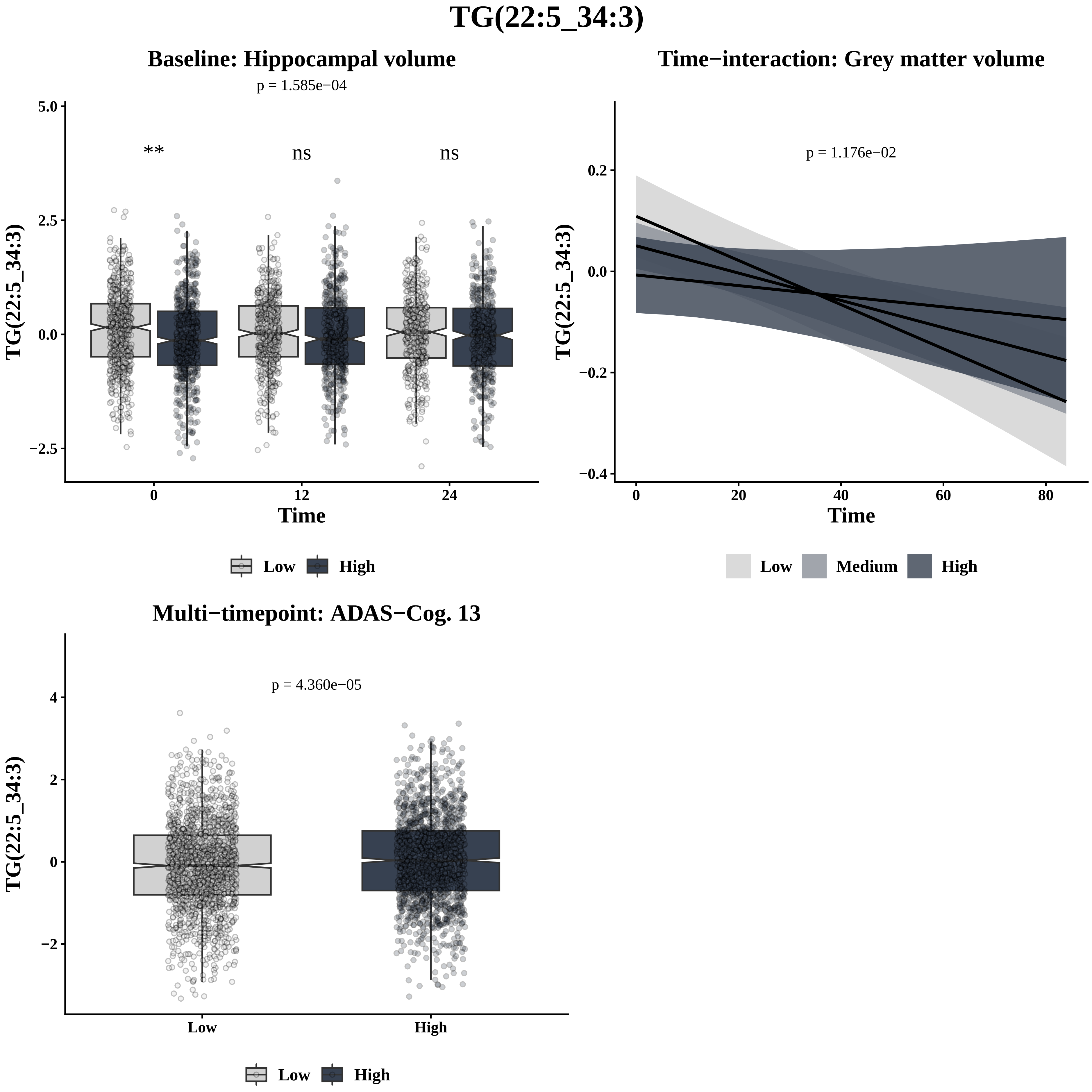
TG(22:5_34:3)
Baseline: Hippocampal volume
p = 1.585e−04
**
ns
ns
5.0
2.5
0.0
−2.5
TG(22:5_34:3)
0
12
24
Time
Low
High
Time−interaction: Grey matter volume
p = 1.176e−02
0.2
0.0
−0.2
−0.4
TG(22:5_34:3)
0
20
40
60
80
Time
Low
Medium
High
Multi−timepoint: ADAS−Cog. 13
p = 4.360e−05
4
2
0
−2
TG(22:5_34:3)
Low
High
Low
High

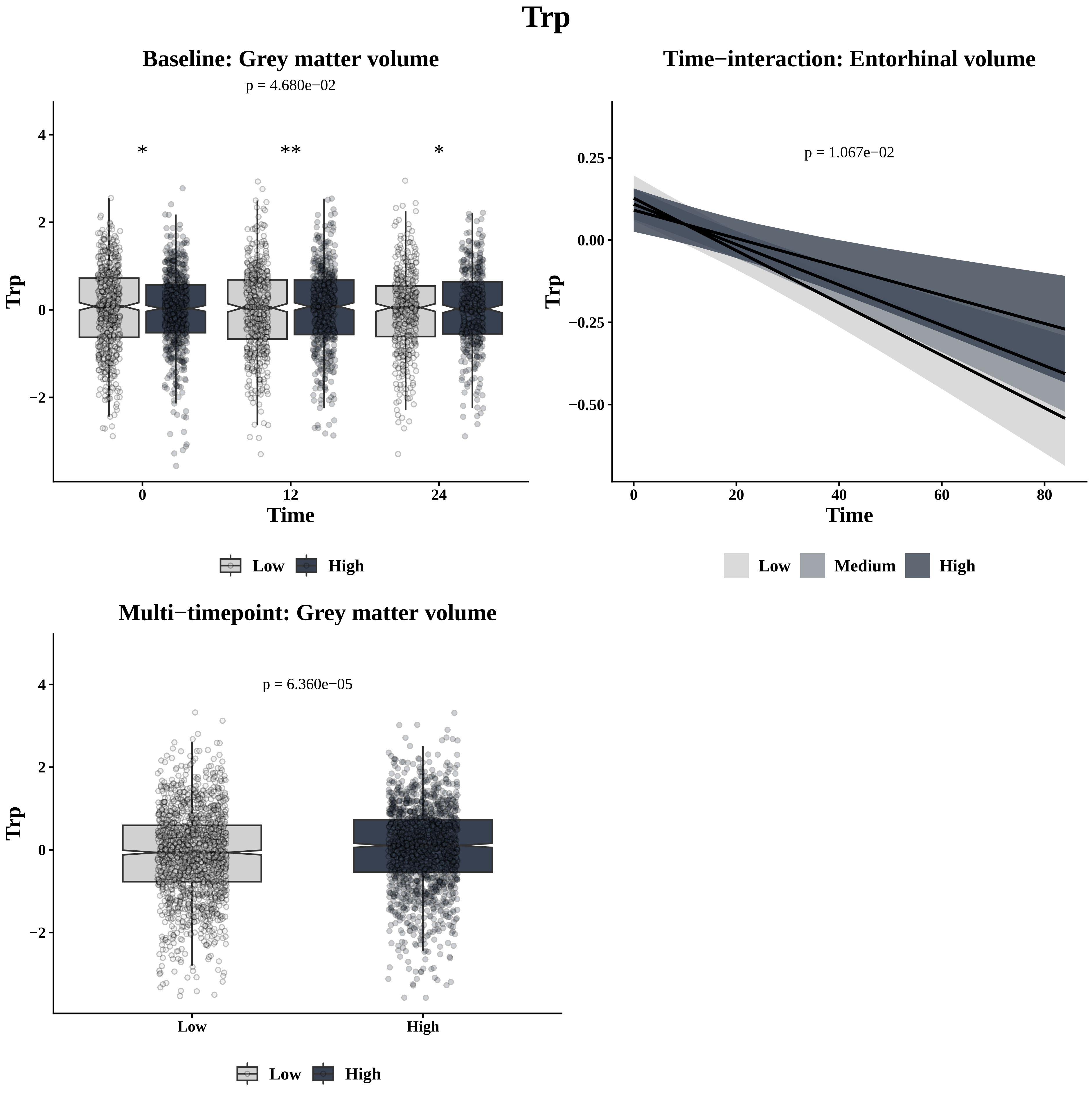
Trp
Baseline: Grey matter volume
p = 4.680e−02
*
**
*
Trp
Time
0
12
24
4
2
0
−2
Low
High
Time−interaction: Entorhinal volume
p = 1.067e−02
Trp
Time
0.25
0.00
−0.25
−0.50
0
20
40
60
80
Low
Medium
High
Multi−timepoint: Grey matter volume
p = 6.360e−05
Trp
4
2
0
−2
Low
High
Low
High

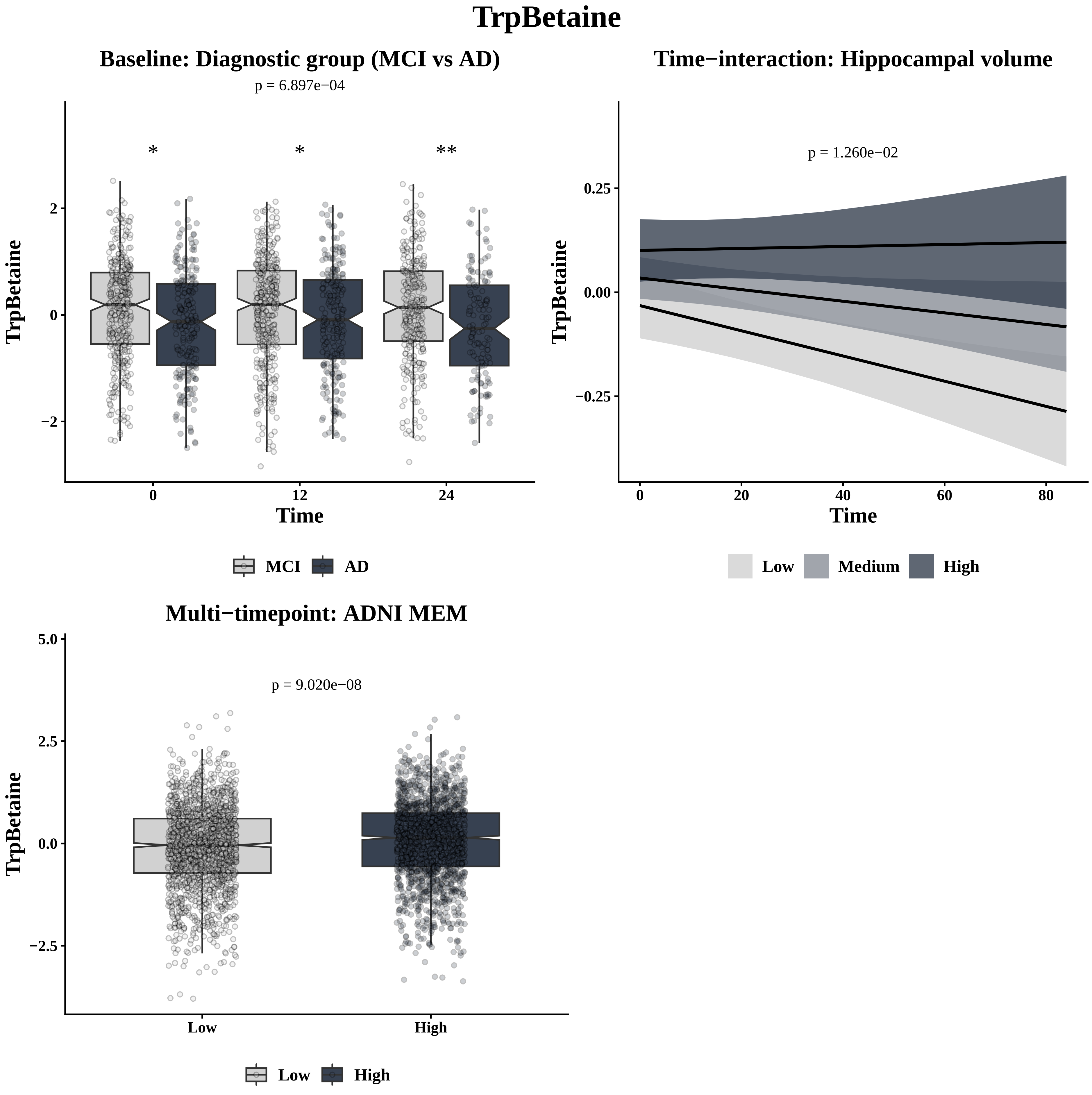

TrpBetaine
Baseline: Diagnostic group (MCI vs AD)
p = 6.897e−04
*
*
**
TrpBetaine
Time
0
12
24
2
0
−2
MCI
AD
Time−interaction: Hippocampal volume
p = 1.260e−02
TrpBetaine
0.25
0.00
−0.25
Time
0
20
40
60
80
Low
Medium
High
Multi−timepoint: ADNI MEM
p = 9.020e−08
TrpBetaine
5.0
2.5
0.0
−2.5
Low
High
Low
High

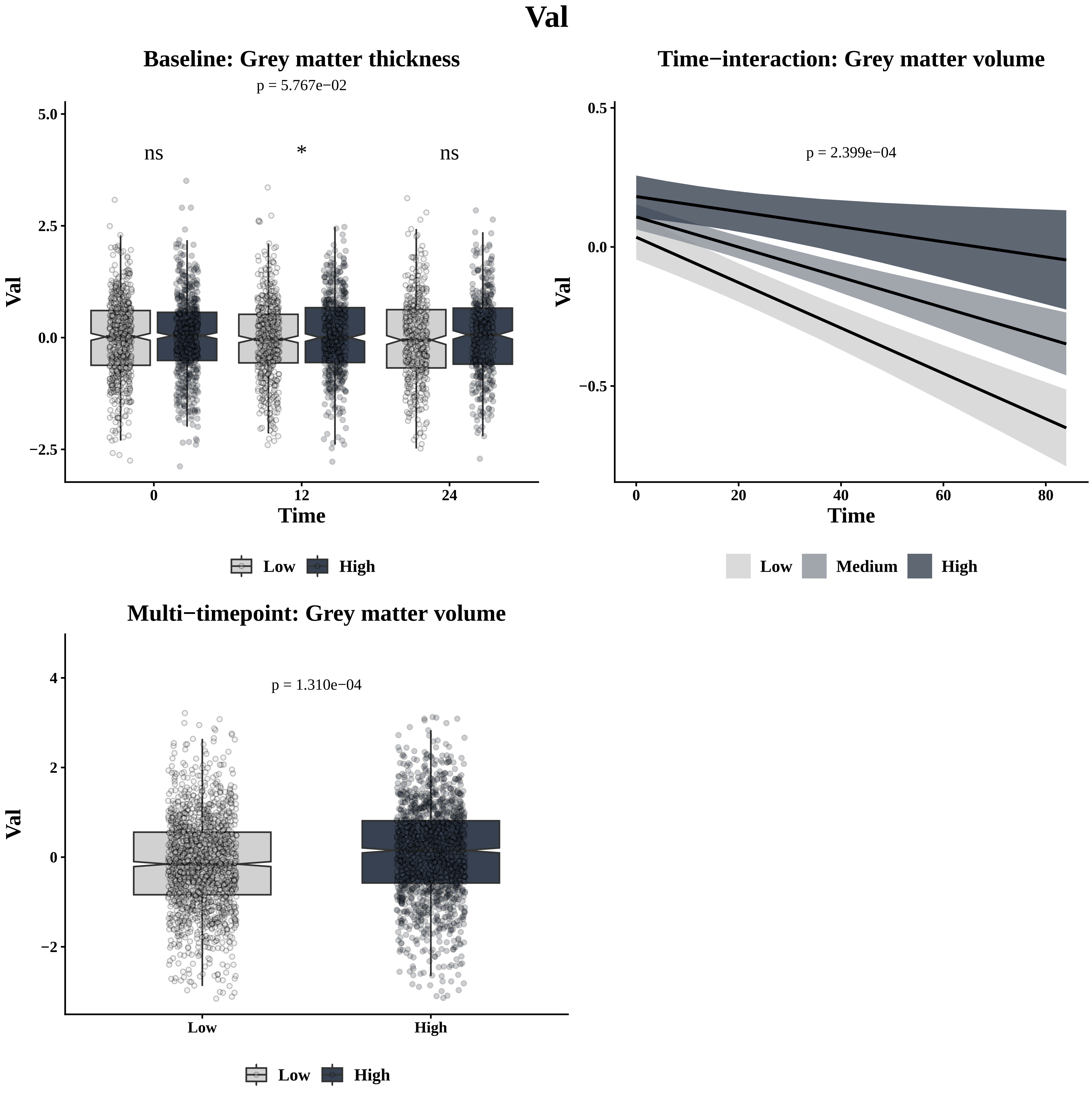

Val
Baseline: Grey matter thickness
p = 5.767e−02
ns
*
ns
5.0
2.5
0.0
−2.5
Val
0
12
24
Time
Low
High
Time−interaction: Grey matter volume
p = 2.399e−04
0.5
0.0
−0.5
Val
0
20
40
60
80
Time
Low
Medium
High
Multi−timepoint: Grey matter volume
p = 1.310e−04
4
2
0
−2
Val
Low
High
Low
High

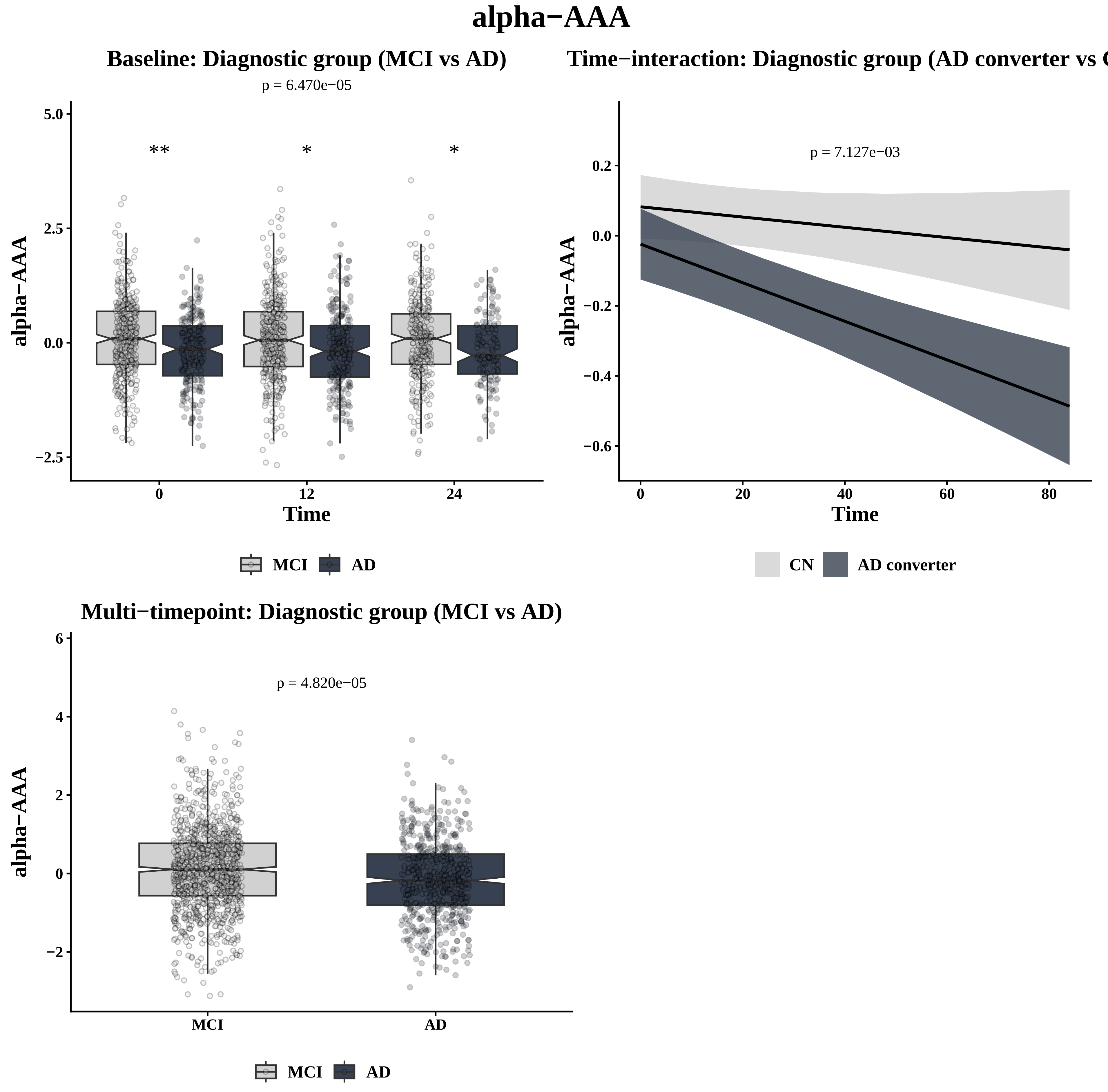
alpha−AAA
Baseline: Diagnostic group (MCI vs AD)
p = 6.470e−05
**
*
*
5.0
2.5
0.0
−2.5
alpha−AAA
0
12
24
Time
MCI
AD
Time−interaction: Diagnostic group (AD converter vs C
p = 7.127e−03
0.2
0.0
−0.2
−0.4
−0.6
alpha−AAA
0
20
40
60
80
Time
CN
AD converter
Multi−timepoint: Diagnostic group (MCI vs AD)
p = 4.820e−05
6
4
2
0
−2
alpha−AAA
MCI
AD
MCI
AD

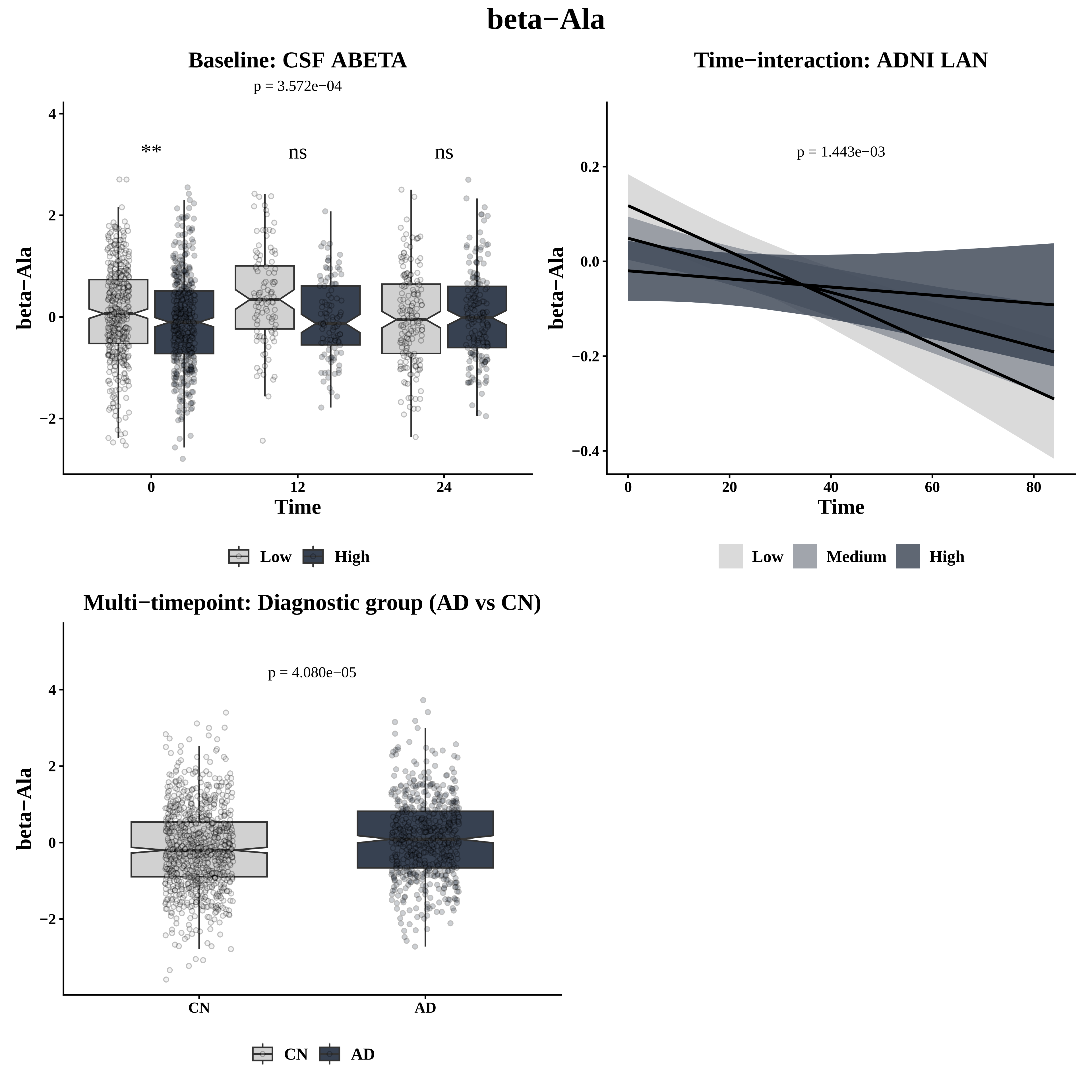
beta−Ala
Baseline: CSF ABETA
p = 3.572e−04
**
ns
ns
beta−Ala
Time
Low
High
Time−interaction: ADNI LAN
p = 1.443e−03
beta−Ala
Time
Low
Medium
High
Multi−timepoint: Diagnostic group (AD vs CN)
p = 4.080e−05
beta−Ala
CN
AD

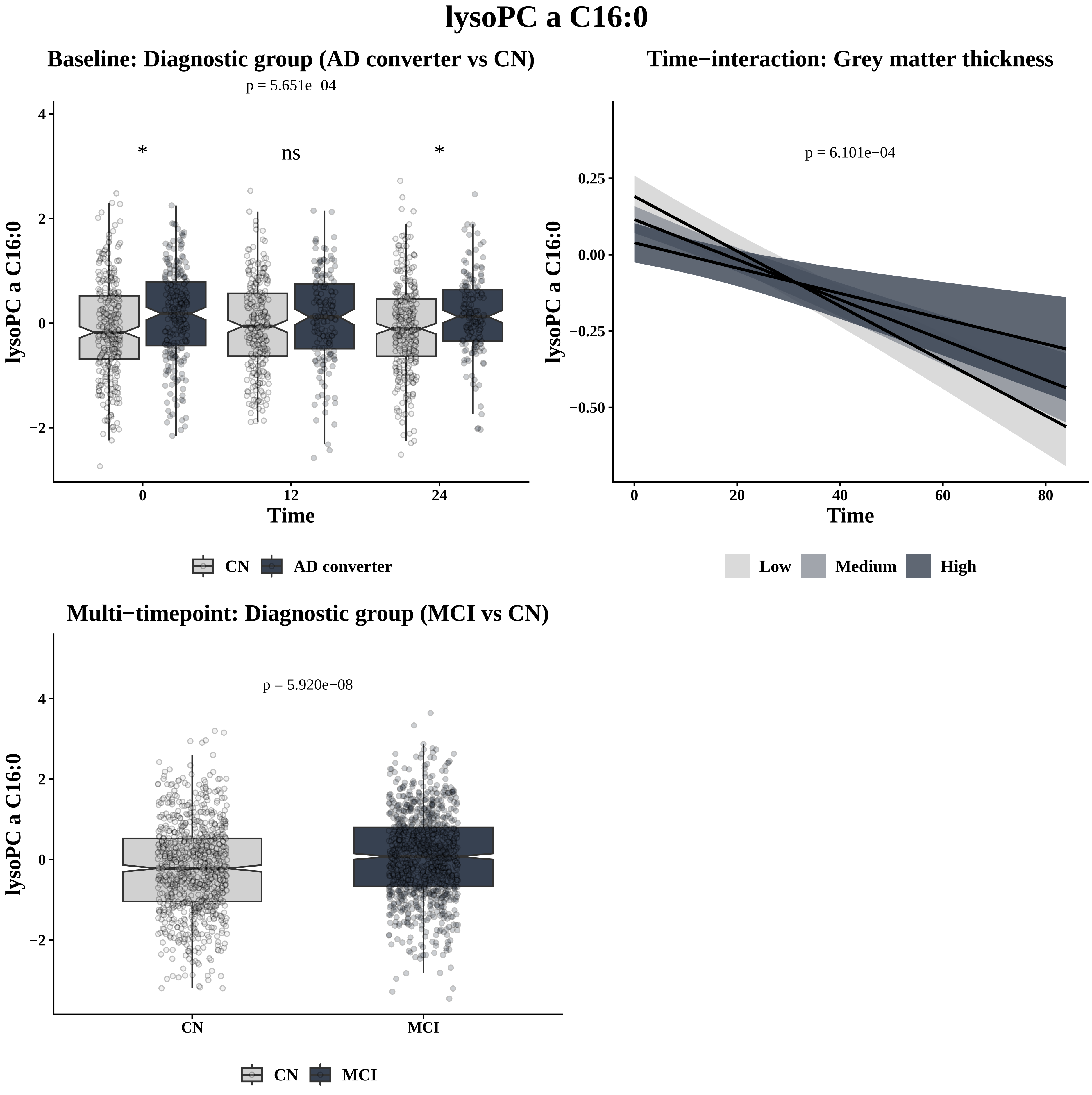
lysoPC a C16:0
Baseline: Diagnostic group (AD converter vs CN)
p = 5.651e−04
*
ns
*
lysoPC a C16:0
4
2
0
−2
0
12
24
Time
CN
AD converter
Time−interaction: Grey matter thickness
p = 6.101e−04
lysoPC a C16:0
0.25
0.00
−0.25
−0.50
0
20
40
60
80
Time
Low
Medium
High
Multi−timepoint: Diagnostic group (MCI vs CN)
p = 5.920e−08
lysoPC a C16:0
4
2
0
−2
CN
MCI
CN
MCI

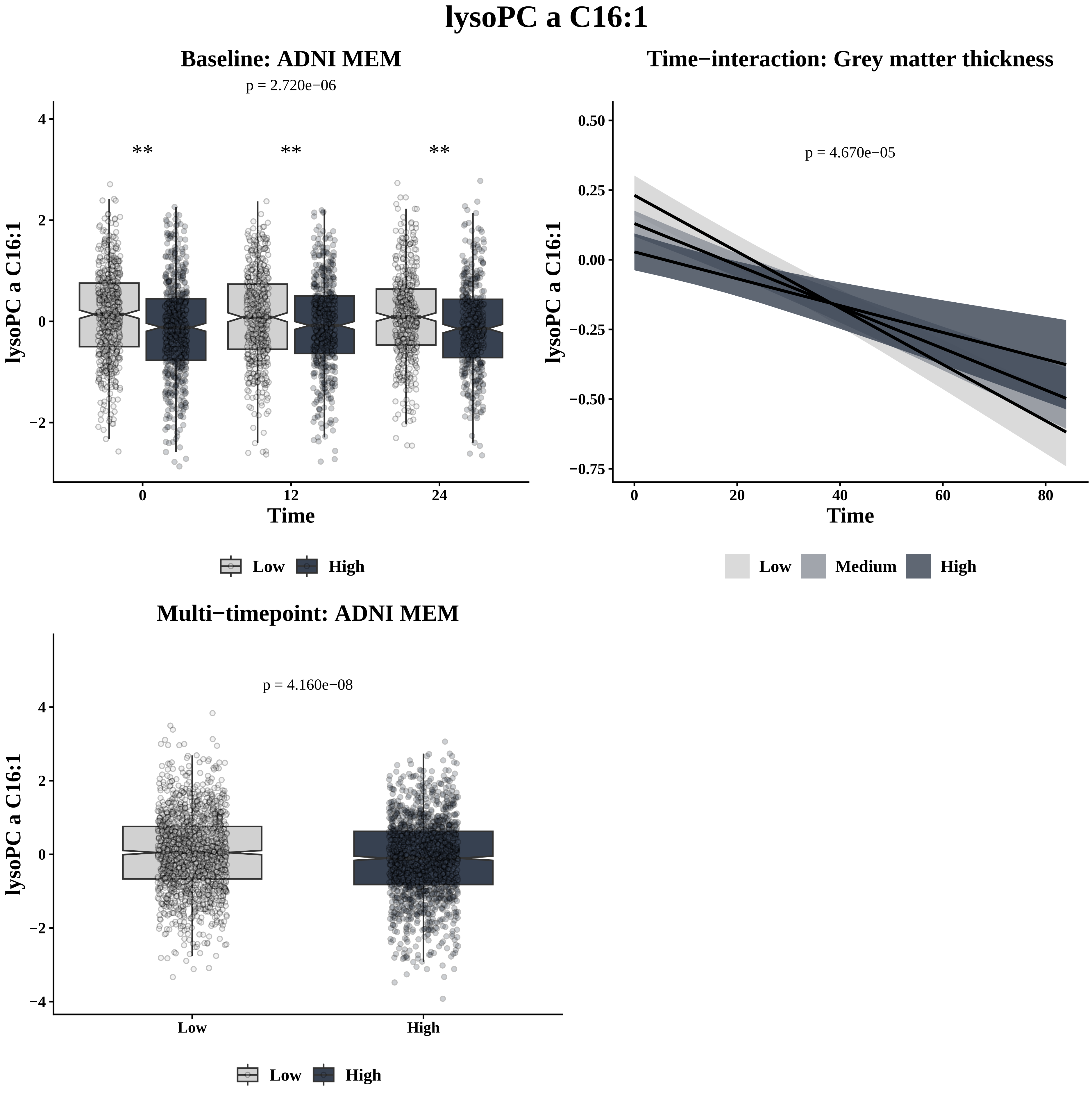
lysoPC a C16:1
Baseline: ADNI MEM
p = 2.720e−06
**
**
**
lysoPC a C16:1
Time
0
12
24
4
2
0
−2
Low
High
Time−interaction: Grey matter thickness
p = 4.670e−05
lysoPC a C16:1
0.50
0.25
0.00
−0.25
−0.50
−0.75
0
20
40
60
80
Time
Low
Medium
High
Multi−timepoint: ADNI MEM
p = 4.160e−08
lysoPC a C16:1
4
2
0
−2
−4
Low
High
Low
High

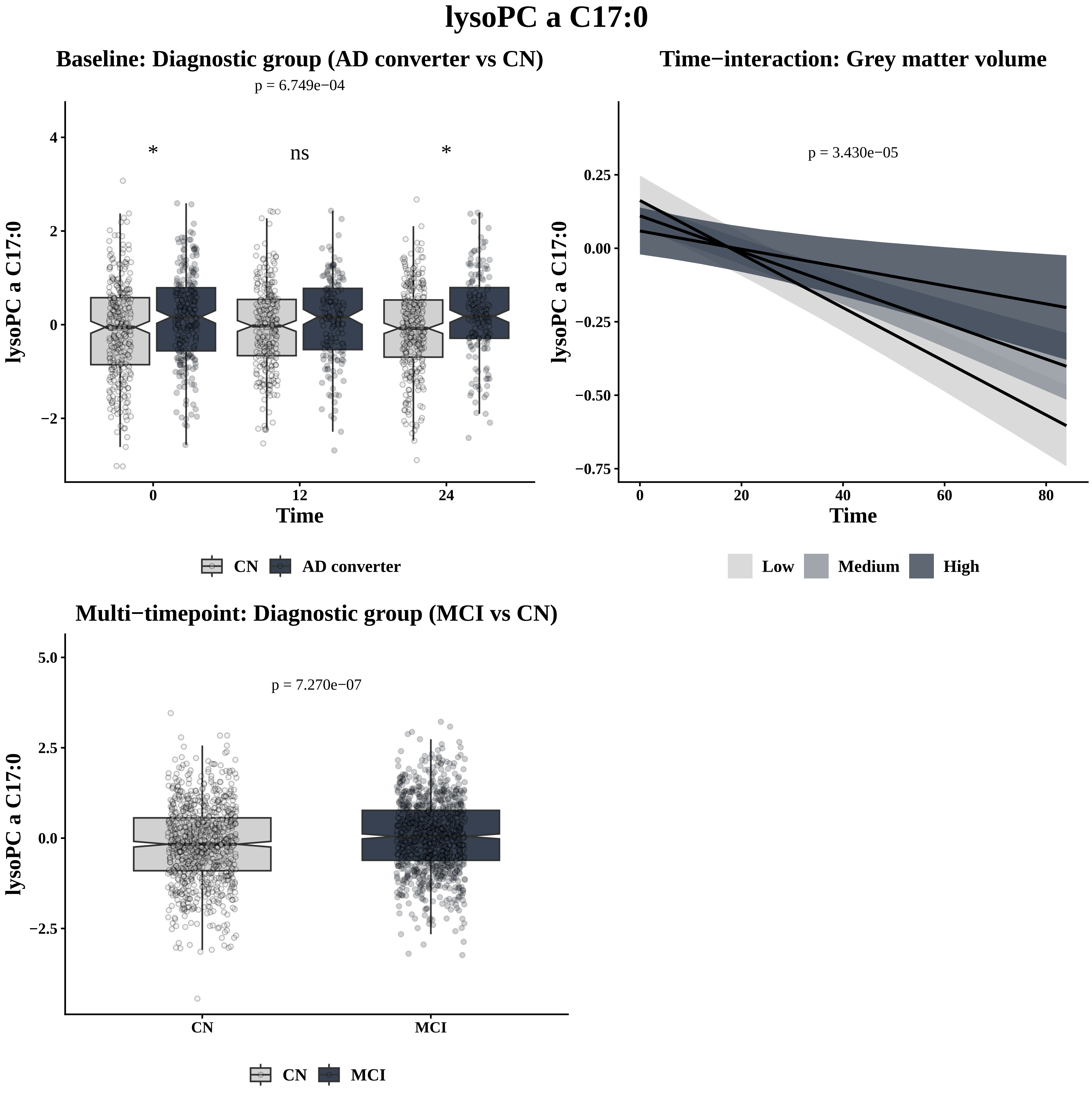
lysoPC a C17:0
Baseline: Diagnostic group (AD converter vs CN)
p = 6.749e−04
*
ns
*
lysoPC a C17:0
4
2
0
−2
0
12
24
Time
CN
AD converter
Time−interaction: Grey matter volume
p = 3.430e−05
lysoPC a C17:0
0.25
0.00
−0.25
−0.50
−0.75
0
20
40
60
80
Time
Low
Medium
High
Multi−timepoint: Diagnostic group (MCI vs CN)
p = 7.270e−07
lysoPC a C17:0
5.0
2.5
0.0
−2.5
CN
MCI
CN
MCI

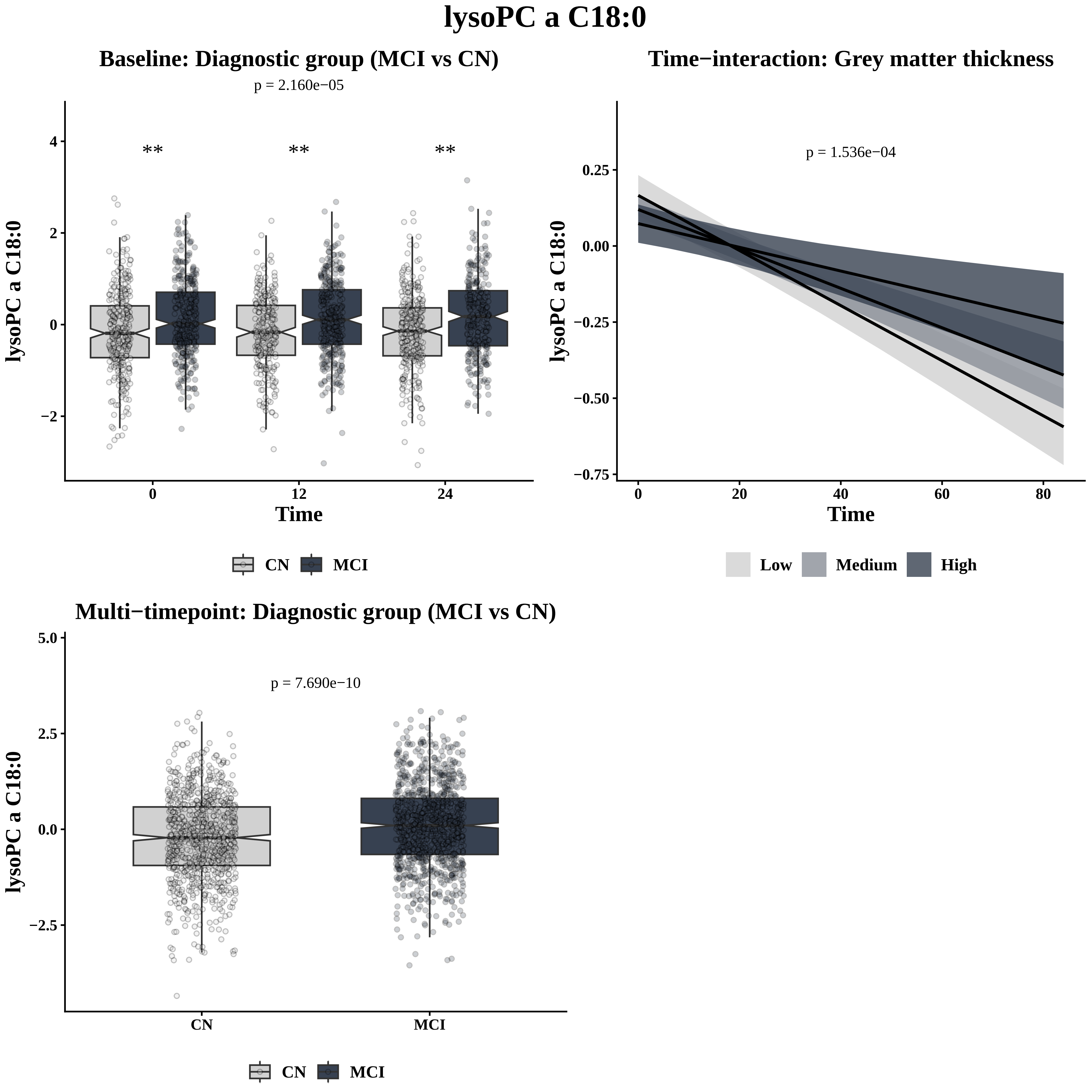
lysoPC a C18:0
Baseline: Diagnostic group (MCI vs CN)
p = 2.160e−05
**
**
**
lysoPC a C18:0
Time
4
2
0
−2
0
12
24
CN
MCI
Time−interaction: Grey matter thickness
p = 1.536e−04
lysoPC a C18:0
Time
0.25
0.00
−0.25
−0.50
−0.75
0
20
40
60
80
Low
Medium
High
Multi−timepoint: Diagnostic group (MCI vs CN)
p = 7.690e−10
lysoPC a C18:0
5.0
2.5
0.0
−2.5
CN
MCI
CN
MCI

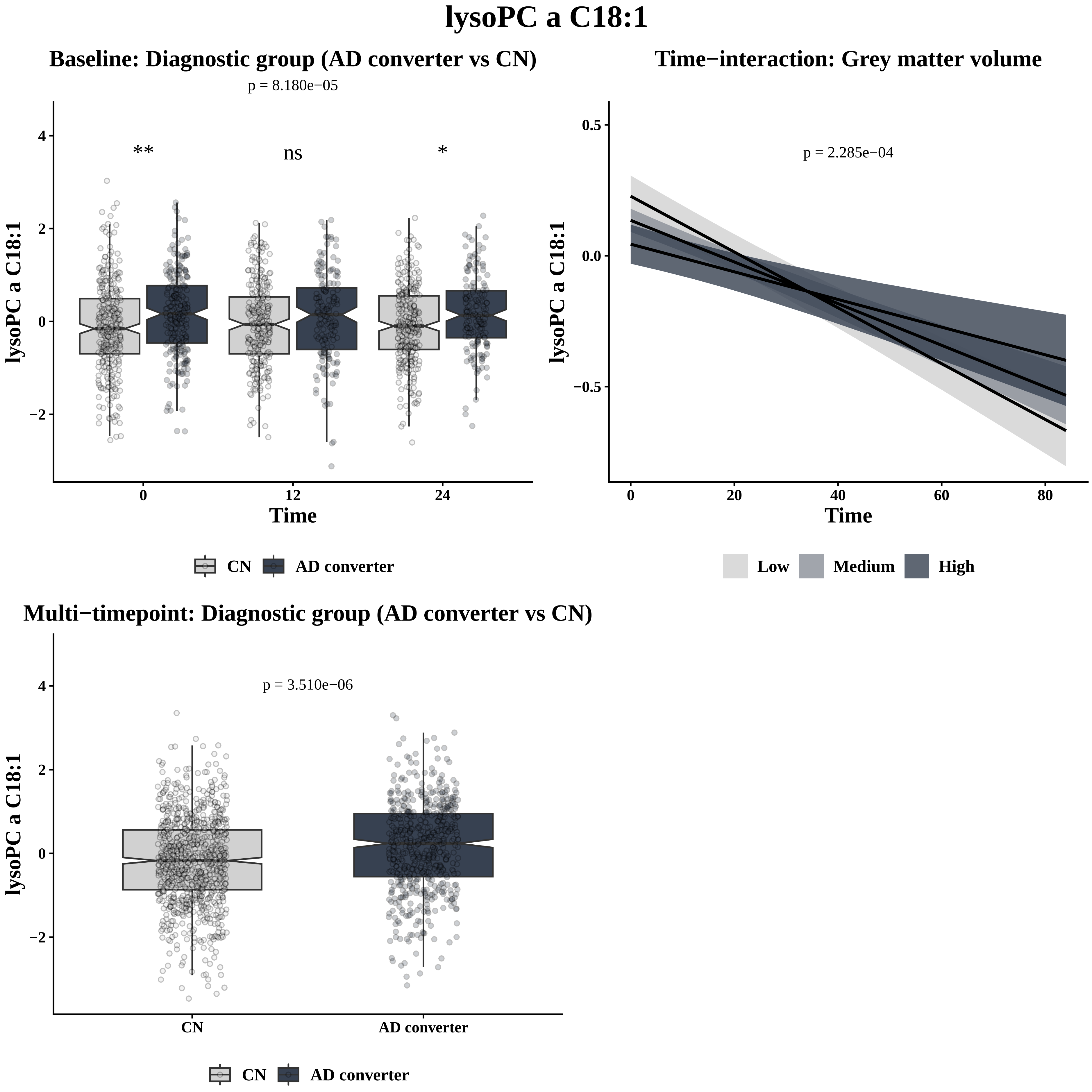
lysoPC a C18:1
Baseline: Diagnostic group (AD converter vs CN)
p = 8.180e−05
**
ns
*
lysoPC a C18:1
4
2
0
−2
0
12
24
Time
CN
AD converter
Time−interaction: Grey matter volume
p = 2.285e−04
lysoPC a C18:1
0.5
0.0
−0.5
0
20
40
60
80
Time
Low
Medium
High
Multi−timepoint: Diagnostic group (AD converter vs CN)
p = 3.510e−06
lysoPC a C18:1
4
2
0
−2
CN
AD converter
CN
AD converter
