## Supplementary Figure S3 for "A seven-year longitudinal study of the Alzheimer’s disease blood metabolome"

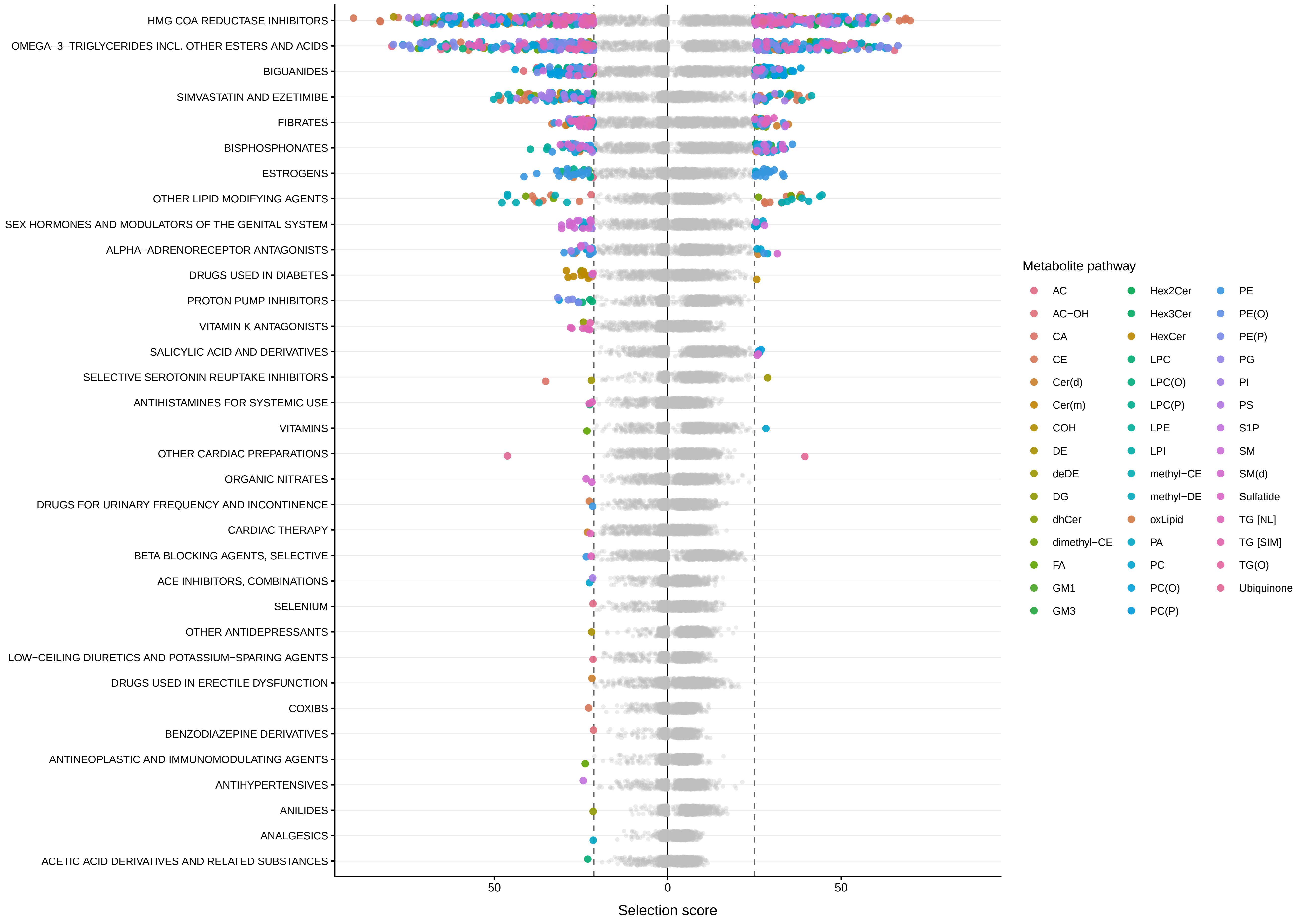
HMG COA REDUCTASE INHIBITORS
OMEGA−3−TRIGLYCERIDES INCL. OTHER ESTERS AND ACIDS
BIGUANIDES
SIMVASTATIN AND EZETIMIBE
FIBRATES
BISPHOSPHONATES
ESTROGENS
OTHER LIPID MODIFYING AGENTS
SEX HORMONES AND MODULATORS OF THE GENITAL SYSTEM
ALPHA−ADRENORECEPTOR ANTAGONISTS
DRUGS USED IN DIABETES
PROTON PUMP INHIBITORS
VITAMIN K ANTAGONISTS
SALICYLIC ACID AND DERIVATIVES
SELECTIVE SEROTONIN REUPTAKE INHIBITORS
ANTIHISTAMINES FOR SYSTEMIC USE
VITAMINS
OTHER CARDIAC PREPARATIONS
ORGANIC NITRATES
DRUGS FOR URINARY FREQUENCY AND INCONTINENCE
CARDIAC THERAPY
BETA BLOCKING AGENTS, SELECTIVE
ACE INHIBITORS, COMBINATIONS
SELENIUM
OTHER ANTIDEPRESSANTS
LOW−CEILING DIURETICS AND POTASSIUM−SPARING AGENTS
DRUGS USED IN ERECTILE DYSFUNCTION
COXIBS
BENZODIAZEPINE DERIVATIVES
ANTINEOPLASTIC AND IMMUNOMODULATING AGENTS
ANTIHYPERTENSIVES
ANILIDES
ANALGESICS
ACETIC ACID DERIVATIVES AND RELATED SUBSTANCES
50
0
50
Selection score
Metabolite pathway
AC
AC−OH
CA
CE
Cer(d)
Cer(m)
COH
DE
deDE
DG
dhCer
dimethyl−CE
FA
GM1
GM3
Hex2Cer
Hex3Cer
HexCer
LPC
LPC(O)
LPC(P)
LPE
LPI
methyl−CE
methyl−DE
oxLipid
PA
PC
PC(O)
PC(P)
PE
PE(O)
PE(P)
PG
PI
PS
S1P
SM
SM(d)
Sulfatide
TG [NL]
TG [SIM]
TG(O)
Ubiquinone

**Supplementary Figure S3: Lipidomics metabolite-medication associations identified by the Boruta random forest algorithm.**

Medication classes are shown on the y-axis and selection scores on the x-axis, with baseline results plotted to the left of zero and longitudinal results to the right. Each point represents a metabolite-medication pair. Selected relationships are colored by metabolite class, whereas unselected relationships are shown in grey. The exact statistics of these associations can be found in **Supplementary Tables S13-S14.**
