## Supplementary Figure S4 for "A seven-year longitudinal study of the Alzheimer’s disease blood metabolome"

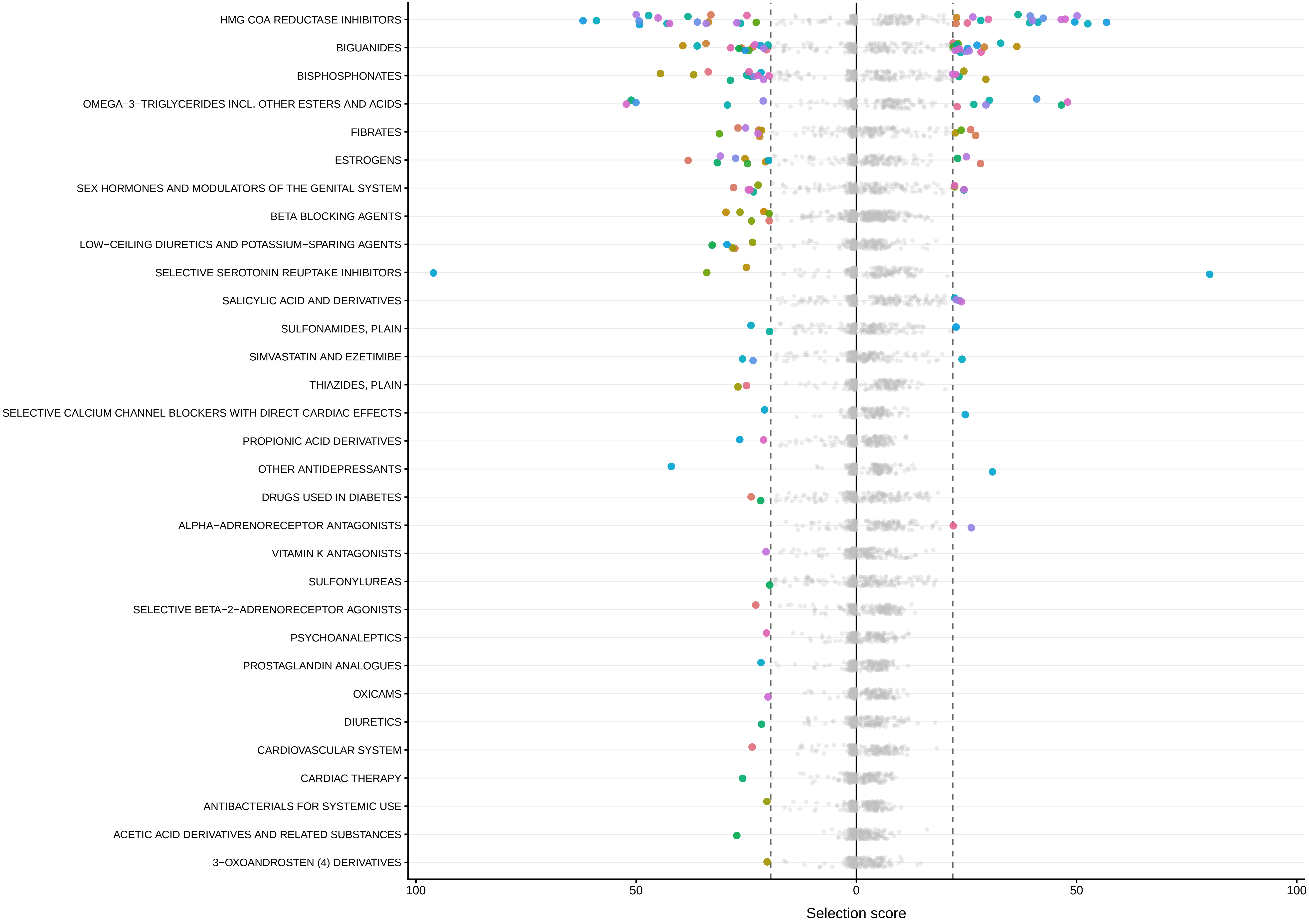

HMG COA REDUCTASE INHIBITORS
BIGUANIDES
BISPHOSPHONATES
OMEGA−3−TRIGLYCERIDES INCL. OTHER ESTERS AND ACIDS
FIBRATES
ESTROGENS
SEX HORMONES AND MODULATORS OF THE GENITAL SYSTEM
BETA BLOCKING AGENTS
LOW−CEILING DIURETICS AND POTASSIUM−SPARING AGENTS
SELECTIVE SEROTONIN REUPTAKE INHIBITORS
SALICYLIC ACID AND DERIVATIVES
SULFONAMIDES, PLAIN
SIMVASTATIN AND EZETIMIBE
THIAZIDES, PLAIN
SELECTIVE CALCIUM CHANNEL BLOCKERS WITH DIRECT CARDIAC EFFECTS
PROPIONIC ACID DERIVATIVES
OTHER ANTIDEPRESSANTS
DRUGS USED IN DIABETES
ALPHA−ADRENORECEPTOR ANTAGONISTS
VITAMIN K ANTAGONISTS
SULFONYLUREAS
SELECTIVE BETA−2−ADRENORECEPTOR AGONISTS
PSYCHOANALEPTICS
PROSTAGLANDIN ANALOGUES
OXICAMS
DIURETICS
CARDIOVASCULAR SYSTEM
CARDIAC THERAPY
ANTIBACTERIALS FOR SYSTEMIC USE
ACETIC ACID DERIVATIVES AND RELATED SUBSTANCES
3−OXOANDROSTEN (4) DERIVATIVES
100
50
0
50
100
Selection score

**Supplementary Figure S4: MetaboINDICATOR™-medication associations identified by the Boruta random forest algorithm.**
Medication classes are shown on the y-axis and selection scores on the x-axis, with baseline results plotted to the left of zero and longitudinal results to the right.
Each point represents a metaboINDICATOR-medication pair. Selected relationships are colored by medication class, whereas unselected relationships are shown in grey.
The exact statistics of these associations can be found in **Supplementary Tables S17-S18.**
